# Associations between obstructive sleep apnea and comorbidities in a large clinical biobank

**DOI:** 10.1101/2025.07.30.25332466

**Authors:** Brian E. Cade, Le Li, Megan Duff, Chen Yang, Syed Moin Hassan, Xinting Yu, Matthew O. Goodman, Raichel M. Alex, Ali Azarbarzin, Salma Batool-Anwar, Farhad R. Nezami, Amin Ramezani, Scott A. Sands, Heming Wang, H. Lester Kirchner, Neomi A. Shah, Lawrence J. Epstein, Milena K. Pavlova, Susan Redline

**Affiliations:** Division of Sleep and Circadian Disorders, Brigham and Women’s Hospital, Boston, MA 02115, USA; Division of Sleep Medicine, Harvard Medical School, Boston, MA 02115, USA; Division of Cardiac Surgery, Brigham and Women’s Hospital and Division of Surgery, Harvard Medical School, Boston, MA 02115, USA; Department of Population Health Sciences, Geisinger Health, Danville, PA 17822, USA; Icahn School of Medicine at Mount Sinai, New York, NY 10029, USA

**Author notes:** Please address all correspondence and requests for reprints to: Brian Cade, Division of Sleep and Circadian Disorders, Brigham and Women’s Hospital, Harvard Medical School 221 Longwood Avenue, Boston, MA 02115, USA, 1-857-307-0353 (phone) (e-mail).

**Keywords:** Sleep-disordered breathing, sleep apnea, polysomnography, multimorbidity

## Abstract

**Background:** Obstructive sleep apnea (OSA) is associated with a wide range of comorbidities, but large-scale phenome-wide analyses in clinical biobanks remain under-reported. In this study, we identified common comorbidities enriched in patients with OSA, tested the temporality of these associations, and analyzed relevant associations with summary sleep recording data.

**Methods:** 48,251 participants with OSA in the Mass General Brigham healthcare system were identified using a natural language processing phenotyping algorithm and/or evidence of an elevated apnea-hypopnea index (AHI). Controls were matched (2:1) on demographics, body mass index (BMI), and healthcare utilization. Associations with 358 incident and 563 cross-sectional diseases were tested using Modified Poisson regression, adjusting for covariates. Sensitivity analyses examined timing by binning data in years relative to the first OSA diagnosis. Selected laboratory results were obtained based on associated diseases. Associated diseases were tested with sleep recording statistics (n ≤18,348).

**Findings:** 179 incident and 421 cross-sectional diseases were associated with OSA at Bonferroni significance. 37 diseases had Bonferroni-significant sex interactions. Several associations were significant years before the first recorded OSA diagnosis. Four red blood cell laboratory measures were significant ten years prior to the first diagnosis. One incident and 47 cross-sectional diseases were associated with the AHI and/or chronic hypoxemia.

**Interpretation:** Obstructive sleep apnea is associated with enrichment of hundreds of diseases, several of which are supported by orthogonal polysomnographic evidence. Leveraging early signs of OSA in clinical data may help to identify at-risk patients.

**Funding:** National Institutes of Health and the American Academy of Sleep Medicine Foundation.

## Research in context

### Evidence before this study

Epidemiological studies have implicated obstructive sleep apnea (OSA) with risk of developing diseases across a broad range of pathophysiology. Several studies used quantitative polysomnography statistics, typically using sample sizes in the hundreds to low thousands. A notable recent study has associated polysomnographic measures to largely self-reported characteristics in 15 body systems.

Recent work has begun to investigate larger clinical biobanks, which requires specialized approaches to appropriately repurpose data collected for clinical care but can enable “phenome-wide” analyses of all common diseases, many of which have never been assessed in cohort studies. We searched PubMed and identified a gap in the literature for phenome-wide analyses that have comprehensively investigated the associations between OSA and common diseases.

### Added value of this study

To our knowledge, this is the largest study of OSA associations with comorbidities to date that is supported by polysomnographic follow-up. We used a natural language processing (NLP) algorithm to improve OSA phenotyping and demonstrated improved positive predictive value in chart reviews and an approximate doubling of OSA genetic heritability when compared to a single ICD diagnosis-based definition. We also performed additional NLP-informed phenotyping on all comorbidities to further improve accuracy. We identified associations between OSA and hundreds of diseases, following rigorous accounting for obesity, healthcare utilization effects, and other covariates using propensity scored matching and further covariate adjustment. Several disorders had significant sex interaction effects. Fifty cross-sectional associations were associated with OSA status 5 years before the first OSA diagnosis at p < 1 × 10^-10^, consistent with delayed clinical recognition of OSA. Several disease associations were supported by polysomnographic analyses.

### Implications of all the available evidence

Collectively, these results increase our understanding of OSA contributions to a broad range of pathophysiology. Individual comorbid associations can be prioritized for future research based on the level of additional evidence. The relative timing of associations with lead diseases suggests signatures of undiagnosed OSA that may help identify at-risk patients.

## Introduction

Obstructive sleep apnea (OSA) is a common, multifactorial disease featuring repetitive upper airway obstruction and chronic intermittent hypoxemia. A wide range of disorders have been shown to be prevalent in patients with OSA, including cardiometabolic, neurocognitive, and other disorders, typically based on data from cohort studies.^1^ While prior research has been mostly based on epidemiological cohort studies, those studies have potential limitations that may preclude their use for obtaining a comprehensive understanding of OSA and associated comorbidities. Notably, prospective research cohort studies practically cannot address hundreds of individual diseases, often rely on self-report data, and often include relatively small proportions of individuals with symptomatic OSA and comorbidities-limiting generalizability to clinic-based samples. Additionally, such studies have variable ascertainment procedures resulting in non nationally-representative samples and only moderate sample sizes that limit power for multiple comparisons with a full spectrum of comorbidities.

Clinical biobanks may complement research cohort studies by providing large-scale, inexpensive data from patients with a broad range of comorbidities. There are separate challenges to analyzing electronic health record (EHR) data however, and multiple steps should be taken to minimize bias when repurposing clinical and billing data for research, including accounting for rule-out diagnoses, biases in healthcare utilization, and open healthcare systems.^2^ Phenotyping algorithms attempting to mitigate these potential biases can be evaluated with chart review, and augmented with orthogonal evidence.

Diseases associated with OSA case-control status can be further evaluated with clinical sleep recording summary statistics from diagnostic, split-night, and/or home sleep tests. Phenome-wide analyses remain underexplored. Properly-interpreted, high-dimensional clinical data have the added potential benefit of producing clinically actionable information, including OSA prediction and risk stratification using clinical data, diagnoses, and models that may be more relevant than those derived from research cohorts.

In this study, we identified a large set of adult participants with OSA in a clinical biobank, leveraging a natural language processing (NLP)-informed phenotyping algorithm, and/or evidence of an elevated AHI. We measured algorithm performance using chart review and genetic heritability analysis. We investigated OSA associations with common-frequency comorbidities using matched controls. Next, recognizing that OSA diagnosis is often delayed, we assessed the relative timing of the first diagnoses of OSA relative to the potential causes and consequences of OSA.^3^ We further queried whether comorbidities associated with case/control OSA status were also associated with quantitative metrics from sleep studies in a smaller set of participants with available clinical sleep study recordings. This study extends our prior report by considering an order of magnitude more participants.^4^

## Methods

### Participant selection and OSA phenotyping

Participant data from adults aged >18 were obtained from the Mass General Brigham (MGB) Research Patient Data Registry (RPDR), a clinical data warehouse of patients from 10 hospitals in the greater Boston, MA, USA region.^5^ This study received ethical approval (MGB protocols 2024P002360, 2021P003546, and 2010P001765). Case-control study data were censored on March 6, 2023, apart from data from participants with genetic and/or sleep study recordings, which were censored on January 9, 2025. Clinical data included date-stamped ICD-9 and ICD-10 based diagnoses grouped into PheCodes^6^ and clinical notes. Potential cases and controls were selected by having either ≥1 or 0 PheCode counts for 327·3 (sleep apnea) and/or 327·32 (obstructive sleep apnea), respectively. Data from patients with diagnostic, split-night, and home sleep recordings interpreted at Brigham and Women’s Faulkner Hospital (BWFH) were also used, irrespective of OSA diagnoses and prioritizing the first available diagnostic, split-night, or home sleep test, respectively. Genotype chip and whole-genome sequencing (WGS) data were generated by the MGB Biobank.^7^ Patients without available covariates (*e.g.* BMI) or a minimal amount of healthcare utilization (“data floor”; ≥3 diagnoses, ≥3 clinical encounters, and ≥3 notes) were excluded.

We used multiple techniques to improve the selection of patients with true OSA among our initial pool of diagnosed participants. The PheVis algorithm, which combines diagnostic codes and clinical note terms to improve true case selection^8^, identified the bulk (89.7%) of our cases. Similar to our previous algorithm^4^, note terms were extracted using cTAKES^9^, coded using the UMLS ontology^10^, and subsetted to generalizable terms describing OSA using the SAFE procedure and term presence in a majority of seven online OSA articles (**Table S1**). Clinical chart reviews of 484 participants performed by sleep clinicians evaluated ICSD-3 defined OSA criteria^4^ to measure PheVis performance. Participants with an AHI ≥15 via sleep study reports or regular expression-based clinical note extraction (excluding machine-based or stage-specific data) were also set to true cases. Age was calculated using the first OSA diagnosis or sleep study date, while BMI was calculated using the two values closest to this age.

OSA controls were matched to cases using propensity score matching. Matching considered birth date (±5 years), BMI (±5 units), self-reported population and number of clinical encounters^11^, conducted separately by sex at birth. We used 2× controls for each case, with each control participant’s age set to the corresponding case age. While matching for clinical encounter counts help to account for potential healthcare utilization biases (*e.g.* receiving care at outside hospitals), they could also introduce biases by selecting controls enriched with encounters linked to specific PheCodes, as prior work adjusting for clinical encounter counts has demonstrated improved chart review-measured performance but some inverse associations.^11^ We therefore performed a sensitivity analysis using propensity score matching without clinical encounters (but still adjusted for encounters in the analyses) in order to flag disease associations with opposite effect direction relative to the primary analysis.

### Secondary phenotype data

ICD-9 and ICD-10 comorbidities were consolidated into PheCodes and refined using the Multimodal Automated Phenotyping NLP algorithm, which often improves positive predictive value relative to diagnostic codes alone.^12^ Cross-sectional analyses considered all first diagnosis dates, while incident analyses required a first diagnosis ≥1 year after the first OSA diagnosis (or the sleep recording for continuous trait analyses). Analyses required a minimum comorbidity prevalence or incidence of 1%.

Sleep recording statistics considered AHI 3% (i.e., all apneas plus hypopneas requiring ≥3% desaturation) and the percentage of sleep time with oxyhemoglobin saturation <88% (Per88). Summary statistics were extracted from clinic reports. In Per88 values not present for a subset of HST summary reports, and <0·5% of HSTs used 89% saturation thresholds. Per88 was included due to prior reports of more significant associations with chronic hypoxemia compared to the AHI (*e.g.* ^13,14^).

### Statistical analyses

Modified Poisson regression analyses with the rqlm R package^19^ modeled each comorbidity as an outcome with the primary exposure OSA status (or a sleep recording variable, transformed using rank-normalization), adjusting for age, sex, BMI, race and ethnicity, and number of clinical encounters^11^.

Analyses of sleep recording outcomes further adjusted for sleep study type. Potential effect moderation by sex was tested in separate models by including product terms for sex × OSA status (or sleep recording variables, without rank normalization) and reporting interaction p-values. Sensitivity analyses repeated the case-control analyses after censoring data at cut-points from -5 to +5 years relative to the first OSA diagnosis date. An exploratory analysis to identify signatures of undiagnosed OSA used the EHRtemporalVariability R package, a method designed to detect temporal shifts in EHR data patterns^15^, which does not consider covariates.

Genetic heritability analyses are described in the Supplementary Methods.

## Results

### Study sample

An overview of the participant groups is provided in **Table 1**. Collectively there were 48,251 cases and 96,502 matched controls in the main sample, 18,348 participants in the sleep recording sample, and 56,082 participants in the heritability sample. While birth date and, notably, BMI distributions were similar (**Figures S1 and S2**), we further adjusted for covariates in all applicable analyses. At least one OSA PheCode diagnosis was observed in 58·6% of the sleep recording sample. Among these participants, 26·6% had their first OSA diagnosis at least one year before the sleep study date, and 1·0% had their first OSA diagnosis recorded at least one year after the sleep study date. 5 An AHI >15 was found in 51·3% of the sleep recording sample.

**Table 1.**
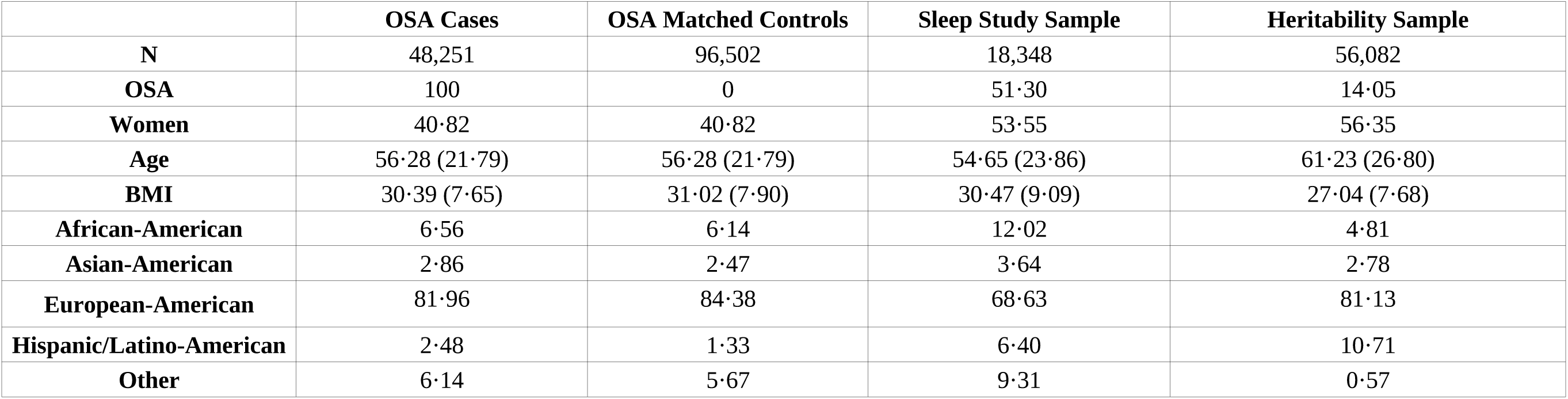
Sample description. Values are presented as counts (N), medians and interquartile ranges (Age and BMI), or otherwise as percentages. Populations are based on self-report, except for the heritability sample which uses inferred 1000 Genomes continental ancestry. The sleep study sample is comprised of diagnostic recordings (38%), split recordings (8%), and home sleep tests (53%), with the first available recording prioritized first for diagnostic then split then home sleep test recordings. OSA status is based on AHI > 15 for the sleep study sample or a combination of PheVis NLP phenotyping and AHI > 15 for the main case and heritability samples. Populations are by self-report apart from the heritability sample, which uses genetically-determined continental ancestry based on the 1000 Genomes Project (1·74% East Asians, 1·04% South Asians).

### Phenotyping algorithm performance and heritability

The NLP phenotyping algorithm, designed to identify high-confidence cases, had improved positive predictive value in 484 chart reviews compared to using one or more PheCodes to define cases (0·876 vs. 0·689). When including additional AHI and CPAP information (available only for patients seen at the BWHF sleep clinic), the positive predictive value ranged from 0·873 – 0·874 (**Table S2**). OSA heritability estimates adjusted for BMI and other covariates were nearly doubled using PheVis compared to using ≥1 PheCode to define cases among European continental ancestry participants (0·097 [0·027] vs. 0·054 [0·011]). Consideration of known, extracted AHI values further increased the pointwise estimate (0·101 [0·027]). After inclusion of AHI information after the PheVis censoring date, the European ancestry heritability estimate was 0·105 (0·027), while the combined sample heritability across all ancestries was 0·130 (0·027). Equivalent estimates in American ancestry participants identified significant heritability, while an unbounded heritability estimate was not significant in African ancestry participants, although both of these analyses were underpowered as evidenced by elevated standard errors (**Table S3**).

### Case-control comorbidity associations

Prospective analyses considered diseases first diagnosed at least one year after the first OSA diagnosis (or matched age for controls). We identified 223 diseases associated with OSA status at Bonferroni significance in combined-sex analyses. We performed a sensitivity analysis where healthcare utilization was not used for control matching (but was still adjusted for in the analysis) in order to identify potential artificial associations with common PheCodes. When considering concordant directionality and nominal significance in the sensitivity analysis, there were 179 higher-confidence PheCodes associated at Bonferroni significance (**Tables 2, S4;** lead results chronic pharyngitis and nasopharyngitis, major depressive disorder, hemorrhoids; p <1·3 × 10^-176^). Full results, including sex-stratified results, are provided in **Table S4**. Six diseases had Bonferroni-significant OSA × sex interaction p-values (lead diseases [sex] with p <1·9 × 10^-5^: substance addiction and disorders [women], morbid obesity [men], synovitis and tenosynovitis [men]).

We performed an equivalent analysis using cross-sectional diagnoses and identified 421 higher-confidence PheCodes associated with OSA at Bonferroni significance in combined-sex analyses (**Table S5, Table 3**; lead results sorted by risk ratios: sleep disorders, chronic pharyngitis and nasopharyngitis, restless legs syndrome p <1·1 × 10^-293^). Multiple associations (e.g. with obesity, essential hypertension, and Type 2 diabetes) are consistent with the literature regarding known OSA comorbidities. Thirty-seven disorders had Bonferroni-significant OSA × sex interaction p-values (lead diseases [sex] with p <2·6 × 10^-22^: allergic rhinitis [men], benign neoplasm of skin [men], chronic sinusitis [men]).

**Table 2.** Lead incident OSA case-control disease associations. An incident diagnosis was defined as the first diagnosis for a potential comorbidity occurring at least one year after the first diagnosis date for OSA. Otherwise, participants with prior diagnoses were excluded. These modified Poisson regression analyses using propensity-matched controls and adjusted for age, sex, BMI, population, and healthcare encounters from age 18 to the first OSA diagnosis (or the equivalent age for controls). 179 combined-sex PheCodes were significantly associated following Bonferroni correction with concordant directionality in sensitivity analyses where controls were not matched for healthcare utilization. Complete results can be found in Table S4.

| PheCode | Disease | OSA Case Incidence | OSA Control Incidence | Risk Ratio | P-value |
| --- | --- | --- | --- | --- | --- |
| 472 | Chronic pharyngitis and nasopharyngitis | 3.33 | 0.54 | 6.19 (5.55 – 6.90) | $3.05 \times 10^{-236}$ |
| 296.22 | Major depressive disorder | 9.87 | 5.61 | 1.87 (1.79 – 1.95) | $2.56 \times 10^{-179}$ |
| 455 | Hemorrhoids | 7.00 | 3.14 | 2.20 (2.09 – 2.33) | $1.23 \times 10^{-176}$ |
| 327 | Sleep disorders | 2.39 | 0.41 | 5.90 (5.20 – 6.70) | $8.71 \times 10^{-166}$ |
| 689 | Disorder of skin and subcutaneous tissue NOS | 6.78 | 3.35 | 1.99 (1.88 – 2.09) | $2.90 \times 10^{-140}$ |
| 327.71 | Restless legs syndrome | 2.14 | 0.48 | 4.50 (4.00 – 5.07) | $5.65 \times 10^{-136}$ |
| 470 | Septal Deviations/Turbinate Hypertrophy | 3.19 | 1.15 | 2.86 (2.63 – 3.11) | $9.43 \times 10^{-129}$ |
| 476 | Allergic rhinitis | 6.87 | 3.67 | 1.93 (1.82 – 2.03) | $7.16 \times 10^{-126}$ |
| 327.4 | Insomnia | 5.40 | 2.92 | 1.83 (1.73 – 1.94) | $1.58 \times 10^{-89}$ |
| 289.9 | Abnormality of red blood cells | 1.07 | 0.14 | 7.36 (6.00 – 9.02) | $4.03 \times 10^{-82}$ |
| 272.1 | Hyperlipidemia | 12.78 | 9.38 | 1.48 (1.42 – 1.54) | $2.31 \times 10^{-76}$ |
| 278 | Overweight, obesity and other hyperalimentation | 6.74 | 4.12 | 1.60 (1.52 – 1.69) | $4.03 \times 10^{-72}$ |
| 292 | Neurological disorders | 1.50 | 0.46 | 3.12 (2.74 – 3.54) | $2.04 \times 10^{-68}$ |
| 196 | Radiotherapy | 2.31 | 4.15 | 0.54 (0.51 – 0.58) | $9.40 \times 10^{-66}$ |
| 379.9 | Pain, swelling or discharge of eye | 1.24 | 0.35 | 3.46 (3.00 – 4.00) | $4.03 \times 10^{-64}$ |
| 292.3 | Memory loss | 3.14 | 1.54 | 1.93 (1.78 – 2.08) | $1.26 \times 10^{-62}$ |
| 379 | Other disorders of eye | 1.09 | 0.03 | 41.54 (26.81 – 64.35) | $1.59 \times 10^{-62}$ |
| 300.11 | Generalized anxiety disorder | 4.82 | 3.09 | 1.62 (1.53 – 1.72) | $1.11 \times 10^{-60}$ |
| 475 | Chronic sinusitis | 2.91 | 1.58 | 1.90 (1.75 – 2.07) | $3.24 \times 10^{-54}$ |
| 550.2 | Diaphragmatic hernia | 2.31 | 1.14 | 2.05 (1.87 – 2.25) | $5.97 \times 10^{-54}$ |
| 306 | Other mental disorder | 4.20 | 2.58 | 1.63 (1.53 – 1.74) | $2.14 \times 10^{-53}$ |
| 278.1 | Obesity | 9.17 | 8.41 | 1.41 (1.35 – 1.48) | $1.05 \times 10^{-52}$ |
| 313.1 | Attention deficit hyperactivity disorder | 1.53 | 0.75 | 2.30 (2.06 – 2.56) | $1.07 \times 10^{-50}$ |
| 208 | Benign neoplasm of colon | 6.99 | 4.70 | 1.47 (1.39 – 1.54) | $1.56 \times 10^{-49}$ |
| 327.41 | Organic or persistent insomnia | 1.55 | 0.65 | 2.32 (2.07 – 2.60) | $1.45 \times 10^{-47}$ |

**Table 3.** Lead cross-sectional OSA case-control disease associations. These modified Poisson regression analyses using propensity-matched controls and adjusted for age, sex, BMI, population, and healthcare encounters from age 18 to the first OSA diagnosis (or the equivalent age for controls). 421 combined-sex PheCodes were significantly associated following Bonferroni correction with concordant directionality in sensitivity analyses where controls were not matched for healthcare utilization. Complete results can be found in Table S5.

| PheCode | Disease | OSA Case Prevalence | OSA Control Prevalence | Risk Ratio | P-value |
| --- | --- | --- | --- | --- | --- |
| 327 | Sleep disorders | 6.05 | 0.82 | 7.19 (6.65 – 7.77) | $<1.04 \times 10^{-293}$ |
| 472 | Chronic pharyngitis and nasopharyngitis | 7.71 | 1.10 | 6.87 (6.42 – 7.35) | $<1.04 \times 10^{-293}$ |
| 327.71 | Restless legs syndrome | 4.34 | 0.69 | 6.27 (5.75 – 6.84) | $<1.04 \times 10^{-293}$ |
| 474.2 | Chronic tonsillitis and adenoiditis | 4.52 | 0.75 | 5.56 (5.11 – 6.06) | $<1.04 \times 10^{-293}$ |
| 470 | Septal Deviations/Turbinate Hypertrophy | 9.90 | 1.97 | 4.90 (4.66 – 5.17) | $<1.04 \times 10^{-293}$ |
| 327.4 | Insomnia | 12.26 | 4.42 | 2.71 (2.61 – 2.81) | $<1.04 \times 10^{-293}$ |
| 296.22 | Major depressive disorder | 20.16 | 9.03 | 2.24 (2.18 – 2.30) | $<1.04 \times 10^{-293}$ |
| 455 | Hemorrhoids | 12.44 | 5.82 | 2.10 (2.03 – 2.18) | $<1.04 \times 10^{-293}$ |
| 476 | Allergic rhinitis | 17.63 | 8.35 | 2.07 (2.01 – 2.13) | $<1.04 \times 10^{-293}$ |
| 689 | Disorder of skin and subcutaneous tissue NOS | 11.25 | 5.42 | 2.06 (1.99 – 2.14) | $<1.04 \times 10^{-293}$ |
| 495 | Asthma | 16.86 | 8.77 | 1.92 (1.87 – 1.98) | $<1.04 \times 10^{-293}$ |
| 244.4 | Hypothyroidism NOS | 12.49 | 6.58 | 1.92 (1.86 – 1.98) | $<1.04 \times 10^{-293}$ |
| 530.11 | GERD | 29.04 | 15.19 | 1.91 (1.87 – 1.95) | $<1.04 \times 10^{-293}$ |
| 272.1 | Hyperlipidemia | 34.31 | 18.20 | 1.90 (1.86 – 1.93) | $<1.04 \times 10^{-293}$ |
| 300.1 | Anxiety disorder | 20.79 | 11.69 | 1.78 (1.74 – 1.83) | $<1.04 \times 10^{-293}$ |
| 278.1 | Obesity | 25.56 | 15.14 | 1.77 (1.73 – 1.80) | $<1.04 \times 10^{-293}$ |
| 272.11 | Hypercholesterolemia | 21.27 | 13.49 | 1.58 (1.55 – 1.62) | $<1.04 \times 10^{-293}$ |
| 401.1 | Essential hypertension | 43.16 | 30.60 | 1.43 (1.41 – 1.45) | $<1.04 \times 10^{-293}$ |
| 512.9 | Other dyspnea | 15.81 | 9.24 | 1.72 (1.67 – 1.77) | $1.04 \times 10^{-293}$ |
| 427.21 | Atrial fibrillation | 12.01 | 6.70 | 1.83 (1.77 – 1.89) | $1.48 \times 10^{-291}$ |
| 278 | Overweight, obesity and other hyperalimentation | 12.53 | 6.56 | 1.87 (1.81 – 1.93) | $7.60 \times 10^{-288}$ |
| 475 | Chronic sinusitis | 8.21 | 3.60 | 2.26 (2.16 – 2.36) | $8.88 \times 10^{-284}$ |
| 250.2 | Type 2 diabetes | 16.52 | 10.75 | 1.56 (1.52 – 1.60) | $3.09 \times 10^{-237}$ |
| 208 | Benign neoplasm of colon | 14.80 | 9.13 | 1.62 (1.57 – 1.67) | $5.92 \times 10^{-233}$ |
| 300.11 | Generalized anxiety disorder | 8.92 | 4.51 | 1.96 (1.89 – 2.05) | $6.82 \times 10^{-230}$ |

**Table 4.** Lead cross-sectional polysomnographic disease associations. Modified Poisson regression analyses adjusted for age, sex, BMI, population, PSG study type, and healthcare encounters using rank-normalization. 47 PheCodes were significantly associated following Bonferroni correction, while 117 PheCodes were significant when applying less stringent false discovery rate criteria. Sensitivity analyses considered only participants with an AHI > 15. Complete results, including additional sensitivity analyses considering sleep and pulmonary disease, are contained in Table S11 and S12.

| PheCode | Disease | PSG Phenotype | Sleep Clinic Prevalence | Risk Ratio | P-value | Risk Ratio (AHI >15) |
| --- | --- | --- | --- | --- | --- | --- |
| 1013 | Asphyxia and hypoxemia | Per88 | 3·84 | 1·07 (1·06 – 1·07) | $1·11 \times 10^{-70}$ | 1·05 (1·04 – 1·06) |
| 496 | Chronic airway obstruction | Per88 | 3·90 | 1·06 (1·05 – 1·06) | $3·19 \times 10^{-45}$ | 1·04 (1·03 – 1·05) |
| 509·1 | Respiratory failure | Per88 | 2·30 | 1·07 (1·06 – 1·08) | $7·01 \times 10^{-40}$ | 1·05 (1·04 – 1·06) |
| 415·2 | Chronic pulmonary heart disease | Per88 | 2·12 | 1·07 (1·05 – 1·08) | $5·48 \times 10^{-38}$ | 1·05 (1·04 – 1·07) |
| 496·21 | Obstructive chronic bronchitis | Per88 | 1·33 | 1·07 (1·06 – 1·09) | $2·36 \times 10^{-34}$ | 1·06 (1·05 – 1·08) |
| 508 | Pulmonary collapse; interstitial and compensatory emphysema | Per88 | 5·29 | 1·04 (1·04 – 1·05) | $3·89 \times 10^{-34}$ | 1·03 (1·02 – 1·04) |
| 503 | Pulmonary congestion and hypostasis | Per88 | 2·95 | 1·06 (1·05 – 1·07) | $6·81 \times 10^{-31}$ | 1·04 (1·03 – 1·05) |
| 496·1 | Emphysema | Per88 | 2·04 | 1·06 (1·05 – 1·07) | $1·14 \times 10^{-30}$ | 1·05 (1·03 – 1·06) |
| 512·7 | Shortness of breath | Per88 | 17·10 | 1·02 (1·02 – 1·03) | $8·56 \times 10^{-26}$ | 1·02 (1·01 – 1·02) |
| 782·3 | Edema | Per88 | 9·48 | 1·03 (1·03 – 1·04) | $6·69 \times 10^{-25}$ | 1·03 (1·02 – 1·03) |
| 428·1 | Congestive heart failure NOS | Per88 | 5·59 | 1·04 (1·03 – 1·05) | $1·25 \times 10^{-21}$ | 1·03 (1·02 – 1·04) |
| 401·1 | Essential hypertension | AHI 3% | 41·76 | 1·00 (1·00 – 1·01) | $7·04 \times 10^{-20}$ | 1·00 (1·00 – 1·00) |
| 512·9 | Other dyspnea | Per88 | 18·42 | 1·02 (1·01 – 1·02) | $5·79 \times 10^{-18}$ | 1·01 (1·01 – 1·02) |
| 428·4 | Heart failure with preserved EF [Diastolic heart failure] | Per88 | 2·41 | 1·05 (1·04 – 1·06) | $2·01 \times 10^{-17}$ | 1·04 (1·03 – 1·05) |
| 480 | Pneumonia | Per88 | 7·85 | 1·03 (1·02 – 1·03) | $5·63 \times 10^{-17}$ | 1·02 (1·01 – 1·03) |
| 505 | Other pulmonary inflammation or edema | Per88 | 1·29 | 1·06 (1·04 – 1·07) | $2·01 \times 10^{-16}$ | 1·04 (1·02 – 1·05) |
| 510 | Other diseases of lung | Per88 | 3·74 | 1·04 (1·03 – 1·04) | $2·04 \times 10^{-15}$ | 1·03 (1·02 – 1·04) |
| 428·2 | Heart failure NOS | Per88 | 2·26 | 1·05 (1·03 – 1·06) | $1·20 \times 10^{-14}$ | 1·03 (1·02 – 1·05) |
| 276·6 | Fluid overload | Per88 | 1·94 | 1·05 (1·04 – 1·06) | $1·27 \times 10^{-14}$ | 1·03 (1·02 – 1·04) |
| 340 | Migraine | AHI 3% | 13·31 | 0·99 (0·99 – 0·99) | $5·60 \times 10^{-14}$ | 1·00 (0·99 – 1·00) |
| 401·21 | Hypertensive heart disease | Per88 | 2·47 | 1·04 (1·03 – 1·06) | $6·22 \times 10^{-13}$ | 1·03 (1·02 – 1·04) |
| 805 | Fracture of vertebral column without mention of spinal cord injury | Per88 | 1·52 | 1·05 (1·03 – 1·06) | $1·16 \times 10^{-12}$ | 1·04 (1·02 – 1·05) |
| 458·9 | Hypotension NOS | Per88 | 3·64 | 1·03 (1·02 – 1·04) | $1·47 \times 10^{-12}$ | 1·02 (1·01 – 1·03) |

**Cade *et al.*: Associations between sleep apnea and comorbidities in a large clinical biobank**
| **PheCode** | **Disease** | **PSG Phenotype** | **Sleep Clinic Prevalence** | **Risk Ratio** | **P-value** | **Risk Ratio (AHI >15)** |
| --- | --- | --- | --- | --- | --- | --- |
| 585·1 | Acute renal failure | Per88 | 4·81 | 1·03 (1·02 – 1·04) | $3·63 \times 10^{-10}$ | 1·02 (1·01 – 1·03) |
| 427·7 | Tachycardia NOS | Per88 | 7·35 | 1·02 (1·01 – 1·03) | $5·57 \times 10^{-10}$ | 1·01 (1·01 – 1·02) |

### Temporal patterns of comorbidities, selected laboratory results, and sleep-related note terms

We examined the strength of the cross-sectional associations by repeating the analyses using data censored at different intervals relative to the first OSA diagnosis date (or the equivalent matching control age) and year-specific covariates (**Table S6, Figure 1**). Fifty diseases were associated with OSA status 5 years before the first OSA diagnosis at p < 1 × 10^-10^ (lead results at -5 years: hyperlipidemia, chronic pharyngitis and nasopharyngitis, and GERD; p <1·7 × 10^-61^). Given early associations with hyperlipidemia and abnormality of red blood cells, we examined median lab values in relative years, adjusting for covariates (**Figure S3, Table S7**). Values were available from a fraction of the sample in a given year. HDL cholesterol associations became significant seven years prior to diagnosis (p = 0.004). Red cell distribution width, hematocrit, hemoglobin, and red blood cell count were significantly associated ten years prior to the first OSA diagnosis (p <9.2 × 10^-19^).

**Figure 1.**
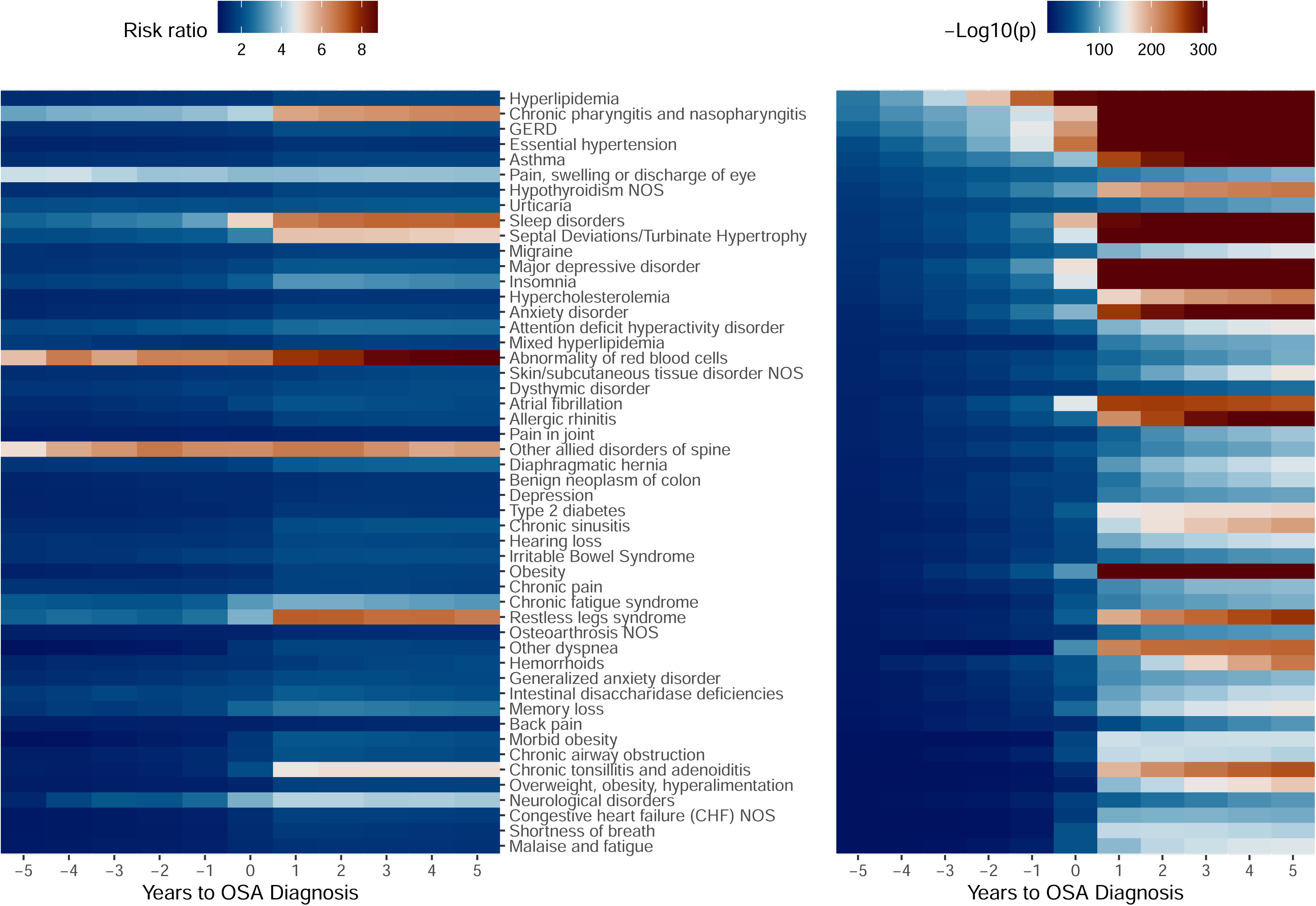
Temporal patterns of associations between sleep apnea and the 50 lead diseases. The lead 50 cross-sectional associations were further tested in a sensitivity analysis censoring data a number of years away from the first sleep apnea results (or the matching age for controls). Future comorbidity cases were classified as controls for a year prior to the initial comorbidity diagnosis. Analyses were adjusted for age, sex, BMI, population, and healthcare encounters since age 18 to a given relative year. Age was set to the relative year for controls or a prior year for cases with an earlier diagnosis. BMIs were based on measurements relative to a given age. The left panel displays risk ratio point estimates, while the right panel displays the negative log values of association p-values. Rows are ordered based on the strength of the association five years prior to the first OSA diagnosis date. Further details are provided in Table S6.

We performed an exploratory analysis to identify EHR data changes in diagnosis and clinical note sleep term patterns that may be associated with undiagnosed OSA using a method designed to detect temporal shifts in EHR data.^15,16^ We examined all 421 and the lead 50 cross-sectionally associated PheCodes as well as 46 sleep-related NLP clinical note terms (**Table S8**), orienting all dates around a relative first OSA diagnosis date of 1/1/2000. Diagnosis and sleep term patterns typically changed by relative year as participants aged, however all three datasets had proportionally large temporal shifts two years prior to the first OSA diagnosis, consistent with potential two-year signatures of undiagnosed OSA (**Figures S4-S6**). Multiple sleep-related terms typically had relatively more occurrences in notes from the years prior to the OSA diagnosis, including “insomnia” (**Figure S7**). Equivalent analyses in the matched controls are presented in **Figures S8-S11**.

### Sleep recording comorbidity associations

Diseases significantly associated in case-control analyses were carried forward for analyses in a sample of participants seen at the BWFH sleep clinic (n up to 18,348). We identified one disease associated with either AHI 3% or Per88 at Bonferroni significance in prospective analyses (**Tables S9-S10**; abnormal involuntary movements / AHI 3% p = 2·0 × 10^-4^). In contrast, 47 PheCodes were significantly associated in cross-sectional analyses (**Tables 4, S11, S12**; lead results were with Per88: asphyxia and hypoxemia, chronic airway obstruction, and respiratory failure; p <7·1 × 10^-40^). 117 PheCodes were associated when using false discovery rate criteria. PSG statistics had protective associations with ten PheCodes, most of which were partially or completely attenuated when considering participants with AHI 3% >15. Notable attenuated but still negative associations remained for migraine and tinnitus (p <2·2 × 10^-4^).

## Discussion

In this study, we used a clinical biobank and clinical sleep laboratory data to identify associations between OSA and comorbidities in what is to our knowledge the largest phenome-wide analysis of clinically-diagnosed OSA and comorbidities to date. We identified associations between several of these diseases and sleep recording summary statistics and examined temporal trends of comorbidity diagnoses. Several lead associations (*e.g.* with obesity, essential hypertension, and Type 2 diabetes) were consistent with the literature regarding known OSA comorbidities. Our analyses further extend the literature by providing new information on the timing of clinical recognition, which for certain comorbidities can occur years before the first OSA diagnosis date and thus represent markers of risk that might be used to identify people at risk for OSA (Figure 1, Table S6). We also identified 37 Bonferroni-significant conditions that differed by sex in cross-sectional analyses (Table S5) and dozens of associations between summary sleep recording statistics and comorbidities (Tables 5 and S11). This work highlights the complex associations of OSA with a broad range of pathophysiology and identifies a potential means of identifying patients for testing earlier using data available in the EHR.

We identified a wide range of associations (Tables 2, 3, S4, and S5), consistent with diverse pathophysiology associated in prior reports^1^ and a recent report linking PSG measures to largely self-reported characteristics in 15 body systems.^17^ Here we extended OSA associations to over 400 clinician-diagnosed diseases using a relatively large sample size (n = 144,753). Among the 36 PheCodes with Bonferroni-significant sex × OSA interactions, women had stronger associations for respiratory failure, chronic pulmonary heart disease, bariatric surgery, and five mental health and substance disorders. Men had stronger associations for three obesity-related and seven airway inflammation-related disorders. Certain PheCodes may have indirect relationships with OSA (*e.g.* via systemic inflammation). A small number of significantly associated PheCodes have inverse associations, including multiple cancers and cancer treatments, which while consistent could conceivably reflect survival or other bias.

Multiple reports have identified underdiagnosis or delayed diagnosis of OSA (the latter resulting in high disease severity at the time of diagnosis); both patterns may result from multiple factors related to recognition or action by patients and clinicians.^3,18,19^ Fifty cross-sectional associations were associated with OSA status 5 years before the first OSA diagnosis at p <1 × 10^-10^ (Table S6, Figure 1). Particularly significant early associations may induce changes in incident association directionality (e.g. for essential hypertension, Table S4). Interestingly, the initial associations with individual comorbidities occurred at different relative dates, which may reflect actual temporal patterns of disease occurrences, as well as be influenced by, differential recognition by clinicians, and/or alignment of individual comorbidities with a classic but incomplete understanding of OSA symptom presentation. While exact temporality is difficult to assess with this study design, our prior epidemiological and genetic associations with cholesterol and lipid synthesis^20,21^ suggest that metabolic abnormalities may precede the occurrence of OSA by five or more years. Similarly, while cross-sectional associations between OSA and hemoglobin are reported, associations ten years prior to the first OSA diagnosis and our previous genetic associations with hexokinase (involved with both metabolism and hemoglobin) suggest potential causal effects.^23–25^ The temporality of these effects may provide a time window for potential prevention.

An exploratory analysis suggests that multiple potential signatures of undiagnosed OSA are detectable in the EHR for two years (Figures S4-S6). Associated diagnosis and sleep-related clinical note concept coding patterns changed gradually as the participants with OSA aged, similar to the controls (Figures S8-S10), apart from the 2-year relative span to the first OSA diagnosis from 1998 - 2000. This period of increased multimorbidity may impact sleep clinic referrals prior to eventual diagnosis. While additional work is required to confirm whether this time window generalizes to other hospitals, this analysis highlights a means of identifying patients for prioritized OSA screening with minimal burden using clinical databases.

Certain sleep-related symptoms and signs found in the clinical notes displayed temporal trends relative to the initial year of OSA diagnosis (Table S8, Figure S7). While “obstructive sleep apnea” and “sleep apnea syndrome” are most frequently found in the notes from the year of diagnosis onward (2000), these terms were also identified in prior years. Other concepts have opposite temporal shifts, including “fatigue”, “insomnia”, “awake”, and “tired”, which decrease near the initial diagnosis year and may indicate a general awareness of disordered sleep symptoms that are not unique to OSA. These results suggest that improved patient recognition of OSA-specific symptoms (as reflected in clinical notes) may affect the time to diagnosis.

An additional analysis of participants with sleep recordings identified 47 diseases cross-sectionally associated with the AHI and/or chronic hypoxemia (Per88), despite a reduced sample size (Tables 4, S11). While power was limited, using false discovery rate criteria we identified 117 cross-sectional and one incident disease association. The latter also may reflect limited follow-up time, unavailable early recordings (27% had an OSA diagnosis over a year before their recording), and/or late sleep clinic referrals (Figures S4, S6). Inverse associations were observed for migraine, consistent with our pilot analysis^4^ and largely driven by women. These results were attenuated but not eliminated when considering participants with AHI >15, highlighting a need to better understand the effects of OSA comorbid with insomnia and/or other sleep disorders. Multiple studies have identified more significant associations using chronic hypoxemia in place of the AHI (*e.g.* ^4,13,14^). While adjustment for overt lung disease typically modestly attenuated our associations (Table S12), these effects could be partially due to associations between OSA and subclinical interstitial lung abnormalities.^25^

Multiple measures were used to mitigate potential biases present in EHR analyses.^2^ We used a NLP-informed phenotyping algorithm to combine ICD diagnoses and commonly accepted descriptors of OSA (Table S1) to improve chart review precision (Table S2) and nearly double the estimated heritability. Comorbidity phenotyping also used a NLP-informed algorithm with demonstrated improvements for multiple diseases.^12^ A data floor was used to reduce the impact of open healthcare systems. While adjusting for healthcare encounters is an important consideration in matched control analyses^11^, we used a sensitivity analysis without encounter count matching to identify potential artificial associations. Propensity score matching demonstrated reasonable concordance for birth year and notably BMI (Figures S1 and S2) and minimized the impact of changes in healthcare delivery over time and the independent effects of obesity on multimorbidity.

Future work is required to understand the potential impact of continuous positive airway pressure and other treatments on disease associations and to replicate our results in outside samples. While laboratory results had non-random ascertainment, results were restricted to commonly-ordered complete blood counts and lipid measures that were averaged into yearly bins and had concordant results with equivalent PheCodes. Understanding and accounting for bias in real-world EHR databases may prove important for the practical and efficient application of precision-based interventions for OSA in a clinical setting. Notable study strengths include a large sample size, a phenotyping algorithm with demonstrated improvements to ICD-based coding in chart review, and careful consideration of obesity and healthcare utilization effects. Limitations included the relatively smaller amount of sleep recording data, limited follow-up time for prospective analyses, and the inherent limitations of a clinical biobank.

In conclusion, we have identified hundreds of associations between diseases and OSA in a large clinical biobank, with many occurring years before diagnosis. Several associations are supported by additional sleep recording evidence. We utilized a relative novelty of methods to perform EHR-based analyses that should be evaluated with future replication and generalizability studies. The wide range of disease associations highlight an opportunity to efficiently target OSA screenings in specific clinics. Understanding the patterns of EHR-derived data will be important for the appropriate application of OSA interventions in clinical environments.

## Contributors

BEC, HLK, NAS, LJE, MKP, and SR contributed to study design and funding. BEC, LL, MD, CY, and MOG contributed to data acquisition. SMH and XY performed clinical chart reviews. BEC and MD analyzed the study. BEC drafted the first version of the article. All authors contributed to analysis interpretation and manuscript review. BEC had full access to the study data and takes responsibility for the integrity of the data and accuracy of analyses.

## Data sharing

The primary data contain protected health information and cannot be shared. All supporting summary analysis results are included in the appendix.

## Acknowledgments

The authors wish to thank the Mass General Brigham RPDR and Biobank and participants for providing samples, genomic data, and health information data. The authors also wish to thank the staff of the BWFH Sleep Testing Center.

## Supplementary Information

### Additional file 1

This file contains supplementary methods, 11 supplementary figures, and 12 supplementary tables.

