## Supplementary Materials for "Associations between obstructive sleep apnea and comorbidities in a large clinical biobank"

#### Supplementary Material


| Item | Page |
|---|---|
| Supplementary Methods. | 2 |
| Figure S1. Case-control BMI density plots. | 3 |
| Figure S2. Case−control birth date density plots. | 4 |
| Figure S3. Relative temporal trends of lipid and red blood cell-related laboratory results. | 5 |
| Figure S4. Relative temporal trends of the 421 associated PheCodes for participants with sleep apnea. | 6 |
| Figure S5. Relative temporal trends of sleep-related clinical note concepts for participants with sleep apnea. | 7 |
| Figure S6. Relative temporal trends of the top 50 associated PheCodes for participants with sleep apnea. | 8 |
| Figure S7. Proportional clinical note occurrences of sleep-related clinical note concepts for participants with sleep apnea. | 9 |
| Figure S8. Relative temporal trends of the 421 associated PheCodes for matched controls without sleep apnea. | 10 |
| Figure S9. Relative temporal trends of sleep-related clinical note concepts for matched controls without sleep apnea. | 11 |
| Figure S10. Relative temporal trends of the top 50 associated PheCodes for matched controls without sleep apnea. | 12 |
| Figure S11. Proportional clinical note occurrences of sleep-related clinical note concepts for matched controls without OSA. | 13 |
| Table S1. PheVis Note Terms. | 14 |
| Table S2. Algorithm Chart Review Performance. | 16 |
| Table S3. Sleep apnea heritability. | 17 |
| Table S4. Incident case/control comorbidity associations. | 18 |
| Table S5. Cross-sectional case/control comorbidity associations. | 34 |
| Table S6. Temporal stability of lead cross-sectional case-control associations. | 59 |
| Table S7. Relative temporal trends of lipid and red blood cell-related laboratory associations with OSA. | 83 |
| Table S8. Sleep-related note term frequencies. | 90 |
| Table S9. Incident sleep statistic associations with comorbidities. | 96 |
| Table S10. Incident sleep statistic associations with comorbidities sensitivity analyses. | 105 |
| Table S11. Cross-sectional sleep statistic associations with comorbidities. | 113 |
| Table S12. Cross-sectional sleep statistic associations with comorbidities sensitivity analyses. | 138 |

#### Supplementary Methods

<u>Genetic data and heritability analyses</u>

Genotype chip data were imputed using the TOPMed r3 imputation panel[1], following quality control procedures addressing participant and variant missingness, Hardy-Weinberg equilibrium, and forward-strand build lift-over, prior to additional TOPMed Imputation Server-based quality control. Autosomal variants with info scores ≥0·95 and minor allele frequencies >0·5% were retained. FRAPOSA[2] was used to obtain principal components and 1000 Genomes Project-defined continental ancestries.

Genetic heritability analyses were performed separately for each continental ancestry (n >1,000). Participants with ICD-coded OSA but PheVis non-case status were excluded, as were participants with a known maximum AHI that fill in the “mild OSA” range (5 – 15). Analyses were performed using PCGC (via LDAK), adjusting for age, sex at birth, BMI, and 10 population principal components. We report marginal results, which accounts for covariate variance and gives lower estimates than conditional results.[3,4] While algorithm heritability comparisons used data censored in 2023, final heritability estimates used additional phenotype data collected through 2025.

Figure S1. Case−control BMI density plots.

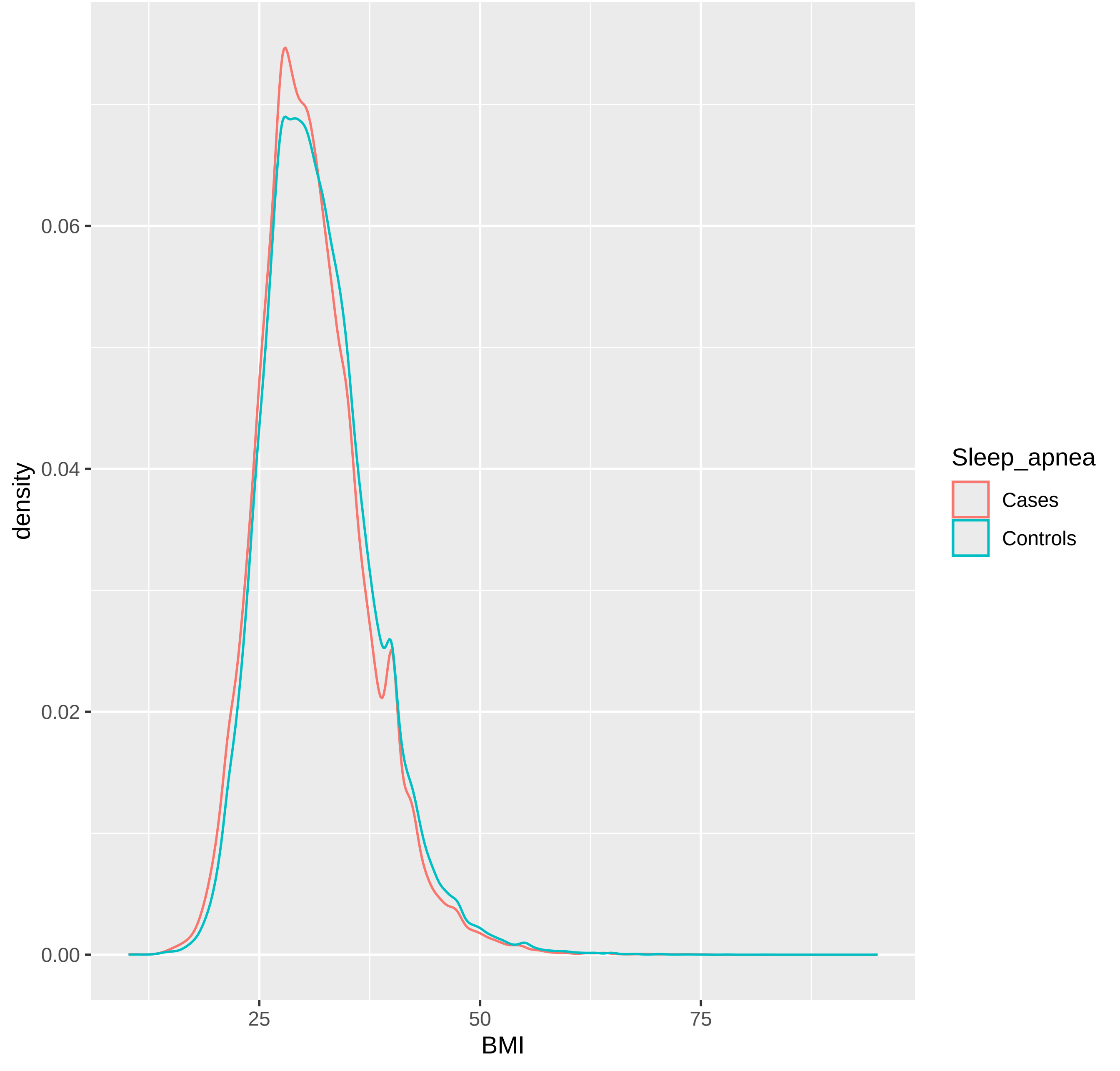

Figure S2. Case−control birth date density plots.

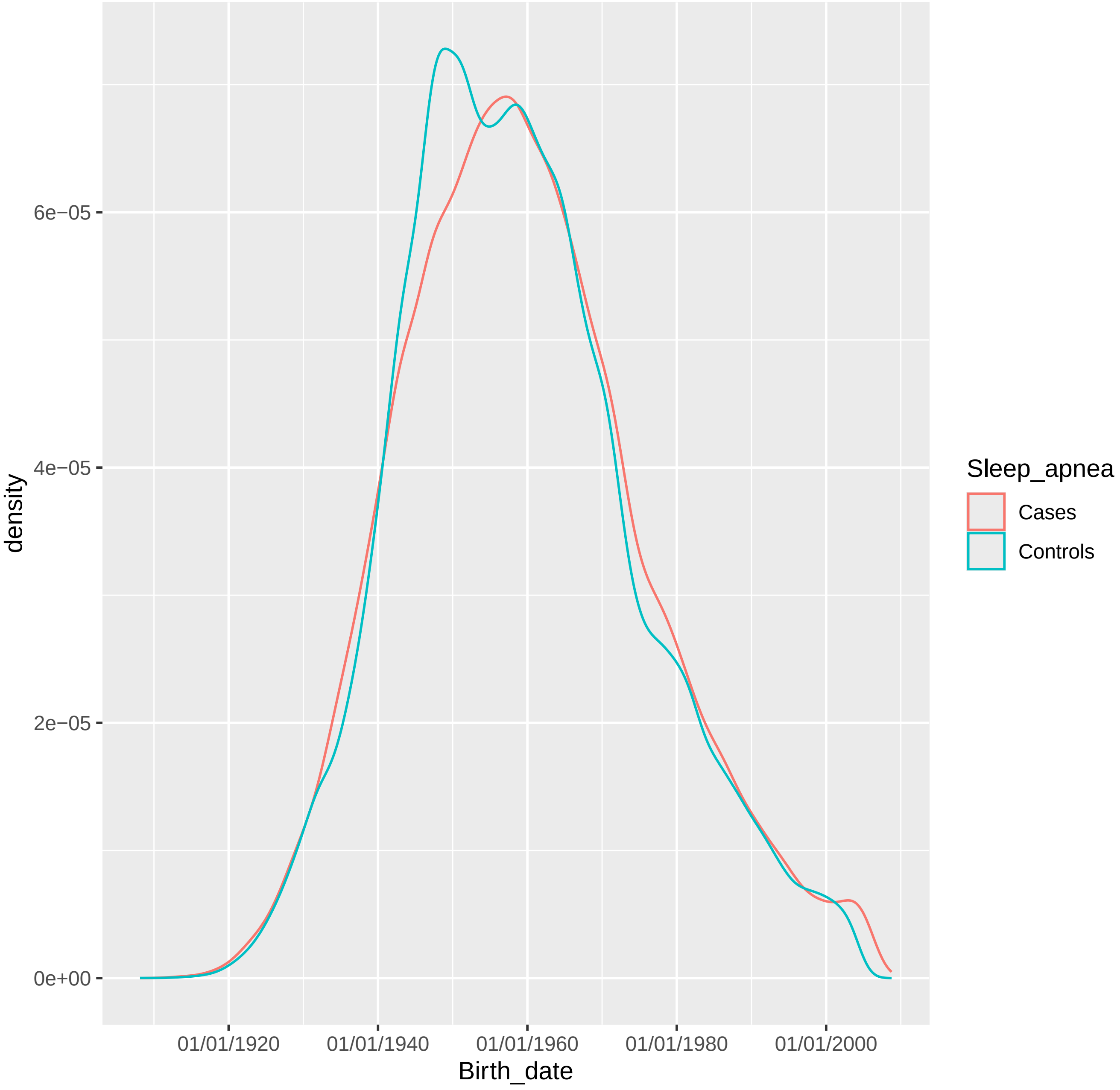

Figure S3. Relative temporal trends of lipid and red blood cell-related laboratory results.

Median values for years relative to the first OSA diagnosis (or equivalent age in controls) were obtained. All available results were used, with the proportion of cases and controls with data varying by year (Table S7). Analyses adjusted for age, sex, BMI, population, and cumulative clinical encounter counts through the given year. Age was specific to the relative year, and the closest two BMIs to this relative date were averaged. The results are grouped by red blood cell then by lipid analysis and ordered by significance ten years prior to the first OSA diagnosis date. Full results are shown in Table S7.

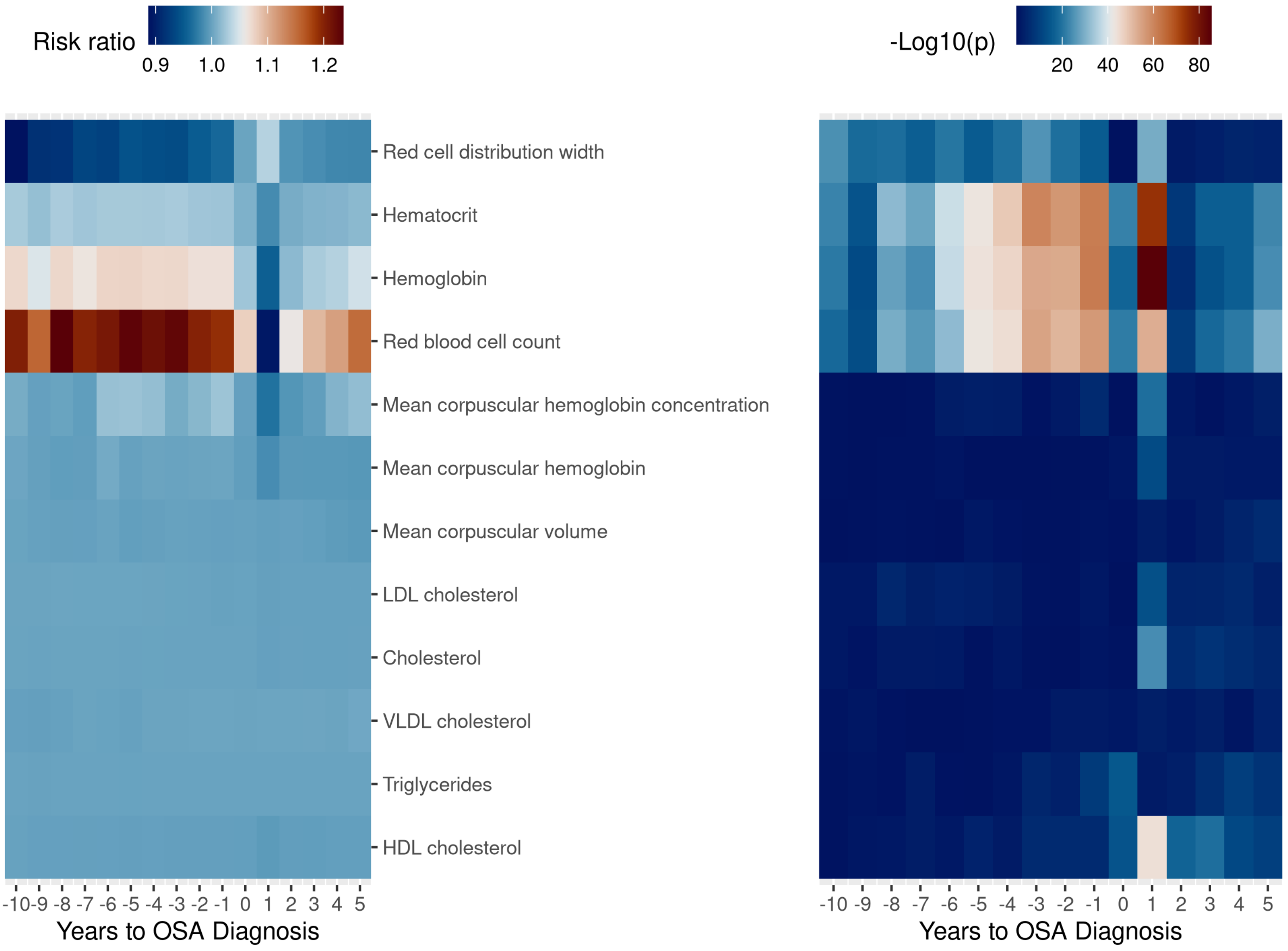

Figure S4. Relative temporal trends of the 421 associated PheCodes for participants with sleep apnea.

Relative PheCode (ICD) diagnosis dates were oriented to 1/1/2000 as the first sleep apnea diagnosis date. The 421 Bonferroni-significant PheCodes are listed in Table S5. All PheCode diagnosis dates were used, in contrast to other analyses using only the first recorded diagnosis date. Positions reflect temporal trends of all variables in year-wide bins, with a longer distance reflecting greater differences between two relative years. Multidimensional scaling is used to visualize the variability among data from relative years. Axis coordinates reflect the components of variance. Relative years are positioned as points where the distance between them represents the Jensen-Shannon probabilistic distance between their distributions. For more information, please see the EHRtemporalVariability references (PMID:30342679 and PMC7391413).

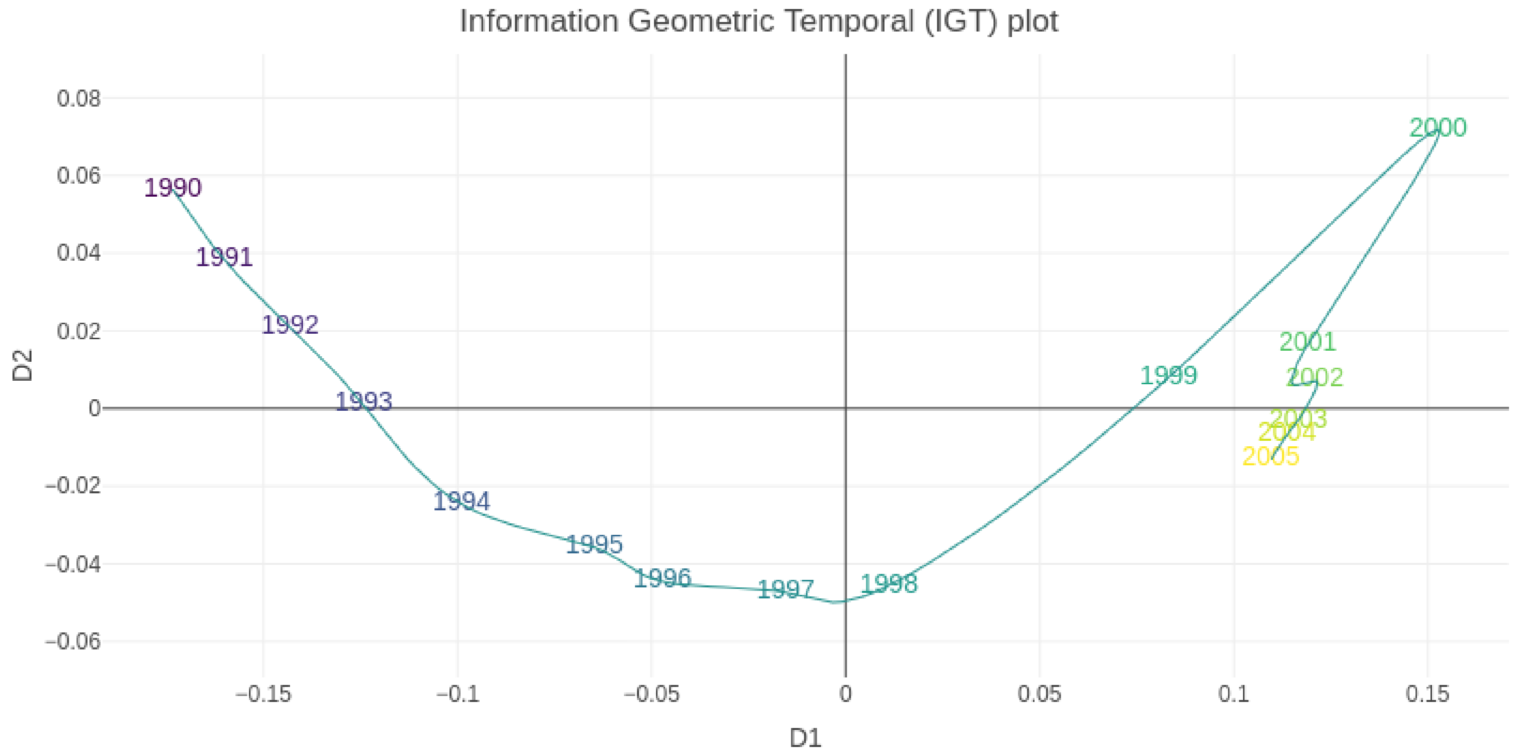

Figure S5. Relative temporal trends of sleep-related clinical note concepts for participants with sleep apnea.

Relative clinical note dates were oriented to 1/1/2000 as the first sleep apnea diagnosis date. The 47 sleep-related terms are defined in Table S8. Positions reflect temporal trends of all variables in year-wide bins, with a longer distance reflecting greater differences between two relative years. Multidimensional scaling is used to visualize the variability among data from relative years. Axis coordinates reflect the components of variance. Relative years are positioned as points where the distance between them represents the Jensen-Shannon probabilistic distance between their distributions. For more information, please see the EHRtemporalVariability references (PMID:30342679 and PMC7391413).

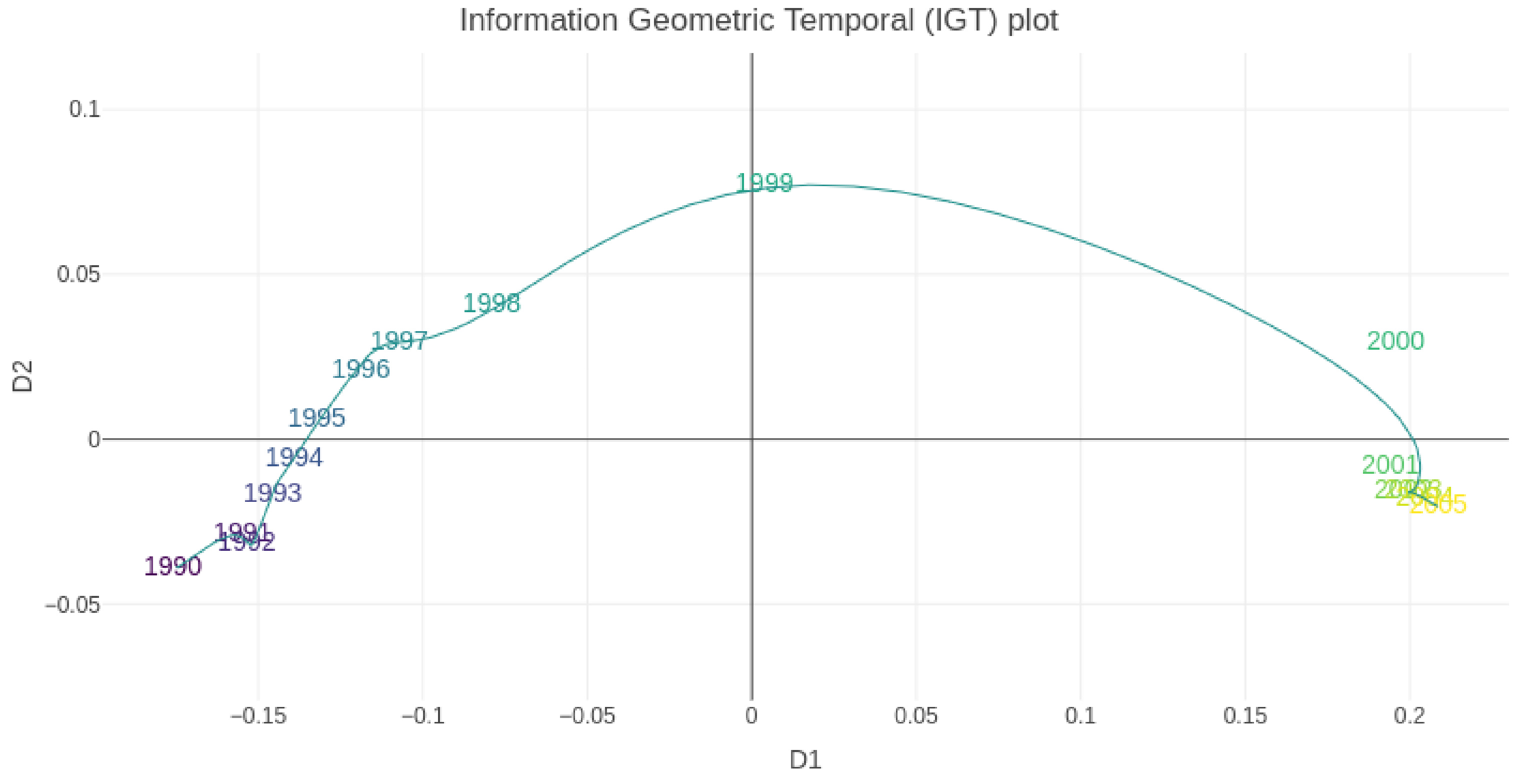

Figure S6. Relative temporal trends of the top 50 associated PheCodes for participants with sleep apnea.

Relative PheCode (ICD) diagnosis dates were oriented to 1/1/2000 as the first sleep apnea diagnosis date. The top 50 PheCodes are listed in Figure S6. All PheCode diagnosis dates were used, in contrast to other analyses using only the first recorded diagnosis date. Positions reflect temporal trends of all variables in year-wide bins, with a longer distance reflecting greater differences between two relative years. Multidimensional scaling is used to visualize the variability among data from relative years. Axis coordinates reflect the components of variance. Relative years are positioned as points where the distance between them represents the Jensen-Shannon probabilistic distance between their distributions. For more information, please see the EHRtemporalVariability references (PMID:30342679 and PMC7391413).

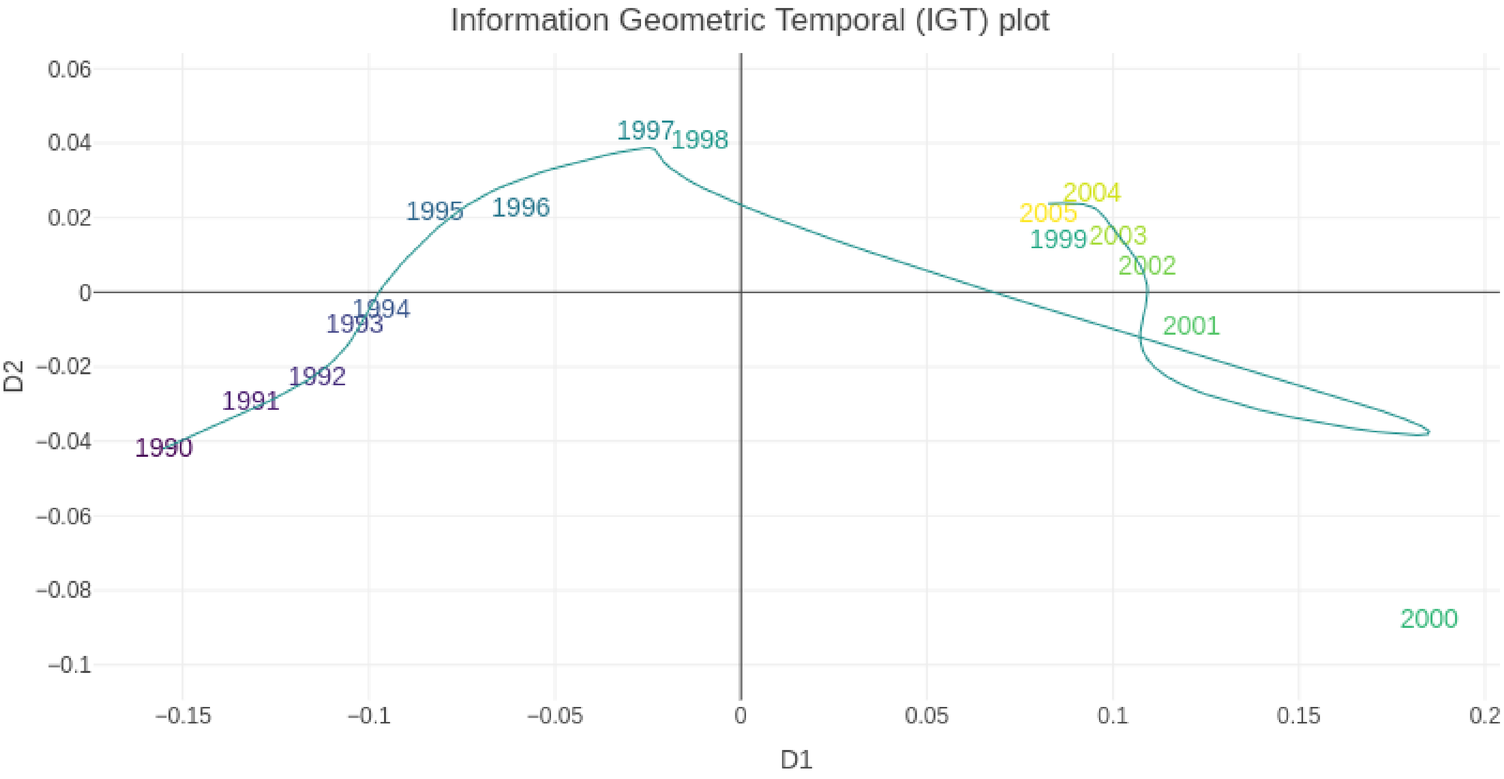

Figure S7. Proportional clinical note occurrences of sleep-related clinical note concepts for participants with sleep apnea.

Relative clinical note dates were oriented to 1/1/2000 as the first sleep apnea diagnosis date. The 47 sleep-related terms are defined in Table S8. The analysis was performed using the EHRtemporalVariability package (PMC7391413).

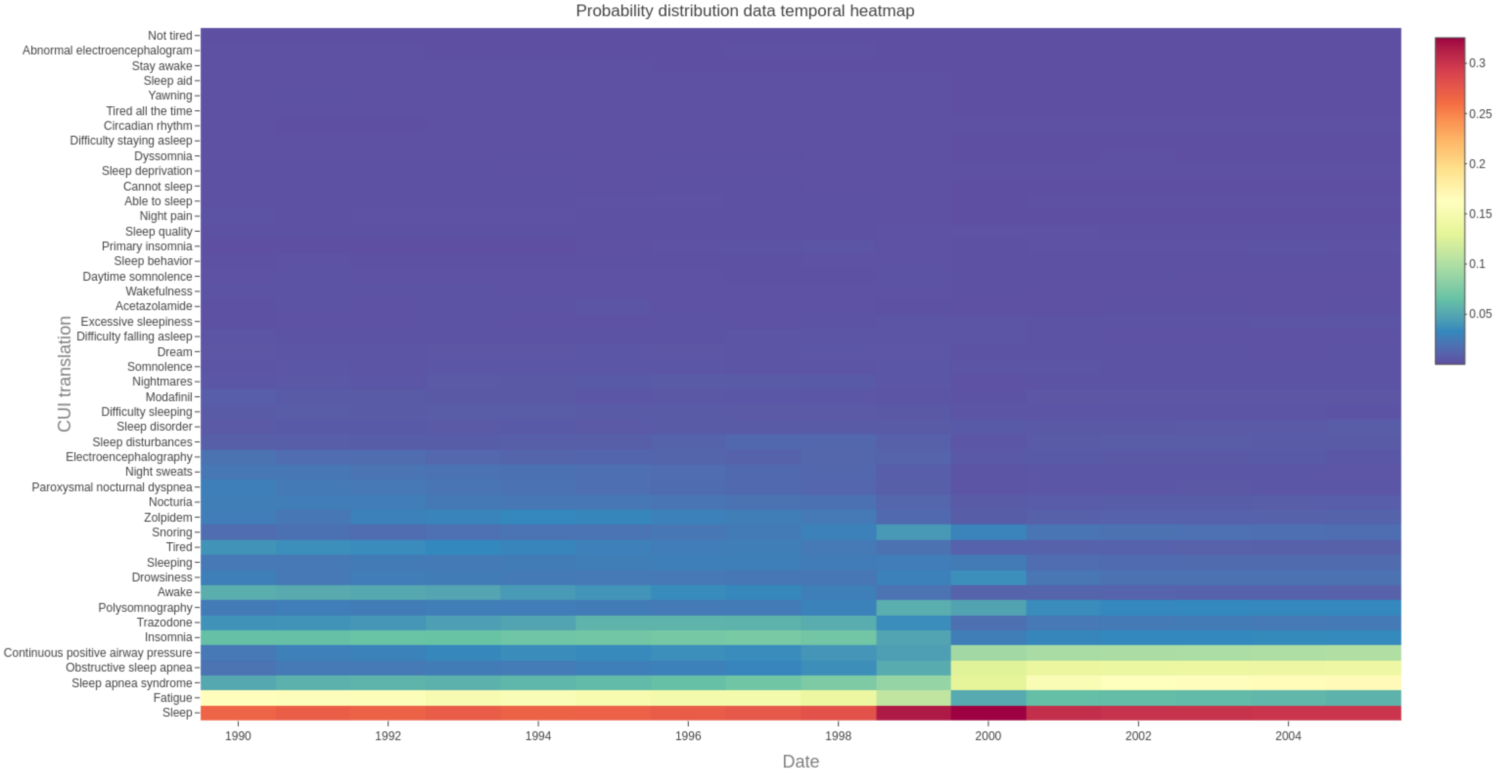

Figure S8. Relative temporal trends of the 421 associated PheCodes for matched controls without sleep apnea.

Relative PheCode (ICD) diagnosis dates were oriented to 1/1/2000 as the first sleep apnea diagnosis date. The 421 Bonferroni-significant PheCodes are listed in Table S5. All PheCode diagnosis dates were used, in contrast to other analyses using only the first recorded diagnosis date. Positions reflect temporal trends of all variables in year-wide bins, with a longer distance reflecting greater differences between two relative years. Multidimensional scaling is used to visualize the variability among data from relative years. Axis coordinates reflect the components of variance. Relative years are positioned as points where the distance between them represents the Jensen-Shannon probabilistic distance between their distributions. For more information, please see the EHRtemporalVariability references (PMID:30342679 and PMC7391413).

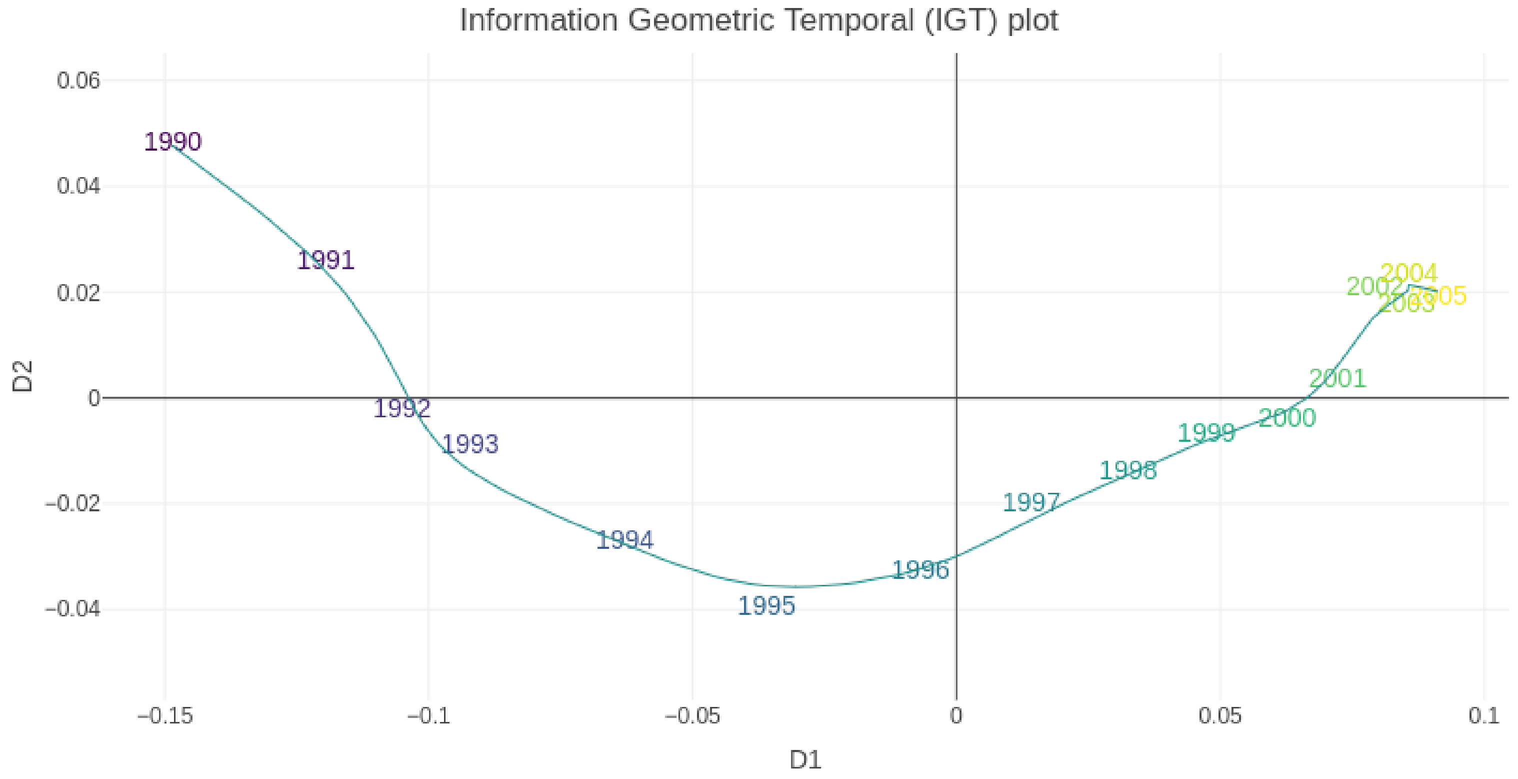

Figure S9. Relative temporal trends of sleep-related clinical note concepts for matched controls without sleep apnea.

Relative clinical note dates were oriented to 1/1/2000 as the first sleep apnea diagnosis date. The 47 sleep-related terms are defined in Table S8. Positions reflect temporal trends of all variables in year-wide bins, with a longer distance reflecting greater differences between two relative years. Multidimensional scaling is used to visualize the variability among data from relative years. Axis coordinates reflect the components of variance. Relative years are positioned as points where the distance between them represents the Jensen-Shannon probabilistic distance between their distributions. For more information, please see the EHRtemporalVariability references (PMID:30342679 and PMC7391413).

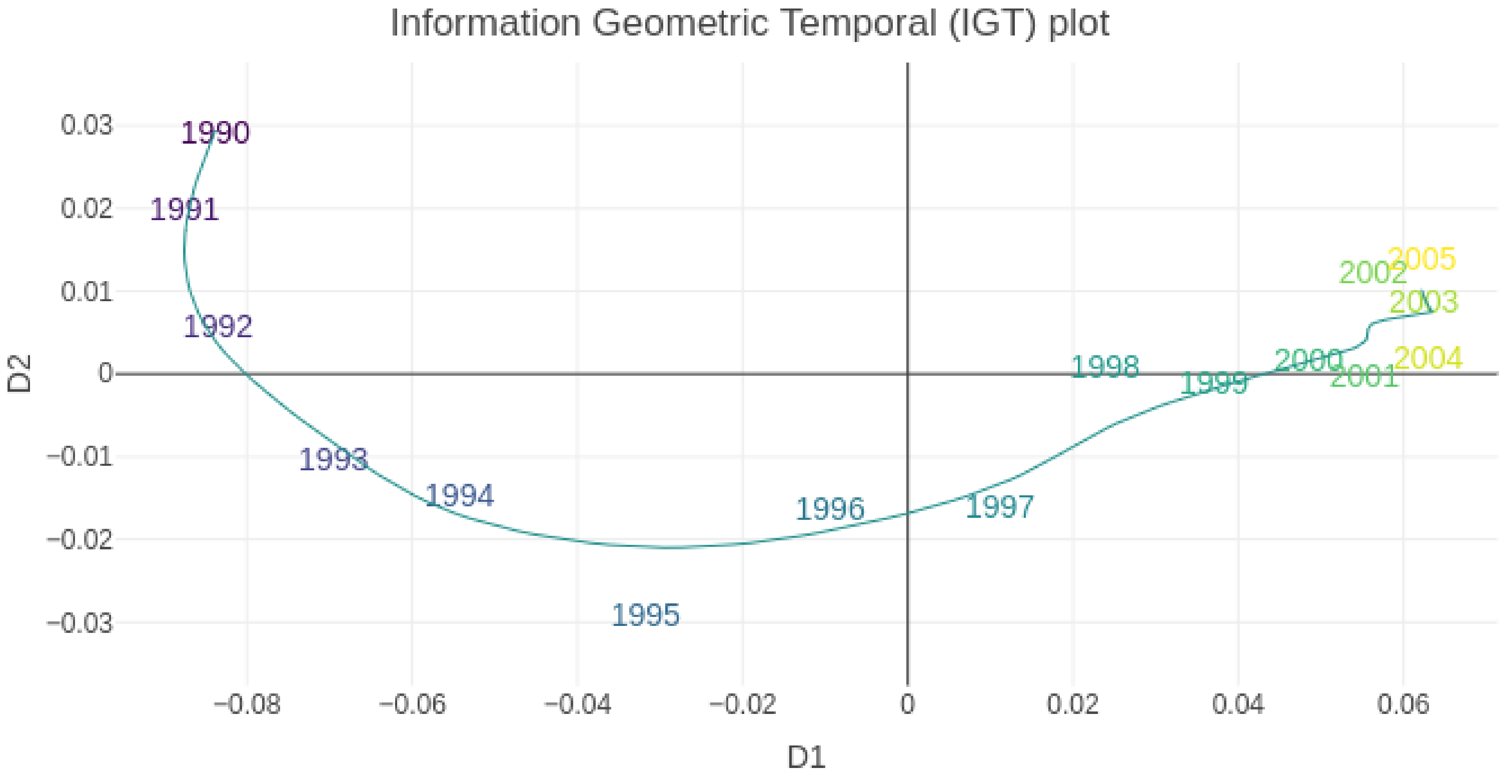

Figure S10. Relative temporal trends of the top 50 associated PheCodes for matched controls without sleep apnea.

Relative PheCode (ICD) diagnosis dates were oriented to 1/1/2000 as the first sleep apnea diagnosis date. The top 50 PheCodes are listed in Figure S6. All PheCode diagnosis dates were used, in contrast to other analyses using only the first recorded diagnosis date. Positions reflect temporal trends of all variables in year-wide bins, with a longer distance reflecting greater differences between two relative years. Multidimensional scaling is used to visualize the variability among data from relative years. Axis coordinates reflect the components of variance. Relative years are positioned as points where the distance between them represents the Jensen-Shannon probabilistic distance between their distributions. For more information, please see the EHRtemporalVariability references (PMID:30342679 and PMC7391413).

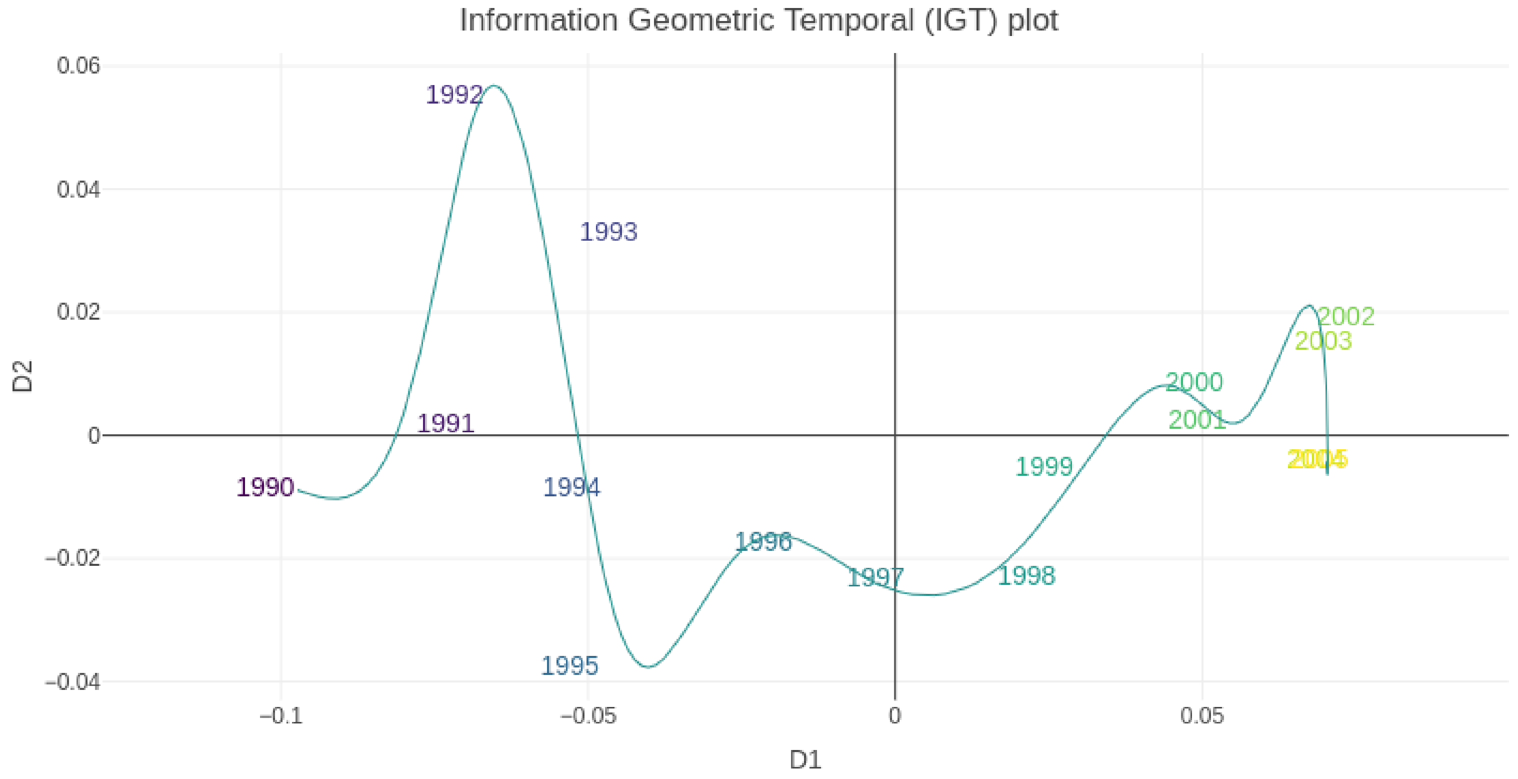

Figure S11. Proportional clinical note occurrences of sleep-related clinical note concepts for matched controls without OSA.

Relative clinical note dates were oriented to 1/1/2000 as the first sleep apnea diagnosis date. The 47 sleep-related terms are defined in Table S8. The analysis was performed using the EHRtemporalVariability package (PMC7391413).

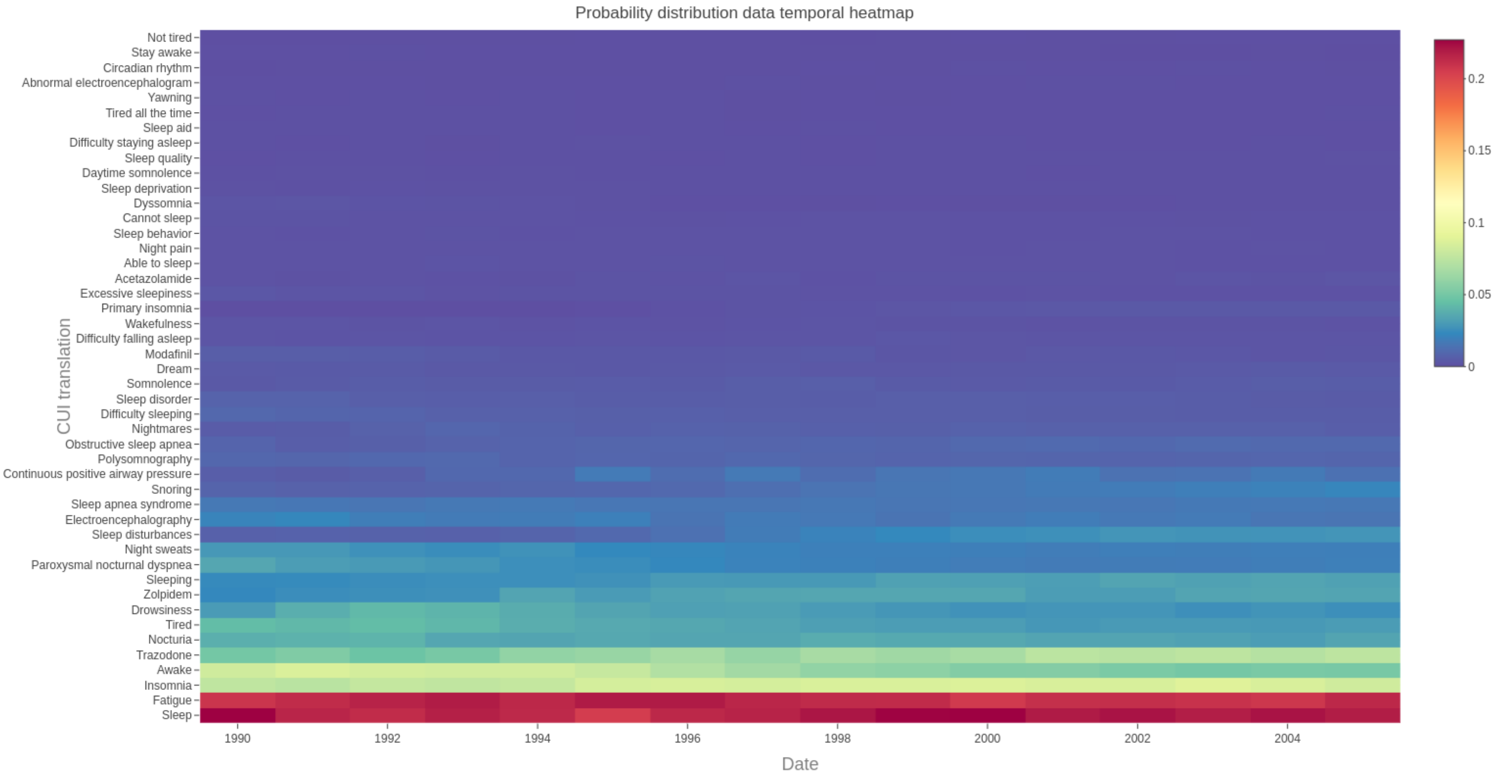

#### Supplemental Tables

##### Table S1. PheVis Note Terms.

The SAFE procedure (PMC6080726) was used to identify these clinical note terms (UMLS, https://www.nlm.nih.gov/research/umls/index.html) for use in PheVis. Terms were found in a majority of online sleep apnea reviews and were present in our clinical notes as extracted using cTAKES. Reviews are further described in Table S1 of PMC8826997 and included NLP extraction of online information from: American Thoracic Society Patient Series, Mayo Clinic, Medline, Medscape, Merck Manuals, Nature Reviews Disease Primers, and Wikipedia. Excluded status terms were removed prior to PheVis calculations due to non-specificity (e.g. “does pull”) or false-positive matches (e.g. the metal “lead” versus PSG electrode leads). MainCUI refers to a prioritized joint term consisting of the sum of the "sleep apnea" and "obstructive sleep apnea syndrome" term counts.

| Status | CUI | Translation |
|---|---|---|
| Retained | [mainCUI] | mainCUI = C0037315 (sleep apnea); C0520679 (obstructive sleep apnea syndrome) |
| Retained | C0001425 | Adenoidectomy |
| Retained | C0003578 | Apnea |
| Retained | C0003811 | Cardiac Arrhythmia |
| Retained | C0004238 | Atrial Fibrillation |
| Retained | C0007222 | Cardiovascular Diseases |
| Retained | C0008301 | Choking |
| Retained | C0013144 | Drowsiness |
| Retained | C0013839 | Electromyography |
| Retained | C0018801 | Heart failure |
| Retained | C0018802 | Congestive heart failure |
| Retained | C0024117 | Chronic Obstructive Airway Disease |
| Retained | C0027051 | Myocardial Infarction |
| Retained | C0027424 | Nasal congestion (finding) |
| Retained | C0028734 | Nocturia |
| Retained | C0028754 | Obesity |
| Retained | C0036557 | Sedatives |
| Retained | C0036974 | Shock |
| Retained | C0037317 | Sleep disturbances |
| Retained | C0037384 | Snoring |
| Retained | C0043352 | Xerostomia |
| Retained | C0085631 | Agitation |
| Retained | C0085633 | Mood swings |

| Status | CUI | Translation |
|---|---|---|
| Retained | C0162701 | Polysomnography |
| Retained | C0199451 | Continuous Positive Airway Pressure |
| Retained | C0233660 | Mental blocking |
| Retained | C0233794 | Memory impairment |
| Retained | C0242184 | Hypoxia |
| Retained | C0338656 | Impaired cognition |
| Retained | C0344329 | Collapse (finding) |
| Retained | C0424295 | Hyperactive behavior |
| Retained | C0424563 | Quality of sleep |
| Retained | C0425449 | Gasping for breath |
| Retained | C0425466 | Respiratory effort |
| Retained | C0497247 | Increase in blood pressure |
| Retained | C0497406 | Overweight |
| Retained | C0517967 | Respiratory pattern |
| Retained | C0542476 | Forgetful |
| Retained | C0700292 | Hypoxemia |
| Retained | C0746961 | Oxygen saturation below reference range |
| Retained | C0851578 | Sleep Disorders |
| Retained | C0917801 | Sleeplessness |
| Retained | C1272641 | Systemic arterial pressure |
| Retained | C2830004 | Somnolence |
| Retained | C3854333 | Narrowing |
| Retained | C3887611 | Restlessness |
| Excluded | C0013216 | Pharmacotherapy |
| Excluded | C0013658 | Educational Status |
| Excluded | C0023175 | Lead |
| Excluded | C0242402 | Opioids |
| Excluded | C0332155 | Did not receive therapy or drug for |
| Excluded | C0436331 | Symptom aggravating factors |
| Excluded | C0541749 | Does turn (finding) |
| Excluded | C0580846 | Does pull |
| Excluded | C1292856 | Stimulation procedure |
| Excluded | C1705232 | Death Related to Adverse Event |

#### Table S2. Algorithm Chart Review Performance.

| Method | Total | True positives | False positives | True negatives | False negatives | Accuracy | Precision | Negative predictive value | Recall | Specificity | F1 |
|---|---|---|---|---|---|---|---|---|---|---|---|
| 1+ PheCode | 484 | 333 | 150 | 0 | 1 | 0.688 | 0.689 | 0.000 | 0.997 | 0.000 | 0.815 |
| PheVis | 484 | 262 | 37 | 113 | 72 | 0.775 | 0.876 | 0.611 | 0.784 | 0.753 | 0.828 |
| PheVis + AHI (PSG) | 481 | 263 | 38 | 110 | 70 | 0.775 | 0.874 | 0.611 | 0.790 | 0.743 | 0.830 |
| PheVis + AHI (PSG and NLP) | 476 | 268 | 39 | 109 | 60 | 0.792 | 0.873 | 0.645 | 0.817 | 0.736 | 0.844 |
| PheVis + AHI (PSG and NLP) + CPAP | 477 | 270 | 39 | 109 | 59 | 0.795 | 0.874 | 0.649 | 0.821 | 0.736 | 0.846 |

The PheVis algorithm was chosen to optimize high-confidence cases (i.e. precision / positive predictive value) for use with controls defined by 0 OSA PheCodes. 484 chart reviews were performed for participants with one or more sleep apnea PheCode codings. Therefore PheCode-only values lack negative predictive values by definition. The base NLP prediction PheVis was supplemented with available AHI data obtained from sleep reports (PSG) and extracted from clinical notes (NLP). A PheVis control would be converted to a case if AHI >= 15 or nulled if the AHI was between 5 - 15. Note that this could still classify a patient as a case with sleep apnea using ICSD-3 criteria with available comorbidities. Finally, evidence of a CPAP prescription could convert a PheVis control to case, but these data were only available for participants seen in the Faulkner clinic and affected 2 participants in the chart review. AHI and CPAP information were masked if the corresponding date occurred after the chart review, which lowers the total count.

Accuracy = (True positives + True negatives) / (True positives + False positives + False negatives + True negatives)
Precision = True positives / (True positives + False positives)
Negative predictive value = True negatives / (True negatives + False negatives)
Recall = True positives / (True positives + False negatives)
Specificity = True negatives / (True negatives + False positives)
F1 = (2 * Recall * Precision) / (Recall + Precision)

#### Table S3. Sleep apnea heritability.

| Continental ancestry | Cases | Controls | BMI adjusted estimate (SE) | BMI unadjusted estimate (SE) |
|---|---|---|---|---|
| AFR | 527 | 2171 | -0.064 (0.263) | 0.157 (0.219) |
| AMR | 844 | 5162 | 0.266 (0.162) | 0.152 (0.152) |
| EUR | 6336 | 39162 | 0.105 (0.027) | 0.072 (0.020) |
| Combined populations | 7878 | 48204 | 0.130 (0.027) | 0.088 (0.018) |

Marginal PCGC-based heritability estimates are shown, adjusted for age, sex, BMI, and 10 population principal components. Marginal estimates account for variance introduced by the covariates and may be lower than conditional heritability estimates often reported using other methods. Continental ancestry was calculated based on 1000 Genomes Project template data. Ancestry groups with n > 1,000 (African, American, and European) were directly tested. Negative heritability estimates are possible when using unbounded calculations and can be seen in results with relatively high standard error values. The final choice of PheVis + AHI information was based on the maximizing the point-wise estimate of the largest sample (EUR). Inclusion of CPAP prescription data decreased the pointwise estimate in EUR, although there were further marginal improvements in post-hoc analyses of the combined population sample (0.133 [0.027]). No BMI values were used to construct the PheVis algorithm estimates.

#### Table S4. Incident case/control comorbidity associations.

Incident analyses considered diagnoses at least one year after the initial sleep apnea diagnosis. Primary analyses considered joint-sex PheCodes with >1% incidence rate and were adjusted for age, sex, BMI, population, and healthcare encounters. Bonferroni-significant p-values are bolded. A sensitivity analysis using alternative control propensity score matching without healthcare encounters (but still adjusting for encounters in the analysis) was performed. PheCodes with discordant results (n=32) or p > 0.05 (n=187) in either analysis are displayed at the end of the table (from PheCode 721.8 onwards, highlighted in green). 179 PheCodes were both Bonferroni-level significant and had concordant directionality in the sensitivity analysis. Sleep apnea × sex interaction analyses were performed separately when incidence was >1% in men and women. Equivalent sex-stratified analyses are shown.

| PheCode | Disease | Incidence OSA case / control | Risk ratio | P-value | Sensitivity analysis risk ratio | OSA x sex interaction p-value | Risk ratio (men) | Risk ratio (women) |
|---|---|---|---|---|---|---|---|---|
| 472 | Chronic pharyngitis and nasopharyngitis | 3.3 / 0.5 | 6.19 (5.55 – 6.90) | **3.0E-236** | 2.81 (2.56 – 3.09) | 1.0E-01 | 6.72 (5.80 – 7.78) | 5.55 (4.72 – 6.54) |
| 296.22 | Major depressive disorder | 9.9 / 5.6 | 1.87 (1.79 – 1.95) | **2.6E-179** | 2.22 (2.12 – 2.33) | 5.8E-04 | 1.73 (1.63 – 1.84) | 2.00 (1.89 – 2.12) |
| 455 | Hemorrhoids | 7.0 / 3.1 | 2.20 (2.09 – 2.33) | **1.2E-176** | 1.09 (1.03 – 1.15) | **1.5E-04** | 2.41 (2.25 – 2.59) | 1.92 (1.76 – 2.09) |
| 327 | Sleep disorders | 2.4 / 0.4 | 5.90 (5.20 – 6.70) | **8.7E-166** | 2.24 (2.01 – 2.49) | 8.0E-01 | 5.86 (4.97 – 6.91) | 5.96 (4.89 – 7.26) |
| 689 | Disorder of skin and subcutaneous tissue NOS | 6.8 / 3.3 | 1.99 (1.88 – 2.09) | **2.9E-140** | 1.23 (1.16 – 1.29) | 4.9E-03 | 2.10 (1.97 – 2.25) | 1.80 (1.65 – 1.96) |
| 327.71 | Restless legs syndrome | 2.1 / 0.5 | 4.50 (4.00 – 5.07) | **5.6E-136** | 5.38 (4.65 – 6.23) | 8.6E-01 | 4.37 (3.68 – 5.19) | 4.59 (3.89 – 5.41) |
| 470 | Septal Deviations/Turbinate Hypertrophy | 3.2 / 1.1 | 2.86 (2.63 – 3.11) | **9.4E-129** | 3.08 (2.78 – 3.42) | **2.6E-05** | 3.30 (2.96 – 3.68) | 2.23 (1.94 – 2.57) |
| 476 | Allergic rhinitis | 6.9 / 3.7 | 1.93 (1.82 – 2.03) | **7.2E-126** | 1.56 (1.46 – 1.66) | **6.8E-05** | 2.15 (2.00 – 2.31) | 1.69 (1.56 – 1.83) |
| 327.4 | Insomnia | 5.4 / 2.9 | 1.83 (1.73 – 1.94) | **1.6E-89** | 2.16 (2.02 – 2.32) | 2.6E-01 | 1.78 (1.64 – 1.93) | 1.89 (1.74 – 2.06) |
| 289.9 | Abnormality of red blood cells | 1.1 / 0.1 | 7.36 (6.00 – 9.02) | **4.0E-82** | 1.36 (1.19 – 1.56) | NA | NA | 11.26 (7.98 – 15.89) |
| 272.1 | Hyperlipidemia | 12.8 / 9.4 | 1.48 (1.42 – 1.54) | **2.3E-76** | 1.08 (1.03 – 1.13) | 3.3E-01 | 1.48 (1.40 – 1.55) | 1.49 (1.39 – 1.60) |
| 278 | Overweight, obesity and other hyperalimentation | 6.7 / 4.1 | 1.60 (1.52 – 1.69) | **4.0E-72** | 1.62 (1.52 – 1.73) | 1.7E-02 | 1.69 (1.58 – 1.80) | 1.49 (1.38 – 1.62) |
| 292 | Neurological disorders | 1.5 / 0.5 | 3.12 (2.74 – 3.54) | **2.0E-68** | 1.97 (1.74 – 2.22) | 3.1E-01 | 3.27 (2.77 – 3.86) | 2.89 (2.36 – 3.53) |
| 196 | Radiotherapy | 2.3 / 4.1 | 0.54 (0.51 – 0.58) | **9.4E-66** | 0.65 (0.61 – 0.70) | 5.6E-02 | 0.51 (0.47 – 0.56) | 0.60 (0.54 – 0.68) |
| 379.9 | Pain, swelling or discharge of eye | 1.2 / 0.4 | 3.46 (3.00 – 4.00) | **4.0E-64** | 2.74 (2.28 – 3.28) | 4.9E-02 | 3.04 (2.50 – 3.71) | 4.00 (3.24 – 4.93) |
| 292.3 | Memory loss | 3.1 / 1.5 | 1.93 (1.78 – 2.08) | **1.3E-62** | 1.74 (1.59 – 1.91) | 6.2E-03 | 2.11 (1.90 – 2.34) | 1.72 (1.54 – 1.93) |

| PheCode | Disease | Incidence OSA case / control | Risk ratio | P-value | Sensitivity analysis risk ratio | OSA x sex interaction p-value | Risk ratio (men) | Risk ratio (women) |
|---|---|---|---|---|---|---|---|---|
| 379 | Other disorders of eye | 1.1 / 0.0 | 41.54 (26.81 – 64.35) | **1.6E-62** | 1.40 (1.16 – 1.69) | 3.3E-01 | 52.08 (26.77 – 101.32) | 33.45 (18.69 – 59.88) |
| 300.11 | Generalized anxiety disorder | 4.8 / 3.1 | 1.62 (1.53 – 1.72) | **1.1E-60** | 2.09 (1.95 – 2.23) | 4.9E-01 | 1.59 (1.46 – 1.73) | 1.65 (1.53 – 1.79) |
| 475 | Chronic sinusitis | 2.9 / 1.6 | 1.90 (1.75 – 2.07) | **3.2E-54** | 1.63 (1.49 – 1.79) | 2.5E-04 | 2.19 (1.96 – 2.44) | 1.59 (1.40 – 1.80) |
| 550.2 | Diaphragmatic hernia | 2.3 / 1.1 | 2.05 (1.87 – 2.25) | **6.0E-54** | 1.34 (1.20 – 1.49) | 1.6E-01 | 2.16 (1.88 – 2.48) | 1.95 (1.72 – 2.20) |
| 306 | Other mental disorder | 4.2 / 2.6 | 1.63 (1.53 – 1.74) | **2.1E-53** | 1.62 (1.51 – 1.74) | 5.1E-01 | 1.66 (1.53 – 1.80) | 1.60 (1.46 – 1.76) |
| 278.1 | Obesity | 9.2 / 8.4 | 1.41 (1.35 – 1.48) | **1.1E-52** | 1.53 (1.46 – 1.60) | 2.4E-01 | 1.53 (1.44 – 1.63) | 1.32 (1.23 – 1.40) |
| 313.1 | Attention deficit hyperactivity disorder | 1.5 / 0.7 | 2.30 (2.06 – 2.56) | **1.1E-50** | 2.65 (2.32 – 3.01) | 1.4E-01 | 2.44 (2.12 – 2.80) | 2.08 (1.75 – 2.49) |
| 208 | Benign neoplasm of colon | 7.0 / 4.7 | 1.47 (1.39 – 1.54) | **1.6E-49** | 1.31 (1.24 – 1.38) | 9.8E-01 | 1.48 (1.39 – 1.58) | 1.45 (1.33 – 1.58) |
| 327.41 | Organic or persistent insomnia | 1.5 / 0.7 | 2.32 (2.07 – 2.60) | **1.5E-47** | 2.15 (1.89 – 2.46) | 9.6E-01 | 2.30 (1.95 – 2.71) | 2.33 (1.99 – 2.73) |
| 300.4 | Dysthymic disorder | 1.4 / 0.7 | 2.22 (1.98 – 2.50) | **5.4E-40** | 5.10 (3.82 – 6.81) | 5.9E-01 | 2.14 (1.81 – 2.54) | 2.29 (1.94 – 2.70) |
| 197 | Chemotherapy | 2.1 / 3.5 | 0.61 (0.56 – 0.65) | **1.1E-39** | 0.57 (0.53 – 0.62) | 3.4E-01 | 0.59 (0.53 – 0.64) | 0.64 (0.57 – 0.72) |
| 389 | Hearing loss | 5.5 / 3.7 | 1.44 (1.36 – 1.52) | **2.3E-39** | 1.72 (1.61 – 1.83) | 2.9E-01 | 1.46 (1.37 – 1.57) | 1.39 (1.27 – 1.52) |
| 798.1 | Chronic fatigue syndrome | 1.5 / 0.7 | 2.11 (1.88 – 2.36) | **2.1E-38** | 2.69 (2.34 – 3.10) | 7.4E-01 | 2.02 (1.71 – 2.39) | 2.17 (1.86 – 2.54) |
| 300.1 | Anxiety disorder | 8.7 / 7.0 | 1.33 (1.28 – 1.39) | **7.0E-38** | 1.56 (1.48 – 1.63) | 8.5E-01 | 1.33 (1.25 – 1.41) | 1.34 (1.26 – 1.42) |
| 564.1 | Irritable Bowel Syndrome | 1.8 / 1.0 | 1.91 (1.73 – 2.12) | **2.1E-37** | 1.85 (1.65 – 2.09) | 1.3E-01 | 2.12 (1.79 – 2.52) | 1.81 (1.60 – 2.04) |
| 706.8 | Other specified diseases of sebaceous glands | 2.4 / 1.3 | 1.72 (1.57 – 1.87) | **5.5E-34** | 1.36 (1.25 – 1.48) | 7.8E-01 | 1.72 (1.54 – 1.93) | 1.69 (1.48 – 1.94) |
| 172.22 | Squamous cell carcinoma | 1.9 / 1.0 | 1.82 (1.65 – 2.01) | **2.5E-33** | 1.34 (1.18 – 1.52) | 7.0E-01 | 1.80 (1.60 – 2.02) | 1.87 (1.56 – 2.25) |
| 530.11 | GERD | 10.9 / 8.8 | 1.28 (1.23 – 1.34) | **2.6E-32** | 1.33 (1.28 – 1.40) | 2.9E-01 | 1.26 (1.20 – 1.34) | 1.31 (1.23 – 1.39) |
| 761 | Cervicalgia | 5.0 / 3.5 | 1.43 (1.35 – 1.52) | **4.3E-32** | 1.55 (1.45 – 1.66) | 3.1E-01 | 1.40 (1.29 – 1.52) | 1.47 (1.35 – 1.61) |
| 701.2 | Scar conditions and fibrosis of skin | 2.8 / 1.7 | 1.58 (1.46 – 1.71) | **1.5E-30** | 1.20 (1.10 – 1.30) | 4.6E-01 | 1.60 (1.45 – 1.77) | 1.54 (1.35 – 1.75) |
| 272.12 | Hyperglyceridemia | 1.8 / 1.1 | 1.74 (1.58 – 1.92) | **1.5E-28** | 1.67 (1.50 – 1.86) | 1.3E-01 | 1.66 (1.48 – 1.87) | 1.92 (1.62 – 2.28) |
| 338.2 | Chronic pain | 6.8 / 5.2 | 1.31 (1.25 – 1.37) | **1.6E-27** | 1.21 (1.15 – 1.28) | 2.7E-01 | 1.35 (1.27 – 1.44) | 1.26 (1.17 – 1.35) |
| 599.5 | Frequency of urination and polyuria | 5.6 / 4.0 | 1.34 (1.27 – 1.42) | **2.7E-27** | 1.57 (1.48 – 1.67) | 3.4E-03 | 1.40 (1.32 – 1.49) | 1.18 (1.06 – 1.32) |
| 348.7 | Coma | 1.1 / 0.5 | 2.01 (1.77 – 2.28) | **1.5E-26** | 2.74 (2.38 – 3.16) | 3.6E-02 | 1.81 (1.55 – 2.12) | 2.43 (1.94 – 3.03) |
| 357 | Inflammatory and toxic neuropathy | 3.5 / 2.4 | 1.44 (1.34 – 1.54) | **3.5E-26** | 1.78 (1.65 – 1.91) | 5.3E-02 | 1.36 (1.24 – 1.48) | 1.57 (1.41 – 1.74) |

| PheCode | Disease | Incidence OSA case / control | Risk ratio | P-value | Sensitivity analysis risk ratio | OSA x sex interaction p-value | Risk ratio (men) | Risk ratio (women) |
|---|---|---|---|---|---|---|---|---|
| 599.9 | Other abnormality of urination | 3.0 / 2.0 | 1.47 (1.37 – 1.59) | **4.8E-25** | 1.61 (1.48 – 1.75) | 8.5E-02 | 1.53 (1.40 – 1.67) | 1.36 (1.19 – 1.54) |
| 300.12 | Agorophobia, social phobia, and panic disorder | 1.2 / 0.7 | 1.90 (1.68 – 2.14) | **8.0E-25** | 1.91 (1.67 – 2.18) | NA | NA | 2.21 (1.87 – 2.61) |
| 262 | Mineral deficiency NEC | 1.4 / 0.8 | 1.77 (1.59 – 1.97) | **2.0E-24** | 2.44 (2.16 – 2.76) | NA | NA | 1.88 (1.64 – 2.16) |
| 271.3 | Intestinal disaccharidase deficiencies and disaccharide malabsorption | 5.0 / 3.8 | 1.33 (1.25 – 1.40) | **7.7E-24** | 1.36 (1.29 – 1.44) | 2.5E-02 | 1.42 (1.31 – 1.53) | 1.24 (1.15 – 1.34) |
| 740.1 | Osteoarthritis; localized | 1.0 / 1.8 | 0.58 (0.52 – 0.64) | **1.2E-23** | 0.86 (0.78 – 0.94) | NA | NA | 0.57 (0.49 – 0.67) |
| 599.4 | Urinary incontinence | 2.9 / 2.0 | 1.46 (1.35 – 1.57) | **3.0E-23** | 1.56 (1.43 – 1.69) | 1.1E-01 | 1.32 (1.16 – 1.49) | 1.54 (1.41 – 1.69) |
| 296 | Mood disorders | 1.7 / 1.1 | 1.66 (1.50 – 1.83) | **3.4E-23** | 1.86 (1.67 – 2.07) | 6.4E-01 | 1.62 (1.42 – 1.84) | 1.71 (1.46 – 1.99) |
| 563 | Constipation | 5.1 / 3.7 | 1.33 (1.25 – 1.40) | **1.1E-22** | 1.77 (1.66 – 1.88) | 2.0E-04 | 1.19 (1.10 – 1.28) | 1.49 (1.38 – 1.62) |
| 375.1 | Dry eyes | 2.1 / 1.3 | 1.55 (1.42 – 1.69) | **2.2E-22** | 1.49 (1.34 – 1.64) | 5.9E-01 | 1.60 (1.39 – 1.83) | 1.51 (1.35 – 1.70) |
| 379.2 | Disorders of vitreous body | 2.0 / 1.2 | 1.58 (1.44 – 1.73) | **2.8E-22** | 1.28 (1.15 – 1.42) | 7.8E-02 | 1.69 (1.49 – 1.91) | 1.44 (1.25 – 1.66) |
| 277 | Other disorders of metabolism | 2.7 / 1.9 | 1.45 (1.35 – 1.57) | **7.6E-22** | 1.61 (1.48 – 1.74) | 6.9E-03 | 1.27 (1.12 – 1.43) | 1.59 (1.44 – 1.76) |
| 272.11 | Hypercholesterolemia | 9.1 / 7.4 | 1.24 (1.19 – 1.30) | **2.4E-21** | 1.09 (1.04 – 1.15) | 3.0E-01 | 1.28 (1.21 – 1.36) | 1.19 (1.11 – 1.28) |
| 340 | Migraine | 2.5 / 1.9 | 1.46 (1.35 – 1.58) | **8.1E-21** | 1.58 (1.45 – 1.73) | 9.9E-01 | 1.44 (1.25 – 1.65) | 1.47 (1.33 – 1.61) |
| 272.13 | Mixed hyperlipidemia | 5.3 / 4.1 | 1.29 (1.23 – 1.37) | **8.7E-21** | 1.11 (1.05 – 1.17) | 5.3E-02 | 1.35 (1.26 – 1.44) | 1.19 (1.08 – 1.31) |
| 278.4 | Abnormal weight gain | 2.0 / 1.4 | 1.54 (1.41 – 1.69) | **9.5E-21** | 2.04 (1.86 – 2.23) | 8.9E-02 | 1.75 (1.51 – 2.03) | 1.44 (1.29 – 1.61) |
| 481 | Influenza | 2.2 / 1.5 | 1.49 (1.37 – 1.62) | **2.0E-20** | 2.87 (2.39 – 3.46) | 4.2E-01 | 1.54 (1.37 – 1.73) | 1.44 (1.27 – 1.63) |
| 289.4 | Lymphadenitis | 2.6 / 3.6 | 0.72 (0.67 – 0.77) | **2.2E-20** | 0.70 (0.65 – 0.75) | 3.9E-02 | 0.67 (0.61 – 0.74) | 0.78 (0.70 – 0.87) |
| 475.9 | Postnasal drip | 2.4 / 1.6 | 1.46 (1.35 – 1.58) | **1.2E-19** | 1.56 (1.42 – 1.72) | 4.1E-03 | 1.64 (1.46 – 1.84) | 1.29 (1.15 – 1.45) |
| 701 | Other hypertrophic and atrophic conditions of skin | 3.2 / 2.3 | 1.39 (1.29 – 1.49) | **1.4E-19** | 1.48 (1.37 – 1.61) | 3.6E-03 | 1.53 (1.40 – 1.68) | 1.21 (1.08 – 1.36) |
| 594.1 | Calculus of kidney | 2.9 / 2.1 | 1.40 (1.30 – 1.50) | **5.2E-19** | 1.39 (1.29 – 1.50) | 3.7E-01 | 1.37 (1.25 – 1.49) | 1.46 (1.28 – 1.68) |
| 428.2 | Heart failure NOS | 1.5 / 0.9 | 1.61 (1.45 – 1.79) | **5.6E-19** | 2.42 (2.16 – 2.71) | 1.7E-04 | 1.42 (1.25 – 1.61) | 2.15 (1.78 – 2.61) |
| 304 | Adjustment reaction | 2.5 / 1.7 | 1.46 (1.34 – 1.58) | **5.6E-19** | 1.46 (1.33 – 1.60) | 7.8E-02 | 1.56 (1.39 – 1.75) | 1.35 (1.19 – 1.52) |
| 429.3 | Symptoms involving cardiovascular system | 4.0 / 3.0 | 1.32 (1.24 – 1.40) | **1.6E-18** | 1.55 (1.44 – 1.66) | 3.1E-01 | 1.35 (1.24 – 1.46) | 1.27 (1.16 – 1.40) |
| 402 | Elevated blood pressure reading without diagnosis of hypertension | 3.4 / 2.6 | 1.36 (1.27 – 1.45) | **2.3E-18** | 1.09 (1.01 – 1.18) | 7.8E-03 | 1.49 (1.36 – 1.63) | 1.19 (1.07 – 1.33) |
| 495 | Asthma | 4.4 / 3.8 | 1.30 (1.23 – 1.38) | **6.8E-18** | 1.28 (1.20 – 1.36) | 8.6E-01 | 1.30 (1.19 – 1.42) | 1.30 (1.20 – 1.41) |

| PheCode | Disease | Incidence OSA case / control | Risk ratio | P-value | Sensitivity analysis risk ratio | OSA x sex interaction p-value | Risk ratio (men) | Risk ratio (women) |
|---|---|---|---|---|---|---|---|---|
| 380.4 | Impacted cerumen | 2.7 / 1.8 | 1.41 (1.30 – 1.53) | **8.5E-18** | 1.43 (1.30 – 1.57) | 6.8E-01 | 1.43 (1.30 – 1.58) | 1.37 (1.20 – 1.57) |
| 579.8 | Nonspecific abnormal findings in stool contents | 2.6 / 1.8 | 1.41 (1.30 – 1.52) | **8.9E-18** | 1.39 (1.27 – 1.53) | 7.1E-01 | 1.43 (1.28 – 1.58) | 1.38 (1.23 – 1.55) |
| 764 | Sciatica | 4.0 / 3.0 | 1.31 (1.23 – 1.39) | **2.7E-17** | 1.41 (1.32 – 1.51) | 7.7E-01 | 1.33 (1.22 – 1.44) | 1.29 (1.18 – 1.42) |
| 292.2 | Mild cognitive impairment | 1.2 / 0.7 | 1.66 (1.48 – 1.87) | **5.5E-17** | 2.04 (1.77 – 2.36) | 3.8E-01 | 1.73 (1.48 – 2.03) | 1.57 (1.31 – 1.88) |
| 250.41 | Impaired fasting glucose | 2.4 / 1.7 | 1.41 (1.30 – 1.53) | **9.0E-17** | 1.43 (1.31 – 1.56) | 2.2E-01 | 1.48 (1.33 – 1.65) | 1.33 (1.18 – 1.50) |
| 389.1 | Sensorineural hearing loss | 3.3 / 2.4 | 1.34 (1.25 – 1.44) | **1.0E-16** | 1.35 (1.24 – 1.46) | 1.2E-01 | 1.39 (1.28 – 1.52) | 1.25 (1.12 – 1.40) |
| 726 | Peripheral enthesopathies and allied syndromes | 3.2 / 2.3 | 1.36 (1.26 – 1.46) | **1.6E-16** | 1.20 (1.10 – 1.30) | 4.2E-01 | 1.41 (1.29 – 1.54) | 1.28 (1.13 – 1.44) |
| 912 | Insect bite | 1.2 / 0.7 | 1.65 (1.46 – 1.86) | **1.9E-16** | 2.26 (1.96 – 2.61) | 6.0E-01 | 1.61 (1.37 – 1.89) | 1.70 (1.43 – 2.02) |
| 508 | Pulmonary collapse; interstitial and compensatory emphysema | 3.4 / 4.3 | 0.77 (0.72 – 0.82) | **4.1E-16** | 0.75 (0.70 – 0.80) | 4.0E-02 | 0.73 (0.68 – 0.79) | 0.84 (0.76 – 0.94) |
| 797 | Shock | 1.2 / 0.7 | 1.63 (1.45 – 1.83) | **7.1E-16** | 1.23 (1.11 – 1.36) | NA | 1.51 (1.31 – 1.74) | NA |
| 599.8 | Other symptoms involving urinary system | 2.5 / 1.7 | 1.38 (1.28 – 1.50) | **9.5E-16** | 1.89 (1.72 – 2.08) | 5.2E-01 | 1.35 (1.23 – 1.48) | 1.47 (1.27 – 1.71) |
| 427.6 | Premature beats | 2.0 / 1.3 | 1.44 (1.31 – 1.57) | **1.6E-15** | 1.48 (1.33 – 1.63) | 1.8E-01 | 1.49 (1.34 – 1.65) | 1.32 (1.12 – 1.55) |
| 765 | Cervical radiculitis | 2.2 / 1.6 | 1.41 (1.29 – 1.53) | **1.8E-15** | 1.61 (1.46 – 1.78) | 2.0E-01 | 1.35 (1.21 – 1.51) | 1.49 (1.31 – 1.69) |
| 519.8 | Other diseases of respiratory system, NEC | 1.4 / 0.9 | 1.55 (1.39 – 1.73) | **2.0E-15** | 2.00 (1.78 – 2.26) | 5.8E-01 | 1.51 (1.30 – 1.75) | 1.60 (1.36 – 1.88) |
| 401.1 | Essential hypertension | 11.9 / 17.3 | 0.84 (0.81 – 0.88) | **2.1E-15** | 0.77 (0.74 – 0.81) | 5.7E-02 | 0.85 (0.81 – 0.89) | 0.86 (0.80 – 0.92) |
| 427.9 | Palpitations | 4.3 / 3.4 | 1.29 (1.21 – 1.37) | **2.3E-15** | 1.35 (1.26 – 1.45) | 8.6E-01 | 1.29 (1.18 – 1.41) | 1.28 (1.18 – 1.40) |
| 386.9 | Dizziness and giddiness (Light-headedness and vertigo) | 6.1 / 4.9 | 1.24 (1.17 – 1.30) | **2.9E-15** | 1.31 (1.24 – 1.39) | 7.2E-01 | 1.25 (1.16 – 1.34) | 1.22 (1.13 – 1.32) |
| 726.1 | Enthesopathy | 3.9 / 3.0 | 1.30 (1.21 – 1.38) | **3.6E-15** | 1.28 (1.20 – 1.38) | 9.0E-02 | 1.38 (1.26 – 1.52) | 1.22 (1.12 – 1.34) |
| 110.11 | Dermatophytosis of nail | 1.7 / 1.1 | 1.48 (1.34 – 1.63) | **5.1E-15** | 1.36 (1.22 – 1.51) | 2.5E-02 | 1.60 (1.42 – 1.80) | 1.24 (1.04 – 1.47) |
| 507 | Pleurisy; pleural effusion | 2.7 / 3.5 | 0.76 (0.71 – 0.82) | **1.2E-14** | 0.66 (0.61 – 0.72) | 9.3E-01 | 0.76 (0.69 – 0.82) | 0.76 (0.68 – 0.86) |
| 537 | Other disorders of stomach and duodenum | 2.9 / 2.2 | 1.33 (1.24 – 1.43) | **1.4E-14** | 1.42 (1.32 – 1.53) | 5.2E-03 | 1.21 (1.10 – 1.33) | 1.49 (1.34 – 1.67) |
| 740.9 | Osteoarthrosis NOS | 6.1 / 5.2 | 1.23 (1.17 – 1.30) | **3.4E-14** | 1.16 (1.10 – 1.22) | 7.7E-01 | 1.24 (1.15 – 1.33) | 1.22 (1.13 – 1.32) |
| 728.7 | Fasciitis | 1.8 / 1.3 | 1.43 (1.30 – 1.58) | **8.5E-14** | 1.41 (1.27 – 1.56) | 4.7E-02 | 1.58 (1.38 – 1.80) | 1.30 (1.13 – 1.49) |
| 527.7 | Disturbance of salivary | 1.3 / 0.8 | 1.53 (1.37 – 1.71) | **1.5E-13** | 2.27 (1.94 – 2.66) | NA | NA | 1.69 (1.45 – 1.98) |

| PheCode | Disease | Incidence OSA case / control | Risk ratio | P-value | Sensitivity analysis risk ratio | OSA x sex interaction p-value | Risk ratio (men) | Risk ratio (women) |
|---|---|---|---|---|---|---|---|---|
| | secretion | | | | | | | |
| 464 | Acute sinusitis | 1.7 / 1.2 | 1.45 (1.31 – 1.60) | **1.9E-13** | 1.41 (1.26 – 1.58) | 6.9E-04 | 1.75 (1.52 – 2.03) | 1.22 (1.07 – 1.40) |
| 596.5 | Functional disorders of bladder | 1.2 / 0.7 | 1.56 (1.38 – 1.75) | **2.1E-13** | 1.39 (1.22 – 1.59) | 8.3E-01 | 1.51 (1.28 – 1.78) | 1.60 (1.35 – 1.89) |
| 382 | Otalgia | 3.1 / 2.4 | 1.30 (1.21 – 1.39) | **3.9E-13** | 1.18 (1.09 – 1.28) | 1.2E-02 | 1.43 (1.29 – 1.58) | 1.19 (1.08 – 1.31) |
| 586.4 | Stricture/obstruction of ureter | 1.5 / 0.9 | 1.46 (1.32 – 1.62) | **7.3E-13** | 1.21 (1.08 – 1.36) | NA | 1.52 (1.37 – 1.69) | NA |
| 296.2 | Depression | 2.2 / 1.7 | 1.39 (1.27 – 1.52) | **9.4E-13** | 2.02 (1.83 – 2.23) | 9.5E-01 | 1.40 (1.23 – 1.58) | 1.38 (1.21 – 1.57) |
| 228.1 | Hemangioma of skin and subcutaneous tissue | 2.3 / 1.7 | 1.34 (1.24 – 1.46) | **2.2E-12** | 1.38 (1.25 – 1.51) | 1.0E-03 | 1.48 (1.34 – 1.64) | 1.12 (0.97 – 1.29) |
| 512.9 | Other dyspnea | 6.5 / 5.2 | 1.21 (1.15 – 1.28) | **2.3E-12** | 1.99 (1.88 – 2.10) | 1.1E-02 | 1.15 (1.07 – 1.23) | 1.32 (1.21 – 1.44) |
| 261.4 | Vitamin D deficiency | 4.7 / 4.0 | 1.23 (1.16 – 1.30) | **2.3E-12** | 1.44 (1.36 – 1.53) | 1.0E-02 | 1.33 (1.22 – 1.45) | 1.15 (1.07 – 1.24) |
| 807 | Fracture of ribs | 1.1 / 0.7 | 1.55 (1.37 – 1.76) | **3.0E-12** | 1.16 (1.01 – 1.35) | 3.5E-03 | 1.37 (1.17 – 1.60) | 1.95 (1.59 – 2.40) |
| 173 | Neoplasm of uncertain behavior of skin | 2.4 / 1.7 | 1.34 (1.23 – 1.46) | **3.1E-12** | 1.65 (1.51 – 1.81) | 6.6E-01 | 1.31 (1.18 – 1.45) | 1.38 (1.21 – 1.59) |
| 290.1 | Dementias | 2.0 / 1.3 | 1.37 (1.25 – 1.49) | **4.0E-12** | 1.58 (1.42 – 1.76) | 2.5E-03 | 1.52 (1.36 – 1.71) | 1.16 (1.01 – 1.33) |
| 741.2 | Stiffness of joint | 1.2 / 0.8 | 1.51 (1.34 – 1.70) | **4.3E-12** | 1.29 (1.13 – 1.47) | 3.6E-01 | 1.60 (1.36 – 1.89) | 1.42 (1.21 – 1.68) |
| 465 | Acute upper respiratory infections of multiple or unspecified sites | 3.8 / 3.1 | 1.27 (1.18 – 1.36) | **1.3E-11** | 1.55 (1.44 – 1.67) | 3.6E-02 | 1.36 (1.24 – 1.50) | 1.17 (1.05 – 1.29) |
| 380.1 | Otitis externa | 1.3 / 0.9 | 1.46 (1.31 – 1.63) | **1.5E-11** | 1.65 (1.46 – 1.87) | 2.7E-01 | 1.54 (1.34 – 1.77) | 1.35 (1.13 – 1.61) |
| 365 | Glaucoma | 2.7 / 2.0 | 1.29 (1.20 – 1.39) | **3.2E-11** | 1.33 (1.23 – 1.45) | 9.1E-01 | 1.29 (1.17 – 1.43) | 1.29 (1.15 – 1.44) |
| 760 | Back pain | 8.5 / 7.5 | 1.17 (1.12 – 1.23) | **3.9E-11** | 1.15 (1.10 – 1.21) | 4.0E-01 | 1.17 (1.10 – 1.24) | 1.18 (1.10 – 1.28) |
| 386.2 | Peripheral or central vertigo | 1.5 / 1.0 | 1.42 (1.28 – 1.57) | **4.1E-11** | 1.50 (1.33 – 1.69) | 5.2E-01 | 1.36 (1.16 – 1.59) | 1.46 (1.28 – 1.68) |
| 292.1 | Aphasia/speech disturbance | 1.3 / 0.9 | 1.45 (1.30 – 1.63) | **5.7E-11** | 1.37 (1.21 – 1.55) | 2.0E-01 | 1.53 (1.33 – 1.76) | 1.34 (1.11 – 1.61) |
| 704 | Diseases of hair and hair follicles | 1.5 / 1.1 | 1.40 (1.27 – 1.55) | **6.5E-11** | 1.60 (1.44 – 1.79) | 3.8E-01 | 1.44 (1.28 – 1.63) | 1.31 (1.10 – 1.57) |
| 704.1 | Alopecia | 1.7 / 1.3 | 1.37 (1.25 – 1.50) | **7.2E-11** | 1.43 (1.28 – 1.60) | NA | NA | 1.26 (1.13 – 1.41) |
| 840 | Sprains and strains | 3.9 / 3.2 | 1.24 (1.16 – 1.32) | **8.8E-11** | 1.12 (1.04 – 1.20) | 6.4E-01 | 1.27 (1.17 – 1.37) | 1.20 (1.08 – 1.33) |
| 278.11 | Morbid obesity | 3.3 / 4.1 | 1.29 (1.19 – 1.39) | **9.0E-11** | 1.52 (1.42 – 1.63) | **1.4E-07** | 1.74 (1.56 – 1.94) | 1.01 (0.92 – 1.12) |
| 742.9 | Other derangement of joint | 2.5 / 2.0 | 1.29 (1.19 – 1.39) | **1.2E-10** | 1.33 (1.23 – 1.45) | 1.7E-02 | 1.17 (1.04 – 1.31) | 1.40 (1.26 – 1.55) |
| 798 | Malaise and fatigue | 9.4 / 7.9 | 1.16 (1.11 – 1.21) | **1.3E-10** | 1.48 (1.41 – 1.56) | 7.4E-01 | 1.14 (1.08 – 1.21) | 1.18 (1.10 – 1.27) |
| 809 | Fracture of unspecified bones | 1.8 / 2.3 | 0.76 (0.70 – 0.83) | **1.4E-10** | 0.85 (0.78 – 0.92) | 1.3E-01 | 0.72 (0.64 – 0.81) | 0.80 (0.71 – 0.91) |

| PheCode | Disease | Incidence OSA case / control | Risk ratio | P-value | Sensitivity analysis risk ratio | OSA x sex interaction p-value | Risk ratio (men) | Risk ratio (women) |
|---|---|---|---|---|---|---|---|---|
| 440.9 | Atherosclerosis of aorta | 1.2 / 0.8 | 1.46 (1.30 – 1.64) | **2.6E-10** | 1.63 (1.43 – 1.86) | 8.2E-01 | 1.43 (1.22 – 1.67) | 1.48 (1.24 – 1.76) |
| 350.1 | Abnormal involuntary movements | 2.6 / 2.0 | 1.28 (1.18 – 1.38) | **3.0E-10** | 1.57 (1.44 – 1.71) | 5.4E-01 | 1.25 (1.13 – 1.38) | 1.31 (1.17 – 1.47) |
| 427.3 | Other specified cardiac dysrhythmias | 3.8 / 2.9 | 1.23 (1.15 – 1.31) | **3.3E-10** | 1.33 (1.24 – 1.43) | 1.8E-01 | 1.25 (1.16 – 1.35) | 1.17 (1.04 – 1.31) |
| 593.2 | Microscopic hematuria | 1.8 / 1.3 | 1.34 (1.22 – 1.46) | **5.5E-10** | 1.18 (1.07 – 1.32) | 2.4E-01 | 1.27 (1.12 – 1.45) | 1.41 (1.24 – 1.61) |
| 285 | Other anemias | 6.7 / 5.7 | 1.17 (1.11 – 1.23) | **5.9E-10** | 1.29 (1.23 – 1.37) | 7.0E-04 | 1.07 (1.00 – 1.14) | 1.32 (1.22 – 1.43) |
| 415.2 | Chronic pulmonary heart disease | 1.2 / 0.9 | 1.42 (1.27 – 1.59) | **7.5E-10** | 1.99 (1.75 – 2.26) | 9.1E-04 | 1.19 (1.02 – 1.39) | 1.74 (1.48 – 2.05) |
| 536.8 | Dyspepsia and other specified disorders of function of stomach | 1.4 / 1.0 | 1.39 (1.25 – 1.55) | **7.7E-10** | 1.26 (1.12 – 1.42) | 7.3E-02 | 1.53 (1.32 – 1.77) | 1.26 (1.08 – 1.47) |
| 480.11 | Pneumococcal pneumonia | 1.4 / 1.0 | 1.39 (1.25 – 1.54) | **8.1E-10** | 1.75 (1.55 – 1.97) | 9.3E-01 | 1.37 (1.20 – 1.57) | 1.41 (1.20 – 1.67) |
| 512.2 | Painful respiration | 3.0 / 2.3 | 1.24 (1.16 – 1.34) | **2.5E-09** | 1.28 (1.18 – 1.38) | 8.1E-01 | 1.23 (1.12 – 1.36) | 1.25 (1.13 – 1.40) |
| 740.11 | Osteoarthrosis, localized, primary | 6.0 / 6.9 | 0.86 (0.82 – 0.91) | **3.4E-09** | 0.82 (0.79 – 0.87) | 3.3E-02 | 0.92 (0.86 – 0.98) | 0.81 (0.75 – 0.87) |
| 394.7 | Disease of tricuspid valve | 1.3 / 0.9 | 1.40 (1.25 – 1.57) | **4.8E-09** | 1.22 (1.07 – 1.38) | 2.8E-02 | 1.25 (1.07 – 1.45) | 1.62 (1.37 – 1.92) |
| 561 | Symptoms involving digestive system | 2.0 / 1.5 | 1.30 (1.19 – 1.42) | **4.9E-09** | 1.50 (1.35 – 1.66) | 4.7E-01 | 1.25 (1.09 – 1.43) | 1.34 (1.19 – 1.51) |
| 965 | Poisoning by analgesics, antipyretics, and antirheumatics | 2.2 / 1.7 | 1.28 (1.18 – 1.39) | **5.3E-09** | 1.40 (1.29 – 1.53) | 9.3E-01 | 1.28 (1.12 – 1.46) | 1.28 (1.15 – 1.42) |
| 695.3 | Rosacea | 1.1 / 0.8 | 1.42 (1.26 – 1.60) | **6.1E-09** | 1.26 (1.10 – 1.43) | NA | NA | 1.36 (1.16 – 1.60) |
| 727.1 | Synovitis and tenosynovitis | 2.0 / 1.5 | 1.30 (1.19 – 1.42) | **9.6E-09** | 1.14 (1.04 – 1.26) | **1.8E-05** | 1.56 (1.38 – 1.77) | 1.05 (0.93 – 1.20) |
| 781 | Symptoms involving nervous and musculoskeletal systems | 2.6 / 2.0 | 1.25 (1.16 – 1.35) | **1.0E-08** | 1.88 (1.72 – 2.05) | 4.9E-02 | 1.16 (1.05 – 1.29) | 1.37 (1.22 – 1.53) |
| 261.2 | Vitamin B-complex deficiencies | 1.3 / 0.9 | 1.38 (1.24 – 1.55) | **1.1E-08** | 1.58 (1.40 – 1.79) | 3.3E-01 | 1.27 (1.08 – 1.49) | 1.49 (1.28 – 1.74) |
| 766 | Neuralgia, neuritis, and radiculitis NOS | 1.7 / 1.3 | 1.32 (1.20 – 1.45) | **1.2E-08** | 1.36 (1.22 – 1.51) | 3.5E-01 | 1.36 (1.20 – 1.55) | 1.26 (1.09 – 1.44) |
| 351 | Other peripheral nerve disorders | 3.0 / 2.4 | 1.23 (1.15 – 1.33) | **2.2E-08** | 1.12 (1.03 – 1.21) | 2.9E-04 | 1.40 (1.27 – 1.54) | 1.05 (0.94 – 1.17) |
| 427.11 | Paroxysmal supraventricular tachycardia | 1.9 / 1.5 | 1.29 (1.18 – 1.41) | **2.2E-08** | 1.52 (1.38 – 1.68) | 9.4E-01 | 1.28 (1.13 – 1.44) | 1.30 (1.14 – 1.48) |

| PheCode | Disease | Incidence OSA case / control | Risk ratio | P-value | Sensitivity analysis risk ratio | OSA x sex interaction p-value | Risk ratio (men) | Risk ratio (women) |
|---|---|---|---|---|---|---|---|---|
| 459.9 | Circulatory disease NEC | 2.4 / 1.8 | 1.26 (1.16 – 1.36) | **2.6E-08** | 1.48 (1.35 – 1.61) | 5.2E-03 | 1.36 (1.24 – 1.50) | 1.07 (0.93 – 1.23) |
| 599.2 | Retention of urine | 2.6 / 2.0 | 1.23 (1.14 – 1.33) | **6.4E-08** | 1.18 (1.08 – 1.28) | 2.1E-02 | 1.17 (1.07 – 1.27) | 1.52 (1.28 – 1.81) |
| 395.1 | Nonrheumatic mitral valve disorders | 2.1 / 1.6 | 1.27 (1.16 – 1.39) | **8.1E-08** | 1.13 (1.03 – 1.25) | 9.6E-01 | 1.27 (1.13 – 1.43) | 1.27 (1.11 – 1.46) |
| 565 | Anal and rectal conditions | 1.5 / 1.1 | 1.32 (1.19 – 1.46) | **8.1E-08** | 1.21 (1.08 – 1.35) | 2.6E-01 | 1.26 (1.11 – 1.44) | 1.41 (1.20 – 1.67) |
| 389.4 | Tinnitus | 2.6 / 2.0 | 1.23 (1.14 – 1.33) | **8.5E-08** | 1.25 (1.15 – 1.37) | 2.3E-02 | 1.33 (1.21 – 1.47) | 1.10 (0.97 – 1.24) |
| 276.5 | Hypovolemia | 3.7 / 4.2 | 0.85 (0.80 – 0.90) | **1.2E-07** | 0.80 (0.74 – 0.85) | 7.0E-01 | 0.85 (0.79 – 0.92) | 0.84 (0.76 – 0.93) |
| 425.1 | Primary/intrinsic cardiomyopathies | 1.1 / 1.5 | 0.75 (0.67 – 0.83) | **1.3E-07** | 0.89 (0.80 – 0.99) | NA | 0.66 (0.58 – 0.75) | NA |
| 696.41 | Psoriasis vulgaris | 1.1 / 0.8 | 1.37 (1.22 – 1.54) | **1.7E-07** | 1.26 (1.11 – 1.43) | 7.4E-01 | 1.37 (1.17 – 1.60) | 1.39 (1.15 – 1.67) |
| 530.9 | Heartburn | 2.1 / 1.6 | 1.25 (1.15 – 1.37) | **1.9E-07** | 1.31 (1.19 – 1.44) | 3.2E-01 | 1.30 (1.15 – 1.48) | 1.21 (1.08 – 1.36) |
| 136 | Other infectious and parasitic diseases | 2.1 / 1.6 | 1.25 (1.15 – 1.36) | **2.0E-07** | 1.45 (1.33 – 1.58) | 1.1E-03 | 1.10 (0.98 – 1.23) | 1.48 (1.30 – 1.67) |
| 401.21 | Hypertensive heart disease | 1.7 / 1.2 | 1.29 (1.17 – 1.42) | **2.1E-07** | 2.03 (1.83 – 2.25) | 2.5E-02 | 1.19 (1.05 – 1.35) | 1.45 (1.25 – 1.68) |
| 366.2 | Senile cataract | 3.6 / 2.9 | 1.18 (1.11 – 1.26) | **4.6E-07** | 1.16 (1.08 – 1.25) | 1.4E-01 | 1.22 (1.12 – 1.33) | 1.12 (1.01 – 1.23) |
| 244.2 | Acquired hypothyroidism | 1.5 / 1.2 | 1.29 (1.17 – 1.43) | **5.7E-07** | 1.45 (1.30 – 1.62) | NA | NA | 1.25 (1.11 – 1.42) |
| 269 | Proteinuria | 1.6 / 1.2 | 1.28 (1.16 – 1.42) | **6.3E-07** | 1.31 (1.18 – 1.45) | 2.1E-01 | 1.35 (1.20 – 1.53) | 1.17 (0.99 – 1.38) |
| 496.1 | Emphysema | 1.4 / 1.0 | 1.30 (1.17 – 1.44) | **1.1E-06** | 1.44 (1.27 – 1.62) | 4.0E-01 | 1.24 (1.08 – 1.43) | 1.38 (1.17 – 1.62) |
| 530.14 | Reflux esophagitis | 1.3 / 0.9 | 1.32 (1.18 – 1.48) | **1.1E-06** | 1.58 (1.39 – 1.80) | 4.7E-02 | 1.46 (1.26 – 1.71) | 1.17 (0.99 – 1.38) |
| 054 | Herpes simplex | 1.3 / 1.0 | 1.31 (1.17 – 1.46) | **1.5E-06** | 1.33 (1.17 – 1.51) | NA | NA | 1.31 (1.14 – 1.51) |
| 008 | Intestinal infection | 1.9 / 1.5 | 1.24 (1.14 – 1.35) | **1.6E-06** | 1.41 (1.28 – 1.56) | 1.3E-02 | 1.38 (1.22 – 1.56) | 1.11 (0.98 – 1.26) |
| 286.9 | Abnormal coagulation profile | 1.2 / 0.9 | 1.31 (1.17 – 1.47) | **1.8E-06** | 1.59 (1.42 – 1.78) | 4.3E-01 | 1.26 (1.09 – 1.46) | 1.39 (1.17 – 1.66) |
| 172.21 | Basal cell carcinoma | 3.0 / 2.4 | 1.18 (1.10 – 1.27) | **2.8E-06** | 1.13 (1.05 – 1.22) | 9.4E-01 | 1.18 (1.08 – 1.28) | 1.18 (1.05 – 1.34) |
| 369.5 | Conjunctivitis, infectious | 1.7 / 1.3 | 1.26 (1.14 – 1.38) | **2.9E-06** | 1.63 (1.46 – 1.81) | 3.4E-01 | 1.33 (1.15 – 1.52) | 1.20 (1.05 – 1.37) |
| 565.1 | Anal and rectal polyp | 1.1 / 0.9 | 1.30 (1.16 – 1.47) | **8.7E-06** | 1.19 (1.04 – 1.35) | NA | 1.37 (1.19 – 1.58) | NA |
| 418.1 | Precordial pain | 1.5 / 1.2 | 1.25 (1.13 – 1.38) | **9.2E-06** | 1.48 (1.33 – 1.65) | 2.6E-01 | 1.20 (1.05 – 1.36) | 1.34 (1.15 – 1.56) |
| 280.1 | Iron deficiency anemias, unspecified or not due to blood loss | 2.3 / 1.9 | 1.20 (1.11 – 1.30) | **9.9E-06** | 1.45 (1.33 – 1.58) | 3.3E-01 | 1.11 (0.98 – 1.26) | 1.27 (1.14 – 1.42) |
| 483 | Acute bronchitis and | 1.7 / 1.4 | 1.24 (1.12 – 1.36) | **1.4E-05** | 1.37 (1.23 – 1.52) | 1.5E-03 | 1.43 (1.26 – 1.63) | 1.03 (0.89 – 1.19) |

| PheCode | Disease | Incidence OSA case / control | Risk ratio | P-value | Sensitivity analysis risk ratio | OSA x sex interaction p-value | Risk ratio (men) | Risk ratio (women) |
|---|---|---|---|---|---|---|---|---|
| | bronchiolitis | | | | | | | |
| 585.3 | Chronic renal failure [CKD] | 3.8 / 3.2 | 1.14 (1.08 – 1.22) | **1.6E-05** | 1.52 (1.42 – 1.62) | 4.8E-04 | 1.06 (0.98 – 1.14) | 1.34 (1.20 – 1.50) |
| 550.4 | Umbilical hernia | 1.1 / 0.9 | 1.28 (1.14 – 1.44) | **2.9E-05** | 1.50 (1.33 – 1.68) | NA | 1.29 (1.13 – 1.48) | NA |
| 510 | Other diseases of lung | 1.6 / 1.3 | 1.24 (1.12 – 1.37) | **2.9E-05** | 1.43 (1.28 – 1.60) | 2.0E-02 | 1.12 (0.99 – 1.28) | 1.44 (1.23 – 1.69) |
| 562.2 | Diverticulitis | 1.7 / 1.4 | 1.22 (1.11 – 1.33) | **3.2E-05** | 1.17 (1.06 – 1.29) | 2.8E-01 | 1.29 (1.14 – 1.47) | 1.13 (0.99 – 1.30) |
| 290.2 | Delirium due to conditions classified elsewhere | 1.3 / 1.0 | 1.26 (1.13 – 1.40) | **3.2E-05** | 1.34 (1.20 – 1.50) | 5.4E-03 | 1.14 (1.00 – 1.30) | 1.56 (1.28 – 1.89) |
| 480 | Pneumonia | 4.4 / 3.8 | 1.13 (1.07 – 1.20) | **4.2E-05** | 1.45 (1.36 – 1.55) | 4.7E-01 | 1.10 (1.03 – 1.19) | 1.17 (1.06 – 1.29) |
| 585.4 | Chronic kidney disease, Stage I or II | 1.2 / 0.9 | 1.26 (1.13 – 1.41) | **4.6E-05** | 1.66 (1.46 – 1.87) | 6.4E-02 | 1.17 (1.02 – 1.35) | 1.46 (1.21 – 1.77) |
| 720 | Spinal stenosis | 2.1 / 1.7 | 1.19 (1.09 – 1.30) | **5.5E-05** | 1.30 (1.19 – 1.42) | 4.5E-01 | 1.17 (1.05 – 1.30) | 1.23 (1.07 – 1.40) |
| 367.4 | Presbyopia | 1.1 / 0.9 | 1.27 (1.13 – 1.42) | **5.6E-05** | 1.55 (1.37 – 1.76) | 8.6E-01 | 1.27 (1.09 – 1.49) | 1.26 (1.06 – 1.49) |
| 316 | Substance addiction and disorders | 1.8 / 1.6 | 1.20 (1.10 – 1.31) | **6.4E-05** | 1.36 (1.24 – 1.49) | **3.4E-10** | 0.99 (0.89 – 1.10) | 1.86 (1.58 – 2.18) |
| 743.11 | Osteoporosis NOS | 3.0 / 2.5 | 1.15 (1.08 – 1.24) | **6.7E-05** | 1.25 (1.15 – 1.36) | 7.5E-03 | 1.37 (1.18 – 1.58) | 1.09 (1.01 – 1.18) |
| 217.1 | Nevus, non-neoplastic | 2.2 / 1.9 | 1.18 (1.09 – 1.28) | **7.0E-05** | 1.24 (1.13 – 1.36) | 2.6E-04 | 1.35 (1.21 – 1.50) | 0.99 (0.87 – 1.12) |
| 528 | Diseases of the oral soft tissues, excluding lesions specific for gingiva and tongue | 1.2 / 1.0 | 1.25 (1.12 – 1.40) | **7.1E-05** | 1.58 (1.38 – 1.80) | 4.5E-01 | 1.30 (1.12 – 1.50) | 1.18 (1.00 – 1.40) |
| 571.5 | Other chronic nonalcoholic liver disease | 2.1 / 1.9 | 1.18 (1.09 – 1.29) | **7.3E-05** | 1.20 (1.10 – 1.30) | 9.9E-02 | 1.29 (1.15 – 1.44) | 1.08 (0.95 – 1.22) |
| 573.7 | Abnormal results of function study of liver | 2.1 / 1.8 | 1.19 (1.09 – 1.29) | **8.2E-05** | 1.36 (1.24 – 1.49) | 2.5E-01 | 1.25 (1.12 – 1.39) | 1.10 (0.97 – 1.26) |
| 687.1 | Rash and other nonspecific skin eruption | 4.6 / 4.1 | 1.12 (1.06 – 1.19) | **9.4E-05** | 1.71 (1.60 – 1.82) | 5.6E-02 | 1.06 (0.97 – 1.14) | 1.19 (1.10 – 1.30) |
| 727 | Other disorders of synovium, tendon, and bursa | 1.3 / 1.1 | 1.23 (1.11 – 1.37) | **1.1E-04** | 1.57 (1.38 – 1.77) | 7.6E-01 | 1.26 (1.09 – 1.46) | 1.21 (1.04 – 1.40) |
| 770 | Myalgia and myositis unspecified | 2.0 / 1.8 | 1.19 (1.09 – 1.30) | **1.2E-04** | 1.49 (1.36 – 1.64) | 6.1E-01 | 1.20 (1.04 – 1.39) | 1.18 (1.05 – 1.31) |
| 110.12 | Althete's foot | 1.0 / 0.8 | 1.27 (1.13 – 1.44) | **1.2E-04** | 1.24 (1.08 – 1.42) | NA | 1.44 (1.25 – 1.67) | NA |
| 274.11 | Gouty arthropathy | 1.3 / 1.1 | 1.22 (1.10 – 1.36) | 1.9E-04 | 1.24 (1.12 – 1.39) | NA | 1.24 (1.11 – 1.39) | NA |
| 550.1 | Inguinal hernia | 1.9 / 1.5 | 1.18 (1.08 – 1.29) | 2.2E-04 | 1.34 (1.20 – 1.49) | NA | 1.20 (1.10 – 1.32) | NA |
| 371.3 | Inflammation of eyelids | 1.4 / 1.2 | 1.21 (1.09 – 1.34) | 2.3E-04 | 1.38 (1.24 – 1.55) | 8.7E-03 | 1.08 (0.94 – 1.24) | 1.40 (1.20 – 1.63) |

| PheCode | Disease | Incidence OSA case / control | Risk ratio | P-value | Sensitivity analysis risk ratio | OSA x sex interaction p-value | Risk ratio (men) | Risk ratio (women) |
|---|---|---|---|---|---|---|---|---|
| 414 | Other forms of chronic heart disease | 1.4 / 1.1 | 1.22 (1.10 – 1.35) | 2.3E-04 | 1.72 (1.54 – 1.93) | NA | 1.25 (1.10 – 1.41) | NA |
| 690.1 | Seborrheic dermatitis | 1.3 / 1.1 | 1.22 (1.10 – 1.36) | 2.4E-04 | 1.36 (1.21 – 1.54) | 6.6E-03 | 1.36 (1.19 – 1.55) | 1.00 (0.83 – 1.20) |
| 772.2 | Spasm of muscle | 1.2 / 1.0 | 1.23 (1.10 – 1.37) | 2.5E-04 | 2.05 (1.80 – 2.34) | NA | NA | 1.38 (1.19 – 1.61) |
| 561.1 | Diarrhea | 4.4 / 3.9 | 1.12 (1.05 – 1.19) | 3.0E-04 | 1.40 (1.31 – 1.49) | 1.8E-01 | 1.07 (0.98 – 1.16) | 1.17 (1.07 – 1.28) |
| 426.31 | Right bundle branch block | 1.3 / 1.1 | 1.21 (1.09 – 1.35) | 3.5E-04 | 1.29 (1.15 – 1.45) | NA | 1.28 (1.13 – 1.44) | NA |
| 427.22 | Atrial flutter | 1.6 / 1.3 | 1.19 (1.08 – 1.31) | 4.3E-04 | 1.21 (1.09 – 1.35) | NA | 1.22 (1.10 – 1.37) | NA |
| 053 | Herpes zoster | 2.5 / 2.1 | 1.15 (1.06 – 1.24) | 5.0E-04 | 1.27 (1.17 – 1.39) | 2.9E-02 | 1.24 (1.11 – 1.38) | 1.06 (0.95 – 1.18) |
| 350.2 | Abnormality of gait | 3.2 / 2.7 | 1.12 (1.05 – 1.20) | 7.7E-04 | 1.19 (1.10 – 1.29) | 9.1E-01 | 1.12 (1.03 – 1.22) | 1.12 (1.01 – 1.25) |
| 250.42 | Other abnormal glucose | 4.8 / 4.3 | 1.10 (1.04 – 1.16) | 7.9E-04 | 1.29 (1.22 – 1.37) | 8.1E-01 | 1.11 (1.03 – 1.19) | 1.09 (1.00 – 1.19) |
| 574.3 | Cholecystitis without cholelithiasis | 1.1 / 0.9 | 1.22 (1.08 – 1.37) | 1.0E-03 | 1.35 (1.19 – 1.53) | 2.4E-03 | 1.41 (1.21 – 1.65) | 1.01 (0.84 – 1.21) |
| 704.8 | Other specified diseases of hair and hair follicles | 1.1 / 0.9 | 1.21 (1.08 – 1.36) | 1.0E-03 | 1.42 (1.24 – 1.62) | NA | 1.37 (1.19 – 1.57) | NA |
| 793.2 | Nonspecific abnormal findings on radiological and other examination of other intrathoracic organs (echocardiogram, etc) | 1.8 / 1.5 | 1.16 (1.06 – 1.27) | 1.2E-03 | 1.29 (1.18 – 1.42) | 4.2E-01 | 1.20 (1.06 – 1.35) | 1.12 (0.98 – 1.28) |
| 840.3 | Joint/ligament sprain | 1.7 / 1.5 | 1.16 (1.06 – 1.28) | 1.3E-03 | 1.17 (1.05 – 1.31) | 2.7E-01 | 1.23 (1.09 – 1.39) | 1.09 (0.95 – 1.25) |
| 501 | Pneumonitis due to inhalation of food or vomitus | 1.1 / 0.8 | 1.21 (1.08 – 1.36) | 1.5E-03 | 1.47 (1.27 – 1.69) | NA | 1.16 (1.01 – 1.33) | NA |
| 274.1 | Gout | 2.5 / 2.2 | 1.13 (1.05 – 1.22) | 1.6E-03 | 1.23 (1.14 – 1.33) | 8.9E-02 | 1.11 (1.02 – 1.21) | 1.25 (1.04 – 1.50) |
| 512.8 | Cough | 8.0 / 7.5 | 1.08 (1.03 – 1.13) | 2.3E-03 | 1.26 (1.19 – 1.32) | 1.0E-01 | 1.04 (0.98 – 1.11) | 1.13 (1.05 – 1.21) |
| 505 | Other pulmonary inflamation or edema | 1.2 / 0.9 | 1.19 (1.06 – 1.33) | 3.2E-03 | 1.44 (1.27 – 1.62) | 5.9E-03 | 1.05 (0.91 – 1.22) | 1.45 (1.21 – 1.75) |
| 458.1 | Orthostatic hypotension | 1.1 / 0.9 | 1.19 (1.06 – 1.33) | 3.6E-03 | 1.26 (1.11 – 1.43) | NA | 1.13 (0.98 – 1.30) | NA |
| 280.2 | Iron deficiency anemia secondary to blood loss (chronic) | 1.2 / 1.0 | 1.17 (1.05 – 1.31) | 4.9E-03 | 1.40 (1.24 – 1.59) | 1.8E-01 | 1.24 (1.06 – 1.45) | 1.10 (0.94 – 1.29) |
| 599.3 | Dysuria | 2.9 / 2.6 | 1.11 (1.03 – 1.19) | 5.1E-03 | 1.50 (1.39 – 1.63) | 2.3E-01 | 1.16 (1.04 – 1.30) | 1.07 (0.97 – 1.18) |
| 772.3 | Muscle weakness | 1.2 / 1.0 | 1.17 (1.05 – 1.30) | 5.9E-03 | 2.01 (1.74 – 2.32) | 6.2E-02 | 1.06 (0.91 – 1.23) | 1.31 (1.12 – 1.55) |

| PheCode | Disease | Incidence OSA case / control | Risk ratio | P-value | Sensitivity analysis risk ratio | OSA x sex interaction p-value | Risk ratio (men) | Risk ratio (women) |
|---|---|---|---|---|---|---|---|---|
| 428.4 | Heart failure with preserved EF [Diastolic heart failure] | 1.8 / 1.6 | 1.13 (1.03 – 1.23) | 6.9E-03 | 1.67 (1.51 – 1.84) | 2.5E-01 | 1.09 (0.97 – 1.22) | 1.19 (1.04 – 1.36) |
| 564 | Functional digestive disorders | 1.1 / 0.9 | 1.17 (1.04 – 1.32) | 7.7E-03 | 1.50 (1.32 – 1.71) | NA | NA | 1.24 (1.05 – 1.47) |
| 716.9 | Arthropathy NOS | 2.2 / 1.9 | 1.12 (1.03 – 1.22) | 8.8E-03 | 1.37 (1.26 – 1.50) | 5.4E-01 | 1.09 (0.98 – 1.23) | 1.14 (1.01 – 1.29) |
| 569 | Other disorders of intestine | 3.3 / 2.9 | 1.09 (1.02 – 1.17) | 1.0E-02 | 1.16 (1.08 – 1.25) | 9.5E-02 | 1.04 (0.95 – 1.14) | 1.16 (1.05 – 1.29) |
| 477 | Epistaxis or throat hemorrhage | 1.3 / 1.1 | 1.14 (1.03 – 1.27) | 1.2E-02 | 1.37 (1.22 – 1.54) | 1.3E-01 | 1.22 (1.07 – 1.39) | 1.03 (0.87 – 1.22) |
| 941 | Adverse reaction to serum or vaccine | 1.1 / 0.9 | 1.16 (1.03 – 1.30) | 1.3E-02 | 1.36 (1.20 – 1.54) | NA | NA | 1.18 (1.02 – 1.37) |
| 720.1 | Spinal stenosis of lumbar region | 2.2 / 2.4 | 0.90 (0.84 – 0.98) | 1.3E-02 | 0.87 (0.80 – 0.94) | 8.2E-01 | 0.92 (0.83 – 1.02) | 0.89 (0.78 – 1.00) |
| 938.2 | Chronic dermatitis due to solar radiation | 1.8 / 1.6 | 1.12 (1.02 – 1.22) | 1.6E-02 | 1.25 (1.12 – 1.38) | 2.5E-01 | 1.15 (1.03 – 1.29) | 1.04 (0.89 – 1.23) |
| 550.5 | Ventral hernia | 1.0 / 0.9 | 1.15 (1.02 – 1.29) | 2.2E-02 | 1.38 (1.22 – 1.56) | NA | NA | 1.21 (1.01 – 1.45) |
| 593.1 | Gross hematuria | 1.8 / 1.6 | 1.11 (1.01 – 1.21) | 2.5E-02 | 1.23 (1.12 – 1.35) | NA | 1.08 (0.98 – 1.20) | NA |
| 416 | Cardiomegaly | 1.1 / 1.0 | 1.14 (1.01 – 1.28) | 3.0E-02 | 1.19 (1.05 – 1.34) | NA | 1.07 (0.93 – 1.23) | NA |
| 525 | Other diseases of the teeth and supporting structures | 1.0 / 0.9 | 1.14 (1.01 – 1.29) | 3.0E-02 | 1.26 (1.10 – 1.43) | NA | NA | 1.30 (1.09 – 1.55) |
| 532 | Dysphagia | 4.0 / 3.6 | 1.06 (1.00 – 1.13) | 4.6E-02 | 1.25 (1.17 – 1.34) | 1.1E-02 | 0.99 (0.91 – 1.07) | 1.18 (1.07 – 1.30) |
| 721.8 | Other allied disorders of spine | 1.2 / 0.3 | 4.53 (3.87 – 5.31) | 4.3E-79 | 1.05 (0.93 – 1.20) | 3.2E-01 | 4.28 (3.50 – 5.25) | 4.94 (3.86 – 6.34) |
| 318 | Tobacco use disorder | 2.4 / 4.3 | 0.58 (0.53 – 0.62) | 1.6E-45 | 0.95 (0.88 – 1.03) | 3.9E-02 | 0.54 (0.49 – 0.60) | 0.64 (0.57 – 0.73) |
| 947 | Urticaria | 1.7 / 0.8 | 2.09 (1.88 – 2.33) | 1.8E-42 | 1.02 (0.92 – 1.14) | 6.5E-01 | 2.02 (1.70 – 2.40) | 2.14 (1.87 – 2.45) |
| 530.2 | Esophageal bleeding (varices/hemorrhage) | 1.0 / 0.5 | 1.87 (1.64 – 2.13) | 1.6E-20 | 0.89 (0.76 – 1.06) | 1.6E-04 | 1.56 (1.32 – 1.84) | 2.59 (2.07 – 3.24) |
| 773 | Pain in limb | 8.7 / 7.1 | 1.25 (1.19 – 1.31) | 2.4E-20 | 0.95 (0.90 – 1.01) | 5.5E-01 | 1.28 (1.21 – 1.37) | 1.21 (1.12 – 1.31) |
| 733 | Other disorders of bone and cartilage | 5.6 / 4.3 | 1.26 (1.20 – 1.33) | 2.1E-19 | 1.02 (0.96 – 1.09) | 4.6E-03 | 1.46 (1.31 – 1.63) | 1.21 (1.14 – 1.28) |
| 694.2 | Other dyschromia | 3.6 / 2.7 | 1.32 (1.23 – 1.41) | 1.6E-15 | 1.02 (0.94 – 1.10) | 3.5E-05 | 1.47 (1.35 – 1.61) | 1.10 (0.99 – 1.23) |
| 244.4 | Hypothyroidism NOS | 3.3 / 2.7 | 1.29 (1.21 – 1.38) | 3.6E-13 | 1.02 (0.95 – 1.10) | 3.0E-03 | 1.44 (1.30 – 1.60) | 1.19 (1.08 – 1.30) |
| 426.21 | First degree AV block | 1.0 / 1.4 | 0.69 (0.62 – 0.77) | 5.0E-11 | 1.11 (0.97 – 1.28) | NA | 0.68 (0.60 – 0.78) | NA |
| 427.7 | Tachycardia NOS | 2.8 / 3.5 | 0.80 (0.74 – 0.85) | 1.4E-10 | 0.98 (0.91 – 1.06) | 1.4E-01 | 0.76 (0.69 – 0.83) | 0.85 (0.77 – 0.94) |
| 695.9 | Unspecified erythematous | 1.2 / 1.6 | 0.71 (0.64 – 0.79) | 1.6E-10 | 1.00 (0.90 – 1.11) | 3.0E-01 | 0.75 (0.66 – 0.86) | 0.67 (0.57 – 0.78) |

| PheCode | Disease | Incidence OSA case / control | Risk ratio | P-value | Sensitivity analysis risk ratio | OSA x sex interaction p-value | Risk ratio (men) | Risk ratio (women) |
|---|---|---|---|---|---|---|---|---|
| | condition | | | | | | | |
| 433.21 | Cerebral artery occlusion, with cerebral infarction | 2.2 / 2.7 | 0.79 (0.74 – 0.86) | 2.6E-09 | 1.01 (0.93 – 1.09) | 7.7E-01 | 0.79 (0.71 – 0.86) | 0.81 (0.71 – 0.91) |
| 285.1 | Acute posthemorrhagic anemia | 1.3 / 1.7 | 0.75 (0.68 – 0.82) | 6.4E-09 | 0.95 (0.86 – 1.06) | 1.2E-01 | 0.70 (0.62 – 0.80) | 0.83 (0.70 – 0.97) |
| 078 | Viral warts & HPV | 1.5 / 1.1 | 1.35 (1.22 – 1.50) | 9.8E-09 | 0.99 (0.88 – 1.12) | 1.5E-01 | 1.44 (1.26 – 1.65) | 1.23 (1.05 – 1.44) |
| 276.41 | Acidosis | 1.1 / 1.5 | 0.76 (0.69 – 0.85) | 5.0E-07 | 0.92 (0.82 – 1.02) | 4.8E-05 | 0.64 (0.56 – 0.74) | 1.01 (0.85 – 1.20) |
| 851 | Complications of transplants and reattached limbs | 1.0 / 1.4 | 0.77 (0.69 – 0.86) | 3.5E-06 | 1.11 (1.00 – 1.24) | NA | 0.76 (0.66 – 0.88) | NA |
| 596 | Other disorders of bladder | 1.2 / 0.9 | 1.31 (1.17 – 1.47) | 4.7E-06 | 1.12 (0.99 – 1.27) | NA | 1.30 (1.14 – 1.48) | NA |
| 506 | Empyema and pneumothorax | 1.3 / 1.6 | 0.80 (0.72 – 0.88) | 7.0E-06 | 0.92 (0.83 – 1.03) | 2.8E-01 | 0.76 (0.68 – 0.86) | 0.87 (0.73 – 1.04) |
| 079.9 | Viremia, NOS | 1.3 / 1.6 | 0.80 (0.72 – 0.88) | 9.7E-06 | 1.07 (0.96 – 1.18) | NA | NA | 0.83 (0.72 – 0.95) |
| 585.1 | Acute renal failure | 3.1 / 3.5 | 0.86 (0.81 – 0.92) | 1.0E-05 | 0.94 (0.87 – 1.00) | 3.4E-02 | 0.82 (0.76 – 0.89) | 0.95 (0.85 – 1.07) |
| 721.1 | Spondylosis without myelopathy | 4.3 / 3.7 | 1.14 (1.08 – 1.22) | 1.5E-05 | 0.97 (0.90 – 1.04) | 4.9E-01 | 1.18 (1.08 – 1.28) | 1.11 (1.01 – 1.22) |
| 702.1 | Actinic keratosis | 2.1 / 1.7 | 1.20 (1.11 – 1.31) | 2.1E-05 | 1.03 (0.94 – 1.14) | 2.4E-01 | 1.24 (1.12 – 1.37) | 1.11 (0.94 – 1.31) |
| 276.13 | Hyperpotassemia | 1.4 / 1.6 | 0.83 (0.75 – 0.91) | 1.4E-04 | 0.95 (0.85 – 1.05) | 6.0E-01 | 0.84 (0.75 – 0.95) | 0.79 (0.66 – 0.94) |
| 411.4 | Coronary atherosclerosis | 3.9 / 4.5 | 0.89 (0.84 – 0.95) | 2.7E-04 | 0.95 (0.89 – 1.01) | 5.0E-02 | 0.87 (0.81 – 0.93) | 0.98 (0.86 – 1.11) |
| 428.3 | Heart failure with reduced EF [Systolic or combined heart failure] | 1.1 / 1.3 | 0.82 (0.73 – 0.91) | 2.9E-04 | 1.01 (0.89 – 1.14) | NA | 0.77 (0.68 – 0.87) | NA |
| 452.2 | Deep vein thrombosis [DVT] | 2.0 / 2.3 | 0.86 (0.80 – 0.94) | 3.4E-04 | 1.01 (0.94 – 1.10) | 2.1E-02 | 0.80 (0.72 – 0.89) | 0.97 (0.85 – 1.11) |
| 509.1 | Respiratory failure | 1.8 / 2.1 | 0.86 (0.79 – 0.94) | 4.4E-04 | 1.02 (0.93 – 1.12) | 5.8E-04 | 0.77 (0.69 – 0.85) | 1.04 (0.90 – 1.19) |
| 695.8 | Other specified erythematous conditions | 1.0 / 0.9 | 1.23 (1.09 – 1.38) | 6.4E-04 | 1.09 (0.96 – 1.24) | NA | NA | 1.14 (0.98 – 1.33) |
| 418 | Nonspecific chest pain | 8.1 / 7.7 | 1.09 (1.04 – 1.14) | 6.4E-04 | 0.99 (0.94 – 1.05) | 1.4E-01 | 1.07 (1.01 – 1.14) | 1.13 (1.04 – 1.23) |
| 514.2 | Solitary pulmonary nodule | 3.6 / 3.1 | 1.11 (1.04 – 1.19) | 8.9E-04 | 1.06 (0.99 – 1.14) | 4.0E-01 | 1.14 (1.05 – 1.24) | 1.08 (0.98 – 1.19) |
| 785 | Abdominal pain | 7.5 / 7.4 | 1.08 (1.03 – 1.14) | 1.8E-03 | 0.95 (0.90 – 1.00) | 2.4E-02 | 1.03 (0.96 – 1.10) | 1.16 (1.07 – 1.25) |
| 763 | Thoracic or lumbosacral neuritis or radiculitis, unspecified | 3.3 / 3.0 | 1.11 (1.04 – 1.19) | 1.9E-03 | 1.01 (0.94 – 1.09) | 8.8E-01 | 1.13 (1.03 – 1.23) | 1.09 (0.99 – 1.21) |

| PheCode | Disease | Incidence OSA case / control | Risk ratio | P-value | Sensitivity analysis risk ratio | OSA x sex interaction p-value | Risk ratio (men) | Risk ratio (women) |
|---|---|---|---|---|---|---|---|---|
| 252.1 | Hyperparathyroidism | 1.0 / 0.9 | 1.20 (1.07 – 1.35) | 2.5E-03 | 1.05 (0.92 – 1.19) | NA | NA | 1.16 (0.99 – 1.35) |
| 172.2 | Other non-epithelial cancer of skin | 2.3 / 2.0 | 1.12 (1.03 – 1.21) | 5.8E-03 | 1.03 (0.94 – 1.12) | 5.0E-01 | 1.10 (0.99 – 1.21) | 1.17 (1.01 – 1.35) |
| 722.1 | Displacement of intervertebral disc | 1.6 / 1.4 | 1.14 (1.04 – 1.26) | 8.0E-03 | 1.11 (1.00 – 1.23) | 5.2E-01 | 1.19 (1.05 – 1.34) | 1.08 (0.92 – 1.27) |
| 250.2 | Type 2 diabetes | 5.2 / 5.5 | 1.07 (1.01 – 1.13) | 1.6E-02 | 1.00 (0.95 – 1.06) | 3.8E-04 | 1.03 (0.96 – 1.10) | 1.17 (1.07 – 1.28) |
| 558 | Noninfectious gastroenteritis | 2.0 / 1.8 | 1.11 (1.02 – 1.21) | 2.0E-02 | 1.06 (0.96 – 1.17) | 9.2E-01 | 1.09 (0.97 – 1.23) | 1.12 (0.99 – 1.27) |
| 830 | Dislocation | 1.2 / 1.1 | 1.14 (1.02 – 1.27) | 2.1E-02 | 1.07 (0.95 – 1.20) | 9.6E-02 | 1.26 (1.09 – 1.45) | 1.00 (0.84 – 1.19) |
| 401.3 | Other hypertensive complications | 1.2 / 1.3 | 0.88 (0.79 – 0.98) | 2.1E-02 | 1.09 (0.98 – 1.22) | 8.8E-01 | 0.88 (0.77 – 1.01) | 0.89 (0.75 – 1.06) |
| 241.1 | Nontoxic uninodular goiter | 1.9 / 1.8 | 1.10 (1.01 – 1.20) | 2.3E-02 | 1.06 (0.96 – 1.17) | 8.9E-02 | 1.00 (0.86 – 1.16) | 1.17 (1.05 – 1.30) |
| 433.31 | Transient cerebral ischemia | 2.4 / 2.1 | 1.09 (1.01 – 1.18) | 2.8E-02 | 1.02 (0.94 – 1.11) | 9.2E-01 | 1.09 (0.99 – 1.21) | 1.09 (0.96 – 1.23) |
| 560.4 | Other intestinal obstruction | 1.1 / 1.0 | 1.13 (1.01 – 1.26) | 3.5E-02 | 1.03 (0.91 – 1.17) | 6.1E-01 | 1.09 (0.93 – 1.27) | 1.17 (0.99 – 1.38) |
| 706.2 | Sebaceous cyst | 1.9 / 1.7 | 1.10 (1.00 – 1.20) | 3.9E-02 | 0.95 (0.86 – 1.05) | 2.3E-02 | 1.21 (1.08 – 1.35) | 0.96 (0.84 – 1.11) |
| 411.2 | Myocardial infarction | 2.3 / 2.4 | 0.92 (0.85 – 1.00) | 3.9E-02 | 0.96 (0.89 – 1.04) | 4.0E-01 | 0.91 (0.83 – 1.00) | 0.96 (0.82 – 1.11) |
| 454.1 | Varicose veins of lower extremity | 1.1 / 1.0 | 1.12 (1.00 – 1.26) | 5.1E-02 | 1.25 (1.10 – 1.41) | NA | NA | 1.06 (0.90 – 1.23) |
| 426.32 | Left bundle branch block | 1.0 / 0.9 | 1.12 (1.00 – 1.27) | 5.5E-02 | 1.12 (0.98 – 1.28) | NA | 1.14 (0.98 – 1.32) | NA |
| 199 | Neoplasm of uncertain behavior | 1.4 / 1.6 | 0.91 (0.82 – 1.00) | 5.9E-02 | 0.84 (0.77 – 0.92) | 2.6E-01 | 0.86 (0.76 – 0.98) | 0.98 (0.84 – 1.14) |
| 038 | Septicemia | 1.8 / 1.9 | 0.92 (0.85 – 1.01) | 6.7E-02 | 1.16 (1.06 – 1.27) | 7.0E-01 | 0.91 (0.82 – 1.00) | 0.96 (0.82 – 1.11) |
| 805 | Fracture of vertebral column without mention of spinal cord injury | 1.3 / 1.2 | 1.10 (0.99 – 1.22) | 7.5E-02 | 1.03 (0.92 – 1.16) | 2.3E-01 | 1.05 (0.91 – 1.21) | 1.16 (1.00 – 1.35) |
| 593 | Hematuria | 3.5 / 3.2 | 1.06 (0.99 – 1.13) | 7.6E-02 | 1.18 (1.10 – 1.27) | 2.5E-01 | 1.03 (0.95 – 1.11) | 1.14 (1.01 – 1.28) |
| 979 | Adverse drug events and drug allergies | 3.1 / 2.9 | 1.06 (0.99 – 1.13) | 1.1E-01 | 0.97 (0.90 – 1.05) | 6.2E-01 | 1.07 (0.97 – 1.17) | 1.04 (0.94 – 1.15) |
| 535.9 | Gastritis and duodenitis, NOS | 1.6 / 1.7 | 0.93 (0.85 – 1.02) | 1.2E-01 | 1.42 (1.29 – 1.55) | 4.5E-02 | 0.85 (0.75 – 0.97) | 1.02 (0.89 – 1.17) |
| 574.1 | Cholelithiasis | 2.1 / 2.0 | 1.06 (0.98 – 1.16) | 1.3E-01 | 1.08 (0.99 – 1.18) | 9.6E-01 | 1.05 (0.93 – 1.18) | 1.08 (0.96 – 1.21) |
| 591 | Urinary tract infection | 3.4 / 3.2 | 1.05 (0.98 – 1.13) | 1.4E-01 | 1.01 (0.94 – 1.09) | 5.8E-01 | 1.01 (0.91 – 1.11) | 1.09 (1.00 – 1.20) |
| 512.7 | Shortness of breath | 7.9 / 8.2 | 0.97 (0.92 – 1.01) | 1.4E-01 | 1.37 (1.31 – 1.44) | 1.1E-03 | 0.91 (0.86 – 0.96) | 1.06 (0.99 – 1.14) |

| PheCode | Disease | Incidence OSA case / control | Risk ratio | P-value | Sensitivity analysis risk ratio | OSA x sex interaction p-value | Risk ratio (men) | Risk ratio (women) |
|---|---|---|---|---|---|---|---|---|
| 427.61 | Supraventricular premature beats | 1.8 / 1.6 | 1.07 (0.98 – 1.16) | 1.6E-01 | 1.19 (1.07 – 1.33) | 8.2E-01 | 1.07 (0.96 – 1.20) | 1.05 (0.91 – 1.22) |
| 687.2 | Localized superficial swelling, mass, or lump | 3.5 / 3.3 | 1.05 (0.98 – 1.11) | 1.7E-01 | 1.38 (1.29 – 1.48) | 5.2E-01 | 1.03 (0.94 – 1.12) | 1.07 (0.97 – 1.16) |
| 415.11 | Pulmonary embolism and infarction, acute | 1.5 / 1.6 | 0.94 (0.85 – 1.03) | 1.7E-01 | 0.97 (0.88 – 1.07) | 2.0E-01 | 0.89 (0.78 – 1.00) | 1.01 (0.87 – 1.17) |
| 1011 | Complications of surgical and medical procedures | 3.2 / 3.0 | 1.05 (0.98 – 1.12) | 1.7E-01 | 0.86 (0.80 – 0.92) | 7.0E-01 | 1.05 (0.96 – 1.15) | 1.04 (0.94 – 1.15) |
| 578.9 | Hemorrhage of gastrointestinal tract | 1.9 / 1.7 | 1.06 (0.97 – 1.16) | 2.1E-01 | 1.26 (1.15 – 1.39) | 7.3E-01 | 1.06 (0.96 – 1.18) | 1.04 (0.89 – 1.22) |
| 599 | Other symptoms/disorders or the urinary system | 1.4 / 1.3 | 1.06 (0.96 – 1.18) | 2.2E-01 | 1.23 (1.11 – 1.37) | 7.6E-01 | 1.07 (0.94 – 1.21) | 1.05 (0.90 – 1.24) |
| 473.4 | Voice disturbance | 1.8 / 1.6 | 1.06 (0.97 – 1.16) | 2.2E-01 | 1.35 (1.22 – 1.48) | 9.5E-01 | 1.04 (0.92 – 1.18) | 1.07 (0.94 – 1.22) |
| 586 | Other disorders of the kidney and ureters | 3.0 / 2.8 | 1.04 (0.97 – 1.12) | 2.3E-01 | 1.34 (1.25 – 1.44) | 3.8E-01 | 1.06 (0.98 – 1.16) | 1.00 (0.88 – 1.13) |
| 916 | Contusion | 3.9 / 4.0 | 0.96 (0.91 – 1.03) | 2.4E-01 | 0.87 (0.81 – 0.94) | 3.1E-01 | 0.94 (0.86 – 1.02) | 1.00 (0.91 – 1.09) |
| 686 | Other local infections of skin and subcutaneous tissue | 1.0 / 1.1 | 0.93 (0.83 – 1.05) | 2.5E-01 | 1.82 (1.66 – 2.00) | NA | 0.94 (0.81 – 1.08) | NA |
| 428.1 | Congestive heart failure (CHF) NOS | 2.8 / 2.7 | 1.04 (0.97 – 1.12) | 2.7E-01 | 1.49 (1.38 – 1.60) | 3.0E-03 | 0.97 (0.89 – 1.05) | 1.20 (1.07 – 1.36) |
| 1013 | Asphyxia and hypoxemia | 1.8 / 1.9 | 0.95 (0.87 – 1.04) | 2.9E-01 | 1.34 (1.22 – 1.48) | 2.2E-01 | 0.91 (0.81 – 1.02) | 1.02 (0.89 – 1.17) |
| 835 | Internal derangement of knee | 1.1 / 1.1 | 1.06 (0.95 – 1.19) | 2.9E-01 | 1.09 (0.97 – 1.23) | 8.3E-01 | 1.08 (0.93 – 1.26) | 1.04 (0.87 – 1.25) |
| 396 | Abnormal heart sounds | 2.4 / 2.4 | 0.96 (0.89 – 1.04) | 3.0E-01 | 1.39 (1.28 – 1.51) | 4.8E-01 | 0.93 (0.84 – 1.03) | 1.00 (0.89 – 1.11) |
| 339 | Other headache syndromes | 3.2 / 3.4 | 0.96 (0.90 – 1.03) | 3.0E-01 | 1.22 (1.13 – 1.31) | 8.2E-04 | 0.85 (0.76 – 0.94) | 1.08 (0.98 – 1.19) |
| 458.9 | Hypotension NOS | 2.8 / 2.8 | 0.96 (0.90 – 1.03) | 3.1E-01 | 1.27 (1.17 – 1.37) | 1.2E-01 | 0.91 (0.84 – 1.00) | 1.06 (0.94 – 1.19) |
| 367.2 | Astigmatism | 1.2 / 1.1 | 1.06 (0.95 – 1.18) | 3.2E-01 | 1.50 (1.33 – 1.70) | 6.3E-01 | 1.03 (0.89 – 1.19) | 1.09 (0.93 – 1.29) |
| 041 | Bacterial infection NOS | 1.1 / 1.2 | 0.95 (0.85 – 1.06) | 3.2E-01 | 1.19 (1.06 – 1.33) | 4.2E-01 | 0.90 (0.78 – 1.04) | 1.01 (0.85 – 1.21) |
| 592.11 | Acute cystitis | 1.7 / 1.6 | 1.05 (0.96 – 1.14) | 3.2E-01 | 1.34 (1.21 – 1.48) | 4.3E-01 | 0.98 (0.85 – 1.14) | 1.08 (0.97 – 1.21) |
| 771.1 | Swelling of limb | 2.2 / 2.3 | 0.96 (0.88 – 1.04) | 3.2E-01 | 1.67 (1.54 – 1.81) | 9.1E-01 | 0.96 (0.87 – 1.07) | 0.95 (0.84 – 1.08) |
| 395.2 | Nonrheumatic aortic valve disorders | 1.7 / 1.8 | 0.96 (0.87 – 1.05) | 3.3E-01 | 0.98 (0.89 – 1.08) | 2.1E-01 | 0.92 (0.83 – 1.03) | 1.03 (0.88 – 1.22) |
| 228 | Hemangioma and lymphangioma, any site | 1.1 / 1.2 | 0.95 (0.85 – 1.06) | 3.3E-01 | 0.95 (0.83 – 1.07) | 3.6E-02 | 1.05 (0.90 – 1.22) | 0.84 (0.72 – 0.99) |

| PheCode | Disease | Incidence OSA case / control | Risk ratio | P-value | Sensitivity analysis risk ratio | OSA x sex interaction p-value | Risk ratio (men) | Risk ratio (women) |
|---|---|---|---|---|---|---|---|---|
| 276.12 | Hyposmolality and/or hyponatremia | 1.8 / 1.8 | 0.96 (0.88 – 1.05) | 3.3E-01 | 0.91 (0.82 – 1.00) | 9.7E-01 | 0.96 (0.85 – 1.07) | 0.96 (0.84 – 1.10) |
| 698 | Pruritus and related conditions | 2.4 / 2.2 | 1.04 (0.96 – 1.12) | 3.3E-01 | 1.53 (1.40 – 1.67) | 1.4E-02 | 0.91 (0.81 – 1.03) | 1.14 (1.03 – 1.27) |
| 454.11 | Varicose veins of lower extremity, symptomtic | 1.0 / 1.0 | 1.05 (0.94 – 1.19) | 3.7E-01 | 1.07 (0.95 – 1.21) | NA | NA | 1.05 (0.89 – 1.23) |
| 427.21 | Atrial fibrillation | 3.9 / 3.9 | 1.03 (0.97 – 1.09) | 3.7E-01 | 1.12 (1.06 – 1.20) | 7.1E-01 | 1.03 (0.96 – 1.10) | 1.03 (0.92 – 1.15) |
| 038.3 | Bacteremia | 1.6 / 1.7 | 0.96 (0.88 – 1.05) | 3.8E-01 | 1.12 (1.02 – 1.23) | 2.4E-01 | 0.92 (0.83 – 1.03) | 1.05 (0.89 – 1.24) |
| 801 | Fracture of ankle and foot | 1.1 / 1.1 | 1.05 (0.94 – 1.17) | 4.1E-01 | 1.04 (0.92 – 1.18) | NA | NA | 0.97 (0.83 – 1.13) |
| 789 | Nausea and vomiting | 5.0 / 5.3 | 0.98 (0.93 – 1.03) | 4.4E-01 | 1.09 (1.03 – 1.15) | 6.4E-09 | 0.82 (0.75 – 0.89) | 1.14 (1.06 – 1.22) |
| 598.9 | Other nonspecific findings on examination of urine | 1.7 / 1.6 | 1.04 (0.95 – 1.13) | 4.4E-01 | 1.27 (1.15 – 1.40) | 2.0E-01 | 0.96 (0.85 – 1.10) | 1.10 (0.97 – 1.25) |
| 112 | Candidiasis | 1.9 / 1.8 | 1.03 (0.95 – 1.13) | 4.6E-01 | 1.11 (1.01 – 1.23) | 4.0E-02 | 0.91 (0.80 – 1.04) | 1.13 (1.01 – 1.26) |
| 573 | Other disorders of liver | 2.0 / 2.1 | 0.97 (0.89 – 1.05) | 4.6E-01 | 0.95 (0.87 – 1.04) | 9.0E-02 | 0.91 (0.82 – 1.02) | 1.06 (0.93 – 1.20) |
| 743.9 | Osteopenia or other disorder of bone and cartilage | 1.2 / 1.2 | 1.04 (0.93 – 1.16) | 4.8E-01 | 0.95 (0.84 – 1.08) | NA | NA | 0.98 (0.84 – 1.14) |
| 288.2 | Elevated white blood cell count | 2.5 / 2.5 | 0.97 (0.91 – 1.05) | 5.0E-01 | 1.06 (0.98 – 1.15) | 1.6E-02 | 0.90 (0.81 – 0.99) | 1.10 (0.98 – 1.23) |
| 788 | Syncope and collapse | 3.7 / 3.5 | 1.02 (0.96 – 1.09) | 5.1E-01 | 1.18 (1.10 – 1.27) | 5.9E-01 | 1.03 (0.95 – 1.11) | 1.00 (0.90 – 1.12) |
| 260 | Protein-calorie malnutrition | 1.3 / 1.2 | 1.04 (0.93 – 1.15) | 5.2E-01 | 1.20 (1.06 – 1.36) | 2.9E-02 | 0.93 (0.81 – 1.06) | 1.21 (1.02 – 1.42) |
| 287.3 | Thrombocytopenia | 1.8 / 1.7 | 1.03 (0.94 – 1.12) | 5.2E-01 | 1.03 (0.94 – 1.14) | 3.9E-01 | 1.05 (0.95 – 1.16) | 0.97 (0.82 – 1.15) |
| 705.8 | Hyperhidrosis | 1.8 / 1.8 | 1.03 (0.94 – 1.12) | 5.7E-01 | 1.20 (1.09 – 1.33) | 1.7E-01 | 1.09 (0.97 – 1.23) | 0.96 (0.84 – 1.09) |
| 411.3 | Angina pectoris | 2.9 / 2.8 | 1.02 (0.95 – 1.09) | 5.9E-01 | 1.18 (1.09 – 1.27) | 7.1E-02 | 1.06 (0.98 – 1.16) | 0.92 (0.81 – 1.05) |
| 782.3 | Edema | 5.0 / 4.9 | 1.01 (0.96 – 1.07) | 6.1E-01 | 1.47 (1.39 – 1.55) | 9.8E-01 | 1.02 (0.95 – 1.09) | 1.01 (0.93 – 1.10) |
| 276.14 | Hypopotassemia | 1.7 / 1.7 | 0.98 (0.89 – 1.07) | 6.3E-01 | 1.22 (1.10 – 1.35) | 3.9E-01 | 1.02 (0.90 – 1.16) | 0.94 (0.83 – 1.07) |
| 456 | Chronic venous insufficiency [CVI] | 1.4 / 1.3 | 1.02 (0.93 – 1.13) | 6.4E-01 | 0.99 (0.89 – 1.09) | 2.0E-02 | 1.16 (1.01 – 1.33) | 0.89 (0.76 – 1.03) |
| 595 | Hydronephrosis | 1.2 / 1.2 | 0.98 (0.88 – 1.09) | 7.2E-01 | 0.88 (0.78 – 0.99) | 8.0E-01 | 0.99 (0.86 – 1.13) | 0.97 (0.81 – 1.16) |
| 276.6 | Fluid overload | 1.3 / 1.3 | 1.02 (0.92 – 1.13) | 7.3E-01 | 1.52 (1.37 – 1.68) | 1.7E-01 | 0.97 (0.85 – 1.10) | 1.13 (0.95 – 1.34) |
| 317.1 | Alcoholism | 1.2 / 1.1 | 1.02 (0.91 – 1.14) | 7.4E-01 | 0.92 (0.83 – 1.03) | NA | 0.86 (0.76 – 0.98) | NA |
| 496 | Chronic airway obstruction | 1.8 / 1.9 | 0.99 (0.90 – 1.08) | 7.7E-01 | 1.19 (1.08 – 1.31) | 7.9E-04 | 0.87 (0.77 – 0.98) | 1.18 (1.03 – 1.35) |
| 443.9 | Peripheral vascular | 1.4 / 1.3 | 1.01 (0.92 – 1.12) | 7.9E-01 | 1.13 (1.01 – 1.26) | 1.3E-02 | 0.94 (0.83 – 1.06) | 1.23 (1.02 – 1.49) |

| PheCode | Disease | Incidence OSA case / control | Risk ratio | P-value | Sensitivity analysis risk ratio | OSA x sex interaction p-value | Risk ratio (men) | Risk ratio (women) |
|---|---|---|---|---|---|---|---|---|
| | disease, unspecified | | | | | | | |
| 368 | Visual disturbances | 3.0 / 3.0 | 0.99 (0.93 – 1.06) | 8.0E-01 | 1.07 (0.99 – 1.15) | 7.5E-01 | 1.00 (0.91 – 1.10) | 0.98 (0.89 – 1.08) |
| 741.4 | Joint effusions | 3.9 / 4.0 | 0.99 (0.94 – 1.05) | 8.2E-01 | 0.89 (0.84 – 0.96) | 6.0E-02 | 1.05 (0.97 – 1.14) | 0.92 (0.84 – 1.01) |
| 427.5 | Arrhythmia (cardiac) NOS | 4.7 / 4.6 | 0.99 (0.94 – 1.05) | 8.3E-01 | 1.10 (1.03 – 1.17) | 1.2E-01 | 0.96 (0.90 – 1.03) | 1.07 (0.96 – 1.19) |
| 348.8 | Encephalopathy, not elsewhere classified | 1.0 / 1.0 | 0.99 (0.88 – 1.11) | 8.3E-01 | 1.15 (1.02 – 1.29) | NA | 0.97 (0.83 – 1.12) | NA |
| 726.3 | Bursitis | 1.1 / 1.1 | 1.01 (0.90 – 1.13) | 8.3E-01 | 1.27 (1.14 – 1.43) | NA | 1.07 (0.93 – 1.23) | NA |
| 452 | Other venous embolism and thrombosis | 1.9 / 1.9 | 0.99 (0.91 – 1.08) | 8.7E-01 | 1.01 (0.93 – 1.11) | 1.1E-03 | 0.89 (0.80 – 0.99) | 1.19 (1.03 – 1.36) |
| 738.4 | Acquired spondylolisthesis | 1.3 / 1.3 | 1.00 (0.90 – 1.11) | 9.7E-01 | 0.83 (0.74 – 0.93) | NA | NA | 1.03 (0.90 – 1.19) |
| 442.1 | Aortic aneurysm | 1.1 / 1.1 | 1.00 (0.89 – 1.12) | 9.8E-01 | 0.98 (0.88 – 1.10) | NA | 0.96 (0.85 – 1.08) | NA |
| 338.1 | Acute pain | 6.1 / 11.0 | 0.55 (0.52 – 0.57) | 1.3E-156 | 1.13 (1.08 – 1.17) | 1.1E-07 | 0.49 (0.46 – 0.52) | 0.62 (0.58 – 0.66) |
| 949 | Allergies, other | 4.2 / 5.9 | 0.71 (0.67 – 0.75) | 5.9E-36 | 1.68 (1.58 – 1.80) | 7.8E-01 | 0.71 (0.65 – 0.77) | 0.70 (0.66 – 0.76) |
| 1002 | Symptoms concerning nutrition, metabolism, and development | 1.5 / 2.4 | 0.60 (0.54 – 0.65) | 4.8E-28 | 3.28 (2.94 – 3.64) | 2.5E-04 | 0.51 (0.45 – 0.57) | 0.72 (0.63 – 0.83) |
| 695.7 | Prurigo and Lichen | 1.3 / 0.7 | 1.94 (1.72 – 2.18) | 6.1E-28 | 0.87 (0.77 – 0.99) | 1.3E-01 | 2.16 (1.81 – 2.57) | 1.78 (1.51 – 2.09) |
| 479 | Other upper respiratory disease | 4.6 / 6.0 | 0.77 (0.73 – 0.81) | 1.1E-21 | 1.60 (1.52 – 1.69) | 1.2E-05 | 0.86 (0.80 – 0.92) | 0.67 (0.62 – 0.73) |
| 512.1 | Wheezing | 1.7 / 2.4 | 0.71 (0.65 – 0.77) | 2.8E-15 | 1.26 (1.17 – 1.37) | 3.6E-01 | 0.68 (0.60 – 0.77) | 0.74 (0.66 – 0.83) |
| 561.2 | Flatulence | 3.0 / 3.7 | 0.80 (0.75 – 0.85) | 1.2E-11 | 1.44 (1.33 – 1.56) | 6.3E-01 | 0.81 (0.74 – 0.89) | 0.79 (0.72 – 0.87) |
| 915 | Superficial injury without mention of infection | 1.7 / 2.3 | 0.75 (0.69 – 0.81) | 2.5E-11 | 1.63 (1.47 – 1.81) | 2.2E-01 | 0.72 (0.65 – 0.80) | 0.80 (0.69 – 0.92) |
| 939 | Atopic/contact dermatitis due to other or unspecified | 3.6 / 4.4 | 0.82 (0.77 – 0.87) | 8.1E-10 | 1.18 (1.09 – 1.27) | 9.4E-03 | 0.88 (0.81 – 0.96) | 0.75 (0.68 – 0.82) |
| 578.2 | Blood in stool | 2.4 / 2.9 | 0.79 (0.74 – 0.85) | 8.2E-10 | 1.51 (1.39 – 1.65) | 9.9E-01 | 0.79 (0.72 – 0.87) | 0.79 (0.70 – 0.90) |
| 745 | Pain in joint | 14.8 / 13.9 | 1.12 (1.08 – 1.17) | 1.7E-09 | 0.89 (0.85 – 0.92) | 7.1E-01 | 1.14 (1.08 – 1.19) | 1.11 (1.04 – 1.18) |
| 562.1 | Diverticulosis | 6.2 / 5.3 | 1.16 (1.10 – 1.22) | 6.0E-09 | 0.89 (0.84 – 0.94) | 8.3E-06 | 1.28 (1.20 – 1.36) | 0.98 (0.90 – 1.07) |
| 503 | Pulmonary congestion and hypostasis | 2.1 / 2.5 | 0.80 (0.73 – 0.86) | 1.8E-08 | 1.13 (1.05 – 1.23) | 4.4E-03 | 0.73 (0.66 – 0.81) | 0.93 (0.81 – 1.06) |
| 366 | Cataract | 2.5 / 2.9 | 0.82 (0.76 – 0.88) | 5.1E-08 | 1.84 (1.70 – 1.99) | 4.6E-01 | 0.80 (0.72 – 0.88) | 0.84 (0.76 – 0.94) |
| 702.2 | Seborrheic keratosis | 3.2 / 2.6 | 1.21 (1.12 – 1.29) | 2.9E-07 | 0.87 (0.80 – 0.94) | 9.3E-04 | 1.31 (1.20 – 1.43) | 1.03 (0.91 – 1.16) |
| 687.4 | Disturbance of skin sensation | 5.0 / 5.7 | 0.87 (0.83 – 0.92) | 6.4E-07 | 1.53 (1.44 – 1.63) | 8.4E-01 | 0.88 (0.82 – 0.95) | 0.87 (0.80 – 0.94) |

| PheCode | Disease | Incidence OSA case / control | Risk ratio | P-value | Sensitivity analysis risk ratio | OSA x sex interaction p-value | Risk ratio (men) | Risk ratio (women) |
|---|---|---|---|---|---|---|---|---|
| 497 | Bronchitis | 2.3 / 1.9 | 1.23 (1.13 – 1.34) | 8.1E-07 | 0.88 (0.80 – 0.97) | 4.4E-02 | 1.33 (1.19 – 1.49) | 1.13 (1.00 – 1.27) |
| 292.4 | Altered mental status | 3.3 / 3.7 | 0.86 (0.81 – 0.92) | 2.1E-06 | 1.14 (1.07 – 1.22) | 4.0E-01 | 0.84 (0.78 – 0.91) | 0.89 (0.80 – 0.98) |
| 741 | Symptoms and disorders of the joints | 1.4 / 1.7 | 0.80 (0.72 – 0.88) | 9.9E-06 | 1.56 (1.40 – 1.74) | 9.8E-01 | 0.80 (0.69 – 0.92) | 0.80 (0.70 – 0.92) |
| 465.2 | Acute pharyngitis | 3.5 / 4.1 | 0.87 (0.82 – 0.93) | 5.2E-05 | 1.23 (1.15 – 1.32) | 9.0E-01 | 0.87 (0.80 – 0.95) | 0.88 (0.80 – 0.96) |
| 756.5 | Congenital osteodystrophies | 1.4 / 1.7 | 0.82 (0.75 – 0.91) | 7.1E-05 | 1.28 (1.17 – 1.41) | 3.4E-01 | 0.79 (0.70 – 0.90) | 0.87 (0.75 – 1.00) |
| 783 | Fever of unknown origin | 4.1 / 4.5 | 0.89 (0.84 – 0.95) | 1.9E-04 | 1.08 (1.01 – 1.15) | 8.4E-03 | 0.83 (0.77 – 0.90) | 1.00 (0.91 – 1.11) |
| 279 | Disorders involving the immune mechanism | 2.1 / 2.4 | 0.87 (0.80 – 0.94) | 6.9E-04 | 1.40 (1.29 – 1.52) | 8.7E-01 | 0.86 (0.78 – 0.95) | 0.88 (0.77 – 1.01) |
| 286.7 | Other and unspecified coagulation defects | 1.5 / 1.7 | 0.86 (0.78 – 0.94) | 1.3E-03 | 1.13 (1.03 – 1.24) | 5.4E-02 | 0.78 (0.69 – 0.89) | 0.97 (0.83 – 1.13) |
| 216 | Benign neoplasm of skin | 3.5 / 3.2 | 1.12 (1.04 – 1.20) | 1.4E-03 | 0.87 (0.80 – 0.94) | 3.1E-08 | 1.30 (1.19 – 1.41) | 0.85 (0.76 – 0.96) |
| 780 | Hypothermia/Chills | 1.1 / 1.3 | 0.85 (0.77 – 0.95) | 4.6E-03 | 1.26 (1.12 – 1.41) | NA | NA | 0.86 (0.73 – 1.00) |
| 681 | Superficial cellulitis and abscess | 2.1 / 2.4 | 0.89 (0.82 – 0.97) | 5.1E-03 | 1.23 (1.13 – 1.33) | 1.9E-01 | 0.86 (0.77 – 0.95) | 0.95 (0.83 – 1.09) |
| 539 | Bariatric surgery | 4.7 / 5.3 | 0.93 (0.88 – 0.98) | 5.3E-03 | 1.59 (1.51 – 1.68) | 3.4E-04 | 0.82 (0.75 – 0.88) | 1.05 (0.97 – 1.13) |
| 459.1 | Hemorrhage NOS | 1.3 / 1.4 | 0.88 (0.80 – 0.97) | 1.4E-02 | 1.89 (1.66 – 2.16) | 2.2E-01 | 0.83 (0.72 – 0.95) | 0.95 (0.82 – 1.11) |
| 722.6 | Degeneration of intervertebral disc | 2.3 / 2.1 | 1.10 (1.01 – 1.19) | 2.7E-02 | 0.87 (0.79 – 0.95) | 1.0E-01 | 1.17 (1.05 – 1.30) | 1.00 (0.88 – 1.14) |
| 241.2 | Nontoxic multinodular goiter | 1.1 / 1.0 | 1.14 (1.01 – 1.27) | 2.8E-02 | 0.84 (0.73 – 0.95) | NA | NA | 1.07 (0.93 – 1.22) |
| 079 | Viral infection | 1.8 / 1.6 | 1.10 (1.00 – 1.20) | 4.9E-02 | 0.87 (0.79 – 0.97) | 2.2E-02 | 1.20 (1.06 – 1.35) | 0.98 (0.85 – 1.12) |

### Table S5. Cross-sectional case/control comorbidity associations.

Primary analyses considered joint-sex PheCodes with >1% prevalence rate and were adjusted for age, sex, BMI, population, and healthcare encounters. Bonferroni-significant p-values are bolded. A sensitivity analysis using alternative control propensity score matching without healthcare encounters (but still adjusting for encounters in the analysis) was performed. PheCodes with discordant results (n=27) or p > 0.05 (n=72) in either analysis are displayed at the end of the table (from PheCode 530.2 onwards, highlighted in green). 421 PheCodes were both Bonferroni-level significant and had concordant directionality in the sensitivity analysis. Sleep apnea × sex interaction analyses were performed separately when prevalence was >1% in men and women. Equivalent sex-stratified analyses are shown.

| PheCode | Disease | Prevalence OSA Case / Control | Risk ratio | P-value | Sensitivity analysis risk ratio | OSA x sex interaction p-value | Risk ratio (men) | Risk ratio (women) |
|---|---|---|---|---|---|---|---|---|
| 244.4 | Hypothyroidism NOS | 12.5 / 6.6 | 1.92 (1.86 – 1.98) | **0.0E+00** | 1.84 (1.77 – 1.91) | 1.2E-03 | 2.08 (1.96 – 2.21) | 1.85 (1.78 – 1.92) |
| 272.1 | Hyperlipidemia | 34.3 / 18.2 | 1.90 (1.86 – 1.93) | **0.0E+00** | 1.68 (1.65 – 1.72) | 7.5E-01 | 1.89 (1.85 – 1.94) | 1.91 (1.85 – 1.97) |
| 272.11 | Hypercholesterolemia | 21.3 / 13.5 | 1.58 (1.55 – 1.62) | **0.0E+00** | 1.48 (1.44 – 1.52) | 7.4E-01 | 1.59 (1.54 – 1.64) | 1.58 (1.52 – 1.64) |
| 278.1 | Obesity | 25.6 / 15.1 | 1.77 (1.73 – 1.80) | **0.0E+00** | 1.97 (1.92 – 2.01) | **7.9E-14** | 1.97 (1.90 – 2.04) | 1.62 (1.58 – 1.67) |
| 296.22 | Major depressive disorder | 20.2 / 9.0 | 2.24 (2.18 – 2.30) | **0.0E+00** | 2.70 (2.61 – 2.78) | **2.9E-05** | 2.11 (2.02 – 2.19) | 2.36 (2.28 – 2.44) |
| 300.1 | Anxiety disorder | 20.8 / 11.7 | 1.78 (1.74 – 1.83) | **0.0E+00** | 2.10 (2.04 – 2.16) | 1.2E-01 | 1.82 (1.75 – 1.89) | 1.76 (1.70 – 1.81) |
| 327 | Sleep disorders | 6.1 / 0.8 | 7.19 (6.65 – 7.77) | **0.0E+00** | 2.77 (2.59 – 2.96) | 3.5E-01 | 7.44 (6.70 – 8.26) | 6.90 (6.14 – 7.75) |
| 327.4 | Insomnia | 12.3 / 4.4 | 2.71 (2.61 – 2.81) | **0.0E+00** | 3.06 (2.92 – 3.21) | 4.8E-01 | 2.75 (2.60 – 2.90) | 2.68 (2.54 – 2.82) |
| 327.71 | Restless legs syndrome | 4.3 / 0.7 | 6.27 (5.75 – 6.84) | **0.0E+00** | 6.94 (6.21 – 7.76) | 7.9E-02 | 6.75 (5.93 – 7.70) | 5.88 (5.24 – 6.60) |
| 401.1 | Essential hypertension | 43.2 / 30.6 | 1.43 (1.41 – 1.45) | **0.0E+00** | 1.62 (1.59 – 1.64) | 4.9E-01 | 1.43 (1.40 – 1.45) | 1.43 (1.40 – 1.46) |
| 455 | Hemorrhoids | 12.4 / 5.8 | 2.10 (2.03 – 2.18) | **0.0E+00** | 1.29 (1.24 – 1.34) | **5.6E-07** | 2.27 (2.17 – 2.37) | 1.88 (1.79 – 1.99) |
| 470 | Septal Deviations/Turbinate Hypertrophy | 9.9 / 2.0 | 4.90 (4.66 – 5.17) | **0.0E+00** | 4.88 (4.56 – 5.22) | **4.4E-13** | 5.66 (5.30 – 6.05) | 3.80 (3.49 – 4.14) |
| 472 | Chronic pharyngitis and nasopharyngitis | 7.7 / 1.1 | 6.87 (6.42 – 7.35) | **0.0E+00** | 3.12 (2.94 – 3.32) | 1.6E-03 | 7.56 (6.90 – 8.29) | 6.08 (5.50 – 6.72) |
| 474.2 | Chronic tonsillitis and adenoiditis | 4.5 / 0.7 | 5.56 (5.11 – 6.06) | **0.0E+00** | 6.89 (6.12 – 7.77) | **2.8E-12** | 7.44 (6.59 – 8.41) | 4.01 (3.55 – 4.52) |
| 476 | Allergic rhinitis | 17.6 / 8.4 | 2.07 (2.01 – 2.13) | **0.0E+00** | 1.74 (1.68 – 1.80) | **9.3E-30** | 2.43 (2.34 – 2.54) | 1.76 (1.69 – 1.83) |
| 495 | Asthma | 16.9 / 8.8 | 1.92 (1.87 – 1.98) | **0.0E+00** | 2.08 (2.02 – 2.15) | 3.4E-01 | 1.88 (1.80 – 1.97) | 1.95 (1.88 – 2.02) |

| PheCode | Disease | Prevalence OSA Case / Control | Risk ratio | P-value | Sensitivity analysis risk ratio | OSA x sex interaction p-value | Risk ratio (men) | Risk ratio (women) |
|---|---|---|---|---|---|---|---|---|
| 530.11 | GERD | 29.0 / 15.2 | 1.91 (1.87 – 1.95) | **0.0E+00** | 2.16 (2.11 – 2.21) | 4.2E-01 | 1.92 (1.86 – 1.97) | 1.90 (1.85 – 1.95) |
| 689 | Disorder of skin and subcutaneous tissue NOS | 11.3 / 5.4 | 2.06 (1.99 – 2.14) | **0.0E+00** | 1.26 (1.21 – 1.30) | 3.3E-04 | 2.18 (2.08 – 2.29) | 1.91 (1.80 – 2.02) |
| 512.9 | Other dyspnea | 15.8 / 9.2 | 1.72 (1.67 – 1.77) | **1.0E-293** | 2.94 (2.84 – 3.04) | 4.4E-01 | 1.70 (1.64 – 1.77) | 1.75 (1.67 – 1.82) |
| 427.21 | Atrial fibrillation | 12.0 / 6.7 | 1.83 (1.77 – 1.89) | **1.5E-291** | 1.93 (1.86 – 2.01) | 5.1E-01 | 1.84 (1.77 – 1.92) | 1.80 (1.69 – 1.92) |
| 278 | Overweight, obesity and other hyperalimentation | 12.5 / 6.6 | 1.87 (1.81 – 1.93) | **7.6E-288** | 1.73 (1.66 – 1.81) | **5.8E-11** | 2.07 (1.98 – 2.17) | 1.65 (1.57 – 1.74) |
| 475 | Chronic sinusitis | 8.2 / 3.6 | 2.26 (2.16 – 2.36) | **8.9E-284** | 1.98 (1.88 – 2.09) | **2.6E-22** | 2.76 (2.60 – 2.94) | 1.79 (1.68 – 1.91) |
| 250.2 | Type 2 diabetes | 16.5 / 10.8 | 1.56 (1.52 – 1.60) | **3.1E-237** | 1.56 (1.51 – 1.60) | **6.4E-07** | 1.49 (1.44 – 1.54) | 1.69 (1.62 – 1.77) |
| 208 | Benign neoplasm of colon | 14.8 / 9.1 | 1.62 (1.57 – 1.67) | **5.9E-233** | 1.49 (1.44 – 1.54) | 6.5E-02 | 1.66 (1.60 – 1.72) | 1.56 (1.49 – 1.64) |
| 300.11 | Generalized anxiety disorder | 8.9 / 4.5 | 1.96 (1.89 – 2.05) | **6.8E-230** | 2.46 (2.35 – 2.58) | 5.9E-01 | 1.99 (1.87 – 2.12) | 1.95 (1.84 – 2.06) |
| 296.2 | Depression | 12.4 / 7.1 | 1.73 (1.67 – 1.79) | **1.4E-221** | 2.43 (2.33 – 2.52) | 1.8E-02 | 1.64 (1.56 – 1.73) | 1.79 (1.72 – 1.87) |
| 340 | Migraine | 8.6 / 4.4 | 1.94 (1.86 – 2.02) | **5.2E-221** | 2.21 (2.10 – 2.32) | 1.0E-02 | 2.11 (1.94 – 2.29) | 1.88 (1.79 – 1.97) |
| 313.1 | Attention deficit hyperactivity disorder | 4.4 / 1.5 | 2.87 (2.69 – 3.06) | **2.6E-220** | 3.44 (3.17 – 3.73) | 4.1E-02 | 2.72 (2.50 – 2.96) | 3.14 (2.82 – 3.49) |
| 292.3 | Memory loss | 5.4 / 2.1 | 2.51 (2.37 – 2.66) | **4.0E-219** | 2.21 (2.05 – 2.38) | 3.6E-04 | 2.76 (2.55 – 2.98) | 2.24 (2.06 – 2.43) |
| 798 | Malaise and fatigue | 19.4 / 13.0 | 1.48 (1.44 – 1.51) | **1.0E-215** | 1.82 (1.77 – 1.87) | 8.3E-04 | 1.53 (1.48 – 1.58) | 1.41 (1.36 – 1.47) |
| 389 | Hearing loss | 9.6 / 5.1 | 1.84 (1.77 – 1.92) | **3.1E-215** | 2.01 (1.92 – 2.11) | 5.0E-01 | 1.86 (1.77 – 1.95) | 1.81 (1.70 – 1.93) |
| 740.9 | Osteoarthrosis NOS | 17.5 / 12.1 | 1.48 (1.44 – 1.52) | **3.8E-206** | 1.42 (1.38 – 1.46) | 1.2E-01 | 1.45 (1.40 – 1.51) | 1.50 (1.45 – 1.55) |
| 550.2 | Diaphragmatic hernia | 5.2 / 2.1 | 2.45 (2.32 – 2.60) | **1.9E-205** | 1.76 (1.64 – 1.89) | 1.2E-01 | 2.56 (2.34 – 2.79) | 2.37 (2.19 – 2.56) |
| 745 | Pain in joint | 29.0 / 22.0 | 1.32 (1.29 – 1.34) | **8.1E-190** | 1.14 (1.12 – 1.16) | 3.6E-01 | 1.31 (1.27 – 1.34) | 1.33 (1.29 – 1.36) |
| 379.9 | Pain, swelling or discharge of eye | 2.5 / 0.6 | 4.14 (3.75 – 4.57) | **2.4E-175** | 2.57 (2.26 – 2.92) | 4.8E-02 | 3.75 (3.26 – 4.32) | 4.56 (3.97 – 5.23) |
| 338.2 | Chronic pain | 11.0 / 6.7 | 1.65 (1.59 – 1.71) | **3.5E-175** | 1.47 (1.41 – 1.53) | 6.6E-01 | 1.67 (1.59 – 1.75) | 1.63 (1.55 – 1.72) |
| 721.8 | Other allied disorders of spine | 2.0 / 0.3 | 5.89 (5.20 – 6.68) | **7.3E-170** | 1.20 (1.09 – 1.33) | 9.9E-01 | 5.93 (5.02 – 7.01) | 5.86 (4.85 – 7.09) |
| 760 | Back pain | 20.9 / 15.0 | 1.38 (1.35 – 1.42) | **3.5E-168** | 1.45 (1.41 – 1.48) | 6.4E-01 | 1.38 (1.33 – 1.42) | 1.39 (1.35 – 1.44) |

| PheCode | Disease | Prevalence OSA Case / Control | Risk ratio | P-value | Sensitivity analysis risk ratio | OSA x sex interaction p-value | Risk ratio (men) | Risk ratio (women) |
|---|---|---|---|---|---|---|---|---|
| 289.9 | Abnormality of red blood cells | 1.8 / 0.2 | 8.49 (7.30 – 9.89) | **1.7E-167** | 1.45 (1.30 – 1.61) | 9.8E-04 | 6.85 (5.66 – 8.30) | 11.87 (9.19 – 15.32) |
| 947 | Urticaria | 4.2 / 1.7 | 2.43 (2.28 – 2.60) | **9.3E-161** | 1.13 (1.06 – 1.20) | 3.1E-01 | 2.53 (2.27 – 2.82) | 2.38 (2.20 – 2.58) |
| 564.1 | Irritable Bowel Syndrome | 4.6 / 2.1 | 2.24 (2.11 – 2.37) | **4.9E-155** | 2.37 (2.20 – 2.55) | 6.5E-04 | 2.57 (2.32 – 2.85) | 2.08 (1.94 – 2.24) |
| 292 | Neurological disorders | 2.3 / 0.6 | 3.90 (3.52 – 4.31) | **1.2E-152** | 2.39 (2.16 – 2.65) | 1.1E-01 | 4.15 (3.64 – 4.73) | 3.54 (3.01 – 4.16) |
| 496 | Chronic airway obstruction | 6.5 / 3.6 | 1.87 (1.79 – 1.96) | **1.3E-152** | 2.17 (2.05 – 2.29) | 5.0E-04 | 1.74 (1.64 – 1.86) | 2.06 (1.92 – 2.22) |
| 428.1 | Congestive heart failure (CHF) NOS | 8.4 / 5.2 | 1.68 (1.62 – 1.75) | **2.3E-152** | 2.11 (2.02 – 2.20) | 1.3E-03 | 1.61 (1.54 – 1.69) | 1.84 (1.72 – 1.96) |
| 271.3 | Intestinal disaccharidase deficiencies and disaccharide malabsorption | 7.8 / 4.4 | 1.77 (1.70 – 1.85) | **1.3E-151** | 1.82 (1.74 – 1.91) | 2.0E-02 | 1.87 (1.76 – 1.99) | 1.69 (1.59 – 1.79) |
| 300.4 | Dysthymic disorder | 4.3 / 1.8 | 2.35 (2.20 – 2.50) | **1.9E-149** | 4.28 (3.64 – 5.02) | 7.0E-01 | 2.39 (2.16 – 2.64) | 2.33 (2.15 – 2.53) |
| 278.11 | Morbid obesity | 10.1 / 6.2 | 1.79 (1.71 – 1.86) | **6.5E-149** | 1.92 (1.85 – 2.00) | **2.3E-16** | 2.40 (2.20 – 2.61) | 1.53 (1.46 – 1.61) |
| 512.7 | Shortness of breath | 16.4 / 11.7 | 1.41 (1.38 – 1.45) | **1.2E-146** | 1.94 (1.88 – 2.00) | 1.5E-02 | 1.37 (1.33 – 1.42) | 1.47 (1.41 – 1.53) |
| 798.1 | Chronic fatigue syndrome | 2.8 / 0.9 | 2.97 (2.73 – 3.23) | **1.5E-143** | 3.74 (3.35 – 4.17) | 6.4E-01 | 3.02 (2.66 – 3.43) | 2.93 (2.62 – 3.28) |
| 272.13 | Mixed hyperlipidemia | 8.9 / 5.5 | 1.65 (1.59 – 1.72) | **4.2E-142** | 1.38 (1.33 – 1.45) | 3.1E-01 | 1.67 (1.60 – 1.75) | 1.61 (1.50 – 1.72) |
| 427.9 | Palpitations | 10.4 / 6.6 | 1.56 (1.51 – 1.62) | **1.6E-134** | 1.66 (1.59 – 1.73) | 2.3E-02 | 1.63 (1.55 – 1.72) | 1.51 (1.44 – 1.58) |
| 418 | Nonspecific chest pain | 19.4 / 14.4 | 1.34 (1.31 – 1.37) | **1.1E-129** | 1.36 (1.32 – 1.40) | 9.4E-02 | 1.37 (1.33 – 1.41) | 1.31 (1.26 – 1.36) |
| 261.4 | Vitamin D deficiency | 9.4 / 5.9 | 1.59 (1.53 – 1.65) | **1.2E-129** | 1.84 (1.76 – 1.92) | 1.2E-03 | 1.72 (1.62 – 1.82) | 1.51 (1.44 – 1.58) |
| 563 | Constipation | 9.5 / 5.8 | 1.58 (1.53 – 1.64) | **4.5E-127** | 2.00 (1.91 – 2.09) | **1.3E-05** | 1.45 (1.37 – 1.53) | 1.71 (1.63 – 1.80) |
| 599.5 | Frequency of urination and polyuria | 9.5 / 6.0 | 1.56 (1.50 – 1.62) | **1.5E-123** | 1.73 (1.65 – 1.80) | **9.9E-07** | 1.65 (1.58 – 1.73) | 1.35 (1.26 – 1.45) |
| 427.3 | Other specified cardiac dysrhythmias | 6.7 / 3.9 | 1.73 (1.65 – 1.81) | **5.3E-123** | 1.74 (1.65 – 1.84) | 6.2E-01 | 1.71 (1.62 – 1.80) | 1.77 (1.63 – 1.91) |
| 374 | Other disorders of eyelids | 1.4 / 0.2 | 7.00 (5.95 – 8.23) | **5.2E-122** | 1.90 (1.70 – 2.13) | 2.3E-02 | 8.70 (6.74 – 11.23) | 5.93 (4.80 – 7.32) |
| 285 | Other anemias | 13.8 / 9.7 | 1.42 (1.38 – 1.46) | **1.3E-120** | 1.54 (1.49 – 1.60) | 3.7E-02 | 1.38 (1.33 – 1.44) | 1.46 (1.40 – 1.53) |
| 761 | Cervicalgia | 10.4 / 6.7 | 1.52 (1.47 – 1.58) | **6.1E-119** | 1.63 (1.56 – 1.70) | 5.1E-01 | 1.51 (1.43 – 1.59) | 1.54 (1.46 – 1.61) |
| 327.41 | Organic or persistent insomnia | 2.3 / 0.8 | 2.93 (2.67 – 3.21) | **2.9E-117** | 2.72 (2.44 – 3.05) | 9.1E-01 | 2.92 (2.56 – 3.34) | 2.93 (2.59 – 3.33) |
| 306 | Other mental disorder | 6.1 / 3.4 | 1.79 (1.70 – 1.88) | **2.0E-115** | 1.76 (1.66 – 1.86) | 8.6E-01 | 1.78 (1.67 – 1.91) | 1.80 (1.67 – 1.94) |
| 388 | Other disorders of ear | 1.3 / 0.2 | 7.12 (6.02 – 8.43) | **2.9E-115** | 1.75 (1.48 – 2.08) | 7.2E-02 | 6.22 (5.01 – 7.74) | 8.56 (6.55 – 11.19) |
| 259 | Other endocrine disorders | 1.3 / 0.2 | 6.11 (5.23 – 7.15) | **1.4E-113** | 2.75 (2.42 – 3.14) | 4.5E-01 | 5.72 (4.64 – 7.06) | 6.60 (5.22 – 8.36) |

| PheCode | Disease | Prevalence OSA Case / Control | Risk ratio | P-value | Sensitivity analysis risk ratio | OSA x sex interaction p-value | Risk ratio (men) | Risk ratio (women) |
|---|---|---|---|---|---|---|---|---|
| 389.2 | Conductive hearing loss | 1.5 / 0.3 | 4.77 (4.17 – 5.47) | **4.0E-113** | 2.04 (1.68 – 2.48) | 5.5E-01 | 4.63 (3.88 – 5.52) | 5.00 (4.04 – 6.18) |
| 415.2 | Chronic pulmonary heart disease | 3.0 / 1.3 | 2.34 (2.18 – 2.53) | **5.7E-111** | 2.92 (2.67 – 3.20) | **5.5E-05** | 2.04 (1.84 – 2.26) | 2.75 (2.46 – 3.06) |
| 773 | Pain in limb | 17.0 / 12.6 | 1.34 (1.31 – 1.38) | **1.2E-109** | 1.11 (1.08 – 1.14) | **2.2E-06** | 1.43 (1.38 – 1.48) | 1.26 (1.21 – 1.31) |
| 293 | Symptoms involving head and neck | 1.3 / 0.2 | 6.56 (5.56 – 7.75) | **3.6E-109** | 1.70 (1.53 – 1.88) | 1.2E-02 | 8.07 (6.35 – 10.27) | 5.28 (4.19 – 6.66) |
| 379 | Other disorders of eye | 1.8 / 0.0 | 55.83 (39.03 – 79.86) | **1.7E-107** | 1.38 (1.19 – 1.59) | 1.6E-01 | 74.43 (41.94 – 132.09) | 44.13 (27.89 – 69.81) |
| 427.5 | Arrhythmia (cardiac) NOS | 11.5 / 8.1 | 1.43 (1.39 – 1.48) | **5.8E-105** | 1.46 (1.41 – 1.52) | 5.4E-01 | 1.44 (1.39 – 1.50) | 1.41 (1.33 – 1.50) |
| 389.1 | Sensorineural hearing loss | 5.9 / 3.4 | 1.72 (1.64 – 1.80) | **4.2E-104** | 1.60 (1.51 – 1.69) | 3.2E-01 | 1.75 (1.64 – 1.86) | 1.67 (1.54 – 1.80) |
| 357 | Inflammatory and toxic neuropathy | 5.4 / 3.1 | 1.76 (1.67 – 1.85) | **1.9E-102** | 2.04 (1.92 – 2.16) | 1.3E-02 | 1.67 (1.56 – 1.78) | 1.89 (1.75 – 2.05) |
| 386.9 | Dizziness and giddiness (Light-headedness and vertigo) | 11.8 / 8.2 | 1.42 (1.37 – 1.46) | **1.6E-100** | 1.50 (1.44 – 1.56) | 2.0E-02 | 1.48 (1.41 – 1.55) | 1.36 (1.30 – 1.42) |
| 733 | Other disorders of bone and cartilage | 8.5 / 5.5 | 1.50 (1.45 – 1.56) | **3.1E-99** | 1.18 (1.12 – 1.23) | 1.6E-03 | 1.70 (1.56 – 1.85) | 1.46 (1.40 – 1.52) |
| 706.8 | Other specified diseases of sebaceous glands | 4.6 / 2.5 | 1.85 (1.75 – 1.96) | **4.9E-98** | 1.42 (1.34 – 1.51) | 1.7E-01 | 1.93 (1.78 – 2.08) | 1.77 (1.63 – 1.93) |
| 701.2 | Scar conditions and fibrosis of skin | 5.6 / 3.3 | 1.71 (1.63 – 1.80) | **8.1E-97** | 1.26 (1.19 – 1.33) | 9.8E-01 | 1.71 (1.60 – 1.83) | 1.71 (1.58 – 1.85) |
| 173 | Neoplasm of uncertain behavior of skin | 5.4 / 3.1 | 1.73 (1.64 – 1.82) | **1.4E-96** | 1.74 (1.64 – 1.85) | 4.3E-01 | 1.75 (1.64 – 1.87) | 1.69 (1.56 – 1.84) |
| 599.4 | Urinary incontinence | 5.9 / 3.5 | 1.67 (1.59 – 1.75) | **7.7E-96** | 1.80 (1.70 – 1.90) | 7.9E-01 | 1.68 (1.54 – 1.84) | 1.66 (1.57 – 1.76) |
| 512.8 | Cough | 17.7 / 13.4 | 1.30 (1.27 – 1.34) | **1.4E-94** | 1.58 (1.53 – 1.62) | 1.0E-01 | 1.33 (1.28 – 1.38) | 1.28 (1.23 – 1.32) |
| 278.4 | Abnormal weight gain | 5.0 / 3.0 | 1.72 (1.63 – 1.82) | **2.1E-90** | 2.24 (2.12 – 2.37) | 4.5E-01 | 1.81 (1.65 – 1.99) | 1.69 (1.59 – 1.80) |
| 402 | Elevated blood pressure reading without diagnosis of hypertension | 7.3 / 4.7 | 1.55 (1.48 – 1.62) | **2.1E-89** | 1.42 (1.35 – 1.49) | **7.7E-06** | 1.69 (1.60 – 1.79) | 1.38 (1.29 – 1.47) |
| 395.1 | Nonrheumatic mitral valve disorders | 5.8 / 3.5 | 1.64 (1.56 – 1.72) | **2.3E-88** | 1.38 (1.30 – 1.46) | 6.1E-01 | 1.65 (1.55 – 1.76) | 1.62 (1.50 – 1.74) |
| 770 | Myalgia and myositis unspecified | 6.3 / 3.9 | 1.59 (1.52 – 1.67) | **1.8E-85** | 1.91 (1.81 – 2.02) | 2.0E-02 | 1.46 (1.34 – 1.59) | 1.66 (1.57 – 1.75) |
| 272.1 | Hyperglyceridemia | 3.4 / 1.8 | 1.96 (1.83 – 2.10) | **1.9E-85** | 1.91 (1.77 – 2.06) | 7.1E-01 | 1.98 (1.83 – 2.15) | 1.92 (1.71 – 2.17) |

| PheCode | Disease | Prevalence OSA Case / Control | Risk ratio | P-value | Sensitivity analysis risk ratio | OSA x sex interaction p-value | Risk ratio (men) | Risk ratio (women) |
|---|---|---|---|---|---|---|---|---|
| 2 | | | | | | | | |
| 464 | Acute sinusitis | 5.2 / 3.0 | 1.69 (1.61 – 1.78) | **3.6E-85** | 1.62 (1.53 – 1.73) | **6.5E-07** | 2.00 (1.84 – 2.17) | 1.52 (1.42 – 1.62) |
| 594.1 | Calculus of kidney | 6.6 / 4.2 | 1.56 (1.49 – 1.64) | **5.7E-84** | 1.62 (1.55 – 1.71) | 3.0E-01 | 1.54 (1.45 – 1.62) | 1.62 (1.49 – 1.75) |
| 428.4 | Heart failure with preserved EF [Diastolic heart failure] | 3.5 / 1.9 | 1.87 (1.76 – 2.00) | **5.1E-82** | 2.39 (2.22 – 2.58) | 1.0E+00 | 1.89 (1.73 – 2.06) | 1.86 (1.69 – 2.05) |
| 250.41 | Impaired fasting glucose | 4.6 / 2.6 | 1.73 (1.63 – 1.83) | **3.6E-81** | 1.73 (1.63 – 1.84) | 4.7E-01 | 1.77 (1.64 – 1.91) | 1.68 (1.55 – 1.83) |
| 367.1 | Myopia | 1.5 / 0.4 | 3.20 (2.84 – 3.60) | **9.9E-81** | 1.84 (1.63 – 2.08) | 3.3E-01 | 3.35 (2.85 – 3.95) | 3.02 (2.54 – 3.60) |
| 172.22 | Squamous cell carcinoma | 2.7 / 1.3 | 2.11 (1.95 – 2.28) | **1.3E-80** | 1.46 (1.31 – 1.61) | 4.1E-01 | 2.06 (1.88 – 2.26) | 2.22 (1.93 – 2.56) |
| 428.2 | Heart failure NOS | 3.3 / 1.8 | 1.90 (1.77 – 2.03) | **1.6E-76** | 2.64 (2.44 – 2.86) | 4.2E-04 | 1.76 (1.62 – 1.90) | 2.30 (2.03 – 2.60) |
| 764 | Sciatica | 6.7 / 4.3 | 1.53 (1.46 – 1.60) | **3.0E-76** | 1.57 (1.49 – 1.65) | 5.9E-01 | 1.51 (1.42 – 1.61) | 1.54 (1.45 – 1.64) |
| 261 | Vitamin deficiency | 1.6 / 0.6 | 2.79 (2.50 – 3.12) | **1.9E-75** | 2.02 (1.79 – 2.29) | NA | NA | 2.72 (2.41 – 3.08) |
| 537 | Other disorders of stomach and duodenum | 5.1 / 3.1 | 1.64 (1.56 – 1.73) | **3.7E-75** | 1.55 (1.46 – 1.64) | 1.4E-02 | 1.54 (1.44 – 1.66) | 1.77 (1.63 – 1.91) |
| 411.4 | Coronary atherosclerosis | 12.3 / 9.5 | 1.32 (1.28 – 1.36) | **8.0E-75** | 1.45 (1.40 – 1.50) | 5.1E-03 | 1.29 (1.25 – 1.33) | 1.43 (1.34 – 1.53) |
| 480 | Pneumonia | 8.8 / 6.2 | 1.42 (1.37 – 1.48) | **5.6E-74** | 1.72 (1.64 – 1.79) | 2.1E-01 | 1.39 (1.33 – 1.46) | 1.48 (1.39 – 1.57) |
| 539 | Bariatric surgery | 9.2 / 6.7 | 1.40 (1.35 – 1.45) | **8.3E-74** | 2.06 (1.98 – 2.14) | **2.8E-11** | 1.21 (1.14 – 1.28) | 1.56 (1.49 – 1.63) |
| 296 | Mood disorders | 3.9 / 2.2 | 1.79 (1.68 – 1.90) | **1.3E-73** | 2.02 (1.88 – 2.17) | 3.8E-03 | 1.65 (1.52 – 1.79) | 1.99 (1.81 – 2.19) |
| 274.1 | Gout | 6.1 / 4.1 | 1.53 (1.46 – 1.60) | **4.0E-73** | 1.62 (1.54 – 1.70) | 3.2E-01 | 1.51 (1.44 – 1.59) | 1.63 (1.45 – 1.84) |
| 375.1 | Dry eyes | 3.3 / 1.7 | 1.87 (1.75 – 2.01) | **8.4E-73** | 1.75 (1.61 – 1.89) | 4.3E-01 | 1.94 (1.74 – 2.16) | 1.84 (1.68 – 2.00) |
| 694.2 | Other dyschromia | 8.5 / 5.8 | 1.43 (1.38 – 1.49) | **1.1E-72** | 1.16 (1.10 – 1.21) | **5.6E-09** | 1.60 (1.52 – 1.69) | 1.27 (1.20 – 1.34) |
| 427.22 | Atrial flutter | 3.6 / 2.0 | 1.78 (1.67 – 1.90) | **4.1E-71** | 1.77 (1.64 – 1.91) | 1.9E-01 | 1.82 (1.70 – 1.96) | 1.65 (1.44 – 1.89) |
| 475.9 | Postnasal drip | 3.7 / 2.1 | 1.77 (1.66 – 1.88) | **9.1E-71** | 1.83 (1.70 – 1.98) | **1.0E-04** | 2.00 (1.83 – 2.19) | 1.56 (1.43 – 1.71) |
| 304 | Adjustment reaction | 5.7 / 3.5 | 1.57 (1.49 – 1.65) | **3.9E-70** | 1.52 (1.43 – 1.61) | 1.2E-02 | 1.69 (1.57 – 1.81) | 1.48 (1.39 – 1.59) |
| 599.9 | Other abnormality of urination | 4.5 / 2.7 | 1.66 (1.57 – 1.75) | **9.7E-70** | 1.69 (1.59 – 1.81) | 3.4E-02 | 1.73 (1.61 – 1.85) | 1.53 (1.38 – 1.68) |
| 250.42 | Other abnormal glucose | 8.5 / 6.0 | 1.41 (1.36 – 1.47) | **1.7E-69** | 1.59 (1.52 – 1.66) | 3.0E-01 | 1.39 (1.32 – 1.47) | 1.45 (1.36 – 1.53) |
| 427.6 | Premature beats | 3.5 / 2.0 | 1.79 (1.67 – 1.91) | **6.3E-69** | 1.79 (1.65 – 1.93) | 9.2E-01 | 1.80 (1.66 – 1.94) | 1.77 (1.58 – 1.99) |
| 585.3 | Chronic renal failure [CKD] | 7.4 / 5.3 | 1.44 (1.38 – 1.50) | **9.3E-69** | 1.77 (1.69 – 1.85) | 3.4E-02 | 1.40 (1.33 – 1.47) | 1.54 (1.43 – 1.66) |
| 380.4 | Impacted cerumen | 5.4 / 3.4 | 1.57 (1.49 – 1.65) | **1.1E-68** | 1.55 (1.46 – 1.65) | 1.0E-01 | 1.61 (1.52 – 1.72) | 1.49 (1.37 – 1.62) |
| 290.3 | Other persistent mental | 1.3 / 0.4 | 3.13 (2.75 – 3.56) | **6.2E-68** | 2.31 (2.01 – 2.66) | 1.1E-01 | 3.38 (2.86 – 3.99) | 2.77 (2.26 – 3.40) |

| PheCode | Disease | Prevalence OSA Case / Control | Risk ratio | P-value | Sensitivity analysis risk ratio | OSA x sex interaction p-value | Risk ratio (men) | Risk ratio (women) |
|---|---|---|---|---|---|---|---|---|
| | disorders due to conditions classified elsewhere | | | | | | | |
| 519.8 | Other diseases of respiratory system, NEC | 2.5 / 1.2 | 2.07 (1.91 – 2.25) | **6.3E-68** | 2.51 (2.28 – 2.77) | 6.9E-01 | 2.05 (1.83 – 2.29) | 2.12 (1.88 – 2.40) |
| 471 | Nasal polyps | 1.6 / 0.6 | 2.54 (2.29 – 2.82) | **1.1E-67** | 2.01 (1.79 – 2.27) | NA | 2.76 (2.44 – 3.12) | NA |
| 277 | Other disorders of metabolism | 4.2 / 2.5 | 1.67 (1.57 – 1.77) | **1.3E-65** | 1.76 (1.65 – 1.88) | 6.6E-03 | 1.52 (1.38 – 1.66) | 1.79 (1.66 – 1.93) |
| 379.2 | Disorders of vitreous body | 3.1 / 1.6 | 1.84 (1.71 – 1.97) | **8.0E-65** | 1.52 (1.40 – 1.66) | 2.8E-01 | 1.90 (1.72 – 2.09) | 1.77 (1.59 – 1.96) |
| 557 | Intestinal malabsorption (non-celiac) | 1.5 / 0.6 | 2.65 (2.37 – 2.96) | **8.7E-65** | 2.57 (2.24 – 2.95) | NA | NA | 2.68 (2.34 – 3.06) |
| 366.2 | Senile cataract | 6.3 / 4.2 | 1.48 (1.41 – 1.55) | **1.3E-64** | 1.39 (1.32 – 1.47) | 6.0E-01 | 1.50 (1.41 – 1.59) | 1.46 (1.36 – 1.56) |
| 292.2 | Mild cognitive impairment | 1.8 / 0.8 | 2.29 (2.08 – 2.53) | **3.0E-64** | 2.54 (2.24 – 2.87) | 2.7E-02 | 2.51 (2.21 – 2.84) | 2.01 (1.72 – 2.33) |
| 701 | Other hypertrophic and atrophic conditions of skin | 6.7 / 4.6 | 1.47 (1.40 – 1.53) | **4.2E-64** | 1.54 (1.46 – 1.61) | **3.2E-09** | 1.68 (1.58 – 1.79) | 1.27 (1.19 – 1.36) |
| 350.1 | Abnormal involuntary movements | 4.6 / 2.8 | 1.61 (1.52 – 1.70) | **4.3E-64** | 1.90 (1.78 – 2.02) | 3.4E-01 | 1.64 (1.53 – 1.77) | 1.56 (1.44 – 1.70) |
| 365 | Glaucoma | 5.0 / 3.1 | 1.57 (1.49 – 1.65) | **4.8E-64** | 1.62 (1.53 – 1.72) | 9.7E-01 | 1.57 (1.47 – 1.69) | 1.56 (1.45 – 1.69) |
| 300.9 | Posttraumatic stress disorder | 2.8 / 1.5 | 1.90 (1.76 – 2.04) | **5.8E-64** | 2.08 (1.91 – 2.25) | **9.0E-05** | 1.58 (1.41 – 1.78) | 2.16 (1.96 – 2.38) |
| 429.3 | Symptoms involving cardiovascular system | 6.2 / 4.3 | 1.47 (1.41 – 1.54) | **6.0E-62** | 1.66 (1.58 – 1.76) | 6.4E-01 | 1.49 (1.40 – 1.58) | 1.46 (1.35 – 1.56) |
| 782.3 | Edema | 9.4 / 7.2 | 1.34 (1.30 – 1.39) | **1.4E-61** | 1.81 (1.74 – 1.88) | 8.4E-01 | 1.34 (1.28 – 1.41) | 1.35 (1.28 – 1.42) |
| 532 | Dysphagia | 7.6 / 5.3 | 1.41 (1.36 – 1.47) | **2.7E-61** | 1.56 (1.48 – 1.64) | 4.0E-03 | 1.33 (1.26 – 1.41) | 1.52 (1.43 – 1.62) |
| 496.1 | Emphysema | 3.1 / 1.7 | 1.79 (1.67 – 1.92) | **6.6E-61** | 1.90 (1.74 – 2.07) | 1.9E-01 | 1.72 (1.57 – 1.89) | 1.90 (1.71 – 2.12) |
| 389.4 | Tinnitus | 4.8 / 3.0 | 1.57 (1.48 – 1.65) | **1.5E-60** | 1.48 (1.38 – 1.57) | 1.1E-02 | 1.66 (1.55 – 1.78) | 1.44 (1.32 – 1.56) |
| 262 | Mineral deficiency NEC | 2.0 / 0.9 | 2.15 (1.96 – 2.36) | **3.2E-60** | 2.74 (2.47 – 3.05) | 8.6E-01 | 2.10 (1.80 – 2.45) | 2.18 (1.94 – 2.44) |
| 726 | Peripheral enthesopathies and allied syndromes | 7.1 / 4.9 | 1.43 (1.37 – 1.49) | **2.0E-59** | 1.27 (1.21 – 1.34) | 7.7E-04 | 1.53 (1.44 – 1.62) | 1.30 (1.22 – 1.39) |
| 726.1 | Enthesopathy | 8.2 / 5.9 | 1.39 (1.33 – 1.44) | **2.9E-59** | 1.40 (1.34 – 1.47) | 9.1E-04 | 1.49 (1.40 – 1.58) | 1.31 (1.24 – 1.38) |
| 415.21 | Primary pulmonary hypertension | 1.2 / 0.4 | 2.84 (2.50 – 3.23) | **2.5E-58** | 3.37 (2.90 – 3.91) | NA | NA | 3.58 (2.99 – 4.28) |
| 785 | Abdominal pain | 16.7 / 13.3 | 1.23 (1.20 – 1.26) | **1.3E-57** | 1.20 (1.17 – 1.24) | 6.6E-01 | 1.24 (1.19 – 1.28) | 1.23 (1.19 – 1.27) |
| 743.11 | Osteoporosis NOS | 7.1 / 4.9 | 1.38 (1.33 – 1.44) | **1.3E-56** | 1.40 (1.33 – 1.48) | **2.6E-05** | 1.66 (1.51 – 1.83) | 1.32 (1.27 – 1.38) |
| 300.1 | Agorophobia, social | 2.9 / 1.6 | 1.81 (1.68 – 1.95) | **2.9E-56** | 2.08 (1.92 – 2.26) | 6.4E-03 | 1.62 (1.45 – 1.81) | 1.99 (1.80 – 2.19) |

| PheCode | Disease | Prevalence OSA Case / Control | Risk ratio | P-value | Sensitivity analysis risk ratio | OSA x sex interaction p-value | Risk ratio (men) | Risk ratio (women) |
|---|---|---|---|---|---|---|---|---|
| 2 | phobia, and panic disorder | | | | | | | |
| 530.13 | Barrett's esophagus | 2.2 / 1.1 | 1.96 (1.80 – 2.13) | **1.7E-55** | 1.92 (1.74 – 2.12) | 7.2E-01 | 1.93 (1.74 – 2.14) | 2.01 (1.73 – 2.33) |
| 765 | Cervical radiculitis | 4.0 / 2.5 | 1.61 (1.51 – 1.70) | **2.1E-55** | 1.78 (1.66 – 1.92) | 2.0E-01 | 1.56 (1.44 – 1.69) | 1.67 (1.53 – 1.82) |
| 481 | Influenza | 3.9 / 2.3 | 1.63 (1.54 – 1.74) | **2.6E-55** | 3.39 (2.92 – 3.93) | 1.7E-01 | 1.70 (1.56 – 1.85) | 1.57 (1.44 – 1.71) |
| 433.31 | Transient cerebral ischemia | 5.0 / 3.3 | 1.51 (1.43 – 1.59) | **4.9E-55** | 1.34 (1.26 – 1.42) | 4.4E-01 | 1.53 (1.43 – 1.64) | 1.48 (1.36 – 1.60) |
| 530.14 | Reflux esophagitis | 3.2 / 1.8 | 1.72 (1.61 – 1.84) | **5.6E-55** | 2.02 (1.86 – 2.20) | 1.0E-03 | 1.91 (1.74 – 2.10) | 1.53 (1.39 – 1.69) |
| 381.1 | Otitis media | 2.1 / 1.0 | 2.02 (1.85 – 2.21) | **1.8E-54** | 2.31 (2.09 – 2.55) | 6.4E-01 | 2.07 (1.84 – 2.34) | 1.97 (1.73 – 2.25) |
| 946 | Anaphylactic shock NOS | 1.4 / 0.6 | 2.42 (2.16 – 2.70) | **3.1E-53** | 1.31 (1.17 – 1.46) | 4.3E-01 | 2.55 (2.15 – 3.03) | 2.32 (2.00 – 2.69) |
| 721.1 | Spondylosis without myelopathy | 8.8 / 6.6 | 1.34 (1.29 – 1.39) | **8.8E-53** | 1.18 (1.13 – 1.23) | 8.4E-02 | 1.38 (1.31 – 1.46) | 1.29 (1.22 – 1.36) |
| 1013 | Asphyxia and hypoxemia | 4.1 / 2.7 | 1.57 (1.48 – 1.66) | **2.4E-52** | 1.98 (1.85 – 2.13) | 2.9E-03 | 1.45 (1.35 – 1.57) | 1.75 (1.60 – 1.91) |
| 781 | Symptoms involving nervous and musculoskeletal systems | 3.8 / 2.4 | 1.59 (1.50 – 1.69) | **8.9E-52** | 2.22 (2.07 – 2.38) | 5.6E-01 | 1.57 (1.44 – 1.70) | 1.62 (1.49 – 1.78) |
| 427.11 | Paroxysmal supraventricular tachycardia | 3.5 / 2.1 | 1.63 (1.53 – 1.74) | **2.9E-51** | 1.80 (1.67 – 1.93) | 7.7E-01 | 1.62 (1.48 – 1.76) | 1.65 (1.51 – 1.81) |
| 720 | Spinal stenosis | 4.2 / 2.8 | 1.53 (1.45 – 1.62) | **2.1E-50** | 1.56 (1.47 – 1.66) | 2.6E-01 | 1.50 (1.39 – 1.61) | 1.59 (1.46 – 1.73) |
| 401.21 | Hypertensive heart disease | 2.7 / 1.6 | 1.73 (1.61 – 1.86) | **4.1E-50** | 2.40 (2.21 – 2.61) | 3.7E-01 | 1.70 (1.55 – 1.86) | 1.80 (1.60 – 2.03) |
| 593.2 | Microscopic hematuria | 3.0 / 1.7 | 1.70 (1.58 – 1.82) | **1.1E-49** | 1.46 (1.34 – 1.58) | 2.3E-01 | 1.63 (1.48 – 1.80) | 1.77 (1.60 – 1.95) |
| 348.7 | Coma | 1.5 / 0.6 | 2.26 (2.03 – 2.51) | **2.0E-49** | 2.82 (2.49 – 3.19) | 3.2E-02 | 2.07 (1.81 – 2.37) | 2.66 (2.21 – 3.20) |
| 514.2 | Solitary pulmonary nodule | 5.7 / 4.0 | 1.43 (1.36 – 1.50) | **2.5E-49** | 1.30 (1.23 – 1.38) | 1.4E-01 | 1.48 (1.39 – 1.58) | 1.37 (1.28 – 1.47) |
| 561.1 | Diarrhea | 8.5 / 6.4 | 1.33 (1.28 – 1.39) | **3.5E-49** | 1.67 (1.60 – 1.75) | 4.7E-01 | 1.30 (1.23 – 1.38) | 1.36 (1.29 – 1.44) |
| 302.1 | Decreased libido | 1.5 / 0.7 | 2.20 (1.98 – 2.44) | **3.1E-48** | 2.08 (1.84 – 2.35) | NA | 2.32 (2.05 – 2.62) | NA |
| 386.2 | Peripheral or central vertigo | 2.6 / 1.4 | 1.75 (1.63 – 1.89) | **1.1E-47** | 1.63 (1.49 – 1.78) | 7.5E-01 | 1.78 (1.58 – 2.00) | 1.74 (1.57 – 1.92) |
| 797 | Shock | 1.7 / 0.8 | 2.04 (1.85 – 2.25) | **2.1E-47** | 1.39 (1.28 – 1.52) | 2.9E-01 | 1.96 (1.75 – 2.21) | 2.23 (1.87 – 2.65) |
| 579.8 | Nonspecific abnormal findings in stool contents | 4.4 / 2.9 | 1.51 (1.43 – 1.60) | **3.3E-47** | 1.41 (1.32 – 1.51) | 1.5E-01 | 1.46 (1.35 – 1.57) | 1.58 (1.45 – 1.72) |
| 136 | Other infectious and parasitic diseases | 3.7 / 2.3 | 1.57 (1.48 – 1.67) | **8.1E-47** | 1.61 (1.50 – 1.72) | 1.5E-03 | 1.44 (1.33 – 1.57) | 1.77 (1.62 – 1.94) |
| 411.2 | Myocardial infarction | 6.6 / 4.9 | 1.37 (1.31 – 1.43) | **1.3E-46** | 1.35 (1.29 – 1.41) | 1.0E-01 | 1.34 (1.28 – 1.41) | 1.46 (1.34 – 1.60) |

| PheCode | Disease | Prevalence OSA Case / Control | Risk ratio | P-value | Sensitivity analysis risk ratio | OSA x sex interaction p-value | Risk ratio (men) | Risk ratio (women) |
|---|---|---|---|---|---|---|---|---|
| 280.1 | Iron deficiency anemias, unspecified or not due to blood loss | 4.7 / 3.1 | 1.48 (1.41 – 1.57) | **1.6E-46** | 1.72 (1.61 – 1.82) | 3.2E-01 | 1.43 (1.31 – 1.57) | 1.51 (1.42 – 1.62) |
| 411.3 | Angina pectoris | 6.3 / 4.5 | 1.39 (1.33 – 1.45) | **4.1E-46** | 1.55 (1.47 – 1.63) | 2.7E-02 | 1.44 (1.36 – 1.51) | 1.28 (1.18 – 1.39) |
| 696.41 | Psoriasis vulgaris | 3.2 / 2.0 | 1.62 (1.52 – 1.73) | **4.2E-46** | 1.60 (1.49 – 1.73) | 9.8E-01 | 1.61 (1.48 – 1.75) | 1.63 (1.47 – 1.81) |
| 110.11 | Dermatophytosis of nail | 3.6 / 2.2 | 1.58 (1.48 – 1.68) | **6.5E-46** | 1.43 (1.33 – 1.54) | **6.8E-05** | 1.74 (1.61 – 1.88) | 1.32 (1.18 – 1.47) |
| 440 | Atherosclerosis | 1.1 / 0.5 | 2.47 (2.18 – 2.80) | **6.7E-45** | 2.24 (1.88 – 2.68) | 2.0E-01 | 2.62 (2.23 – 3.08) | 2.24 (1.83 – 2.74) |
| 763 | Thoracic or lumbosacral neuritis or radiculitis, unspecified | 6.1 / 4.4 | 1.39 (1.32 – 1.45) | **1.9E-44** | 1.24 (1.18 – 1.31) | 9.7E-01 | 1.39 (1.31 – 1.48) | 1.38 (1.29 – 1.48) |
| 339 | Other headache syndromes | 10.7 / 8.2 | 1.27 (1.23 – 1.31) | **2.7E-44** | 1.59 (1.53 – 1.65) | 5.1E-01 | 1.25 (1.19 – 1.32) | 1.29 (1.23 – 1.34) |
| 496.2 | Chronic bronchitis | 1.4 / 0.6 | 2.19 (1.96 – 2.44) | **3.0E-44** | 2.72 (2.37 – 3.12) | 6.7E-01 | 2.13 (1.84 – 2.47) | 2.25 (1.91 – 2.66) |
| 290.1 | Dementias | 2.8 / 1.7 | 1.64 (1.53 – 1.76) | **1.5E-43** | 1.87 (1.71 – 2.05) | **2.6E-06** | 1.88 (1.71 – 2.06) | 1.34 (1.20 – 1.49) |
| 599.8 | Other symptoms involving urinary system | 3.2 / 2.0 | 1.59 (1.49 – 1.70) | **6.3E-43** | 1.95 (1.80 – 2.11) | 2.9E-01 | 1.54 (1.43 – 1.67) | 1.70 (1.50 – 1.92) |
| 300.3 | Obsessive-compulsive disorders | 1.1 / 0.5 | 2.37 (2.09 – 2.68) | **9.4E-43** | 2.48 (2.14 – 2.86) | 4.3E-01 | 2.46 (2.09 – 2.89) | 2.26 (1.87 – 2.73) |
| 172.21 | Basal cell carcinoma | 4.6 / 3.2 | 1.44 (1.37 – 1.52) | **3.9E-42** | 1.32 (1.24 – 1.41) | 2.1E-01 | 1.41 (1.32 – 1.50) | 1.51 (1.39 – 1.66) |
| 395.2 | Nonrheumatic aortic valve disorders | 4.7 / 3.4 | 1.43 (1.36 – 1.50) | **5.8E-42** | 1.40 (1.32 – 1.48) | 2.6E-02 | 1.37 (1.29 – 1.46) | 1.57 (1.43 – 1.73) |
| 296.1 | Bipolar | 3.2 / 2.0 | 1.58 (1.48 – 1.69) | **1.4E-41** | 1.62 (1.51 – 1.74) | **5.3E-08** | 1.31 (1.19 – 1.44) | 1.92 (1.75 – 2.11) |
| 246 | Other disorders of thyroid | 2.5 / 1.4 | 1.70 (1.57 – 1.83) | **1.6E-41** | 1.38 (1.25 – 1.52) | 4.2E-01 | 1.62 (1.41 – 1.87) | 1.73 (1.58 – 1.90) |
| 350.2 | Abnormality of gait | 5.4 / 3.9 | 1.39 (1.33 – 1.46) | **2.1E-41** | 1.37 (1.29 – 1.45) | 8.0E-02 | 1.44 (1.35 – 1.54) | 1.33 (1.24 – 1.43) |
| 196 | Radiotherapy | 4.3 / 6.0 | 0.72 (0.68 – 0.75) | **1.3E-40** | 0.80 (0.76 – 0.85) | 1.2E-02 | 0.68 (0.64 – 0.73) | 0.78 (0.72 – 0.85) |
| 741.2 | Stiffness of joint | 2.1 / 1.2 | 1.78 (1.63 – 1.93) | **1.9E-40** | 1.41 (1.28 – 1.56) | 3.0E-02 | 1.95 (1.73 – 2.21) | 1.62 (1.44 – 1.82) |
| 261.2 | Vitamin B-complex deficiencies | 2.4 / 1.4 | 1.69 (1.56 – 1.82) | **2.2E-40** | 1.77 (1.62 – 1.94) | 5.6E-01 | 1.63 (1.45 – 1.83) | 1.73 (1.56 – 1.92) |
| 292.1 | Aphasia/speech disturbance | 2.2 / 1.2 | 1.74 (1.60 – 1.88) | **1.6E-39** | 1.63 (1.48 – 1.79) | 2.9E-01 | 1.79 (1.62 – 1.99) | 1.65 (1.44 – 1.88) |
| 704 | Diseases of hair and hair follicles | 2.3 / 1.4 | 1.71 (1.58 – 1.85) | **2.4E-39** | 1.92 (1.75 – 2.11) | 6.3E-01 | 1.73 (1.57 – 1.91) | 1.67 (1.46 – 1.92) |
| 599.2 | Retention of urine | 4.3 / 3.0 | 1.44 (1.36 – 1.52) | **1.9E-38** | 1.27 (1.19 – 1.36) | 3.4E-03 | 1.37 (1.29 – 1.46) | 1.69 (1.50 – 1.91) |

| PheCode | Disease | Prevalence OSA Case / Control | Risk ratio | P-value | Sensitivity analysis risk ratio | OSA x sex interaction p-value | Risk ratio (men) | Risk ratio (women) |
|---|---|---|---|---|---|---|---|---|
| 807 | Fracture of ribs | 1.7 / 0.9 | 1.86 (1.69 – 2.04) | **2.8E-38** | 1.27 (1.13 – 1.43) | 1.3E-02 | 1.71 (1.52 – 1.92) | 2.18 (1.86 – 2.56) |
| 216 | Benign neoplasm of skin | 10.8 / 8.6 | 1.24 (1.20 – 1.29) | **2.9E-38** | 1.05 (1.01 – 1.09) | **5.8E-23** | 1.44 (1.38 – 1.50) | 1.03 (0.98 – 1.08) |
| 571.5 | Other chronic nonalcoholic liver disease | 4.5 / 3.2 | 1.43 (1.35 – 1.51) | **3.6E-38** | 1.51 (1.42 – 1.60) | 2.0E-02 | 1.53 (1.42 – 1.64) | 1.33 (1.23 – 1.44) |
| 596.5 | Functional disorders of bladder | 2.1 / 1.2 | 1.74 (1.60 – 1.89) | **5.9E-38** | 1.50 (1.36 – 1.65) | 7.8E-01 | 1.74 (1.54 – 1.96) | 1.73 (1.54 – 1.95) |
| 459.9 | Circulatory disease NEC | 4.0 / 2.8 | 1.46 (1.38 – 1.55) | **6.3E-38** | 1.62 (1.52 – 1.73) | 1.3E-04 | 1.60 (1.49 – 1.72) | 1.26 (1.14 – 1.39) |
| 340.1 | Migraine with aura | 1.9 / 1.1 | 1.79 (1.63 – 1.95) | **9.4E-38** | 1.67 (1.49 – 1.87) | 7.9E-02 | 2.02 (1.71 – 2.38) | 1.70 (1.53 – 1.88) |
| 742.9 | Other derangement of joint | 4.3 / 3.0 | 1.44 (1.36 – 1.52) | **2.8E-37** | 1.47 (1.38 – 1.56) | 2.8E-01 | 1.39 (1.28 – 1.51) | 1.48 (1.37 – 1.60) |
| 740.1 | Osteoarthritis; localized | 2.8 / 4.2 | 0.68 (0.64 – 0.72) | **5.2E-37** | 0.92 (0.87 – 0.97) | 4.2E-01 | 0.66 (0.60 – 0.72) | 0.69 (0.63 – 0.75) |
| 427.8 | Sinoatrial node dysfunction (Bradycardia) | 1.9 / 1.1 | 1.74 (1.60 – 1.90) | **1.5E-36** | 1.93 (1.73 – 2.16) | 2.1E-01 | 1.80 (1.63 – 2.00) | 1.61 (1.37 – 1.89) |
| 788 | Syncope and collapse | 7.4 / 5.6 | 1.30 (1.25 – 1.35) | **1.6E-36** | 1.46 (1.40 – 1.54) | 2.0E-02 | 1.35 (1.28 – 1.42) | 1.22 (1.14 – 1.31) |
| 465 | Acute upper respiratory infections of multiple or unspecified sites | 10.0 / 7.8 | 1.25 (1.21 – 1.30) | **2.9E-36** | 1.56 (1.50 – 1.63) | **5.7E-08** | 1.39 (1.32 – 1.47) | 1.15 (1.10 – 1.21) |
| 287.1 | Spontaneous ecchymoses | 1.0 / 0.4 | 2.31 (2.02 – 2.63) | **4.0E-36** | 1.51 (1.33 – 1.72) | NA | NA | 2.25 (1.88 – 2.70) |
| 562.2 | Diverticulitis | 3.6 / 2.5 | 1.47 (1.39 – 1.57) | **1.0E-35** | 1.44 (1.34 – 1.54) | 5.3E-01 | 1.51 (1.39 – 1.64) | 1.44 (1.32 – 1.57) |
| 531.2 | Gastric ulcer | 1.4 / 0.7 | 1.96 (1.76 – 2.17) | **1.5E-35** | 1.36 (1.21 – 1.53) | 1.8E-01 | 1.81 (1.56 – 2.10) | 2.12 (1.83 – 2.46) |
| 054 | Herpes simplex | 3.0 / 1.9 | 1.54 (1.44 – 1.65) | **1.8E-35** | 1.65 (1.52 – 1.80) | 7.9E-01 | 1.52 (1.36 – 1.69) | 1.56 (1.43 – 1.70) |
| 277.7 | Dysmetabolic syndrome X | 1.2 / 0.6 | 2.08 (1.86 – 2.34) | **2.5E-35** | 2.13 (1.89 – 2.39) | 2.3E-01 | 2.26 (1.91 – 2.67) | 1.94 (1.65 – 2.28) |
| 420.3 | Endocarditis | 1.3 / 0.6 | 2.04 (1.82 – 2.28) | **2.6E-35** | 1.46 (1.25 – 1.71) | 1.5E-04 | 1.76 (1.53 – 2.01) | 2.91 (2.36 – 3.59) |
| 241.1 | Nontoxic uninodular goiter | 4.5 / 3.2 | 1.40 (1.33 – 1.47) | **6.9E-35** | 1.40 (1.31 – 1.49) | 6.9E-02 | 1.30 (1.18 – 1.44) | 1.44 (1.36 – 1.54) |
| 728.7 | Fasciitis | 3.6 / 2.4 | 1.47 (1.38 – 1.56) | **1.5E-34** | 1.45 (1.35 – 1.55) | 1.1E-02 | 1.61 (1.47 – 1.76) | 1.37 (1.26 – 1.49) |
| 394.7 | Disease of tricuspid valve | 3.4 / 2.2 | 1.49 (1.40 – 1.59) | **3.0E-34** | 1.33 (1.23 – 1.44) | 1.2E-03 | 1.36 (1.25 – 1.48) | 1.69 (1.54 – 1.86) |
| 426.31 | Right bundle branch block | 2.3 / 1.4 | 1.62 (1.49 – 1.75) | **4.9E-33** | 1.61 (1.47 – 1.77) | 5.8E-02 | 1.69 (1.54 – 1.84) | 1.40 (1.18 – 1.65) |
| 561 | Symptoms involving digestive system | 3.2 / 2.1 | 1.49 (1.40 – 1.59) | **9.1E-33** | 1.67 (1.54 – 1.80) | 4.5E-01 | 1.54 (1.39 – 1.71) | 1.46 (1.34 – 1.59) |
| 527.7 | Disturbance of salivary secretion | 1.8 / 1.0 | 1.73 (1.58 – 1.89) | **2.9E-32** | 2.39 (2.10 – 2.73) | 9.3E-02 | 1.59 (1.39 – 1.81) | 1.86 (1.64 – 2.10) |
| 516.1 | Hemoptysis | 1.7 / 1.0 | 1.76 (1.60 – 1.93) | **3.0E-32** | 2.17 (1.95 – 2.41) | 2.0E-01 | 1.69 (1.51 – 1.90) | 1.92 (1.63 – 2.26) |
| 228.1 | Hemangioma of skin and subcutaneous tissue | 4.5 / 3.2 | 1.38 (1.31 – 1.46) | **3.1E-32** | 1.43 (1.34 – 1.52) | 7.0E-04 | 1.49 (1.39 – 1.60) | 1.24 (1.14 – 1.35) |
| 840 | Sprains and strains | 7.7 / 6.0 | 1.27 (1.22 – 1.33) | **4.2E-32** | 1.14 (1.09 – 1.20) | 5.5E-04 | 1.35 (1.28 – 1.42) | 1.17 (1.09 – 1.25) |

| PheCode | Disease | Prevalence OSA Case / Control | Risk ratio | P-value | Sensitivity analysis risk ratio | OSA x sex interaction p-value | Risk ratio (men) | Risk ratio (women) |
|---|---|---|---|---|---|---|---|---|
| 912 | Insect bite | 2.4 / 1.5 | 1.59 (1.47 – 1.72) | **4.3E-32** | 2.30 (2.09 – 2.53) | 3.5E-01 | 1.53 (1.38 – 1.71) | 1.65 (1.48 – 1.84) |
| 596 | Other disorders of bladder | 2.0 / 1.2 | 1.67 (1.53 – 1.81) | **4.8E-32** | 1.25 (1.13 – 1.37) | 7.9E-01 | 1.66 (1.51 – 1.83) | 1.64 (1.38 – 1.94) |
| 505 | Other pulmonary inflamation or edema | 2.1 / 1.3 | 1.64 (1.51 – 1.78) | **1.3E-31** | 1.88 (1.71 – 2.07) | 5.2E-02 | 1.55 (1.39 – 1.71) | 1.83 (1.60 – 2.11) |
| 502 | Postinflammatory pulmonary fibrosis | 1.4 / 0.8 | 1.85 (1.67 – 2.06) | **1.8E-31** | 1.53 (1.35 – 1.74) | 6.6E-02 | 1.71 (1.50 – 1.96) | 2.08 (1.77 – 2.45) |
| 291.4 | Specific nonpsychotic mental disorders due to brain damage | 1.2 / 0.6 | 2.00 (1.78 – 2.25) | **2.1E-31** | 2.55 (2.20 – 2.96) | 9.2E-01 | 2.00 (1.71 – 2.35) | 2.00 (1.69 – 2.37) |
| 269 | Proteinuria | 2.8 / 1.8 | 1.52 (1.41 – 1.63) | **3.5E-31** | 1.56 (1.45 – 1.69) | 1.3E-01 | 1.60 (1.46 – 1.74) | 1.41 (1.25 – 1.58) |
| 380.1 | Otitis externa | 2.5 / 1.6 | 1.55 (1.44 – 1.67) | **6.3E-31** | 1.63 (1.50 – 1.77) | 1.4E-01 | 1.64 (1.48 – 1.81) | 1.45 (1.29 – 1.63) |
| 382 | Otalgia | 6.6 / 4.9 | 1.30 (1.24 – 1.36) | **8.6E-31** | 1.17 (1.11 – 1.23) | 2.3E-04 | 1.43 (1.34 – 1.53) | 1.21 (1.14 – 1.28) |
| 480.11 | Pneumococcal pneumonia | 2.0 / 1.2 | 1.65 (1.51 – 1.79) | **1.2E-30** | 1.93 (1.74 – 2.13) | 6.3E-01 | 1.67 (1.50 – 1.87) | 1.62 (1.41 – 1.85) |
| 414 | Other forms of chronic heart disease | 3.2 / 2.2 | 1.46 (1.37 – 1.55) | **3.4E-30** | 1.95 (1.81 – 2.10) | 1.4E-01 | 1.51 (1.40 – 1.63) | 1.35 (1.20 – 1.53) |
| 704.1 | Alopecia | 3.4 / 2.3 | 1.44 (1.35 – 1.54) | **7.4E-30** | 1.50 (1.38 – 1.62) | **4.3E-05** | 1.80 (1.59 – 2.04) | 1.34 (1.24 – 1.44) |
| 351 | Other peripheral nerve disorders | 6.6 / 5.1 | 1.29 (1.23 – 1.34) | **8.1E-30** | 1.21 (1.15 – 1.27) | **3.7E-05** | 1.42 (1.34 – 1.51) | 1.17 (1.10 – 1.24) |
| 443.1 | Raynaud's syndrome | 1.4 / 0.7 | 1.84 (1.66 – 2.05) | **9.6E-30** | 1.71 (1.50 – 1.96) | NA | NA | 1.68 (1.48 – 1.91) |
| 965 | Poisoning by analgesics, antipyretics, and antirheumatics | 2.9 / 1.9 | 1.49 (1.39 – 1.60) | **1.1E-29** | 1.54 (1.43 – 1.66) | 7.7E-01 | 1.48 (1.32 – 1.66) | 1.51 (1.38 – 1.65) |
| 286.9 | Abnormal coagulation profile | 2.5 / 1.6 | 1.55 (1.43 – 1.67) | **1.6E-29** | 1.56 (1.44 – 1.69) | 8.2E-01 | 1.54 (1.39 – 1.70) | 1.57 (1.40 – 1.76) |
| 263 | Other nutritional deficiency | 1.2 / 0.6 | 1.93 (1.72 – 2.16) | **2.0E-29** | 2.54 (2.26 – 2.87) | NA | NA | 2.23 (1.93 – 2.56) |
| 565 | Anal and rectal conditions | 3.2 / 2.1 | 1.46 (1.37 – 1.56) | **2.1E-29** | 1.39 (1.29 – 1.51) | 5.3E-01 | 1.44 (1.32 – 1.56) | 1.51 (1.35 – 1.68) |
| 473 | Diseases of the larynx and vocal cords | 2.0 / 1.2 | 1.63 (1.49 – 1.77) | **6.0E-29** | 1.82 (1.65 – 2.02) | 4.5E-01 | 1.67 (1.49 – 1.88) | 1.57 (1.38 – 1.78) |
| 244.2 | Acquired hypothyroidism | 2.4 / 1.6 | 1.54 (1.43 – 1.67) | **6.1E-29** | 1.68 (1.54 – 1.84) | 9.1E-01 | 1.53 (1.34 – 1.75) | 1.55 (1.42 – 1.70) |
| 727.1 | Synovitis and tenosynovitis | 4.3 / 3.1 | 1.37 (1.30 – 1.45) | **6.4E-29** | 1.19 (1.12 – 1.27) | **2.0E-06** | 1.59 (1.47 – 1.72) | 1.21 (1.12 – 1.30) |
| 586.4 | Stricture/obstruction of ureter | 2.0 / 1.3 | 1.60 (1.47 – 1.73) | **2.3E-28** | 1.34 (1.22 – 1.48) | NA | 1.70 (1.56 – 1.86) | NA |
| 766 | Neuralgia, neuritis, and radiculitis NOS | 3.2 / 2.2 | 1.44 (1.35 – 1.54) | **3.4E-28** | 1.42 (1.32 – 1.53) | 1.6E-01 | 1.51 (1.38 – 1.66) | 1.38 (1.25 – 1.51) |
| 416 | Cardiomegaly | 3.8 / 2.8 | 1.38 (1.31 – 1.47) | **5.5E-28** | 1.48 (1.39 – 1.58) | 8.1E-02 | 1.34 (1.25 – 1.43) | 1.51 (1.36 – 1.68) |

| PheCode | Disease | Prevalence OSA Case / Control | Risk ratio | P-value | Sensitivity analysis risk ratio | OSA x sex interaction p-value | Risk ratio (men) | Risk ratio (women) |
|---|---|---|---|---|---|---|---|---|
| 716.9 | Arthropathy NOS | 5.8 / 4.5 | 1.30 (1.24 – 1.36) | **9.3E-28** | 1.45 (1.38 – 1.53) | 8.6E-01 | 1.29 (1.21 – 1.39) | 1.30 (1.22 – 1.39) |
| 706.1 | Acne | 3.0 / 2.0 | 1.45 (1.36 – 1.56) | **5.4E-27** | 1.59 (1.46 – 1.73) | 3.1E-03 | 1.66 (1.49 – 1.85) | 1.34 (1.23 – 1.46) |
| 245.21 | Chronic lymphocytic thyroiditis | 1.5 / 0.9 | 1.72 (1.56 – 1.90) | **9.3E-27** | 1.54 (1.36 – 1.74) | NA | NA | 1.65 (1.47 – 1.84) |
| 427.12 | Paroxysmal ventricular tachycardia | 2.3 / 1.6 | 1.52 (1.41 – 1.64) | **1.1E-26** | 1.42 (1.30 – 1.55) | 5.6E-01 | 1.54 (1.41 – 1.68) | 1.45 (1.22 – 1.72) |
| 008.5 | Bacterial enteritis | 1.4 / 0.7 | 1.78 (1.60 – 1.97) | **1.3E-26** | 1.80 (1.60 – 2.01) | 4.7E-01 | 1.72 (1.47 – 2.01) | 1.84 (1.60 – 2.12) |
| 214 | Lipoma | 2.3 / 1.5 | 1.52 (1.41 – 1.64) | **3.4E-26** | 1.59 (1.46 – 1.73) | 1.1E-03 | 1.69 (1.53 – 1.87) | 1.31 (1.17 – 1.48) |
| 415 | Pulmonary heart disease | 1.2 / 0.7 | 1.82 (1.63 – 2.03) | **3.5E-26** | 1.67 (1.49 – 1.89) | 8.6E-01 | 1.78 (1.53 – 2.08) | 1.85 (1.57 – 2.17) |
| 695.3 | Rosacea | 2.9 / 2.0 | 1.44 (1.35 – 1.55) | **1.1E-25** | 1.40 (1.29 – 1.51) | 1.5E-01 | 1.52 (1.37 – 1.68) | 1.38 (1.26 – 1.52) |
| 591 | Urinary tract infection | 8.0 / 6.4 | 1.23 (1.18 – 1.27) | **2.0E-25** | 1.17 (1.12 – 1.23) | 1.2E-01 | 1.28 (1.20 – 1.37) | 1.19 (1.14 – 1.25) |
| 724.8 | Other symptoms referable to back | 1.4 / 0.8 | 1.71 (1.55 – 1.90) | **2.8E-25** | 1.77 (1.58 – 1.98) | 8.4E-01 | 1.73 (1.50 – 2.00) | 1.70 (1.47 – 1.96) |
| 350.6 | Disturbances of sensation of smell and taste | 1.1 / 0.6 | 1.85 (1.65 – 2.08) | **3.6E-25** | 2.33 (2.02 – 2.67) | 5.3E-01 | 1.91 (1.62 – 2.25) | 1.78 (1.51 – 2.10) |
| 425.1 | Primary/intrinsic cardiomyopathies | 4.0 / 3.0 | 1.35 (1.27 – 1.43) | **4.1E-25** | 1.53 (1.43 – 1.62) | 4.5E-02 | 1.30 (1.22 – 1.39) | 1.49 (1.33 – 1.67) |
| 197 | Chemotherapy | 4.4 / 5.7 | 0.77 (0.74 – 0.81) | **5.3E-25** | 0.75 (0.71 – 0.79) | 4.1E-01 | 0.76 (0.71 – 0.81) | 0.80 (0.74 – 0.86) |
| 172.2 | Other non-epithelial cancer of skin | 5.7 / 4.5 | 1.27 (1.22 – 1.33) | **5.9E-25** | 1.15 (1.09 – 1.21) | 9.2E-01 | 1.27 (1.20 – 1.35) | 1.28 (1.18 – 1.38) |
| 702.1 | Actinic keratosis | 5.5 / 4.3 | 1.28 (1.22 – 1.34) | **7.5E-25** | 1.09 (1.03 – 1.15) | 1.2E-02 | 1.33 (1.26 – 1.41) | 1.17 (1.07 – 1.27) |
| 687.1 | Rash and other nonspecific skin eruption | 8.7 / 7.1 | 1.21 (1.17 – 1.26) | **1.4E-24** | 1.81 (1.74 – 1.89) | 7.6E-03 | 1.14 (1.08 – 1.21) | 1.28 (1.22 – 1.35) |
| 345 | Epilepsy, recurrent seizures, convulsions | 2.4 / 1.6 | 1.48 (1.37 – 1.60) | **1.5E-24** | 1.58 (1.45 – 1.72) | 8.6E-02 | 1.39 (1.26 – 1.54) | 1.62 (1.44 – 1.83) |
| 530.9 | Heartburn | 3.9 / 2.8 | 1.36 (1.28 – 1.44) | **1.7E-24** | 1.45 (1.35 – 1.55) | 1.7E-02 | 1.47 (1.34 – 1.60) | 1.28 (1.19 – 1.39) |
| 473.4 | Voice disturbance | 3.5 / 2.5 | 1.37 (1.29 – 1.46) | **1.9E-24** | 1.66 (1.54 – 1.78) | 3.3E-01 | 1.41 (1.30 – 1.53) | 1.33 (1.22 – 1.46) |
| 512.2 | Painful respiration | 5.7 / 4.4 | 1.28 (1.22 – 1.34) | **2.5E-24** | 1.28 (1.21 – 1.35) | 9.6E-01 | 1.28 (1.20 – 1.37) | 1.29 (1.20 – 1.38) |
| 217.1 | Nevus, non-neoplastic | 4.1 / 3.0 | 1.34 (1.27 – 1.42) | **2.7E-24** | 1.35 (1.26 – 1.44) | 7.3E-04 | 1.48 (1.37 – 1.60) | 1.21 (1.12 – 1.32) |
| 477 | Epistaxis or throat hemorrhage | 2.6 / 1.8 | 1.45 (1.35 – 1.56) | **6.3E-24** | 1.53 (1.41 – 1.67) | 1.4E-01 | 1.52 (1.39 – 1.67) | 1.36 (1.21 – 1.52) |
| 290.2 | Delirium due to conditions classified elsewhere | 2.2 / 1.5 | 1.49 (1.38 – 1.62) | **6.7E-24** | 1.42 (1.31 – 1.54) | 5.2E-03 | 1.38 (1.26 – 1.52) | 1.77 (1.54 – 2.04) |
| 251.1 | Hypoglycemia | 1.4 / 0.9 | 1.69 (1.52 – 1.87) | **7.9E-24** | 2.24 (1.99 – 2.52) | 1.5E-02 | 1.48 (1.28 – 1.72) | 1.92 (1.66 – 2.22) |
| 433 | Cerebrovascular disease | 1.3 / 0.8 | 1.72 (1.55 – 1.91) | **1.3E-23** | 1.33 (1.17 – 1.52) | 4.6E-01 | 1.77 (1.55 – 2.03) | 1.64 (1.38 – 1.94) |

| PheCode | Disease | Prevalence OSA Case / Control | Risk ratio | P-value | Sensitivity analysis risk ratio | OSA x sex interaction p-value | Risk ratio (men) | Risk ratio (women) |
|---|---|---|---|---|---|---|---|---|
| 550.1 | Inguinal hernia | 3.8 / 2.7 | 1.34 (1.27 – 1.42) | **2.3E-23** | 1.41 (1.31 – 1.51) | NA | 1.35 (1.27 – 1.43) | NA |
| 274.11 | Gouty arthropathy | 2.4 / 1.7 | 1.46 (1.35 – 1.57) | **3.9E-23** | 1.38 (1.28 – 1.50) | NA | 1.46 (1.35 – 1.59) | NA |
| 418.1 | Precordial pain | 3.0 / 2.2 | 1.40 (1.31 – 1.49) | **8.5E-23** | 1.57 (1.46 – 1.69) | 5.4E-01 | 1.37 (1.26 – 1.50) | 1.43 (1.29 – 1.59) |
| 772.3 | Muscle weakness | 2.2 / 1.5 | 1.48 (1.37 – 1.60) | **1.1E-22** | 2.30 (2.06 – 2.57) | 2.2E-01 | 1.42 (1.27 – 1.58) | 1.56 (1.39 – 1.76) |
| 789 | Nausea and vomiting | 10.4 / 8.7 | 1.18 (1.14 – 1.22) | **1.4E-22** | 1.33 (1.28 – 1.38) | **1.1E-10** | 1.03 (0.97 – 1.08) | 1.30 (1.24 – 1.35) |
| 367.4 | Presbyopia | 1.7 / 1.0 | 1.58 (1.44 – 1.73) | **1.7E-22** | 1.92 (1.73 – 2.14) | 7.0E-01 | 1.61 (1.42 – 1.83) | 1.55 (1.36 – 1.77) |
| 316 | Substance addiction and disorders | 4.3 / 3.3 | 1.31 (1.24 – 1.38) | **2.6E-22** | 1.50 (1.42 – 1.59) | **4.9E-20** | 1.11 (1.04 – 1.18) | 1.96 (1.77 – 2.16) |
| 550.4 | Umbilical hernia | 2.1 / 1.4 | 1.48 (1.37 – 1.61) | **6.8E-22** | 1.69 (1.55 – 1.84) | 9.1E-01 | 1.49 (1.35 – 1.63) | 1.48 (1.26 – 1.74) |
| 241.2 | Nontoxic multinodular goiter | 2.9 / 2.1 | 1.38 (1.29 – 1.48) | **1.2E-21** | 1.10 (1.01 – 1.19) | 3.9E-02 | 1.57 (1.37 – 1.79) | 1.33 (1.23 – 1.43) |
| 350.3 | Lack of coordination | 1.7 / 1.0 | 1.57 (1.43 – 1.72) | **1.5E-21** | 1.46 (1.30 – 1.64) | 5.3E-01 | 1.61 (1.42 – 1.81) | 1.51 (1.31 – 1.74) |
| 565.1 | Anal and rectal polyp | 1.9 / 1.3 | 1.51 (1.39 – 1.65) | **2.1E-21** | 1.34 (1.21 – 1.48) | 2.2E-02 | 1.64 (1.47 – 1.82) | 1.31 (1.13 – 1.52) |
| 452 | Other venous embolism and thrombosis | 4.4 / 3.4 | 1.30 (1.23 – 1.37) | **2.7E-21** | 1.30 (1.23 – 1.38) | 3.3E-01 | 1.27 (1.18 – 1.36) | 1.34 (1.24 – 1.46) |
| 276.6 | Fluid overload | 2.5 / 1.8 | 1.42 (1.32 – 1.53) | **3.7E-21** | 1.84 (1.70 – 2.00) | 2.5E-02 | 1.35 (1.23 – 1.47) | 1.59 (1.41 – 1.81) |
| 496.21 | Obstructive chronic bronchitis | 1.2 / 0.7 | 1.69 (1.51 – 1.88) | **5.4E-21** | 2.87 (2.52 – 3.27) | NA | NA | 1.91 (1.63 – 2.23) |
| 573.7 | Abnormal results of function study of liver | 4.0 / 3.0 | 1.32 (1.24 – 1.39) | **5.9E-21** | 1.49 (1.40 – 1.59) | 2.4E-04 | 1.45 (1.35 – 1.56) | 1.16 (1.07 – 1.27) |
| 333.1 | Essential tremor | 1.0 / 0.6 | 1.77 (1.57 – 2.00) | **7.6E-21** | 1.77 (1.55 – 2.04) | NA | 1.97 (1.68 – 2.30) | NA |
| 793.2 | Nonspecific abnormal findings on radiological and other examination of other intrathoracic organs (echocardiogram, etc) | 3.0 / 2.2 | 1.37 (1.28 – 1.46) | **9.4E-21** | 1.50 (1.39 – 1.62) | 8.6E-01 | 1.38 (1.26 – 1.51) | 1.36 (1.23 – 1.50) |
| 501 | Pneumonitis due to inhalation of food or vomitus | 1.7 / 1.1 | 1.54 (1.41 – 1.68) | **1.1E-20** | 1.65 (1.48 – 1.85) | 5.5E-01 | 1.50 (1.35 – 1.67) | 1.63 (1.38 – 1.93) |
| 356 | Hereditary and idiopathic peripheral neuropathy | 2.5 / 1.8 | 1.41 (1.31 – 1.52) | **1.7E-20** | 1.66 (1.53 – 1.81) | 7.4E-01 | 1.40 (1.28 – 1.54) | 1.44 (1.28 – 1.62) |
| 724.9 | Other unspecified back disorders | 1.6 / 1.0 | 1.56 (1.42 – 1.71) | **2.0E-20** | 1.43 (1.29 – 1.58) | 9.8E-01 | 1.55 (1.37 – 1.75) | 1.56 (1.36 – 1.80) |
| 735.3 | Hallux valgus (Bunion) | 2.2 / 1.4 | 1.46 (1.35 – 1.58) | **2.0E-20** | 1.35 (1.22 – 1.49) | 1.9E-01 | 1.58 (1.38 – 1.81) | 1.40 (1.27 – 1.55) |
| 722.1 | Displacement of intervertebral disc | 4.9 / 3.9 | 1.27 (1.21 – 1.34) | **2.1E-20** | 1.21 (1.14 – 1.28) | 9.4E-01 | 1.28 (1.19 – 1.36) | 1.27 (1.17 – 1.37) |

| PheCode | Disease | Prevalence OSA Case / Control | Risk ratio | P-value | Sensitivity analysis risk ratio | OSA x sex interaction p-value | Risk ratio (men) | Risk ratio (women) |
|---|---|---|---|---|---|---|---|---|
| 290.11 | Alzheimer's disease | 1.1 / 0.6 | 1.73 (1.54 – 1.94) | **2.3E-20** | 1.65 (1.42 – 1.92) | NA | 2.13 (1.83 – 2.48) | NA |
| 723 | Other disorders of cervical region | 1.1 / 0.6 | 1.74 (1.55 – 1.96) | **3.0E-20** | 1.76 (1.53 – 2.04) | NA | NA | 1.80 (1.56 – 2.09) |
| 737.3 | Kyphoscoliosis and scoliosis | 1.8 / 1.2 | 1.51 (1.38 – 1.65) | **3.4E-20** | 1.51 (1.35 – 1.69) | 7.9E-01 | 1.52 (1.31 – 1.76) | 1.50 (1.34 – 1.67) |
| 306.9 | Tension headache | 1.5 / 0.8 | 1.61 (1.46 – 1.79) | **3.5E-20** | 1.63 (1.46 – 1.81) | NA | NA | 1.49 (1.31 – 1.69) |
| 292.6 | Hallucinations | 1.0 / 0.6 | 1.76 (1.56 – 1.99) | **4.1E-20** | 1.46 (1.30 – 1.64) | NA | NA | 1.96 (1.63 – 2.35) |
| 528 | Diseases of the oral soft tissues, excluding lesions specific for gingiva and tongue | 2.2 / 1.5 | 1.45 (1.34 – 1.58) | **4.3E-20** | 1.63 (1.48 – 1.80) | 1.2E-02 | 1.59 (1.43 – 1.76) | 1.30 (1.15 – 1.47) |
| 394.2 | Mitral valve disease | 1.3 / 0.8 | 1.63 (1.47 – 1.81) | **1.3E-19** | 1.63 (1.42 – 1.86) | 8.4E-01 | 1.61 (1.40 – 1.85) | 1.66 (1.41 – 1.96) |
| 440.9 | Atherosclerosis of aorta | 1.5 / 1.0 | 1.56 (1.42 – 1.72) | **1.3E-19** | 1.75 (1.57 – 1.96) | 6.2E-01 | 1.52 (1.34 – 1.73) | 1.60 (1.39 – 1.85) |
| 078 | Viral warts & HPV | 3.9 / 2.9 | 1.31 (1.24 – 1.39) | **1.8E-19** | 1.10 (1.03 – 1.18) | 1.5E-01 | 1.36 (1.26 – 1.47) | 1.25 (1.15 – 1.37) |
| 426.32 | Left bundle branch block | 1.9 / 1.3 | 1.48 (1.36 – 1.61) | **2.2E-19** | 1.36 (1.23 – 1.50) | 8.2E-01 | 1.49 (1.34 – 1.66) | 1.45 (1.26 – 1.68) |
| 536.8 | Dyspepsia and other specified disorders of function of stomach | 4.0 / 2.9 | 1.31 (1.23 – 1.39) | **3.5E-19** | 1.07 (1.00 – 1.14) | 3.3E-04 | 1.46 (1.35 – 1.59) | 1.19 (1.10 – 1.29) |
| 369.5 | Conjunctivitis, infectious | 3.8 / 2.8 | 1.31 (1.23 – 1.39) | **1.2E-18** | 1.52 (1.42 – 1.63) | 2.9E-02 | 1.41 (1.30 – 1.54) | 1.23 (1.13 – 1.34) |
| 496.3 | Bronchiectasis | 1.1 / 0.6 | 1.68 (1.50 – 1.89) | **1.3E-18** | 1.78 (1.53 – 2.07) | NA | NA | 1.67 (1.41 – 1.97) |
| 569 | Other disorders of intestine | 6.4 / 5.2 | 1.22 (1.17 – 1.27) | **1.3E-18** | 1.25 (1.19 – 1.31) | 2.5E-01 | 1.25 (1.18 – 1.33) | 1.19 (1.11 – 1.27) |
| 458.1 | Orthostatic hypotension | 1.8 / 1.2 | 1.48 (1.36 – 1.62) | **1.5E-18** | 1.54 (1.39 – 1.71) | 5.4E-01 | 1.47 (1.32 – 1.63) | 1.52 (1.31 – 1.77) |
| 443.9 | Peripheral vascular disease, unspecified | 3.1 / 2.3 | 1.32 (1.24 – 1.41) | **1.2E-17** | 1.39 (1.29 – 1.49) | 5.3E-02 | 1.27 (1.18 – 1.37) | 1.46 (1.30 – 1.65) |
| 428.3 | Heart failure with reduced EF [Systolic or combined heart failure] | 2.4 / 1.7 | 1.38 (1.28 – 1.49) | **3.6E-17** | 1.54 (1.41 – 1.68) | 4.2E-01 | 1.36 (1.25 – 1.48) | 1.45 (1.25 – 1.69) |
| 772 | Symptoms of the muscles | 1.2 / 0.7 | 1.62 (1.45 – 1.81) | **4.1E-17** | 1.70 (1.48 – 1.96) | NA | NA | 1.60 (1.37 – 1.86) |
| 447 | Other disorders of arteries and arterioles | 1.3 / 0.8 | 1.57 (1.41 – 1.74) | **6.3E-17** | 1.47 (1.30 – 1.66) | 9.2E-01 | 1.59 (1.38 – 1.82) | 1.55 (1.32 – 1.83) |
| 447.7 | Aortic ectasia | 1.4 / 1.0 | 1.52 (1.38 – 1.67) | **6.9E-17** | 1.62 (1.45 – 1.81) | NA | 1.54 (1.38 – 1.72) | NA |
| 585.4 | Chronic kidney disease, Stage I or II | 2.1 / 1.5 | 1.41 (1.30 – 1.53) | **8.6E-17** | 1.64 (1.50 – 1.80) | 4.4E-01 | 1.45 (1.31 – 1.60) | 1.36 (1.18 – 1.56) |
| 362.29 | Macular degeneration (senile) of retina NOS | 1.1 / 0.7 | 1.61 (1.44 – 1.80) | **1.0E-16** | 1.74 (1.52 – 2.00) | 2.7E-01 | 1.70 (1.45 – 1.98) | 1.51 (1.29 – 1.78) |

| PheCode | Disease | Prevalence OSA Case / Control | Risk ratio | P-value | Sensitivity analysis risk ratio | OSA x sex interaction p-value | Risk ratio (men) | Risk ratio (women) |
|---|---|---|---|---|---|---|---|---|
| 772.2 | Spasm of muscle | 2.3 / 1.6 | 1.38 (1.28 – 1.49) | **1.2E-16** | 2.16 (1.96 – 2.37) | 1.1E-01 | 1.28 (1.14 – 1.44) | 1.46 (1.32 – 1.62) |
| 738.4 | Acquired spondylolisthesis | 2.4 / 1.8 | 1.36 (1.26 – 1.46) | **1.5E-16** | 1.14 (1.05 – 1.23) | 6.6E-01 | 1.33 (1.19 – 1.48) | 1.38 (1.25 – 1.52) |
| 483 | Acute bronchitis and bronchiolitis | 5.2 / 4.2 | 1.23 (1.17 – 1.30) | **2.2E-16** | 1.28 (1.21 – 1.35) | **2.8E-05** | 1.37 (1.28 – 1.47) | 1.11 (1.04 – 1.20) |
| 586 | Other disorders of the kidney and ureters | 5.6 / 4.7 | 1.21 (1.16 – 1.27) | **6.5E-16** | 1.45 (1.38 – 1.52) | 7.3E-02 | 1.25 (1.18 – 1.32) | 1.14 (1.05 – 1.24) |
| 165.1 | Cancer of bronchus; lung | 1.8 / 2.4 | 0.73 (0.67 – 0.79) | **1.2E-15** | 0.67 (0.61 – 0.73) | 8.3E-01 | 0.74 (0.66 – 0.82) | 0.72 (0.64 – 0.81) |
| 361 | Retinal detachments and defects | 1.1 / 0.7 | 1.60 (1.43 – 1.80) | **1.6E-15** | 1.41 (1.23 – 1.61) | NA | 1.58 (1.37 – 1.81) | NA |
| 510 | Other diseases of lung | 5.6 / 4.6 | 1.21 (1.15 – 1.27) | **1.8E-15** | 1.27 (1.20 – 1.34) | 1.6E-02 | 1.15 (1.08 – 1.23) | 1.29 (1.21 – 1.39) |
| 277.51 | Lipoprotein disorders | 1.5 / 1.0 | 1.48 (1.34 – 1.63) | **3.3E-15** | 1.73 (1.55 – 1.93) | 8.3E-01 | 1.49 (1.32 – 1.67) | 1.46 (1.23 – 1.73) |
| 368.2 | Diplopia and disorders of binocular vision | 1.6 / 1.1 | 1.45 (1.32 – 1.59) | **3.5E-15** | 1.69 (1.52 – 1.88) | 2.8E-01 | 1.39 (1.23 – 1.57) | 1.55 (1.34 – 1.79) |
| 288.2 | Elevated white blood cell count | 4.1 / 3.3 | 1.25 (1.18 – 1.32) | **4.0E-15** | 1.32 (1.24 – 1.40) | 7.5E-02 | 1.19 (1.11 – 1.28) | 1.33 (1.22 – 1.45) |
| 593 | Hematuria | 7.0 / 5.9 | 1.18 (1.13 – 1.23) | **4.3E-15** | 1.26 (1.20 – 1.32) | 7.4E-01 | 1.18 (1.12 – 1.24) | 1.19 (1.10 – 1.28) |
| 371.3 | Inflammation of eyelids | 2.5 / 1.9 | 1.33 (1.24 – 1.43) | **5.7E-15** | 1.54 (1.41 – 1.67) | 8.1E-02 | 1.26 (1.15 – 1.39) | 1.43 (1.28 – 1.59) |
| 110.12 | Althete's foot | 2.1 / 1.5 | 1.38 (1.27 – 1.50) | **5.8E-15** | 1.28 (1.17 – 1.40) | 1.6E-04 | 1.53 (1.39 – 1.68) | 1.07 (0.91 – 1.25) |
| 727 | Other disorders of synovium, tendon, and bursa | 1.9 / 1.3 | 1.40 (1.29 – 1.53) | **6.3E-15** | 1.78 (1.60 – 1.97) | 7.0E-01 | 1.38 (1.22 – 1.56) | 1.43 (1.27 – 1.61) |
| 526.41 | Temporomandibular joint disorder, unspecified | 1.2 / 0.8 | 1.54 (1.38 – 1.71) | **7.9E-15** | 2.62 (2.24 – 3.07) | NA | NA | 1.54 (1.35 – 1.75) |
| 690.1 | Seborrheic dermatitis | 2.7 / 2.0 | 1.32 (1.23 – 1.41) | **8.3E-15** | 1.46 (1.34 – 1.58) | 3.0E-03 | 1.42 (1.31 – 1.55) | 1.15 (1.02 – 1.29) |
| 465.4 | Acute laryngitis and tracheitis | 1.3 / 0.9 | 1.50 (1.36 – 1.67) | **9.5E-15** | 1.77 (1.56 – 2.01) | 3.4E-01 | 1.59 (1.36 – 1.86) | 1.44 (1.25 – 1.65) |
| 558 | Noninfectious gastroenteritis | 4.3 / 3.4 | 1.24 (1.18 – 1.31) | **1.2E-14** | 1.18 (1.10 – 1.25) | 6.9E-02 | 1.30 (1.20 – 1.41) | 1.19 (1.11 – 1.29) |
| 252.1 | Hyperparathyroidism | 2.0 / 1.5 | 1.37 (1.26 – 1.49) | **1.8E-14** | 1.29 (1.18 – 1.42) | 1.8E-01 | 1.48 (1.29 – 1.69) | 1.32 (1.19 – 1.46) |
| 008.6 | Viral Enteritis | 1.3 / 0.8 | 1.52 (1.36 – 1.69) | **1.9E-14** | 2.73 (2.42 – 3.08) | 1.7E-01 | 1.42 (1.22 – 1.66) | 1.65 (1.42 – 1.91) |
| 840.3 | Joint/ligament sprain | 3.2 / 2.5 | 1.27 (1.20 – 1.36) | **5.2E-14** | 1.31 (1.21 – 1.41) | 1.6E-01 | 1.33 (1.22 – 1.44) | 1.21 (1.10 – 1.33) |
| 593.1 | Gross hematuria | 2.5 / 1.9 | 1.31 (1.22 – 1.41) | **1.1E-13** | 1.36 (1.26 – 1.48) | 2.5E-01 | 1.34 (1.23 – 1.45) | 1.20 (1.03 – 1.41) |
| 599.3 | Dysuria | 5.1 / 4.1 | 1.21 (1.15 – 1.27) | **1.6E-13** | 1.62 (1.53 – 1.71) | 1.8E-02 | 1.30 (1.20 – 1.41) | 1.15 (1.08 – 1.23) |
| 442.1 | Aortic aneurysm | 2.8 / 2.2 | 1.29 (1.21 – 1.38) | **1.7E-13** | 1.25 (1.16 – 1.34) | 8.0E-02 | 1.26 (1.17 – 1.36) | 1.44 (1.24 – 1.68) |

| PheCode | Disease | Prevalence OSA Case / Control | Risk ratio | P-value | Sensitivity analysis risk ratio | OSA x sex interaction p-value | Risk ratio (men) | Risk ratio (women) |
|---|---|---|---|---|---|---|---|---|
| 411.1 | Unstable angina (intermediate coronary syndrome) | 2.3 / 1.7 | 1.33 (1.23 – 1.43) | **1.7E-13** | 1.29 (1.19 – 1.41) | 3.7E-01 | 1.35 (1.24 – 1.47) | 1.25 (1.06 – 1.47) |
| 458.9 | Hypotension NOS | 4.9 / 4.1 | 1.21 (1.15 – 1.27) | **2.0E-13** | 1.43 (1.35 – 1.52) | 8.0E-01 | 1.20 (1.13 – 1.27) | 1.22 (1.12 – 1.33) |
| 819 | Skull and face fracture and other intercranial injury | 1.7 / 1.2 | 1.40 (1.28 – 1.52) | **2.0E-13** | 1.22 (1.10 – 1.35) | 8.2E-03 | 1.30 (1.17 – 1.44) | 1.69 (1.43 – 1.99) |
| 415.11 | Pulmonary embolism and infarction, acute | 3.1 / 2.5 | 1.27 (1.19 – 1.35) | **2.7E-13** | 1.32 (1.23 – 1.41) | 3.1E-01 | 1.23 (1.13 – 1.33) | 1.32 (1.20 – 1.46) |
| 008 | Intestinal infection | 3.1 / 2.3 | 1.28 (1.20 – 1.37) | **3.4E-13** | 1.42 (1.32 – 1.53) | 6.0E-02 | 1.37 (1.25 – 1.50) | 1.21 (1.10 – 1.33) |
| 426 | Cardiac conduction disorders | 1.4 / 1.0 | 1.43 (1.30 – 1.57) | **5.8E-13** | 1.65 (1.47 – 1.86) | NA | 1.43 (1.28 – 1.60) | NA |
| 250.1 | Type 1 diabetes | 1.3 / 0.9 | 1.46 (1.31 – 1.61) | **5.9E-13** | 1.51 (1.35 – 1.68) | 2.4E-01 | 1.39 (1.22 – 1.59) | 1.58 (1.34 – 1.86) |
| 575.7 | Other disorders of gallbladder | 1.1 / 0.8 | 1.50 (1.34 – 1.68) | **9.2E-13** | 6.20 (4.18 – 9.19) | NA | NA | 1.61 (1.38 – 1.89) |
| 242 | Thyrotoxicosis with or without goiter | 1.9 / 1.4 | 1.35 (1.24 – 1.46) | **1.9E-12** | 1.25 (1.14 – 1.38) | 1.6E-04 | 1.69 (1.47 – 1.94) | 1.19 (1.08 – 1.32) |
| 535.2 | Atrophic gastritis | 1.4 / 0.9 | 1.44 (1.30 – 1.59) | **2.0E-12** | 1.49 (1.34 – 1.65) | 5.9E-01 | 1.39 (1.20 – 1.62) | 1.48 (1.29 – 1.69) |
| 280.2 | Iron deficiency anemia secondary to blood loss (chronic) | 1.7 / 1.2 | 1.38 (1.26 – 1.50) | **2.8E-12** | 1.65 (1.49 – 1.83) | 3.8E-02 | 1.53 (1.34 – 1.74) | 1.25 (1.11 – 1.42) |
| 735.1 | Flat foot | 1.2 / 0.8 | 1.47 (1.32 – 1.64) | **3.2E-12** | 1.61 (1.42 – 1.82) | 3.2E-02 | 1.64 (1.41 – 1.90) | 1.31 (1.12 – 1.54) |
| 715 | Other inflammatory spondylopathies | 1.1 / 0.8 | 1.48 (1.33 – 1.65) | **3.3E-12** | 1.51 (1.34 – 1.71) | 9.7E-03 | 1.72 (1.47 – 2.00) | 1.27 (1.08 – 1.49) |
| 355.1 | Chronic pain syndrome | 1.1 / 0.7 | 1.50 (1.34 – 1.68) | **4.1E-12** | 1.59 (1.40 – 1.80) | NA | NA | 1.43 (1.23 – 1.67) |
| 367.8 | Hypermetropia | 1.1 / 0.7 | 1.49 (1.33 – 1.66) | **4.1E-12** | 1.74 (1.50 – 2.02) | NA | NA | 1.49 (1.27 – 1.74) |
| 368 | Visual disturbances | 5.4 / 4.5 | 1.18 (1.13 – 1.24) | **4.5E-12** | 1.25 (1.19 – 1.32) | 1.5E-01 | 1.23 (1.15 – 1.31) | 1.15 (1.07 – 1.23) |
| 805 | Fracture of vertebral column without mention of spinal cord injury | 2.2 / 1.7 | 1.31 (1.22 – 1.42) | **4.6E-12** | 1.14 (1.04 – 1.25) | 5.7E-01 | 1.29 (1.16 – 1.43) | 1.35 (1.20 – 1.51) |
| 480.1 | Bacterial pneumonia | 1.3 / 0.9 | 1.45 (1.30 – 1.61) | **4.9E-12** | 1.51 (1.34 – 1.70) | 6.3E-01 | 1.42 (1.25 – 1.62) | 1.52 (1.27 – 1.81) |
| 277.5 | Other disorders of lipoid metabolism | 1.0 / 0.7 | 1.50 (1.34 – 1.69) | **6.0E-12** | 1.44 (1.27 – 1.63) | NA | 1.64 (1.42 – 1.89) | NA |
| 721.2 | Spondylosis with myelopathy | 1.0 / 0.7 | 1.50 (1.34 – 1.68) | **6.8E-12** | 1.32 (1.15 – 1.51) | 5.5E-01 | 1.45 (1.25 – 1.68) | 1.57 (1.31 – 1.88) |
| 112 | Candidiasis | 3.9 / 3.1 | 1.23 (1.16 – 1.30) | **6.9E-12** | 1.18 (1.11 – 1.27) | 1.8E-01 | 1.16 (1.05 – 1.28) | 1.27 (1.18 – 1.36) |
| 345.1 | Epilepsy | 1.1 / 0.7 | 1.48 (1.32 – 1.65) | **7.1E-12** | 1.75 (1.55 – 1.97) | 1.8E-01 | 1.39 (1.20 – 1.60) | 1.64 (1.37 – 1.95) |

| PheCode | Disease | Prevalence OSA Case / Control | Risk ratio | P-value | Sensitivity analysis risk ratio | OSA x sex interaction p-value | Risk ratio (men) | Risk ratio (women) |
|---|---|---|---|---|---|---|---|---|
| 110.2 | Dermatomycoses | 1.2 / 0.8 | 1.45 (1.30 – 1.61) | **7.6E-12** | 1.69 (1.49 – 1.91) | 2.1E-02 | 1.59 (1.40 – 1.82) | 1.24 (1.03 – 1.48) |
| 287.3 | Thrombocytopenia | 3.1 / 2.5 | 1.25 (1.17 – 1.33) | **1.0E-11** | 1.21 (1.12 – 1.30) | 3.5E-01 | 1.27 (1.18 – 1.37) | 1.19 (1.06 – 1.34) |
| 276.12 | Hyposmolality and/or hyponatremia | 3.2 / 2.5 | 1.24 (1.17 – 1.33) | **1.1E-11** | 1.10 (1.02 – 1.19) | 6.1E-01 | 1.26 (1.16 – 1.37) | 1.23 (1.12 – 1.35) |
| 726.2 | Synoviopathy | 1.5 / 1.1 | 1.39 (1.26 – 1.53) | **1.4E-11** | 1.16 (1.04 – 1.29) | 9.2E-01 | 1.41 (1.24 – 1.59) | 1.37 (1.18 – 1.60) |
| 979 | Adverse drug events and drug allergies | 5.4 / 4.5 | 1.18 (1.12 – 1.24) | **1.8E-11** | 1.07 (1.02 – 1.14) | 6.2E-01 | 1.16 (1.09 – 1.24) | 1.20 (1.12 – 1.29) |
| 687.2 | Localized superficial swelling, mass, or lump | 5.9 / 5.0 | 1.17 (1.12 – 1.22) | **1.8E-11** | 1.48 (1.41 – 1.56) | 5.6E-01 | 1.15 (1.08 – 1.23) | 1.19 (1.11 – 1.26) |
| 300.13 | Phobia | 1.5 / 1.1 | 1.39 (1.26 – 1.53) | **1.9E-11** | 1.88 (1.66 – 2.14) | 2.7E-03 | 1.65 (1.43 – 1.91) | 1.22 (1.07 – 1.38) |
| 348.8 | Encephalopathy, not elsewhere classified | 2.0 / 1.5 | 1.32 (1.22 – 1.43) | **1.9E-11** | 1.35 (1.24 – 1.47) | 5.1E-01 | 1.29 (1.17 – 1.43) | 1.37 (1.20 – 1.57) |
| 427.61 | Supraventricular premature beats | 2.6 / 2.0 | 1.27 (1.19 – 1.36) | **2.0E-11** | 1.35 (1.24 – 1.47) | 9.4E-01 | 1.27 (1.16 – 1.39) | 1.28 (1.14 – 1.43) |
| 578.9 | Hemorrhage of gastrointestinal tract | 3.8 / 3.1 | 1.22 (1.15 – 1.29) | **2.2E-11** | 1.30 (1.21 – 1.38) | 1.8E-01 | 1.24 (1.16 – 1.33) | 1.16 (1.05 – 1.29) |
| 747.11 | Cardiac shunt/ heart septal defect | 1.5 / 1.0 | 1.39 (1.26 – 1.53) | **2.4E-11** | 1.63 (1.45 – 1.82) | 9.4E-01 | 1.38 (1.21 – 1.56) | 1.40 (1.21 – 1.62) |
| 594.8 | Renal colic | 1.0 / 0.7 | 1.50 (1.33 – 1.68) | **3.0E-11** | 1.18 (1.04 – 1.35) | NA | NA | 1.55 (1.29 – 1.87) |
| 374.3 | Ptosis of eyelid | 1.1 / 0.7 | 1.46 (1.31 – 1.63) | **3.1E-11** | 1.67 (1.46 – 1.91) | NA | NA | 1.50 (1.29 – 1.75) |
| 720.1 | Spinal stenosis of lumbar region | 4.5 / 3.9 | 1.19 (1.13 – 1.25) | **3.1E-11** | 1.15 (1.09 – 1.22) | 3.3E-01 | 1.21 (1.13 – 1.30) | 1.16 (1.07 – 1.25) |
| 279.2 | Autoimmune disease NEC | 3.0 / 2.4 | 1.25 (1.17 – 1.33) | **3.4E-11** | 1.19 (1.11 – 1.29) | 1.7E-01 | 1.33 (1.20 – 1.46) | 1.20 (1.10 – 1.31) |
| 260 | Protein-calorie malnutrition | 2.3 / 1.7 | 1.29 (1.20 – 1.39) | **3.5E-11** | 1.31 (1.20 – 1.44) | 3.1E-01 | 1.24 (1.12 – 1.36) | 1.37 (1.21 – 1.54) |
| 564 | Functional digestive disorders | 2.5 / 1.9 | 1.28 (1.19 – 1.38) | **3.8E-11** | 1.49 (1.37 – 1.62) | 6.4E-02 | 1.36 (1.23 – 1.50) | 1.20 (1.08 – 1.34) |
| 735 | Acquired foot deformities | 1.1 / 0.7 | 1.46 (1.30 – 1.64) | **6.4E-11** | 1.58 (1.39 – 1.80) | NA | 1.45 (1.26 – 1.68) | NA |
| 585.1 | Acute renal failure | 5.6 / 4.9 | 1.17 (1.11 – 1.22) | **6.9E-11** | 1.21 (1.15 – 1.28) | 2.3E-01 | 1.14 (1.08 – 1.21) | 1.22 (1.13 – 1.32) |
| 367.2 | Astigmatism | 1.8 / 1.3 | 1.32 (1.21 – 1.44) | **1.7E-10** | 1.83 (1.65 – 2.02) | 9.0E-01 | 1.33 (1.18 – 1.49) | 1.31 (1.16 – 1.48) |
| 324 | Other CNS infection and poliomyelitis | 1.1 / 0.8 | 1.43 (1.28 – 1.60) | **1.9E-10** | 1.80 (1.53 – 2.11) | 1.3E-01 | 1.32 (1.14 – 1.54) | 1.58 (1.34 – 1.86) |
| 317.1 | Alcoholism | 3.0 / 2.4 | 1.23 (1.16 – 1.32) | **2.0E-10** | 1.19 (1.11 – 1.28) | **7.7E-12** | 1.09 (1.01 – 1.17) | 1.86 (1.63 – 2.13) |
| 250.7 | Diabetic retinopathy | 1.4 / 1.0 | 1.38 (1.25 – 1.52) | **2.3E-10** | 1.39 (1.26 – 1.54) | 3.3E-02 | 1.27 (1.12 – 1.44) | 1.58 (1.34 – 1.85) |
| 722.9 | Other and unspecified disc | 1.7 / 1.3 | 1.33 (1.22 – 1.45) | **2.4E-10** | 1.54 (1.40 – 1.70) | 8.8E-01 | 1.34 (1.19 – 1.51) | 1.32 (1.15 – 1.50) |

| PheCode | Disease | Prevalence OSA Case / Control | Risk ratio | P-value | Sensitivity analysis risk ratio | OSA x sex interaction p-value | Risk ratio (men) | Risk ratio (women) |
|---|---|---|---|---|---|---|---|---|
| | disorder | | | | | | | |
| 941 | Adverse reaction to serum or vaccine | 1.4 / 1.0 | 1.37 (1.25 – 1.52) | **2.8E-10** | 1.53 (1.37 – 1.71) | NA | NA | 1.33 (1.17 – 1.51) |
| 433.8 | Late effects of cerebrovascular disease | 1.6 / 1.2 | 1.33 (1.22 – 1.45) | **4.0E-10** | 1.29 (1.16 – 1.42) | 8.1E-02 | 1.41 (1.26 – 1.57) | 1.20 (1.03 – 1.40) |
| 433.21 | Cerebral artery occlusion, with cerebral infarction | 4.7 / 4.0 | 1.17 (1.12 – 1.23) | **4.7E-10** | 1.38 (1.30 – 1.45) | 3.5E-01 | 1.19 (1.12 – 1.27) | 1.14 (1.05 – 1.24) |
| 841 | Sprains and strains of back and neck | 1.4 / 1.0 | 1.37 (1.24 – 1.51) | **5.9E-10** | 1.41 (1.27 – 1.57) | NA | NA | 1.43 (1.26 – 1.62) |
| 110.13 | Dermatophytosis of the body | 1.5 / 1.1 | 1.35 (1.23 – 1.48) | **6.3E-10** | 1.60 (1.45 – 1.77) | NA | 1.40 (1.25 – 1.56) | NA |
| 727.6 | Rupture of tendon, nontraumatic | 2.1 / 1.6 | 1.28 (1.18 – 1.38) | **9.8E-10** | 1.12 (1.03 – 1.22) | 5.2E-01 | 1.31 (1.19 – 1.44) | 1.23 (1.07 – 1.41) |
| 681.2 | Cellulitis and abscess of face/neck | 1.3 / 1.0 | 1.37 (1.24 – 1.52) | **1.4E-09** | 1.54 (1.34 – 1.77) | 6.0E-01 | 1.41 (1.24 – 1.61) | 1.34 (1.14 – 1.57) |
| 153.2 | Colon cancer | 1.3 / 1.8 | 0.76 (0.69 – 0.83) | **1.5E-09** | 0.76 (0.69 – 0.83) | 2.6E-01 | 0.73 (0.65 – 0.82) | 0.81 (0.70 – 0.93) |
| 504 | Other alveolar and parietoalveolar pneumonopathy | 1.2 / 0.9 | 1.37 (1.24 – 1.52) | **2.5E-09** | 1.59 (1.41 – 1.79) | 5.3E-01 | 1.42 (1.24 – 1.62) | 1.32 (1.12 – 1.54) |
| 531.4 | Peptic ulcer, site unspecified | 1.3 / 1.0 | 1.35 (1.22 – 1.50) | **3.0E-09** | 1.47 (1.31 – 1.64) | 9.0E-01 | 1.34 (1.17 – 1.53) | 1.38 (1.19 – 1.61) |
| 695.8 | Other specified erythematous conditions | 1.8 / 1.4 | 1.29 (1.19 – 1.40) | **3.3E-09** | 1.17 (1.06 – 1.28) | 2.5E-01 | 1.36 (1.19 – 1.55) | 1.25 (1.12 – 1.39) |
| 598.9 | Other nonspecific findings on examination of urine | 2.6 / 2.1 | 1.23 (1.15 – 1.32) | **5.0E-09** | 1.41 (1.31 – 1.53) | 2.7E-01 | 1.19 (1.07 – 1.31) | 1.28 (1.16 – 1.41) |
| 427.7 | Tachycardia NOS | 5.7 / 4.9 | 1.15 (1.10 – 1.20) | **5.6E-09** | 1.33 (1.26 – 1.40) | 3.3E-01 | 1.12 (1.05 – 1.19) | 1.18 (1.10 – 1.27) |
| 592.11 | Acute cystitis | 2.2 / 1.8 | 1.25 (1.16 – 1.34) | **1.1E-08** | 1.48 (1.35 – 1.61) | 3.5E-01 | 1.31 (1.16 – 1.49) | 1.21 (1.10 – 1.33) |
| 594.3 | Calculus of ureter | 1.6 / 1.3 | 1.29 (1.18 – 1.41) | **1.3E-08** | 1.14 (1.04 – 1.27) | 3.2E-02 | 1.38 (1.24 – 1.53) | 1.11 (0.94 – 1.32) |
| 285.21 | Anemia in chronic kidney disease | 1.0 / 0.8 | 1.39 (1.24 – 1.56) | **1.4E-08** | 1.59 (1.40 – 1.80) | NA | 1.40 (1.22 – 1.61) | NA |
| 1004 | Other signs and symptoms involving emotional state | 1.1 / 0.8 | 1.38 (1.24 – 1.55) | **1.5E-08** | 1.60 (1.41 – 1.82) | 9.1E-03 | 1.58 (1.36 – 1.83) | 1.18 (1.00 – 1.40) |
| 571.51 | Cirrhosis of liver without mention of alcohol | 1.2 / 1.6 | 0.76 (0.69 – 0.84) | **2.5E-08** | 0.79 (0.72 – 0.87) | NA | 0.67 (0.60 – 0.75) | NA |
| 286.81 | Primary hypercoagulable state | 1.0 / 0.8 | 1.38 (1.23 – 1.55) | **2.7E-08** | 1.49 (1.32 – 1.68) | NA | NA | 1.39 (1.19 – 1.63) |

| PheCode | Disease | Prevalence OSA Case / Control | Risk ratio | P-value | Sensitivity analysis risk ratio | OSA x sex interaction p-value | Risk ratio (men) | Risk ratio (women) |
|---|---|---|---|---|---|---|---|---|
| 704.8 | Other specified diseases of hair and hair follicles | 2.3 / 1.8 | 1.24 (1.15 – 1.34) | **2.8E-08** | 1.42 (1.30 – 1.55) | **9.0E-05** | 1.38 (1.26 – 1.52) | 1.01 (0.88 – 1.15) |
| 366 | Cataract | 4.4 / 3.8 | 1.16 (1.10 – 1.22) | **4.1E-08** | 2.13 (2.00 – 2.26) | 4.2E-01 | 1.14 (1.06 – 1.22) | 1.18 (1.09 – 1.27) |
| 526 | Diseases of the jaws | 1.3 / 1.0 | 1.33 (1.20 – 1.47) | **4.8E-08** | 1.39 (1.23 – 1.55) | NA | NA | 1.33 (1.16 – 1.52) |
| 574.3 | Cholecystitis without cholelithiasis | 1.9 / 1.5 | 1.26 (1.16 – 1.37) | **5.9E-08** | 1.26 (1.15 – 1.38) | 4.7E-04 | 1.45 (1.29 – 1.63) | 1.08 (0.96 – 1.22) |
| 508 | Pulmonary collapse; interstitial and compensatory emphysema | 8.2 / 7.5 | 1.11 (1.07 – 1.15) | **6.3E-08** | 1.05 (1.01 – 1.10) | 1.1E-01 | 1.09 (1.04 – 1.14) | 1.16 (1.09 – 1.24) |
| 727.4 | Ganglion and cyst of synovium, tendon, and bursa | 1.6 / 1.2 | 1.28 (1.17 – 1.40) | **1.1E-07** | 1.32 (1.18 – 1.47) | 5.9E-02 | 1.40 (1.23 – 1.60) | 1.18 (1.04 – 1.34) |
| 574.1 | Cholelithiasis | 4.3 / 3.7 | 1.15 (1.09 – 1.22) | **1.9E-07** | 1.09 (1.02 – 1.15) | 2.5E-01 | 1.19 (1.10 – 1.29) | 1.12 (1.04 – 1.21) |
| 454.1 | Varicose veins of lower extremity | 2.3 / 1.8 | 1.22 (1.13 – 1.31) | **2.9E-07** | 1.31 (1.20 – 1.42) | 8.9E-01 | 1.25 (1.11 – 1.41) | 1.20 (1.09 – 1.32) |
| 772.6 | Facial weakness | 1.1 / 0.8 | 1.34 (1.20 – 1.50) | **3.0E-07** | 1.47 (1.28 – 1.69) | 7.0E-01 | 1.36 (1.18 – 1.57) | 1.30 (1.09 – 1.55) |
| 509.1 | Respiratory failure | 3.4 / 3.0 | 1.17 (1.10 – 1.24) | **3.1E-07** | 1.31 (1.22 – 1.41) | **1.1E-05** | 1.06 (0.98 – 1.14) | 1.39 (1.26 – 1.53) |
| 703.1 | Ingrowing nail | 1.2 / 0.9 | 1.33 (1.19 – 1.48) | **3.5E-07** | 1.72 (1.53 – 1.93) | 1.6E-01 | 1.44 (1.24 – 1.67) | 1.23 (1.05 – 1.44) |
| 698 | Pruritus and related conditions | 4.3 / 3.6 | 1.15 (1.09 – 1.21) | **3.6E-07** | 1.60 (1.50 – 1.70) | 8.6E-01 | 1.13 (1.04 – 1.23) | 1.16 (1.08 – 1.25) |
| 577.3 | Cyst and pseudocyst of pancreas | 1.1 / 0.8 | 1.33 (1.19 – 1.48) | **3.8E-07** | 1.24 (1.07 – 1.42) | NA | NA | 1.25 (1.07 – 1.46) |
| 452.2 | Deep vein thrombosis [DVT] | 3.8 / 3.3 | 1.16 (1.09 – 1.23) | **4.2E-07** | 1.31 (1.23 – 1.39) | 2.2E-01 | 1.12 (1.04 – 1.21) | 1.21 (1.11 – 1.33) |
| 411.8 | Other chronic ischemic heart disease, unspecified | 2.9 / 2.5 | 1.18 (1.11 – 1.26) | **5.1E-07** | 1.12 (1.04 – 1.21) | 9.8E-01 | 1.18 (1.10 – 1.27) | 1.18 (1.00 – 1.39) |
| 560.4 | Other intestinal obstruction | 2.1 / 1.7 | 1.22 (1.13 – 1.32) | **5.7E-07** | 1.14 (1.04 – 1.25) | 2.6E-01 | 1.26 (1.13 – 1.40) | 1.17 (1.05 – 1.32) |
| 053 | Herpes zoster | 4.5 / 3.9 | 1.14 (1.08 – 1.20) | **6.5E-07** | 1.25 (1.18 – 1.33) | 4.3E-02 | 1.20 (1.12 – 1.30) | 1.09 (1.01 – 1.17) |
| 509.3 | Pulmonary insufficiency or respiratory failure following trauma and surgery | 1.2 / 0.9 | 1.31 (1.18 – 1.45) | **7.0E-07** | 1.20 (1.07 – 1.35) | NA | 1.29 (1.14 – 1.46) | NA |
| 130.1 | Lyme disease | 1.4 / 1.1 | 1.27 (1.15 – 1.40) | **1.0E-06** | 1.28 (1.16 – 1.42) | 8.8E-01 | 1.26 (1.11 – 1.42) | 1.29 (1.11 – 1.50) |
| 503 | Pulmonary congestion and hypostasis | 4.6 / 4.2 | 1.13 (1.08 – 1.19) | **1.1E-06** | 1.40 (1.33 – 1.48) | 6.8E-04 | 1.06 (1.00 – 1.13) | 1.29 (1.18 – 1.40) |
| 550.5 | Ventral hernia | 1.9 / 1.6 | 1.23 (1.13 – 1.33) | **1.3E-06** | 1.37 (1.26 – 1.50) | 7.4E-01 | 1.21 (1.08 – 1.36) | 1.24 (1.10 – 1.40) |
| 938.2 | Chronic dermatitis due to | 3.8 / 3.3 | 1.15 (1.08 – 1.21) | **1.6E-06** | 1.21 (1.13 – 1.29) | 3.0E-01 | 1.17 (1.09 – 1.26) | 1.11 (1.01 – 1.21) |

| PheCode | Disease | Prevalence OSA Case / Control | Risk ratio | P-value | Sensitivity analysis risk ratio | OSA x sex interaction p-value | Risk ratio (men) | Risk ratio (women) |
|---|---|---|---|---|---|---|---|---|
| | solar radiation | | | | | | | |
| 550.6 | Incisional hernia | 1.3 / 1.0 | 1.28 (1.16 – 1.41) | **1.7E-06** | 1.20 (1.08 – 1.34) | 2.1E-02 | 1.13 (0.99 – 1.31) | 1.45 (1.26 – 1.68) |
| 420.2 | Pericarditis | 1.8 / 1.5 | 1.23 (1.13 – 1.34) | **2.1E-06** | 1.21 (1.10 – 1.34) | 7.0E-01 | 1.25 (1.12 – 1.40) | 1.20 (1.05 – 1.37) |
| 315 | Develomental delays and disorders | 1.4 / 1.0 | 1.28 (1.15 – 1.41) | **2.4E-06** | 1.36 (1.25 – 1.49) | 1.3E-02 | 1.16 (1.02 – 1.31) | 1.51 (1.28 – 1.79) |
| 401.3 | Other hypertensive complications | 2.1 / 1.8 | 1.20 (1.11 – 1.30) | **2.9E-06** | 1.33 (1.22 – 1.44) | 4.0E-01 | 1.24 (1.12 – 1.37) | 1.16 (1.03 – 1.31) |
| 1010.1 | Screening for infectious and parasitic diseases | 1.2 / 0.9 | 1.28 (1.16 – 1.43) | **3.3E-06** | 1.25 (1.10 – 1.41) | NA | NA | 1.15 (1.01 – 1.30) |
| 456 | Chronic venous insufficiency [CVI] | 2.7 / 2.4 | 1.17 (1.09 – 1.25) | **4.4E-06** | 1.12 (1.05 – 1.21) | 1.1E-02 | 1.30 (1.19 – 1.43) | 1.06 (0.96 – 1.16) |
| 288.1 | Decreased white blood cell count | 1.2 / 0.9 | 1.26 (1.14 – 1.40) | **8.2E-06** | 1.27 (1.12 – 1.44) | 6.1E-02 | 1.15 (1.00 – 1.33) | 1.40 (1.21 – 1.62) |
| 334 | Degenerative disease of the spinal cord | 1.4 / 1.1 | 1.24 (1.13 – 1.37) | **9.9E-06** | 1.13 (1.02 – 1.26) | 8.7E-01 | 1.25 (1.10 – 1.42) | 1.23 (1.06 – 1.43) |
| 300.8 | Acute reaction to stress | 1.2 / 1.0 | 1.26 (1.13 – 1.39) | **1.5E-05** | 1.40 (1.25 – 1.57) | NA | NA | 1.12 (0.98 – 1.28) |
| 686.1 | Carbuncle and furuncle | 1.2 / 0.9 | 1.26 (1.14 – 1.40) | **1.7E-05** | 1.43 (1.27 – 1.60) | NA | NA | 1.22 (1.07 – 1.40) |
| 276.14 | Hypopotassemia | 3.0 / 2.6 | 1.15 (1.08 – 1.22) | **2.0E-05** | 1.39 (1.29 – 1.50) | 2.6E-01 | 1.20 (1.09 – 1.32) | 1.11 (1.02 – 1.21) |
| 705.8 | Hyperhidrosis | 3.5 / 3.0 | 1.14 (1.07 – 1.21) | **2.5E-05** | 1.27 (1.18 – 1.36) | 3.7E-01 | 1.17 (1.08 – 1.27) | 1.11 (1.02 – 1.21) |
| 454.11 | Varicose veins of lower extremity, symptomtic | 2.2 / 1.9 | 1.17 (1.08 – 1.26) | **5.7E-05** | 1.14 (1.06 – 1.24) | 8.9E-01 | 1.19 (1.06 – 1.34) | 1.16 (1.05 – 1.28) |
| 041 | Bacterial infection NOS | 1.9 / 1.6 | 1.18 (1.09 – 1.28) | **6.0E-05** | 1.33 (1.22 – 1.45) | 3.6E-01 | 1.14 (1.03 – 1.27) | 1.24 (1.09 – 1.40) |
| 279 | Disorders involving the immune mechanism | 5.5 / 5.0 | 1.10 (1.05 – 1.16) | **6.0E-05** | 1.51 (1.44 – 1.59) | 9.6E-01 | 1.10 (1.04 – 1.18) | 1.11 (1.04 – 1.20) |
| 288 | Diseases of white blood cells | 1.1 / 0.8 | 1.25 (1.12 – 1.39) | 9.4E-05 | 1.66 (1.47 – 1.88) | NA | NA | 1.31 (1.11 – 1.55) |
| 276.11 | Hyperosmolality and/or hypernatremia | 1.0 / 0.8 | 1.25 (1.12 – 1.40) | 1.0E-04 | 1.73 (1.50 – 2.00) | NA | 1.20 (1.04 – 1.39) | NA |
| 279.1 | Immunity deficiency | 1.2 / 1.0 | 1.23 (1.11 – 1.37) | 1.1E-04 | 1.47 (1.30 – 1.66) | 5.6E-01 | 1.20 (1.04 – 1.38) | 1.29 (1.11 – 1.51) |
| 599 | Other symptoms/disorders or the urinary system | 2.5 / 2.2 | 1.15 (1.07 – 1.23) | 1.2E-04 | 1.22 (1.13 – 1.32) | 7.7E-01 | 1.16 (1.05 – 1.28) | 1.13 (1.02 – 1.26) |
| 289.4 | Lymphadenitis | 5.5 / 6.0 | 0.92 (0.88 – 0.96) | 1.2E-04 | 0.82 (0.78 – 0.87) | 7.4E-02 | 0.89 (0.83 – 0.94) | 0.96 (0.90 – 1.02) |
| 714.1 | Rheumatoid arthritis | 2.0 / 1.8 | 1.16 (1.07 – 1.25) | 1.8E-04 | 1.12 (1.03 – 1.22) | 5.8E-01 | 1.19 (1.04 – 1.37) | 1.14 (1.04 – 1.26) |
| 525 | Other diseases of the teeth | 1.8 / 1.5 | 1.17 (1.08 – 1.28) | 2.6E-04 | 1.13 (1.03 – 1.25) | 3.2E-03 | 1.04 (0.93 – 1.17) | 1.36 (1.20 – 1.55) |

| PheCode | Disease | Prevalence OSA Case / Control | Risk ratio | P-value | Sensitivity analysis risk ratio | OSA x sex interaction p-value | Risk ratio (men) | Risk ratio (women) |
|---|---|---|---|---|---|---|---|---|
| | and supporting structures | | | | | | | |
| 038 | Septicemia | 2.8 / 2.5 | 1.13 (1.06 – 1.21) | 2.8E-04 | 1.25 (1.16 – 1.34) | 3.1E-01 | 1.15 (1.07 – 1.25) | 1.08 (0.97 – 1.22) |
| 195.1 | Malignant neoplasm, other | 1.8 / 1.5 | 1.17 (1.07 – 1.27) | 2.8E-04 | 1.43 (1.30 – 1.57) | 1.2E-01 | 1.10 (0.99 – 1.23) | 1.27 (1.11 – 1.44) |
| 349 | Other and unspecified disorders of the nervous system | 1.1 / 0.9 | 1.22 (1.09 – 1.36) | 3.1E-04 | 1.80 (1.45 – 2.24) | 9.8E-01 | 1.21 (1.06 – 1.39) | 1.22 (1.03 – 1.45) |
| 783 | Fever of unknown origin | 8.2 / 7.5 | 1.07 (1.03 – 1.11) | 3.9E-04 | 1.32 (1.26 – 1.37) | 3.6E-01 | 1.06 (1.01 – 1.11) | 1.10 (1.03 – 1.17) |
| 110.1 | Dermatophytosis | 1.1 / 0.8 | 1.23 (1.10 – 1.37) | 4.1E-04 | 1.55 (1.37 – 1.75) | 1.0E-01 | 1.33 (1.15 – 1.55) | 1.11 (0.93 – 1.32) |
| 700 | Corns and callosities | 1.0 / 0.9 | 1.22 (1.09 – 1.36) | 5.6E-04 | 1.80 (1.59 – 2.03) | NA | NA | 1.18 (1.00 – 1.39) |
| 577.1 | Acute pancreatitis | 1.7 / 1.4 | 1.16 (1.07 – 1.27) | 7.0E-04 | 1.20 (1.09 – 1.32) | 8.2E-01 | 1.17 (1.04 – 1.31) | 1.16 (1.01 – 1.33) |
| 726.3 | Bursitis | 2.1 / 1.9 | 1.13 (1.05 – 1.22) | 1.2E-03 | 1.33 (1.23 – 1.44) | 3.2E-01 | 1.16 (1.05 – 1.28) | 1.09 (0.96 – 1.23) |
| 717 | Polymyalgia Rheumatica | 1.1 / 1.0 | 1.19 (1.07 – 1.32) | 1.4E-03 | 1.46 (1.29 – 1.65) | 4.6E-02 | 1.30 (1.12 – 1.50) | 1.07 (0.92 – 1.24) |
| 396 | Abnormal heart sounds | 4.8 / 4.4 | 1.08 (1.03 – 1.14) | 1.5E-03 | 1.54 (1.45 – 1.62) | 9.8E-01 | 1.08 (1.01 – 1.16) | 1.09 (1.01 – 1.17) |
| 817 | Concussion | 1.2 / 1.0 | 1.18 (1.06 – 1.30) | 1.9E-03 | 1.62 (1.44 – 1.82) | NA | NA | 1.33 (1.16 – 1.54) |
| 585.31 | Renal dialysis | 1.3 / 1.1 | 1.17 (1.06 – 1.29) | 2.2E-03 | 1.32 (1.19 – 1.47) | NA | 1.14 (1.01 – 1.28) | NA |
| 381.2 | Eustachian tube disorders | 1.9 / 1.6 | 1.14 (1.05 – 1.23) | 2.2E-03 | 2.10 (1.91 – 2.31) | 9.0E-02 | 1.22 (1.09 – 1.37) | 1.05 (0.93 – 1.19) |
| 681.3 | Cellulitis and abscess of arm/hand | 1.8 / 1.6 | 1.14 (1.05 – 1.23) | 2.6E-03 | 1.31 (1.20 – 1.44) | 4.7E-01 | 1.11 (1.00 – 1.23) | 1.19 (1.04 – 1.36) |
| 590 | Pyelonephritis | 1.4 / 1.2 | 1.16 (1.05 – 1.27) | 3.0E-03 | 1.12 (1.01 – 1.24) | NA | NA | 1.14 (1.01 – 1.29) |
| 506 | Empyema and pneumothorax | 3.0 / 2.7 | 1.10 (1.03 – 1.17) | 3.2E-03 | 1.17 (1.09 – 1.26) | 1.7E-01 | 1.07 (0.99 – 1.15) | 1.18 (1.06 – 1.32) |
| 681.1 | Cellulitis and abscess of fingers/toes | 1.7 / 1.5 | 1.14 (1.04 – 1.24) | 3.2E-03 | 1.15 (1.05 – 1.27) | 3.2E-01 | 1.18 (1.06 – 1.32) | 1.09 (0.95 – 1.24) |
| 573 | Other disorders of liver | 4.0 / 3.7 | 1.08 (1.03 – 1.15) | 3.5E-03 | 1.07 (1.00 – 1.14) | 3.7E-04 | 0.99 (0.92 – 1.07) | 1.22 (1.12 – 1.32) |
| 038.3 | Bacteremia | 2.5 / 2.3 | 1.11 (1.03 – 1.19) | 4.5E-03 | 1.19 (1.10 – 1.28) | 9.9E-01 | 1.10 (1.01 – 1.20) | 1.12 (0.99 – 1.27) |
| 479 | Other upper respiratory disease | 9.0 / 8.3 | 1.05 (1.01 – 1.09) | 5.9E-03 | 2.00 (1.92 – 2.08) | **2.8E-09** | 1.16 (1.11 – 1.22) | 0.93 (0.89 – 0.99) |
| 244.1 | Secondary hypothyroidism | 1.3 / 1.1 | 1.15 (1.04 – 1.26) | 6.0E-03 | 1.17 (1.05 – 1.31) | NA | NA | 1.14 (1.01 – 1.29) |
| 612.2 | Hypertrophy of breast (Gynecomastia) | 1.6 / 1.4 | 1.13 (1.04 – 1.23) | 6.0E-03 | 1.44 (1.30 – 1.59) | 1.0E-01 | 1.24 (1.08 – 1.43) | 1.07 (0.96 – 1.20) |
| 442.11 | Abdominal aortic aneurysm | 1.2 / 1.1 | 1.15 (1.04 – 1.27) | 6.9E-03 | 1.30 (1.16 – 1.46) | NA | 1.16 (1.04 – 1.30) | NA |
| 368.9 | Subjective visual disturbances | 1.4 / 1.2 | 1.14 (1.03 – 1.25) | 7.9E-03 | 1.27 (1.14 – 1.41) | 7.0E-01 | 1.16 (1.01 – 1.34) | 1.12 (0.98 – 1.27) |
| 275.3 | Disorders of magnesium | 1.2 / 1.1 | 1.15 (1.04 – 1.27) | 8.5E-03 | 1.29 (1.15 – 1.45) | 1.1E-01 | 1.06 (0.92 – 1.22) | 1.27 (1.09 – 1.47) |

| PheCode | Disease | Prevalence OSA Case / Control | Risk ratio | P-value | Sensitivity analysis risk ratio | OSA x sex interaction p-value | Risk ratio (men) | Risk ratio (women) |
|---|---|---|---|---|---|---|---|---|
| | metabolism | | | | | | | |
| 916 | Contusion | 7.7 / 8.0 | 0.95 (0.92 – 0.99) | 1.2E-02 | 0.81 (0.77 – 0.84) | 1.8E-01 | 0.98 (0.93 – 1.03) | 0.93 (0.88 – 0.98) |
| 276.13 | Hyperpotassemia | 2.4 / 2.2 | 1.09 (1.02 – 1.17) | 1.8E-02 | 1.14 (1.06 – 1.24) | 5.4E-01 | 1.11 (1.02 – 1.21) | 1.06 (0.93 – 1.20) |
| 292.4 | Altered mental status | 5.2 / 4.9 | 1.06 (1.01 – 1.11) | 1.8E-02 | 1.29 (1.22 – 1.36) | 4.4E-01 | 1.07 (1.01 – 1.14) | 1.04 (0.96 – 1.12) |
| 241 | Nontoxic nodular goiter | 1.3 / 1.1 | 1.13 (1.02 – 1.24) | 1.9E-02 | 1.46 (1.31 – 1.62) | NA | NA | 1.12 (1.00 – 1.25) |
| 550 | Abdominal hernia | 2.1 / 2.0 | 1.09 (1.01 – 1.17) | 2.9E-02 | 1.34 (1.24 – 1.46) | 1.6E-01 | 1.03 (0.94 – 1.13) | 1.20 (1.05 – 1.36) |
| 686 | Other local infections of skin and subcutaneous tissue | 1.6 / 1.5 | 1.10 (1.01 – 1.20) | 3.3E-02 | 1.81 (1.69 – 1.95) | 5.6E-01 | 1.13 (1.01 – 1.26) | 1.06 (0.92 – 1.22) |
| 585.2 | Renal failure NOS | 1.9 / 1.8 | 1.09 (1.01 – 1.18) | 3.7E-02 | 1.40 (1.28 – 1.52) | 9.2E-01 | 1.09 (0.99 – 1.20) | 1.10 (0.95 – 1.28) |
| 835 | Internal derangement of knee | 3.6 / 3.3 | 1.06 (1.00 – 1.13) | 3.9E-02 | 1.12 (1.05 – 1.19) | 8.3E-03 | 1.14 (1.06 – 1.23) | 0.98 (0.90 – 1.07) |
| 275.6 | Hypercalcemia | 1.3 / 1.2 | 1.10 (1.00 – 1.22) | 4.5E-02 | 1.14 (1.03 – 1.27) | NA | NA | 1.09 (0.96 – 1.23) |
| 454 | Varicose veins | 1.1 / 1.0 | 1.11 (1.00 – 1.24) | 4.7E-02 | 1.12 (1.03 – 1.23) | NA | 1.12 (0.98 – 1.27) | NA |
| 851 | Complications of transplants and reattached limbs | 2.0 / 1.9 | 1.08 (1.00 – 1.17) | 5.0E-02 | 1.28 (1.17 – 1.39) | 7.0E-01 | 1.10 (0.99 – 1.21) | 1.07 (0.95 – 1.22) |
| 530.2 | Esophageal bleeding (varices/hemorrhage) | 1.7 / 0.8 | 2.21 (2.00 – 2.43) | 7.2E-56 | 1.08 (0.95 – 1.22) | 2.5E-06 | 1.87 (1.65 – 2.11) | 3.06 (2.58 – 3.63) |
| 702.2 | Seborrheic keratosis | 8.6 / 6.5 | 1.33 (1.28 – 1.38) | 1.0E-49 | 1.02 (0.97 – 1.06) | 4.2E-06 | 1.43 (1.36 – 1.50) | 1.20 (1.13 – 1.27) |
| 562.1 | Diverticulosis | 10.9 / 8.7 | 1.27 (1.23 – 1.31) | 6.7E-47 | 1.03 (0.99 – 1.07) | 4.2E-09 | 1.37 (1.32 – 1.43) | 1.12 (1.07 – 1.18) |
| 1011 | Complications of surgical and medical procedures | 6.1 / 4.8 | 1.28 (1.22 – 1.34) | 8.1E-27 | 0.96 (0.91 – 1.01) | 2.1E-02 | 1.35 (1.27 – 1.43) | 1.21 (1.13 – 1.30) |
| 240 | Simple and unspecified goiter | 1.6 / 1.0 | 1.67 (1.52 – 1.84) | 2.0E-26 | 1.02 (0.93 – 1.12) | NA | NA | 1.61 (1.44 – 1.79) |
| 741.3 | Difficulty in walking | 1.8 / 2.7 | 0.69 (0.64 – 0.75) | 2.0E-21 | 0.94 (0.87 – 1.00) | 8.2E-01 | 0.70 (0.63 – 0.77) | 0.69 (0.61 – 0.77) |
| 722.6 | Degeneration of intervertebral disc | 5.9 / 4.8 | 1.25 (1.19 – 1.31) | 2.8E-21 | 1.05 (0.99 – 1.10) | 5.0E-03 | 1.33 (1.25 – 1.42) | 1.16 (1.08 – 1.24) |
| 800 | Fracture of lower limb | 1.3 / 0.9 | 1.53 (1.38 – 1.69) | 8.9E-16 | 1.03 (0.93 – 1.13) | 8.7E-02 | 1.40 (1.20 – 1.63) | 1.66 (1.44 – 1.92) |
| 743.9 | Osteopenia or other disorder of bone and cartilage | 5.5 / 4.4 | 1.20 (1.15 – 1.26) | 7.6E-15 | 1.05 (1.00 – 1.12) | 3.1E-01 | 1.26 (1.15 – 1.39) | 1.19 (1.13 – 1.25) |
| 938 | Dermatitis due to solar radiation | 1.5 / 2.0 | 0.72 (0.66 – 0.79) | 1.2E-13 | 1.08 (0.99 – 1.18) | 8.5E-01 | 0.72 (0.63 – 0.81) | 0.73 (0.65 – 0.82) |

| PheCode | Disease | Prevalence OSA Case / Control | Risk ratio | P-value | Sensitivity analysis risk ratio | OSA x sex interaction p-value | Risk ratio (men) | Risk ratio (women) |
|---|---|---|---|---|---|---|---|---|
| 706.2 | Sebaceous cyst | 4.7 / 3.9 | 1.20 (1.14 – 1.26) | 3.2E-12 | 1.05 (0.99 – 1.11) | 7.5E-03 | 1.28 (1.20 – 1.37) | 1.11 (1.03 – 1.20) |
| 727.5 | Rupture of synovium | 1.4 / 1.0 | 1.36 (1.23 – 1.50) | 1.1E-09 | 1.03 (0.92 – 1.15) | 1.2E-01 | 1.48 (1.28 – 1.71) | 1.26 (1.10 – 1.45) |
| 592.1 | Cystitis | 1.3 / 0.9 | 1.33 (1.20 – 1.47) | 8.1E-08 | 0.98 (0.88 – 1.10) | NA | NA | 1.40 (1.23 – 1.60) |
| 595 | Hydronephrosis | 2.1 / 1.7 | 1.21 (1.12 – 1.30) | 1.7E-06 | 0.97 (0.89 – 1.06) | 3.2E-01 | 1.24 (1.13 – 1.37) | 1.15 (1.01 – 1.30) |
| 292.11 | Aphasia | 1.0 / 0.8 | 1.24 (1.11 – 1.39) | 1.3E-04 | 1.06 (0.94 – 1.21) | NA | 1.25 (1.08 – 1.44) | NA |
| 740.11 | Osteoarthrosis, localized, primary | 10.6 / 10.2 | 1.06 (1.03 – 1.10) | 1.4E-04 | 1.00 (0.96 – 1.03) | 6.5E-03 | 1.12 (1.07 – 1.17) | 1.01 (0.97 – 1.06) |
| 695.9 | Unspecified erythematous condition | 2.1 / 2.4 | 0.86 (0.80 – 0.93) | 1.7E-04 | 1.05 (0.97 – 1.14) | 3.1E-01 | 0.90 (0.81 – 1.00) | 0.84 (0.75 – 0.93) |
| 270.32 | Paraproteinemia | 1.5 / 1.3 | 1.18 (1.08 – 1.29) | 4.1E-04 | 1.06 (0.95 – 1.17) | 7.6E-01 | 1.17 (1.04 – 1.30) | 1.19 (1.02 – 1.39) |
| 317 | Alcohol-related disorders | 1.9 / 1.6 | 1.16 (1.07 – 1.25) | 4.7E-04 | 0.99 (0.91 – 1.09) | 8.9E-06 | 1.05 (0.95 – 1.15) | 1.64 (1.39 – 1.95) |
| 800.1 | Fracture of neck of femur | 1.2 / 1.0 | 1.18 (1.06 – 1.30) | 2.1E-03 | 1.09 (0.96 – 1.23) | 8.0E-01 | 1.16 (1.00 – 1.34) | 1.20 (1.03 – 1.40) |
| 801 | Fracture of ankle and foot | 2.1 / 1.9 | 1.12 (1.03 – 1.21) | 4.7E-03 | 1.07 (0.98 – 1.17) | 2.9E-01 | 1.18 (1.04 – 1.33) | 1.08 (0.98 – 1.20) |
| 228 | Hemangioma and lymphangioma, any site | 2.4 / 2.1 | 1.10 (1.03 – 1.19) | 7.8E-03 | 1.08 (1.00 – 1.18) | 1.5E-02 | 1.21 (1.09 – 1.34) | 1.01 (0.91 – 1.12) |
| 585.32 | End stage renal disease | 1.5 / 1.3 | 1.11 (1.01 – 1.22) | 2.7E-02 | 0.99 (0.90 – 1.09) | 4.9E-01 | 1.09 (0.98 – 1.22) | 1.18 (1.00 – 1.39) |
| 567 | Peritonitis and retroperitoneal infections | 1.0 / 0.9 | 1.13 (1.01 – 1.26) | 3.1E-02 | 1.10 (0.97 – 1.24) | NA | 1.09 (0.95 – 1.25) | NA |
| 577 | Diseases of pancreas | 1.3 / 1.1 | 1.11 (1.01 – 1.23) | 3.3E-02 | 1.02 (0.92 – 1.14) | 1.0E+00 | 1.10 (0.97 – 1.25) | 1.12 (0.96 – 1.31) |
| 255 | Disorders of adrenal glands | 1.1 / 1.0 | 1.11 (1.00 – 1.23) | 5.3E-02 | 1.28 (1.14 – 1.44) | NA | NA | 1.18 (1.02 – 1.36) |
| 426.91 | Cardiac pacemaker in situ | 1.4 / 1.6 | 0.92 (0.84 – 1.01) | 6.6E-02 | 1.70 (1.51 – 1.92) | 9.3E-01 | 0.92 (0.83 – 1.02) | 0.91 (0.77 – 1.08) |
| 202.2 | Non-Hodgkins lymphoma | 1.5 / 1.6 | 0.92 (0.84 – 1.01) | 7.2E-02 | 0.88 (0.80 – 0.97) | 1.4E-01 | 0.88 (0.79 – 0.98) | 1.02 (0.87 – 1.19) |
| 276.41 | Acidosis | 2.1 / 1.9 | 1.07 (0.99 – 1.16) | 8.9E-02 | 1.06 (0.98 – 1.16) | 1.3E-03 | 0.97 (0.88 – 1.07) | 1.27 (1.12 – 1.44) |
| 478 | Throat pain | 2.0 / 1.8 | 1.07 (0.99 – 1.16) | 9.2E-02 | 1.81 (1.64 – 1.99) | 1.5E-01 | 1.14 (1.01 – 1.28) | 1.02 (0.91 – 1.13) |
| 703 | Diseases of nail, NOS | 1.3 / 1.2 | 1.09 (0.99 – 1.20) | 9.2E-02 | 1.51 (1.36 – 1.67) | 2.2E-01 | 1.16 (1.02 – 1.31) | 1.01 (0.87 – 1.18) |
| 530 | Diseases of esophagus | 2.0 / 1.8 | 1.06 (0.98 – 1.15) | 1.4E-01 | 1.18 (1.08 – 1.29) | 6.4E-03 | 1.16 (1.04 – 1.29) | 0.95 (0.84 – 1.07) |
| 540.11 | Acute appendicitis | 1.2 / 1.1 | 1.08 (0.97 – 1.19) | 1.5E-01 | 1.21 (1.08 – 1.37) | 7.2E-01 | 1.07 (0.93 – 1.22) | 1.10 (0.94 – 1.29) |
| 318 | Tobacco use disorder | 7.7 / 7.9 | 0.97 (0.94 – 1.01) | 1.5E-01 | 1.55 (1.48 – 1.62) | 1.1E-05 | 0.90 (0.86 – 0.95) | 1.09 (1.02 – 1.15) |
| 215 | Other benign neoplasm of connective and other soft tissue | 2.0 / 1.9 | 1.06 (0.98 – 1.14) | 1.6E-01 | 1.27 (1.17 – 1.38) | 7.3E-01 | 1.08 (0.96 – 1.21) | 1.04 (0.94 – 1.16) |

| PheCode | Disease | Prevalence OSA Case / Control | Risk ratio | P-value | Sensitivity analysis risk ratio | OSA x sex interaction p-value | Risk ratio (men) | Risk ratio (women) |
|---|---|---|---|---|---|---|---|---|
| 804 | Fracture of hand or wrist | 1.7 / 1.8 | 0.94 (0.87 – 1.02) | 1.6E-01 | 1.13 (1.03 – 1.24) | 3.1E-02 | 0.88 (0.78 – 0.98) | 1.04 (0.92 – 1.18) |
| 041.2 | Streptococcus infection | 1.1 / 1.0 | 1.08 (0.97 – 1.20) | 1.7E-01 | 1.68 (1.44 – 1.95) | 7.5E-01 | 1.09 (0.95 – 1.24) | 1.06 (0.89 – 1.26) |
| 276.1 | Electrolyte imbalance | 1.6 / 1.5 | 1.06 (0.97 – 1.16) | 1.7E-01 | 1.40 (1.27 – 1.54) | 6.1E-01 | 1.05 (0.94 – 1.16) | 1.10 (0.95 – 1.29) |
| 727.2 | Bursitis disorders | 1.3 / 1.3 | 0.93 (0.85 – 1.03) | 1.7E-01 | 1.05 (0.94 – 1.16) | 4.8E-02 | 1.03 (0.89 – 1.19) | 0.86 (0.75 – 0.98) |
| 535.9 | Gastritis and duodenitis, NOS | 4.6 / 4.2 | 1.04 (0.98 – 1.09) | 1.9E-01 | 1.43 (1.36 – 1.51) | 2.6E-02 | 0.98 (0.91 – 1.05) | 1.10 (1.02 – 1.18) |
| 803.2 | Fracture of radius and ulna | 1.4 / 1.3 | 1.06 (0.97 – 1.16) | 2.2E-01 | 1.00 (0.90 – 1.11) | 3.4E-01 | 1.14 (0.98 – 1.31) | 1.02 (0.90 – 1.15) |
| 288.11 | Neutropenia | 1.6 / 1.6 | 0.95 (0.87 – 1.03) | 2.3E-01 | 0.93 (0.84 – 1.03) | 3.3E-01 | 0.98 (0.88 – 1.10) | 0.90 (0.78 – 1.03) |
| 687.4 | Disturbance of skin sensation | 9.1 / 9.2 | 0.98 (0.95 – 1.01) | 2.4E-01 | 1.75 (1.67 – 1.83) | 9.7E-01 | 0.98 (0.94 – 1.03) | 0.98 (0.93 – 1.03) |
| 291.8 | Alteration of consciousness | 1.7 / 1.7 | 0.95 (0.87 – 1.04) | 2.5E-01 | 3.82 (3.41 – 4.28) | 4.7E-02 | 0.89 (0.80 – 0.99) | 1.07 (0.94 – 1.23) |
| 426.21 | First degree AV block | 1.6 / 1.7 | 0.95 (0.88 – 1.04) | 2.6E-01 | 1.28 (1.15 – 1.43) | NA | 0.95 (0.87 – 1.05) | NA |
| 535.1 | Acute gastritis | 1.0 / 1.0 | 0.94 (0.84 – 1.05) | 2.8E-01 | 1.24 (1.11 – 1.38) | NA | NA | 0.89 (0.77 – 1.04) |
| 579.2 | Splenomegaly | 1.5 / 1.4 | 1.05 (0.96 – 1.15) | 2.9E-01 | 1.07 (0.97 – 1.18) | 6.5E-02 | 1.00 (0.90 – 1.11) | 1.22 (1.03 – 1.46) |
| 990 | Effects radiation NOS | 1.6 / 1.7 | 0.96 (0.88 – 1.04) | 3.0E-01 | 0.95 (0.86 – 1.04) | 6.8E-01 | 0.97 (0.87 – 1.09) | 0.94 (0.82 – 1.07) |
| 079.9 | Viremia, NOS | 1.9 / 1.9 | 0.96 (0.88 – 1.04) | 3.0E-01 | 1.24 (1.13 – 1.35) | 8.2E-01 | 0.95 (0.84 – 1.07) | 0.97 (0.88 – 1.09) |
| 803.1 | Fracture of humerus | 1.1 / 1.1 | 1.05 (0.95 – 1.17) | 3.2E-01 | 1.05 (0.94 – 1.18) | NA | NA | 1.08 (0.94 – 1.24) |
| 572 | Ascites (non malignant) | 1.7 / 1.7 | 0.96 (0.88 – 1.04) | 3.3E-01 | 0.78 (0.71 – 0.85) | 6.7E-01 | 0.95 (0.85 – 1.05) | 0.99 (0.86 – 1.14) |
| 870 | Open wounds of head; neck; and trunk | 1.6 / 1.7 | 0.96 (0.88 – 1.05) | 3.5E-01 | 1.13 (1.03 – 1.24) | 1.7E-01 | 0.91 (0.81 – 1.01) | 1.05 (0.92 – 1.21) |
| 286.7 | Other and unspecified coagulation defects | 3.8 / 3.7 | 1.03 (0.97 – 1.09) | 3.5E-01 | 1.24 (1.16 – 1.31) | 9.1E-01 | 1.02 (0.95 – 1.10) | 1.04 (0.95 – 1.13) |
| 199 | Neoplasm of uncertain behavior | 4.5 / 4.4 | 1.02 (0.97 – 1.08) | 3.9E-01 | 0.98 (0.93 – 1.02) | 1.4E-01 | 1.06 (0.99 – 1.14) | 0.98 (0.91 – 1.06) |
| 780 | Hypothermia/Chills | 1.8 / 1.7 | 1.03 (0.95 – 1.12) | 4.6E-01 | 1.35 (1.24 – 1.48) | 2.4E-01 | 1.08 (0.96 – 1.21) | 0.99 (0.88 – 1.12) |
| 738 | Other acquired musculoskeletal deformity | 1.2 / 1.1 | 1.03 (0.93 – 1.14) | 5.4E-01 | 2.47 (1.89 – 3.22) | 9.3E-01 | 1.04 (0.91 – 1.19) | 1.03 (0.88 – 1.20) |
| 345.3 | Convulsions | 3.4 / 3.3 | 1.02 (0.96 – 1.08) | 5.5E-01 | 1.57 (1.46 – 1.68) | 1.9E-01 | 0.98 (0.91 – 1.06) | 1.08 (0.98 – 1.19) |
| 295.3 | Psychosis | 1.2 / 1.2 | 1.03 (0.93 – 1.14) | 5.7E-01 | 1.24 (1.11 – 1.38) | 2.4E-02 | 0.94 (0.82 – 1.07) | 1.21 (1.03 – 1.42) |
| 372 | Disorders of conjunctiva | 1.3 / 1.2 | 1.03 (0.93 – 1.13) | 5.9E-01 | 1.38 (1.26 – 1.52) | 2.9E-01 | 0.98 (0.86 – 1.11) | 1.09 (0.95 – 1.26) |
| 381.11 | Suppurative and unspecified otitis media | 3.0 / 2.8 | 1.02 (0.95 – 1.09) | 6.2E-01 | 1.52 (1.41 – 1.65) | 2.6E-01 | 1.06 (0.97 – 1.16) | 0.98 (0.89 – 1.07) |
| 756.5 | Congenital osteodystrophies | 2.0 / 1.9 | 1.02 (0.94 – 1.10) | 6.4E-01 | 1.47 (1.36 – 1.60) | 4.1E-01 | 0.99 (0.89 – 1.10) | 1.06 (0.94 – 1.19) |

| PheCode | Disease | Prevalence OSA Case / Control | Risk ratio | P-value | Sensitivity analysis risk ratio | OSA x sex interaction p-value | Risk ratio (men) | Risk ratio (women) |
|---|---|---|---|---|---|---|---|---|
| 080 | Postoperative infection | 1.4 / 1.4 | 1.02 (0.93 – 1.12) | 6.4E-01 | 1.14 (1.04 – 1.26) | 9.6E-01 | 1.02 (0.90 – 1.15) | 1.03 (0.90 – 1.18) |
| 509.2 | Respiratory insufficiency | 1.2 / 1.2 | 1.02 (0.92 – 1.13) | 6.8E-01 | 1.69 (1.53 – 1.87) | 1.7E-04 | 0.88 (0.78 – 1.01) | 1.32 (1.12 – 1.55) |
| 459.1 | Hemorrhage NOS | 2.0 / 1.9 | 1.02 (0.94 – 1.10) | 6.9E-01 | 1.96 (1.76 – 2.17) | 6.9E-01 | 1.03 (0.93 – 1.15) | 1.00 (0.89 – 1.13) |
| 189.21 | Malignant neoplasm of bladder | 1.2 / 1.2 | 1.02 (0.92 – 1.12) | 7.3E-01 | 1.03 (0.93 – 1.15) | NA | 1.06 (0.95 – 1.18) | NA |
| 285.1 | Acute posthemorrhagic anemia | 2.5 / 2.5 | 1.01 (0.94 – 1.08) | 7.9E-01 | 1.15 (1.07 – 1.24) | 6.3E-01 | 1.02 (0.93 – 1.11) | 1.00 (0.89 – 1.12) |
| 193 | Thyroid cancer | 1.1 / 1.1 | 0.99 (0.89 – 1.10) | 8.3E-01 | 0.99 (0.89 – 1.11) | NA | NA | 1.03 (0.90 – 1.18) |
| 743.2 | Pathologic fracture | 1.1 / 1.1 | 0.99 (0.89 – 1.10) | 8.6E-01 | 1.01 (0.89 – 1.14) | NA | NA | 1.03 (0.90 – 1.18) |
| 809 | Fracture of unspecified bones | 3.4 / 3.4 | 1.00 (0.95 – 1.06) | 9.3E-01 | 1.03 (0.97 – 1.09) | 3.7E-03 | 0.92 (0.85 – 1.00) | 1.10 (1.01 – 1.19) |
| 771.1 | Swelling of limb | 7.4 / 7.4 | 1.00 (0.96 – 1.04) | 9.5E-01 | 1.41 (1.36 – 1.47) | 3.8E-01 | 0.99 (0.94 – 1.04) | 1.02 (0.97 – 1.08) |
| 297.1 | Suicidal ideation | 1.1 / 1.1 | 1.00 (0.90 – 1.11) | 9.5E-01 | 1.37 (1.22 – 1.53) | NA | NA | 1.52 (1.30 – 1.79) |
| 070.9 | Hepatitis NOS | 2.1 / 2.0 | 1.00 (0.92 – 1.08) | 9.7E-01 | 1.06 (0.98 – 1.15) | 4.9E-02 | 0.94 (0.86 – 1.04) | 1.12 (0.99 – 1.27) |
| 338.1 | Acute pain | 9.5 / 13.2 | 0.72 (0.70 – 0.75) | 2.9E-88 | 1.38 (1.34 – 1.42) | 1.2E-10 | 0.65 (0.62 – 0.68) | 0.80 (0.77 – 0.84) |
| 695.7 | Prurigo and Lichen | 2.3 / 1.1 | 2.11 (1.94 – 2.30) | 1.7E-67 | 0.91 (0.83 – 0.99) | 5.6E-04 | 2.47 (2.18 – 2.79) | 1.85 (1.65 – 2.07) |
| 726.4 | Calcaneal spur; Exostosis NOS | 1.4 / 2.9 | 0.49 (0.45 – 0.53) | 2.9E-63 | 1.14 (1.04 – 1.26) | NA | NA | 0.50 (0.45 – 0.56) |
| 289 | Other diseases of blood and blood-forming organs | 1.2 / 2.4 | 0.51 (0.47 – 0.56) | 8.5E-47 | 1.43 (1.32 – 1.54) | 4.4E-01 | 0.53 (0.47 – 0.60) | 0.50 (0.43 – 0.57) |
| 512.3 | Abnormal chest sounds | 1.0 / 2.0 | 0.51 (0.46 – 0.56) | 9.0E-39 | 1.70 (1.51 – 1.93) | 1.1E-02 | 0.58 (0.50 – 0.66) | 0.44 (0.38 – 0.52) |
| 1002 | Symptoms concerning nutrition, metabolism, and development | 2.4 / 3.6 | 0.64 (0.60 – 0.69) | 1.5E-38 | 3.18 (2.94 – 3.45) | 7.2E-03 | 0.59 (0.54 – 0.64) | 0.71 (0.65 – 0.79) |
| 741 | Symptoms and disorders of the joints | 3.1 / 4.2 | 0.73 (0.68 – 0.77) | 3.9E-25 | 1.38 (1.29 – 1.47) | 6.9E-01 | 0.72 (0.66 – 0.78) | 0.74 (0.68 – 0.80) |
| 497 | Bronchitis | 6.2 / 5.0 | 1.24 (1.19 – 1.30) | 3.9E-21 | 0.91 (0.87 – 0.96) | 7.1E-07 | 1.39 (1.31 – 1.48) | 1.11 (1.04 – 1.19) |
| 949 | Allergies, other | 7.2 / 8.3 | 0.85 (0.81 – 0.88) | 1.8E-17 | 1.96 (1.87 – 2.06) | 7.0E-01 | 0.85 (0.80 – 0.90) | 0.84 (0.80 – 0.88) |
| 578.2 | Blood in stool | 4.2 / 5.1 | 0.80 (0.76 – 0.84) | 6.2E-17 | 1.54 (1.45 – 1.64) | 2.4E-02 | 0.84 (0.79 – 0.90) | 0.75 (0.69 – 0.81) |
| 740.2 | Osteoarthrosis, generalized | 1.1 / 1.7 | 0.66 (0.60 – 0.73) | 6.8E-17 | 1.34 (1.23 – 1.47) | NA | NA | 0.69 (0.61 – 0.77) |
| 915 | Superficial injury without mention of infection | 3.1 / 4.0 | 0.78 (0.73 – 0.82) | 9.6E-17 | 1.61 (1.49 – 1.73) | 9.4E-01 | 0.78 (0.72 – 0.84) | 0.78 (0.70 – 0.86) |
| 939 | Atopic/contact dermatitis due to other or unspecified | 9.2 / 10.2 | 0.88 (0.85 – 0.91) | 3.7E-13 | 1.19 (1.14 – 1.24) | 1.6E-05 | 0.95 (0.90 – 1.00) | 0.82 (0.78 – 0.86) |
| 512.1 | Wheezing | 3.6 / 4.4 | 0.82 (0.77 – 0.86) | 1.6E-12 | 1.41 (1.33 – 1.49) | 1.3E-01 | 0.78 (0.72 – 0.84) | 0.85 (0.79 – 0.92) |

| PheCode | Disease | Prevalence OSA Case / Control | Risk ratio | P-value | Sensitivity analysis risk ratio | OSA x sex interaction p-value | Risk ratio (men) | Risk ratio (women) |
|---|---|---|---|---|---|---|---|---|
| 530.1 | Esophagitis, GERD and related diseases | 2.1 / 2.5 | 0.82 (0.77 – 0.89) | 3.0E-07 | 1.16 (1.08 – 1.24) | 3.5E-02 | 0.88 (0.80 – 0.97) | 0.76 (0.67 – 0.85) |
| 681 | Superficial cellulitis and abscess | 4.3 / 4.9 | 0.88 (0.83 – 0.92) | 6.5E-07 | 1.17 (1.11 – 1.23) | 3.7E-02 | 0.92 (0.86 – 0.98) | 0.83 (0.76 – 0.90) |
| 507 | Pleurisy; pleural effusion | 6.6 / 6.0 | 1.11 (1.06 – 1.16) | 8.9E-07 | 0.94 (0.89 – 0.99) | 3.8E-01 | 1.13 (1.07 – 1.18) | 1.08 (1.01 – 1.17) |
| 465.2 | Acute pharyngitis | 9.5 / 10.1 | 0.92 (0.89 – 0.95) | 4.1E-06 | 1.29 (1.25 – 1.34) | 1.7E-01 | 0.95 (0.90 – 1.00) | 0.90 (0.86 – 0.95) |
| 561.2 | Flatulence | 5.1 / 5.5 | 0.90 (0.86 – 0.94) | 6.2E-06 | 1.59 (1.50 – 1.70) | 3.8E-01 | 0.92 (0.86 – 0.99) | 0.88 (0.83 – 0.94) |
| 681.5 | Cellulitis and abscess of leg, except foot | 1.5 / 1.9 | 0.82 (0.76 – 0.90) | 7.9E-06 | 1.37 (1.26 – 1.49) | 7.5E-01 | 0.82 (0.73 – 0.91) | 0.84 (0.73 – 0.97) |
| 741.4 | Joint effusions | 8.2 / 7.6 | 1.09 (1.05 – 1.13) | 1.7E-05 | 0.94 (0.90 – 0.98) | 3.2E-03 | 1.15 (1.09 – 1.21) | 1.02 (0.96 – 1.08) |
| 272.9 | Unspecified disorder of lipoid metabolism | 1.2 / 1.4 | 0.87 (0.79 – 0.96) | 5.0E-03 | 1.60 (1.43 – 1.78) | 9.2E-01 | 0.87 (0.77 – 0.99) | 0.87 (0.73 – 1.02) |
| 297 | Suicidal ideation or attempt | 2.2 / 2.4 | 0.91 (0.85 – 0.98) | 1.1E-02 | 1.22 (1.12 – 1.33) | 2.3E-02 | 0.85 (0.78 – 0.93) | 1.04 (0.92 – 1.17) |
| 276.5 | Hypovolemia | 7.5 / 7.1 | 1.04 (1.00 – 1.09) | 3.0E-02 | 0.91 (0.87 – 0.95) | 2.0E-01 | 1.07 (1.01 – 1.12) | 1.02 (0.96 – 1.08) |
| 830 | Dislocation | 3.6 / 3.4 | 1.07 (1.01 – 1.13) | 3.0E-02 | 0.91 (0.85 – 0.97) | 3.2E-04 | 1.16 (1.08 – 1.25) | 0.95 (0.86 – 1.04) |
| 079 | Viral infection | 5.0 / 4.4 | 1.06 (1.00 – 1.12) | 3.3E-02 | 0.80 (0.76 – 0.85) | 1.1E-05 | 1.19 (1.11 – 1.28) | 0.95 (0.88 – 1.02) |
| 401.22 | Hypertensive chronic kidney disease | 2.3 / 2.5 | 0.93 (0.87 – 1.00) | 4.3E-02 | 1.49 (1.39 – 1.61) | 2.5E-01 | 0.90 (0.83 – 0.98) | 0.99 (0.88 – 1.12) |

**Table S6. Temporal stability of lead cross-sectional case-control associations.**

Cross-sectional associations were further tested in a sensitivity analysis censoring data a number of years away from the first sleep apnea results (or the matching age for controls). Data were censored at a given relative year, meaning that future comorbidity cases were classified as controls for that year prior to the initial comorbidity diagnosis. Analyses were adjusted for age, sex, BMI, population, and healthcare encounters since age 18 to a given relative year. BMIs were based on measurements relative to a given age. Data from the lead 50 Table S5 results and other PheCodes with p <1e-10 at year -5 are shown for practicality, given a PDF table format and >4,600 total rows. Results are ordered by p-value at year -5.

| PheCode | Disease | Years to OSA diagnosis | Risk ratio | P-value | N |
|---|---|---|---|---|---|
| 272.1 | Hyperlipidemia | -5 | 1.36 (1.31 – 1.40) | 2.21E-74 | 85312 |
| 272.1 | Hyperlipidemia | -4 | 1.39 (1.35 – 1.44) | 1.10E-98 | 90932 |
| 272.1 | Hyperlipidemia | -3 | 1.44 (1.39 – 1.48) | 1.13E-130 | 96622 |
| 272.1 | Hyperlipidemia | -2 | 1.49 (1.45 – 1.53) | 7.13E-179 | 102600 |
| 272.1 | Hyperlipidemia | -1 | 1.55 (1.51 – 1.59) | 6.82E-242 | 108657 |
| 272.1 | Hyperlipidemia | 0 | 1.58 (1.54 – 1.62) | 2.94E-301 | 117551 |
| 272.1 | Hyperlipidemia | 1 | 1.83 (1.79 – 1.87) | 0.00E+00 | 124868 |
| 272.1 | Hyperlipidemia | 2 | 1.85 (1.81 – 1.89) | 0.00E+00 | 127897 |
| 272.1 | Hyperlipidemia | 3 | 1.85 (1.81 – 1.89) | 0.00E+00 | 130482 |
| 272.1 | Hyperlipidemia | 4 | 1.85 (1.81 – 1.89) | 0.00E+00 | 132711 |
| 272.1 | Hyperlipidemia | 5 | 1.85 (1.81 – 1.89) | 0.00E+00 | 134656 |
| 472 | Chronic pharyngitis and nasopharyngitis | -5 | 3.40 (2.98 – 3.88) | 1.39E-72 | 85312 |
| 472 | Chronic pharyngitis and nasopharyngitis | -4 | 3.59 (3.17 – 4.07) | 3.51E-88 | 90932 |
| 472 | Chronic pharyngitis and nasopharyngitis | -3 | 3.65 (3.24 – 4.11) | 3.23E-100 | 96622 |
| 472 | Chronic pharyngitis and nasopharyngitis | -2 | 3.66 (3.27 – 4.10) | 2.87E-114 | 102600 |
| 472 | Chronic pharyngitis and nasopharyngitis | -1 | 3.83 (3.45 – 4.25) | 2.97E-139 | 108657 |
| 472 | Chronic pharyngitis and nasopharyngitis | 0 | 4.14 (3.76 – 4.56) | 5.95E-180 | 117551 |
| 472 | Chronic pharyngitis and nasopharyngitis | 1 | 5.90 (5.41 – 6.44) | 0.00E+00 | 124868 |
| 472 | Chronic pharyngitis and nasopharyngitis | 2 | 6.10 (5.62 – 6.63) | 0.00E+00 | 127897 |
| 472 | Chronic pharyngitis and nasopharyngitis | 3 | 6.22 (5.74 – 6.74) | 0.00E+00 | 130482 |
| 472 | Chronic pharyngitis and nasopharyngitis | 4 | 6.38 (5.90 – 6.90) | 0.00E+00 | 132711 |
| 472 | Chronic pharyngitis and nasopharyngitis | 5 | 6.44 (5.97 – 6.95) | 0.00E+00 | 134656 |
| 530.11 | GERD | -5 | 1.44 (1.38 – 1.51) | 1.66E-61 | 85312 |
| 530.11 | GERD | -4 | 1.47 (1.41 – 1.53) | 4.49E-77 | 90932 |
| 530.11 | GERD | -3 | 1.48 (1.43 – 1.54) | 2.55E-93 | 96622 |
| 530.11 | GERD | -2 | 1.50 (1.45 – 1.56) | 9.83E-113 | 102600 |

| PheCode | Disease | Years to OSA diagnosis | Risk ratio | P-value | N |
|---|---|---|---|---|---|
| 530.11 | GERD | -1 | 1.55 (1.50 – 1.60) | 5.77E-149 | 108657 |
| 530.11 | GERD | 0 | 1.61 (1.56 – 1.66) | 1.05E-206 | 117551 |
| 530.11 | GERD | 1 | 2.00 (1.95 – 2.06) | 0.00E+00 | 124868 |
| 530.11 | GERD | 2 | 2.01 (1.96 – 2.06) | 0.00E+00 | 127897 |
| 530.11 | GERD | 3 | 1.98 (1.94 – 2.03) | 0.00E+00 | 130482 |
| 530.11 | GERD | 4 | 1.96 (1.91 – 2.01) | 0.00E+00 | 132711 |
| 530.11 | GERD | 5 | 1.94 (1.89 – 1.98) | 0.00E+00 | 134656 |
| 401.1 | Essential hypertension | -5 | 1.20 (1.17 – 1.23) | 7.64E-45 | 85312 |
| 401.1 | Essential hypertension | -4 | 1.22 (1.19 – 1.25) | 8.72E-61 | 90932 |
| 401.1 | Essential hypertension | -3 | 1.24 (1.21 – 1.27) | 9.65E-79 | 96622 |
| 401.1 | Essential hypertension | -2 | 1.26 (1.23 – 1.29) | 2.90E-104 | 102600 |
| 401.1 | Essential hypertension | -1 | 1.29 (1.27 – 1.32) | 1.19E-143 | 108657 |
| 401.1 | Essential hypertension | 0 | 1.35 (1.32 – 1.37) | 5.99E-231 | 117551 |
| 401.1 | Essential hypertension | 1 | 1.50 (1.48 – 1.53) | 0.00E+00 | 124868 |
| 401.1 | Essential hypertension | 2 | 1.49 (1.46 – 1.51) | 0.00E+00 | 127897 |
| 401.1 | Essential hypertension | 3 | 1.47 (1.45 – 1.49) | 0.00E+00 | 130482 |
| 401.1 | Essential hypertension | 4 | 1.45 (1.43 – 1.47) | 0.00E+00 | 132711 |
| 401.1 | Essential hypertension | 5 | 1.43 (1.41 – 1.46) | 0.00E+00 | 134656 |
| 495 | Asthma | -5 | 1.40 (1.33 – 1.47) | 8.74E-41 | 85312 |
| 495 | Asthma | -4 | 1.43 (1.36 – 1.49) | 2.03E-50 | 90932 |
| 495 | Asthma | -3 | 1.47 (1.41 – 1.54) | 1.52E-66 | 96622 |
| 495 | Asthma | -2 | 1.48 (1.42 – 1.55) | 1.10E-76 | 102600 |
| 495 | Asthma | -1 | 1.50 (1.45 – 1.57) | 2.17E-90 | 108657 |
| 495 | Asthma | 0 | 1.55 (1.49 – 1.60) | 5.54E-117 | 117551 |
| 495 | Asthma | 1 | 1.80 (1.74 – 1.86) | 1.33E-257 | 124868 |
| 495 | Asthma | 2 | 1.83 (1.77 – 1.89) | 5.29E-288 | 127897 |
| 495 | Asthma | 3 | 1.83 (1.77 – 1.89) | 1.07E-305 | 130482 |
| 495 | Asthma | 4 | 1.82 (1.77 – 1.88) | 0.00E+00 | 132711 |
| 495 | Asthma | 5 | 1.82 (1.76 – 1.87) | 0.00E+00 | 134656 |
| 379.9 | Pain, swelling or discharge of eye | -5 | 4.37 (3.47 – 5.50) | 6.98E-36 | 85312 |
| 379.9 | Pain, swelling or discharge of eye | -4 | 4.42 (3.58 – 5.46) | 6.34E-43 | 90932 |
| 379.9 | Pain, swelling or discharge of eye | -3 | 4.15 (3.43 – 5.03) | 4.05E-48 | 96622 |
| 379.9 | Pain, swelling or discharge of eye | -2 | 3.91 (3.29 – 4.65) | 9.20E-54 | 102600 |
| 379.9 | Pain, swelling or discharge of eye | -1 | 3.92 (3.33 – 4.63) | 2.80E-59 | 108657 |

| PheCode | Disease | Years to OSA diagnosis | Risk ratio | P-value | N |
|---|---|---|---|---|---|
| 379.9 | Pain, swelling or discharge of eye | 0 | 3.73 (3.20 – 4.35) | 3.09E-63 | 117551 |
| 379.9 | Pain, swelling or discharge of eye | 1 | 3.78 (3.28 – 4.35) | 4.61E-75 | 124868 |
| 379.9 | Pain, swelling or discharge of eye | 2 | 3.82 (3.34 – 4.37) | 1.61E-84 | 127897 |
| 379.9 | Pain, swelling or discharge of eye | 3 | 3.88 (3.41 – 4.42) | 5.44E-94 | 130482 |
| 379.9 | Pain, swelling or discharge of eye | 4 | 3.85 (3.41 – 4.36) | 8.39E-102 | 132711 |
| 379.9 | Pain, swelling or discharge of eye | 5 | 3.83 (3.40 – 4.31) | 9.44E-110 | 134656 |
| 173 | Neoplasm of uncertain behavior of skin | -5 | 1.83 (1.66 – 2.01) | 4.27E-35 | 85312 |
| 173 | Neoplasm of uncertain behavior of skin | -4 | 1.85 (1.69 – 2.03) | 8.78E-41 | 90932 |
| 173 | Neoplasm of uncertain behavior of skin | -3 | 1.83 (1.68 – 2.00) | 1.50E-43 | 96622 |
| 173 | Neoplasm of uncertain behavior of skin | -2 | 1.80 (1.65 – 1.95) | 1.61E-44 | 102600 |
| 173 | Neoplasm of uncertain behavior of skin | -1 | 1.73 (1.60 – 1.87) | 1.59E-43 | 108657 |
| 173 | Neoplasm of uncertain behavior of skin | 0 | 1.63 (1.52 – 1.76) | 1.82E-39 | 117551 |
| 173 | Neoplasm of uncertain behavior of skin | 1 | 1.60 (1.49 – 1.72) | 2.88E-39 | 124868 |
| 173 | Neoplasm of uncertain behavior of skin | 2 | 1.61 (1.51 – 1.72) | 1.67E-43 | 127897 |
| 173 | Neoplasm of uncertain behavior of skin | 3 | 1.62 (1.52 – 1.74) | 1.09E-46 | 130482 |
| 173 | Neoplasm of uncertain behavior of skin | 4 | 1.64 (1.53 – 1.75) | 4.78E-50 | 132711 |
| 173 | Neoplasm of uncertain behavior of skin | 5 | 1.65 (1.54 – 1.75) | 7.48E-54 | 134656 |
| 293 | Symptoms involving head and neck | -5 | 6.45 (4.79 – 8.67) | 6.92E-35 | 85312 |
| 293 | Symptoms involving head and neck | -4 | 6.43 (4.85 – 8.54) | 6.67E-38 | 90932 |
| 293 | Symptoms involving head and neck | -3 | 6.19 (4.74 – 8.08) | 8.65E-41 | 96622 |
| 293 | Symptoms involving head and neck | -2 | 5.75 (4.47 – 7.39) | 2.81E-42 | 102600 |
| 293 | Symptoms involving head and neck | -1 | 5.49 (4.33 – 6.96) | 4.91E-45 | 108657 |
| 293 | Symptoms involving head and neck | 0 | 5.06 (4.05 – 6.32) | 4.64E-46 | 117551 |
| 293 | Symptoms involving head and neck | 1 | 5.68 (4.61 – 7.00) | 4.28E-60 | 124868 |
| 293 | Symptoms involving head and neck | 2 | 5.77 (4.73 – 7.05) | 2.81E-66 | 127897 |
| 293 | Symptoms involving head and neck | 3 | 5.93 (4.88 – 7.21) | 1.92E-71 | 130482 |
| 293 | Symptoms involving head and neck | 4 | 6.13 (5.06 – 7.42) | 7.99E-77 | 132711 |
| 293 | Symptoms involving head and neck | 5 | 6.27 (5.20 – 7.57) | 1.74E-81 | 134656 |
| 197 | Chemotherapy | -5 | 0.44 (0.38 – 0.50) | 5.67E-32 | 85312 |
| 197 | Chemotherapy | -4 | 0.43 (0.38 – 0.49) | 1.84E-39 | 90932 |
| 197 | Chemotherapy | -3 | 0.43 (0.38 – 0.48) | 1.33E-47 | 96622 |
| 197 | Chemotherapy | -2 | 0.45 (0.40 – 0.50) | 1.61E-52 | 102600 |
| 197 | Chemotherapy | -1 | 0.45 (0.41 – 0.50) | 1.08E-60 | 108657 |
| 197 | Chemotherapy | 0 | 0.54 (0.50 – 0.59) | 1.86E-50 | 117551 |

| PheCode | Disease | Years to OSA diagnosis | Risk ratio | P-value | N |
|---|---|---|---|---|---|
| 197 | Chemotherapy | 1 | 0.74 (0.70 – 0.79) | 2.41E-18 | 124868 |
| 197 | Chemotherapy | 2 | 0.77 (0.73 – 0.82) | 1.13E-15 | 127897 |
| 197 | Chemotherapy | 3 | 0.78 (0.74 – 0.83) | 3.36E-15 | 130482 |
| 197 | Chemotherapy | 4 | 0.78 (0.73 – 0.82) | 3.19E-17 | 132711 |
| 197 | Chemotherapy | 5 | 0.77 (0.73 – 0.82) | 4.67E-19 | 134656 |
| 244.4 | Hypothyroidism NOS | -5 | 1.42 (1.34 – 1.50) | 9.39E-32 | 85312 |
| 244.4 | Hypothyroidism NOS | -4 | 1.44 (1.36 – 1.52) | 9.50E-39 | 90932 |
| 244.4 | Hypothyroidism NOS | -3 | 1.48 (1.40 – 1.56) | 3.12E-49 | 96622 |
| 244.4 | Hypothyroidism NOS | -2 | 1.51 (1.44 – 1.59) | 2.02E-61 | 102600 |
| 244.4 | Hypothyroidism NOS | -1 | 1.56 (1.49 – 1.63) | 2.19E-78 | 108657 |
| 244.4 | Hypothyroidism NOS | 0 | 1.59 (1.52 – 1.66) | 4.87E-97 | 117551 |
| 244.4 | Hypothyroidism NOS | 1 | 1.83 (1.76 – 1.91) | 1.45E-193 | 124868 |
| 244.4 | Hypothyroidism NOS | 2 | 1.85 (1.78 – 1.93) | 3.18E-209 | 127897 |
| 244.4 | Hypothyroidism NOS | 3 | 1.86 (1.79 – 1.93) | 1.10E-216 | 130482 |
| 244.4 | Hypothyroidism NOS | 4 | 1.85 (1.78 – 1.92) | 1.20E-222 | 132711 |
| 244.4 | Hypothyroidism NOS | 5 | 1.84 (1.78 – 1.91) | 1.80E-227 | 134656 |
| 740.1 | Osteoarthritis; localized | -5 | 0.51 (0.45 – 0.57) | 2.05E-29 | 85312 |
| 740.1 | Osteoarthritis; localized | -4 | 0.49 (0.44 – 0.55) | 2.50E-36 | 90932 |
| 740.1 | Osteoarthritis; localized | -3 | 0.51 (0.46 – 0.57) | 7.63E-38 | 96622 |
| 740.1 | Osteoarthritis; localized | -2 | 0.53 (0.48 – 0.58) | 1.34E-39 | 102600 |
| 740.1 | Osteoarthritis; localized | -1 | 0.52 (0.48 – 0.57) | 8.59E-46 | 108657 |
| 740.1 | Osteoarthritis; localized | 0 | 0.51 (0.47 – 0.55) | 3.74E-57 | 117551 |
| 740.1 | Osteoarthritis; localized | 1 | 0.58 (0.54 – 0.63) | 1.93E-45 | 124868 |
| 740.1 | Osteoarthritis; localized | 2 | 0.59 (0.55 – 0.64) | 6.38E-44 | 127897 |
| 740.1 | Osteoarthritis; localized | 3 | 0.60 (0.56 – 0.64) | 1.46E-43 | 130482 |
| 740.1 | Osteoarthritis; localized | 4 | 0.61 (0.57 – 0.66) | 2.91E-42 | 132711 |
| 740.1 | Osteoarthritis; localized | 5 | 0.62 (0.58 – 0.67) | 6.24E-41 | 134656 |
| 947 | Urticaria | -5 | 1.99 (1.76 – 2.24) | 8.33E-29 | 85312 |
| 947 | Urticaria | -4 | 2.03 (1.81 – 2.28) | 9.99E-34 | 90932 |
| 947 | Urticaria | -3 | 2.07 (1.86 – 2.31) | 9.96E-40 | 96622 |
| 947 | Urticaria | -2 | 2.05 (1.86 – 2.27) | 9.67E-44 | 102600 |
| 947 | Urticaria | -1 | 2.06 (1.87 – 2.27) | 6.95E-49 | 108657 |
| 947 | Urticaria | 0 | 2.03 (1.86 – 2.22) | 4.45E-53 | 117551 |
| 947 | Urticaria | 1 | 2.09 (1.92 – 2.27) | 2.55E-65 | 124868 |

| PheCode | Disease | Years to OSA diagnosis | Risk ratio | P-value | N |
|---|---|---|---|---|---|
| 947 | Urticaria | 2 | 2.16 (1.99 – 2.35) | 1.85E-76 | 127897 |
| 947 | Urticaria | 3 | 2.21 (2.05 – 2.39) | 3.15E-87 | 130482 |
| 947 | Urticaria | 4 | 2.22 (2.06 – 2.40) | 3.56E-94 | 132711 |
| 947 | Urticaria | 5 | 2.22 (2.06 – 2.39) | 8.50E-100 | 134656 |
| 327 | Sleep disorders | -5 | 2.42 (2.05 – 2.84) | 2.63E-26 | 85312 |
| 327 | Sleep disorders | -4 | 2.57 (2.20 – 3.00) | 4.23E-33 | 90932 |
| 327 | Sleep disorders | -3 | 2.80 (2.42 – 3.24) | 4.87E-43 | 96622 |
| 327 | Sleep disorders | -2 | 2.90 (2.53 – 3.32) | 2.60E-52 | 102600 |
| 327 | Sleep disorders | -1 | 3.35 (2.95 – 3.80) | 6.07E-78 | 108657 |
| 327 | Sleep disorders | 0 | 5.14 (4.60 – 5.74) | 1.25E-186 | 117551 |
| 327 | Sleep disorders | 1 | 6.62 (5.99 – 7.31) | 1.79E-300 | 124868 |
| 327 | Sleep disorders | 2 | 6.85 (6.23 – 7.54) | 0.00E+00 | 127897 |
| 327 | Sleep disorders | 3 | 7.02 (6.39 – 7.70) | 0.00E+00 | 130482 |
| 327 | Sleep disorders | 4 | 6.97 (6.37 – 7.63) | 0.00E+00 | 132711 |
| 327 | Sleep disorders | 5 | 7.08 (6.48 – 7.74) | 0.00E+00 | 134656 |
| 470 | Septal Deviations/Turbinate Hypertrophy | -5 | 1.98 (1.75 – 2.25) | 3.08E-26 | 85312 |
| 470 | Septal Deviations/Turbinate Hypertrophy | -4 | 2.00 (1.78 – 2.24) | 3.08E-31 | 90932 |
| 470 | Septal Deviations/Turbinate Hypertrophy | -3 | 2.07 (1.86 – 2.31) | 5.19E-40 | 96622 |
| 470 | Septal Deviations/Turbinate Hypertrophy | -2 | 2.18 (1.97 – 2.41) | 4.06E-52 | 102600 |
| 470 | Septal Deviations/Turbinate Hypertrophy | -1 | 2.27 (2.07 – 2.49) | 1.26E-67 | 108657 |
| 470 | Septal Deviations/Turbinate Hypertrophy | 0 | 2.88 (2.66 – 3.13) | 6.77E-142 | 117551 |
| 470 | Septal Deviations/Turbinate Hypertrophy | 1 | 5.45 (5.09 – 5.84) | 0.00E+00 | 124868 |
| 470 | Septal Deviations/Turbinate Hypertrophy | 2 | 5.43 (5.09 – 5.80) | 0.00E+00 | 127897 |
| 470 | Septal Deviations/Turbinate Hypertrophy | 3 | 5.40 (5.07 – 5.75) | 0.00E+00 | 130482 |
| 470 | Septal Deviations/Turbinate Hypertrophy | 4 | 5.29 (4.98 – 5.63) | 0.00E+00 | 132711 |
| 470 | Septal Deviations/Turbinate Hypertrophy | 5 | 5.17 (4.87 – 5.49) | 0.00E+00 | 134656 |
| 340 | Migraine | -5 | 1.47 (1.36 – 1.58) | 3.35E-25 | 85312 |
| 340 | Migraine | -4 | 1.45 (1.35 – 1.55) | 4.70E-26 | 90932 |
| 340 | Migraine | -3 | 1.49 (1.39 – 1.59) | 7.00E-33 | 96622 |
| 340 | Migraine | -2 | 1.52 (1.43 – 1.62) | 4.20E-41 | 102600 |
| 340 | Migraine | -1 | 1.58 (1.49 – 1.67) | 2.74E-54 | 108657 |
| 340 | Migraine | 0 | 1.59 (1.51 – 1.68) | 1.14E-64 | 117551 |
| 340 | Migraine | 1 | 1.75 (1.66 – 1.84) | 1.32E-108 | 124868 |
| 340 | Migraine | 2 | 1.78 (1.69 – 1.86) | 6.59E-123 | 127897 |

| PheCode | Disease | Years to OSA diagnosis | Risk ratio | P-value | N |
|---|---|---|---|---|---|
| 340 | Migraine | 3 | 1.77 (1.69 – 1.86) | 2.94E-131 | 130482 |
| 340 | Migraine | 4 | 1.77 (1.70 – 1.86) | 4.76E-140 | 132711 |
| 340 | Migraine | 5 | 1.77 (1.70 – 1.85) | 1.23E-145 | 134656 |
| 296.22 | Major depressive disorder | -5 | 1.48 (1.37 – 1.59) | 4.03E-25 | 85312 |
| 296.22 | Major depressive disorder | -4 | 1.52 (1.42 – 1.63) | 4.09E-33 | 90932 |
| 296.22 | Major depressive disorder | -3 | 1.57 (1.48 – 1.68) | 1.32E-44 | 96622 |
| 296.22 | Major depressive disorder | -2 | 1.62 (1.53 – 1.71) | 2.82E-59 | 102600 |
| 296.22 | Major depressive disorder | -1 | 1.70 (1.62 – 1.79) | 3.47E-89 | 108657 |
| 296.22 | Major depressive disorder | 0 | 1.88 (1.79 – 1.97) | 1.06E-158 | 117551 |
| 296.22 | Major depressive disorder | 1 | 2.18 (2.09 – 2.27) | 0.00E+00 | 124868 |
| 296.22 | Major depressive disorder | 2 | 2.21 (2.13 – 2.29) | 0.00E+00 | 127897 |
| 296.22 | Major depressive disorder | 3 | 2.20 (2.12 – 2.28) | 0.00E+00 | 130482 |
| 296.22 | Major depressive disorder | 4 | 2.18 (2.10 – 2.25) | 0.00E+00 | 132711 |
| 296.22 | Major depressive disorder | 5 | 2.16 (2.09 – 2.23) | 0.00E+00 | 134656 |
| 327.4 | Insomnia | -5 | 1.70 (1.53 – 1.89) | 3.23E-22 | 85312 |
| 327.4 | Insomnia | -4 | 1.77 (1.60 – 1.95) | 1.17E-30 | 90932 |
| 327.4 | Insomnia | -3 | 1.82 (1.66 – 1.99) | 8.78E-40 | 96622 |
| 327.4 | Insomnia | -2 | 1.84 (1.70 – 2.00) | 7.72E-49 | 102600 |
| 327.4 | Insomnia | -1 | 1.94 (1.81 – 2.09) | 5.45E-70 | 108657 |
| 327.4 | Insomnia | 0 | 2.30 (2.16 – 2.45) | 5.44E-145 | 117551 |
| 327.4 | Insomnia | 1 | 3.17 (3.01 – 3.35) | 0.00E+00 | 124868 |
| 327.4 | Insomnia | 2 | 3.17 (3.01 – 3.34) | 0.00E+00 | 127897 |
| 327.4 | Insomnia | 3 | 3.08 (2.93 – 3.24) | 0.00E+00 | 130482 |
| 327.4 | Insomnia | 4 | 3.00 (2.86 – 3.15) | 0.00E+00 | 132711 |
| 327.4 | Insomnia | 5 | 2.93 (2.80 – 3.06) | 0.00E+00 | 134656 |
| 508 | Pulmonary collapse; interstitial and compensatory emphysema | -5 | 0.64 (0.58 – 0.70) | 3.24E-22 | 85312 |
| 508 | Pulmonary collapse; interstitial and compensatory emphysema | -4 | 0.64 (0.59 – 0.70) | 1.32E-24 | 90932 |
| 508 | Pulmonary collapse; interstitial and compensatory emphysema | -3 | 0.64 (0.59 – 0.69) | 1.68E-29 | 96622 |
| 508 | Pulmonary collapse; interstitial and compensatory emphysema | -2 | 0.64 (0.59 – 0.69) | 1.85E-33 | 102600 |
| 508 | Pulmonary collapse; interstitial and compensatory emphysema | -1 | 0.65 (0.61 – 0.70) | 1.16E-35 | 108657 |

| PheCode | Disease | Years to OSA diagnosis | Risk ratio | P-value | N |
|---|---|---|---|---|---|
| 508 | Pulmonary collapse; interstitial and compensatory emphysema | 0 | 0.76 (0.72 – 0.80) | 3.55E-20 | 117551 |
| 508 | Pulmonary collapse; interstitial and compensatory emphysema | 1 | 1.11 (1.06 – 1.17) | 1.98E-05 | 124868 |
| 508 | Pulmonary collapse; interstitial and compensatory emphysema | 2 | 1.12 (1.07 – 1.18) | 2.45E-06 | 127897 |
| 508 | Pulmonary collapse; interstitial and compensatory emphysema | 3 | 1.11 (1.06 – 1.16) | 1.51E-05 | 130482 |
| 508 | Pulmonary collapse; interstitial and compensatory emphysema | 4 | 1.10 (1.05 – 1.16) | 3.87E-05 | 132711 |
| 508 | Pulmonary collapse; interstitial and compensatory emphysema | 5 | 1.10 (1.05 – 1.15) | 6.57E-05 | 134656 |
| 272.11 | Hypercholesterolemia | -5 | 1.24 (1.19 – 1.30) | 2.54E-21 | 85312 |
| 272.11 | Hypercholesterolemia | -4 | 1.27 (1.22 – 1.33) | 6.18E-29 | 90932 |
| 272.11 | Hypercholesterolemia | -3 | 1.28 (1.23 – 1.34) | 2.91E-35 | 96622 |
| 272.11 | Hypercholesterolemia | -2 | 1.30 (1.25 – 1.34) | 2.35E-42 | 102600 |
| 272.11 | Hypercholesterolemia | -1 | 1.31 (1.27 – 1.36) | 3.43E-53 | 108657 |
| 272.11 | Hypercholesterolemia | 0 | 1.32 (1.28 – 1.36) | 3.00E-62 | 117551 |
| 272.11 | Hypercholesterolemia | 1 | 1.52 (1.47 – 1.56) | 2.08E-168 | 124868 |
| 272.11 | Hypercholesterolemia | 2 | 1.54 (1.50 – 1.59) | 2.01E-192 | 127897 |
| 272.11 | Hypercholesterolemia | 3 | 1.55 (1.51 – 1.59) | 7.99E-207 | 130482 |
| 272.11 | Hypercholesterolemia | 4 | 1.54 (1.50 – 1.58) | 7.52E-213 | 132711 |
| 272.11 | Hypercholesterolemia | 5 | 1.54 (1.50 – 1.58) | 2.03E-222 | 134656 |
| 389.2 | Conductive hearing loss | -5 | 4.59 (3.30 – 6.39) | 1.36E-19 | 85312 |
| 389.2 | Conductive hearing loss | -4 | 4.50 (3.34 – 6.06) | 3.59E-23 | 90932 |
| 389.2 | Conductive hearing loss | -3 | 4.57 (3.49 – 5.99) | 4.38E-28 | 96622 |
| 389.2 | Conductive hearing loss | -2 | 4.64 (3.60 – 5.99) | 3.07E-32 | 102600 |
| 389.2 | Conductive hearing loss | -1 | 4.41 (3.51 – 5.56) | 1.36E-36 | 108657 |
| 389.2 | Conductive hearing loss | 0 | 4.35 (3.53 – 5.37) | 6.60E-43 | 117551 |
| 389.2 | Conductive hearing loss | 1 | 4.76 (3.93 – 5.76) | 6.45E-58 | 124868 |
| 389.2 | Conductive hearing loss | 2 | 4.59 (3.84 – 5.48) | 1.23E-63 | 127897 |
| 389.2 | Conductive hearing loss | 3 | 4.64 (3.92 – 5.49) | 1.04E-70 | 130482 |
| 389.2 | Conductive hearing loss | 4 | 4.65 (3.95 – 5.48) | 4.74E-76 | 132711 |
| 389.2 | Conductive hearing loss | 5 | 4.49 (3.84 – 5.25) | 1.93E-78 | 134656 |
| 503 | Pulmonary congestion and hypostasis | -5 | 0.51 (0.44 – 0.59) | 4.10E-18 | 85312 |

| PheCode | Disease | Years to OSA diagnosis | Risk ratio | P-value | N |
|---|---|---|---|---|---|
| 503 | Pulmonary congestion and hypostasis | -4 | 0.53 (0.46 – 0.61) | 9.62E-20 | 90932 |
| 503 | Pulmonary congestion and hypostasis | -3 | 0.52 (0.46 – 0.59) | 3.27E-24 | 96622 |
| 503 | Pulmonary congestion and hypostasis | -2 | 0.55 (0.49 – 0.62) | 5.80E-25 | 102600 |
| 503 | Pulmonary congestion and hypostasis | -1 | 0.57 (0.52 – 0.63) | 2.96E-26 | 108657 |
| 503 | Pulmonary congestion and hypostasis | 0 | 0.72 (0.66 – 0.78) | 8.97E-14 | 117551 |
| 503 | Pulmonary congestion and hypostasis | 1 | 1.16 (1.08 – 1.24) | 2.80E-05 | 124868 |
| 503 | Pulmonary congestion and hypostasis | 2 | 1.17 (1.09 – 1.25) | 5.95E-06 | 127897 |
| 503 | Pulmonary congestion and hypostasis | 3 | 1.16 (1.09 – 1.24) | 1.09E-05 | 130482 |
| 503 | Pulmonary congestion and hypostasis | 4 | 1.16 (1.08 – 1.23) | 9.56E-06 | 132711 |
| 503 | Pulmonary congestion and hypostasis | 5 | 1.14 (1.07 – 1.21) | 4.25E-05 | 134656 |
| 300.1 | Anxiety disorder | -5 | 1.28 (1.21 – 1.35) | 1.39E-17 | 85312 |
| 300.1 | Anxiety disorder | -4 | 1.31 (1.24 – 1.38) | 4.13E-24 | 90932 |
| 300.1 | Anxiety disorder | -3 | 1.36 (1.29 – 1.43) | 5.42E-36 | 96622 |
| 300.1 | Anxiety disorder | -2 | 1.40 (1.34 – 1.46) | 4.30E-49 | 102600 |
| 300.1 | Anxiety disorder | -1 | 1.46 (1.40 – 1.52) | 2.53E-72 | 108657 |
| 300.1 | Anxiety disorder | 0 | 1.53 (1.47 – 1.59) | 3.77E-110 | 117551 |
| 300.1 | Anxiety disorder | 1 | 1.79 (1.74 – 1.85) | 3.27E-265 | 124868 |
| 300.1 | Anxiety disorder | 2 | 1.80 (1.74 – 1.86) | 3.55E-294 | 127897 |
| 300.1 | Anxiety disorder | 3 | 1.79 (1.74 – 1.85) | 0.00E+00 | 130482 |
| 300.1 | Anxiety disorder | 4 | 1.77 (1.72 – 1.82) | 0.00E+00 | 132711 |
| 300.1 | Anxiety disorder | 5 | 1.74 (1.69 – 1.79) | 0.00E+00 | 134656 |
| 706.8 | Other specified diseases of sebaceous glands | -5 | 1.65 (1.47 – 1.85) | 3.25E-17 | 85312 |
| 706.8 | Other specified diseases of sebaceous glands | -4 | 1.60 (1.44 – 1.79) | 1.86E-17 | 90932 |
| 706.8 | Other specified diseases of sebaceous glands | -3 | 1.63 (1.48 – 1.81) | 4.58E-21 | 96622 |
| 706.8 | Other specified diseases of sebaceous glands | -2 | 1.63 (1.48 – 1.79) | 4.17E-23 | 102600 |
| 706.8 | Other specified diseases of sebaceous glands | -1 | 1.61 (1.47 – 1.76) | 1.45E-24 | 108657 |
| 706.8 | Other specified diseases of sebaceous glands | 0 | 1.53 (1.40 – 1.67) | 2.08E-22 | 117551 |
| 706.8 | Other specified diseases of sebaceous glands | 1 | 1.55 (1.43 – 1.68) | 1.83E-26 | 124868 |
| 706.8 | Other specified diseases of sebaceous glands | 2 | 1.62 (1.50 – 1.75) | 4.75E-33 | 127897 |
| 706.8 | Other specified diseases of sebaceous glands | 3 | 1.65 (1.53 – 1.78) | 5.03E-37 | 130482 |
| 706.8 | Other specified diseases of sebaceous glands | 4 | 1.67 (1.55 – 1.80) | 1.13E-40 | 132711 |
| 706.8 | Other specified diseases of sebaceous glands | 5 | 1.66 (1.54 – 1.78) | 5.40E-42 | 134656 |
| 313.1 | Attention deficit hyperactivity disorder | -5 | 1.87 (1.62 – 2.17) | 6.97E-17 | 85312 |
| 313.1 | Attention deficit hyperactivity disorder | -4 | 1.91 (1.66 – 2.18) | 1.55E-20 | 90932 |

| PheCode | Disease | Years to OSA diagnosis | Risk ratio | P-value | N |
|---|---|---|---|---|---|
| 313.1 | Attention deficit hyperactivity disorder | -3 | 1.96 (1.73 – 2.22) | 6.52E-26 | 96622 |
| 313.1 | Attention deficit hyperactivity disorder | -2 | 2.10 (1.87 – 2.35) | 1.54E-36 | 102600 |
| 313.1 | Attention deficit hyperactivity disorder | -1 | 2.17 (1.95 – 2.41) | 1.87E-47 | 108657 |
| 313.1 | Attention deficit hyperactivity disorder | 0 | 2.25 (2.04 – 2.48) | 1.06E-60 | 117551 |
| 313.1 | Attention deficit hyperactivity disorder | 1 | 2.53 (2.33 – 2.75) | 1.00E-108 | 124868 |
| 313.1 | Attention deficit hyperactivity disorder | 2 | 2.61 (2.41 – 2.82) | 1.24E-126 | 127897 |
| 313.1 | Attention deficit hyperactivity disorder | 3 | 2.59 (2.40 – 2.79) | 1.71E-137 | 130482 |
| 313.1 | Attention deficit hyperactivity disorder | 4 | 2.59 (2.40 – 2.78) | 7.17E-146 | 132711 |
| 313.1 | Attention deficit hyperactivity disorder | 5 | 2.56 (2.39 – 2.75) | 5.09E-152 | 134656 |
| 272.13 | Mixed hyperlipidemia | -5 | 1.66 (1.47 – 1.88) | 1.88E-16 | 85312 |
| 272.13 | Mixed hyperlipidemia | -4 | 1.61 (1.44 – 1.80) | 1.98E-17 | 90932 |
| 272.13 | Mixed hyperlipidemia | -3 | 1.53 (1.39 – 1.69) | 4.21E-17 | 96622 |
| 272.13 | Mixed hyperlipidemia | -2 | 1.49 (1.37 – 1.63) | 5.56E-19 | 102600 |
| 272.13 | Mixed hyperlipidemia | -1 | 1.43 (1.32 – 1.55) | 3.18E-19 | 108657 |
| 272.13 | Mixed hyperlipidemia | 0 | 1.48 (1.38 – 1.58) | 1.82E-29 | 117551 |
| 272.13 | Mixed hyperlipidemia | 1 | 1.75 (1.65 – 1.85) | 1.87E-76 | 124868 |
| 272.13 | Mixed hyperlipidemia | 2 | 1.75 (1.66 – 1.85) | 8.79E-88 | 127897 |
| 272.13 | Mixed hyperlipidemia | 3 | 1.76 (1.67 – 1.85) | 1.12E-98 | 130482 |
| 272.13 | Mixed hyperlipidemia | 4 | 1.73 (1.64 – 1.81) | 9.18E-101 | 132711 |
| 272.13 | Mixed hyperlipidemia | 5 | 1.70 (1.62 – 1.79) | 1.88E-104 | 134656 |
| 946 | Anaphylactic shock NOS | -5 | 2.19 (1.81 – 2.65) | 8.38E-16 | 85312 |
| 946 | Anaphylactic shock NOS | -4 | 2.05 (1.71 – 2.45) | 6.88E-15 | 90932 |
| 946 | Anaphylactic shock NOS | -3 | 2.18 (1.84 – 2.58) | 1.72E-19 | 96622 |
| 946 | Anaphylactic shock NOS | -2 | 2.01 (1.71 – 2.37) | 1.76E-17 | 102600 |
| 946 | Anaphylactic shock NOS | -1 | 1.93 (1.66 – 2.25) | 3.34E-17 | 108657 |
| 946 | Anaphylactic shock NOS | 0 | 1.98 (1.71 – 2.29) | 4.60E-20 | 117551 |
| 946 | Anaphylactic shock NOS | 1 | 2.05 (1.78 – 2.35) | 2.43E-24 | 124868 |
| 946 | Anaphylactic shock NOS | 2 | 2.14 (1.87 – 2.45) | 1.07E-28 | 127897 |
| 946 | Anaphylactic shock NOS | 3 | 2.18 (1.92 – 2.49) | 1.22E-31 | 130482 |
| 946 | Anaphylactic shock NOS | 4 | 2.18 (1.92 – 2.47) | 2.79E-33 | 132711 |
| 946 | Anaphylactic shock NOS | 5 | 2.20 (1.95 – 2.49) | 7.39E-36 | 134656 |
| 510 | Other diseases of lung | -5 | 0.73 (0.67 – 0.79) | 1.06E-15 | 85312 |
| 510 | Other diseases of lung | -4 | 0.74 (0.68 – 0.79) | 4.46E-16 | 90932 |
| 510 | Other diseases of lung | -3 | 0.76 (0.71 – 0.82) | 2.09E-14 | 96622 |

| PheCode | Disease | Years to OSA diagnosis | Risk ratio | P-value | N |
|---|---|---|---|---|---|
| 510 | Other diseases of lung | -2 | 0.77 (0.72 – 0.82) | 9.78E-15 | 102600 |
| 510 | Other diseases of lung | -1 | 0.78 (0.73 – 0.84) | 8.67E-14 | 108657 |
| 510 | Other diseases of lung | 0 | 0.83 (0.78 – 0.88) | 1.11E-09 | 117551 |
| 510 | Other diseases of lung | 1 | 0.97 (0.92 – 1.02) | 2.34E-01 | 124868 |
| 510 | Other diseases of lung | 2 | 1.01 (0.96 – 1.07) | 7.50E-01 | 127897 |
| 510 | Other diseases of lung | 3 | 1.02 (0.97 – 1.08) | 3.85E-01 | 130482 |
| 510 | Other diseases of lung | 4 | 1.04 (0.98 – 1.10) | 1.75E-01 | 132711 |
| 510 | Other diseases of lung | 5 | 1.06 (1.00 – 1.11) | 4.68E-02 | 134656 |
| 289.9 | Abnormality of red blood cells | -5 | 5.51 (3.62 – 8.39) | 1.77E-15 | 85312 |
| 289.9 | Abnormality of red blood cells | -4 | 6.62 (4.45 – 9.83) | 8.08E-21 | 90932 |
| 289.9 | Abnormality of red blood cells | -3 | 5.93 (4.18 – 8.41) | 1.58E-23 | 96622 |
| 289.9 | Abnormality of red blood cells | -2 | 6.51 (4.73 – 8.96) | 1.14E-30 | 102600 |
| 289.9 | Abnormality of red blood cells | -1 | 6.48 (4.85 – 8.65) | 1.05E-36 | 108657 |
| 289.9 | Abnormality of red blood cells | 0 | 6.60 (5.09 – 8.55) | 3.76E-46 | 117551 |
| 289.9 | Abnormality of red blood cells | 1 | 7.71 (6.09 – 9.76) | 1.19E-64 | 124868 |
| 289.9 | Abnormality of red blood cells | 2 | 7.89 (6.32 – 9.85) | 1.81E-74 | 127897 |
| 289.9 | Abnormality of red blood cells | 3 | 8.62 (6.97 – 10.67) | 3.59E-87 | 130482 |
| 289.9 | Abnormality of red blood cells | 4 | 8.71 (7.11 – 10.69) | 4.33E-96 | 132711 |
| 289.9 | Abnormality of red blood cells | 5 | 8.79 (7.23 – 10.68) | 9.47E-106 | 134656 |
| 689 | Skin/subcutaneous tissue disorder NOS | -5 | 1.40 (1.29 – 1.52) | 2.70E-15 | 85312 |
| 689 | Skin/subcutaneous tissue disorder NOS | -4 | 1.42 (1.32 – 1.54) | 4.15E-19 | 90932 |
| 689 | Skin/subcutaneous tissue disorder NOS | -3 | 1.44 (1.34 – 1.55) | 8.31E-23 | 96622 |
| 689 | Skin/subcutaneous tissue disorder NOS | -2 | 1.47 (1.37 – 1.57) | 3.79E-29 | 102600 |
| 689 | Skin/subcutaneous tissue disorder NOS | -1 | 1.54 (1.44 – 1.64) | 3.06E-41 | 108657 |
| 689 | Skin/subcutaneous tissue disorder NOS | 0 | 1.56 (1.47 – 1.65) | 5.11E-50 | 117551 |
| 689 | Skin/subcutaneous tissue disorder NOS | 1 | 1.67 (1.58 – 1.77) | 7.56E-75 | 124868 |
| 689 | Skin/subcutaneous tissue disorder NOS | 2 | 1.78 (1.69 – 1.88) | 3.55E-101 | 127897 |
| 689 | Skin/subcutaneous tissue disorder NOS | 3 | 1.84 (1.75 – 1.94) | 1.06E-121 | 130482 |
| 689 | Skin/subcutaneous tissue disorder NOS | 4 | 1.88 (1.79 – 1.98) | 7.88E-140 | 132711 |
| 689 | Skin/subcutaneous tissue disorder NOS | 5 | 1.91 (1.82 – 2.00) | 1.61E-156 | 134656 |
| 701.2 | Scar conditions and fibrosis of skin | -5 | 1.48 (1.34 – 1.63) | 3.26E-15 | 85312 |
| 701.2 | Scar conditions and fibrosis of skin | -4 | 1.50 (1.37 – 1.65) | 1.76E-18 | 90932 |
| 701.2 | Scar conditions and fibrosis of skin | -3 | 1.47 (1.35 – 1.60) | 1.56E-18 | 96622 |
| 701.2 | Scar conditions and fibrosis of skin | -2 | 1.48 (1.37 – 1.61) | 2.21E-21 | 102600 |

| PheCode | Disease | Years to OSA diagnosis | Risk ratio | P-value | N |
|---|---|---|---|---|---|
| 701.2 | Scar conditions and fibrosis of skin | -1 | 1.49 (1.38 – 1.61) | 3.76E-24 | 108657 |
| 701.2 | Scar conditions and fibrosis of skin | 0 | 1.39 (1.29 – 1.50) | 9.46E-19 | 117551 |
| 701.2 | Scar conditions and fibrosis of skin | 1 | 1.40 (1.31 – 1.50) | 3.07E-21 | 124868 |
| 701.2 | Scar conditions and fibrosis of skin | 2 | 1.45 (1.35 – 1.55) | 8.29E-27 | 127897 |
| 701.2 | Scar conditions and fibrosis of skin | 3 | 1.49 (1.39 – 1.59) | 4.42E-32 | 130482 |
| 701.2 | Scar conditions and fibrosis of skin | 4 | 1.52 (1.43 – 1.62) | 3.81E-38 | 132711 |
| 701.2 | Scar conditions and fibrosis of skin | 5 | 1.53 (1.44 – 1.63) | 1.53E-41 | 134656 |
| 585.1 | Acute renal failure | -5 | 0.49 (0.41 – 0.59) | 3.84E-15 | 85312 |
| 585.1 | Acute renal failure | -4 | 0.51 (0.44 – 0.60) | 9.93E-17 | 90932 |
| 585.1 | Acute renal failure | -3 | 0.56 (0.49 – 0.64) | 6.23E-17 | 96622 |
| 585.1 | Acute renal failure | -2 | 0.60 (0.53 – 0.68) | 5.77E-17 | 102600 |
| 585.1 | Acute renal failure | -1 | 0.66 (0.60 – 0.74) | 1.00E-14 | 108657 |
| 585.1 | Acute renal failure | 0 | 0.86 (0.79 – 0.94) | 6.08E-04 | 117551 |
| 585.1 | Acute renal failure | 1 | 1.25 (1.17 – 1.34) | 2.79E-10 | 124868 |
| 585.1 | Acute renal failure | 2 | 1.26 (1.18 – 1.35) | 1.21E-11 | 127897 |
| 585.1 | Acute renal failure | 3 | 1.24 (1.16 – 1.33) | 1.07E-10 | 130482 |
| 585.1 | Acute renal failure | 4 | 1.22 (1.14 – 1.30) | 1.46E-09 | 132711 |
| 585.1 | Acute renal failure | 5 | 1.19 (1.12 – 1.26) | 5.87E-08 | 134656 |
| 300.4 | Dysthymic disorder | -5 | 1.54 (1.38 – 1.72) | 1.07E-14 | 85312 |
| 300.4 | Dysthymic disorder | -4 | 1.54 (1.39 – 1.71) | 8.56E-16 | 90932 |
| 300.4 | Dysthymic disorder | -3 | 1.60 (1.45 – 1.78) | 1.22E-19 | 96622 |
| 300.4 | Dysthymic disorder | -2 | 1.64 (1.49 – 1.81) | 2.83E-23 | 102600 |
| 300.4 | Dysthymic disorder | -1 | 1.72 (1.57 – 1.90) | 7.59E-30 | 108657 |
| 300.4 | Dysthymic disorder | 0 | 1.71 (1.56 – 1.87) | 2.25E-31 | 117551 |
| 300.4 | Dysthymic disorder | 1 | 1.90 (1.74 – 2.07) | 3.29E-48 | 124868 |
| 300.4 | Dysthymic disorder | 2 | 1.93 (1.78 – 2.10) | 4.42E-52 | 127897 |
| 300.4 | Dysthymic disorder | 3 | 1.96 (1.81 – 2.13) | 1.97E-57 | 130482 |
| 300.4 | Dysthymic disorder | 4 | 1.99 (1.84 – 2.15) | 9.25E-65 | 132711 |
| 300.4 | Dysthymic disorder | 5 | 2.00 (1.85 – 2.16) | 1.87E-69 | 134656 |
| 367.1 | Myopia | -5 | 4.34 (2.99 – 6.30) | 1.15E-14 | 85312 |
| 367.1 | Myopia | -4 | 3.62 (2.59 – 5.04) | 3.18E-14 | 90932 |
| 367.1 | Myopia | -3 | 2.98 (2.26 – 3.93) | 1.37E-14 | 96622 |
| 367.1 | Myopia | -2 | 2.73 (2.15 – 3.46) | 1.84E-16 | 102600 |
| 367.1 | Myopia | -1 | 2.54 (2.05 – 3.15) | 1.08E-17 | 108657 |

| PheCode | Disease | Years to OSA diagnosis | Risk ratio | P-value | N |
|---|---|---|---|---|---|
| 367.1 | Myopia | 0 | 2.50 (2.07 – 3.02) | 2.40E-21 | 117551 |
| 367.1 | Myopia | 1 | 2.60 (2.19 – 3.07) | 7.45E-29 | 124868 |
| 367.1 | Myopia | 2 | 2.74 (2.34 – 3.21) | 1.98E-36 | 127897 |
| 367.1 | Myopia | 3 | 2.81 (2.42 – 3.27) | 1.72E-41 | 130482 |
| 367.1 | Myopia | 4 | 2.83 (2.45 – 3.26) | 1.02E-45 | 132711 |
| 367.1 | Myopia | 5 | 2.93 (2.55 – 3.36) | 6.93E-52 | 134656 |
| 427.21 | Atrial fibrillation | -5 | 1.35 (1.25 – 1.46) | 1.43E-14 | 85312 |
| 427.21 | Atrial fibrillation | -4 | 1.37 (1.28 – 1.47) | 1.85E-18 | 90932 |
| 427.21 | Atrial fibrillation | -3 | 1.45 (1.36 – 1.55) | 6.13E-30 | 96622 |
| 427.21 | Atrial fibrillation | -2 | 1.51 (1.43 – 1.61) | 6.70E-43 | 102600 |
| 427.21 | Atrial fibrillation | -1 | 1.54 (1.46 – 1.63) | 3.47E-55 | 108657 |
| 427.21 | Atrial fibrillation | 0 | 1.87 (1.78 – 1.96) | 1.16E-147 | 117551 |
| 427.21 | Atrial fibrillation | 1 | 2.13 (2.04 – 2.23) | 6.55E-262 | 124868 |
| 427.21 | Atrial fibrillation | 2 | 2.08 (1.99 – 2.17) | 3.36E-263 | 127897 |
| 427.21 | Atrial fibrillation | 3 | 2.03 (1.95 – 2.11) | 2.42E-259 | 130482 |
| 427.21 | Atrial fibrillation | 4 | 1.98 (1.91 – 2.06) | 1.25E-255 | 132711 |
| 427.21 | Atrial fibrillation | 5 | 1.94 (1.87 – 2.01) | 1.08E-249 | 134656 |
| 476 | Allergic rhinitis | -5 | 1.23 (1.17 – 1.30) | 1.66E-14 | 85312 |
| 476 | Allergic rhinitis | -4 | 1.24 (1.18 – 1.31) | 1.07E-17 | 90932 |
| 476 | Allergic rhinitis | -3 | 1.29 (1.23 – 1.35) | 4.32E-26 | 96622 |
| 476 | Allergic rhinitis | -2 | 1.33 (1.27 – 1.39) | 9.48E-34 | 102600 |
| 476 | Allergic rhinitis | -1 | 1.37 (1.31 – 1.43) | 2.00E-45 | 108657 |
| 476 | Allergic rhinitis | 0 | 1.41 (1.35 – 1.47) | 3.14E-62 | 117551 |
| 476 | Allergic rhinitis | 1 | 1.76 (1.70 – 1.82) | 6.63E-213 | 124868 |
| 476 | Allergic rhinitis | 2 | 1.83 (1.77 – 1.89) | 2.09E-258 | 127897 |
| 476 | Allergic rhinitis | 3 | 1.87 (1.81 – 1.93) | 5.51E-296 | 130482 |
| 476 | Allergic rhinitis | 4 | 1.89 (1.83 – 1.96) | 0.00E+00 | 132711 |
| 476 | Allergic rhinitis | 5 | 1.91 (1.85 – 1.97) | 0.00E+00 | 134656 |
| 509.1 | Respiratory failure | -5 | 0.37 (0.29 – 0.48) | 1.76E-14 | 85312 |
| 509.1 | Respiratory failure | -4 | 0.42 (0.33 – 0.52) | 2.27E-15 | 90932 |
| 509.1 | Respiratory failure | -3 | 0.41 (0.34 – 0.50) | 6.84E-19 | 96622 |
| 509.1 | Respiratory failure | -2 | 0.43 (0.36 – 0.51) | 6.57E-22 | 102600 |
| 509.1 | Respiratory failure | -1 | 0.48 (0.41 – 0.56) | 4.59E-22 | 108657 |
| 509.1 | Respiratory failure | 0 | 0.82 (0.74 – 0.92) | 4.56E-04 | 117551 |

| PheCode | Disease | Years to OSA diagnosis | Risk ratio | P-value | N |
|---|---|---|---|---|---|
| 509.1 | Respiratory failure | 1 | 1.25 (1.14 – 1.36) | 7.56E-07 | 124868 |
| 509.1 | Respiratory failure | 2 | 1.24 (1.14 – 1.35) | 2.81E-07 | 127897 |
| 509.1 | Respiratory failure | 3 | 1.25 (1.16 – 1.35) | 2.53E-08 | 130482 |
| 509.1 | Respiratory failure | 4 | 1.23 (1.14 – 1.32) | 1.29E-07 | 132711 |
| 509.1 | Respiratory failure | 5 | 1.20 (1.11 – 1.29) | 1.36E-06 | 134656 |
| 196 | Radiotherapy | -5 | 0.55 (0.47 – 0.64) | 1.89E-14 | 85312 |
| 196 | Radiotherapy | -4 | 0.55 (0.49 – 0.63) | 6.27E-18 | 90932 |
| 196 | Radiotherapy | -3 | 0.53 (0.47 – 0.59) | 1.14E-25 | 96622 |
| 196 | Radiotherapy | -2 | 0.54 (0.49 – 0.60) | 8.25E-30 | 102600 |
| 196 | Radiotherapy | -1 | 0.56 (0.51 – 0.62) | 2.00E-32 | 108657 |
| 196 | Radiotherapy | 0 | 0.61 (0.57 – 0.67) | 4.44E-31 | 117551 |
| 196 | Radiotherapy | 1 | 0.74 (0.69 – 0.79) | 8.88E-17 | 124868 |
| 196 | Radiotherapy | 2 | 0.75 (0.70 – 0.80) | 1.01E-17 | 127897 |
| 196 | Radiotherapy | 3 | 0.75 (0.70 – 0.80) | 8.82E-19 | 130482 |
| 196 | Radiotherapy | 4 | 0.75 (0.70 – 0.80) | 2.69E-20 | 132711 |
| 196 | Radiotherapy | 5 | 0.74 (0.70 – 0.78) | 1.05E-23 | 134656 |
| 745 | Pain in joint | -5 | 1.14 (1.10 – 1.17) | 2.30E-14 | 85312 |
| 745 | Pain in joint | -4 | 1.15 (1.11 – 1.18) | 4.53E-18 | 90932 |
| 745 | Pain in joint | -3 | 1.15 (1.12 – 1.19) | 7.45E-22 | 96622 |
| 745 | Pain in joint | -2 | 1.16 (1.13 – 1.19) | 3.24E-25 | 102600 |
| 745 | Pain in joint | -1 | 1.16 (1.13 – 1.19) | 5.05E-30 | 108657 |
| 745 | Pain in joint | 0 | 1.16 (1.13 – 1.19) | 3.80E-33 | 117551 |
| 745 | Pain in joint | 1 | 1.21 (1.19 – 1.24) | 4.79E-62 | 124868 |
| 745 | Pain in joint | 2 | 1.25 (1.22 – 1.28) | 3.28E-87 | 127897 |
| 745 | Pain in joint | 3 | 1.26 (1.24 – 1.29) | 5.05E-103 | 130482 |
| 745 | Pain in joint | 4 | 1.27 (1.24 – 1.29) | 1.82E-112 | 132711 |
| 745 | Pain in joint | 5 | 1.27 (1.24 – 1.30) | 8.12E-121 | 134656 |
| 366.2 | Senile cataract | -5 | 1.59 (1.41 – 1.80) | 2.81E-14 | 85312 |
| 366.2 | Senile cataract | -4 | 1.64 (1.48 – 1.83) | 6.91E-20 | 90932 |
| 366.2 | Senile cataract | -3 | 1.59 (1.44 – 1.75) | 2.17E-20 | 96622 |
| 366.2 | Senile cataract | -2 | 1.60 (1.46 – 1.74) | 4.98E-25 | 102600 |
| 366.2 | Senile cataract | -1 | 1.57 (1.45 – 1.70) | 1.63E-27 | 108657 |
| 366.2 | Senile cataract | 0 | 1.53 (1.42 – 1.64) | 1.15E-29 | 117551 |
| 366.2 | Senile cataract | 1 | 1.49 (1.39 – 1.59) | 2.07E-30 | 124868 |

| PheCode | Disease | Years to OSA diagnosis | Risk ratio | P-value | N |
|---|---|---|---|---|---|
| 366.2 | Senile cataract | 2 | 1.50 (1.40 – 1.60) | 1.06E-34 | 127897 |
| 366.2 | Senile cataract | 3 | 1.49 (1.40 – 1.59) | 9.03E-37 | 130482 |
| 366.2 | Senile cataract | 4 | 1.49 (1.40 – 1.58) | 3.81E-39 | 132711 |
| 366.2 | Senile cataract | 5 | 1.49 (1.40 – 1.57) | 2.61E-42 | 134656 |
| 427.3 | Other specified cardiac dysrhythmias | -5 | 1.68 (1.47 – 1.93) | 4.41E-14 | 85312 |
| 427.3 | Other specified cardiac dysrhythmias | -4 | 1.62 (1.43 – 1.83) | 1.80E-14 | 90932 |
| 427.3 | Other specified cardiac dysrhythmias | -3 | 1.51 (1.35 – 1.69) | 8.27E-13 | 96622 |
| 427.3 | Other specified cardiac dysrhythmias | -2 | 1.51 (1.36 – 1.67) | 4.14E-15 | 102600 |
| 427.3 | Other specified cardiac dysrhythmias | -1 | 1.48 (1.35 – 1.63) | 3.45E-17 | 108657 |
| 427.3 | Other specified cardiac dysrhythmias | 0 | 1.68 (1.55 – 1.82) | 2.54E-36 | 117551 |
| 427.3 | Other specified cardiac dysrhythmias | 1 | 2.04 (1.90 – 2.19) | 1.13E-88 | 124868 |
| 427.3 | Other specified cardiac dysrhythmias | 2 | 2.01 (1.88 – 2.14) | 7.10E-96 | 127897 |
| 427.3 | Other specified cardiac dysrhythmias | 3 | 1.94 (1.82 – 2.07) | 1.49E-96 | 130482 |
| 427.3 | Other specified cardiac dysrhythmias | 4 | 1.90 (1.79 – 2.01) | 8.72E-99 | 132711 |
| 427.3 | Other specified cardiac dysrhythmias | 5 | 1.83 (1.73 – 1.94) | 1.54E-95 | 134656 |
| 375.1 | Dry eyes | -5 | 2.64 (2.05 – 3.40) | 6.26E-14 | 85312 |
| 375.1 | Dry eyes | -4 | 2.36 (1.90 – 2.93) | 1.12E-14 | 90932 |
| 375.1 | Dry eyes | -3 | 2.20 (1.81 – 2.67) | 1.23E-15 | 96622 |
| 375.1 | Dry eyes | -2 | 1.95 (1.65 – 2.31) | 3.87E-15 | 102600 |
| 375.1 | Dry eyes | -1 | 2.05 (1.77 – 2.37) | 1.07E-21 | 108657 |
| 375.1 | Dry eyes | 0 | 2.04 (1.79 – 2.32) | 2.65E-27 | 117551 |
| 375.1 | Dry eyes | 1 | 1.97 (1.76 – 2.21) | 2.45E-31 | 124868 |
| 375.1 | Dry eyes | 2 | 1.94 (1.75 – 2.16) | 6.49E-35 | 127897 |
| 375.1 | Dry eyes | 3 | 1.94 (1.76 – 2.14) | 5.43E-39 | 130482 |
| 375.1 | Dry eyes | 4 | 1.91 (1.74 – 2.10) | 4.63E-41 | 132711 |
| 375.1 | Dry eyes | 5 | 1.91 (1.75 – 2.09) | 1.21E-45 | 134656 |
| 789 | Nausea and vomiting | -5 | 0.74 (0.68 – 0.80) | 4.90E-13 | 85312 |
| 789 | Nausea and vomiting | -4 | 0.78 (0.72 – 0.84) | 2.34E-11 | 90932 |
| 789 | Nausea and vomiting | -3 | 0.81 (0.76 – 0.87) | 2.13E-09 | 96622 |
| 789 | Nausea and vomiting | -2 | 0.84 (0.79 – 0.90) | 1.10E-07 | 102600 |
| 789 | Nausea and vomiting | -1 | 0.86 (0.81 – 0.91) | 9.56E-07 | 108657 |
| 789 | Nausea and vomiting | 0 | 0.91 (0.86 – 0.96) | 1.12E-03 | 117551 |
| 789 | Nausea and vomiting | 1 | 1.11 (1.06 – 1.17) | 8.72E-06 | 124868 |
| 789 | Nausea and vomiting | 2 | 1.16 (1.10 – 1.21) | 4.55E-10 | 127897 |

| PheCode | Disease | Years to OSA diagnosis | Risk ratio | P-value | N |
|---|---|---|---|---|---|
| 789 | Nausea and vomiting | 3 | 1.18 (1.13 – 1.23) | 4.01E-13 | 130482 |
| 789 | Nausea and vomiting | 4 | 1.18 (1.13 – 1.23) | 5.33E-15 | 132711 |
| 789 | Nausea and vomiting | 5 | 1.17 (1.13 – 1.22) | 1.21E-14 | 134656 |
| 721.8 | Other allied disorders of spine | -5 | 5.00 (3.22 – 7.78) | 9.39E-13 | 85312 |
| 721.8 | Other allied disorders of spine | -4 | 5.85 (3.91 – 8.74) | 7.43E-18 | 90932 |
| 721.8 | Other allied disorders of spine | -3 | 6.18 (4.36 – 8.77) | 1.91E-24 | 96622 |
| 721.8 | Other allied disorders of spine | -2 | 6.69 (4.90 – 9.14) | 6.33E-33 | 102600 |
| 721.8 | Other allied disorders of spine | -1 | 6.31 (4.81 – 8.29) | 3.92E-40 | 108657 |
| 721.8 | Other allied disorders of spine | 0 | 6.29 (4.94 – 8.02) | 3.34E-50 | 117551 |
| 721.8 | Other allied disorders of spine | 1 | 6.63 (5.35 – 8.21) | 5.14E-67 | 124868 |
| 721.8 | Other allied disorders of spine | 2 | 6.63 (5.46 – 8.05) | 2.62E-81 | 127897 |
| 721.8 | Other allied disorders of spine | 3 | 6.27 (5.25 – 7.48) | 2.78E-92 | 130482 |
| 721.8 | Other allied disorders of spine | 4 | 5.86 (4.97 – 6.91) | 1.04E-98 | 132711 |
| 721.8 | Other allied disorders of spine | 5 | 6.04 (5.15 – 7.07) | 4.94E-110 | 134656 |
| 379 | Other disorders of eye | -5 | 62.06 (19.49 – 197.59) | 2.83E-12 | 85312 |
| 379 | Other disorders of eye | -4 | 85.24 (26.78 – 271.34) | 5.29E-14 | 90932 |
| 379 | Other disorders of eye | -3 | 54.16 (23.58 – 124.37) | 4.90E-21 | 96622 |
| 379 | Other disorders of eye | -2 | 64.76 (28.51 – 147.08) | 2.18E-23 | 102600 |
| 379 | Other disorders of eye | -1 | 58.44 (28.70 – 118.99) | 3.52E-29 | 108657 |
| 379 | Other disorders of eye | 0 | 52.24 (27.59 – 98.90) | 5.91E-34 | 117551 |
| 379 | Other disorders of eye | 1 | 65.36 (34.84 – 122.60) | 9.14E-39 | 124868 |
| 379 | Other disorders of eye | 2 | 58.64 (33.76 – 101.85) | 2.45E-47 | 127897 |
| 379 | Other disorders of eye | 3 | 63.19 (37.16 – 107.43) | 6.51E-53 | 130482 |
| 379 | Other disorders of eye | 4 | 70.92 (41.75 – 120.46) | 5.32E-56 | 132711 |
| 379 | Other disorders of eye | 5 | 56.42 (35.75 – 89.03) | 2.91E-67 | 134656 |
| 550.2 | Diaphragmatic hernia | -5 | 1.56 (1.37 – 1.77) | 3.72E-12 | 85312 |
| 550.2 | Diaphragmatic hernia | -4 | 1.56 (1.39 – 1.76) | 7.22E-14 | 90932 |
| 550.2 | Diaphragmatic hernia | -3 | 1.63 (1.47 – 1.82) | 8.06E-19 | 96622 |
| 550.2 | Diaphragmatic hernia | -2 | 1.63 (1.47 – 1.80) | 5.04E-21 | 102600 |
| 550.2 | Diaphragmatic hernia | -1 | 1.66 (1.51 – 1.82) | 5.02E-26 | 108657 |
| 550.2 | Diaphragmatic hernia | 0 | 1.80 (1.65 – 1.96) | 1.04E-40 | 117551 |
| 550.2 | Diaphragmatic hernia | 1 | 2.25 (2.08 – 2.43) | 3.28E-93 | 124868 |
| 550.2 | Diaphragmatic hernia | 2 | 2.34 (2.18 – 2.52) | 1.13E-112 | 127897 |
| 550.2 | Diaphragmatic hernia | 3 | 2.38 (2.21 – 2.55) | 1.12E-123 | 130482 |

| PheCode | Disease | Years to OSA diagnosis | Risk ratio | P-value | N |
|---|---|---|---|---|---|
| 550.2 | Diaphragmatic hernia | 4 | 2.40 (2.24 – 2.57) | 5.33E-135 | 132711 |
| 550.2 | Diaphragmatic hernia | 5 | 2.41 (2.25 – 2.57) | 8.86E-144 | 134656 |
| 165.1 | Cancer of bronchus; lung | -5 | 0.49 (0.40 – 0.60) | 5.61E-12 | 85312 |
| 165.1 | Cancer of bronchus; lung | -4 | 0.46 (0.38 – 0.55) | 3.64E-16 | 90932 |
| 165.1 | Cancer of bronchus; lung | -3 | 0.46 (0.39 – 0.55) | 4.67E-19 | 96622 |
| 165.1 | Cancer of bronchus; lung | -2 | 0.47 (0.40 – 0.55) | 5.82E-21 | 102600 |
| 165.1 | Cancer of bronchus; lung | -1 | 0.47 (0.40 – 0.54) | 2.84E-25 | 108657 |
| 165.1 | Cancer of bronchus; lung | 0 | 0.58 (0.52 – 0.66) | 3.09E-18 | 117551 |
| 165.1 | Cancer of bronchus; lung | 1 | 0.71 (0.64 – 0.79) | 3.44E-10 | 124868 |
| 165.1 | Cancer of bronchus; lung | 2 | 0.72 (0.65 – 0.79) | 6.20E-11 | 127897 |
| 165.1 | Cancer of bronchus; lung | 3 | 0.71 (0.65 – 0.79) | 4.96E-12 | 130482 |
| 165.1 | Cancer of bronchus; lung | 4 | 0.73 (0.67 – 0.80) | 2.72E-11 | 132711 |
| 165.1 | Cancer of bronchus; lung | 5 | 0.72 (0.66 – 0.79) | 3.98E-13 | 134656 |
| 208 | Benign neoplasm of colon | -5 | 1.22 (1.15 – 1.30) | 1.92E-11 | 85312 |
| 208 | Benign neoplasm of colon | -4 | 1.25 (1.18 – 1.31) | 3.26E-15 | 90932 |
| 208 | Benign neoplasm of colon | -3 | 1.26 (1.19 – 1.32) | 1.68E-18 | 96622 |
| 208 | Benign neoplasm of colon | -2 | 1.30 (1.24 – 1.36) | 2.37E-27 | 102600 |
| 208 | Benign neoplasm of colon | -1 | 1.32 (1.26 – 1.38) | 3.94E-35 | 108657 |
| 208 | Benign neoplasm of colon | 0 | 1.31 (1.25 – 1.36) | 3.53E-37 | 117551 |
| 208 | Benign neoplasm of colon | 1 | 1.42 (1.36 – 1.47) | 6.61E-72 | 124868 |
| 208 | Benign neoplasm of colon | 2 | 1.48 (1.43 – 1.54) | 3.64E-97 | 127897 |
| 208 | Benign neoplasm of colon | 3 | 1.51 (1.45 – 1.56) | 3.94E-111 | 130482 |
| 208 | Benign neoplasm of colon | 4 | 1.52 (1.46 – 1.57) | 1.28E-120 | 132711 |
| 208 | Benign neoplasm of colon | 5 | 1.53 (1.48 – 1.59) | 1.02E-133 | 134656 |
| 571.51 | Cirrhosis of liver without mention of alcohol | -5 | 0.39 (0.30 – 0.51) | 1.92E-11 | 85312 |
| 571.51 | Cirrhosis of liver without mention of alcohol | -4 | 0.40 (0.31 – 0.52) | 5.89E-13 | 90932 |
| 571.51 | Cirrhosis of liver without mention of alcohol | -3 | 0.39 (0.31 – 0.49) | 4.45E-16 | 96622 |
| 571.51 | Cirrhosis of liver without mention of alcohol | -2 | 0.41 (0.33 – 0.50) | 1.15E-17 | 102600 |
| 571.51 | Cirrhosis of liver without mention of alcohol | -1 | 0.44 (0.36 – 0.52) | 5.45E-19 | 108657 |
| 571.51 | Cirrhosis of liver without mention of alcohol | 0 | 0.54 (0.47 – 0.63) | 3.72E-15 | 117551 |
| 571.51 | Cirrhosis of liver without mention of alcohol | 1 | 0.67 (0.59 – 0.77) | 3.09E-09 | 124868 |
| 571.51 | Cirrhosis of liver without mention of alcohol | 2 | 0.71 (0.63 – 0.80) | 5.31E-08 | 127897 |
| 571.51 | Cirrhosis of liver without mention of alcohol | 3 | 0.72 (0.64 – 0.81) | 4.01E-08 | 130482 |
| 571.51 | Cirrhosis of liver without mention of alcohol | 4 | 0.72 (0.65 – 0.81) | 3.95E-08 | 132711 |

| PheCode | Disease | Years to OSA diagnosis | Risk ratio | P-value | N |
|---|---|---|---|---|---|
| 571.51 | Cirrhosis of liver without mention of alcohol | 5 | 0.72 (0.65 – 0.81) | 1.09E-08 | 134656 |
| 296.2 | Depression | -5 | 1.19 (1.13 – 1.25) | 5.26E-11 | 85312 |
| 296.2 | Depression | -4 | 1.21 (1.15 – 1.27) | 9.68E-15 | 90932 |
| 296.2 | Depression | -3 | 1.24 (1.18 – 1.30) | 5.64E-20 | 96622 |
| 296.2 | Depression | -2 | 1.27 (1.21 – 1.33) | 1.05E-25 | 102600 |
| 296.2 | Depression | -1 | 1.30 (1.24 – 1.36) | 8.67E-33 | 108657 |
| 296.2 | Depression | 0 | 1.31 (1.26 – 1.37) | 7.03E-38 | 117551 |
| 296.2 | Depression | 1 | 1.46 (1.40 – 1.52) | 1.28E-77 | 124868 |
| 296.2 | Depression | 2 | 1.49 (1.43 – 1.55) | 1.48E-87 | 127897 |
| 296.2 | Depression | 3 | 1.51 (1.45 – 1.57) | 1.53E-92 | 130482 |
| 296.2 | Depression | 4 | 1.51 (1.45 – 1.57) | 1.24E-97 | 132711 |
| 296.2 | Depression | 5 | 1.51 (1.46 – 1.57) | 6.04E-100 | 134656 |
| 250.2 | Type 2 diabetes | -5 | 1.19 (1.13 – 1.26) | 8.07E-11 | 85312 |
| 250.2 | Type 2 diabetes | -4 | 1.20 (1.15 – 1.27) | 2.94E-13 | 90932 |
| 250.2 | Type 2 diabetes | -3 | 1.22 (1.16 – 1.28) | 7.45E-17 | 96622 |
| 250.2 | Type 2 diabetes | -2 | 1.26 (1.20 – 1.31) | 4.23E-25 | 102600 |
| 250.2 | Type 2 diabetes | -1 | 1.28 (1.23 – 1.34) | 9.76E-33 | 108657 |
| 250.2 | Type 2 diabetes | 0 | 1.35 (1.30 – 1.40) | 6.43E-56 | 117551 |
| 250.2 | Type 2 diabetes | 1 | 1.58 (1.52 – 1.63) | 9.49E-153 | 124868 |
| 250.2 | Type 2 diabetes | 2 | 1.57 (1.52 – 1.62) | 2.58E-158 | 127897 |
| 250.2 | Type 2 diabetes | 3 | 1.57 (1.52 – 1.62) | 3.88E-163 | 130482 |
| 250.2 | Type 2 diabetes | 4 | 1.55 (1.50 – 1.60) | 1.22E-163 | 132711 |
| 250.2 | Type 2 diabetes | 5 | 1.54 (1.50 – 1.59) | 8.86E-167 | 134656 |
| 475 | Chronic sinusitis | -5 | 1.30 (1.20 – 1.41) | 2.21E-10 | 85312 |
| 475 | Chronic sinusitis | -4 | 1.31 (1.21 – 1.41) | 6.42E-12 | 90932 |
| 475 | Chronic sinusitis | -3 | 1.33 (1.24 – 1.43) | 1.80E-14 | 96622 |
| 475 | Chronic sinusitis | -2 | 1.37 (1.28 – 1.46) | 6.17E-19 | 102600 |
| 475 | Chronic sinusitis | -1 | 1.45 (1.36 – 1.55) | 6.35E-29 | 108657 |
| 475 | Chronic sinusitis | 0 | 1.53 (1.44 – 1.63) | 3.33E-43 | 117551 |
| 475 | Chronic sinusitis | 1 | 1.97 (1.86 – 2.08) | 1.20E-132 | 124868 |
| 475 | Chronic sinusitis | 2 | 2.05 (1.95 – 2.16) | 4.03E-159 | 127897 |
| 475 | Chronic sinusitis | 3 | 2.09 (1.98 – 2.20) | 1.52E-176 | 130482 |
| 475 | Chronic sinusitis | 4 | 2.10 (2.00 – 2.21) | 6.34E-190 | 132711 |
| 475 | Chronic sinusitis | 5 | 2.11 (2.01 – 2.22) | 1.28E-201 | 134656 |

| PheCode | Disease | Years to OSA diagnosis | Risk ratio | P-value | N |
|---|---|---|---|---|---|
| 389 | Hearing loss | -5 | 1.43 (1.28 – 1.60) | 3.19E-10 | 85312 |
| 389 | Hearing loss | -4 | 1.49 (1.35 – 1.65) | 5.09E-15 | 90932 |
| 389 | Hearing loss | -3 | 1.48 (1.35 – 1.62) | 2.99E-17 | 96622 |
| 389 | Hearing loss | -2 | 1.52 (1.40 – 1.65) | 6.34E-24 | 102600 |
| 389 | Hearing loss | -1 | 1.51 (1.40 – 1.63) | 6.96E-28 | 108657 |
| 389 | Hearing loss | 0 | 1.57 (1.47 – 1.68) | 1.76E-40 | 117551 |
| 389 | Hearing loss | 1 | 1.90 (1.79 – 2.02) | 9.06E-104 | 124868 |
| 389 | Hearing loss | 2 | 1.93 (1.82 – 2.04) | 4.77E-121 | 127897 |
| 389 | Hearing loss | 3 | 1.90 (1.81 – 2.01) | 9.89E-129 | 130482 |
| 389 | Hearing loss | 4 | 1.88 (1.79 – 1.98) | 5.63E-136 | 132711 |
| 389 | Hearing loss | 5 | 1.85 (1.77 – 1.94) | 4.55E-140 | 134656 |
| 564.1 | Irritable Bowel Syndrome | -5 | 1.45 (1.29 – 1.63) | 3.80E-10 | 85312 |
| 564.1 | Irritable Bowel Syndrome | -4 | 1.49 (1.34 – 1.67) | 7.90E-13 | 90932 |
| 564.1 | Irritable Bowel Syndrome | -3 | 1.54 (1.39 – 1.70) | 2.02E-16 | 96622 |
| 564.1 | Irritable Bowel Syndrome | -2 | 1.62 (1.47 – 1.79) | 4.67E-22 | 102600 |
| 564.1 | Irritable Bowel Syndrome | -1 | 1.75 (1.59 – 1.91) | 3.44E-33 | 108657 |
| 564.1 | Irritable Bowel Syndrome | 0 | 1.78 (1.64 – 1.94) | 1.99E-40 | 117551 |
| 564.1 | Irritable Bowel Syndrome | 1 | 1.97 (1.83 – 2.13) | 6.40E-66 | 124868 |
| 564.1 | Irritable Bowel Syndrome | 2 | 2.02 (1.87 – 2.18) | 1.24E-74 | 127897 |
| 564.1 | Irritable Bowel Syndrome | 3 | 2.04 (1.89 – 2.19) | 2.43E-82 | 130482 |
| 564.1 | Irritable Bowel Syndrome | 4 | 2.05 (1.91 – 2.19) | 4.36E-89 | 132711 |
| 564.1 | Irritable Bowel Syndrome | 5 | 2.03 (1.89 – 2.17) | 1.66E-91 | 134656 |
| 278.1 | Obesity | -5 | 1.16 (1.10 – 1.21) | 1.40E-09 | 85312 |
| 278.1 | Obesity | -4 | 1.18 (1.13 – 1.24) | 5.09E-14 | 90932 |
| 278.1 | Obesity | -3 | 1.23 (1.19 – 1.29) | 2.44E-24 | 96622 |
| 278.1 | Obesity | -2 | 1.25 (1.21 – 1.30) | 3.02E-31 | 102600 |
| 278.1 | Obesity | -1 | 1.30 (1.25 – 1.34) | 2.12E-48 | 108657 |
| 278.1 | Obesity | 0 | 1.39 (1.34 – 1.43) | 3.21E-91 | 117551 |
| 278.1 | Obesity | 1 | 1.78 (1.73 – 1.83) | 0.00E+00 | 124868 |
| 278.1 | Obesity | 2 | 1.82 (1.77 – 1.87) | 0.00E+00 | 127897 |
| 278.1 | Obesity | 3 | 1.81 (1.76 – 1.86) | 0.00E+00 | 130482 |
| 278.1 | Obesity | 4 | 1.79 (1.75 – 1.84) | 0.00E+00 | 132711 |
| 278.1 | Obesity | 5 | 1.77 (1.73 – 1.82) | 0.00E+00 | 134656 |
| 338.2 | Chronic pain | -5 | 1.51 (1.31 – 1.74) | 1.35E-08 | 85312 |

| PheCode | Disease | Years to OSA diagnosis | Risk ratio | P-value | N |
|---|---|---|---|---|---|
| 338.2 | Chronic pain | -4 | 1.50 (1.33 – 1.68) | 1.91E-11 | 90932 |
| 338.2 | Chronic pain | -3 | 1.52 (1.37 – 1.67) | 1.94E-16 | 96622 |
| 338.2 | Chronic pain | -2 | 1.47 (1.35 – 1.60) | 6.55E-19 | 102600 |
| 338.2 | Chronic pain | -1 | 1.50 (1.40 – 1.62) | 8.97E-27 | 108657 |
| 338.2 | Chronic pain | 0 | 1.58 (1.48 – 1.69) | 5.64E-43 | 117551 |
| 338.2 | Chronic pain | 1 | 1.79 (1.69 – 1.90) | 6.62E-85 | 124868 |
| 338.2 | Chronic pain | 2 | 1.80 (1.70 – 1.90) | 2.48E-97 | 127897 |
| 338.2 | Chronic pain | 3 | 1.81 (1.72 – 1.91) | 6.14E-111 | 130482 |
| 338.2 | Chronic pain | 4 | 1.75 (1.67 – 1.84) | 1.10E-111 | 132711 |
| 338.2 | Chronic pain | 5 | 1.72 (1.64 – 1.80) | 1.71E-114 | 134656 |
| 798.1 | Chronic fatigue syndrome | -5 | 2.24 (1.67 – 2.99) | 5.19E-08 | 85312 |
| 798.1 | Chronic fatigue syndrome | -4 | 2.19 (1.67 – 2.86) | 1.14E-08 | 90932 |
| 798.1 | Chronic fatigue syndrome | -3 | 2.12 (1.66 – 2.70) | 1.33E-09 | 96622 |
| 798.1 | Chronic fatigue syndrome | -2 | 2.12 (1.71 – 2.62) | 4.88E-12 | 102600 |
| 798.1 | Chronic fatigue syndrome | -1 | 2.29 (1.91 – 2.76) | 7.76E-19 | 108657 |
| 798.1 | Chronic fatigue syndrome | 0 | 3.24 (2.78 – 3.77) | 1.85E-51 | 117551 |
| 798.1 | Chronic fatigue syndrome | 1 | 3.54 (3.10 – 4.04) | 9.98E-79 | 124868 |
| 798.1 | Chronic fatigue syndrome | 2 | 3.56 (3.15 – 4.02) | 1.54E-91 | 127897 |
| 798.1 | Chronic fatigue syndrome | 3 | 3.41 (3.04 – 3.83) | 1.07E-96 | 130482 |
| 798.1 | Chronic fatigue syndrome | 4 | 3.32 (2.98 – 3.70) | 8.92E-104 | 132711 |
| 798.1 | Chronic fatigue syndrome | 5 | 3.20 (2.89 – 3.55) | 1.49E-107 | 134656 |
| 327.71 | Restless legs syndrome | -5 | 2.38 (1.74 – 3.26) | 5.25E-08 | 85312 |
| 327.71 | Restless legs syndrome | -4 | 2.58 (1.96 – 3.39) | 1.57E-11 | 90932 |
| 327.71 | Restless legs syndrome | -3 | 2.45 (1.93 – 3.11) | 1.85E-13 | 96622 |
| 327.71 | Restless legs syndrome | -2 | 2.35 (1.91 – 2.91) | 2.00E-15 | 102600 |
| 327.71 | Restless legs syndrome | -1 | 2.64 (2.20 – 3.18) | 6.99E-25 | 108657 |
| 327.71 | Restless legs syndrome | 0 | 3.60 (3.08 – 4.22) | 8.88E-58 | 117551 |
| 327.71 | Restless legs syndrome | 1 | 7.12 (6.25 – 8.10) | 2.10E-193 | 124868 |
| 327.71 | Restless legs syndrome | 2 | 7.09 (6.29 – 8.01) | 1.10E-221 | 127897 |
| 327.71 | Restless legs syndrome | 3 | 6.86 (6.12 – 7.69) | 7.23E-239 | 130482 |
| 327.71 | Restless legs syndrome | 4 | 6.87 (6.15 – 7.68) | 7.27E-256 | 132711 |
| 327.71 | Restless legs syndrome | 5 | 6.62 (5.95 – 7.36) | 7.34E-268 | 134656 |
| 740.9 | Osteoarthrosis NOS | -5 | 1.13 (1.08 – 1.18) | 1.17E-07 | 85312 |
| 740.9 | Osteoarthrosis NOS | -4 | 1.14 (1.10 – 1.19) | 3.36E-10 | 90932 |

| PheCode | Disease | Years to OSA diagnosis | Risk ratio | P-value | N |
|---|---|---|---|---|---|
| 740.9 | Osteoarthrosis NOS | -3 | 1.16 (1.11 – 1.21) | 2.41E-13 | 96622 |
| 740.9 | Osteoarthrosis NOS | -2 | 1.17 (1.13 – 1.21) | 2.97E-16 | 102600 |
| 740.9 | Osteoarthrosis NOS | -1 | 1.19 (1.14 – 1.23) | 6.74E-21 | 108657 |
| 740.9 | Osteoarthrosis NOS | 0 | 1.19 (1.15 – 1.23) | 5.28E-24 | 117551 |
| 740.9 | Osteoarthrosis NOS | 1 | 1.32 (1.28 – 1.36) | 6.00E-66 | 124868 |
| 740.9 | Osteoarthrosis NOS | 2 | 1.36 (1.32 – 1.41) | 6.69E-82 | 127897 |
| 740.9 | Osteoarthrosis NOS | 3 | 1.38 (1.33 – 1.42) | 1.54E-87 | 130482 |
| 740.9 | Osteoarthrosis NOS | 4 | 1.38 (1.34 – 1.43) | 3.36E-91 | 132711 |
| 740.9 | Osteoarthrosis NOS | 5 | 1.38 (1.34 – 1.43) | 1.08E-94 | 134656 |
| 512.9 | Other dyspnea | -5 | 0.83 (0.78 – 0.89) | 3.39E-07 | 85312 |
| 512.9 | Other dyspnea | -4 | 0.89 (0.83 – 0.95) | 2.48E-04 | 90932 |
| 512.9 | Other dyspnea | -3 | 0.93 (0.87 – 0.98) | 1.16E-02 | 96622 |
| 512.9 | Other dyspnea | -2 | 0.98 (0.93 – 1.04) | 5.08E-01 | 102600 |
| 512.9 | Other dyspnea | -1 | 1.08 (1.03 – 1.14) | 1.89E-03 | 108657 |
| 512.9 | Other dyspnea | 0 | 1.52 (1.46 – 1.58) | 1.23E-86 | 117551 |
| 512.9 | Other dyspnea | 1 | 1.83 (1.76 – 1.90) | 8.77E-220 | 124868 |
| 512.9 | Other dyspnea | 2 | 1.84 (1.78 – 1.91) | 1.60E-235 | 127897 |
| 512.9 | Other dyspnea | 3 | 1.82 (1.76 – 1.89) | 3.69E-236 | 130482 |
| 512.9 | Other dyspnea | 4 | 1.80 (1.74 – 1.86) | 8.43E-238 | 132711 |
| 512.9 | Other dyspnea | 5 | 1.78 (1.72 – 1.85) | 2.38E-241 | 134656 |
| 455 | Hemorrhoids | -5 | 1.21 (1.13 – 1.31) | 3.74E-07 | 85312 |
| 455 | Hemorrhoids | -4 | 1.31 (1.23 – 1.41) | 1.08E-14 | 90932 |
| 455 | Hemorrhoids | -3 | 1.34 (1.26 – 1.43) | 7.90E-19 | 96622 |
| 455 | Hemorrhoids | -2 | 1.40 (1.32 – 1.49) | 1.80E-28 | 102600 |
| 455 | Hemorrhoids | -1 | 1.43 (1.35 – 1.51) | 2.26E-35 | 108657 |
| 455 | Hemorrhoids | 0 | 1.48 (1.40 – 1.55) | 1.50E-48 | 117551 |
| 455 | Hemorrhoids | 1 | 1.64 (1.56 – 1.72) | 1.15E-89 | 124868 |
| 455 | Hemorrhoids | 2 | 1.76 (1.68 – 1.84) | 4.00E-130 | 127897 |
| 455 | Hemorrhoids | 3 | 1.85 (1.77 – 1.93) | 2.95E-167 | 130482 |
| 455 | Hemorrhoids | 4 | 1.90 (1.82 – 1.98) | 1.73E-196 | 132711 |
| 455 | Hemorrhoids | 5 | 1.95 (1.87 – 2.03) | 1.15E-226 | 134656 |
| 300.11 | Generalized anxiety disorder | -5 | 1.32 (1.17 – 1.50) | 1.28E-05 | 85312 |
| 300.11 | Generalized anxiety disorder | -4 | 1.39 (1.24 – 1.55) | 1.66E-08 | 90932 |
| 300.11 | Generalized anxiety disorder | -3 | 1.45 (1.31 – 1.61) | 5.47E-13 | 96622 |

| PheCode | Disease | Years to OSA diagnosis | Risk ratio | P-value | N |
|---|---|---|---|---|---|
| 300.11 | Generalized anxiety disorder | -2 | 1.53 (1.40 – 1.68) | 6.79E-20 | 102600 |
| 300.11 | Generalized anxiety disorder | -1 | 1.62 (1.49 – 1.76) | 3.89E-30 | 108657 |
| 300.11 | Generalized anxiety disorder | 0 | 1.74 (1.62 – 1.88) | 2.26E-49 | 117551 |
| 300.11 | Generalized anxiety disorder | 1 | 1.95 (1.82 – 2.08) | 7.41E-87 | 124868 |
| 300.11 | Generalized anxiety disorder | 2 | 1.97 (1.85 – 2.10) | 1.18E-96 | 127897 |
| 300.11 | Generalized anxiety disorder | 3 | 1.97 (1.85 – 2.09) | 2.52E-102 | 130482 |
| 300.11 | Generalized anxiety disorder | 4 | 1.94 (1.82 – 2.05) | 5.71E-106 | 132711 |
| 300.11 | Generalized anxiety disorder | 5 | 1.93 (1.82 – 2.04) | 1.67E-112 | 134656 |
| 271.3 | Intestinal disaccharidase deficiencies | -5 | 1.63 (1.29 – 2.05) | 3.27E-05 | 85312 |
| 271.3 | Intestinal disaccharidase deficiencies | -4 | 1.72 (1.42 – 2.09) | 4.18E-08 | 90932 |
| 271.3 | Intestinal disaccharidase deficiencies | -3 | 1.89 (1.61 – 2.21) | 6.59E-15 | 96622 |
| 271.3 | Intestinal disaccharidase deficiencies | -2 | 1.79 (1.56 – 2.05) | 4.19E-17 | 102600 |
| 271.3 | Intestinal disaccharidase deficiencies | -1 | 1.85 (1.66 – 2.07) | 3.41E-27 | 108657 |
| 271.3 | Intestinal disaccharidase deficiencies | 0 | 1.90 (1.73 – 2.08) | 2.08E-42 | 117551 |
| 271.3 | Intestinal disaccharidase deficiencies | 1 | 2.28 (2.12 – 2.47) | 5.10E-99 | 124868 |
| 271.3 | Intestinal disaccharidase deficiencies | 2 | 2.23 (2.08 – 2.40) | 5.15E-114 | 127897 |
| 271.3 | Intestinal disaccharidase deficiencies | 3 | 2.15 (2.02 – 2.30) | 4.43E-121 | 130482 |
| 271.3 | Intestinal disaccharidase deficiencies | 4 | 2.12 (1.99 – 2.25) | 1.04E-132 | 132711 |
| 271.3 | Intestinal disaccharidase deficiencies | 5 | 2.03 (1.92 – 2.15) | 1.57E-134 | 134656 |
| 292.3 | Memory loss | -5 | 1.52 (1.23 – 1.88) | 8.62E-05 | 85312 |
| 292.3 | Memory loss | -4 | 1.69 (1.41 – 2.03) | 1.40E-08 | 90932 |
| 292.3 | Memory loss | -3 | 1.63 (1.39 – 1.91) | 2.57E-09 | 96622 |
| 292.3 | Memory loss | -2 | 1.70 (1.48 – 1.95) | 1.09E-13 | 102600 |
| 292.3 | Memory loss | -1 | 1.85 (1.64 – 2.09) | 6.78E-23 | 108657 |
| 292.3 | Memory loss | 0 | 2.44 (2.20 – 2.70) | 2.98E-66 | 117551 |
| 292.3 | Memory loss | 1 | 2.76 (2.52 – 3.02) | 2.27E-109 | 124868 |
| 292.3 | Memory loss | 2 | 2.82 (2.59 – 3.07) | 3.67E-130 | 127897 |
| 292.3 | Memory loss | 3 | 2.76 (2.55 – 2.99) | 4.17E-142 | 130482 |
| 292.3 | Memory loss | 4 | 2.70 (2.50 – 2.91) | 1.36E-148 | 132711 |
| 292.3 | Memory loss | 5 | 2.65 (2.47 – 2.85) | 6.36E-155 | 134656 |
| 760 | Back pain | -5 | 1.07 (1.03 – 1.12) | 6.49E-04 | 85312 |
| 760 | Back pain | -4 | 1.09 (1.05 – 1.14) | 4.61E-06 | 90932 |
| 760 | Back pain | -3 | 1.13 (1.09 – 1.17) | 2.08E-11 | 96622 |
| 760 | Back pain | -2 | 1.15 (1.11 – 1.19) | 1.14E-14 | 102600 |

| PheCode | Disease | Years to OSA diagnosis | Risk ratio | P-value | N |
|---|---|---|---|---|---|
| 760 | Back pain | -1 | 1.16 (1.12 – 1.19) | 5.49E-18 | 108657 |
| 760 | Back pain | 0 | 1.15 (1.12 – 1.19) | 2.97E-19 | 117551 |
| 760 | Back pain | 1 | 1.23 (1.20 – 1.27) | 4.58E-44 | 124868 |
| 760 | Back pain | 2 | 1.27 (1.24 – 1.31) | 1.95E-62 | 127897 |
| 760 | Back pain | 3 | 1.30 (1.26 – 1.33) | 3.64E-75 | 130482 |
| 760 | Back pain | 4 | 1.31 (1.27 – 1.34) | 4.26E-85 | 132711 |
| 760 | Back pain | 5 | 1.31 (1.28 – 1.35) | 3.87E-92 | 134656 |
| 278.11 | Morbid obesity | -5 | 0.86 (0.76 – 0.98) | 1.88E-02 | 85312 |
| 278.11 | Morbid obesity | -4 | 0.89 (0.79 – 0.99) | 3.71E-02 | 90932 |
| 278.11 | Morbid obesity | -3 | 0.96 (0.87 – 1.06) | 4.57E-01 | 96622 |
| 278.11 | Morbid obesity | -2 | 1.06 (0.97 – 1.15) | 2.37E-01 | 102600 |
| 278.11 | Morbid obesity | -1 | 1.13 (1.04 – 1.22) | 2.47E-03 | 108657 |
| 278.11 | Morbid obesity | 0 | 1.57 (1.47 – 1.68) | 1.42E-39 | 117551 |
| 278.11 | Morbid obesity | 1 | 2.19 (2.06 – 2.32) | 1.99E-139 | 124868 |
| 278.11 | Morbid obesity | 2 | 2.20 (2.06 – 2.34) | 7.28E-135 | 127897 |
| 278.11 | Morbid obesity | 3 | 2.14 (2.02 – 2.27) | 9.40E-138 | 130482 |
| 278.11 | Morbid obesity | 4 | 2.07 (1.96 – 2.19) | 2.15E-138 | 132711 |
| 278.11 | Morbid obesity | 5 | 2.02 (1.91 – 2.14) | 1.77E-138 | 134656 |
| 496 | Chronic airway obstruction | -5 | 1.08 (0.97 – 1.20) | 1.58E-01 | 85312 |
| 496 | Chronic airway obstruction | -4 | 1.10 (1.00 – 1.22) | 5.31E-02 | 90932 |
| 496 | Chronic airway obstruction | -3 | 1.19 (1.09 – 1.30) | 1.47E-04 | 96622 |
| 496 | Chronic airway obstruction | -2 | 1.23 (1.14 – 1.34) | 7.27E-07 | 102600 |
| 496 | Chronic airway obstruction | -1 | 1.33 (1.24 – 1.44) | 8.02E-14 | 108657 |
| 496 | Chronic airway obstruction | 0 | 1.60 (1.49 – 1.71) | 2.71E-43 | 117551 |
| 496 | Chronic airway obstruction | 1 | 2.10 (1.98 – 2.23) | 3.24E-134 | 124868 |
| 496 | Chronic airway obstruction | 2 | 2.07 (1.96 – 2.20) | 1.70E-137 | 127897 |
| 496 | Chronic airway obstruction | 3 | 2.02 (1.91 – 2.14) | 1.10E-134 | 130482 |
| 496 | Chronic airway obstruction | 4 | 1.98 (1.88 – 2.10) | 1.76E-133 | 132711 |
| 496 | Chronic airway obstruction | 5 | 1.93 (1.83 – 2.04) | 1.68E-128 | 134656 |
| 474.2 | Chronic tonsillitis and adenoiditis | -5 | 1.12 (0.89 – 1.41) | 3.15E-01 | 85312 |
| 474.2 | Chronic tonsillitis and adenoiditis | -4 | 1.08 (0.87 – 1.34) | 4.98E-01 | 90932 |
| 474.2 | Chronic tonsillitis and adenoiditis | -3 | 1.11 (0.91 – 1.36) | 2.93E-01 | 96622 |
| 474.2 | Chronic tonsillitis and adenoiditis | -2 | 1.18 (0.98 – 1.42) | 7.67E-02 | 102600 |
| 474.2 | Chronic tonsillitis and adenoiditis | -1 | 1.31 (1.11 – 1.55) | 1.61E-03 | 108657 |

| PheCode | Disease | Years to OSA diagnosis | Risk ratio | P-value | N |
|---|---|---|---|---|---|
| 474.2 | Chronic tonsillitis and adenoiditis | 0 | 2.01 (1.75 – 2.31) | 7.27E-23 | 117551 |
| 474.2 | Chronic tonsillitis and adenoiditis | 1 | 4.84 (4.36 – 5.39) | 8.33E-187 | 124868 |
| 474.2 | Chronic tonsillitis and adenoiditis | 2 | 5.00 (4.52 – 5.53) | 4.33E-212 | 127897 |
| 474.2 | Chronic tonsillitis and adenoiditis | 3 | 5.02 (4.55 – 5.53) | 6.43E-230 | 130482 |
| 474.2 | Chronic tonsillitis and adenoiditis | 4 | 5.01 (4.56 – 5.51) | 3.02E-243 | 132711 |
| 474.2 | Chronic tonsillitis and adenoiditis | 5 | 4.98 (4.54 – 5.46) | 6.85E-252 | 134656 |
| 278 | Overweight, obesity, hyperalimentation | -5 | 1.05 (0.95 – 1.16) | 3.22E-01 | 85312 |
| 278 | Overweight, obesity, hyperalimentation | -4 | 1.05 (0.96 – 1.15) | 2.54E-01 | 90932 |
| 278 | Overweight, obesity, hyperalimentation | -3 | 1.07 (0.99 – 1.16) | 8.02E-02 | 96622 |
| 278 | Overweight, obesity, hyperalimentation | -2 | 1.12 (1.04 – 1.20) | 2.99E-03 | 102600 |
| 278 | Overweight, obesity, hyperalimentation | -1 | 1.13 (1.06 – 1.21) | 2.31E-04 | 108657 |
| 278 | Overweight, obesity, hyperalimentation | 0 | 1.23 (1.16 – 1.31) | 4.80E-12 | 117551 |
| 278 | Overweight, obesity, hyperalimentation | 1 | 1.75 (1.67 – 1.84) | 7.79E-115 | 124868 |
| 278 | Overweight, obesity, hyperalimentation | 2 | 1.77 (1.69 – 1.85) | 3.42E-132 | 127897 |
| 278 | Overweight, obesity, hyperalimentation | 3 | 1.80 (1.72 – 1.88) | 5.60E-153 | 130482 |
| 278 | Overweight, obesity, hyperalimentation | 4 | 1.79 (1.71 – 1.86) | 7.93E-162 | 132711 |
| 278 | Overweight, obesity, hyperalimentation | 5 | 1.79 (1.72 – 1.87) | 1.21E-175 | 134656 |
| 292 | Neurological disorders | -5 | 1.27 (0.67 – 2.39) | 4.64E-01 | 85312 |
| 292 | Neurological disorders | -4 | 1.86 (1.14 – 3.04) | 1.29E-02 | 90932 |
| 292 | Neurological disorders | -3 | 2.21 (1.49 – 3.28) | 8.82E-05 | 96622 |
| 292 | Neurological disorders | -2 | 2.16 (1.58 – 2.94) | 1.23E-06 | 102600 |
| 292 | Neurological disorders | -1 | 2.51 (1.96 – 3.22) | 4.00E-13 | 108657 |
| 292 | Neurological disorders | 0 | 3.60 (2.94 – 4.40) | 7.77E-36 | 117551 |
| 292 | Neurological disorders | 1 | 4.17 (3.51 – 4.95) | 8.69E-60 | 124868 |
| 292 | Neurological disorders | 2 | 4.18 (3.56 – 4.89) | 3.07E-70 | 127897 |
| 292 | Neurological disorders | 3 | 4.04 (3.49 – 4.67) | 7.16E-78 | 130482 |
| 292 | Neurological disorders | 4 | 4.08 (3.55 – 4.68) | 3.59E-88 | 132711 |
| 292 | Neurological disorders | 5 | 4.00 (3.50 – 4.56) | 3.17E-93 | 134656 |
| 428.1 | Congestive heart failure (CHF) NOS | -5 | 0.98 (0.89 – 1.07) | 6.41E-01 | 85312 |
| 428.1 | Congestive heart failure (CHF) NOS | -4 | 1.00 (0.92 – 1.09) | 9.77E-01 | 90932 |
| 428.1 | Congestive heart failure (CHF) NOS | -3 | 1.04 (0.96 – 1.12) | 3.69E-01 | 96622 |
| 428.1 | Congestive heart failure (CHF) NOS | -2 | 1.09 (1.01 – 1.17) | 2.02E-02 | 102600 |
| 428.1 | Congestive heart failure (CHF) NOS | -1 | 1.14 (1.07 – 1.22) | 5.26E-05 | 108657 |
| 428.1 | Congestive heart failure (CHF) NOS | 0 | 1.41 (1.33 – 1.49) | 1.50E-32 | 117551 |

| PheCode | Disease | Years to OSA diagnosis | Risk ratio | P-value | N |
|---|---|---|---|---|---|
| 428.1 | Congestive heart failure (CHF) NOS | 1 | 1.76 (1.67 – 1.85) | 8.68E-106 | 124868 |
| 428.1 | Congestive heart failure (CHF) NOS | 2 | 1.75 (1.67 – 1.84) | 3.37E-108 | 127897 |
| 428.1 | Congestive heart failure (CHF) NOS | 3 | 1.74 (1.66 – 1.83) | 2.59E-109 | 130482 |
| 428.1 | Congestive heart failure (CHF) NOS | 4 | 1.71 (1.63 – 1.80) | 8.64E-107 | 132711 |
| 428.1 | Congestive heart failure (CHF) NOS | 5 | 1.69 (1.61 – 1.77) | 9.37E-105 | 134656 |
| 512.7 | Shortness of breath | -5 | 0.99 (0.93 – 1.07) | 8.83E-01 | 85312 |
| 512.7 | Shortness of breath | -4 | 1.01 (0.94 – 1.07) | 8.64E-01 | 90932 |
| 512.7 | Shortness of breath | -3 | 1.03 (0.97 – 1.10) | 2.67E-01 | 96622 |
| 512.7 | Shortness of breath | -2 | 1.05 (1.00 – 1.11) | 6.32E-02 | 102600 |
| 512.7 | Shortness of breath | -1 | 1.12 (1.07 – 1.17) | 6.78E-06 | 108657 |
| 512.7 | Shortness of breath | 0 | 1.37 (1.31 – 1.43) | 6.18E-49 | 117551 |
| 512.7 | Shortness of breath | 1 | 1.61 (1.55 – 1.67) | 2.41E-134 | 124868 |
| 512.7 | Shortness of breath | 2 | 1.59 (1.53 – 1.65) | 1.95E-135 | 127897 |
| 512.7 | Shortness of breath | 3 | 1.57 (1.52 – 1.63) | 4.76E-134 | 130482 |
| 512.7 | Shortness of breath | 4 | 1.54 (1.49 – 1.59) | 1.98E-131 | 132711 |
| 512.7 | Shortness of breath | 5 | 1.51 (1.46 – 1.56) | 7.56E-128 | 134656 |
| 798 | Malaise and fatigue | -5 | 1.00 (0.95 – 1.06) | 9.48E-01 | 85312 |
| 798 | Malaise and fatigue | -4 | 1.03 (0.98 – 1.08) | 2.19E-01 | 90932 |
| 798 | Malaise and fatigue | -3 | 1.07 (1.03 – 1.12) | 2.45E-03 | 96622 |
| 798 | Malaise and fatigue | -2 | 1.10 (1.05 – 1.15) | 2.10E-05 | 102600 |
| 798 | Malaise and fatigue | -1 | 1.16 (1.11 – 1.20) | 1.81E-12 | 108657 |
| 798 | Malaise and fatigue | 0 | 1.31 (1.26 – 1.36) | 6.03E-49 | 117551 |
| 798 | Malaise and fatigue | 1 | 1.47 (1.42 – 1.52) | 2.94E-113 | 124868 |
| 798 | Malaise and fatigue | 2 | 1.50 (1.45 – 1.55) | 1.79E-132 | 127897 |
| 798 | Malaise and fatigue | 3 | 1.50 (1.45 – 1.55) | 2.45E-138 | 130482 |
| 798 | Malaise and fatigue | 4 | 1.49 (1.45 – 1.54) | 1.16E-143 | 132711 |
| 798 | Malaise and fatigue | 5 | 1.48 (1.44 – 1.53) | 5.16E-146 | 134656 |

#### Table S7. Relative temporal trends of lipid and red blood cell-related laboratory associations with OSA.

Median values for years relative to the first OSA diagnosis (or equivalent age in controls) were obtained. All available results were used, with the proportion of cases and controls with data varying by year. Analyses adjusted for age, sex, BMI, population, and cumulative clinical encounter counts through the given year. Age was specific to the relative year, and the closest two BMIs to this relative date were averaged. The results are grouped by red blood cell then by lipid analysis and ordered by significance ten years prior to the first OSA diagnosis date.

| Lab | Years to OSA Diagnosis | Cases percentage with lab in year | Controls percentage with lab in year | Risk ratio | P-value | N |
|---|---|---|---|---|---|---|
| Red cell distribution width | -10 | 12.7 | 13.7 | 0.89 (0.87 – 0.91) | 3.50E-25 | 19303 |
| Red cell distribution width | -9 | 14.7 | 15.2 | 0.91 (0.90 – 0.93) | 5.47E-19 | 21752 |
| Red cell distribution width | -8 | 16.3 | 16.6 | 0.92 (0.90 – 0.93) | 1.24E-19 | 23880 |
| Red cell distribution width | -7 | 18.7 | 18.3 | 0.93 (0.92 – 0.95) | 7.81E-17 | 26721 |
| Red cell distribution width | -6 | 20.9 | 19.7 | 0.93 (0.92 – 0.94) | 5.34E-21 | 29036 |
| Red cell distribution width | -5 | 23.1 | 21.0 | 0.94 (0.93 – 0.96) | 2.93E-16 | 31454 |
| Red cell distribution width | -4 | 25.5 | 22.4 | 0.94 (0.93 – 0.95) | 6.14E-20 | 33926 |
| Red cell distribution width | -3 | 28.7 | 23.7 | 0.94 (0.93 – 0.95) | 7.12E-26 | 36677 |
| Red cell distribution width | -2 | 32.1 | 25.4 | 0.95 (0.94 – 0.96) | 5.97E-20 | 40018 |
| Red cell distribution width | -1 | 36.0 | 26.4 | 0.96 (0.95 – 0.97) | 2.76E-16 | 42799 |
| Red cell distribution width | 0 | 55.0 | 27.0 | 1.00 (0.99 – 1.01) | 0.86 | 52541 |
| Red cell distribution width | 1 | 50.7 | 26.3 | 1.03 (1.03 – 1.04) | 8.13E-30 | 49793 |
| Red cell distribution width | 2 | 41.5 | 25.0 | 0.99 (0.98 – 1.00) | 0.01 | 44133 |
| Red cell distribution width | 3 | 35.5 | 23.4 | 0.99 (0.98 – 0.99) | 0 | 39627 |
| Red cell distribution width | 4 | 30.7 | 21.9 | 0.98 (0.97 – 0.99) | 0 | 35929 |
| Red cell distribution width | 5 | 25.9 | 19.8 | 0.98 (0.97 – 0.99) | 0 | 31540 |
| Hematocrit | -10 | 13.7 | 14.7 | 1.03 (1.02 – 1.03) | 3.88E-23 | 20754 |
| Hematocrit | -9 | 15.7 | 16.2 | 1.02 (1.01 – 1.02) | 9.51E-15 | 23167 |
| Hematocrit | -8 | 17.3 | 17.4 | 1.03 (1.02 – 1.03) | 3.03E-32 | 25174 |
| Hematocrit | -7 | 19.7 | 19.1 | 1.02 (1.02 – 1.03) | 9.25E-29 | 27934 |
| Hematocrit | -6 | 21.8 | 20.3 | 1.03 (1.02 – 1.03) | 1.13E-38 | 30147 |
| Hematocrit | -5 | 24.0 | 21.7 | 1.03 (1.02 – 1.03) | 8.36E-44 | 32482 |
| Hematocrit | -4 | 26.4 | 23.0 | 1.03 (1.02 – 1.03) | 4.33E-49 | 34899 |
| Hematocrit | -3 | 29.5 | 24.2 | 1.03 (1.02 – 1.03) | 4.32E-61 | 37543 |
| Hematocrit | -2 | 32.9 | 25.8 | 1.02 (1.02 – 1.03) | 1.18E-57 | 40760 |
| Hematocrit | -1 | 36.7 | 26.7 | 1.02 (1.02 – 1.03) | 2.02E-62 | 43465 |

| Lab | Years to OSA Diagnosis | Cases percentage with lab in year | Controls percentage with lab in year | Risk ratio | P-value | N |
|---|---|---|---|---|---|---|
| Hematocrit | 0 | 55.7 | 27.3 | 1.01 (1.01 – 1.01) | 6.68E-23 | 53187 |
| Hematocrit | 1 | 51.3 | 26.5 | 0.98 (0.98 – 0.99) | 5.72E-75 | 50345 |
| Hematocrit | 2 | 41.9 | 25.2 | 1.01 (1.00 – 1.01) | 7.81E-09 | 44521 |
| Hematocrit | 3 | 35.7 | 23.5 | 1.01 (1.01 – 1.01) | 6.89E-17 | 39925 |
| Hematocrit | 4 | 30.9 | 22.1 | 1.01 (1.01 – 1.01) | 6.84E-17 | 36181 |
| Hematocrit | 5 | 26.0 | 19.9 | 1.02 (1.01 – 1.02) | 7.12E-24 | 31741 |
| Hemoglobin | -10 | 13.6 | 14.6 | 1.07 (1.06 – 1.09) | 1.87E-21 | 20717 |
| Hemoglobin | -9 | 15.7 | 16.1 | 1.05 (1.04 – 1.06) | 1.01E-12 | 23107 |
| Hemoglobin | -8 | 17.3 | 17.4 | 1.07 (1.06 – 1.09) | 5.88E-28 | 25119 |
| Hemoglobin | -7 | 19.6 | 19.1 | 1.06 (1.05 – 1.08) | 7.43E-25 | 27877 |
| Hemoglobin | -6 | 21.8 | 20.3 | 1.08 (1.06 – 1.09) | 3.31E-38 | 30088 |
| Hemoglobin | -5 | 24.0 | 21.6 | 1.08 (1.07 – 1.09) | 2.05E-44 | 32421 |
| Hemoglobin | -4 | 26.4 | 23.0 | 1.07 (1.06 – 1.08) | 7.43E-47 | 34846 |
| Hemoglobin | -3 | 29.5 | 24.1 | 1.07 (1.06 – 1.08) | 1.34E-54 | 37501 |
| Hemoglobin | -2 | 32.8 | 25.8 | 1.07 (1.06 – 1.08) | 2.87E-54 | 40707 |
| Hemoglobin | -1 | 36.6 | 26.7 | 1.07 (1.06 – 1.07) | 1.13E-62 | 43420 |
| Hemoglobin | 0 | 55.7 | 27.3 | 1.02 (1.02 – 1.03) | 8.57E-18 | 53151 |
| Hemoglobin | 1 | 51.2 | 26.5 | 0.95 (0.95 – 0.96) | 4.38E-86 | 50260 |
| Hemoglobin | 2 | 41.9 | 25.2 | 1.02 (1.01 – 1.02) | 1.57E-06 | 44482 |
| Hemoglobin | 3 | 35.7 | 23.5 | 1.03 (1.02 – 1.03) | 2.86E-14 | 39890 |
| Hemoglobin | 4 | 30.9 | 22.1 | 1.03 (1.02 – 1.04) | 1.23E-16 | 36149 |
| Hemoglobin | 5 | 26.0 | 19.9 | 1.04 (1.04 – 1.05) | 9.35E-25 | 31709 |
| Red blood cell count | -10 | 13.6 | 14.6 | 1.20 (1.16 – 1.25) | 9.11E-19 | 20664 |
| Red blood cell count | -9 | 15.6 | 16.1 | 1.16 (1.11 – 1.20) | 1.53E-13 | 23073 |
| Red blood cell count | -8 | 17.2 | 17.4 | 1.23 (1.19 – 1.28) | 4.94E-30 | 25064 |
| Red blood cell count | -7 | 19.6 | 19.0 | 1.20 (1.16 – 1.24) | 1.49E-26 | 27817 |
| Red blood cell count | -6 | 21.7 | 20.3 | 1.21 (1.17 – 1.25) | 1.98E-32 | 30024 |
| Red blood cell count | -5 | 23.9 | 21.6 | 1.23 (1.19 – 1.27) | 1.21E-43 | 32340 |
| Red blood cell count | -4 | 26.3 | 22.9 | 1.22 (1.19 – 1.25) | 2.00E-45 | 34761 |
| Red blood cell count | -3 | 29.4 | 24.1 | 1.23 (1.20 – 1.26) | 2.08E-55 | 37408 |
| Red blood cell count | -2 | 32.7 | 25.7 | 1.20 (1.17 – 1.23) | 2.05E-52 | 40622 |
| Red blood cell count | -1 | 36.5 | 26.7 | 1.19 (1.17 – 1.22) | 3.05E-57 | 43337 |
| Red blood cell count | 0 | 55.6 | 27.2 | 1.08 (1.06 – 1.10) | 4.43E-22 | 53057 |

| Lab | Years to OSA Diagnosis | Cases percentage with lab in year | Controls percentage with lab in year | Risk ratio | P-value | N |
|---|---|---|---|---|---|---|
| Red blood cell count | 1 | 51.1 | 26.5 | 0.89 (0.88 – 0.91) | 1.11E-53 | 50179 |
| Red blood cell count | 2 | 41.8 | 25.2 | 1.06 (1.04 – 1.08) | 6.68E-10 | 44416 |
| Red blood cell count | 3 | 35.6 | 23.5 | 1.10 (1.07 – 1.12) | 8.92E-19 | 39830 |
| Red blood cell count | 4 | 30.8 | 22.0 | 1.11 (1.09 – 1.14) | 1.81E-21 | 36102 |
| Red blood cell count | 5 | 26.0 | 19.9 | 1.15 (1.12 – 1.18) | 2.39E-30 | 31669 |
| Mean corpuscular hemoglobin concentration | -10 | 13.6 | 14.6 | 1.01 (0.99 – 1.03) | 0.51 | 20660 |
| Mean corpuscular hemoglobin concentration | -9 | 15.6 | 16.1 | 1.00 (0.98 – 1.02) | 0.89 | 23066 |
| Mean corpuscular hemoglobin concentration | -8 | 17.2 | 17.4 | 1.00 (0.98 – 1.02) | 0.97 | 25059 |
| Mean corpuscular hemoglobin concentration | -7 | 19.6 | 19.0 | 1.00 (0.98 – 1.01) | 0.64 | 27812 |
| Mean corpuscular hemoglobin concentration | -6 | 21.7 | 20.3 | 1.02 (1.01 – 1.03) | 0 | 30033 |
| Mean corpuscular hemoglobin concentration | -5 | 23.9 | 21.6 | 1.02 (1.01 – 1.03) | 0 | 32340 |
| Mean corpuscular hemoglobin concentration | -4 | 26.3 | 22.9 | 1.02 (1.01 – 1.03) | 0 | 34766 |
| Mean corpuscular hemoglobin concentration | -3 | 29.4 | 24.1 | 1.01 (0.99 – 1.02) | 0.33 | 37402 |
| Mean corpuscular hemoglobin concentration | -2 | 32.7 | 25.7 | 1.01 (1.00 – 1.02) | 0.01 | 40617 |
| Mean corpuscular hemoglobin concentration | -1 | 36.5 | 26.7 | 1.02 (1.01 – 1.03) | 1.03E-05 | 43341 |
| Mean corpuscular hemoglobin concentration | 0 | 55.6 | 27.2 | 1.00 (0.99 – 1.00) | 0.71 | 53059 |
| Mean corpuscular hemoglobin concentration | 1 | 51.1 | 26.5 | 0.97 (0.96 – 0.97) | 7.44E-20 | 50180 |
| Mean corpuscular hemoglobin concentration | 2 | 41.8 | 25.2 | 0.99 (0.98 – 1.00) | 0.03 | 44415 |
| Mean corpuscular hemoglobin concentration | 3 | 35.6 | 23.5 | 1.00 (0.99 – 1.00) | 0.4 | 39830 |
| Mean corpuscular hemoglobin concentration | 4 | 30.8 | 22.0 | 1.01 (1.00 – 1.02) | 0.04 | 36104 |
| Mean corpuscular hemoglobin concentration | 5 | 26.0 | 19.9 | 1.02 (1.01 – 1.03) | 0 | 31674 |
| Mean corpuscular hemoglobin | -10 | 13.6 | 14.6 | 1.00 (0.99 – 1.01) | 0.63 | 20657 |
| Mean corpuscular hemoglobin | -9 | 15.6 | 16.1 | 1.00 (0.99 – 1.01) | 0.69 | 23064 |
| Mean corpuscular hemoglobin | -8 | 17.2 | 17.4 | 1.00 (0.99 – 1.00) | 0.39 | 25054 |
| Mean corpuscular hemoglobin | -7 | 19.6 | 19.0 | 1.00 (0.99 – 1.00) | 0.46 | 27809 |
| Mean corpuscular hemoglobin | -6 | 21.7 | 20.3 | 1.00 (1.00 – 1.01) | 0.18 | 30024 |
| Mean corpuscular hemoglobin | -5 | 23.9 | 21.6 | 1.00 (0.99 – 1.01) | 0.81 | 32337 |
| Mean corpuscular hemoglobin | -4 | 26.3 | 22.9 | 1.00 (0.99 – 1.01) | 0.74 | 34760 |
| Mean corpuscular hemoglobin | -3 | 29.4 | 24.1 | 1.00 (0.99 – 1.01) | 0.97 | 37402 |
| Mean corpuscular hemoglobin | -2 | 32.7 | 25.7 | 1.00 (1.00 – 1.01) | 0.67 | 40621 |
| Mean corpuscular hemoglobin | -1 | 36.5 | 26.7 | 1.00 (1.00 – 1.01) | 0.44 | 43335 |
| Mean corpuscular hemoglobin | 0 | 55.6 | 27.2 | 1.00 (0.99 – 1.00) | 0.1 | 53057 |
| Mean corpuscular hemoglobin | 1 | 51.1 | 26.5 | 0.99 (0.98 – 0.99) | 6.75E-13 | 50178 |

| Lab | Years to OSA Diagnosis | Cases percentage with lab in year | Controls percentage with lab in year | Risk ratio | P-value | N |
|---|---|---|---|---|---|---|
| Mean corpuscular hemoglobin | 2 | 41.8 | 25.2 | 0.99 (0.99 – 1.00) | 0.01 | 44416 |
| Mean corpuscular hemoglobin | 3 | 35.6 | 23.5 | 0.99 (0.99 – 1.00) | 0.01 | 39830 |
| Mean corpuscular hemoglobin | 4 | 30.8 | 22.0 | 0.99 (0.99 – 1.00) | 0.02 | 36103 |
| Mean corpuscular hemoglobin | 5 | 26.0 | 19.9 | 0.99 (0.99 – 1.00) | 0.02 | 31669 |
| Mean corpuscular volume | -10 | 13.6 | 14.6 | 1.00 (1.00 – 1.00) | 0.85 | 20664 |
| Mean corpuscular volume | -9 | 15.6 | 16.1 | 1.00 (1.00 – 1.00) | 0.52 | 23074 |
| Mean corpuscular volume | -8 | 17.2 | 17.4 | 1.00 (1.00 – 1.00) | 0.28 | 25064 |
| Mean corpuscular volume | -7 | 19.6 | 19.0 | 1.00 (1.00 – 1.00) | 0.44 | 27816 |
| Mean corpuscular volume | -6 | 21.7 | 20.3 | 1.00 (1.00 – 1.00) | 0.93 | 30024 |
| Mean corpuscular volume | -5 | 23.9 | 21.6 | 1.00 (0.99 – 1.00) | 0.03 | 32341 |
| Mean corpuscular volume | -4 | 26.3 | 22.9 | 1.00 (1.00 – 1.00) | 0.24 | 34761 |
| Mean corpuscular volume | -3 | 29.4 | 24.1 | 1.00 (1.00 – 1.00) | 0.63 | 37407 |
| Mean corpuscular volume | -2 | 32.7 | 25.7 | 1.00 (1.00 – 1.00) | 0.38 | 40622 |
| Mean corpuscular volume | -1 | 36.5 | 26.7 | 1.00 (1.00 – 1.00) | 0.18 | 43337 |
| Mean corpuscular volume | 0 | 55.6 | 27.2 | 1.00 (1.00 – 1.00) | 0.31 | 53057 |
| Mean corpuscular volume | 1 | 51.1 | 26.5 | 1.00 (1.00 – 1.00) | 0 | 50179 |
| Mean corpuscular volume | 2 | 41.8 | 25.2 | 1.00 (1.00 – 1.00) | 0.14 | 44416 |
| Mean corpuscular volume | 3 | 35.6 | 23.5 | 1.00 (1.00 – 1.00) | 0.01 | 39830 |
| Mean corpuscular volume | 4 | 30.8 | 22.0 | 1.00 (0.99 – 1.00) | 0 | 36102 |
| Mean corpuscular volume | 5 | 26.0 | 19.9 | 0.99 (0.99 – 1.00) | 4.91E-06 | 31669 |
| LDL cholesterol | -10 | 4.6 | 6.5 | 1.00 (1.00 – 1.00) | 0.05 | 8525 |
| LDL cholesterol | -9 | 5.2 | 7.1 | 1.00 (1.00 – 1.00) | 0.06 | 9356 |
| LDL cholesterol | -8 | 5.7 | 7.6 | 1.00 (1.00 – 1.00) | 2.66E-05 | 10116 |
| LDL cholesterol | -7 | 6.5 | 8.1 | 1.00 (1.00 – 1.00) | 0 | 11003 |
| LDL cholesterol | -6 | 7.1 | 8.6 | 1.00 (1.00 – 1.00) | 0 | 11771 |
| LDL cholesterol | -5 | 8.1 | 9.1 | 1.00 (1.00 – 1.00) | 0 | 12673 |
| LDL cholesterol | -4 | 9.1 | 9.5 | 1.00 (1.00 – 1.00) | 0.01 | 13604 |
| LDL cholesterol | -3 | 10.3 | 10.1 | 1.00 (1.00 – 1.00) | 0.26 | 14764 |
| LDL cholesterol | -2 | 12.0 | 10.9 | 1.00 (1.00 – 1.00) | 0.61 | 16305 |
| LDL cholesterol | -1 | 14.0 | 11.3 | 1.00 (1.00 – 1.00) | 0.05 | 17663 |
| LDL cholesterol | 0 | 21.3 | 12.0 | 1.00 (1.00 – 1.00) | 0.96 | 21807 |
| LDL cholesterol | 1 | 20.1 | 12.4 | 1.00 (1.00 – 1.00) | 5.62E-14 | 21669 |
| LDL cholesterol | 2 | 20.3 | 13.0 | 1.00 (1.00 – 1.00) | 0 | 22310 |

| Lab | Years to OSA Diagnosis | Cases percentage with lab in year | Controls percentage with lab in year | Risk ratio | P-value | N |
|---|---|---|---|---|---|---|
| LDL cholesterol | 3 | 20.3 | 13.2 | 1.00 (1.00 – 1.00) | 7.40E-05 | 22472 |
| LDL cholesterol | 4 | 19.9 | 13.4 | 1.00 (1.00 – 1.00) | 1.15E-05 | 22545 |
| LDL cholesterol | 5 | 19.0 | 12.7 | 1.00 (1.00 – 1.00) | 0 | 21402 |
| Cholesterol | -10 | 11.7 | 11.9 | 1.00 (1.00 – 1.00) | 0.05 | 17149 |
| Cholesterol | -9 | 13.3 | 13.1 | 1.00 (1.00 – 1.00) | 0.21 | 19091 |
| Cholesterol | -8 | 15.0 | 14.2 | 1.00 (1.00 – 1.00) | 0.01 | 20933 |
| Cholesterol | -7 | 16.8 | 15.4 | 1.00 (1.00 – 1.00) | 0.01 | 22902 |
| Cholesterol | -6 | 18.7 | 16.2 | 1.00 (1.00 – 1.00) | 0.02 | 24690 |
| Cholesterol | -5 | 20.8 | 17.3 | 1.00 (1.00 – 1.00) | 0.41 | 26700 |
| Cholesterol | -4 | 23.1 | 18.1 | 1.00 (1.00 – 1.00) | 0.05 | 28575 |
| Cholesterol | -3 | 25.7 | 19.0 | 1.00 (1.00 – 1.00) | 0.46 | 30732 |
| Cholesterol | -2 | 29.0 | 20.2 | 1.00 (1.00 – 1.00) | 0.77 | 33473 |
| Cholesterol | -1 | 32.8 | 20.8 | 1.00 (1.00 – 1.00) | 0.11 | 35955 |
| Cholesterol | 0 | 46.7 | 21.7 | 1.00 (1.00 – 1.00) | 0.29 | 43443 |
| Cholesterol | 1 | 42.4 | 22.6 | 1.00 (1.00 – 1.00) | 9.16E-25 | 42275 |
| Cholesterol | 2 | 43.8 | 23.2 | 1.00 (1.00 – 1.00) | 1.72E-06 | 43512 |
| Cholesterol | 3 | 41.6 | 23.7 | 1.00 (1.00 – 1.00) | 5.80E-08 | 42871 |
| Cholesterol | 4 | 38.8 | 23.7 | 1.00 (1.00 – 1.00) | 1.15E-06 | 41625 |
| Cholesterol | 5 | 35.1 | 22.6 | 1.00 (1.00 – 1.00) | 2.61E-05 | 38742 |
| VLDL cholesterol | -10 | 3.5 | 2.8 | 1.00 (1.00 – 1.00) | 0.27 | 4375 |
| VLDL cholesterol | -9 | 3.9 | 3.1 | 1.00 (1.00 – 1.00) | 0.08 | 4851 |
| VLDL cholesterol | -8 | 4.4 | 3.4 | 1.00 (1.00 – 1.00) | 0.33 | 5365 |
| VLDL cholesterol | -7 | 4.8 | 3.7 | 1.00 (1.00 – 1.00) | 0.69 | 5860 |
| VLDL cholesterol | -6 | 5.2 | 3.9 | 1.00 (1.00 – 1.00) | 0.98 | 6253 |
| VLDL cholesterol | -5 | 5.7 | 4.0 | 1.00 (1.00 – 1.00) | 0.38 | 6666 |
| VLDL cholesterol | -4 | 6.2 | 4.4 | 1.00 (1.00 – 1.00) | 0.19 | 7226 |
| VLDL cholesterol | -3 | 6.8 | 4.5 | 1.00 (1.00 – 1.00) | 0.2 | 7657 |
| VLDL cholesterol | -2 | 7.8 | 4.8 | 1.00 (1.00 – 1.00) | 0.01 | 8424 |
| VLDL cholesterol | -1 | 8.6 | 5.1 | 1.00 (1.00 – 1.00) | 0.01 | 9037 |
| VLDL cholesterol | 0 | 12.6 | 5.2 | 1.00 (1.00 – 1.00) | 0.05 | 11076 |
| VLDL cholesterol | 1 | 11.4 | 5.4 | 1.00 (1.00 – 1.00) | 0 | 10757 |
| VLDL cholesterol | 2 | 11.6 | 5.6 | 1.00 (1.00 – 1.00) | 0.01 | 11014 |
| VLDL cholesterol | 3 | 10.7 | 5.8 | 1.00 (1.00 – 1.00) | 0 | 10732 |

| Lab | Years to OSA Diagnosis | Cases percentage with lab in year | Controls percentage with lab in year | Risk ratio | P-value | N |
|---|---|---|---|---|---|---|
| VLDL cholesterol | 4 | 9.7 | 5.8 | 1.00 (1.00 – 1.00) | 0.14 | 10266 |
| VLDL cholesterol | 5 | 8.6 | 5.5 | 1.00 (1.00 – 1.00) | 0 | 9447 |
| Triglycerides | -10 | 10.9 | 11.1 | 1.00 (1.00 – 1.00) | 0.51 | 15952 |
| Triglycerides | -9 | 12.5 | 12.4 | 1.00 (1.00 – 1.00) | 0.18 | 17963 |
| Triglycerides | -8 | 14.3 | 13.5 | 1.00 (1.00 – 1.00) | 0.32 | 19905 |
| Triglycerides | -7 | 16.0 | 14.7 | 1.00 (1.00 – 1.00) | 0.01 | 21885 |
| Triglycerides | -6 | 17.9 | 15.6 | 1.00 (1.00 – 1.00) | 0.37 | 23726 |
| Triglycerides | -5 | 20.0 | 16.7 | 1.00 (1.00 – 1.00) | 0.55 | 25770 |
| Triglycerides | -4 | 22.4 | 17.5 | 1.00 (1.00 – 1.00) | 0.08 | 27698 |
| Triglycerides | -3 | 25.0 | 18.5 | 1.00 (1.00 – 1.00) | 5.03E-05 | 29895 |
| Triglycerides | -2 | 28.3 | 19.7 | 1.00 (1.00 – 1.00) | 0 | 32668 |
| Triglycerides | -1 | 32.2 | 20.4 | 1.00 (1.00 – 1.00) | 1.41E-09 | 35248 |
| Triglycerides | 0 | 46.0 | 21.3 | 1.00 (1.00 – 1.00) | 1.44E-15 | 42781 |
| Triglycerides | 1 | 42.3 | 22.2 | 1.00 (1.00 – 1.00) | 0.01 | 41851 |
| Triglycerides | 2 | 43.3 | 23.0 | 1.00 (1.00 – 1.00) | 0 | 43063 |
| Triglycerides | 3 | 41.1 | 23.4 | 1.00 (1.00 – 1.00) | 7.77E-07 | 42446 |
| Triglycerides | 4 | 38.5 | 23.6 | 1.00 (1.00 – 1.00) | 2.96E-10 | 41275 |
| Triglycerides | 5 | 34.8 | 22.5 | 1.00 (1.00 – 1.00) | 1.07E-07 | 38453 |
| HDL cholesterol | -10 | 11.2 | 11.5 | 1.00 (1.00 – 1.00) | 0.6 | 16508 |
| HDL cholesterol | -9 | 12.9 | 12.8 | 1.00 (1.00 – 1.00) | 0.08 | 18516 |
| HDL cholesterol | -8 | 14.6 | 13.9 | 1.00 (1.00 – 1.00) | 0.05 | 20441 |
| HDL cholesterol | -7 | 16.4 | 15.0 | 1.00 (1.00 – 1.00) | 0 | 22419 |
| HDL cholesterol | -6 | 18.3 | 15.9 | 1.00 (1.00 – 1.00) | 0.04 | 24210 |
| HDL cholesterol | -5 | 20.4 | 17.0 | 1.00 (1.00 – 1.00) | 0 | 26289 |
| HDL cholesterol | -4 | 22.8 | 17.8 | 1.00 (1.00 – 1.00) | 0.01 | 28212 |
| HDL cholesterol | -3 | 25.4 | 18.8 | 1.00 (1.00 – 1.00) | 8.51E-06 | 30341 |
| HDL cholesterol | -2 | 28.7 | 20.0 | 1.00 (1.00 – 1.00) | 6.61E-06 | 33113 |
| HDL cholesterol | -1 | 32.5 | 20.7 | 1.00 (1.00 – 1.00) | 6.52E-06 | 35635 |
| HDL cholesterol | 0 | 46.4 | 21.5 | 1.00 (1.00 – 1.00) | 9.35E-15 | 43119 |
| HDL cholesterol | 1 | 42.1 | 22.4 | 0.99 (0.99 – 1.00) | 4.69E-45 | 41958 |
| HDL cholesterol | 2 | 43.5 | 23.1 | 1.00 (1.00 – 1.00) | 1.19E-17 | 43286 |
| HDL cholesterol | 3 | 41.4 | 23.5 | 1.00 (1.00 – 1.00) | 7.65E-20 | 42670 |
| HDL cholesterol | 4 | 38.7 | 23.7 | 1.00 (1.00 – 1.00) | 2.26E-12 | 41488 |

| Lab | Years to OSA Diagnosis | Cases percentage with lab in year | Controls percentage with lab in year | Risk ratio | P-value | N |
|---|---|---|---|---|---|---|
| HDL cholesterol | 5 | 35.0 | 22.5 | 1.00 (1.00 – 1.00) | 1.98E-10 | 38625 |

#### Table S8. Sleep-related note term frequencies.

Clinical note terms were extracted using cTAKES (PMC2995668) using a default clinical pipeline and a Unified Medical Language System (UMLS) ontology (PMC308795). “Prevalence” indicates the proportion of participants with at least one term occurrence in any note. Individual CUI concepts have one or more exact phrases in UMLS.

| CUI | Case prevalence | Control prevalence | Concept | UMLS Phrases |
|---|---|---|---|---|
| C0000981 | 2.9 | 1.2 | acetazolamide | 2-acetylamino-1,3,4-thiadiazole-5-sulfonamide\|5-acetamido-1,3,4-thiadiazole-2-sulfonamide\|5-acetylamino-1,3,4-thiadiazole-2-sulfonamide\|acetamide, n-(5-(aminosulfonyl)-1,3,4-thiadiazol-2-yl)-\|acetazolamid\|acetazolamida\|acetazolamide\|acetazolamide (medication)\|acetazolamide (product)\|acetazolamide (substance)\|acetazolamide - chemical\|acetazolamide - chemical (substance)\|acetazolamide [chemical/ingredient]\|acetazolamide [epilepsy]\|acetazolamide [epilepsy] (product)\|acetazolamide [epilepsy] (substance)\|acetazolamide [glaucoma]\|acetazolamide [glaucoma] (product)\|acetazolamide [glaucoma] (substance)\|acetazolamide product\|acetazolamide-containing product\|acetazolamides\|acetazolamidum\|acã©tazolamide\|n-(5-(aminosulfonyl)-1,3,4-thiadiazol-2-yl)acetamide\|n-[5-(aminosulfonyl)-1,3,4-thiadiazol-2-yl]acetamide\|n-[5-(aminosulfonyl)-1,3,5-thiadiazol-2-yl]acetamide\|product containing acetazolamide\|product containing acetazolamide (medicinal product) |
| C0008810 | 4.9 | 1.0 | circadian rhythm | circadian process\|circadian rhythm\|circadian rhythm (function)\|circadian rhythm, function\|circadian rhythm, function (observable entity)\|circadian rhythm, nos\|circadian rhythms\|daily biorhythm\|daily biorhythm, nos\|diurnal rhythm\|diurnal rhythms\|diurnal variation\|diurnal variations\|nycthemeral rhythm\|nycthemeral rhythms\|nyctohemeral rhythm\|nyctohemeral rhythms\|rhythm, circadian\|rhythm, nycthemeral\|rhythm, nyctohemeral\|rhythm, twenty-four hour\|rhythms, circadian\|rhythms, nycthemeral\|rhythms, nyctohemeral\|rhythms, twenty-four hour\|twenty four hour rhythm\|twenty-four hour rhythm\|twenty-four hour rhythms\|variation diurnal |
| C0013117 | 22.9 | 6.9 | dream | dream\|dream (function)\|dream, function\|dream, function (observable entity)\|dream, nos\|dreaming\|dreams\|dreamy |
| C0013144 | 79.4 | 36.1 | drowsiness | [d]drowsiness\|[d]drowsiness (context-dependent category)\|[d]drowsiness (situation)\|drowsiness\|drowsiness (function)\|drowsiness (symptom)\|drowsiness was observed\|drowsiness, function\|drowsiness, function (observable entity)\|drowsy\|drowsy (finding)\|drowsy (physical finding)\|mental status, drowsy\|sleepiness\|sleepy\|sleepy (finding)\|somnolence\|somnolence (sleepiness)\|somnolent |
| C0013819 | 26.9 | 8.6 | electroencephalography | eeg\|eeg (electrophysiology)\|eeg - electroencephalography\|eeg electroencephalography\|eeg, nos\|electrical tracings;neurolo\|electroencephalogr\|electroencephalogram\|electroencephalogram (eeg)\|electroencephalogram (procedure)\|electroencephalogram, nos\|electroencephalograms\|electroencephalography\|electroencephalography (eeg)\|electroencephalography (procedure)\|electroencephalography [eeg] |
| C0015672 | 94.1 | 78.6 | fatigue | [d]fatigue\|[d]fatigue (context-dependent category)\|[d]fatigue (situation)\|[d]tiredness\| |

| CUI | Case prevalence | Control prevalence | Concept | UMLS Phrases |
|---|---|---|---|---|
| | | | | [d]tiredness (context-dependent category)\|[d]tiredness (situation)\|decrease in energy\|decreased energy\|energy decreased\|energy loss\|extreme fatigue\|fatigue\|fatigue (diagnosis)\|fatigue (finding)\|fatigue [disease/finding]\|fatigue extreme\|fatigue nos\|fatigued\|fatigues\|fatiguing\|feeling of total lack of energy\|lack (of);energy\|lack of energy\|lack of energy (finding)\|lacking energy\|lacking in energy\|lacking in energy (finding)\|loss of energy\|rndx fatigue\|rndx fatigue (diagnosis)\|tatt\|time tired\|tired\|tired all the time\|tired out\|tired time\|tiredness\|tiredness (finding)\|weariness |
| C0028081 | 48.9 | 33.2 | nights sweats | night sweat\|night sweating\|night sweats\|night sweats (finding)\|nights sweats\|nocturnal hyperhidrosis\|nocturnal perspiration\|nocturnal; sweating\|problem;sweating;night\|sweat; night sweats\|sweating heavily at night\|sweating heavily at night (symptom)\|sweating; night |
| C0028084 | 14.2 | 6.1 | nightmares | [x]nightmares\|frightening dreams\|nightmare\|nightmares\|nightmares (finding)\|nightmares (symptom)\|terrifying dreams |
| C0028734 | 49.0 | 29.6 | nocturia | [d]nocturia\|[d]nocturia (context-dependent category)\|[d]nocturia (situation)\|finding of nocturia\|finding of nocturia (finding)\|nocturia\|nocturia (finding)\|nocturia - finding\|nocturia [disease/finding]\|nocturnal frequency\|nycturia\|urinary frequency more than once at night\|urinary frequency more than once at night (nocturia)\|urinary frequency more than once at night (symptom) |
| C0033139 | 5.6 | 1.7 | primary insomnia | insomnia, primary\|insomnia; primary\|primary insomnia\|primary insomnia (diagnosis)\|primary insomnia (disorder)\|primary; insomnia |
| C0037313 | 99.9 | 76.0 | sleep | sleep\|sleep (biological function) (observable entity)\|sleep (biological function) [ambiguous]\|sleep (qualifier value)\|sleep functions\|sleep functions, unspecified\|sleep, function\|sleep, function (observable entity)\|sleep, nos\|sleeping\|sleeps |
| C0037315 | 99.1 | 21.7 | sleep apnea syndrome | [d]sleep apnea syndrome\|[d]sleep apnea syndrome (context-dependent category)\|[d]sleep apnea syndrome (situation)\|[d]sleep apnoea syndrome\|[d]syndrome sleep apnea\|[d]syndrome sleep apnoea\|apnea syndrome\|apnea syndrome, sleep\|apnea syndromes, sleep\|apnea, sleep\|apnea; sleep\|apneas, sleep\|apnoea syndrome\|hypopnea, sleep\|hypopneas, sleep\|periods of not breathing while asleep\|periods of not breathing while asleep (sleep apnea)\|periods of not breathing while asleep (symptom)\|sas - sleep apnea syndrome\|sas - sleep apnoea syndrome\|sleep apnea\|sleep apnea (disorder)\|sleep apnea (finding)\|sleep apnea nos\|sleep apnea syndrome\|sleep apnea syndromes\|sleep apnea syndromes [disease/finding]\|sleep apnea, nos\|sleep apnea, unspecified\|sleep apneas\|sleep apnoea\|sleep apnoea (disorder)\|sleep apnoea syndrome\|sleep apnoea syndromes\|sleep apnoea, nos\|sleep apnoea, unspecified\|sleep apnoeas\|sleep hypopnea\|sleep hypopnea (diagnosis)\|sleep hypopneas\|sleep hypopnoea\|sleep; apnea\|syndrome sleep apnea\|syndrome sleep apnoea\|unspecified sleep apnea |
| C0037316 | 7.5 | 1.9 | sleep deprivation | deprivation, sleep\|deprivations, sleep\|lack of adequate sleep\|not enough sleeping\|rndx sleep deprivation\|rndx sleep deprivation (diagnosis)\|sleep deficit\|sleep deprivation\|sleep deprivation (finding)\|sleep deprivation [disease/finding]\|sleep |

| CUI | Case prevalence | Control prevalence | Concept | UMLS Phrases |
|---|---|---|---|---|
| | | | | deprivations |
| C0037317 | 57.2 | 32.4 | sleep disturbances | [d]sleep disturbance, unspecif\|[d]sleep disturbance, unspecified\|[d]sleep disturbance, unspecified (context-dependent category)\|[d]sleep disturbance, unspecified (situation)\|[d]sleep disturbances\|[d]sleep disturbances (context-dependent category)\|[d]sleep disturbances (situation)\|difficulty sleeping\|disturbance in sleep behavior\|disturbance in sleep behavior (finding)\|disturbance in sleep behavior, nos\|disturbance in sleep behaviour\|disturbance in sleep behaviour, nos\|disturbances, sleep\|disturbed sleep\|disturbed;sleep\|problems with sleeping\|sleep disturbance\|sleep disturbance nos\|sleep disturbance, unspecif\|sleep disturbance, unspecified\|sleep disturbances\|sleep disturbances (incl subtypes)\|sleep disturbances (including subtypes)\|sleep disturbances (symptom)\|sleep disturbances [d]\|sleep disturbed\|sleep dysfunction\|sleeping disturbances\|trouble sleeping\|unspecified sleep disturbance |
| C0037384 | 77.5 | 17.3 | snoring | [d]snoring\|[d]snoring (context-dependent category)\|[d]snoring (situation)\|finding of snoring\|finding of snoring (finding)\|observation of snoring\|snore\|snored\|snores\|snoring\|snoring (finding)\|snoring (physical finding)\|snoring (symptom)\|snoring [disease/finding]\|snoring symptom\|snoring symptom nos\|snoring symptom nos (context-dependent category)\|snoring symptom nos (finding)\|snoring symptoms\|snoring symptoms (context-dependent category)\|snoring symptoms (finding)\|snoring was observed\|snorings |
| C0040805 | 26.4 | 15.5 | trazodone | 1,2,4-triazolo(4,3-a)pyridin-3(2h)-one, 2-(3-(4-(3-chlorophenyl)-1-piperazinyl)propyl)-\|2-(3-[4-(3-chlorophenyl)-1-piperazinyl]propyl)[1,2,4]triazolo[4,3-a]pyridin-3(2h)-one\|antidepressants trazodone\|product containing trazodone\|product containing trazodone (medicinal product)\|searle brand of trazodone hydrochloride\|tradozone\|trazodona\|trazodone\|trazodone (medication)\|trazodone (product)\|trazodone (substance)\|trazodone [chemical/ingredient]\|trazodone product\|trazodone-containing product\|trazodonum |
| C0043012 | 18.7 | 6.5 | wakefulness | aroused\|arousing\|vigilance\|vigilant\|wakefulness\|wakefulness (observable entity)\|wakefulnesses\|watchfulness |
| C0043387 | 3.8 | 1.7 | yawning | [d]yawning\|[d]yawning (context-dependent category)\|[d]yawning (situation)\|finding of yawning\|finding of yawning (finding)\|observation of yawning\|yawn\|yawning\|yawning (finding)\|yawning (function)\|yawning (symptom)\|yawning, function\|yawning, function (observable entity)\|yawnings\|yawns |
| C0078839 | 15.5 | 7.8 | zolpidem | hypnotics zolpidem\|imidazo(1,2-a)pyridine-3-acetamide, n,n,6-trimethyl-2-(4-methylphenyl)-\|n,n,6-trimethyl-2-(4-methylphenyl)imidazo(1,2-a)pyridine-3-acetamide\|n,n,6-trimethyl-2-(4-methylphenyl)imidazo(1,2a)pyridine-3-acetamide hemitartrate\|product containing zolpidem\|product containing zolpidem (medicinal product)\|zolpidem\|zolpidem (medication)\|zolpidem (product)\|zolpidem (substance)\|zolpidem [chemical/ingredient]\|zolpidem product\|zolpidem-containing product\|zolpidemum\|zpd |
| C0151611 | 2.2 | 1.1 | abnormal | [d]eeg/electroencephalogr.abn.\|[d]electroencephalogram (eeg) abnormal\| |

| CUI | Case prevalence | Control prevalence | Concept | UMLS Phrases |
|---|---|---|---|---|
| | | | electroencephalogram | [d]electroencephalogram (eeg) abnormal (context-dependent category)\|[d]electroencephalogram (eeg) abnormal (situation)\|abnormal eeg\|abnormal electroencephalogram\|abnormal electroencephalogram [eeg]\|abnormal; electroencephalogram\|abnormality eeg\|disorder eeg (nos)\|eeg abnormal\|eeg abnormalities\|eeg abnormality\|eeg abnormality (finding)\|eeg abnormality, nos\|eeg/electroencephalogr.abn.\|electroencephalogram (eeg) abnormal\|electroencephalogram (eeg) abnormalities\|electroencephalogram abnormal\|electroencephalogram abnormal (finding)\|electroencephalogram abnormal (situation)\|electroencephalogram abnormalities\|electroencephalogram abnormality\|electroencephalogram abnormality (finding)\|electroencephalogram; abnormal |
| C0162701 | 86.3 | 11.8 | polysomnography | 2-95 sleep disorder tests and procedures\|full sleep study\|monitor sleep\|monitoring, sleep\|polysomnogr\|polysomnogram\|polysomnogram, nos\|polysomnograms\|polysomnographies\|polysomnography\|polysomnography (procedure)\|polysomnography, nos\|psg\|sleep disorder tests and procedures\|sleep measurement\|sleep monitoring\|sleep recording\|sleep studies\|sleep studies (procedure)\|sleep study\|sleep study (procedure)\|sleep test\|sleep testing\|sleep testing (procedure)\|sleep tests\|sleeping studying\|somnogr\|somnographies\|somnography\|study sleep |
| C0199451 | 93.5 | 14.0 | continuous positive airway pressure | cont pos airways pres therapy\|cont positive airways pressure\|continuous cpap\|continuous cpap (procedure)\|continuous cpap (regime/therapy)\|continuous positive airway pressure\|continuous positive airway pressure (cpap)\|continuous positive airway pressure [cpap]\|continuous positive airway pressure ventilation\|continuous positive airway pressure ventilation (treatment)\|continuous positive airway pressure ventilation treatment\|continuous positive airway pressure ventilation treatment (procedure)\|continuous positive airway pressure ventilation treatment (regime/therapy)\|continuous positive airways pressure\|continuous positive airways pressure (observable entity)\|continuous positive airways pressure therapy\|cpap\|cpap (continuous positive airway pressure ventilation)\|cpap (continuous positive airway pressure)\|cpap - cont pos air pres ther\|cpap - continuous positive airways pressure\|cpap - continuous positive airways pressure therapy\|cpap treatment\|cpap ventilation\|pressure.continuous positive airway\|ventilation, cpap |
| C0234255 | 8.1 | 5.3 | night pain | night pain\|night pain (finding)\|night pains\|nyctalgia\|pain night |
| C0234422 | 79.5 | 62.2 | awake | awake\|awake (finding)\|awakes\|clear consciousness\|wide awake |
| C0235162 | 37.9 | 13.1 | difficulty sleeping | difficult sleeping\|difficulties sleep\|difficulties sleeping\|difficulty sleep\|difficulty sleeping\|difficulty sleeping (finding)\|poor sleep\|sleep difficult\|sleep difficulties\|sleeping difficulties\|sleeping difficulty |
| C0393760 | 31.2 | 6.9 | difficulty falling asleep | cannot get off to sleep\|difficulty falling asleep\|difficulty falling asleep (symptom)\|difficulty getting to sleep\|difficulty in sleep initiation\|initial insomnia\|initial insomnia (disorder)\|initial insomnia (finding)\|insomnia, vesperal\|trouble falling asleep\|vesperal insomnia |
| C0393761 | 5.3 | 1.6 | difficulty staying asleep | broken sleep\|difficulty in sleep maintenanc\|difficulty in sleep maintenance\|difficulty |

| CUI | Case prevalence | Control prevalence | Concept | UMLS Phrases |
|---|---|---|---|---|
| | | | | staying asleep\|fitful sleep\|interrupted sleep\|interrupting sleep\|keeps waking up\|middle insomnia\|middle insomnia (disorder)\|middle insomnia (finding)\|night wake\|night wakes\|night waking\|sleep maintenance insomnia\|waking night |
| C0424522 | 81.2 | 42.8 | sleeping | activity, sleeping\|asleep\|asleep (finding)\|sleeping |
| C0424563 | 21.1 | 2.5 | sleep quality | quality of sleep\|quality of sleep (finding)\|quality of sleep (observable entity)\|sleep quality |
| C0424565 | 8.8 | 4.7 | cannot sleep | cannot sleep\|cannot sleep at all\|cannot sleep at all (finding)\|inability to sleep\|unable (to);sleep\|unable to sleep |
| C0424583 | 3.0 | 1.4 | not tired | not tired\|not tired (context-dependent category)\|not tired (finding)\|not tired (situation) |
| C0439055 | 5.8 | 1.7 | tired all the time | "tatt" - tired all the time\|c/o - "tired all the time"\|c/o - "tired all the time" (context-dependent category)\|complaining of "tired all the time"\|complaining of "tired all the time" (finding)\|tatt\|tatt - tired all the time\|tired all the time\|tired all the time (& [c/o -])\|tired all the time (finding) |
| C0474396 | 14.1 | 4.0 | sleep behavior | behavior sleep\|behaviors sleep\|behaviour sleeping\|pattern sleep\|patterns sleep\|patterns sleeping\|sleep behavior\|sleep behavior (observable entity)\|sleep behavior finding\|sleep behavior finding (finding)\|sleep behavior observations\|sleep behavior, nos\|sleep behaviour\|sleep behaviour finding\|sleep behaviour observations\|sleep behaviour, nos\|sleep pattern\|sleep pattern (finding)\|sleep pattern (history)\|sleep pattern finding\|sleep pattern finding (finding)\|sleeping pattern |
| C0520679 | 96.9 | 11.9 | obstructive sleep apnea | apnea obstructive sleep\|apnea obstructive sleeping\|apnea, obstructive\|apnea, obstructive sleep\|apneas, obstructive sleep\|obstructive apnea\|obstructive sleep apnea\|obstructive sleep apnea (adult) (pediatric)\|obstructive sleep apnea (adult) (pediatric)\|obstructive sleep apnea (diagnosis)\|obstructive sleep apnea syndrome\|obstructive sleep apnea syndrome (disorder)\|obstructive sleep apneas\|obstructive sleep apnoea\|obstructive sleep apnoea (disorder)\|obstructive sleep apnoea syndrome\|obstructive sleep osa apnea\|osa\|osa - obstructive sleep apnea\|osa - obstructive sleep apnoea\|osahs\|osas\|sahs\|sleep apnea (& [obstructive])\|sleep apnea hypopnea syndrome\|sleep apnea obstructive syndrome\|sleep apnea syndrome, obstructive\|sleep apnea, obstructive\|sleep apnea, obstructive [disease/finding]\|sleep apnea/hypopnea syndrome\|sleep apneas, obstructive\|sleep apnoea (& [obstructive])\|sleep apnoea (& [obstructive]) (disorder)\|syndrome, obstructive sleep apnea\|syndrome, sleep apnea, obstructive |
| C0557875 | 75.7 | 46.0 | tired | tired\|tired (finding) |
| C0700201 | 5.4 | 1.0 | dyssomnia | dyssomnia\|dyssomnia (disorder)\|dyssomnia nos\|dyssomnia, nos\|dyssomnias\|dyssomnias [disease/finding]\|sleep disturbance\|sleep problem |
| C0718303 | 4.2 | 1.8 | sleep aid | aid sleep\|aid sleeping\|aid to sleep\|aiding sleep\|aids sleep\|aids sleeping\|sleep aid\|to aid sleep |
| C0722882 | 5.7 | 1.1 | modafinil | modafinil (provigil)\|provigil |
| C0723446 | 4.9 | 1.4 | stay awake | awake stay\|awake staying\|stay awake |

| CUI | Case prevalence | Control prevalence | Concept | UMLS Phrases |
|---|---|---|---|---|
| C0851578 | 36.8 | 3.6 | sleep disorder | 9-74 sleep disorders\|[d]sleep dysfunction nos\|[d]sleep dysfunction nos (context-dependent category)\|[d]sleep dysfunction nos (situation)\|disorder sleep\|disorder, sleep\|disorder;sleep\|disorders, sleep\|disturbances, sleep\|parasomnia\|problem;sleep\|sleep dis\|sleep disorder\|sleep disorder (diagnosis)\|sleep disorder (disorder)\|sleep disorder (nos)\|sleep disorder nos\|sleep disorder, nos\|sleep disorder, unspecified\|sleep disorders\|sleep disorders and disturbances\|sleep disturbance\|sleep disturbances\|sleep dysfunction nos\|sleep problem\|sleep wake disorders\|sleep wake disorders [disease/finding]\|sleep-wake disorder\|sleep-wake disorder (disorder)\|sleeping disorders |
| C0917799 | 19.2 | 3.2 | excessive sleepiness | [d]hypersomnia\|[d]hypersomnia (context-dependent category)\|[d]hypersomnia (situation)\|[d]hypersomnia nos\|[d]hypersomnia nos (context-dependent category)\|[d]hypersomnia nos (situation)\|excessive sleep\|excessive sleepiness\|excessive sleeping\|hypersomnia\|hypersomnia (diagnosis)\|hypersomnia (disorder)\|hypersomnia (excessive sleeping)\|hypersomnia [d]\|hypersomnia nos\|hypersomnia nos (disorder)\|hypersomnia, nos\|hypersomnia, unspecified\|hypersomnias\|increased sleep\|increased sleeping\|much sleep too\|much sleeping too\|sleep disorder hypersomnia\|sleep excessive\|sleep too much\|sleeping excessive\|sleeping too much\|sleeping too much (hypersomnia)\|sleeping too much (symptom)\|sleeps too much\|too much sleep |
| C0917801 | 62.3 | 33.4 | insomnia | [d]insomnia\|[d]insomnia (context-dependent category)\|[d]insomnia (situation)\|[d]insomnia - symptom\|[d]insomnia nos\|[d]insomnia nos (context-dependent category)\|[d]insomnia nos (situation)\|cannot sleep - insomnia\|disturbances of sleep/insomnia\|insomnia\|insomnia (disorder)\|insomnia (disorder) [ambiguous]\|insomnia (symptom)\|insomnia - symptom\|insomnia [ambiguous] (disorder)\|insomnia [d]\|insomnia dis\|insomnia disorder\|insomnia disorder (diagnosis)\|insomnia disorders\|insomnia nos\|insomnia nos (disorder)\|insomnia symptom\|insomnia, nos\|insomnia, unspecified\|insomnia: [transient] or [nos]\|insomnia: [transient] or [nos] (disorder)\|insomnias\|sleep disorder insomnia\|sleeplessness |
| C1956415 | 49.1 | 31.4 | paroxysmal nocturnal dyspnea | cardiac asthma\|dyspnea nocturnal paroxysmal\|dyspnea paroxysmal\|dyspnea paroxysmal nocturnal\|dyspnea, nocturnal paroxysmal\|dyspnea, paroxysmal nocturnal\|dyspnoea paroxysmal nocturnal\|paroxysmal dyspnea\|paroxysmal nocturnal dyspnea\|paroxysmal nocturnal dyspnea (finding)\|paroxysmal nocturnal dyspnea pnd\|paroxysmal nocturnal dyspnoea\|paroxysmal nocturnal dyspnoea (finding)\|pnd\|pnd - paroxys nocturn dyspnea\|pnd - paroxys nocturn dyspnoea\|pnd - paroxysmal nocturnal dyspnea\|pnd - paroxysmal nocturnal dyspnoea\|pnds |
| C2219848 | 14.8 | 2.1 | daytime somnolence | daytime ce\|daytime somnolence (finding)\|excessive sleepiness during day\|excessive sleepiness during day (symptom)\|excessive sleepiness during the day (daytime somnolence) |
| C2364286 | 11.4 | 6.5 | able to sleep | able to sleep\|able to sleep (finding)\|actual positive sleep\|adequate sleep\|good sleep pattern\|good sleep pattern (finding) |
| C2830004 | 26.9 | 8.8 | somnolence | [d]somnolence\|[d]somnolence (context-dependent category)\|[d]somnolence (situation)\|drowsiness\|sleepiness\|somnolence\|somnolent |

#### Table S9. Incident sleep statistic associations with comorbidities.

Incident analyses considered diagnoses at least one year after the first sleep recording (prioritizing diagnostics, then splits, then home sleep tests) and are ordered by p-values. Primary analyses considered joint-sex PheCodes with >1% incidence rate and were rank-normalized after adjusting for age, sex, BMI, population, sleep study type, and healthcare encounters. The primary analysis considered all available sleep laboratory data (i.e. including participants being screened for other sleep disorders).

1 PheCode was Bonferroni-level significant (**bolded**) and FDR-level significant (in red, uncorrected p-values).

Due to PDF width constraints, sensitivity analyses are presented in two tables, with primary columns repeated (apart from incidence and n):

**Table S9 further includes:**
A sensitivity analysis considering only participants with AHI >15.
A sensitivity analysis further adjusting for an indicator variable for other common sleep disorders (PheCodes 327, 327.1, 327.4, 327.41, 327.5, 327.6, 327.7, 327.71) in the full sleep recording sample.

**Table S10 further includes:**
A sensitivity analysis further adjusting for an indicator variable for common pulmonary disorders (PheCodes 415.1, 415.11, 415.2, 480.1, 480.11, 480.2, 480, 495, 495.2, 496.1, 496.2, 496.21, 496.3, 497, 502, 503, 505, 506, 509.2).
P-values from sleep statistic × sex interaction analyses (not rank-normalized) performed separately.
Equivalent sex-stratified results.

| PheCode | Disease | Sleep statistic | Incidence | Risk ratio | P-value | N | Risk ratio (AHI ≥15) | N (AHI ≥15) | Risk ratio (adj sleep disorders) |
|---|---|---|---|---|---|---|---|---|---|
| 350.1 | Abnormal involuntary movements | AHI 3% | 1.6 | 0.99 (0.98 – 0.99) | **2.0E-04** | 14830 | 0.99 (0.98 – 1.00) | 7699 | 0.99 (0.98 – 0.99) |
| 427.9 | Palpitations | AHI 3% | 1.8 | 0.99 (0.98 – 1.00) | 8.1E-04 | 13710 | 0.99 (0.98 – 1.00) | 7205 | 0.99 (0.98 – 1.00) |
| 285 | Other anemias | Per88 | 2.6 | 1.02 (1.01 – 1.04) | 1.4E-03 | 8330 | 1.02 (1.01 – 1.04) | 4035 | 1.02 (1.01 – 1.04) |
| 530.9 | Heartburn | Per88 | 1.6 | 0.97 (0.95 – 0.99) | 2.1E-03 | 9237 | 0.97 (0.95 – 0.99) | 4476 | 0.97 (0.95 – 0.99) |
| 508 | Pulmonary collapse; interstitial and compensatory emphysema | Per88 | 1.3 | 1.03 (1.01 – 1.06) | 2.8E-03 | 9271 | 1.02 (1.00 – 1.04) | 4447 | 1.03 (1.01 – 1.06) |
| 585.3 | Chronic renal failure [CKD] | Per88 | 1.3 | 1.03 (1.01 – 1.05) | 3.8E-03 | 9252 | 1.03 (1.01 – 1.05) | 4411 | 1.03 (1.01 – 1.05) |
| 519.8 | Other diseases of respiratory system, NEC | Per88 | 1.4 | 1.03 (1.01 – 1.05) | 4.3E-03 | 9444 | 1.02 (1.00 – 1.04) | 4578 | 1.03 (1.01 – 1.05) |
| 292.3 | Memory loss | AHI 3% | 1.6 | 0.99 (0.98 – 1.00) | 4.6E-03 | 14731 | 0.99 (0.98 – 1.00) | 7606 | 0.99 (0.98 – 1.00) |
| 512.9 | Other dyspnea | Per88 | 1.6 | 1.03 (1.01 – 1.05) | 6.1E-03 | 7969 | 1.03 (1.01 – 1.05) | 3789 | 1.03 (1.01 – 1.05) |
| 760 | Back pain | AHI 3% | 1.9 | 0.99 (0.98 – 1.00) | 6.3E-03 | 11884 | 0.99 (0.98 – 1.00) | 6096 | 0.99 (0.98 – 1.00) |
| 340 | Migraine | AHI 3% | 1.1 | 0.99 (0.99 – 1.00) | 8.0E-03 | 13482 | 0.99 (0.98 – 1.00) | 7329 | 0.99 (0.99 – 1.00) |

| PheCode | Disease | Sleep statistic | Incidence | Risk ratio | P-value | N | Risk ratio (AHI ≥15) | N (AHI ≥15) | Risk ratio (adj sleep disorders) |
|---|---|---|---|---|---|---|---|---|---|
| 840 | Sprains and strains | AHI 3% | 2.0 | 0.99 (0.99 – 1.00) | 1.0E-02 | 14207 | 0.99 (0.98 – 1.00) | 7355 | 0.99 (0.99 – 1.00) |
| 382 | Otalgia | Per88 | 1.9 | 0.98 (0.96 – 0.99) | 1.0E-02 | 9060 | 0.99 (0.97 – 1.01) | 4419 | 0.98 (0.96 – 0.99) |
| 455 | Hemorrhoids | AHI 3% | 2.1 | 0.99 (0.99 – 1.00) | 1.5E-02 | 13979 | 0.99 (0.99 – 1.00) | 7233 | 0.99 (0.99 – 1.00) |
| 599.9 | Other abnormality of urination | AHI 3% | 1.6 | 0.99 (0.98 – 1.00) | 1.7E-02 | 14972 | 0.99 (0.98 – 1.00) | 7763 | 0.99 (0.98 – 1.00) |
| 599.5 | Frequency of urination and polyuria | AHI 3% | 1.8 | 0.99 (0.98 – 1.00) | 1.8E-02 | 14148 | 0.99 (0.98 – 1.00) | 7223 | 0.99 (0.98 – 1.00) |
| 781 | Symptoms involving nervous and musculoskeletal systems | Per88 | 1.8 | 1.02 (1.00 – 1.04) | 2.0E-02 | 9286 | 1.02 (1.00 – 1.04) | 4516 | 1.02 (1.00 – 1.04) |
| 563 | Constipation | AHI 3% | 2.4 | 0.99 (0.99 – 1.00) | 2.1E-02 | 14061 | 1.00 (0.99 – 1.00) | 7418 | 0.99 (0.99 – 1.00) |
| 382 | Otalgia | AHI 3% | 1.9 | 0.99 (0.99 – 1.00) | 2.4E-02 | 14490 | 1.00 (0.99 – 1.00) | 7560 | 0.99 (0.99 – 1.00) |
| 480 | Pneumonia | Per88 | 2.0 | 1.02 (1.00 – 1.04) | 2.4E-02 | 9080 | 1.01 (0.99 – 1.03) | 4353 | 1.02 (1.00 – 1.04) |
| 512.9 | Other dyspnea | AHI 3% | 1.6 | 1.01 (1.00 – 1.02) | 2.7E-02 | 12850 | 1.01 (1.00 – 1.02) | 6549 | 1.01 (1.00 – 1.02) |
| 465 | Acute upper respiratory infections of multiple or unspecified sites | AHI 3% | 1.3 | 1.01 (1.00 – 1.02) | 3.2E-02 | 13885 | 1.01 (1.00 – 1.02) | 7221 | 1.01 (1.00 – 1.02) |
| 599.4 | Urinary incontinence | AHI 3% | 1.3 | 0.99 (0.98 – 1.00) | 3.2E-02 | 14569 | 0.99 (0.98 – 1.00) | 7531 | 0.99 (0.98 – 1.00) |
| 561 | Symptoms involving digestive system | AHI 3% | 1.2 | 0.99 (0.98 – 1.00) | 3.3E-02 | 15072 | 0.99 (0.98 – 1.00) | 7818 | 0.99 (0.98 – 1.00) |
| 720 | Spinal stenosis | AHI 3% | 1.1 | 0.99 (0.98 – 1.00) | 3.8E-02 | 14898 | 1.00 (0.99 – 1.01) | 7647 | 0.99 (0.98 – 1.00) |
| 173 | Neoplasm of uncertain behavior of skin | Per88 | 2.0 | 0.98 (0.96 – 1.00) | 3.9E-02 | 8914 | 0.98 (0.96 – 1.00) | 4288 | 0.98 (0.96 – 1.00) |
| 272.13 | Mixed hyperlipidemia | AHI 3% | 1.4 | 1.01 (1.00 – 1.02) | 4.1E-02 | 14573 | 1.01 (1.00 – 1.02) | 7434 | 1.01 (1.00 – 1.02) |
| 350.1 | Abnormal involuntary movements | Per88 | 1.6 | 0.98 (0.96 – 1.00) | 4.3E-02 | 9249 | 0.98 (0.96 – 1.01) | 4505 | 0.98 (0.96 – 1.00) |
| 278.4 | Abnormal weight gain | AHI 3% | 1.3 | 0.99 (0.98 – 1.00) | 4.6E-02 | 14603 | 1.00 (0.99 – 1.01) | 7580 | 0.99 (0.98 – 1.00) |
| 726.1 | Enthesopathy | AHI 3% | 1.6 | 0.99 (0.99 – 1.00) | 4.6E-02 | 13881 | 0.99 (0.98 – 1.00) | 7147 | 0.99 (0.99 – 1.00) |
| 300.11 | Generalized anxiety disorder | AHI 3% | 1.6 | 0.99 (0.99 – 1.00) | 5.1E-02 | 14485 | 0.99 (0.98 – 1.00) | 7657 | 0.99 (0.99 – 1.00) |
| 136 | Other infectious and parasitic diseases | Per88 | 1.6 | 1.02 (1.00 – 1.03) | 5.3E-02 | 9347 | 1.01 (1.00 – 1.03) | 4527 | 1.02 (1.00 – 1.03) |
| 292.3 | Memory loss | Per88 | 1.6 | 0.98 (0.96 – 1.00) | 5.7E-02 | 9172 | 0.99 (0.97 – 1.01) | 4420 | 0.98 (0.96 – 1.00) |
| 740.9 | Osteoarthrosis NOS | AHI 3% | 1.5 | 0.99 (0.98 – 1.00) | 6.1E-02 | 12965 | 1.00 (0.99 – 1.00) | 6461 | 0.99 (0.99 – 1.00) |
| 357 | Inflammatory and toxic neuropathy | AHI 3% | 1.6 | 0.99 (0.98 – 1.00) | 6.3E-02 | 14758 | 1.00 (0.99 – 1.01) | 7605 | 0.99 (0.98 – 1.00) |
| 599.5 | Frequency of urination and polyuria | Per88 | 1.8 | 0.98 (0.96 – 1.00) | 6.4E-02 | 8852 | 0.99 (0.97 – 1.00) | 4207 | 0.98 (0.96 – 1.00) |
| 530.11 | GERD | AHI 3% | 3.9 | 1.00 (0.99 – 1.00) | 6.7E-02 | 10866 | 1.00 (0.99 – 1.00) | 5447 | 1.00 (0.99 – 1.00) |
| 427.3 | Other specified cardiac dysrhythmias | AHI 3% | 1.9 | 0.99 (0.99 – 1.00) | 7.6E-02 | 14582 | 1.00 (0.99 – 1.00) | 7508 | 0.99 (0.99 – 1.00) |
| 760 | Back pain | Per88 | 1.9 | 1.01 (1.00 – 1.03) | 8.1E-02 | 7385 | 1.02 (1.00 – 1.04) | 3544 | 1.01 (1.00 – 1.03) |
| 742.9 | Other derangement of joint | Per88 | 1.5 | 0.98 (0.97 – 1.00) | 8.6E-02 | 9321 | 0.98 (0.96 – 1.00) | 4522 | 0.98 (0.97 – 1.00) |
| 262 | Mineral deficiency NEC | AHI 3% | 1.4 | 0.99 (0.99 – 1.00) | 8.9E-02 | 15118 | 1.00 (0.99 – 1.01) | 7856 | 0.99 (0.99 – 1.00) |

| PheCode | Disease | Sleep statistic | Incidence | Risk ratio | P-value | N | Risk ratio (AHI ≥15) | N (AHI ≥15) | Risk ratio (adj sleep disorders) |
|---|---|---|---|---|---|---|---|---|---|
| 208 | Benign neoplasm of colon | AHI 3% | 1.9 | 0.99 (0.99 – 1.00) | 9.7E-02 | 13336 | 0.99 (0.99 – 1.00) | 6697 | 0.99 (0.99 – 1.00) |
| 276.5 | Hypovolemia | Per88 | 1.5 | 1.02 (1.00 – 1.04) | 9.9E-02 | 9233 | 1.01 (0.99 – 1.03) | 4483 | 1.02 (1.00 – 1.04) |
| 689 | Disorder of skin and subcutaneous tissue NOS | Per88 | 4.8 | 0.99 (0.98 – 1.00) | 1.1E-01 | 8700 | 0.99 (0.98 – 1.00) | 4179 | 0.99 (0.98 – 1.00) |
| 366.2 | Senile cataract | AHI 3% | 1.6 | 0.99 (0.99 – 1.00) | 1.2E-01 | 14630 | 0.99 (0.98 – 1.00) | 7484 | 0.99 (0.99 – 1.00) |
| 338.2 | Chronic pain | AHI 3% | 3.0 | 1.00 (0.99 – 1.00) | 1.2E-01 | 13500 | 0.99 (0.99 – 1.00) | 6997 | 1.00 (0.99 – 1.00) |
| 476 | Allergic rhinitis | Per88 | 1.5 | 0.99 (0.97 – 1.00) | 1.3E-01 | 8332 | 0.98 (0.95 – 1.00) | 4074 | 0.99 (0.97 – 1.00) |
| 550.2 | Diaphragmatic hernia | AHI 3% | 1.3 | 0.99 (0.99 – 1.00) | 1.4E-01 | 14748 | 0.99 (0.98 – 1.00) | 7592 | 0.99 (0.99 – 1.00) |
| 701.2 | Scar conditions and fibrosis of skin | Per88 | 1.3 | 0.98 (0.96 – 1.01) | 1.5E-01 | 9193 | 0.99 (0.96 – 1.01) | 4468 | 0.98 (0.96 – 1.01) |
| 365 | Glaucoma | Per88 | 1.2 | 0.98 (0.95 – 1.01) | 1.5E-01 | 9252 | 0.98 (0.96 – 1.01) | 4441 | 0.98 (0.95 – 1.01) |
| 495 | Asthma | Per88 | 1.5 | 1.01 (1.00 – 1.03) | 1.5E-01 | 7746 | 1.02 (0.99 – 1.04) | 3795 | 1.01 (1.00 – 1.03) |
| 687.1 | Rash and other nonspecific skin eruption | Per88 | 2.6 | 1.01 (1.00 – 1.02) | 1.5E-01 | 8724 | 1.01 (0.99 – 1.02) | 4254 | 1.01 (1.00 – 1.02) |
| 480 | Pneumonia | AHI 3% | 2.0 | 1.00 (1.00 – 1.01) | 1.5E-01 | 14512 | 1.00 (0.99 – 1.01) | 7429 | 1.01 (1.00 – 1.01) |
| 402 | Elevated blood pressure reading without diagnosis of hypertension | Per88 | 1.7 | 0.98 (0.96 – 1.01) | 1.6E-01 | 8895 | 0.99 (0.96 – 1.01) | 4277 | 0.98 (0.96 – 1.01) |
| 701 | Other hypertrophic and atrophic conditions of skin | Per88 | 2.6 | 0.99 (0.97 – 1.00) | 1.7E-01 | 8800 | 0.99 (0.97 – 1.00) | 4210 | 0.99 (0.97 – 1.00) |
| 375.1 | Dry eyes | AHI 3% | 1.5 | 1.00 (0.99 – 1.00) | 1.8E-01 | 14923 | 0.99 (0.99 – 1.00) | 7751 | 1.00 (0.99 – 1.00) |
| 743.11 | Osteoporosis NOS | Per88 | 1.3 | 1.01 (0.99 – 1.03) | 1.8E-01 | 9023 | 1.01 (1.00 – 1.03) | 4358 | 1.01 (0.99 – 1.03) |
| 472 | Chronic pharyngitis and nasopharyngitis | Per88 | 1.6 | 1.01 (0.99 – 1.03) | 1.8E-01 | 9020 | 1.00 (0.98 – 1.03) | 4366 | 1.01 (0.99 – 1.03) |
| 726 | Peripheral enthesopathies and allied syndromes | AHI 3% | 1.1 | 1.00 (0.99 – 1.00) | 1.9E-01 | 14304 | 0.99 (0.98 – 1.00) | 7397 | 1.00 (0.99 – 1.00) |
| 304 | Adjustment reaction | AHI 3% | 1.5 | 1.00 (0.99 – 1.00) | 2.0E-01 | 14422 | 0.99 (0.98 – 1.00) | 7502 | 1.00 (0.99 – 1.00) |
| 280.1 | Iron deficiency anemias, unspecified or not due to blood loss | AHI 3% | 1.3 | 0.99 (0.99 – 1.00) | 2.0E-01 | 14672 | 1.00 (0.99 – 1.01) | 7608 | 1.00 (0.99 – 1.00) |
| 764 | Sciatica | AHI 3% | 1.8 | 1.00 (0.99 – 1.00) | 2.0E-01 | 14416 | 0.99 (0.99 – 1.00) | 7411 | 1.00 (0.99 – 1.00) |
| 389 | Hearing loss | Per88 | 2.2 | 1.01 (0.99 – 1.03) | 2.0E-01 | 8972 | 1.01 (0.99 – 1.02) | 4281 | 1.01 (0.99 – 1.03) |
| 550.2 | Diaphragmatic hernia | Per88 | 1.3 | 0.98 (0.96 – 1.01) | 2.1E-01 | 9230 | 0.98 (0.96 – 1.01) | 4433 | 0.98 (0.96 – 1.01) |
| 366.2 | Senile cataract | Per88 | 1.6 | 0.99 (0.96 – 1.01) | 2.1E-01 | 9140 | 0.99 (0.97 – 1.01) | 4363 | 0.99 (0.96 – 1.01) |
| 594.1 | Calculus of kidney | AHI 3% | 1.3 | 1.00 (1.00 – 1.01) | 2.2E-01 | 14492 | 1.00 (0.99 – 1.01) | 7429 | 1.00 (1.00 – 1.01) |
| 571.5 | Other chronic nonalcoholic liver disease | Per88 | 1.2 | 0.98 (0.95 – 1.01) | 2.3E-01 | 9327 | 0.98 (0.95 – 1.02) | 4468 | 0.98 (0.95 – 1.01) |
| 565.1 | Anal and rectal polyp | AHI 3% | 1.3 | 1.00 (1.00 – 1.01) | 2.3E-01 | 15043 | 1.00 (1.00 – 1.01) | 7746 | 1.00 (1.00 – 1.01) |
| 706.8 | Other specified diseases of | AHI 3% | 1.9 | 1.00 (0.99 – 1.00) | 2.4E-01 | 14368 | 1.00 (0.99 – 1.01) | 7414 | 1.00 (0.99 – 1.00) |

| PheCode | Disease | Sleep statistic | Incidence | Risk ratio | P-value | N | Risk ratio (AHI ≥15) | N (AHI ≥15) | Risk ratio (adj sleep disorders) |
|---|---|---|---|---|---|---|---|---|---|
| | sebaceous glands | | | | | | | | |
| 401.1 | Essential hypertension | AHI 3% | 2.8 | 1.00 (1.00 – 1.01) | 2.4E-01 | 8968 | 1.00 (0.99 – 1.00) | 3740 | 1.00 (1.00 – 1.01) |
| 781 | Symptoms involving nervous and musculoskeletal systems | AHI 3% | 1.8 | 1.00 (1.00 – 1.01) | 2.5E-01 | 14904 | 1.00 (1.00 – 1.01) | 7718 | 1.00 (1.00 – 1.01) |
| 579.8 | Nonspecific abnormal findings in stool contents | Per88 | 1.7 | 0.99 (0.97 – 1.01) | 2.6E-01 | 9351 | 0.98 (0.96 – 1.00) | 4519 | 0.99 (0.97 – 1.01) |
| 272.13 | Mixed hyperlipidemia | Per88 | 1.4 | 0.98 (0.95 – 1.01) | 2.7E-01 | 9150 | 0.99 (0.96 – 1.02) | 4362 | 0.98 (0.95 – 1.01) |
| 272.1 | Hyperlipidemia | AHI 3% | 3.6 | 1.00 (0.99 – 1.00) | 2.7E-01 | 10935 | 0.99 (0.99 – 1.00) | 5099 | 1.00 (0.99 – 1.00) |
| 563 | Constipation | Per88 | 2.4 | 1.01 (0.99 – 1.02) | 2.7E-01 | 8682 | 1.01 (0.99 – 1.02) | 4321 | 1.01 (0.99 – 1.02) |
| 465 | Acute upper respiratory infections of multiple or unspecified sites | Per88 | 1.3 | 1.01 (0.99 – 1.03) | 2.8E-01 | 8709 | 1.01 (1.00 – 1.03) | 4238 | 1.01 (0.99 – 1.03) |
| 519.8 | Other diseases of respiratory system, NEC | AHI 3% | 1.4 | 1.00 (1.00 – 1.01) | 2.9E-01 | 15116 | 1.01 (1.00 – 1.02) | 7821 | 1.00 (1.00 – 1.01) |
| 689 | Disorder of skin and subcutaneous tissue NOS | AHI 3% | 4.8 | 1.00 (0.99 – 1.00) | 2.9E-01 | 13949 | 1.00 (0.99 – 1.00) | 7176 | 1.00 (0.99 – 1.00) |
| 306 | Other mental disorder | Per88 | 3.0 | 1.01 (0.99 – 1.02) | 3.0E-01 | 9016 | 1.00 (0.99 – 1.02) | 4380 | 1.01 (0.99 – 1.02) |
| 365 | Glaucoma | AHI 3% | 1.2 | 1.00 (0.99 – 1.00) | 3.0E-01 | 14762 | 0.99 (0.98 – 1.00) | 7573 | 1.00 (0.99 – 1.00) |
| 386.9 | Dizziness and giddiness (Light-headedness and vertigo) | AHI 3% | 2.1 | 1.00 (1.00 – 1.01) | 3.0E-01 | 13602 | 1.00 (0.99 – 1.01) | 7051 | 1.00 (1.00 – 1.01) |
| 427.9 | Palpitations | Per88 | 1.8 | 0.99 (0.97 – 1.01) | 3.0E-01 | 8525 | 0.99 (0.96 – 1.02) | 4178 | 0.99 (0.97 – 1.01) |
| 593.2 | Microscopic hematuria | Per88 | 1.2 | 0.99 (0.97 – 1.01) | 3.0E-01 | 9354 | 1.00 (0.98 – 1.01) | 4519 | 0.99 (0.97 – 1.01) |
| 357 | Inflammatory and toxic neuropathy | Per88 | 1.6 | 1.01 (0.99 – 1.03) | 3.1E-01 | 9212 | 1.01 (0.99 – 1.03) | 4441 | 1.01 (0.99 – 1.03) |
| 476 | Allergic rhinitis | AHI 3% | 1.5 | 1.00 (0.99 – 1.00) | 3.2E-01 | 13345 | 1.00 (0.99 – 1.01) | 6985 | 1.00 (0.99 – 1.00) |
| 300.1 | Anxiety disorder | Per88 | 2.8 | 1.01 (0.99 – 1.02) | 3.2E-01 | 7210 | 1.00 (0.99 – 1.02) | 3645 | 1.01 (0.99 – 1.02) |
| 379.9 | Pain, swelling or discharge of eye | Per88 | 1.1 | 0.99 (0.97 – 1.01) | 3.3E-01 | 9327 | 1.00 (0.97 – 1.02) | 4528 | 0.99 (0.97 – 1.01) |
| 272.11 | Hypercholesterolemia | Per88 | 2.3 | 1.01 (0.99 – 1.03) | 3.4E-01 | 7915 | 1.01 (0.99 – 1.03) | 3602 | 1.01 (0.99 – 1.03) |
| 427.6 | Premature beats | Per88 | 1.2 | 1.01 (0.99 – 1.03) | 3.4E-01 | 9463 | 1.01 (0.99 – 1.03) | 4572 | 1.01 (0.99 – 1.03) |
| 389.4 | Tinnitus | AHI 3% | 1.6 | 1.00 (0.99 – 1.00) | 3.5E-01 | 14710 | 1.00 (0.99 – 1.01) | 7641 | 1.00 (0.99 – 1.00) |
| 704 | Diseases of hair and hair follicles | AHI 3% | 1.1 | 1.00 (0.99 – 1.00) | 3.7E-01 | 15044 | 1.00 (0.99 – 1.01) | 7805 | 1.00 (0.99 – 1.00) |
| 340 | Migraine | Per88 | 1.1 | 0.99 (0.98 – 1.01) | 3.7E-01 | 8315 | 0.99 (0.97 – 1.01) | 4260 | 0.99 (0.98 – 1.01) |
| 798 | Malaise and fatigue | Per88 | 2.5 | 1.01 (0.99 – 1.02) | 3.8E-01 | 7852 | 1.00 (0.98 – 1.02) | 3908 | 1.01 (0.99 – 1.02) |
| 278.11 | Morbid obesity | AHI 3% | 1.8 | 1.00 (1.00 – 1.01) | 3.8E-01 | 13104 | 1.00 (0.99 – 1.01) | 6429 | 1.00 (1.00 – 1.01) |
| 455 | Hemorrhoids | Per88 | 2.1 | 0.99 (0.97 – 1.01) | 3.8E-01 | 8739 | 1.00 (0.97 – 1.02) | 4229 | 0.99 (0.97 – 1.01) |
| 316 | Substance addiction and disorders | Per88 | 1.1 | 1.01 (0.99 – 1.03) | 3.8E-01 | 9188 | 1.00 (0.97 – 1.03) | 4433 | 1.01 (0.99 – 1.03) |
| 278.11 | Morbid obesity | Per88 | 1.8 | 1.01 (0.99 – 1.04) | 3.9E-01 | 8273 | 1.01 (0.99 – 1.04) | 3806 | 1.01 (0.99 – 1.04) |
| 470 | Septal Deviations/Turbinate | AHI 3% | 1.2 | 1.00 (1.00 – 1.01) | 3.9E-01 | 14480 | 1.00 (0.99 – 1.01) | 7550 | 1.00 (1.00 – 1.01) |

| PheCode | Disease | Sleep statistic | Incidence | Risk ratio | P-value | N | Risk ratio (AHI ≥15) | N (AHI ≥15) | Risk ratio (adj sleep disorders) |
|---|---|---|---|---|---|---|---|---|---|
| | Hypertrophy | | | | | | | | |
| 278.1 | Obesity | Per88 | 3.5 | 1.01 (0.99 – 1.03) | 4.0E-01 | 6442 | 1.00 (0.98 – 1.02) | 2787 | 1.01 (0.99 – 1.03) |
| 740.9 | Osteoarthrosis NOS | Per88 | 1.5 | 0.99 (0.96 – 1.01) | 4.0E-01 | 8113 | 1.00 (0.97 – 1.02) | 3759 | 0.99 (0.96 – 1.01) |
| 561 | Symptoms involving digestive system | Per88 | 1.2 | 0.99 (0.97 – 1.01) | 4.0E-01 | 9424 | 1.00 (0.98 – 1.02) | 4576 | 0.99 (0.97 – 1.01) |
| 327.4 | Insomnia | Per88 | 1.7 | 1.01 (0.99 – 1.02) | 4.0E-01 | 7978 | 1.00 (0.98 – 1.02) | 4022 | 1.01 (0.99 – 1.02) |
| 585.3 | Chronic renal failure [CKD] | AHI 3% | 1.3 | 1.00 (0.99 – 1.01) | 4.2E-01 | 14754 | 1.00 (0.99 – 1.01) | 7515 | 1.00 (0.99 – 1.01) |
| 687.1 | Rash and other nonspecific skin eruption | AHI 3% | 2.6 | 1.00 (0.99 – 1.00) | 4.2E-01 | 14007 | 0.99 (0.99 – 1.00) | 7306 | 1.00 (0.99 – 1.00) |
| 475.9 | Postnasal drip | Per88 | 1.7 | 1.01 (0.99 – 1.02) | 4.3E-01 | 9341 | 1.01 (0.99 – 1.02) | 4533 | 1.01 (0.99 – 1.02) |
| 278 | Overweight, obesity and other hyperalimentation | Per88 | 2.3 | 0.99 (0.98 – 1.01) | 4.3E-01 | 8440 | 1.00 (0.98 – 1.02) | 4046 | 0.99 (0.98 – 1.01) |
| 280.1 | Iron deficiency anemias, unspecified or not due to blood loss | Per88 | 1.3 | 0.99 (0.97 – 1.02) | 4.4E-01 | 9144 | 1.00 (0.96 – 1.03) | 4444 | 0.99 (0.97 – 1.02) |
| 809 | Fracture of unspecified bones | AHI 3% | 1.4 | 1.00 (0.99 – 1.00) | 4.4E-01 | 14935 | 1.00 (0.99 – 1.01) | 7687 | 1.00 (0.99 – 1.00) |
| 277 | Other disorders of metabolism | Per88 | 2.6 | 0.99 (0.98 – 1.01) | 4.4E-01 | 9230 | 1.00 (0.98 – 1.01) | 4486 | 0.99 (0.98 – 1.01) |
| 761 | Cervicalgia | AHI 3% | 2.0 | 1.00 (1.00 – 1.01) | 4.5E-01 | 13801 | 1.01 (1.00 – 1.01) | 7215 | 1.00 (1.00 – 1.01) |
| 304 | Adjustment reaction | Per88 | 1.5 | 1.01 (0.99 – 1.02) | 4.5E-01 | 8964 | 1.01 (0.99 – 1.02) | 4365 | 1.01 (0.99 – 1.02) |
| 761 | Cervicalgia | Per88 | 2.0 | 0.99 (0.98 – 1.01) | 4.6E-01 | 8551 | 0.99 (0.96 – 1.01) | 4192 | 0.99 (0.98 – 1.01) |
| 369.5 | Conjunctivitis, infectious | Per88 | 1.2 | 1.01 (0.99 – 1.02) | 4.6E-01 | 9265 | 1.01 (0.99 – 1.03) | 4511 | 1.01 (0.99 – 1.02) |
| 316 | Substance addiction and disorders | AHI 3% | 1.1 | 1.00 (0.99 – 1.01) | 4.6E-01 | 14760 | 1.00 (0.98 – 1.01) | 7616 | 1.00 (0.99 – 1.01) |
| 389 | Hearing loss | AHI 3% | 2.2 | 1.00 (1.00 – 1.01) | 4.8E-01 | 14350 | 1.00 (0.99 – 1.01) | 7343 | 1.00 (1.00 – 1.01) |
| 306 | Other mental disorder | AHI 3% | 3.0 | 1.00 (1.00 – 1.01) | 4.8E-01 | 14521 | 1.00 (1.00 – 1.01) | 7533 | 1.00 (1.00 – 1.01) |
| 196 | Radiotherapy | AHI 3% | 1.1 | 1.00 (0.99 – 1.01) | 4.8E-01 | 15183 | 1.00 (0.99 – 1.01) | 7836 | 1.00 (0.99 – 1.01) |
| 173 | Neoplasm of uncertain behavior of skin | AHI 3% | 2.0 | 1.00 (1.00 – 1.01) | 4.9E-01 | 14201 | 1.01 (1.00 – 1.01) | 7318 | 1.00 (1.00 – 1.01) |
| 742.9 | Other derangement of joint | AHI 3% | 1.5 | 1.00 (0.99 – 1.00) | 5.1E-01 | 14927 | 0.99 (0.98 – 1.00) | 7742 | 1.00 (0.99 – 1.00) |
| 271.3 | Intestinal disaccharidase deficiencies and disaccharide malabsorption | Per88 | 4.8 | 1.00 (0.98 – 1.01) | 5.1E-01 | 8633 | 0.99 (0.98 – 1.01) | 4076 | 1.00 (0.98 – 1.01) |
| 351 | Other peripheral nerve disorders | Per88 | 1.3 | 1.01 (0.99 – 1.03) | 5.1E-01 | 9024 | 1.01 (0.99 – 1.04) | 4365 | 1.01 (0.99 – 1.03) |
| 292 | Neurological disorders | Per88 | 1.2 | 0.99 (0.97 – 1.02) | 5.3E-01 | 9398 | 0.99 (0.97 – 1.02) | 4569 | 0.99 (0.97 – 1.02) |
| 296.22 | Major depressive disorder | AHI 3% | 2.7 | 1.00 (0.99 – 1.00) | 5.4E-01 | 12139 | 1.00 (0.99 – 1.01) | 6418 | 1.00 (0.99 – 1.00) |
| 272.11 | Hypercholesterolemia | AHI 3% | 2.3 | 1.00 (0.99 – 1.00) | 5.5E-01 | 12454 | 1.00 (0.99 – 1.00) | 6101 | 1.00 (0.99 – 1.00) |
| 196 | Radiotherapy | Per88 | 1.1 | 0.99 (0.96 – 1.02) | 5.5E-01 | 9498 | 1.00 (0.97 – 1.02) | 4586 | 0.99 (0.96 – 1.02) |
| 530.9 | Heartburn | AHI 3% | 1.6 | 1.00 (0.99 – 1.00) | 5.5E-01 | 14782 | 1.00 (0.99 – 1.00) | 7669 | 1.00 (0.99 – 1.01) |

| PheCode | Disease | Sleep statistic | Incidence | Risk ratio | P-value | N | Risk ratio (AHI ≥15) | N (AHI ≥15) | Risk ratio (adj sleep disorders) |
|---|---|---|---|---|---|---|---|---|---|
| 726 | Peripheral enthesopathies and allied syndromes | Per88 | 1.1 | 0.99 (0.97 – 1.02) | 5.6E-01 | 8959 | 0.99 (0.96 – 1.02) | 4335 | 0.99 (0.97 – 1.02) |
| 338.2 | Chronic pain | Per88 | 3.0 | 1.00 (0.98 – 1.01) | 5.6E-01 | 8348 | 1.00 (0.98 – 1.02) | 4067 | 1.00 (0.98 – 1.01) |
| 965 | Poisoning by analgesics, antipyretics, and antirheumatics | AHI 3% | 1.8 | 1.00 (0.99 – 1.01) | 5.8E-01 | 15050 | 1.00 (0.99 – 1.01) | 7786 | 1.00 (0.99 – 1.01) |
| 008 | Intestinal infection | Per88 | 1.4 | 1.00 (0.99 – 1.02) | 5.8E-01 | 9384 | 1.00 (0.98 – 1.02) | 4556 | 1.00 (0.99 – 1.02) |
| 475.9 | Postnasal drip | AHI 3% | 1.7 | 1.00 (0.99 – 1.00) | 5.9E-01 | 14921 | 0.99 (0.99 – 1.00) | 7719 | 1.00 (0.99 – 1.00) |
| 351 | Other peripheral nerve disorders | AHI 3% | 1.3 | 1.00 (0.99 – 1.01) | 6.0E-01 | 14432 | 1.01 (1.00 – 1.02) | 7451 | 1.00 (0.99 – 1.01) |
| 594.1 | Calculus of kidney | Per88 | 1.3 | 0.99 (0.97 – 1.02) | 6.0E-01 | 9096 | 0.99 (0.97 – 1.01) | 4369 | 0.99 (0.97 – 1.02) |
| 470 | Septal Deviations/Turbinate Hypertrophy | Per88 | 1.2 | 0.99 (0.97 – 1.02) | 6.0E-01 | 9085 | 1.00 (0.97 – 1.03) | 4432 | 0.99 (0.97 – 1.02) |
| 289.4 | Lymphadenitis | AHI 3% | 1.1 | 1.00 (0.99 – 1.01) | 6.0E-01 | 14770 | 1.00 (0.99 – 1.01) | 7624 | 1.00 (0.99 – 1.01) |
| 740.11 | Osteoarthrosis, localized, primary | Per88 | 2.0 | 1.01 (0.99 – 1.03) | 6.1E-01 | 8732 | 1.01 (1.00 – 1.03) | 4093 | 1.01 (0.98 – 1.03) |
| 599.2 | Retention of urine | AHI 3% | 1.4 | 1.00 (0.99 – 1.01) | 6.1E-01 | 15042 | 1.01 (1.00 – 1.01) | 7756 | 1.00 (0.99 – 1.01) |
| 537 | Other disorders of stomach and duodenum | Per88 | 2.0 | 1.00 (0.98 – 1.01) | 6.2E-01 | 9046 | 1.00 (0.98 – 1.02) | 4401 | 1.00 (0.98 – 1.01) |
| 740.11 | Osteoarthrosis, localized, primary | AHI 3% | 2.0 | 1.00 (0.99 – 1.01) | 6.2E-01 | 13892 | 1.00 (0.99 – 1.01) | 6988 | 1.00 (0.99 – 1.01) |
| 726.1 | Enthesopathy | Per88 | 1.6 | 0.99 (0.97 – 1.02) | 6.3E-01 | 8665 | 1.00 (0.98 – 1.02) | 4158 | 0.99 (0.97 – 1.02) |
| 599.2 | Retention of urine | Per88 | 1.4 | 0.99 (0.97 – 1.02) | 6.4E-01 | 9410 | 0.99 (0.97 – 1.02) | 4547 | 0.99 (0.97 – 1.02) |
| 809 | Fracture of unspecified bones | Per88 | 1.4 | 0.99 (0.97 – 1.02) | 6.4E-01 | 9348 | 0.99 (0.97 – 1.01) | 4502 | 0.99 (0.97 – 1.02) |
| 512.2 | Painful respiration | Per88 | 2.4 | 1.00 (0.98 – 1.01) | 6.5E-01 | 8902 | 1.00 (0.99 – 1.02) | 4303 | 1.00 (0.98 – 1.01) |
| 764 | Sciatica | Per88 | 1.8 | 1.00 (0.98 – 1.02) | 6.5E-01 | 8968 | 0.99 (0.97 – 1.02) | 4307 | 1.00 (0.98 – 1.02) |
| 296.22 | Major depressive disorder | Per88 | 2.7 | 1.00 (0.99 – 1.02) | 6.6E-01 | 7372 | 1.00 (0.98 – 1.02) | 3637 | 1.00 (0.99 – 1.02) |
| 537 | Other disorders of stomach and duodenum | AHI 3% | 2.0 | 1.00 (0.99 – 1.00) | 6.6E-01 | 14486 | 1.00 (0.99 – 1.01) | 7511 | 1.00 (0.99 – 1.01) |
| 495 | Asthma | AHI 3% | 1.5 | 1.00 (0.99 – 1.01) | 6.6E-01 | 12524 | 1.00 (0.99 – 1.01) | 6565 | 1.00 (0.99 – 1.01) |
| 327.4 | Insomnia | AHI 3% | 1.7 | 1.00 (0.99 – 1.01) | 6.6E-01 | 13047 | 1.00 (0.99 – 1.01) | 6983 | 1.00 (0.99 – 1.01) |
| 512.2 | Painful respiration | AHI 3% | 2.4 | 1.00 (0.99 – 1.00) | 6.7E-01 | 14285 | 1.00 (0.99 – 1.01) | 7397 | 1.00 (0.99 – 1.01) |
| 289.4 | Lymphadenitis | Per88 | 1.1 | 1.00 (0.97 – 1.02) | 6.7E-01 | 9237 | 0.99 (0.97 – 1.01) | 4465 | 1.00 (0.97 – 1.02) |
| 579.8 | Nonspecific abnormal findings in stool contents | AHI 3% | 1.7 | 1.00 (0.99 – 1.01) | 6.7E-01 | 14957 | 1.00 (0.99 – 1.01) | 7723 | 1.00 (0.99 – 1.01) |
| 599.8 | Other symptoms involving urinary system | AHI 3% | 1.4 | 1.00 (0.99 – 1.01) | 6.7E-01 | 15157 | 1.00 (0.99 – 1.01) | 7849 | 1.00 (0.99 – 1.01) |
| 300.1 | Anxiety disorder | AHI 3% | 2.8 | 1.00 (0.99 – 1.00) | 6.8E-01 | 11881 | 1.00 (0.99 – 1.01) | 6407 | 1.00 (0.99 – 1.00) |
| 706.8 | Other specified diseases of sebaceous glands | Per88 | 1.9 | 1.00 (0.98 – 1.02) | 6.8E-01 | 8986 | 1.00 (0.98 – 1.02) | 4340 | 1.00 (0.98 – 1.02) |

| PheCode | Disease | Sleep statistic | Incidence | Risk ratio | P-value | N | Risk ratio (AHI ≥15) | N (AHI ≥15) | Risk ratio (adj sleep disorders) |
|---|---|---|---|---|---|---|---|---|---|
| 508 | Pulmonary collapse; interstitial and compensatory emphysema | AHI 3% | 1.3 | 1.00 (0.99 – 1.01) | 6.8E-01 | 14829 | 1.01 (1.00 – 1.02) | 7589 | 1.00 (0.99 – 1.01) |
| 427.3 | Other specified cardiac dysrhythmias | Per88 | 1.9 | 1.00 (0.98 – 1.02) | 6.8E-01 | 9086 | 0.99 (0.97 – 1.01) | 4373 | 1.00 (0.98 – 1.02) |
| 459.9 | Circulatory disease NEC | AHI 3% | 1.5 | 1.00 (0.99 – 1.01) | 6.9E-01 | 14955 | 1.01 (1.00 – 1.02) | 7694 | 1.00 (0.99 – 1.01) |
| 296 | Mood disorders | AHI 3% | 1.2 | 1.00 (0.99 – 1.01) | 6.9E-01 | 14650 | 1.00 (0.99 – 1.01) | 7604 | 1.00 (0.99 – 1.01) |
| 720 | Spinal stenosis | Per88 | 1.1 | 1.00 (0.97 – 1.02) | 6.9E-01 | 9327 | 1.00 (0.98 – 1.02) | 4476 | 1.00 (0.97 – 1.02) |
| 701.2 | Scar conditions and fibrosis of skin | AHI 3% | 1.3 | 1.00 (0.99 – 1.01) | 7.0E-01 | 14684 | 1.00 (0.99 – 1.01) | 7621 | 1.00 (0.99 – 1.01) |
| 429.3 | Symptoms involving cardiovascular system | AHI 3% | 2.3 | 1.00 (0.99 – 1.00) | 7.1E-01 | 14709 | 1.00 (0.99 – 1.00) | 7572 | 1.00 (0.99 – 1.00) |
| 599.9 | Other abnormality of urination | Per88 | 1.6 | 1.00 (0.98 – 1.02) | 7.1E-01 | 9351 | 0.99 (0.97 – 1.01) | 4539 | 1.00 (0.98 – 1.02) |
| 008 | Intestinal infection | AHI 3% | 1.4 | 1.00 (0.99 – 1.01) | 7.1E-01 | 15014 | 1.00 (0.99 – 1.01) | 7783 | 1.00 (0.99 – 1.01) |
| 389.4 | Tinnitus | Per88 | 1.6 | 1.00 (0.98 – 1.01) | 7.2E-01 | 9178 | 1.00 (0.98 – 1.01) | 4459 | 1.00 (0.98 – 1.01) |
| 704 | Diseases of hair and hair follicles | Per88 | 1.1 | 1.00 (0.98 – 1.02) | 7.5E-01 | 9423 | 1.00 (0.97 – 1.02) | 4574 | 1.00 (0.98 – 1.02) |
| 261.4 | Vitamin D deficiency | AHI 3% | 1.9 | 1.00 (0.99 – 1.01) | 7.5E-01 | 13875 | 1.00 (0.99 – 1.01) | 7194 | 1.00 (0.99 – 1.01) |
| 271.3 | Intestinal disaccharidase deficiencies and disaccharide malabsorption | AHI 3% | 4.8 | 1.00 (1.00 – 1.00) | 7.5E-01 | 13830 | 1.00 (0.99 – 1.00) | 7019 | 1.00 (1.00 – 1.00) |
| 840 | Sprains and strains | Per88 | 2.0 | 1.00 (0.99 – 1.02) | 7.6E-01 | 8923 | 0.99 (0.97 – 1.02) | 4318 | 1.00 (0.99 – 1.02) |
| 375.1 | Dry eyes | Per88 | 1.5 | 1.00 (0.99 – 1.02) | 7.6E-01 | 9322 | 1.00 (0.98 – 1.01) | 4529 | 1.00 (0.99 – 1.02) |
| 704.1 | Alopecia | Per88 | 1.5 | 1.00 (0.98 – 1.02) | 7.6E-01 | 9207 | 1.00 (0.98 – 1.03) | 4520 | 1.00 (0.98 – 1.02) |
| 296 | Mood disorders | Per88 | 1.2 | 1.00 (0.98 – 1.02) | 7.7E-01 | 9084 | 1.01 (0.98 – 1.03) | 4410 | 1.00 (0.98 – 1.02) |
| 272.1 | Hyperlipidemia | Per88 | 3.6 | 1.00 (0.99 – 1.02) | 7.7E-01 | 6985 | 0.99 (0.98 – 1.01) | 3060 | 1.00 (0.99 – 1.02) |
| 389.1 | Sensorineural hearing loss | Per88 | 1.1 | 1.00 (0.98 – 1.03) | 7.8E-01 | 9180 | 1.00 (0.98 – 1.03) | 4404 | 1.00 (0.98 – 1.03) |
| 278.4 | Abnormal weight gain | Per88 | 1.3 | 1.00 (0.98 – 1.02) | 7.8E-01 | 9141 | 1.00 (0.97 – 1.03) | 4432 | 1.00 (0.98 – 1.02) |
| 599.4 | Urinary incontinence | Per88 | 1.3 | 1.00 (0.97 – 1.02) | 7.8E-01 | 9075 | 1.00 (0.97 – 1.02) | 4396 | 1.00 (0.97 – 1.02) |
| 765 | Cervical radiculitis | AHI 3% | 1.1 | 1.00 (0.99 – 1.01) | 7.8E-01 | 14936 | 1.00 (0.99 – 1.01) | 7731 | 1.00 (0.99 – 1.01) |
| 277 | Other disorders of metabolism | AHI 3% | 2.6 | 1.00 (1.00 – 1.01) | 7.9E-01 | 14796 | 1.00 (0.99 – 1.01) | 7682 | 1.00 (1.00 – 1.01) |
| 261.4 | Vitamin D deficiency | Per88 | 1.9 | 1.00 (0.98 – 1.02) | 7.9E-01 | 8642 | 1.00 (0.98 – 1.02) | 4182 | 1.00 (0.98 – 1.02) |
| 701 | Other hypertrophic and atrophic conditions of skin | AHI 3% | 2.6 | 1.00 (0.99 – 1.01) | 7.9E-01 | 13954 | 1.01 (1.00 – 1.01) | 7162 | 1.00 (0.99 – 1.01) |
| 276.5 | Hypovolemia | AHI 3% | 1.5 | 1.00 (0.99 – 1.01) | 8.0E-01 | 14763 | 1.00 (1.00 – 1.01) | 7640 | 1.00 (0.99 – 1.01) |
| 262 | Mineral deficiency NEC | Per88 | 1.4 | 1.00 (0.98 – 1.02) | 8.0E-01 | 9447 | 1.00 (0.97 – 1.02) | 4597 | 1.00 (0.98 – 1.02) |
| 965 | Poisoning by analgesics, antipyretics, and antirheumatics | Per88 | 1.8 | 1.00 (0.98 – 1.02) | 8.0E-01 | 9409 | 0.99 (0.97 – 1.01) | 4559 | 1.00 (0.98 – 1.02) |
| 571.5 | Other chronic nonalcoholic liver | AHI 3% | 1.2 | 1.00 (0.99 – 1.01) | 8.1E-01 | 14818 | 1.00 (0.99 – 1.01) | 7594 | 1.00 (0.99 – 1.01) |

| PheCode | Disease | Sleep statistic | Incidence | Risk ratio | P-value | N | Risk ratio (AHI ≥15) | N (AHI ≥15) | Risk ratio (adj sleep disorders) |
|---|---|---|---|---|---|---|---|---|---|
| | disease | | | | | | | | |
| 427.6 | Premature beats | AHI 3% | 1.2 | 1.00 (0.99 – 1.01) | 8.1E-01 | 15116 | 0.99 (0.98 – 1.00) | 7807 | 1.00 (0.99 – 1.01) |
| 278.1 | Obesity | AHI 3% | 3.5 | 1.00 (0.99 – 1.01) | 8.2E-01 | 10143 | 1.00 (0.99 – 1.01) | 4689 | 1.00 (0.99 – 1.01) |
| 481 | Influenza | AHI 3% | 1.5 | 1.00 (0.99 – 1.01) | 8.4E-01 | 14534 | 1.00 (0.99 – 1.01) | 7509 | 1.00 (0.99 – 1.01) |
| 278 | Overweight, obesity and other hyperalimentation | AHI 3% | 2.3 | 1.00 (0.99 – 1.01) | 8.4E-01 | 13389 | 1.00 (1.00 – 1.01) | 6871 | 1.00 (0.99 – 1.01) |
| 136 | Other infectious and parasitic diseases | AHI 3% | 1.6 | 1.00 (0.99 – 1.01) | 8.4E-01 | 14960 | 1.00 (0.99 – 1.01) | 7747 | 1.00 (0.99 – 1.01) |
| 300.11 | Generalized anxiety disorder | Per88 | 1.6 | 1.00 (0.98 – 1.01) | 8.5E-01 | 8948 | 0.99 (0.96 – 1.01) | 4442 | 1.00 (0.98 – 1.01) |
| 481 | Influenza | Per88 | 1.5 | 1.00 (0.98 – 1.03) | 8.5E-01 | 9080 | 1.01 (0.98 – 1.03) | 4391 | 1.00 (0.98 – 1.03) |
| 401.1 | Essential hypertension | Per88 | 2.8 | 1.00 (0.98 – 1.03) | 8.6E-01 | 5799 | 1.00 (0.98 – 1.03) | 2253 | 1.00 (0.98 – 1.03) |
| 798 | Malaise and fatigue | AHI 3% | 2.5 | 1.00 (0.99 – 1.01) | 8.6E-01 | 12781 | 1.00 (0.99 – 1.00) | 6748 | 1.00 (0.99 – 1.01) |
| 530.11 | GERD | Per88 | 3.9 | 1.00 (0.98 – 1.01) | 8.6E-01 | 6759 | 1.01 (0.99 – 1.02) | 3161 | 1.00 (0.98 – 1.01) |
| 743.11 | Osteoporosis NOS | AHI 3% | 1.3 | 1.00 (0.99 – 1.01) | 8.7E-01 | 14481 | 1.00 (0.99 – 1.00) | 7482 | 1.00 (0.99 – 1.01) |
| 459.9 | Circulatory disease NEC | Per88 | 1.5 | 1.00 (0.98 – 1.03) | 9.0E-01 | 9377 | 1.00 (0.98 – 1.02) | 4514 | 1.00 (0.98 – 1.03) |
| 704.1 | Alopecia | AHI 3% | 1.5 | 1.00 (0.99 – 1.01) | 9.0E-01 | 14763 | 1.00 (0.99 – 1.01) | 7730 | 1.00 (0.99 – 1.01) |
| 208 | Benign neoplasm of colon | Per88 | 1.9 | 1.00 (0.97 – 1.02) | 9.2E-01 | 8415 | 0.99 (0.96 – 1.01) | 3956 | 1.00 (0.97 – 1.02) |
| 472 | Chronic pharyngitis and nasopharyngitis | AHI 3% | 1.6 | 1.00 (0.99 – 1.01) | 9.2E-01 | 14413 | 1.00 (0.99 – 1.01) | 7468 | 1.00 (0.99 – 1.01) |
| 727.1 | Synovitis and tenosynovitis | Per88 | 1.3 | 1.00 (0.98 – 1.03) | 9.2E-01 | 9191 | 1.00 (0.98 – 1.02) | 4436 | 1.00 (0.98 – 1.03) |
| 593.2 | Microscopic hematuria | AHI 3% | 1.2 | 1.00 (0.99 – 1.01) | 9.2E-01 | 14947 | 1.01 (1.00 – 1.01) | 7730 | 1.00 (0.99 – 1.01) |
| 386.9 | Dizziness and giddiness (Light-headedness and vertigo) | Per88 | 2.1 | 1.00 (0.98 – 1.02) | 9.2E-01 | 8415 | 1.00 (0.98 – 1.01) | 4091 | 1.00 (0.98 – 1.02) |
| 565.1 | Anal and rectal polyp | Per88 | 1.3 | 1.00 (0.98 – 1.03) | 9.2E-01 | 9431 | 0.99 (0.96 – 1.02) | 4549 | 1.00 (0.98 – 1.03) |
| 765 | Cervical radiculitis | Per88 | 1.1 | 1.00 (0.98 – 1.02) | 9.3E-01 | 9353 | 1.01 (0.99 – 1.03) | 4530 | 1.00 (0.98 – 1.02) |
| 402 | Elevated blood pressure reading without diagnosis of hypertension | AHI 3% | 1.7 | 1.00 (0.99 – 1.01) | 9.3E-01 | 14115 | 1.00 (0.99 – 1.01) | 7260 | 1.00 (0.99 – 1.01) |
| 369.5 | Conjunctivitis, infectious | AHI 3% | 1.2 | 1.00 (0.99 – 1.01) | 9.5E-01 | 14790 | 1.00 (0.99 – 1.01) | 7679 | 1.00 (0.99 – 1.01) |
| 292 | Neurological disorders | AHI 3% | 1.2 | 1.00 (0.99 – 1.01) | 9.5E-01 | 15056 | 1.00 (0.99 – 1.01) | 7813 | 1.00 (0.99 – 1.01) |
| 379.9 | Pain, swelling or discharge of eye | AHI 3% | 1.1 | 1.00 (0.99 – 1.01) | 9.7E-01 | 14916 | 1.00 (0.99 – 1.01) | 7743 | 1.00 (0.99 – 1.01) |
| 429.3 | Symptoms involving cardiovascular system | Per88 | 2.3 | 1.00 (0.98 – 1.02) | 9.8E-01 | 9191 | 1.00 (0.99 – 1.02) | 4431 | 1.00 (0.98 – 1.02) |
| 285 | Other anemias | AHI 3% | 2.6 | 1.00 (0.99 – 1.01) | 9.9E-01 | 13445 | 1.00 (1.00 – 1.01) | 6944 | 1.00 (0.99 – 1.01) |
| 389.1 | Sensorineural hearing loss | AHI 3% | 1.1 | 1.00 (0.99 – 1.01) | 9.9E-01 | 14658 | 1.00 (0.99 – 1.01) | 7528 | 1.00 (0.99 – 1.01) |
| 727.1 | Synovitis and tenosynovitis | AHI 3% | 1.3 | 1.00 (0.99 – 1.01) | 9.9E-01 | 14689 | 1.00 (0.99 – 1.01) | 7586 | 1.00 (0.99 – 1.01) |
| 599.8 | Other symptoms involving urinary | Per88 | 1.4 | 1.00 (0.98 – 1.02) | 9.9E-01 | 9490 | 1.00 (0.98 – 1.03) | 4608 | 1.00 (0.98 – 1.02) |

| PheCode | Disease | Sleep statistic | Incidence | Risk ratio | P-value | N | Risk ratio (AHI ≥15) | N (AHI ≥15) | Risk ratio (adj sleep disorders) |
|---|---|---|---|---|---|---|---|---|---|
| | system | | | | | | | | |

#### Table S10. Incident sleep statistic associations with comorbidities sensitivity analyses.

Incident analyses considered diagnoses at least one year after the first sleep recording (prioritizing diagnostics, then splits, then home sleep tests) and are ordered by p-values. Primary analyses considered joint-sex PheCodes with >1% incidence rate and were rank-normalized after adjusting for age, sex, BMI, population, sleep study type, and healthcare encounters. The primary analysis considered all available sleep laboratory data (i.e. including participants being screened for other sleep disorders).

1 PheCode was Bonferroni-level significant (**bolded**) and FDR-level significant (in red, uncorrected p-values).

Due to PDF width constraints, sensitivity analyses are presented in two tables, with primary columns repeated (apart from incidence and n):

**Table S9 further includes:**
A sensitivity analysis considering only participants with AHI >15.
A sensitivity analysis further adjusting for an indicator variable for other common sleep disorders (PheCodes 327, 327.1, 327.4, 327.41, 327.5, 327.6, 327.7, 327.71) in the full sleep recording sample.

**Table S10 further includes:**
A sensitivity analysis further adjusting for an indicator variable for common pulmonary disorders (PheCodes 415.1, 415.11, 415.2, 480.1, 480.11, 480.2, 480, 495, 495.2, 496.1, 496.2, 496.21, 496.3, 497, 502, 503, 505, 506, 509.2).
P-values from sleep statistic × sex interaction analyses (not rank-normalized) performed separately.
Equivalent sex-stratified results.

| PheCode | Disease | Sleep statistic | Risk ratio | P-value | Risk ratio (adj pulmonary disorders) | Sleep statistic x sex interaction p-value | Risk ratio (men) | Risk ratio (women) |
|---|---|---|---|---|---|---|---|---|
| 350.1 | Abnormal involuntary movements | AHI 3% | 0.99 (0.98 – 0.99) | **2.0E-04** | 0.99 (0.98 – 0.99) | 8.0E-01 | 0.98 (0.97 – 0.99) | 0.99 (0.98 – 1.00) |
| 427.9 | Palpitations | AHI 3% | 0.99 (0.98 – 1.00) | 8.1E-04 | 0.99 (0.98 – 1.00) | 3.4E-01 | 0.98 (0.97 – 1.00) | 0.99 (0.98 – 1.00) |
| 285 | Other anemias | Per88 | 1.02 (1.01 – 1.04) | 1.4E-03 | 1.02 (1.01 – 1.04) | 3.9E-01 | 1.01 (0.99 – 1.04) | 1.03 (1.01 – 1.05) |
| 530.9 | Heartburn | Per88 | 0.97 (0.95 – 0.99) | 2.1E-03 | 0.97 (0.95 – 0.99) | 9.2E-01 | 0.98 (0.95 – 1.01) | 0.97 (0.94 – 0.99) |
| 508 | Pulmonary collapse; interstitial and compensatory emphysema | Per88 | 1.03 (1.01 – 1.06) | 2.8E-03 | 1.03 (1.01 – 1.06) | 6.7E-01 | 1.02 (1.00 – 1.05) | 1.05 (1.02 – 1.08) |
| 585.3 | Chronic renal failure [CKD] | Per88 | 1.03 (1.01 – 1.05) | 3.8E-03 | 1.03 (1.01 – 1.05) | 6.7E-01 | 1.03 (1.00 – 1.06) | 1.03 (1.00 – 1.07) |
| 519.8 | Other diseases of respiratory system, NEC | Per88 | 1.03 (1.01 – 1.05) | 4.3E-03 | 1.02 (1.01 – 1.04) | 6.2E-01 | 1.02 (0.98 – 1.05) | 1.03 (1.01 – 1.06) |
| 292.3 | Memory loss | AHI 3% | 0.99 (0.98 – 1.00) | 4.6E-03 | 0.99 (0.98 – 1.00) | 7.7E-03 | 0.99 (0.98 – 1.00) | 0.99 (0.98 – 1.00) |
| 512.9 | Other dyspnea | Per88 | 1.03 (1.01 – 1.05) | 6.1E-03 | 1.02 (1.00 – 1.05) | 6.8E-01 | 1.03 (1.01 – 1.06) | 1.02 (0.98 – 1.06) |
| 760 | Back pain | AHI 3% | 0.99 (0.98 – 1.00) | 6.3E-03 | 0.99 (0.98 – 1.00) | 3.7E-01 | 0.99 (0.98 – 1.00) | 0.98 (0.97 – 1.00) |

| PheCode | Disease | Sleep statistic | Risk ratio | P-value | Risk ratio (adj pulmonary disorders) | Sleep statistic x sex interaction p-value | Risk ratio (men) | Risk ratio (women) |
|---|---|---|---|---|---|---|---|---|
| 340 | Migraine | AHI 3% | 0.99 (0.99 – 1.00) | 8.0E-03 | 0.99 (0.99 – 1.00) | 6.8E-01 | 0.99 (0.98 – 1.00) | 0.99 (0.98 – 1.00) |
| 840 | Sprains and strains | AHI 3% | 0.99 (0.99 – 1.00) | 1.0E-02 | 0.99 (0.99 – 1.00) | 3.1E-01 | 0.99 (0.99 – 1.00) | 0.99 (0.98 – 1.00) |
| 382 | Otalgia | Per88 | 0.98 (0.96 – 0.99) | 1.0E-02 | 0.98 (0.96 – 0.99) | 9.4E-01 | 0.97 (0.93 – 1.01) | 0.98 (0.96 – 1.00) |
| 455 | Hemorrhoids | AHI 3% | 0.99 (0.99 – 1.00) | 1.5E-02 | 0.99 (0.99 – 1.00) | 6.7E-02 | 1.00 (0.99 – 1.00) | 0.99 (0.98 – 1.00) |
| 599.9 | Other abnormality of urination | AHI 3% | 0.99 (0.98 – 1.00) | 1.7E-02 | 0.99 (0.98 – 1.00) | 5.1E-01 | 0.99 (0.98 – 1.00) | 1.00 (0.98 – 1.01) |
| 599.5 | Frequency of urination and polyuria | AHI 3% | 0.99 (0.98 – 1.00) | 1.8E-02 | 0.99 (0.98 – 1.00) | 7.3E-01 | 0.99 (0.99 – 1.00) | 1.00 (0.98 – 1.01) |
| 781 | Symptoms involving nervous and musculoskeletal systems | Per88 | 1.02 (1.00 – 1.04) | 2.0E-02 | 1.02 (1.00 – 1.04) | 1.4E-01 | 1.03 (1.01 – 1.06) | 1.01 (0.98 – 1.04) |
| 563 | Constipation | AHI 3% | 0.99 (0.99 – 1.00) | 2.1E-02 | 0.99 (0.99 – 1.00) | 4.6E-01 | 1.00 (0.99 – 1.01) | 0.99 (0.98 – 1.00) |
| 382 | Otalgia | AHI 3% | 0.99 (0.99 – 1.00) | 2.4E-02 | 0.99 (0.99 – 1.00) | 1.0E-01 | 0.99 (0.98 – 1.01) | 0.99 (0.98 – 1.00) |
| 480 | Pneumonia | Per88 | 1.02 (1.00 – 1.04) | 2.4E-02 | 1.01 (0.99 – 1.03) | 8.9E-01 | 1.03 (1.00 – 1.05) | 1.01 (0.98 – 1.04) |
| 512.9 | Other dyspnea | AHI 3% | 1.01 (1.00 – 1.02) | 2.7E-02 | 1.01 (1.00 – 1.02) | 8.3E-01 | 1.01 (1.00 – 1.02) | 1.01 (1.00 – 1.03) |
| 465 | Acute upper respiratory infections of multiple or unspecified sites | AHI 3% | 1.01 (1.00 – 1.02) | 3.2E-02 | 1.01 (1.00 – 1.02) | 5.7E-01 | 1.01 (1.00 – 1.02) | 1.01 (0.99 – 1.02) |
| 599.4 | Urinary incontinence | AHI 3% | 0.99 (0.98 – 1.00) | 3.2E-02 | 0.99 (0.98 – 1.00) | 8.6E-01 | 0.99 (0.98 – 1.00) | 0.99 (0.98 – 1.00) |
| 561 | Symptoms involving digestive system | AHI 3% | 0.99 (0.98 – 1.00) | 3.3E-02 | 0.99 (0.98 – 1.00) | 2.1E-01 | 0.99 (0.98 – 1.00) | 0.99 (0.98 – 1.00) |
| 720 | Spinal stenosis | AHI 3% | 0.99 (0.98 – 1.00) | 3.8E-02 | 0.99 (0.98 – 1.00) | 4.5E-01 | 0.99 (0.98 – 1.00) | 0.99 (0.98 – 1.01) |
| 173 | Neoplasm of uncertain behavior of skin | Per88 | 0.98 (0.96 – 1.00) | 3.9E-02 | 0.98 (0.96 – 1.00) | 4.0E-01 | 0.96 (0.93 – 0.99) | 0.99 (0.97 – 1.02) |
| 272.13 | Mixed hyperlipidemia | AHI 3% | 1.01 (1.00 – 1.02) | 4.1E-02 | 1.01 (1.00 – 1.02) | 3.4E-01 | 1.01 (1.00 – 1.02) | 1.00 (0.99 – 1.02) |
| 350.1 | Abnormal involuntary movements | Per88 | 0.98 (0.96 – 1.00) | 4.3E-02 | 0.98 (0.95 – 1.00) | 5.7E-01 | 0.98 (0.95 – 1.02) | 0.98 (0.95 – 1.01) |
| 278.4 | Abnormal weight gain | AHI 3% | 0.99 (0.98 – 1.00) | 4.6E-02 | 0.99 (0.98 – 1.00) | 5.5E-02 | 1.00 (0.98 – 1.01) | 0.99 (0.98 – 1.00) |
| 726.1 | Enthesopathy | AHI 3% | 0.99 (0.99 – 1.00) | 4.6E-02 | 0.99 (0.99 – 1.00) | 8.2E-01 | 0.99 (0.98 – 1.00) | 1.00 (0.99 – 1.00) |
| 300.11 | Generalized anxiety disorder | AHI 3% | 0.99 (0.99 – 1.00) | 5.1E-02 | 0.99 (0.99 – 1.00) | 1.3E-01 | 0.99 (0.98 – 1.00) | 1.00 (0.99 – 1.01) |
| 136 | Other infectious and parasitic diseases | Per88 | 1.02 (1.00 – 1.03) | 5.3E-02 | 1.01 (1.00 – 1.03) | 4.4E-03 | 1.05 (1.02 – 1.08) | 1.00 (0.98 – 1.02) |
| 292.3 | Memory loss | Per88 | 0.98 (0.96 – 1.00) | 5.7E-02 | 0.98 (0.96 – 1.00) | 7.1E-01 | 0.98 (0.96 – 1.01) | 0.98 (0.94 – 1.01) |
| 740.9 | Osteoarthrosis NOS | AHI 3% | 0.99 (0.98 – 1.00) | 6.1E-02 | 0.99 (0.98 – 1.00) | 7.2E-01 | 1.00 (0.99 – 1.01) | 0.99 (0.97 – 1.00) |
| 357 | Inflammatory and toxic neuropathy | AHI 3% | 0.99 (0.98 – 1.00) | 6.3E-02 | 0.99 (0.98 – 1.00) | 9.1E-02 | 0.98 (0.97 – 1.00) | 1.00 (0.99 – 1.01) |
| 599.5 | Frequency of urination and polyuria | Per88 | 0.98 (0.96 – 1.00) | 6.4E-02 | 0.98 (0.96 – 1.00) | 9.0E-01 | 0.99 (0.97 – 1.01) | 0.99 (0.95 – 1.02) |
| 530.11 | GERD | AHI 3% | 1.00 (0.99 – 1.00) | 6.7E-02 | 1.00 (0.99 – 1.00) | 7.6E-01 | 1.00 (0.99 – 1.00) | 0.99 (0.99 – 1.00) |
| 427.3 | Other specified cardiac dysrhythmias | AHI 3% | 0.99 (0.99 – 1.00) | 7.6E-02 | 0.99 (0.99 – 1.00) | 5.4E-01 | 0.99 (0.99 – 1.00) | 1.00 (0.99 – 1.02) |
| 760 | Back pain | Per88 | 1.01 (1.00 – 1.03) | 8.1E-02 | 1.01 (1.00 – 1.03) | 3.5E-01 | 1.02 (1.00 – 1.04) | 1.01 (0.99 – 1.04) |
| 742.9 | Other derangement of joint | Per88 | 0.98 (0.97 – 1.00) | 8.6E-02 | 0.98 (0.97 – 1.00) | 8.1E-01 | 0.98 (0.94 – 1.02) | 0.98 (0.96 – 1.00) |
| 262 | Mineral deficiency NEC | AHI 3% | 0.99 (0.99 – 1.00) | 8.9E-02 | 0.99 (0.99 – 1.00) | 5.1E-01 | 0.99 (0.98 – 1.01) | 0.99 (0.98 – 1.00) |
| 208 | Benign neoplasm of colon | AHI 3% | 0.99 (0.99 – 1.00) | 9.7E-02 | 0.99 (0.99 – 1.00) | 6.7E-01 | 0.99 (0.99 – 1.00) | 1.00 (0.99 – 1.01) |
| 276.5 | Hypovolemia | Per88 | 1.02 (1.00 – 1.04) | 9.9E-02 | 1.02 (0.99 – 1.04) | 6.1E-01 | 1.01 (0.98 – 1.04) | 1.03 (1.00 – 1.06) |
| 689 | Disorder of skin and subcutaneous tissue NOS | Per88 | 0.99 (0.98 – 1.00) | 1.1E-01 | 0.99 (0.98 – 1.00) | 8.3E-01 | 1.00 (0.98 – 1.01) | 0.99 (0.97 – 1.01) |

| PheCode | Disease | Sleep statistic | Risk ratio | P-value | Risk ratio (adj pulmonary disorders) | Sleep statistic x sex interaction p-value | Risk ratio (men) | Risk ratio (women) |
|---|---|---|---|---|---|---|---|---|
| 366.2 | Senile cataract | AHI 3% | 0.99 (0.99 – 1.00) | 1.2E-01 | 0.99 (0.99 – 1.00) | 3.1E-01 | 0.99 (0.98 – 1.00) | 0.99 (0.98 – 1.01) |
| 338.2 | Chronic pain | AHI 3% | 1.00 (0.99 – 1.00) | 1.2E-01 | 1.00 (0.99 – 1.00) | 3.9E-01 | 1.00 (0.99 – 1.00) | 1.00 (0.99 – 1.00) |
| 476 | Allergic rhinitis | Per88 | 0.99 (0.97 – 1.00) | 1.3E-01 | 0.98 (0.96 – 1.00) | 8.4E-01 | 0.98 (0.95 – 1.01) | 0.98 (0.96 – 1.01) |
| 550.2 | Diaphragmatic hernia | AHI 3% | 0.99 (0.99 – 1.00) | 1.4E-01 | 0.99 (0.99 – 1.00) | 9.9E-01 | 0.99 (0.98 – 1.01) | 0.99 (0.98 – 1.00) |
| 701.2 | Scar conditions and fibrosis of skin | Per88 | 0.98 (0.96 – 1.01) | 1.5E-01 | 0.98 (0.96 – 1.00) | 4.9E-01 | 0.97 (0.93 – 1.00) | 1.00 (0.96 – 1.03) |
| 365 | Glaucoma | Per88 | 0.98 (0.95 – 1.01) | 1.5E-01 | 0.98 (0.95 – 1.01) | 1.5E-01 | 0.98 (0.95 – 1.01) | 0.98 (0.94 – 1.02) |
| 495 | Asthma | Per88 | 1.01 (1.00 – 1.03) | 1.5E-01 | 1.00 (0.98 – 1.03) | 7.0E-01 | 1.00 (0.97 – 1.04) | 1.02 (1.00 – 1.04) |
| 687.1 | Rash and other nonspecific skin eruption | Per88 | 1.01 (1.00 – 1.02) | 1.5E-01 | 1.01 (0.99 – 1.02) | 8.5E-01 | 1.00 (0.97 – 1.03) | 1.01 (1.00 – 1.03) |
| 480 | Pneumonia | AHI 3% | 1.00 (1.00 – 1.01) | 1.5E-01 | 1.00 (1.00 – 1.01) | 1.2E-01 | 1.01 (1.00 – 1.02) | 1.00 (0.99 – 1.01) |
| 402 | Elevated blood pressure reading without diagnosis of hypertension | Per88 | 0.98 (0.96 – 1.01) | 1.6E-01 | 0.99 (0.97 – 1.01) | 2.6E-01 | 0.98 (0.95 – 1.02) | 0.98 (0.96 – 1.01) |
| 701 | Other hypertrophic and atrophic conditions of skin | Per88 | 0.99 (0.97 – 1.00) | 1.7E-01 | 0.99 (0.97 – 1.00) | 2.1E-01 | 0.97 (0.95 – 1.00) | 1.00 (0.98 – 1.02) |
| 375.1 | Dry eyes | AHI 3% | 1.00 (0.99 – 1.00) | 1.8E-01 | 1.00 (0.99 – 1.00) | 3.7E-01 | 1.00 (0.99 – 1.01) | 0.99 (0.98 – 1.00) |
| 743.11 | Osteoporosis NOS | Per88 | 1.01 (0.99 – 1.03) | 1.8E-01 | 1.01 (0.99 – 1.03) | 8.3E-01 | 1.02 (0.98 – 1.06) | 1.00 (0.97 – 1.03) |
| 472 | Chronic pharyngitis and nasopharyngitis | Per88 | 1.01 (0.99 – 1.03) | 1.8E-01 | 1.01 (0.99 – 1.03) | 4.1E-01 | 1.00 (0.98 – 1.03) | 1.02 (1.00 – 1.04) |
| 726 | Peripheral enthesopathies and allied syndromes | AHI 3% | 1.00 (0.99 – 1.00) | 1.9E-01 | 1.00 (0.99 – 1.00) | 6.9E-01 | 1.00 (0.99 – 1.01) | 0.99 (0.98 – 1.00) |
| 304 | Adjustment reaction | AHI 3% | 1.00 (0.99 – 1.00) | 2.0E-01 | 1.00 (0.99 – 1.00) | 8.0E-01 | 0.99 (0.98 – 1.00) | 1.00 (0.99 – 1.01) |
| 280.1 | Iron deficiency anemias, unspecified or not due to blood loss | AHI 3% | 0.99 (0.99 – 1.00) | 2.0E-01 | 0.99 (0.99 – 1.00) | 2.4E-01 | 0.99 (0.98 – 1.01) | 1.00 (0.99 – 1.01) |
| 764 | Sciatica | AHI 3% | 1.00 (0.99 – 1.00) | 2.0E-01 | 1.00 (0.99 – 1.00) | 2.9E-02 | 1.00 (0.99 – 1.01) | 0.99 (0.98 – 1.00) |
| 389 | Hearing loss | Per88 | 1.01 (0.99 – 1.03) | 2.0E-01 | 1.01 (1.00 – 1.03) | 6.5E-01 | 1.01 (0.99 – 1.03) | 1.01 (0.99 – 1.04) |
| 550.2 | Diaphragmatic hernia | Per88 | 0.98 (0.96 – 1.01) | 2.1E-01 | 0.98 (0.96 – 1.01) | 1.8E-01 | 1.00 (0.94 – 1.05) | 0.97 (0.94 – 1.01) |
| 366.2 | Senile cataract | Per88 | 0.99 (0.96 – 1.01) | 2.1E-01 | 0.98 (0.96 – 1.01) | 2.1E-01 | 1.00 (0.98 – 1.03) | 0.96 (0.93 – 1.00) |
| 594.1 | Calculus of kidney | AHI 3% | 1.00 (1.00 – 1.01) | 2.2E-01 | 1.00 (1.00 – 1.01) | 1.4E-01 | 1.01 (1.00 – 1.02) | 1.00 (0.98 – 1.01) |
| 571.5 | Other chronic nonalcoholic liver disease | Per88 | 0.98 (0.95 – 1.01) | 2.3E-01 | 0.98 (0.95 – 1.01) | 7.0E-02 | 0.96 (0.91 – 1.01) | 0.99 (0.95 – 1.04) |
| 565.1 | Anal and rectal polyp | AHI 3% | 1.00 (1.00 – 1.01) | 2.3E-01 | 1.00 (1.00 – 1.01) | 6.0E-01 | 1.00 (1.00 – 1.01) | 1.00 (0.99 – 1.02) |
| 706.8 | Other specified diseases of sebaceous glands | AHI 3% | 1.00 (0.99 – 1.00) | 2.4E-01 | 1.00 (0.99 – 1.00) | 5.4E-01 | 0.99 (0.98 – 1.00) | 1.00 (0.99 – 1.01) |
| 401.1 | Essential hypertension | AHI 3% | 1.00 (1.00 – 1.01) | 2.4E-01 | 1.00 (1.00 – 1.01) | 7.7E-01 | 1.00 (0.99 – 1.01) | 1.01 (0.99 – 1.02) |
| 781 | Symptoms involving nervous and musculoskeletal systems | AHI 3% | 1.00 (1.00 – 1.01) | 2.5E-01 | 1.00 (1.00 – 1.01) | 1.1E-02 | 1.01 (1.00 – 1.02) | 0.99 (0.98 – 1.01) |
| 579.8 | Nonspecific abnormal findings in stool contents | Per88 | 0.99 (0.97 – 1.01) | 2.6E-01 | 0.99 (0.97 – 1.01) | 5.1E-01 | 0.99 (0.96 – 1.02) | 0.98 (0.96 – 1.01) |
| 272.13 | Mixed hyperlipidemia | Per88 | 0.98 (0.95 – 1.01) | 2.7E-01 | 0.98 (0.95 – 1.01) | 5.8E-01 | 1.00 (0.96 – 1.03) | 0.97 (0.92 – 1.02) |
| 272.1 | Hyperlipidemia | AHI 3% | 1.00 (0.99 – 1.00) | 2.7E-01 | 1.00 (0.99 – 1.00) | 2.5E-01 | 1.00 (0.99 – 1.01) | 1.00 (0.99 – 1.00) |
| 563 | Constipation | Per88 | 1.01 (0.99 – 1.02) | 2.7E-01 | 1.01 (0.99 – 1.02) | 6.7E-01 | 1.00 (0.98 – 1.03) | 1.01 (0.99 – 1.03) |
| 465 | Acute upper respiratory infections of multiple or | Per88 | 1.01 (0.99 – 1.03) | 2.8E-01 | 1.01 (0.99 – 1.03) | 8.4E-01 | 1.03 (1.00 – 1.05) | 1.00 (0.97 – 1.02) |

| PheCode | Disease | Sleep statistic | Risk ratio | P-value | Risk ratio (adj pulmonary disorders) | Sleep statistic x sex interaction p-value | Risk ratio (men) | Risk ratio (women) |
|---|---|---|---|---|---|---|---|---|
| | unspecified sites | | | | | | | |
| 519.8 | Other diseases of respiratory system, NEC | AHI 3% | 1.00 (1.00 – 1.01) | 2.9E-01 | 1.00 (1.00 – 1.01) | 5.4E-01 | 1.01 (1.00 – 1.02) | 1.00 (0.99 – 1.01) |
| 689 | Disorder of skin and subcutaneous tissue NOS | AHI 3% | 1.00 (0.99 – 1.00) | 2.9E-01 | 1.00 (0.99 – 1.00) | 7.7E-01 | 1.00 (0.99 – 1.00) | 1.00 (0.99 – 1.01) |
| 306 | Other mental disorder | Per88 | 1.01 (0.99 – 1.02) | 3.0E-01 | 1.01 (0.99 – 1.02) | 5.0E-01 | 1.01 (1.00 – 1.03) | 1.00 (0.98 – 1.02) |
| 365 | Glaucoma | AHI 3% | 1.00 (0.99 – 1.00) | 3.0E-01 | 1.00 (0.99 – 1.00) | 9.1E-01 | 1.00 (0.99 – 1.01) | 0.99 (0.98 – 1.01) |
| 386.9 | Dizziness and giddiness (Light-headedness and vertigo) | AHI 3% | 1.00 (1.00 – 1.01) | 3.0E-01 | 1.00 (1.00 – 1.01) | 3.6E-01 | 1.00 (0.99 – 1.01) | 1.00 (0.99 – 1.01) |
| 427.9 | Palpitations | Per88 | 0.99 (0.97 – 1.01) | 3.0E-01 | 0.99 (0.97 – 1.01) | 1.3E-01 | 0.97 (0.94 – 1.01) | 0.99 (0.97 – 1.02) |
| 593.2 | Microscopic hematuria | Per88 | 0.99 (0.97 – 1.01) | 3.0E-01 | 0.99 (0.97 – 1.01) | 5.5E-01 | 0.99 (0.96 – 1.02) | 0.99 (0.96 – 1.02) |
| 357 | Inflammatory and toxic neuropathy | Per88 | 1.01 (0.99 – 1.03) | 3.1E-01 | 1.01 (0.99 – 1.03) | 2.6E-01 | 1.02 (0.99 – 1.04) | 1.00 (0.98 – 1.03) |
| 476 | Allergic rhinitis | AHI 3% | 1.00 (0.99 – 1.00) | 3.2E-01 | 1.00 (0.99 – 1.00) | 6.4E-01 | 0.99 (0.98 – 1.00) | 1.00 (0.99 – 1.01) |
| 300.1 | Anxiety disorder | Per88 | 1.01 (0.99 – 1.02) | 3.2E-01 | 1.01 (0.99 – 1.02) | 5.8E-02 | 1.01 (0.99 – 1.04) | 1.01 (0.99 – 1.02) |
| 379.9 | Pain, swelling or discharge of eye | Per88 | 0.99 (0.97 – 1.01) | 3.3E-01 | 0.99 (0.97 – 1.01) | 5.3E-02 | 0.95 (0.92 – 0.98) | 1.00 (0.98 – 1.03) |
| 272.11 | Hypercholesterolemia | Per88 | 1.01 (0.99 – 1.03) | 3.4E-01 | 1.01 (0.99 – 1.03) | 2.8E-01 | 1.02 (1.00 – 1.04) | 0.99 (0.95 – 1.02) |
| 427.6 | Premature beats | Per88 | 1.01 (0.99 – 1.03) | 3.4E-01 | 1.01 (0.99 – 1.03) | 8.0E-01 | 1.01 (0.99 – 1.04) | 1.01 (0.96 – 1.05) |
| 389.4 | Tinnitus | AHI 3% | 1.00 (0.99 – 1.00) | 3.5E-01 | 1.00 (0.99 – 1.00) | 6.6E-01 | 1.00 (0.99 – 1.01) | 1.00 (0.99 – 1.01) |
| 704 | Diseases of hair and hair follicles | AHI 3% | 1.00 (0.99 – 1.00) | 3.7E-01 | 1.00 (0.99 – 1.00) | 2.2E-01 | 1.00 (0.99 – 1.01) | 0.99 (0.98 – 1.00) |
| 340 | Migraine | Per88 | 0.99 (0.98 – 1.01) | 3.7E-01 | 0.99 (0.98 – 1.01) | 1.4E-01 | 0.98 (0.95 – 1.00) | 1.00 (0.98 – 1.02) |
| 798 | Malaise and fatigue | Per88 | 1.01 (0.99 – 1.02) | 3.8E-01 | 1.01 (0.99 – 1.02) | 5.9E-01 | 1.01 (0.99 – 1.03) | 1.00 (0.98 – 1.03) |
| 278.11 | Morbid obesity | AHI 3% | 1.00 (1.00 – 1.01) | 3.8E-01 | 1.00 (1.00 – 1.01) | 5.6E-01 | 1.00 (0.99 – 1.01) | 1.00 (0.99 – 1.02) |
| 455 | Hemorrhoids | Per88 | 0.99 (0.97 – 1.01) | 3.8E-01 | 0.99 (0.97 – 1.01) | 4.5E-02 | 1.01 (0.98 – 1.04) | 0.98 (0.95 – 1.01) |
| 316 | Substance addiction and disorders | Per88 | 1.01 (0.99 – 1.03) | 3.8E-01 | 1.01 (0.98 – 1.03) | 6.7E-01 | 1.00 (0.97 – 1.03) | 1.02 (0.99 – 1.05) |
| 278.11 | Morbid obesity | Per88 | 1.01 (0.99 – 1.04) | 3.9E-01 | 1.01 (0.99 – 1.04) | 1.7E-01 | 0.99 (0.94 – 1.05) | 1.01 (0.98 – 1.05) |
| 470 | Septal Deviations/Turbinate Hypertrophy | AHI 3% | 1.00 (1.00 – 1.01) | 3.9E-01 | 1.00 (1.00 – 1.01) | 7.2E-02 | 1.00 (0.99 – 1.01) | 1.01 (1.00 – 1.02) |
| 278.1 | Obesity | Per88 | 1.01 (0.99 – 1.03) | 4.0E-01 | 1.01 (0.99 – 1.03) | 3.0E-01 | 1.01 (0.97 – 1.05) | 1.00 (0.98 – 1.03) |
| 740.9 | Osteoarthrosis NOS | Per88 | 0.99 (0.96 – 1.01) | 4.0E-01 | 0.99 (0.96 – 1.01) | 1.6E-01 | 0.97 (0.93 – 1.00) | 1.00 (0.97 – 1.04) |
| 561 | Symptoms involving digestive system | Per88 | 0.99 (0.97 – 1.01) | 4.0E-01 | 0.99 (0.97 – 1.01) | 4.0E-01 | 1.00 (0.98 – 1.03) | 0.97 (0.93 – 1.01) |
| 327.4 | Insomnia | Per88 | 1.01 (0.99 – 1.02) | 4.0E-01 | 1.01 (0.99 – 1.02) | 9.3E-01 | 1.01 (0.99 – 1.04) | 1.01 (0.98 – 1.03) |
| 585.3 | Chronic renal failure [CKD] | AHI 3% | 1.00 (0.99 – 1.01) | 4.2E-01 | 1.00 (0.99 – 1.01) | 9.2E-02 | 1.01 (1.00 – 1.02) | 0.99 (0.98 – 1.01) |
| 687.1 | Rash and other nonspecific skin eruption | AHI 3% | 1.00 (0.99 – 1.00) | 4.2E-01 | 1.00 (0.99 – 1.00) | 8.8E-01 | 0.99 (0.99 – 1.00) | 1.00 (0.99 – 1.01) |
| 475.9 | Postnasal drip | Per88 | 1.01 (0.99 – 1.02) | 4.3E-01 | 1.01 (0.99 – 1.02) | 9.8E-01 | 1.01 (0.98 – 1.04) | 1.01 (0.98 – 1.03) |
| 278 | Overweight, obesity and other hyperalimentation | Per88 | 0.99 (0.98 – 1.01) | 4.3E-01 | 0.99 (0.98 – 1.01) | 5.0E-01 | 1.00 (0.98 – 1.02) | 0.99 (0.96 – 1.01) |
| 280.1 | Iron deficiency anemias, unspecified or not due to blood loss | Per88 | 0.99 (0.97 – 1.02) | 4.4E-01 | 0.99 (0.96 – 1.01) | 9.0E-01 | 0.98 (0.93 – 1.03) | 1.00 (0.97 – 1.03) |
| 809 | Fracture of unspecified bones | AHI 3% | 1.00 (0.99 – 1.00) | 4.4E-01 | 1.00 (0.99 – 1.00) | 2.5E-01 | 0.99 (0.98 – 1.01) | 1.00 (0.98 – 1.01) |
| 277 | Other disorders of metabolism | Per88 | 0.99 (0.98 – 1.01) | 4.4E-01 | 0.99 (0.98 – 1.01) | 5.2E-03 | 1.02 (0.99 – 1.05) | 0.98 (0.96 – 1.00) |

| PheCode | Disease | Sleep statistic | Risk ratio | P-value | Risk ratio (adj pulmonary disorders) | Sleep statistic x sex interaction p-value | Risk ratio (men) | Risk ratio (women) |
|---|---|---|---|---|---|---|---|---|
| 761 | Cervicalgia | AHI 3% | 1.00 (1.00 – 1.01) | 4.5E-01 | 1.00 (1.00 – 1.01) | 6.7E-01 | 1.00 (0.99 – 1.01) | 1.00 (0.99 – 1.01) |
| 304 | Adjustment reaction | Per88 | 1.01 (0.99 – 1.02) | 4.5E-01 | 1.01 (0.99 – 1.02) | 9.9E-01 | 0.99 (0.96 – 1.02) | 1.01 (0.99 – 1.03) |
| 761 | Cervicalgia | Per88 | 0.99 (0.98 – 1.01) | 4.6E-01 | 0.99 (0.98 – 1.01) | 9.3E-01 | 0.99 (0.97 – 1.02) | 0.98 (0.96 – 1.01) |
| 369.5 | Conjunctivitis, infectious | Per88 | 1.01 (0.99 – 1.02) | 4.6E-01 | 1.00 (0.99 – 1.02) | 6.0E-02 | 1.03 (1.00 – 1.05) | 0.98 (0.95 – 1.01) |
| 316 | Substance addiction and disorders | AHI 3% | 1.00 (0.99 – 1.01) | 4.6E-01 | 1.00 (0.99 – 1.01) | 5.2E-01 | 1.00 (0.98 – 1.01) | 1.00 (0.98 – 1.01) |
| 389 | Hearing loss | AHI 3% | 1.00 (1.00 – 1.01) | 4.8E-01 | 1.00 (1.00 – 1.01) | 7.9E-01 | 1.00 (0.99 – 1.01) | 1.01 (1.00 – 1.01) |
| 306 | Other mental disorder | AHI 3% | 1.00 (1.00 – 1.01) | 4.8E-01 | 1.00 (1.00 – 1.01) | 9.7E-01 | 1.00 (1.00 – 1.01) | 1.00 (0.99 – 1.01) |
| 196 | Radiotherapy | AHI 3% | 1.00 (0.99 – 1.01) | 4.8E-01 | 1.00 (0.99 – 1.01) | 9.9E-01 | 1.00 (1.00 – 1.01) | 1.00 (0.99 – 1.02) |
| 173 | Neoplasm of uncertain behavior of skin | AHI 3% | 1.00 (1.00 – 1.01) | 4.9E-01 | 1.00 (1.00 – 1.01) | 1.8E-01 | 1.00 (1.00 – 1.01) | 1.00 (0.99 – 1.01) |
| 742.9 | Other derangement of joint | AHI 3% | 1.00 (0.99 – 1.00) | 5.1E-01 | 1.00 (0.99 – 1.00) | 7.1E-01 | 1.00 (0.99 – 1.01) | 0.99 (0.98 – 1.00) |
| 271.3 | Intestinal disaccharidase deficiencies and disaccharide malabsorption | Per88 | 1.00 (0.98 – 1.01) | 5.1E-01 | 1.00 (0.98 – 1.01) | 7.1E-02 | 0.98 (0.96 – 1.00) | 1.00 (0.99 – 1.02) |
| 351 | Other peripheral nerve disorders | Per88 | 1.01 (0.99 – 1.03) | 5.1E-01 | 1.01 (0.99 – 1.03) | 9.2E-02 | 1.01 (0.97 – 1.04) | 1.00 (0.98 – 1.03) |
| 292 | Neurological disorders | Per88 | 0.99 (0.97 – 1.02) | 5.3E-01 | 0.99 (0.97 – 1.02) | 9.0E-01 | 0.99 (0.96 – 1.02) | 0.99 (0.96 – 1.03) |
| 296.22 | Major depressive disorder | AHI 3% | 1.00 (0.99 – 1.00) | 5.4E-01 | 1.00 (0.99 – 1.00) | 6.6E-01 | 1.00 (0.99 – 1.01) | 1.00 (0.99 – 1.01) |
| 272.11 | Hypercholesterolemia | AHI 3% | 1.00 (0.99 – 1.00) | 5.5E-01 | 1.00 (0.99 – 1.00) | 3.0E-01 | 1.00 (0.99 – 1.01) | 0.99 (0.98 – 1.00) |
| 196 | Radiotherapy | Per88 | 0.99 (0.96 – 1.02) | 5.5E-01 | 0.99 (0.96 – 1.02) | 4.7E-01 | 1.00 (0.97 – 1.03) | 0.99 (0.94 – 1.04) |
| 530.9 | Heartburn | AHI 3% | 1.00 (0.99 – 1.00) | 5.5E-01 | 1.00 (0.99 – 1.00) | 3.1E-01 | 1.00 (0.99 – 1.01) | 1.00 (0.99 – 1.01) |
| 726 | Peripheral enthesopathies and allied syndromes | Per88 | 0.99 (0.97 – 1.02) | 5.6E-01 | 0.99 (0.96 – 1.02) | 5.6E-01 | 0.99 (0.96 – 1.03) | 1.00 (0.96 – 1.03) |
| 338.2 | Chronic pain | Per88 | 1.00 (0.98 – 1.01) | 5.6E-01 | 1.00 (0.98 – 1.01) | 5.3E-01 | 0.99 (0.97 – 1.01) | 1.00 (0.98 – 1.02) |
| 965 | Poisoning by analgesics, antipyretics, and antirheumatics | AHI 3% | 1.00 (0.99 – 1.01) | 5.8E-01 | 1.00 (0.99 – 1.01) | 9.8E-01 | 1.00 (0.99 – 1.01) | 1.00 (0.99 – 1.01) |
| 008 | Intestinal infection | Per88 | 1.00 (0.99 – 1.02) | 5.8E-01 | 1.01 (0.99 – 1.02) | 5.7E-01 | 1.01 (0.98 – 1.03) | 1.00 (0.98 – 1.03) |
| 475.9 | Postnasal drip | AHI 3% | 1.00 (0.99 – 1.00) | 5.9E-01 | 1.00 (0.99 – 1.00) | 6.8E-01 | 0.99 (0.98 – 1.00) | 1.00 (0.99 – 1.01) |
| 351 | Other peripheral nerve disorders | AHI 3% | 1.00 (0.99 – 1.01) | 6.0E-01 | 1.00 (0.99 – 1.01) | 6.5E-01 | 1.00 (0.99 – 1.01) | 1.00 (0.99 – 1.02) |
| 594.1 | Calculus of kidney | Per88 | 0.99 (0.97 – 1.02) | 6.0E-01 | 0.99 (0.97 – 1.02) | 4.6E-01 | 0.98 (0.95 – 1.02) | 1.00 (0.97 – 1.03) |
| 470 | Septal Deviations/Turbinate Hypertrophy | Per88 | 0.99 (0.97 – 1.02) | 6.0E-01 | 1.00 (0.98 – 1.02) | 4.4E-01 | 0.99 (0.96 – 1.02) | 1.01 (0.98 – 1.03) |
| 289.4 | Lymphadenitis | AHI 3% | 1.00 (0.99 – 1.01) | 6.0E-01 | 1.00 (0.99 – 1.01) | 8.0E-01 | 1.00 (0.99 – 1.01) | 1.00 (0.99 – 1.01) |
| 740.11 | Osteoarthrosis, localized, primary | Per88 | 1.01 (0.99 – 1.03) | 6.1E-01 | 1.00 (0.98 – 1.03) | 4.4E-01 | 0.99 (0.96 – 1.03) | 1.01 (0.98 – 1.04) |
| 599.2 | Retention of urine | AHI 3% | 1.00 (0.99 – 1.01) | 6.1E-01 | 1.00 (0.99 – 1.01) | 1.1E-02 | 1.00 (0.99 – 1.01) | 0.98 (0.97 – 1.00) |
| 537 | Other disorders of stomach and duodenum | Per88 | 1.00 (0.98 – 1.01) | 6.2E-01 | 0.99 (0.98 – 1.01) | 3.0E-01 | 0.99 (0.96 – 1.02) | 1.00 (0.97 – 1.02) |
| 740.11 | Osteoarthrosis, localized, primary | AHI 3% | 1.00 (0.99 – 1.01) | 6.2E-01 | 1.00 (0.99 – 1.01) | 7.7E-01 | 0.99 (0.99 – 1.00) | 1.00 (0.99 – 1.01) |
| 726.1 | Enthesopathy | Per88 | 0.99 (0.97 – 1.02) | 6.3E-01 | 0.99 (0.97 – 1.02) | 3.9E-01 | 1.01 (0.97 – 1.05) | 0.98 (0.95 – 1.01) |
| 599.2 | Retention of urine | Per88 | 0.99 (0.97 – 1.02) | 6.4E-01 | 0.99 (0.97 – 1.02) | 6.0E-02 | 0.99 (0.96 – 1.01) | 1.02 (0.98 – 1.05) |
| 809 | Fracture of unspecified bones | Per88 | 0.99 (0.97 – 1.02) | 6.4E-01 | 1.00 (0.97 – 1.02) | 2.1E-01 | 1.01 (0.98 – 1.04) | 0.98 (0.95 – 1.02) |
| 512.2 | Painful respiration | Per88 | 1.00 (0.98 – 1.01) | 6.5E-01 | 1.00 (0.98 – 1.01) | 5.4E-01 | 1.02 (0.99 – 1.05) | 0.98 (0.95 – 1.00) |

| PheCode | Disease | Sleep statistic | Risk ratio | P-value | Risk ratio (adj pulmonary disorders) | Sleep statistic x sex interaction p-value | Risk ratio (men) | Risk ratio (women) |
|---|---|---|---|---|---|---|---|---|
| 764 | Sciatica | Per88 | 1.00 (0.98 – 1.02) | 6.5E-01 | 1.00 (0.98 – 1.02) | 8.3E-03 | 1.02 (0.99 – 1.05) | 0.97 (0.95 – 1.00) |
| 296.22 | Major depressive disorder | Per88 | 1.00 (0.99 – 1.02) | 6.6E-01 | 1.00 (0.99 – 1.02) | 5.3E-02 | 0.98 (0.95 – 1.01) | 1.02 (1.00 – 1.04) |
| 537 | Other disorders of stomach and duodenum | AHI 3% | 1.00 (0.99 – 1.00) | 6.6E-01 | 1.00 (0.99 – 1.00) | 2.0E-01 | 1.00 (0.99 – 1.01) | 1.00 (0.99 – 1.01) |
| 495 | Asthma | AHI 3% | 1.00 (0.99 – 1.01) | 6.6E-01 | 1.00 (0.99 – 1.01) | 3.2E-01 | 0.99 (0.98 – 1.00) | 1.00 (0.99 – 1.01) |
| 327.4 | Insomnia | AHI 3% | 1.00 (0.99 – 1.01) | 6.6E-01 | 1.00 (0.99 – 1.01) | 3.4E-01 | 1.00 (0.99 – 1.01) | 1.00 (0.99 – 1.01) |
| 512.2 | Painful respiration | AHI 3% | 1.00 (0.99 – 1.00) | 6.7E-01 | 1.00 (0.99 – 1.00) | 9.4E-01 | 1.00 (0.99 – 1.01) | 1.00 (0.99 – 1.01) |
| 289.4 | Lymphadenitis | Per88 | 1.00 (0.97 – 1.02) | 6.7E-01 | 0.99 (0.97 – 1.02) | 8.8E-01 | 0.99 (0.96 – 1.03) | 1.00 (0.98 – 1.03) |
| 579.8 | Nonspecific abnormal findings in stool contents | AHI 3% | 1.00 (0.99 – 1.01) | 6.7E-01 | 1.00 (0.99 – 1.01) | 4.8E-01 | 1.00 (0.99 – 1.01) | 1.00 (0.99 – 1.01) |
| 599.8 | Other symptoms involving urinary system | AHI 3% | 1.00 (0.99 – 1.01) | 6.7E-01 | 1.00 (0.99 – 1.01) | 2.1E-01 | 1.00 (0.99 – 1.01) | 0.99 (0.98 – 1.01) |
| 300.1 | Anxiety disorder | AHI 3% | 1.00 (0.99 – 1.00) | 6.8E-01 | 1.00 (0.99 – 1.00) | 7.6E-01 | 1.00 (0.99 – 1.01) | 1.00 (0.99 – 1.01) |
| 706.8 | Other specified diseases of sebaceous glands | Per88 | 1.00 (0.98 – 1.02) | 6.8E-01 | 1.00 (0.98 – 1.02) | 9.1E-01 | 0.98 (0.95 – 1.02) | 1.00 (0.98 – 1.03) |
| 508 | Pulmonary collapse; interstitial and compensatory emphysema | AHI 3% | 1.00 (0.99 – 1.01) | 6.8E-01 | 1.00 (0.99 – 1.01) | 7.6E-01 | 1.00 (0.99 – 1.01) | 1.00 (0.98 – 1.02) |
| 427.3 | Other specified cardiac dysrhythmias | Per88 | 1.00 (0.98 – 1.02) | 6.8E-01 | 0.99 (0.97 – 1.01) | 4.2E-01 | 1.00 (0.98 – 1.02) | 1.01 (0.98 – 1.04) |
| 459.9 | Circulatory disease NEC | AHI 3% | 1.00 (0.99 – 1.01) | 6.9E-01 | 1.00 (0.99 – 1.01) | 4.8E-01 | 1.00 (0.99 – 1.01) | 1.00 (0.99 – 1.01) |
| 296 | Mood disorders | AHI 3% | 1.00 (0.99 – 1.01) | 6.9E-01 | 1.00 (0.99 – 1.01) | 4.1E-01 | 1.00 (0.99 – 1.01) | 1.00 (0.99 – 1.02) |
| 720 | Spinal stenosis | Per88 | 1.00 (0.97 – 1.02) | 6.9E-01 | 1.00 (0.97 – 1.02) | 9.3E-01 | 0.99 (0.96 – 1.02) | 1.00 (0.96 – 1.03) |
| 701.2 | Scar conditions and fibrosis of skin | AHI 3% | 1.00 (0.99 – 1.01) | 7.0E-01 | 1.00 (0.99 – 1.01) | 5.0E-01 | 0.99 (0.98 – 1.00) | 1.01 (1.00 – 1.02) |
| 429.3 | Symptoms involving cardiovascular system | AHI 3% | 1.00 (0.99 – 1.00) | 7.1E-01 | 1.00 (0.99 – 1.00) | 5.2E-01 | 1.00 (0.99 – 1.01) | 1.00 (0.99 – 1.01) |
| 599.9 | Other abnormality of urination | Per88 | 1.00 (0.98 – 1.02) | 7.1E-01 | 0.99 (0.97 – 1.02) | 6.8E-01 | 0.98 (0.96 – 1.01) | 1.01 (0.99 – 1.04) |
| 008 | Intestinal infection | AHI 3% | 1.00 (0.99 – 1.01) | 7.1E-01 | 1.00 (0.99 – 1.01) | 7.9E-01 | 1.00 (0.99 – 1.01) | 1.00 (0.99 – 1.01) |
| 389.4 | Tinnitus | Per88 | 1.00 (0.98 – 1.01) | 7.2E-01 | 1.00 (0.98 – 1.01) | 5.8E-01 | 1.00 (0.97 – 1.02) | 0.99 (0.97 – 1.02) |
| 704 | Diseases of hair and hair follicles | Per88 | 1.00 (0.98 – 1.02) | 7.5E-01 | 1.00 (0.98 – 1.02) | 9.4E-02 | 1.00 (0.97 – 1.03) | 0.99 (0.97 – 1.02) |
| 261.4 | Vitamin D deficiency | AHI 3% | 1.00 (0.99 – 1.01) | 7.5E-01 | 1.00 (0.99 – 1.01) | 5.5E-01 | 1.00 (0.98 – 1.01) | 1.00 (0.99 – 1.01) |
| 271.3 | Intestinal disaccharidase deficiencies and disaccharide malabsorption | AHI 3% | 1.00 (1.00 – 1.00) | 7.5E-01 | 1.00 (1.00 – 1.00) | 5.1E-01 | 1.00 (0.99 – 1.01) | 1.00 (0.99 – 1.00) |
| 840 | Sprains and strains | Per88 | 1.00 (0.99 – 1.02) | 7.6E-01 | 1.00 (0.98 – 1.02) | 8.3E-01 | 1.00 (0.98 – 1.03) | 1.00 (0.98 – 1.03) |
| 375.1 | Dry eyes | Per88 | 1.00 (0.99 – 1.02) | 7.6E-01 | 1.00 (0.98 – 1.02) | 2.7E-01 | 1.00 (0.97 – 1.02) | 1.00 (0.98 – 1.03) |
| 704.1 | Alopecia | Per88 | 1.00 (0.98 – 1.02) | 7.6E-01 | 1.00 (0.98 – 1.02) | 7.2E-01 | 1.01 (0.98 – 1.05) | 0.99 (0.96 – 1.02) |
| 296 | Mood disorders | Per88 | 1.00 (0.98 – 1.02) | 7.7E-01 | 1.00 (0.98 – 1.03) | 2.7E-01 | 1.01 (0.98 – 1.04) | 1.00 (0.97 – 1.02) |
| 272.1 | Hyperlipidemia | Per88 | 1.00 (0.99 – 1.02) | 7.7E-01 | 1.00 (0.98 – 1.02) | 4.6E-01 | 1.00 (0.98 – 1.02) | 1.01 (0.98 – 1.03) |
| 389.1 | Sensorineural hearing loss | Per88 | 1.00 (0.98 – 1.03) | 7.8E-01 | 1.00 (0.98 – 1.03) | 5.2E-02 | 1.03 (1.00 – 1.05) | 0.97 (0.93 – 1.02) |
| 278.4 | Abnormal weight gain | Per88 | 1.00 (0.98 – 1.02) | 7.8E-01 | 1.00 (0.98 – 1.02) | 8.5E-01 | 0.98 (0.91 – 1.04) | 1.00 (0.98 – 1.02) |
| 599.4 | Urinary incontinence | Per88 | 1.00 (0.97 – 1.02) | 7.8E-01 | 1.00 (0.98 – 1.02) | 7.0E-01 | 0.99 (0.95 – 1.03) | 1.00 (0.97 – 1.03) |
| 765 | Cervical radiculitis | AHI 3% | 1.00 (0.99 – 1.01) | 7.8E-01 | 1.00 (0.99 – 1.01) | 5.5E-01 | 1.00 (0.98 – 1.01) | 1.01 (1.00 – 1.02) |
| 277 | Other disorders of metabolism | AHI 3% | 1.00 (1.00 – 1.01) | 7.9E-01 | 1.00 (1.00 – 1.01) | 2.5E-01 | 1.00 (0.99 – 1.01) | 1.00 (0.99 – 1.01) |

| PheCode | Disease | Sleep statistic | Risk ratio | P-value | Risk ratio (adj pulmonary disorders) | Sleep statistic x sex interaction p-value | Risk ratio (men) | Risk ratio (women) |
|---|---|---|---|---|---|---|---|---|
| 261.4 | Vitamin D deficiency | Per88 | 1.00 (0.98 – 1.02) | 7.9E-01 | 1.00 (0.98 – 1.02) | 1.6E-01 | 0.98 (0.95 – 1.02) | 1.01 (0.98 – 1.03) |
| 701 | Other hypertrophic and atrophic conditions of skin | AHI 3% | 1.00 (0.99 – 1.01) | 7.9E-01 | 1.00 (0.99 – 1.01) | 5.9E-01 | 1.00 (0.99 – 1.01) | 1.00 (0.99 – 1.01) |
| 276.5 | Hypovolemia | AHI 3% | 1.00 (0.99 – 1.01) | 8.0E-01 | 1.00 (0.99 – 1.01) | 8.3E-01 | 1.00 (0.99 – 1.01) | 1.00 (0.99 – 1.01) |
| 262 | Mineral deficiency NEC | Per88 | 1.00 (0.98 – 1.02) | 8.0E-01 | 1.00 (0.98 – 1.02) | 6.1E-01 | 1.00 (0.96 – 1.04) | 1.00 (0.98 – 1.03) |
| 965 | Poisoning by analgesics, antipyretics, and antirheumatics | Per88 | 1.00 (0.98 – 1.02) | 8.0E-01 | 1.00 (0.98 – 1.02) | 1.8E-01 | 0.97 (0.93 – 1.01) | 1.01 (0.99 – 1.03) |
| 571.5 | Other chronic nonalcoholic liver disease | AHI 3% | 1.00 (0.99 – 1.01) | 8.1E-01 | 1.00 (0.99 – 1.01) | 5.2E-01 | 1.00 (0.99 – 1.02) | 0.99 (0.98 – 1.01) |
| 427.6 | Premature beats | AHI 3% | 1.00 (0.99 – 1.01) | 8.1E-01 | 1.00 (0.99 – 1.01) | 9.6E-01 | 1.00 (0.99 – 1.01) | 1.01 (0.99 – 1.02) |
| 278.1 | Obesity | AHI 3% | 1.00 (0.99 – 1.01) | 8.2E-01 | 1.00 (0.99 – 1.01) | 7.9E-01 | 1.00 (0.99 – 1.01) | 1.00 (0.98 – 1.01) |
| 481 | Influenza | AHI 3% | 1.00 (0.99 – 1.01) | 8.4E-01 | 1.00 (0.99 – 1.01) | 3.2E-01 | 1.00 (0.99 – 1.02) | 1.00 (0.99 – 1.01) |
| 278 | Overweight, obesity and other hyperalimentation | AHI 3% | 1.00 (0.99 – 1.01) | 8.4E-01 | 1.00 (0.99 – 1.01) | 4.1E-02 | 1.00 (1.00 – 1.01) | 0.99 (0.99 – 1.00) |
| 136 | Other infectious and parasitic diseases | AHI 3% | 1.00 (0.99 – 1.01) | 8.4E-01 | 1.00 (0.99 – 1.01) | 2.9E-01 | 1.00 (0.99 – 1.01) | 1.00 (0.99 – 1.01) |
| 300.11 | Generalized anxiety disorder | Per88 | 1.00 (0.98 – 1.01) | 8.5E-01 | 1.00 (0.98 – 1.01) | 7.6E-02 | 0.97 (0.94 – 1.01) | 1.01 (0.99 – 1.02) |
| 481 | Influenza | Per88 | 1.00 (0.98 – 1.03) | 8.5E-01 | 1.00 (0.98 – 1.02) | 5.4E-01 | 0.99 (0.96 – 1.03) | 1.01 (0.99 – 1.04) |
| 401.1 | Essential hypertension | Per88 | 1.00 (0.98 – 1.03) | 8.6E-01 | 1.00 (0.98 – 1.03) | 6.1E-01 | 0.99 (0.95 – 1.03) | 1.01 (0.98 – 1.05) |
| 798 | Malaise and fatigue | AHI 3% | 1.00 (0.99 – 1.01) | 8.6E-01 | 1.00 (0.99 – 1.01) | 5.5E-01 | 1.00 (0.99 – 1.01) | 1.00 (0.99 – 1.01) |
| 530.11 | GERD | Per88 | 1.00 (0.98 – 1.01) | 8.6E-01 | 1.00 (0.98 – 1.01) | 3.4E-01 | 0.99 (0.96 – 1.01) | 1.01 (0.99 – 1.03) |
| 743.11 | Osteoporosis NOS | AHI 3% | 1.00 (0.99 – 1.01) | 8.7E-01 | 1.00 (0.99 – 1.01) | 3.2E-01 | 0.98 (0.97 – 1.00) | 1.00 (0.99 – 1.01) |
| 459.9 | Circulatory disease NEC | Per88 | 1.00 (0.98 – 1.03) | 9.0E-01 | 1.00 (0.98 – 1.03) | 1.7E-01 | 1.02 (0.99 – 1.05) | 0.98 (0.94 – 1.01) |
| 704.1 | Alopecia | AHI 3% | 1.00 (0.99 – 1.01) | 9.0E-01 | 1.00 (0.99 – 1.01) | 2.1E-01 | 1.00 (0.99 – 1.02) | 1.00 (0.99 – 1.01) |
| 208 | Benign neoplasm of colon | Per88 | 1.00 (0.97 – 1.02) | 9.2E-01 | 1.00 (0.97 – 1.03) | 1.3E-01 | 1.02 (0.99 – 1.05) | 0.97 (0.93 – 1.01) |
| 472 | Chronic pharyngitis and nasopharyngitis | AHI 3% | 1.00 (0.99 – 1.01) | 9.2E-01 | 1.00 (0.99 – 1.01) | 3.4E-01 | 1.00 (0.99 – 1.01) | 1.00 (0.99 – 1.01) |
| 727.1 | Synovitis and tenosynovitis | Per88 | 1.00 (0.98 – 1.03) | 9.2E-01 | 1.00 (0.98 – 1.03) | 5.7E-01 | 1.00 (0.96 – 1.04) | 1.00 (0.97 – 1.03) |
| 593.2 | Microscopic hematuria | AHI 3% | 1.00 (0.99 – 1.01) | 9.2E-01 | 1.00 (0.99 – 1.01) | 1.7E-01 | 1.00 (0.99 – 1.02) | 0.99 (0.98 – 1.01) |
| 386.9 | Dizziness and giddiness (Light-headedness and vertigo) | Per88 | 1.00 (0.98 – 1.02) | 9.2E-01 | 1.00 (0.98 – 1.02) | 7.3E-01 | 0.99 (0.96 – 1.02) | 1.01 (0.99 – 1.03) |
| 565.1 | Anal and rectal polyp | Per88 | 1.00 (0.98 – 1.03) | 9.2E-01 | 1.00 (0.98 – 1.03) | 3.8E-02 | 1.01 (0.98 – 1.05) | 0.99 (0.96 – 1.02) |
| 765 | Cervical radiculitis | Per88 | 1.00 (0.98 – 1.02) | 9.3E-01 | 1.00 (0.98 – 1.02) | 1.3E-01 | 0.97 (0.94 – 0.99) | 1.02 (1.00 – 1.05) |
| 402 | Elevated blood pressure reading without diagnosis of hypertension | AHI 3% | 1.00 (0.99 – 1.01) | 9.3E-01 | 1.00 (0.99 – 1.01) | 9.1E-01 | 1.00 (0.99 – 1.01) | 1.00 (0.99 – 1.01) |
| 369.5 | Conjunctivitis, infectious | AHI 3% | 1.00 (0.99 – 1.01) | 9.5E-01 | 1.00 (0.99 – 1.01) | 1.2E-02 | 1.01 (1.00 – 1.02) | 0.99 (0.98 – 1.00) |
| 292 | Neurological disorders | AHI 3% | 1.00 (0.99 – 1.01) | 9.5E-01 | 1.00 (0.99 – 1.01) | 2.3E-01 | 1.00 (0.99 – 1.01) | 0.99 (0.98 – 1.01) |
| 379.9 | Pain, swelling or discharge of eye | AHI 3% | 1.00 (0.99 – 1.01) | 9.7E-01 | 1.00 (0.99 – 1.01) | 5.8E-01 | 1.01 (0.99 – 1.02) | 0.99 (0.98 – 1.00) |
| 429.3 | Symptoms involving cardiovascular system | Per88 | 1.00 (0.98 – 1.02) | 9.8E-01 | 1.00 (0.98 – 1.02) | 6.9E-03 | 0.99 (0.96 – 1.01) | 1.01 (0.99 – 1.04) |
| 285 | Other anemias | AHI 3% | 1.00 (0.99 – 1.01) | 9.9E-01 | 1.00 (0.99 – 1.01) | 6.2E-01 | 1.00 (0.99 – 1.01) | 1.00 (0.99 – 1.01) |

| PheCode | Disease | Sleep statistic | Risk ratio | P-value | Risk ratio (adj pulmonary disorders) | Sleep statistic x sex interaction p-value | Risk ratio (men) | Risk ratio (women) |
|---|---|---|---|---|---|---|---|---|
| 389.1 | Sensorineural hearing loss | AHI 3% | 1.00 (0.99 – 1.01) | 9.9E-01 | 1.00 (0.99 – 1.01) | 9.9E-02 | 1.00 (0.99 – 1.01) | 0.99 (0.98 – 1.01) |
| 727.1 | Synovitis and tenosynovitis | AHI 3% | 1.00 (0.99 – 1.01) | 9.9E-01 | 1.00 (0.99 – 1.01) | 4.8E-01 | 1.01 (1.00 – 1.02) | 0.99 (0.98 – 1.01) |
| 599.8 | Other symptoms involving urinary system | Per88 | 1.00 (0.98 – 1.02) | 9.9E-01 | 1.00 (0.98 – 1.02) | 5.6E-01 | 1.01 (0.98 – 1.04) | 1.00 (0.97 – 1.03) |

#### Table S11. Cross-sectional sleep statistic associations with comorbidities.

Cross-sectional analyses used the first sleep recording (prioritizing diagnostics, then splits, then home sleep tests) and are ordered by p-values. Primary analyses considered joint-sex PheCodes with >1% prevalence rate and were rank-normalized after adjusting for age, sex, BMI, population, sleep study type, and healthcare encounters. The primary analysis considered all available sleep laboratory data (i.e. including participants being screened for other sleep disorders).

47 non-redundant PheCodes were Bonferroni-level significant (**bolded**) and 117 non-redundant PheCodes were FDR-level significant (in red, uncorrected p-values).

Due to PDF width constraints, sensitivity analyses are presented in two tables, with primary columns repeated (apart from incidence and n):

**Table S11 further includes:**
A sensitivity analysis considering only participants with AHI >15.
A sensitivity analysis further adjusting for an indicator variable for other common sleep disorders (PheCodes 327, 327.1, 327.4, 327.41, 327.5, 327.6, 327.7, 327.71) in the full sleep recording sample.

**Table S12 further includes:**
A sensitivity analysis further adjusting for an indicator variable for common pulmonary disorders (PheCodes 415.1, 415.11, 415.2, 480.1, 480.11, 480.2, 480, 495, 495.2, 496.1, 496.2, 496.21, 496.3, 497, 502, 503, 505, 506, 509.2).
P-values from sleep statistic × sex interaction analyses (not rank-normalized) performed separately. Two Bonferroni-level significant values are displayed in green.
Equivalent sex-stratified results.

| PheCode | Disease | Sleep statistic | Prevalence | Risk ratio | P-value | N | Risk ratio (AHI >15) | N (AHI >15) | Risk ratio (adj sleep disorders) |
|---|---|---|---|---|---|---|---|---|---|
| 1013 | Asphyxia and hypoxemia | Per88 | 3.8 | 1.07 (1.06 – 1.07) | **1.1E-70** | 12232 | 1.05 (1.04 – 1.06) | 5845 | 1.07 (1.06 – 1.07) |
| 496 | Chronic airway obstruction | Per88 | 3.9 | 1.06 (1.05 – 1.06) | **3.2E-45** | 12232 | 1.04 (1.04 – 1.05) | 5845 | 1.06 (1.05 – 1.06) |
| 509.1 | Respiratory failure | Per88 | 2.3 | 1.07 (1.06 – 1.08) | **7.0E-40** | 12232 | 1.05 (1.04 – 1.06) | 5845 | 1.07 (1.06 – 1.08) |
| 415.2 | Chronic pulmonary heart disease | Per88 | 2.1 | 1.07 (1.05 – 1.08) | **5.5E-38** | 12232 | 1.05 (1.04 – 1.06) | 5845 | 1.07 (1.05 – 1.08) |
| 496.21 | Obstructive chronic bronchitis | Per88 | 1.3 | 1.07 (1.06 – 1.09) | **2.4E-34** | 12232 | 1.06 (1.05 – 1.07) | 5845 | 1.07 (1.06 – 1.09) |
| 508 | Pulmonary collapse; interstitial and compensatory emphysema | Per88 | 5.3 | 1.04 (1.04 – 1.05) | **3.9E-34** | 12232 | 1.03 (1.02 – 1.04) | 5845 | 1.04 (1.04 – 1.05) |
| 503 | Pulmonary congestion and hypostasis | Per88 | 2.9 | 1.06 (1.05 – 1.07) | **6.8E-31** | 12232 | 1.04 (1.03 – 1.05) | 5845 | 1.06 (1.05 – 1.07) |
| 496.1 | Emphysema | Per88 | 2.0 | 1.06 (1.05 – 1.07) | **1.1E-30** | 12232 | 1.04 (1.03 – 1.06) | 5845 | 1.06 (1.05 – 1.07) |
| 512.7 | Shortness of breath | Per88 | 17.1 | 1.02 (1.02 – 1.03) | **8.6E-26** | 12232 | 1.02 (1.01 – 1.02) | 5845 | 1.02 (1.02 – 1.03) |
| 782.3 | Edema | Per88 | 9.5 | 1.03 (1.03 – 1.04) | **6.7E-25** | 12232 | 1.02 (1.02 – 1.03) | 5845 | 1.03 (1.03 – 1.04) |
| 428.1 | Congestive heart failure (CHF) NOS | Per88 | 5.6 | 1.04 (1.03 – 1.05) | **1.3E-21** | 12232 | 1.03 (1.02 – 1.04) | 5845 | 1.04 (1.03 – 1.05) |

| PheCode | Disease | Sleep statistic | Prevalence | Risk ratio | P-value | N | Risk ratio (AHI >15) | N (AHI >15) | Risk ratio (adj sleep disorders) |
|---|---|---|---|---|---|---|---|---|---|
| 401.1 | Essential hypertension | AHI 3% | 41.8 | 1.00 (1.00 – 1.01) | **7.0E-20** | 18332 | 1.00 (1.00 – 1.00) | 9404 | 1.00 (1.00 – 1.01) |
| 512.9 | Other dyspnea | Per88 | 18.4 | 1.02 (1.01 – 1.02) | **5.8E-18** | 12232 | 1.01 (1.01 – 1.02) | 5845 | 1.02 (1.01 – 1.02) |
| 428.4 | Heart failure with preserved EF [Diastolic heart failure] | Per88 | 2.4 | 1.05 (1.04 – 1.06) | **2.0E-17** | 12232 | 1.04 (1.02 – 1.05) | 5845 | 1.05 (1.04 – 1.06) |
| 480 | Pneumonia | Per88 | 7.9 | 1.03 (1.02 – 1.03) | **5.6E-17** | 12232 | 1.02 (1.01 – 1.03) | 5845 | 1.03 (1.02 – 1.03) |
| 505 | Other pulmonary inflamation or edema | Per88 | 1.3 | 1.06 (1.04 – 1.07) | **2.0E-16** | 12232 | 1.03 (1.02 – 1.05) | 5845 | 1.06 (1.04 – 1.07) |
| 510 | Other diseases of lung | Per88 | 3.7 | 1.04 (1.03 – 1.04) | **2.0E-15** | 12232 | 1.03 (1.02 – 1.04) | 5845 | 1.04 (1.03 – 1.04) |
| 428.2 | Heart failure NOS | Per88 | 2.3 | 1.05 (1.03 – 1.06) | **1.2E-14** | 12232 | 1.03 (1.02 – 1.04) | 5845 | 1.05 (1.03 – 1.06) |
| 276.6 | Fluid overload | Per88 | 1.9 | 1.05 (1.04 – 1.06) | **1.3E-14** | 12232 | 1.03 (1.02 – 1.04) | 5845 | 1.05 (1.04 – 1.06) |
| 340 | Migraine | AHI 3% | 13.3 | 0.99 (0.99 – 0.99) | **5.6E-14** | 18332 | 0.99 (0.99 – 1.00) | 9404 | 0.99 (0.99 – 1.00) |
| 401.21 | Hypertensive heart disease | Per88 | 2.5 | 1.04 (1.03 – 1.06) | **6.2E-13** | 12232 | 1.03 (1.02 – 1.04) | 5845 | 1.04 (1.03 – 1.06) |
| 805 | Fracture of vertebral column without mention of spinal cord injury | Per88 | 1.5 | 1.05 (1.03 – 1.06) | **1.2E-12** | 12232 | 1.03 (1.02 – 1.05) | 5845 | 1.05 (1.03 – 1.06) |
| 458.9 | Hypotension NOS | Per88 | 3.6 | 1.03 (1.02 – 1.04) | **1.5E-12** | 12232 | 1.02 (1.01 – 1.03) | 5845 | 1.03 (1.02 – 1.04) |
| 401.1 | Essential hypertension | Per88 | 41.8 | 1.01 (1.01 – 1.01) | **2.0E-10** | 12232 | 1.00 (1.00 – 1.00) | 5845 | 1.01 (1.01 – 1.01) |
| 585.1 | Acute renal failure | Per88 | 4.8 | 1.03 (1.02 – 1.04) | **3.6E-10** | 12232 | 1.02 (1.01 – 1.03) | 5845 | 1.03 (1.02 – 1.04) |
| 427.7 | Tachycardia NOS | Per88 | 7.4 | 1.02 (1.01 – 1.03) | **5.6E-10** | 12232 | 1.02 (1.01 – 1.02) | 5845 | 1.02 (1.01 – 1.03) |
| 327.4 | Insomnia | AHI 3% | 16.8 | 0.99 (0.99 – 1.00) | **8.3E-10** | 18332 | 1.00 (1.00 – 1.00) | 9404 | 1.00 (1.00 – 1.00) |
| 316 | Substance addiction and disorders | Per88 | 5.5 | 1.02 (1.01 – 1.03) | **4.4E-09** | 12232 | 1.02 (1.01 – 1.02) | 5845 | 1.02 (1.01 – 1.03) |
| 260 | Protein-calorie malnutrition | Per88 | 1.7 | 1.03 (1.02 – 1.04) | **1.6E-08** | 12232 | 1.02 (1.01 – 1.04) | 5845 | 1.03 (1.02 – 1.04) |
| 250.2 | Type 2 diabetes | AHI 3% | 15.4 | 1.01 (1.00 – 1.01) | **1.8E-08** | 18332 | 1.00 (1.00 – 1.01) | 9404 | 1.01 (1.00 – 1.01) |
| 339 | Other headache syndromes | AHI 3% | 13.0 | 0.99 (0.99 – 1.00) | **2.2E-08** | 18332 | 0.99 (0.99 – 1.00) | 9404 | 0.99 (0.99 – 1.00) |
| 317.1 | Alcoholism | Per88 | 2.9 | 1.03 (1.02 – 1.04) | **6.1E-08** | 12232 | 1.02 (1.01 – 1.03) | 5845 | 1.03 (1.02 – 1.04) |
| 250.2 | Type 2 diabetes | Per88 | 15.4 | 1.01 (1.01 – 1.02) | **1.0E-07** | 12232 | 1.01 (1.00 – 1.01) | 5845 | 1.01 (1.01 – 1.02) |
| 401.3 | Other hypertensive complications | AHI 3% | 2.3 | 1.02 (1.01 – 1.02) | **1.3E-07** | 18332 | 1.02 (1.01 – 1.02) | 9404 | 1.02 (1.01 – 1.02) |
| 340.1 | Migraine with aura | AHI 3% | 3.5 | 0.99 (0.99 – 0.99) | **2.0E-07** | 18332 | 0.99 (0.98 – 1.00) | 9404 | 0.99 (0.99 – 0.99) |
| 504 | Other alveolar and parietoalveolar pneumonopathy | Per88 | 1.3 | 1.04 (1.02 – 1.05) | **2.6E-07** | 12232 | 1.02 (1.01 – 1.04) | 5845 | 1.04 (1.02 – 1.05) |
| 261 | Vitamin deficiency | AHI 3% | 3.5 | 0.99 (0.98 – 0.99) | **3.7E-07** | 18332 | 0.99 (0.99 – 1.00) | 9404 | 0.99 (0.98 – 0.99) |
| 505 | Other pulmonary inflamation or edema | AHI 3% | 1.3 | 1.02 (1.01 – 1.03) | **6.5E-07** | 18332 | 1.02 (1.01 – 1.02) | 9404 | 1.02 (1.01 – 1.03) |
| 797 | Shock | Per88 | 1.2 | 1.04 (1.03 – 1.06) | **1.0E-06** | 12232 | 1.03 (1.02 – 1.05) | 5845 | 1.04 (1.03 – 1.06) |
| 269 | Proteinuria | AHI 3% | 3.0 | 1.01 (1.01 – 1.02) | **1.3E-06** | 18332 | 1.01 (1.01 – 1.02) | 9404 | 1.01 (1.01 – 1.02) |
| 585.1 | Acute renal failure | AHI 3% | 4.8 | 1.01 (1.01 – 1.01) | **2.8E-06** | 18332 | 1.01 (1.00 – 1.01) | 9404 | 1.01 (1.01 – 1.01) |
| 389.4 | Tinnitus | Per88 | 6.2 | 0.98 (0.97 – 0.99) | **3.3E-06** | 12232 | 0.98 (0.97 – 0.99) | 5845 | 0.98 (0.97 – 0.99) |
| 539 | Bariatric surgery | AHI 3% | 12.4 | 0.99 (0.99 – 1.00) | **6.7E-06** | 18332 | 1.00 (0.99 – 1.00) | 9404 | 0.99 (0.99 – 1.00) |
| 563 | Constipation | AHI 3% | 10.8 | 0.99 (0.99 – 1.00) | **1.0E-05** | 18332 | 1.00 (1.00 – 1.00) | 9404 | 1.00 (0.99 – 1.00) |
| 300.9 | Posttraumatic stress disorder | AHI 3% | 4.1 | 0.99 (0.99 – 1.00) | **1.1E-05** | 18332 | 0.99 (0.99 – 1.00) | 9404 | 0.99 (0.99 – 1.00) |
| 288.2 | Elevated white blood cell count | Per88 | 4.2 | 1.02 (1.01 – 1.03) | **1.2E-05** | 12232 | 1.02 (1.01 – 1.03) | 5845 | 1.02 (1.01 – 1.03) |

| PheCode | Disease | Sleep statistic | Prevalence | Risk ratio | P-value | N | Risk ratio (AHI >15) | N (AHI >15) | Risk ratio (adj sleep disorders) |
|---|---|---|---|---|---|---|---|---|---|
| 425.1 | Primary/intrinsic cardiomyopathies | Per88 | 3.2 | 1.02 (1.01 – 1.04) | **1.7E-05** | 12232 | 1.02 (1.01 – 1.03) | 5845 | 1.03 (1.01 – 1.04) |
| 503 | Pulmonary congestion and hypostasis | AHI 3% | 2.9 | 1.01 (1.01 – 1.02) | **1.8E-05** | 18332 | 1.01 (1.00 – 1.01) | 9404 | 1.01 (1.01 – 1.02) |
| 340 | Migraine | Per88 | 13.3 | 0.99 (0.99 – 0.99) | **2.0E-05** | 12232 | 0.99 (0.98 – 1.00) | 5845 | 0.99 (0.99 – 0.99) |
| 512.8 | Cough | Per88 | 16.5 | 1.01 (1.01 – 1.01) | **2.5E-05** | 12232 | 1.01 (1.00 – 1.01) | 5845 | 1.01 (1.01 – 1.01) |
| 509.1 | Respiratory failure | AHI 3% | 2.3 | 1.01 (1.01 – 1.02) | **2.6E-05** | 18332 | 1.01 (1.01 – 1.02) | 9404 | 1.01 (1.01 – 1.02) |
| 428.2 | Heart failure NOS | AHI 3% | 2.3 | 1.01 (1.01 – 1.02) | **2.8E-05** | 18332 | 1.01 (1.00 – 1.01) | 9404 | 1.01 (1.01 – 1.02) |
| 041 | Bacterial infection NOS | Per88 | 2.1 | 1.02 (1.01 – 1.04) | **3.2E-05** | 12232 | 1.01 (1.00 – 1.03) | 5845 | 1.02 (1.01 – 1.04) |
| 694.2 | Other dyschromia | Per88 | 12.9 | 0.99 (0.98 – 0.99) | **3.3E-05** | 12232 | 0.99 (0.99 – 1.00) | 5845 | 0.99 (0.98 – 0.99) |
| 425.1 | Primary/intrinsic cardiomyopathies | AHI 3% | 3.2 | 1.01 (1.01 – 1.01) | **4.0E-05** | 18332 | 1.01 (1.00 – 1.01) | 9404 | 1.01 (1.00 – 1.01) |
| 136 | Other infectious and parasitic diseases | Per88 | 4.6 | 1.02 (1.01 – 1.02) | **5.4E-05** | 12232 | 1.01 (1.00 – 1.02) | 5845 | 1.02 (1.01 – 1.02) |
| 290.2 | Delirium due to conditions classified elsewhere | Per88 | 1.2 | 1.04 (1.02 – 1.05) | 7.5E-05 | 12232 | 1.03 (1.02 – 1.05) | 5845 | 1.04 (1.02 – 1.05) |
| 327.41 | Organic or persistent insomnia | AHI 3% | 3.0 | 0.99 (0.99 – 1.00) | 7.6E-05 | 18332 | 0.99 (0.99 – 1.00) | 9404 | 1.00 (0.99 – 1.00) |
| 276.14 | Hypopotassemia | Per88 | 2.8 | 1.02 (1.01 – 1.03) | 7.9E-05 | 12232 | 1.01 (1.00 – 1.02) | 5845 | 1.02 (1.01 – 1.03) |
| 414 | Other forms of chronic heart disease | Per88 | 2.3 | 1.03 (1.01 – 1.04) | 8.4E-05 | 12232 | 1.01 (1.00 – 1.03) | 5845 | 1.03 (1.01 – 1.04) |
| 745 | Pain in joint | AHI 3% | 33.1 | 1.00 (1.00 – 1.00) | 8.8E-05 | 18332 | 1.00 (1.00 – 1.00) | 9404 | 1.00 (1.00 – 1.00) |
| 286.9 | Abnormal coagulation profile | Per88 | 2.8 | 1.02 (1.01 – 1.03) | 1.3E-04 | 12232 | 1.01 (1.00 – 1.03) | 5845 | 1.02 (1.01 – 1.03) |
| 416 | Cardiomegaly | Per88 | 2.6 | 1.03 (1.01 – 1.04) | 1.5E-04 | 12232 | 1.02 (1.00 – 1.03) | 5845 | 1.03 (1.01 – 1.04) |
| 288.2 | Elevated white blood cell count | AHI 3% | 4.2 | 1.01 (1.00 – 1.01) | 1.7E-04 | 18332 | 1.01 (1.00 – 1.01) | 9404 | 1.01 (1.00 – 1.01) |
| 261 | Vitamin deficiency | Per88 | 3.5 | 0.97 (0.96 – 0.99) | 2.0E-04 | 12232 | 0.99 (0.97 – 1.00) | 5845 | 0.97 (0.96 – 0.99) |
| 350.1 | Abnormal involuntary movements | AHI 3% | 5.4 | 0.99 (0.99 – 1.00) | 2.6E-04 | 18332 | 1.00 (0.99 – 1.00) | 9404 | 0.99 (0.99 – 1.00) |
| 428.3 | Heart failure with reduced EF [Systolic or combined heart failure] | Per88 | 1.9 | 1.03 (1.01 – 1.05) | 2.7E-04 | 12232 | 1.02 (1.00 – 1.03) | 5845 | 1.03 (1.01 – 1.05) |
| 496.2 | Chronic bronchitis | Per88 | 1.1 | 1.03 (1.02 – 1.05) | 2.9E-04 | 12232 | 1.02 (1.00 – 1.04) | 5845 | 1.03 (1.02 – 1.05) |
| 274.11 | Gouty arthropathy | AHI 3% | 2.3 | 1.01 (1.00 – 1.02) | 3.2E-04 | 18332 | 1.01 (1.00 – 1.01) | 9404 | 1.01 (1.01 – 1.02) |
| 598.9 | Other nonspecific findings on examination of urine | AHI 3% | 3.0 | 0.99 (0.99 – 1.00) | 3.2E-04 | 18332 | 1.00 (0.99 – 1.00) | 9404 | 0.99 (0.99 – 1.00) |
| 278 | Overweight, obesity and other hyperalimentation | Per88 | 14.8 | 0.99 (0.99 – 1.00) | 3.4E-04 | 12232 | 0.99 (0.99 – 1.00) | 5845 | 0.99 (0.99 – 1.00) |
| 427.9 | Palpitations | AHI 3% | 12.6 | 1.00 (0.99 – 1.00) | 3.8E-04 | 18332 | 1.00 (0.99 – 1.00) | 9404 | 1.00 (0.99 – 1.00) |
| 250.7 | Diabetic retinopathy | AHI 3% | 1.4 | 1.01 (1.01 – 1.02) | 3.8E-04 | 18332 | 1.01 (1.00 – 1.01) | 9404 | 1.01 (1.01 – 1.02) |
| 760 | Back pain | AHI 3% | 23.6 | 1.00 (1.00 – 1.00) | 4.1E-04 | 18332 | 1.00 (0.99 – 1.00) | 9404 | 1.00 (1.00 – 1.00) |
| 726 | Peripheral enthesopathies and allied syndromes | AHI 3% | 8.1 | 1.00 (0.99 – 1.00) | 4.3E-04 | 18332 | 1.00 (0.99 – 1.00) | 9404 | 1.00 (0.99 – 1.00) |
| 411.3 | Angina pectoris | Per88 | 7.0 | 1.01 (1.01 – 1.02) | 4.4E-04 | 12232 | 1.01 (1.00 – 1.01) | 5845 | 1.01 (1.01 – 1.02) |
| 250.7 | Diabetic retinopathy | Per88 | 1.4 | 1.03 (1.01 – 1.05) | 5.0E-04 | 12232 | 1.02 (1.00 – 1.04) | 5845 | 1.03 (1.01 – 1.05) |
| 743.11 | Osteoporosis NOS | Per88 | 7.2 | 1.01 (1.01 – 1.02) | 5.4E-04 | 12232 | 1.01 (1.01 – 1.02) | 5845 | 1.01 (1.01 – 1.02) |
| 514.2 | Solitary pulmonary nodule | Per88 | 4.2 | 1.02 (1.01 – 1.03) | 5.5E-04 | 12232 | 1.01 (1.00 – 1.02) | 5845 | 1.02 (1.01 – 1.03) |
| 415 | Pulmonary heart disease | Per88 | 1.3 | 1.03 (1.01 – 1.04) | 5.6E-04 | 12232 | 1.03 (1.01 – 1.04) | 5845 | 1.03 (1.01 – 1.04) |

| PheCode | Disease | Sleep statistic | Prevalence | Risk ratio | P-value | N | Risk ratio (AHI >15) | N (AHI >15) | Risk ratio (adj sleep disorders) |
|---|---|---|---|---|---|---|---|---|---|
| 285 | Other anemias | Per88 | 14.6 | 1.01 (1.00 – 1.01) | 5.7E-04 | 12232 | 1.01 (1.00 – 1.01) | 5845 | 1.01 (1.00 – 1.01) |
| 278.4 | Abnormal weight gain | AHI 3% | 6.5 | 0.99 (0.99 – 1.00) | 6.2E-04 | 18332 | 1.00 (0.99 – 1.00) | 9404 | 0.99 (0.99 – 1.00) |
| 797 | Shock | AHI 3% | 1.2 | 1.02 (1.01 – 1.02) | 6.4E-04 | 18332 | 1.01 (1.01 – 1.02) | 9404 | 1.02 (1.01 – 1.02) |
| 327 | Sleep disorders | AHI 3% | 8.0 | 1.00 (0.99 – 1.00) | 6.7E-04 | 18332 | 1.00 (1.00 – 1.00) | 9404 | 1.00 (1.00 – 1.00) |
| 395.1 | Nonrheumatic mitral valve disorders | Per88 | 4.0 | 1.02 (1.01 – 1.03) | 7.1E-04 | 12232 | 1.01 (1.01 – 1.02) | 5845 | 1.02 (1.01 – 1.03) |
| 250.42 | Other abnormal glucose | Per88 | 11.1 | 1.01 (1.00 – 1.02) | 7.3E-04 | 12232 | 1.01 (1.00 – 1.02) | 5845 | 1.01 (1.00 – 1.02) |
| 530.9 | Heartburn | Per88 | 5.7 | 0.99 (0.98 – 0.99) | 7.4E-04 | 12232 | 0.99 (0.98 – 1.00) | 5845 | 0.99 (0.98 – 0.99) |
| 840 | Sprains and strains | AHI 3% | 9.8 | 1.00 (0.99 – 1.00) | 7.4E-04 | 18332 | 0.99 (0.99 – 1.00) | 9404 | 1.00 (0.99 – 1.00) |
| 512.8 | Cough | AHI 3% | 16.5 | 1.00 (1.00 – 1.00) | 7.7E-04 | 18332 | 1.00 (1.00 – 1.00) | 9404 | 1.00 (1.00 – 1.00) |
| 785 | Abdominal pain | AHI 3% | 20.0 | 1.00 (1.00 – 1.00) | 7.8E-04 | 18332 | 1.00 (0.99 – 1.00) | 9404 | 1.00 (1.00 – 1.00) |
| 427.3 | Other specified cardiac dysrhythmias | AHI 3% | 7.4 | 0.99 (0.99 – 1.00) | 8.3E-04 | 18332 | 1.00 (0.99 – 1.00) | 9404 | 0.99 (0.99 – 1.00) |
| 586 | Other disorders of the kidney and ureters | AHI 3% | 5.2 | 1.01 (1.00 – 1.01) | 8.5E-04 | 18332 | 1.01 (1.00 – 1.01) | 9404 | 1.01 (1.00 – 1.01) |
| 260 | Protein-calorie malnutrition | AHI 3% | 1.7 | 1.01 (1.00 – 1.02) | 9.5E-04 | 18332 | 1.01 (1.00 – 1.01) | 9404 | 1.01 (1.00 – 1.02) |
| 508 | Pulmonary collapse; interstitial and compensatory emphysema | AHI 3% | 5.3 | 1.01 (1.00 – 1.01) | 1.0E-03 | 18332 | 1.01 (1.00 – 1.01) | 9404 | 1.01 (1.00 – 1.01) |
| 428.1 | Congestive heart failure (CHF) NOS | AHI 3% | 5.6 | 1.01 (1.00 – 1.01) | 1.0E-03 | 18332 | 1.00 (1.00 – 1.01) | 9404 | 1.01 (1.00 – 1.01) |
| 558 | Noninfectious gastroenteritis | Per88 | 5.2 | 1.01 (1.01 – 1.02) | 1.1E-03 | 12232 | 1.01 (1.01 – 1.02) | 5845 | 1.01 (1.01 – 1.02) |
| 585.3 | Chronic renal failure [CKD] | Per88 | 5.6 | 1.02 (1.01 – 1.03) | 1.1E-03 | 12232 | 1.01 (1.00 – 1.02) | 5845 | 1.02 (1.01 – 1.03) |
| 216 | Benign neoplasm of skin | Per88 | 14.0 | 0.99 (0.99 – 1.00) | 1.2E-03 | 12232 | 0.99 (0.99 – 1.00) | 5845 | 0.99 (0.99 – 1.00) |
| 427.21 | Atrial fibrillation | Per88 | 9.2 | 1.01 (1.00 – 1.02) | 1.3E-03 | 12232 | 1.01 (1.00 – 1.01) | 5845 | 1.01 (1.00 – 1.02) |
| 415.11 | Pulmonary embolism and infarction, acute | Per88 | 3.0 | 1.02 (1.01 – 1.03) | 1.3E-03 | 12232 | 1.01 (1.00 – 1.03) | 5845 | 1.02 (1.01 – 1.03) |
| 271.3 | Intestinal disaccharidase deficiencies and disaccharide malabsorption | AHI 3% | 14.0 | 1.00 (1.00 – 1.01) | 1.3E-03 | 18332 | 1.00 (1.00 – 1.00) | 9404 | 1.00 (1.00 – 1.01) |
| 519.8 | Other diseases of respiratory system, NEC | Per88 | 3.5 | 1.02 (1.01 – 1.03) | 1.4E-03 | 12232 | 1.01 (1.00 – 1.02) | 5845 | 1.02 (1.01 – 1.03) |
| 443.9 | Peripheral vascular disease, unspecified | Per88 | 1.6 | 1.03 (1.01 – 1.04) | 1.5E-03 | 12232 | 1.02 (1.00 – 1.03) | 5845 | 1.03 (1.01 – 1.04) |
| 274.1 | Gout | AHI 3% | 4.7 | 1.01 (1.00 – 1.01) | 1.6E-03 | 18332 | 1.00 (1.00 – 1.01) | 9404 | 1.01 (1.00 – 1.01) |
| 272.12 | Hyperglyceridemia | AHI 3% | 2.5 | 1.01 (1.00 – 1.01) | 1.6E-03 | 18332 | 1.00 (1.00 – 1.01) | 9404 | 1.01 (1.00 – 1.01) |
| 357 | Inflammatory and toxic neuropathy | Per88 | 5.8 | 1.01 (1.00 – 1.02) | 1.7E-03 | 12232 | 1.01 (1.00 – 1.02) | 5845 | 1.01 (1.00 – 1.02) |
| 743.11 | Osteoporosis NOS | AHI 3% | 7.2 | 1.00 (1.00 – 1.01) | 1.8E-03 | 18332 | 1.00 (1.00 – 1.01) | 9404 | 1.00 (1.00 – 1.01) |
| 560.4 | Other intestinal obstruction | AHI 3% | 1.9 | 1.01 (1.00 – 1.02) | 1.8E-03 | 18332 | 1.01 (1.00 – 1.01) | 9404 | 1.01 (1.00 – 1.02) |
| 728.7 | Fasciitis | Per88 | 5.3 | 0.98 (0.97 – 0.99) | 1.8E-03 | 12232 | 0.99 (0.98 – 1.00) | 5845 | 0.98 (0.97 – 0.99) |
| 527.7 | Disturbance of salivary secretion | Per88 | 1.5 | 1.02 (1.01 – 1.03) | 1.9E-03 | 12232 | 1.02 (1.01 – 1.04) | 5845 | 1.02 (1.01 – 1.03) |
| 585.3 | Chronic renal failure [CKD] | AHI 3% | 5.6 | 1.01 (1.00 – 1.01) | 1.9E-03 | 18332 | 1.01 (1.00 – 1.01) | 9404 | 1.01 (1.00 – 1.01) |
| 292.3 | Memory loss | AHI 3% | 6.1 | 1.00 (0.99 – 1.00) | 2.0E-03 | 18332 | 1.00 (0.99 – 1.00) | 9404 | 1.00 (0.99 – 1.00) |
| 250.42 | Other abnormal glucose | AHI 3% | 11.1 | 1.00 (1.00 – 1.01) | 2.1E-03 | 18332 | 1.00 (1.00 – 1.01) | 9404 | 1.00 (1.00 – 1.01) |
| 228.1 | Hemangioma of skin and subcutaneous tissue | Per88 | 5.0 | 0.98 (0.97 – 0.99) | 2.4E-03 | 12232 | 0.99 (0.98 – 1.00) | 5845 | 0.98 (0.97 – 0.99) |

| PheCode | Disease | Sleep statistic | Prevalence | Risk ratio | P-value | N | Risk ratio (AHI >15) | N (AHI >15) | Risk ratio (adj sleep disorders) |
|---|---|---|---|---|---|---|---|---|---|
| 938.2 | Chronic dermatitis due to solar radiation | Per88 | 5.9 | 0.98 (0.98 – 0.99) | 2.4E-03 | 12232 | 0.99 (0.98 – 1.00) | 5845 | 0.98 (0.98 – 0.99) |
| 798 | Malaise and fatigue | AHI 3% | 19.1 | 1.00 (1.00 – 1.00) | 2.6E-03 | 18332 | 1.00 (1.00 – 1.00) | 9404 | 1.00 (1.00 – 1.00) |
| 539 | Bariatric surgery | Per88 | 12.4 | 0.99 (0.98 – 1.00) | 2.6E-03 | 12232 | 1.00 (0.99 – 1.00) | 5845 | 0.99 (0.98 – 1.00) |
| 427.7 | Tachycardia NOS | AHI 3% | 7.4 | 1.00 (1.00 – 1.01) | 2.7E-03 | 18332 | 1.00 (1.00 – 1.01) | 9404 | 1.00 (1.00 – 1.01) |
| 272.1 | Hyperlipidemia | AHI 3% | 30.5 | 1.00 (1.00 – 1.00) | 2.8E-03 | 18332 | 1.00 (1.00 – 1.00) | 9404 | 1.00 (1.00 – 1.00) |
| 532 | Dysphagia | Per88 | 7.4 | 1.01 (1.00 – 1.02) | 3.1E-03 | 12232 | 1.01 (1.00 – 1.01) | 5845 | 1.01 (1.00 – 1.02) |
| 571.5 | Other chronic nonalcoholic liver disease | AHI 3% | 5.1 | 1.01 (1.00 – 1.01) | 3.3E-03 | 18332 | 1.00 (1.00 – 1.01) | 9404 | 1.01 (1.00 – 1.01) |
| 456 | Chronic venous insufficiency [CVI] | Per88 | 2.9 | 1.02 (1.01 – 1.03) | 3.3E-03 | 12232 | 1.01 (1.00 – 1.03) | 5845 | 1.02 (1.01 – 1.03) |
| 300.11 | Generalized anxiety disorder | AHI 3% | 7.6 | 1.00 (0.99 – 1.00) | 3.3E-03 | 18332 | 1.00 (0.99 – 1.00) | 9404 | 1.00 (0.99 – 1.00) |
| 798.1 | Chronic fatigue syndrome | AHI 3% | 4.4 | 1.00 (0.99 – 1.00) | 3.3E-03 | 18332 | 0.99 (0.99 – 1.00) | 9404 | 1.00 (0.99 – 1.00) |
| 480.11 | Pneumococcal pneumonia | Per88 | 1.5 | 1.02 (1.01 – 1.04) | 3.5E-03 | 12232 | 1.02 (1.00 – 1.03) | 5845 | 1.02 (1.01 – 1.04) |
| 291.4 | Specific nonpsychotic mental disorders due to brain damage | Per88 | 1.7 | 0.98 (0.96 – 0.99) | 4.9E-03 | 12232 | 0.98 (0.96 – 1.00) | 5845 | 0.98 (0.96 – 0.99) |
| 564 | Functional digestive disorders | Per88 | 1.8 | 1.02 (1.01 – 1.03) | 5.4E-03 | 12232 | 1.01 (0.99 – 1.02) | 5845 | 1.02 (1.01 – 1.03) |
| 480 | Pneumonia | AHI 3% | 7.9 | 1.00 (1.00 – 1.01) | 5.7E-03 | 18332 | 1.00 (1.00 – 1.01) | 9404 | 1.00 (1.00 – 1.01) |
| 411.2 | Myocardial infarction | Per88 | 5.1 | 1.01 (1.00 – 1.02) | 5.7E-03 | 12232 | 1.01 (1.00 – 1.02) | 5845 | 1.01 (1.00 – 1.02) |
| 339 | Other headache syndromes | Per88 | 13.0 | 0.99 (0.99 – 1.00) | 5.7E-03 | 12232 | 1.00 (0.99 – 1.00) | 5845 | 0.99 (0.99 – 1.00) |
| 496.21 | Obstructive chronic bronchitis | AHI 3% | 1.3 | 0.99 (0.98 – 1.00) | 6.2E-03 | 18332 | 0.99 (0.98 – 1.00) | 9404 | 0.99 (0.98 – 1.00) |
| 287.3 | Thrombocytopenia | Per88 | 2.7 | 1.02 (1.00 – 1.03) | 6.3E-03 | 12232 | 1.01 (1.00 – 1.02) | 5845 | 1.02 (1.00 – 1.03) |
| 292.2 | Mild cognitive impairment | Per88 | 2.4 | 0.98 (0.96 – 0.99) | 6.9E-03 | 12232 | 0.99 (0.98 – 1.00) | 5845 | 0.98 (0.96 – 0.99) |
| 375.1 | Dry eyes | AHI 3% | 4.8 | 1.00 (0.99 – 1.00) | 7.0E-03 | 18332 | 0.99 (0.99 – 1.00) | 9404 | 1.00 (0.99 – 1.00) |
| 575.7 | Other disorders of gallbladder | Per88 | 2.1 | 1.02 (1.01 – 1.03) | 7.2E-03 | 12232 | 1.02 (1.00 – 1.03) | 5845 | 1.02 (1.01 – 1.03) |
| 276.14 | Hypopotassemia | AHI 3% | 2.8 | 1.01 (1.00 – 1.01) | 7.3E-03 | 18332 | 1.00 (1.00 – 1.01) | 9404 | 1.01 (1.00 – 1.01) |
| 726.1 | Enthesopathy | AHI 3% | 11.0 | 1.00 (0.99 – 1.00) | 7.7E-03 | 18332 | 1.00 (0.99 – 1.00) | 9404 | 1.00 (0.99 – 1.00) |
| 302.1 | Decreased libido | Per88 | 1.5 | 0.98 (0.96 – 0.99) | 7.9E-03 | 12232 | 0.98 (0.96 – 1.00) | 5845 | 0.98 (0.96 – 0.99) |
| 728.7 | Fasciitis | AHI 3% | 5.3 | 1.00 (0.99 – 1.00) | 7.9E-03 | 18332 | 1.00 (0.99 – 1.00) | 9404 | 1.00 (0.99 – 1.00) |
| 112 | Candidiasis | Per88 | 5.3 | 1.01 (1.00 – 1.02) | 8.7E-03 | 12232 | 1.01 (1.00 – 1.02) | 5845 | 1.01 (1.00 – 1.02) |
| 528 | Diseases of the oral soft tissues, excluding lesions specific for gingiva and tongue | AHI 3% | 2.4 | 0.99 (0.99 – 1.00) | 8.8E-03 | 18332 | 0.99 (0.99 – 1.00) | 9404 | 0.99 (0.99 – 1.00) |
| 165.1 | Cancer of bronchus; lung | Per88 | 1.2 | 1.02 (1.01 – 1.04) | 8.8E-03 | 12232 | 1.01 (1.00 – 1.03) | 5845 | 1.02 (1.01 – 1.04) |
| 261.2 | Vitamin B-complex deficiencies | AHI 3% | 2.6 | 0.99 (0.99 – 1.00) | 8.8E-03 | 18332 | 1.00 (0.99 – 1.00) | 9404 | 0.99 (0.99 – 1.00) |
| 454.11 | Varicose veins of lower extremity, symptomtic | AHI 3% | 2.3 | 0.99 (0.99 – 1.00) | 9.0E-03 | 18332 | 0.99 (0.98 – 1.00) | 9404 | 0.99 (0.99 – 1.00) |
| 495 | Asthma | AHI 3% | 19.3 | 1.00 (1.00 – 1.00) | 9.5E-03 | 18332 | 1.00 (1.00 – 1.00) | 9404 | 1.00 (1.00 – 1.00) |
| 386.9 | Dizziness and giddiness (Light-headedness and vertigo) | Per88 | 13.5 | 0.99 (0.99 – 1.00) | 1.0E-02 | 12232 | 0.99 (0.99 – 1.00) | 5845 | 0.99 (0.99 – 1.00) |
| 701.2 | Scar conditions and fibrosis of skin | Per88 | 6.0 | 0.99 (0.98 – 1.00) | 1.0E-02 | 12232 | 0.99 (0.98 – 1.00) | 5845 | 0.99 (0.98 – 1.00) |
| 418.1 | Precordial pain | Per88 | 3.4 | 1.01 (1.00 – 1.02) | 1.1E-02 | 12232 | 1.01 (1.00 – 1.02) | 5845 | 1.01 (1.00 – 1.02) |

| PheCode | Disease | Sleep statistic | Prevalence | Risk ratio | P-value | N | Risk ratio (AHI >15) | N (AHI >15) | Risk ratio (adj sleep disorders) |
|---|---|---|---|---|---|---|---|---|---|
| 340.1 | Migraine with aura | Per88 | 3.5 | 0.99 (0.98 – 1.00) | 1.1E-02 | 12232 | 1.00 (0.98 – 1.01) | 5845 | 0.99 (0.98 – 1.00) |
| 519.8 | Other diseases of respiratory system, NEC | AHI 3% | 3.5 | 1.01 (1.00 – 1.01) | 1.1E-02 | 18332 | 1.00 (1.00 – 1.01) | 9404 | 1.01 (1.00 – 1.01) |
| 480.11 | Pneumococcal pneumonia | AHI 3% | 1.5 | 1.01 (1.00 – 1.02) | 1.1E-02 | 18332 | 1.01 (1.00 – 1.02) | 9404 | 1.01 (1.00 – 1.02) |
| 306 | Other mental disorder | Per88 | 8.8 | 1.01 (1.00 – 1.01) | 1.1E-02 | 12232 | 1.01 (1.00 – 1.01) | 5845 | 1.01 (1.00 – 1.01) |
| 296 | Mood disorders | AHI 3% | 6.3 | 1.00 (1.00 – 1.01) | 1.1E-02 | 18332 | 1.00 (1.00 – 1.01) | 9404 | 1.00 (1.00 – 1.01) |
| 458.9 | Hypotension NOS | AHI 3% | 3.6 | 1.01 (1.00 – 1.01) | 1.2E-02 | 18332 | 1.00 (1.00 – 1.01) | 9404 | 1.01 (1.00 – 1.01) |
| 368 | Visual disturbances | AHI 3% | 6.7 | 1.00 (0.99 – 1.00) | 1.2E-02 | 18332 | 1.00 (0.99 – 1.00) | 9404 | 1.00 (0.99 – 1.00) |
| 381.1 | Otitis media | Per88 | 2.4 | 1.02 (1.00 – 1.03) | 1.2E-02 | 12232 | 1.02 (1.01 – 1.03) | 5845 | 1.02 (1.00 – 1.03) |
| 840.3 | Joint/ligament sprain | AHI 3% | 5.4 | 1.00 (0.99 – 1.00) | 1.3E-02 | 18332 | 1.00 (0.99 – 1.00) | 9404 | 1.00 (0.99 – 1.00) |
| 278.1 | Obesity | AHI 3% | 35.1 | 1.00 (1.00 – 1.00) | 1.3E-02 | 18332 | 1.00 (1.00 – 1.00) | 9404 | 1.00 (1.00 – 1.00) |
| 411.8 | Other chronic ischemic heart disease, unspecified | AHI 3% | 1.5 | 1.01 (1.00 – 1.02) | 1.4E-02 | 18332 | 1.01 (1.00 – 1.01) | 9404 | 1.01 (1.00 – 1.02) |
| 770 | Myalgia and myositis unspecified | AHI 3% | 6.5 | 1.00 (0.99 – 1.00) | 1.4E-02 | 18332 | 1.00 (0.99 – 1.00) | 9404 | 1.00 (0.99 – 1.00) |
| 427.22 | Atrial flutter | Per88 | 2.6 | 1.02 (1.00 – 1.03) | 1.5E-02 | 12232 | 1.01 (1.00 – 1.02) | 5845 | 1.02 (1.00 – 1.03) |
| 578.9 | Hemorrhage of gastrointestinal tract | Per88 | 2.9 | 1.02 (1.00 – 1.03) | 1.6E-02 | 12232 | 1.01 (1.00 – 1.02) | 5845 | 1.02 (1.00 – 1.03) |
| 531.4 | Peptic ulcer, site unspecified | Per88 | 1.3 | 1.02 (1.00 – 1.04) | 1.6E-02 | 12232 | 1.01 (1.00 – 1.03) | 5845 | 1.02 (1.00 – 1.04) |
| 317.1 | Alcoholism | AHI 3% | 2.9 | 1.01 (1.00 – 1.01) | 1.7E-02 | 18332 | 1.00 (1.00 – 1.01) | 9404 | 1.01 (1.00 – 1.01) |
| 279 | Disorders involving the immune mechanism | Per88 | 2.1 | 1.02 (1.00 – 1.03) | 1.7E-02 | 12232 | 1.01 (1.00 – 1.02) | 5845 | 1.02 (1.00 – 1.03) |
| 416 | Cardiomegaly | AHI 3% | 2.6 | 1.01 (1.00 – 1.01) | 1.7E-02 | 18332 | 1.00 (1.00 – 1.01) | 9404 | 1.01 (1.00 – 1.01) |
| 278.11 | Morbid obesity | AHI 3% | 16.1 | 1.00 (1.00 – 1.00) | 1.7E-02 | 18332 | 1.00 (1.00 – 1.00) | 9404 | 1.00 (1.00 – 1.00) |
| 516.1 | Hemoptysis | Per88 | 1.5 | 1.02 (1.00 – 1.04) | 1.8E-02 | 12232 | 1.02 (1.01 – 1.04) | 5845 | 1.02 (1.00 – 1.04) |
| 250.1 | Type 1 diabetes | AHI 3% | 1.4 | 1.01 (1.00 – 1.02) | 1.9E-02 | 18332 | 1.01 (1.00 – 1.02) | 9404 | 1.01 (1.00 – 1.02) |
| 420.2 | Pericarditis | AHI 3% | 1.5 | 1.01 (1.00 – 1.01) | 1.9E-02 | 18332 | 1.01 (1.00 – 1.01) | 9404 | 1.01 (1.00 – 1.01) |
| 172.21 | Basal cell carcinoma | Per88 | 3.9 | 0.98 (0.97 – 1.00) | 2.0E-02 | 12232 | 0.99 (0.98 – 1.00) | 5845 | 0.98 (0.97 – 1.00) |
| 452.2 | Deep vein thrombosis [DVT] | AHI 3% | 3.5 | 0.99 (0.99 – 1.00) | 2.1E-02 | 18332 | 1.00 (0.99 – 1.00) | 9404 | 0.99 (0.99 – 1.00) |
| 564.1 | Irritable Bowel Syndrome | AHI 3% | 5.6 | 1.00 (0.99 – 1.00) | 2.1E-02 | 18332 | 1.00 (0.99 – 1.00) | 9404 | 1.00 (0.99 – 1.00) |
| 599.9 | Other abnormality of urination | AHI 3% | 4.6 | 1.00 (0.99 – 1.00) | 2.2E-02 | 18332 | 1.00 (0.99 – 1.00) | 9404 | 1.00 (0.99 – 1.00) |
| 452 | Other venous embolism and thrombosis | AHI 3% | 4.3 | 1.00 (0.99 – 1.00) | 2.2E-02 | 18332 | 1.00 (0.99 – 1.00) | 9404 | 1.00 (0.99 – 1.00) |
| 427.12 | Paroxysmal ventricular tachycardia | AHI 3% | 1.5 | 1.01 (1.00 – 1.01) | 2.2E-02 | 18332 | 1.01 (1.00 – 1.01) | 9404 | 1.01 (1.00 – 1.01) |
| 292 | Neurological disorders | Per88 | 3.8 | 0.99 (0.98 – 1.00) | 2.3E-02 | 12232 | 0.99 (0.98 – 1.00) | 5845 | 0.99 (0.98 – 1.00) |
| 807 | Fracture of ribs | AHI 3% | 1.4 | 1.01 (1.00 – 1.02) | 2.3E-02 | 18332 | 1.01 (1.00 – 1.02) | 9404 | 1.01 (1.00 – 1.02) |
| 772.3 | Muscle weakness | AHI 3% | 2.7 | 0.99 (0.99 – 1.00) | 2.3E-02 | 18332 | 0.99 (0.99 – 1.00) | 9404 | 0.99 (0.99 – 1.00) |
| 577.3 | Cyst and pseudocyst of pancreas | Per88 | 1.2 | 1.02 (1.00 – 1.04) | 2.3E-02 | 12232 | 1.02 (1.01 – 1.04) | 5845 | 1.02 (1.00 – 1.04) |
| 290.2 | Delirium due to conditions classified elsewhere | AHI 3% | 1.2 | 1.01 (1.00 – 1.02) | 2.4E-02 | 18332 | 1.01 (1.00 – 1.02) | 9404 | 1.01 (1.00 – 1.02) |
| 427.11 | Paroxysmal supraventricular tachycardia | Per88 | 3.1 | 1.01 (1.00 – 1.02) | 2.5E-02 | 12232 | 1.01 (0.99 – 1.02) | 5845 | 1.01 (1.00 – 1.02) |

| PheCode | Disease | Sleep statistic | Prevalence | Risk ratio | P-value | N | Risk ratio (AHI >15) | N (AHI >15) | Risk ratio (adj sleep disorders) |
|---|---|---|---|---|---|---|---|---|---|
| 427.6 | Premature beats | Per88 | 3.4 | 1.01 (1.00 – 1.02) | 2.5E-02 | 12232 | 1.01 (1.00 – 1.02) | 5845 | 1.01 (1.00 – 1.02) |
| 130.1 | Lyme disease | Per88 | 1.4 | 0.98 (0.96 – 1.00) | 2.5E-02 | 12232 | 0.98 (0.96 – 1.00) | 5845 | 0.98 (0.96 – 1.00) |
| 474.2 | Chronic tonsillitis and adenoiditis | AHI 3% | 2.5 | 1.01 (1.00 – 1.01) | 2.6E-02 | 18332 | 1.01 (1.00 – 1.02) | 9404 | 1.01 (1.00 – 1.01) |
| 550.4 | Umbilical hernia | Per88 | 3.1 | 1.01 (1.00 – 1.03) | 2.6E-02 | 12232 | 1.01 (1.00 – 1.02) | 5845 | 1.01 (1.00 – 1.03) |
| 368 | Visual disturbances | Per88 | 6.7 | 0.99 (0.98 – 1.00) | 2.6E-02 | 12232 | 0.99 (0.98 – 1.00) | 5845 | 0.99 (0.98 – 1.00) |
| 306.9 | Tension headache | Per88 | 3.5 | 0.99 (0.98 – 1.00) | 2.7E-02 | 12232 | 0.99 (0.98 – 1.00) | 5845 | 0.99 (0.98 – 1.00) |
| 414 | Other forms of chronic heart disease | AHI 3% | 2.3 | 1.01 (1.00 – 1.01) | 2.7E-02 | 18332 | 1.00 (1.00 – 1.01) | 9404 | 1.01 (1.00 – 1.01) |
| 338.2 | Chronic pain | AHI 3% | 15.2 | 1.00 (1.00 – 1.00) | 2.7E-02 | 18332 | 1.00 (0.99 – 1.00) | 9404 | 1.00 (1.00 – 1.00) |
| 782.3 | Edema | AHI 3% | 9.5 | 1.00 (1.00 – 1.01) | 2.8E-02 | 18332 | 1.00 (1.00 – 1.01) | 9404 | 1.00 (1.00 – 1.01) |
| 496 | Chronic airway obstruction | AHI 3% | 3.9 | 0.99 (0.99 – 1.00) | 2.9E-02 | 18332 | 1.00 (0.99 – 1.00) | 9404 | 0.99 (0.99 – 1.00) |
| 473 | Diseases of the larynx and vocal cords | AHI 3% | 2.2 | 1.01 (1.00 – 1.01) | 3.0E-02 | 18332 | 1.00 (1.00 – 1.01) | 9404 | 1.01 (1.00 – 1.01) |
| 465 | Acute upper respiratory infections of multiple or unspecified sites | AHI 3% | 10.9 | 1.00 (1.00 – 1.00) | 3.0E-02 | 18332 | 1.00 (1.00 – 1.00) | 9404 | 1.00 (1.00 – 1.00) |
| 269 | Proteinuria | Per88 | 3.0 | 1.01 (1.00 – 1.03) | 3.1E-02 | 12232 | 1.01 (0.99 – 1.02) | 5845 | 1.01 (1.00 – 1.03) |
| 300.4 | Dysthymic disorder | AHI 3% | 5.2 | 1.00 (0.99 – 1.00) | 3.2E-02 | 18332 | 1.00 (0.99 – 1.00) | 9404 | 1.00 (0.99 – 1.00) |
| 565.1 | Anal and rectal polyp | AHI 3% | 3.8 | 1.00 (1.00 – 1.01) | 3.2E-02 | 18332 | 1.00 (1.00 – 1.01) | 9404 | 1.00 (1.00 – 1.01) |
| 427.3 | Other specified cardiac dysrhythmias | Per88 | 7.4 | 0.99 (0.98 – 1.00) | 3.2E-02 | 12232 | 0.99 (0.98 – 1.00) | 5845 | 0.99 (0.98 – 1.00) |
| 041 | Bacterial infection NOS | AHI 3% | 2.1 | 1.01 (1.00 – 1.01) | 3.3E-02 | 18332 | 1.01 (1.00 – 1.01) | 9404 | 1.01 (1.00 – 1.01) |
| 250.1 | Type 1 diabetes | Per88 | 1.4 | 1.02 (1.00 – 1.04) | 3.4E-02 | 12232 | 1.02 (1.00 – 1.04) | 5845 | 1.02 (1.00 – 1.04) |
| 772.2 | Spasm of muscle | AHI 3% | 2.9 | 1.00 (0.99 – 1.00) | 3.5E-02 | 18332 | 1.00 (0.99 – 1.00) | 9404 | 1.00 (0.99 – 1.00) |
| 569 | Other disorders of intestine | Per88 | 8.3 | 1.01 (1.00 – 1.01) | 3.5E-02 | 12232 | 1.01 (1.00 – 1.02) | 5845 | 1.01 (1.00 – 1.01) |
| 296 | Mood disorders | Per88 | 6.3 | 1.01 (1.00 – 1.01) | 3.5E-02 | 12232 | 1.00 (1.00 – 1.01) | 5845 | 1.01 (1.00 – 1.01) |
| 473 | Diseases of the larynx and vocal cords | Per88 | 2.2 | 1.01 (1.00 – 1.02) | 3.5E-02 | 12232 | 1.01 (1.00 – 1.03) | 5845 | 1.01 (1.00 – 1.02) |
| 557 | Intestinal malabsorption (non-celiac) | AHI 3% | 1.9 | 0.99 (0.99 – 1.00) | 3.6E-02 | 18332 | 1.00 (0.99 – 1.00) | 9404 | 0.99 (0.99 – 1.00) |
| 272.13 | Mixed hyperlipidemia | AHI 3% | 6.9 | 1.00 (1.00 – 1.01) | 3.6E-02 | 18332 | 1.00 (1.00 – 1.00) | 9404 | 1.00 (1.00 – 1.01) |
| 327.41 | Organic or persistent insomnia | Per88 | 3.0 | 0.99 (0.98 – 1.00) | 3.6E-02 | 12232 | 1.00 (0.98 – 1.01) | 5845 | 0.99 (0.98 – 1.00) |
| 476 | Allergic rhinitis | Per88 | 14.6 | 0.99 (0.99 – 1.00) | 3.6E-02 | 12232 | 0.99 (0.99 – 1.00) | 5845 | 0.99 (0.99 – 1.00) |
| 558 | Noninfectious gastroenteritis | AHI 3% | 5.2 | 1.00 (1.00 – 1.01) | 3.6E-02 | 18332 | 1.00 (1.00 – 1.01) | 9404 | 1.00 (1.00 – 1.01) |
| 477 | Epistaxis or throat hemorrhage | Per88 | 2.5 | 1.01 (1.00 – 1.03) | 3.6E-02 | 12232 | 1.01 (1.00 – 1.02) | 5845 | 1.01 (1.00 – 1.03) |
| 277.51 | Lipoprotein disorders | Per88 | 1.8 | 0.98 (0.96 – 1.00) | 3.7E-02 | 12232 | 0.98 (0.96 – 1.00) | 5845 | 0.98 (0.96 – 1.00) |
| 276.6 | Fluid overload | AHI 3% | 1.9 | 1.01 (1.00 – 1.01) | 3.7E-02 | 18332 | 1.00 (1.00 – 1.01) | 9404 | 1.01 (1.00 – 1.01) |
| 527.7 | Disturbance of salivary secretion | AHI 3% | 1.5 | 1.01 (1.00 – 1.01) | 3.8E-02 | 18332 | 1.01 (1.01 – 1.02) | 9404 | 1.01 (1.00 – 1.01) |
| 472 | Chronic pharyngitis and nasopharyngitis | AHI 3% | 7.9 | 1.00 (1.00 – 1.01) | 3.9E-02 | 18332 | 1.00 (1.00 – 1.00) | 9404 | 1.00 (1.00 – 1.01) |
| 550.4 | Umbilical hernia | AHI 3% | 3.1 | 1.01 (1.00 – 1.01) | 4.0E-02 | 18332 | 1.00 (1.00 – 1.01) | 9404 | 1.01 (1.00 – 1.01) |
| 452.2 | Deep vein thrombosis [DVT] | Per88 | 3.5 | 1.01 (1.00 – 1.02) | 4.2E-02 | 12232 | 1.01 (1.00 – 1.02) | 5845 | 1.01 (1.00 – 1.02) |
| 704.8 | Other specified diseases of hair and hair follicles | Per88 | 4.7 | 0.99 (0.98 – 1.00) | 4.3E-02 | 12232 | 0.99 (0.98 – 1.00) | 5845 | 0.99 (0.98 – 1.00) |
| 722.1 | Displacement of intervertebral disc | AHI 3% | 4.5 | 1.00 (0.99 – 1.00) | 4.4E-02 | 18332 | 0.99 (0.99 – 1.00) | 9404 | 1.00 (0.99 – 1.00) |

| PheCode | Disease | Sleep statistic | Prevalence | Risk ratio | P-value | N | Risk ratio (AHI >15) | N (AHI >15) | Risk ratio (adj sleep disorders) |
|---|---|---|---|---|---|---|---|---|---|
| 747.11 | Cardiac shunt/ heart septal defect | AHI 3% | 1.2 | 0.99 (0.99 – 1.00) | 4.5E-02 | 18332 | 0.99 (0.99 – 1.00) | 9404 | 0.99 (0.99 – 1.00) |
| 789 | Nausea and vomiting | AHI 3% | 13.3 | 1.00 (1.00 – 1.00) | 4.6E-02 | 18332 | 1.00 (1.00 – 1.00) | 9404 | 1.00 (1.00 – 1.00) |
| 196 | Radiotherapy | AHI 3% | 2.9 | 1.00 (1.00 – 1.01) | 4.6E-02 | 18332 | 1.00 (1.00 – 1.01) | 9404 | 1.00 (1.00 – 1.01) |
| 427.61 | Supraventricular premature beats | Per88 | 1.8 | 1.02 (1.00 – 1.03) | 4.6E-02 | 12232 | 1.01 (1.00 – 1.03) | 5845 | 1.02 (1.00 – 1.03) |
| 278.4 | Abnormal weight gain | Per88 | 6.5 | 0.99 (0.98 – 1.00) | 4.8E-02 | 12232 | 1.00 (0.99 – 1.01) | 5845 | 0.99 (0.98 – 1.00) |
| 452 | Other venous embolism and thrombosis | Per88 | 4.3 | 1.01 (1.00 – 1.02) | 4.8E-02 | 12232 | 1.01 (1.00 – 1.02) | 5845 | 1.01 (1.00 – 1.02) |
| 292.3 | Memory loss | Per88 | 6.1 | 0.99 (0.98 – 1.00) | 4.8E-02 | 12232 | 1.00 (0.99 – 1.00) | 5845 | 0.99 (0.98 – 1.00) |
| 687.1 | Rash and other nonspecific skin eruption | AHI 3% | 11.4 | 1.00 (1.00 – 1.00) | 4.9E-02 | 18332 | 1.00 (1.00 – 1.00) | 9404 | 1.00 (1.00 – 1.00) |
| 455 | Hemorrhoids | AHI 3% | 11.2 | 1.00 (1.00 – 1.00) | 5.0E-02 | 18332 | 1.00 (1.00 – 1.00) | 9404 | 1.00 (1.00 – 1.00) |
| 401.21 | Hypertensive heart disease | AHI 3% | 2.5 | 1.01 (1.00 – 1.01) | 5.0E-02 | 18332 | 1.00 (1.00 – 1.01) | 9404 | 1.01 (1.00 – 1.01) |
| 807 | Fracture of ribs | Per88 | 1.4 | 1.02 (1.00 – 1.03) | 5.0E-02 | 12232 | 1.01 (0.99 – 1.03) | 5845 | 1.02 (1.00 – 1.03) |
| 296.2 | Depression | AHI 3% | 11.9 | 1.00 (1.00 – 1.00) | 5.1E-02 | 18332 | 1.00 (1.00 – 1.00) | 9404 | 1.00 (1.00 – 1.00) |
| 261.2 | Vitamin B-complex deficiencies | Per88 | 2.6 | 0.99 (0.97 – 1.00) | 5.1E-02 | 12232 | 0.99 (0.97 – 1.01) | 5845 | 0.99 (0.97 – 1.00) |
| 345.1 | Epilepsy | AHI 3% | 1.7 | 0.99 (0.99 – 1.00) | 5.1E-02 | 18332 | 1.00 (0.99 – 1.00) | 9404 | 0.99 (0.99 – 1.00) |
| 401.3 | Other hypertensive complications | Per88 | 2.3 | 1.01 (1.00 – 1.03) | 5.2E-02 | 12232 | 1.01 (0.99 – 1.02) | 5845 | 1.01 (1.00 – 1.03) |
| 292 | Neurological disorders | AHI 3% | 3.8 | 1.00 (0.99 – 1.00) | 5.3E-02 | 18332 | 1.00 (1.00 – 1.01) | 9404 | 1.00 (0.99 – 1.00) |
| 367.4 | Presbyopia | AHI 3% | 2.5 | 1.00 (0.99 – 1.00) | 5.5E-02 | 18332 | 0.99 (0.99 – 1.00) | 9404 | 1.00 (0.99 – 1.00) |
| 496.3 | Bronchiectasis | Per88 | 1.2 | 1.02 (1.00 – 1.03) | 5.6E-02 | 12232 | 1.02 (1.00 – 1.04) | 5845 | 1.02 (1.00 – 1.03) |
| 306 | Other mental disorder | AHI 3% | 8.8 | 1.00 (1.00 – 1.01) | 5.7E-02 | 18332 | 1.00 (1.00 – 1.01) | 9404 | 1.00 (1.00 – 1.01) |
| 593.2 | Microscopic hematuria | AHI 3% | 4.2 | 1.00 (1.00 – 1.01) | 5.7E-02 | 18332 | 1.00 (1.00 – 1.01) | 9404 | 1.00 (1.00 – 1.01) |
| 575.7 | Other disorders of gallbladder | AHI 3% | 2.1 | 1.01 (1.00 – 1.01) | 5.8E-02 | 18332 | 1.00 (0.99 – 1.01) | 9404 | 1.01 (1.00 – 1.01) |
| 530.9 | Heartburn | AHI 3% | 5.7 | 1.00 (0.99 – 1.00) | 6.0E-02 | 18332 | 0.99 (0.99 – 1.00) | 9404 | 1.00 (0.99 – 1.00) |
| 427.5 | Arrhythmia (cardiac) NOS | Per88 | 13.9 | 1.01 (1.00 – 1.01) | 6.0E-02 | 12232 | 1.00 (1.00 – 1.01) | 5845 | 1.01 (1.00 – 1.01) |
| 798.1 | Chronic fatigue syndrome | Per88 | 4.4 | 0.99 (0.98 – 1.00) | 6.3E-02 | 12232 | 0.99 (0.98 – 1.00) | 5845 | 0.99 (0.98 – 1.00) |
| 374.3 | Ptosis of eyelid | Per88 | 1.5 | 0.98 (0.97 – 1.00) | 6.3E-02 | 12232 | 0.98 (0.97 – 1.00) | 5845 | 0.98 (0.97 – 1.00) |
| 526.41 | Temporomandibular joint disorder, unspecified | AHI 3% | 1.6 | 1.00 (0.99 – 1.00) | 6.4E-02 | 18332 | 0.99 (0.99 – 1.00) | 9404 | 1.00 (0.99 – 1.00) |
| 781 | Symptoms involving nervous and musculoskeletal systems | Per88 | 5.4 | 1.01 (1.00 – 1.02) | 6.5E-02 | 12232 | 1.00 (0.99 – 1.01) | 5845 | 1.01 (1.00 – 1.02) |
| 382 | Otalgia | Per88 | 7.9 | 0.99 (0.99 – 1.00) | 6.5E-02 | 12232 | 1.00 (0.99 – 1.00) | 5845 | 0.99 (0.99 – 1.00) |
| 291.4 | Specific nonpsychotic mental disorders due to brain damage | AHI 3% | 1.7 | 0.99 (0.99 – 1.00) | 6.5E-02 | 18332 | 1.00 (0.99 – 1.01) | 9404 | 1.00 (0.99 – 1.00) |
| 740.9 | Osteoarthrosis NOS | Per88 | 16.4 | 1.00 (1.00 – 1.01) | 6.5E-02 | 12232 | 1.00 (1.00 – 1.01) | 5845 | 1.00 (1.00 – 1.01) |
| 426.31 | Right bundle branch block | Per88 | 2.2 | 1.01 (1.00 – 1.03) | 6.6E-02 | 12232 | 1.01 (1.00 – 1.02) | 5845 | 1.01 (1.00 – 1.03) |
| 402 | Elevated blood pressure reading without diagnosis of hypertension | AHI 3% | 10.0 | 1.00 (1.00 – 1.00) | 6.6E-02 | 18332 | 1.00 (1.00 – 1.00) | 9404 | 1.00 (1.00 – 1.00) |
| 304 | Adjustment reaction | AHI 3% | 8.0 | 1.00 (0.99 – 1.00) | 6.6E-02 | 18332 | 1.00 (0.99 – 1.00) | 9404 | 1.00 (1.00 – 1.00) |
| 280.2 | Iron deficiency anemia secondary to blood loss (chronic) | Per88 | 2.3 | 1.01 (1.00 – 1.03) | 6.7E-02 | 12232 | 1.01 (1.00 – 1.03) | 5845 | 1.01 (1.00 – 1.03) |

| PheCode | Disease | Sleep statistic | Prevalence | Risk ratio | P-value | N | Risk ratio (AHI >15) | N (AHI >15) | Risk ratio (adj sleep disorders) |
|---|---|---|---|---|---|---|---|---|---|
| 512.9 | Other dyspnea | AHI 3% | 18.4 | 1.00 (1.00 – 1.00) | 6.7E-02 | 18332 | 1.00 (1.00 – 1.00) | 9404 | 1.00 (1.00 – 1.00) |
| 726 | Peripheral enthesopathies and allied syndromes | Per88 | 8.1 | 0.99 (0.99 – 1.00) | 6.8E-02 | 12232 | 1.00 (0.99 – 1.01) | 5845 | 0.99 (0.99 – 1.00) |
| 300.12 | Agorophobia, social phobia, and panic disorder | AHI 3% | 4.2 | 1.00 (0.99 – 1.00) | 6.8E-02 | 18332 | 1.00 (0.99 – 1.00) | 9404 | 1.00 (0.99 – 1.00) |
| 594.1 | Calculus of kidney | Per88 | 7.3 | 0.99 (0.98 – 1.00) | 6.9E-02 | 12232 | 0.99 (0.98 – 1.00) | 5845 | 0.99 (0.98 – 1.00) |
| 561.1 | Diarrhea | Per88 | 9.7 | 1.01 (1.00 – 1.01) | 7.0E-02 | 12232 | 1.01 (1.00 – 1.01) | 5845 | 1.01 (1.00 – 1.01) |
| 715 | Other inflammatory spondylopathies | AHI 3% | 1.0 | 0.99 (0.98 – 1.00) | 7.0E-02 | 18332 | 0.99 (0.99 – 1.00) | 9404 | 0.99 (0.98 – 1.00) |
| 578.9 | Hemorrhage of gastrointestinal tract | AHI 3% | 2.9 | 1.00 (1.00 – 1.01) | 7.1E-02 | 18332 | 1.00 (1.00 – 1.01) | 9404 | 1.00 (1.00 – 1.01) |
| 726.2 | Synoviopathy | Per88 | 1.8 | 0.99 (0.97 – 1.00) | 7.2E-02 | 12232 | 0.99 (0.97 – 1.00) | 5845 | 0.99 (0.97 – 1.00) |
| 345 | Epilepsy, recurrent seizures, convulsions | AHI 3% | 2.7 | 1.00 (0.99 – 1.00) | 7.3E-02 | 18332 | 1.00 (0.99 – 1.00) | 9404 | 1.00 (0.99 – 1.00) |
| 130.1 | Lyme disease | AHI 3% | 1.4 | 0.99 (0.99 – 1.00) | 7.5E-02 | 18332 | 0.99 (0.98 – 1.00) | 9404 | 0.99 (0.99 – 1.00) |
| 747.11 | Cardiac shunt/ heart septal defect | Per88 | 1.2 | 1.01 (1.00 – 1.03) | 7.5E-02 | 12232 | 1.01 (1.00 – 1.03) | 5845 | 1.01 (1.00 – 1.03) |
| 244.4 | Hypothyroidism NOS | Per88 | 11.7 | 1.01 (1.00 – 1.01) | 7.5E-02 | 12232 | 1.00 (1.00 – 1.01) | 5845 | 1.01 (1.00 – 1.01) |
| 586 | Other disorders of the kidney and ureters | Per88 | 5.2 | 1.01 (1.00 – 1.02) | 7.6E-02 | 12232 | 1.01 (1.00 – 1.02) | 5845 | 1.01 (1.00 – 1.02) |
| 689 | Disorder of skin and subcutaneous tissue NOS | Per88 | 13.7 | 1.00 (0.99 – 1.00) | 7.7E-02 | 12232 | 0.99 (0.99 – 1.00) | 5845 | 1.00 (0.99 – 1.00) |
| 720.1 | Spinal stenosis of lumbar region | Per88 | 4.3 | 1.01 (1.00 – 1.02) | 7.9E-02 | 12232 | 1.01 (1.00 – 1.02) | 5845 | 1.01 (1.00 – 1.02) |
| 454.1 | Varicose veins of lower extremity | AHI 3% | 3.1 | 1.00 (0.99 – 1.00) | 8.1E-02 | 18332 | 1.00 (0.99 – 1.00) | 9404 | 1.00 (0.99 – 1.00) |
| 564 | Functional digestive disorders | AHI 3% | 1.8 | 1.01 (1.00 – 1.01) | 8.4E-02 | 18332 | 1.01 (1.00 – 1.01) | 9404 | 1.01 (1.00 – 1.01) |
| 300.8 | Acute reaction to stress | Per88 | 2.2 | 0.99 (0.97 – 1.00) | 8.4E-02 | 12232 | 0.99 (0.98 – 1.01) | 5845 | 0.99 (0.97 – 1.00) |
| 819 | Skull and face fracture and other intercranial injury | AHI 3% | 1.4 | 0.99 (0.99 – 1.00) | 8.5E-02 | 18332 | 0.99 (0.99 – 1.00) | 9404 | 0.99 (0.99 – 1.00) |
| 428.3 | Heart failure with reduced EF [Systolic or combined heart failure] | AHI 3% | 1.9 | 1.01 (1.00 – 1.01) | 8.5E-02 | 18332 | 1.00 (0.99 – 1.01) | 9404 | 1.01 (1.00 – 1.01) |
| 456 | Chronic venous insufficiency [CVI] | AHI 3% | 2.9 | 1.00 (0.99 – 1.00) | 8.7E-02 | 18332 | 1.00 (0.99 – 1.00) | 9404 | 1.00 (0.99 – 1.00) |
| 427.9 | Palpitations | Per88 | 12.6 | 1.00 (0.99 – 1.00) | 8.8E-02 | 12232 | 0.99 (0.99 – 1.00) | 5845 | 1.00 (0.99 – 1.00) |
| 433.21 | Cerebral artery occlusion, with cerebral infarction | AHI 3% | 4.3 | 1.00 (1.00 – 1.01) | 8.8E-02 | 18332 | 1.00 (1.00 – 1.01) | 9404 | 1.00 (1.00 – 1.01) |
| 702.1 | Actinic keratosis | Per88 | 5.3 | 0.99 (0.98 – 1.00) | 8.9E-02 | 12232 | 0.99 (0.98 – 1.00) | 5845 | 0.99 (0.98 – 1.00) |
| 272.11 | Hypercholesterolemia | Per88 | 20.2 | 1.00 (0.99 – 1.00) | 9.0E-02 | 12232 | 1.00 (0.99 – 1.00) | 5845 | 1.00 (0.99 – 1.00) |
| 276.12 | Hyposmolality and/or hyponatremia | Per88 | 2.6 | 1.01 (1.00 – 1.02) | 9.0E-02 | 12232 | 1.01 (1.00 – 1.02) | 5845 | 1.01 (1.00 – 1.02) |
| 290.3 | Other persistent mental disorders due to conditions classified elsewhere | AHI 3% | 1.1 | 0.99 (0.99 – 1.00) | 9.2E-02 | 18332 | 1.00 (0.99 – 1.01) | 9404 | 0.99 (0.99 – 1.00) |
| 573.7 | Abnormal results of function study of liver | AHI 3% | 3.8 | 1.00 (1.00 – 1.01) | 9.2E-02 | 18332 | 1.00 (1.00 – 1.01) | 9404 | 1.00 (1.00 – 1.01) |
| 053 | Herpes zoster | AHI 3% | 4.7 | 1.00 (1.00 – 1.01) | 9.4E-02 | 18332 | 1.00 (1.00 – 1.01) | 9404 | 1.00 (1.00 – 1.01) |
| 773 | Pain in limb | AHI 3% | 21.2 | 1.00 (1.00 – 1.00) | 9.4E-02 | 18332 | 1.00 (1.00 – 1.00) | 9404 | 1.00 (1.00 – 1.00) |
| 355.1 | Chronic pain syndrome | Per88 | 1.5 | 1.01 (1.00 – 1.03) | 9.5E-02 | 12232 | 1.01 (1.00 – 1.03) | 5845 | 1.01 (1.00 – 1.03) |

| PheCode | Disease | Sleep statistic | Prevalence | Risk ratio | P-value | N | Risk ratio (AHI >15) | N (AHI >15) | Risk ratio (adj sleep disorders) |
|---|---|---|---|---|---|---|---|---|---|
| 280.1 | Iron deficiency anemias, unspecified or not due to blood loss | AHI 3% | 6.2 | 1.00 (0.99 – 1.00) | 9.5E-02 | 18332 | 1.00 (0.99 – 1.00) | 9404 | 1.00 (0.99 – 1.00) |
| 389.4 | Tinnitus | AHI 3% | 6.2 | 1.00 (0.99 – 1.00) | 9.5E-02 | 18332 | 1.00 (1.00 – 1.00) | 9404 | 1.00 (0.99 – 1.00) |
| 388 | Other disorders of ear | Per88 | 3.1 | 0.99 (0.98 – 1.00) | 9.6E-02 | 12232 | 0.98 (0.97 – 1.00) | 5845 | 0.99 (0.98 – 1.00) |
| 690.1 | Seborrheic dermatitis | Per88 | 4.1 | 0.99 (0.98 – 1.00) | 9.8E-02 | 12232 | 0.99 (0.98 – 1.00) | 5845 | 0.99 (0.98 – 1.00) |
| 459.9 | Circulatory disease NEC | Per88 | 4.5 | 1.01 (1.00 – 1.02) | 9.9E-02 | 12232 | 1.00 (0.99 – 1.01) | 5845 | 1.01 (1.00 – 1.02) |
| 733 | Other disorders of bone and cartilage | AHI 3% | 10.2 | 1.00 (1.00 – 1.00) | 1.0E-01 | 18332 | 1.00 (1.00 – 1.00) | 9404 | 1.00 (1.00 – 1.00) |
| 288.1 | Decreased white blood cell count | AHI 3% | 1.2 | 0.99 (0.99 – 1.00) | 1.0E-01 | 18332 | 0.99 (0.99 – 1.00) | 9404 | 0.99 (0.99 – 1.00) |
| 366 | Cataract | Per88 | 5.3 | 1.01 (1.00 – 1.02) | 1.0E-01 | 12232 | 1.01 (1.00 – 1.02) | 5845 | 1.01 (1.00 – 1.02) |
| 1013 | Asphyxia and hypoxemia | AHI 3% | 3.8 | 1.00 (1.00 – 1.01) | 1.1E-01 | 18332 | 1.01 (1.00 – 1.01) | 9404 | 1.00 (1.00 – 1.01) |
| 726.1 | Enthesopathy | Per88 | 11.0 | 0.99 (0.99 – 1.00) | 1.1E-01 | 12232 | 1.00 (0.99 – 1.00) | 5845 | 0.99 (0.99 – 1.00) |
| 772 | Symptoms of the muscles | Per88 | 2.0 | 0.99 (0.98 – 1.00) | 1.1E-01 | 12232 | 0.99 (0.97 – 1.01) | 5845 | 0.99 (0.98 – 1.00) |
| 585.4 | Chronic kidney disease, Stage I or II | AHI 3% | 2.0 | 1.01 (1.00 – 1.01) | 1.1E-01 | 18332 | 1.00 (1.00 – 1.01) | 9404 | 1.01 (1.00 – 1.01) |
| 735.3 | Hallux valgus (Bunion) | AHI 3% | 2.9 | 1.00 (0.99 – 1.00) | 1.1E-01 | 18332 | 1.00 (0.99 – 1.00) | 9404 | 1.00 (0.99 – 1.00) |
| 381.1 | Otitis media | AHI 3% | 2.4 | 1.00 (1.00 – 1.01) | 1.1E-01 | 18332 | 1.00 (1.00 – 1.01) | 9404 | 1.00 (1.00 – 1.01) |
| 368.2 | Diplopia and disorders of binocular vision | AHI 3% | 1.9 | 1.00 (0.99 – 1.00) | 1.1E-01 | 18332 | 1.00 (0.99 – 1.00) | 9404 | 1.00 (0.99 – 1.00) |
| 704.8 | Other specified diseases of hair and hair follicles | AHI 3% | 4.7 | 1.00 (0.99 – 1.00) | 1.1E-01 | 18332 | 1.00 (1.00 – 1.01) | 9404 | 1.00 (0.99 – 1.00) |
| 690.1 | Seborrheic dermatitis | AHI 3% | 4.1 | 1.00 (0.99 – 1.00) | 1.1E-01 | 18332 | 1.00 (1.00 – 1.01) | 9404 | 1.00 (0.99 – 1.00) |
| 704.1 | Alopecia | AHI 3% | 5.8 | 1.00 (1.00 – 1.00) | 1.1E-01 | 18332 | 1.00 (1.00 – 1.00) | 9404 | 1.00 (1.00 – 1.00) |
| 737.3 | Kyphoscoliosis and scoliosis | Per88 | 1.8 | 1.01 (1.00 – 1.02) | 1.1E-01 | 12232 | 1.01 (1.00 – 1.03) | 5845 | 1.01 (1.00 – 1.02) |
| 946 | Anaphylactic shock NOS | AHI 3% | 1.9 | 1.00 (0.99 – 1.00) | 1.2E-01 | 18332 | 0.99 (0.99 – 1.00) | 9404 | 1.00 (0.99 – 1.00) |
| 496.2 | Chronic bronchitis | AHI 3% | 1.1 | 0.99 (0.99 – 1.00) | 1.2E-01 | 18332 | 0.99 (0.98 – 1.00) | 9404 | 0.99 (0.99 – 1.00) |
| 350.1 | Abnormal involuntary movements | Per88 | 5.4 | 0.99 (0.98 – 1.00) | 1.2E-01 | 12232 | 1.00 (0.99 – 1.01) | 5845 | 0.99 (0.98 – 1.00) |
| 724.8 | Other symptoms referable to back | AHI 3% | 1.3 | 0.99 (0.99 – 1.00) | 1.2E-01 | 18332 | 0.99 (0.99 – 1.00) | 9404 | 0.99 (0.99 – 1.00) |
| 593 | Hematuria | AHI 3% | 7.9 | 1.00 (1.00 – 1.01) | 1.2E-01 | 18332 | 1.00 (1.00 – 1.01) | 9404 | 1.00 (1.00 – 1.01) |
| 327.71 | Restless legs syndrome | Per88 | 4.3 | 0.99 (0.98 – 1.00) | 1.2E-01 | 12232 | 0.99 (0.98 – 1.00) | 5845 | 0.99 (0.98 – 1.00) |
| 594.8 | Renal colic | Per88 | 1.4 | 0.98 (0.96 – 1.00) | 1.2E-01 | 12232 | 0.98 (0.96 – 1.01) | 5845 | 0.98 (0.96 – 1.00) |
| 740.9 | Osteoarthrosis NOS | AHI 3% | 16.4 | 1.00 (1.00 – 1.00) | 1.2E-01 | 18332 | 1.00 (1.00 – 1.00) | 9404 | 1.00 (1.00 – 1.00) |
| 136 | Other infectious and parasitic diseases | AHI 3% | 4.6 | 1.00 (1.00 – 1.01) | 1.2E-01 | 18332 | 1.00 (1.00 – 1.01) | 9404 | 1.00 (1.00 – 1.01) |
| 840.3 | Joint/ligament sprain | Per88 | 5.4 | 0.99 (0.98 – 1.00) | 1.2E-01 | 12232 | 1.00 (0.99 – 1.01) | 5845 | 0.99 (0.98 – 1.00) |
| 429.3 | Symptoms involving cardiovascular system | Per88 | 6.9 | 1.01 (1.00 – 1.01) | 1.3E-01 | 12232 | 1.00 (1.00 – 1.01) | 5845 | 1.01 (1.00 – 1.01) |
| 772 | Symptoms of the muscles | AHI 3% | 2.0 | 1.00 (0.99 – 1.00) | 1.3E-01 | 18332 | 1.00 (0.99 – 1.01) | 9404 | 1.00 (0.99 – 1.00) |
| 941 | Adverse reaction to serum or vaccine | Per88 | 2.7 | 0.99 (0.98 – 1.00) | 1.3E-01 | 12232 | 0.99 (0.98 – 1.01) | 5845 | 0.99 (0.98 – 1.00) |
| 252.1 | Hyperparathyroidism | AHI 3% | 1.9 | 1.00 (1.00 – 1.01) | 1.3E-01 | 18332 | 1.01 (1.00 – 1.01) | 9404 | 1.00 (1.00 – 1.01) |
| 564.1 | Irritable Bowel Syndrome | Per88 | 5.6 | 0.99 (0.99 – 1.00) | 1.3E-01 | 12232 | 0.99 (0.98 – 1.00) | 5845 | 0.99 (0.99 – 1.00) |

| PheCode | Disease | Sleep statistic | Prevalence | Risk ratio | P-value | N | Risk ratio (AHI >15) | N (AHI >15) | Risk ratio (adj sleep disorders) |
|---|---|---|---|---|---|---|---|---|---|
| 279 | Disorders involving the immune mechanism | AHI 3% | 2.1 | 1.00 (1.00 – 1.01) | 1.3E-01 | 18332 | 1.01 (1.00 – 1.01) | 9404 | 1.00 (1.00 – 1.01) |
| 458.1 | Orthostatic hypotension | AHI 3% | 1.7 | 1.00 (0.99 – 1.00) | 1.3E-01 | 18332 | 0.99 (0.99 – 1.00) | 9404 | 1.00 (0.99 – 1.00) |
| 338.2 | Chronic pain | Per88 | 15.2 | 1.00 (1.00 – 1.01) | 1.3E-01 | 12232 | 1.00 (1.00 – 1.01) | 5845 | 1.00 (1.00 – 1.01) |
| 173 | Neoplasm of uncertain behavior of skin | Per88 | 9.4 | 0.99 (0.99 – 1.00) | 1.3E-01 | 12232 | 1.00 (0.99 – 1.00) | 5845 | 0.99 (0.99 – 1.00) |
| 280.1 | Iron deficiency anemias, unspecified or not due to blood loss | Per88 | 6.2 | 1.01 (1.00 – 1.01) | 1.3E-01 | 12232 | 1.01 (1.00 – 1.01) | 5845 | 1.01 (1.00 – 1.01) |
| 241.2 | Nontoxic multinodular goiter | Per88 | 3.0 | 0.99 (0.98 – 1.00) | 1.3E-01 | 12232 | 0.99 (0.98 – 1.00) | 5845 | 0.99 (0.98 – 1.00) |
| 447 | Other disorders of arteries and arterioles | AHI 3% | 1.3 | 1.01 (1.00 – 1.01) | 1.3E-01 | 18332 | 1.01 (1.00 – 1.01) | 9404 | 1.01 (1.00 – 1.01) |
| 217.1 | Nevus, non-neoplastic | Per88 | 5.2 | 0.99 (0.98 – 1.00) | 1.3E-01 | 12232 | 0.99 (0.98 – 1.00) | 5845 | 0.99 (0.98 – 1.00) |
| 696.41 | Psoriasis vulgaris | AHI 3% | 3.3 | 1.00 (1.00 – 1.01) | 1.3E-01 | 18332 | 1.00 (1.00 – 1.01) | 9404 | 1.00 (1.00 – 1.01) |
| 733 | Other disorders of bone and cartilage | Per88 | 10.2 | 1.00 (1.00 – 1.01) | 1.4E-01 | 12232 | 1.00 (1.00 – 1.01) | 5845 | 1.00 (1.00 – 1.01) |
| 302.1 | Decreased libido | AHI 3% | 1.5 | 1.00 (0.99 – 1.00) | 1.4E-01 | 18332 | 1.00 (0.99 – 1.00) | 9404 | 1.00 (0.99 – 1.00) |
| 426.32 | Left bundle branch block | AHI 3% | 1.3 | 1.01 (1.00 – 1.01) | 1.4E-01 | 18332 | 1.00 (0.99 – 1.01) | 9404 | 1.01 (1.00 – 1.01) |
| 277 | Other disorders of metabolism | AHI 3% | 6.5 | 1.00 (0.99 – 1.00) | 1.4E-01 | 18332 | 1.00 (1.00 – 1.00) | 9404 | 1.00 (0.99 – 1.00) |
| 686.1 | Carbuncle and furuncle | Per88 | 1.6 | 1.01 (1.00 – 1.03) | 1.4E-01 | 12232 | 1.01 (0.99 – 1.03) | 5845 | 1.01 (1.00 – 1.03) |
| 278.11 | Morbid obesity | Per88 | 16.1 | 1.00 (1.00 – 1.01) | 1.5E-01 | 12232 | 1.00 (1.00 – 1.01) | 5845 | 1.00 (1.00 – 1.01) |
| 427.11 | Paroxysmal supraventricular tachycardia | AHI 3% | 3.1 | 1.00 (1.00 – 1.01) | 1.5E-01 | 18332 | 1.00 (1.00 – 1.01) | 9404 | 1.00 (1.00 – 1.01) |
| 442.1 | Aortic aneurysm | Per88 | 1.4 | 1.02 (0.99 – 1.04) | 1.5E-01 | 12232 | 1.01 (0.99 – 1.03) | 5845 | 1.02 (0.99 – 1.04) |
| 196 | Radiotherapy | Per88 | 2.9 | 1.01 (1.00 – 1.02) | 1.5E-01 | 12232 | 1.00 (0.99 – 1.02) | 5845 | 1.01 (1.00 – 1.02) |
| 701 | Other hypertrophic and atrophic conditions of skin | Per88 | 11.5 | 1.00 (0.99 – 1.00) | 1.5E-01 | 12232 | 0.99 (0.99 – 1.00) | 5845 | 1.00 (0.99 – 1.00) |
| 426.32 | Left bundle branch block | Per88 | 1.3 | 1.01 (0.99 – 1.03) | 1.5E-01 | 12232 | 1.01 (0.99 – 1.02) | 5845 | 1.01 (0.99 – 1.03) |
| 537 | Other disorders of stomach and duodenum | Per88 | 7.8 | 0.99 (0.99 – 1.00) | 1.6E-01 | 12232 | 0.99 (0.98 – 1.00) | 5845 | 0.99 (0.99 – 1.00) |
| 586.4 | Stricture/obstruction of ureter | Per88 | 2.5 | 0.99 (0.97 – 1.00) | 1.6E-01 | 12232 | 0.99 (0.98 – 1.01) | 5845 | 0.99 (0.97 – 1.00) |
| 721.1 | Spondylosis without myelopathy | AHI 3% | 8.9 | 1.00 (1.00 – 1.00) | 1.6E-01 | 18332 | 1.00 (0.99 – 1.00) | 9404 | 1.00 (1.00 – 1.00) |
| 531.2 | Gastric ulcer | Per88 | 1.7 | 1.01 (1.00 – 1.03) | 1.6E-01 | 12232 | 1.01 (0.99 – 1.02) | 5845 | 1.01 (1.00 – 1.03) |
| 292.2 | Mild cognitive impairment | AHI 3% | 2.4 | 1.00 (0.99 – 1.00) | 1.6E-01 | 18332 | 1.00 (1.00 – 1.01) | 9404 | 1.00 (0.99 – 1.00) |
| 300.1 | Anxiety disorder | AHI 3% | 24.6 | 1.00 (1.00 – 1.00) | 1.6E-01 | 18332 | 1.00 (1.00 – 1.00) | 9404 | 1.00 (1.00 – 1.00) |
| 965 | Poisoning by analgesics, antipyretics, and antirheumatics | Per88 | 4.3 | 1.01 (1.00 – 1.02) | 1.6E-01 | 12232 | 1.00 (0.99 – 1.01) | 5845 | 1.01 (1.00 – 1.02) |
| 530.13 | Barrett's esophagus | Per88 | 1.9 | 1.01 (1.00 – 1.03) | 1.6E-01 | 12232 | 1.00 (0.98 – 1.01) | 5845 | 1.01 (1.00 – 1.03) |
| 251.1 | Hypoglycemia | Per88 | 1.6 | 1.01 (1.00 – 1.03) | 1.6E-01 | 12232 | 1.01 (0.99 – 1.03) | 5845 | 1.01 (1.00 – 1.03) |
| 599.4 | Urinary incontinence | AHI 3% | 6.7 | 1.00 (0.99 – 1.00) | 1.6E-01 | 18332 | 1.00 (1.00 – 1.00) | 9404 | 1.00 (0.99 – 1.00) |
| 763 | Thoracic or lumbosacral neuritis or radiculitis, unspecified | AHI 3% | 7.2 | 1.00 (1.00 – 1.00) | 1.7E-01 | 18332 | 1.00 (0.99 – 1.00) | 9404 | 1.00 (1.00 – 1.00) |
| 300.4 | Dysthymic disorder | Per88 | 5.2 | 0.99 (0.99 – 1.00) | 1.7E-01 | 12232 | 0.99 (0.98 – 1.00) | 5845 | 0.99 (0.99 – 1.00) |
| 427.12 | Paroxysmal ventricular tachycardia | Per88 | 1.5 | 1.01 (0.99 – 1.03) | 1.7E-01 | 12232 | 1.01 (1.00 – 1.03) | 5845 | 1.01 (1.00 – 1.03) |

| PheCode | Disease | Sleep statistic | Prevalence | Risk ratio | P-value | N | Risk ratio (AHI >15) | N (AHI >15) | Risk ratio (adj sleep disorders) |
|---|---|---|---|---|---|---|---|---|---|
| 727.6 | Rupture of tendon, nontraumatic | Per88 | 2.4 | 0.99 (0.97 – 1.01) | 1.7E-01 | 12232 | 0.99 (0.97 – 1.00) | 5845 | 0.99 (0.97 – 1.01) |
| 785 | Abdominal pain | Per88 | 20.0 | 1.00 (0.99 – 1.00) | 1.7E-01 | 12232 | 0.99 (0.99 – 1.00) | 5845 | 1.00 (0.99 – 1.00) |
| 287.3 | Thrombocytopenia | AHI 3% | 2.7 | 1.00 (1.00 – 1.01) | 1.7E-01 | 18332 | 1.00 (1.00 – 1.01) | 9404 | 1.00 (1.00 – 1.01) |
| 228.1 | Hemangioma of skin and subcutaneous tissue | AHI 3% | 5.0 | 1.00 (0.99 – 1.00) | 1.7E-01 | 18332 | 1.00 (1.00 – 1.01) | 9404 | 1.00 (0.99 – 1.00) |
| 386.9 | Dizziness and giddiness (Light-headedness and vertigo) | AHI 3% | 13.5 | 1.00 (1.00 – 1.00) | 1.7E-01 | 18332 | 1.00 (1.00 – 1.00) | 9404 | 1.00 (1.00 – 1.00) |
| 420.2 | Pericarditis | Per88 | 1.5 | 1.01 (0.99 – 1.03) | 1.7E-01 | 12232 | 1.01 (0.99 – 1.02) | 5845 | 1.01 (0.99 – 1.03) |
| 172.2 | Other non-epithelial cancer of skin | Per88 | 5.7 | 0.99 (0.98 – 1.00) | 1.7E-01 | 12232 | 1.00 (0.99 – 1.01) | 5845 | 0.99 (0.98 – 1.00) |
| 788 | Syncope and collapse | Per88 | 8.1 | 1.00 (1.00 – 1.01) | 1.7E-01 | 12232 | 1.00 (1.00 – 1.01) | 5845 | 1.00 (1.00 – 1.01) |
| 172.21 | Basal cell carcinoma | AHI 3% | 3.9 | 1.00 (0.99 – 1.00) | 1.7E-01 | 18332 | 1.00 (0.99 – 1.00) | 9404 | 1.00 (0.99 – 1.00) |
| 454.11 | Varicose veins of lower extremity, symptomtic | Per88 | 2.3 | 1.01 (1.00 – 1.02) | 1.7E-01 | 12232 | 1.01 (1.00 – 1.02) | 5845 | 1.01 (1.00 – 1.02) |
| 535.2 | Atrophic gastritis | AHI 3% | 2.7 | 1.00 (0.99 – 1.00) | 1.8E-01 | 18332 | 1.00 (0.99 – 1.01) | 9404 | 1.00 (0.99 – 1.00) |
| 380.1 | Otitis externa | AHI 3% | 2.2 | 1.00 (0.99 – 1.00) | 1.8E-01 | 18332 | 1.00 (0.99 – 1.01) | 9404 | 1.00 (0.99 – 1.00) |
| 242 | Thyrotoxicosis with or without goiter | AHI 3% | 2.4 | 1.00 (1.00 – 1.01) | 1.8E-01 | 18332 | 1.01 (1.00 – 1.01) | 9404 | 1.00 (1.00 – 1.01) |
| 703.1 | Ingrowing nail | Per88 | 1.9 | 1.01 (0.99 – 1.03) | 1.8E-01 | 12232 | 1.01 (0.99 – 1.03) | 5845 | 1.01 (0.99 – 1.03) |
| 274.11 | Gouty arthropathy | Per88 | 2.3 | 1.01 (0.99 – 1.03) | 1.8E-01 | 12232 | 1.01 (0.99 – 1.02) | 5845 | 1.01 (0.99 – 1.03) |
| 008.5 | Bacterial enteritis | Per88 | 3.8 | 0.99 (0.98 – 1.00) | 1.9E-01 | 12232 | 1.00 (0.99 – 1.01) | 5845 | 0.99 (0.98 – 1.00) |
| 304 | Adjustment reaction | Per88 | 8.0 | 1.00 (0.99 – 1.00) | 1.9E-01 | 12232 | 1.00 (0.99 – 1.00) | 5845 | 1.00 (0.99 – 1.00) |
| 938.2 | Chronic dermatitis due to solar radiation | AHI 3% | 5.9 | 1.00 (0.99 – 1.00) | 1.9E-01 | 18332 | 1.00 (1.00 – 1.00) | 9404 | 1.00 (0.99 – 1.00) |
| 313.1 | Attention deficit hyperactivity disorder | AHI 3% | 5.7 | 1.00 (1.00 – 1.00) | 1.9E-01 | 18332 | 1.00 (0.99 – 1.00) | 9404 | 1.00 (1.00 – 1.00) |
| 560.4 | Other intestinal obstruction | Per88 | 1.9 | 1.01 (1.00 – 1.02) | 1.9E-01 | 12232 | 1.00 (0.99 – 1.02) | 5845 | 1.01 (1.00 – 1.02) |
| 716.9 | Arthropathy NOS | AHI 3% | 2.2 | 1.00 (0.99 – 1.00) | 1.9E-01 | 18332 | 0.99 (0.98 – 1.00) | 9404 | 1.00 (0.99 – 1.00) |
| 395.2 | Nonrheumatic aortic valve disorders | AHI 3% | 2.9 | 1.00 (1.00 – 1.01) | 1.9E-01 | 18332 | 1.00 (1.00 – 1.01) | 9404 | 1.00 (1.00 – 1.01) |
| 723 | Other disorders of cervical region | Per88 | 1.4 | 0.99 (0.98 – 1.00) | 1.9E-01 | 12232 | 0.98 (0.97 – 1.00) | 5845 | 0.99 (0.98 – 1.00) |
| 289.9 | Abnormality of red blood cells | Per88 | 2.5 | 1.01 (1.00 – 1.02) | 1.9E-01 | 12232 | 1.01 (0.99 – 1.02) | 5845 | 1.01 (1.00 – 1.02) |
| 536.8 | Dyspepsia and other specified disorders of function of stomach | Per88 | 3.9 | 0.99 (0.99 – 1.00) | 1.9E-01 | 12232 | 0.99 (0.98 – 1.00) | 5845 | 0.99 (0.99 – 1.00) |
| 371.3 | Inflammation of eyelids | Per88 | 3.2 | 0.99 (0.98 – 1.00) | 2.0E-01 | 12232 | 1.00 (0.98 – 1.01) | 5845 | 0.99 (0.98 – 1.00) |
| 272.11 | Hypercholesterolemia | AHI 3% | 20.2 | 1.00 (1.00 – 1.00) | 2.0E-01 | 18332 | 1.00 (1.00 – 1.00) | 9404 | 1.00 (1.00 – 1.00) |
| 496.1 | Emphysema | AHI 3% | 2.0 | 1.00 (0.99 – 1.00) | 2.0E-01 | 18332 | 1.00 (0.99 – 1.01) | 9404 | 1.00 (0.99 – 1.00) |
| 278.1 | Obesity | Per88 | 35.1 | 1.00 (0.99 – 1.00) | 2.0E-01 | 12232 | 1.00 (1.00 – 1.00) | 5845 | 1.00 (0.99 – 1.00) |
| 300.9 | Posttraumatic stress disorder | Per88 | 4.1 | 1.01 (1.00 – 1.01) | 2.0E-01 | 12232 | 1.01 (0.99 – 1.02) | 5845 | 1.01 (1.00 – 1.01) |
| 350.2 | Abnormality of gait | Per88 | 4.7 | 0.99 (0.98 – 1.00) | 2.0E-01 | 12232 | 0.99 (0.98 – 1.00) | 5845 | 0.99 (0.98 – 1.00) |
| 300.12 | Agorophobia, social phobia, and panic disorder | Per88 | 4.2 | 1.01 (1.00 – 1.01) | 2.0E-01 | 12232 | 1.00 (0.99 – 1.01) | 5845 | 1.01 (1.00 – 1.01) |
| 290.3 | Other persistent mental disorders due to conditions classified elsewhere | Per88 | 1.1 | 0.99 (0.97 – 1.01) | 2.0E-01 | 12232 | 1.00 (0.98 – 1.02) | 5845 | 0.99 (0.97 – 1.01) |

| PheCode | Disease | Sleep statistic | Prevalence | Risk ratio | P-value | N | Risk ratio (AHI >15) | N (AHI >15) | Risk ratio (adj sleep disorders) |
|---|---|---|---|---|---|---|---|---|---|
| 379.9 | Pain, swelling or discharge of eye | AHI 3% | 4.4 | 1.00 (0.99 – 1.00) | 2.0E-01 | 18332 | 1.00 (0.99 – 1.00) | 9404 | 1.00 (0.99 – 1.00) |
| 078 | Viral warts & HPV | Per88 | 4.9 | 0.99 (0.99 – 1.00) | 2.0E-01 | 12232 | 1.00 (0.99 – 1.01) | 5845 | 0.99 (0.99 – 1.00) |
| 840 | Sprains and strains | Per88 | 9.8 | 1.00 (0.99 – 1.00) | 2.1E-01 | 12232 | 1.00 (0.99 – 1.01) | 5845 | 1.00 (0.99 – 1.00) |
| 530.14 | Reflux esophagitis | Per88 | 3.1 | 0.99 (0.98 – 1.00) | 2.1E-01 | 12232 | 1.00 (0.98 – 1.01) | 5845 | 0.99 (0.98 – 1.00) |
| 327.71 | Restless legs syndrome | AHI 3% | 4.3 | 1.00 (0.99 – 1.00) | 2.1E-01 | 18332 | 1.00 (1.00 – 1.00) | 9404 | 1.00 (1.00 – 1.00) |
| 316 | Substance addiction and disorders | AHI 3% | 5.5 | 1.00 (1.00 – 1.01) | 2.1E-01 | 18332 | 1.00 (1.00 – 1.01) | 9404 | 1.00 (1.00 – 1.01) |
| 112 | Candidiasis | AHI 3% | 5.3 | 1.00 (0.99 – 1.00) | 2.1E-01 | 18332 | 1.00 (1.00 – 1.01) | 9404 | 1.00 (0.99 – 1.00) |
| 110.11 | Dermatophytosis of nail | AHI 3% | 4.4 | 1.00 (0.99 – 1.00) | 2.2E-01 | 18332 | 1.00 (0.99 – 1.00) | 9404 | 1.00 (0.99 – 1.00) |
| 261.4 | Vitamin D deficiency | Per88 | 11.6 | 1.00 (0.99 – 1.00) | 2.2E-01 | 12232 | 1.00 (0.99 – 1.00) | 5845 | 1.00 (0.99 – 1.00) |
| 528 | Diseases of the oral soft tissues, excluding lesions specific for gingiva and tongue | Per88 | 2.4 | 1.01 (1.00 – 1.02) | 2.2E-01 | 12232 | 1.01 (0.99 – 1.02) | 5845 | 1.01 (1.00 – 1.02) |
| 526 | Diseases of the jaws | AHI 3% | 1.6 | 1.00 (0.99 – 1.00) | 2.2E-01 | 18332 | 1.00 (0.99 – 1.00) | 9404 | 1.00 (0.99 – 1.00) |
| 428.4 | Heart failure with preserved EF [Diastolic heart failure] | AHI 3% | 2.4 | 1.00 (1.00 – 1.01) | 2.2E-01 | 18332 | 1.00 (1.00 – 1.01) | 9404 | 1.00 (1.00 – 1.01) |
| 300.13 | Phobia | AHI 3% | 2.4 | 1.00 (0.99 – 1.00) | 2.2E-01 | 18332 | 1.00 (0.99 – 1.00) | 9404 | 1.00 (0.99 – 1.00) |
| 327 | Sleep disorders | Per88 | 8.0 | 1.00 (0.99 – 1.00) | 2.2E-01 | 12232 | 1.00 (0.99 – 1.01) | 5845 | 1.00 (0.99 – 1.00) |
| 594.3 | Calculus of ureter | Per88 | 1.5 | 0.99 (0.96 – 1.01) | 2.2E-01 | 12232 | 0.98 (0.96 – 1.00) | 5845 | 0.99 (0.96 – 1.01) |
| 365 | Glaucoma | Per88 | 5.6 | 1.01 (1.00 – 1.01) | 2.2E-01 | 12232 | 1.01 (1.00 – 1.02) | 5845 | 1.01 (1.00 – 1.01) |
| 415.2 | Chronic pulmonary heart disease | AHI 3% | 2.1 | 1.00 (1.00 – 1.01) | 2.2E-01 | 18332 | 1.00 (1.00 – 1.01) | 9404 | 1.00 (1.00 – 1.01) |
| 382 | Otalgia | AHI 3% | 7.9 | 1.00 (1.00 – 1.00) | 2.3E-01 | 18332 | 1.00 (1.00 – 1.00) | 9404 | 1.00 (1.00 – 1.00) |
| 278 | Overweight, obesity and other hyperalimentation | AHI 3% | 14.8 | 1.00 (1.00 – 1.00) | 2.3E-01 | 18332 | 1.00 (1.00 – 1.00) | 9404 | 1.00 (1.00 – 1.00) |
| 357 | Inflammatory and toxic neuropathy | AHI 3% | 5.8 | 1.00 (0.99 – 1.00) | 2.3E-01 | 18332 | 1.00 (1.00 – 1.00) | 9404 | 1.00 (0.99 – 1.00) |
| 366.2 | Senile cataract | AHI 3% | 6.6 | 1.00 (0.99 – 1.00) | 2.4E-01 | 18332 | 1.00 (1.00 – 1.00) | 9404 | 1.00 (0.99 – 1.00) |
| 565 | Anal and rectal conditions | Per88 | 3.8 | 0.99 (0.98 – 1.00) | 2.4E-01 | 12232 | 1.00 (0.98 – 1.01) | 5845 | 0.99 (0.98 – 1.00) |
| 447.7 | Aortic ectasia | AHI 3% | 1.3 | 1.00 (0.99 – 1.00) | 2.4E-01 | 18332 | 0.99 (0.98 – 1.00) | 9404 | 1.00 (0.99 – 1.00) |
| 367.2 | Astigmatism | AHI 3% | 2.8 | 1.00 (0.99 – 1.00) | 2.4E-01 | 18332 | 0.99 (0.99 – 1.00) | 9404 | 1.00 (0.99 – 1.00) |
| 263 | Other nutritional deficiency | AHI 3% | 1.3 | 1.00 (0.99 – 1.00) | 2.4E-01 | 18332 | 1.00 (0.99 – 1.01) | 9404 | 1.00 (0.99 – 1.00) |
| 761 | Cervicalgia | Per88 | 12.3 | 1.00 (0.99 – 1.00) | 2.4E-01 | 12232 | 1.00 (0.99 – 1.00) | 5845 | 1.00 (0.99 – 1.00) |
| 379.2 | Disorders of vitreous body | Per88 | 3.3 | 0.99 (0.98 – 1.00) | 2.4E-01 | 12232 | 1.00 (0.99 – 1.01) | 5845 | 0.99 (0.98 – 1.00) |
| 695.8 | Other specified erythematous conditions | AHI 3% | 1.9 | 1.00 (0.99 – 1.00) | 2.4E-01 | 18332 | 1.00 (0.99 – 1.00) | 9404 | 1.00 (0.99 – 1.00) |
| 110.13 | Dermatophytosis of the body | Per88 | 2.3 | 0.99 (0.97 – 1.01) | 2.4E-01 | 12232 | 0.99 (0.98 – 1.01) | 5845 | 0.99 (0.97 – 1.01) |
| 334 | Degenerative disease of the spinal cord | AHI 3% | 1.3 | 1.00 (1.00 – 1.01) | 2.4E-01 | 18332 | 1.00 (1.00 – 1.01) | 9404 | 1.00 (1.00 – 1.01) |
| 214 | Lipoma | AHI 3% | 2.6 | 1.00 (0.99 – 1.00) | 2.4E-01 | 18332 | 1.00 (0.99 – 1.00) | 9404 | 1.00 (0.99 – 1.00) |
| 402 | Elevated blood pressure reading without diagnosis of hypertension | Per88 | 10.0 | 1.00 (0.99 – 1.00) | 2.4E-01 | 12232 | 1.00 (0.99 – 1.00) | 5845 | 1.00 (0.99 – 1.00) |
| 351 | Other peripheral nerve disorders | AHI 3% | 7.5 | 1.00 (1.00 – 1.00) | 2.4E-01 | 18332 | 1.00 (0.99 – 1.00) | 9404 | 1.00 (1.00 – 1.00) |
| 165.1 | Cancer of bronchus; lung | AHI 3% | 1.2 | 1.00 (1.00 – 1.01) | 2.4E-01 | 18332 | 1.00 (1.00 – 1.01) | 9404 | 1.00 (1.00 – 1.01) |

| PheCode | Disease | Sleep statistic | Prevalence | Risk ratio | P-value | N | Risk ratio (AHI >15) | N (AHI >15) | Risk ratio (adj sleep disorders) |
|---|---|---|---|---|---|---|---|---|---|
| 726.2 | Synoviopathy | AHI 3% | 1.8 | 1.00 (0.99 – 1.00) | 2.5E-01 | 18332 | 0.99 (0.98 – 1.00) | 9404 | 1.00 (0.99 – 1.00) |
| 569 | Other disorders of intestine | AHI 3% | 8.3 | 1.00 (1.00 – 1.00) | 2.5E-01 | 18332 | 1.00 (1.00 – 1.00) | 9404 | 1.00 (1.00 – 1.00) |
| 947 | Urticaria | AHI 3% | 4.8 | 1.00 (0.99 – 1.00) | 2.5E-01 | 18332 | 1.00 (1.00 – 1.01) | 9404 | 1.00 (0.99 – 1.00) |
| 727.6 | Rupture of tendon, nontraumatic | AHI 3% | 2.4 | 1.00 (0.99 – 1.00) | 2.5E-01 | 18332 | 0.99 (0.99 – 1.00) | 9404 | 1.00 (0.99 – 1.00) |
| 411.2 | Myocardial infarction | AHI 3% | 5.1 | 1.00 (1.00 – 1.01) | 2.5E-01 | 18332 | 1.00 (1.00 – 1.01) | 9404 | 1.00 (1.00 – 1.01) |
| 242 | Thyrotoxicosis with or without goiter | Per88 | 2.4 | 0.99 (0.98 – 1.01) | 2.5E-01 | 12232 | 1.00 (0.98 – 1.01) | 5845 | 0.99 (0.98 – 1.01) |
| 704 | Diseases of hair and hair follicles | Per88 | 3.7 | 0.99 (0.98 – 1.00) | 2.5E-01 | 12232 | 1.00 (0.98 – 1.01) | 5845 | 0.99 (0.98 – 1.00) |
| 241.1 | Nontoxic uninodular goiter | Per88 | 5.6 | 0.99 (0.99 – 1.00) | 2.5E-01 | 12232 | 0.99 (0.99 – 1.00) | 5845 | 0.99 (0.99 – 1.00) |
| 252.1 | Hyperparathyroidism | Per88 | 1.9 | 1.01 (0.99 – 1.03) | 2.6E-01 | 12232 | 1.00 (0.99 – 1.02) | 5845 | 1.01 (0.99 – 1.03) |
| 701.2 | Scar conditions and fibrosis of skin | AHI 3% | 6.0 | 1.00 (1.00 – 1.00) | 2.6E-01 | 18332 | 1.00 (1.00 – 1.00) | 9404 | 1.00 (1.00 – 1.00) |
| 246 | Other disorders of thyroid | AHI 3% | 2.6 | 1.00 (1.00 – 1.01) | 2.6E-01 | 18332 | 1.00 (1.00 – 1.01) | 9404 | 1.00 (1.00 – 1.01) |
| 348.7 | Coma | Per88 | 1.5 | 1.01 (0.99 – 1.02) | 2.6E-01 | 12232 | 1.01 (0.99 – 1.02) | 5845 | 1.01 (0.99 – 1.02) |
| 841 | Sprains and strains of back and neck | Per88 | 2.3 | 0.99 (0.98 – 1.01) | 2.6E-01 | 12232 | 1.00 (0.98 – 1.01) | 5845 | 0.99 (0.98 – 1.01) |
| 593.1 | Gross hematuria | AHI 3% | 2.7 | 1.00 (1.00 – 1.01) | 2.6E-01 | 18332 | 1.00 (1.00 – 1.01) | 9404 | 1.00 (1.00 – 1.01) |
| 727 | Other disorders of synovium, tendon, and bursa | Per88 | 2.6 | 0.99 (0.98 – 1.01) | 2.6E-01 | 12232 | 0.99 (0.97 – 1.00) | 5845 | 0.99 (0.98 – 1.01) |
| 793.2 | Nonspecific abnormal findings on radiological and other examination of other intrathoracic organs (echocardiogram, etc) | Per88 | 4.0 | 1.01 (1.00 – 1.02) | 2.7E-01 | 12232 | 1.00 (0.99 – 1.01) | 5845 | 1.01 (1.00 – 1.02) |
| 244.4 | Hypothyroidism NOS | AHI 3% | 11.7 | 1.00 (1.00 – 1.00) | 2.7E-01 | 18332 | 1.00 (1.00 – 1.00) | 9404 | 1.00 (1.00 – 1.00) |
| 271.3 | Intestinal disaccharidase deficiencies and disaccharide malabsorption | Per88 | 14.0 | 1.00 (1.00 – 1.01) | 2.7E-01 | 12232 | 1.00 (1.00 – 1.01) | 5845 | 1.00 (1.00 – 1.01) |
| 172.22 | Squamous cell carcinoma | AHI 3% | 2.2 | 1.00 (0.99 – 1.00) | 2.7E-01 | 18332 | 1.00 (0.99 – 1.00) | 9404 | 1.00 (0.99 – 1.00) |
| 292.1 | Aphasia/speech disturbance | Per88 | 2.0 | 0.99 (0.98 – 1.01) | 2.8E-01 | 12232 | 0.99 (0.97 – 1.01) | 5845 | 0.99 (0.98 – 1.01) |
| 464 | Acute sinusitis | Per88 | 5.1 | 0.99 (0.99 – 1.00) | 2.8E-01 | 12232 | 1.00 (0.99 – 1.01) | 5845 | 0.99 (0.99 – 1.00) |
| 512.2 | Painful respiration | Per88 | 9.5 | 1.00 (1.00 – 1.01) | 2.8E-01 | 12232 | 1.00 (1.00 – 1.01) | 5845 | 1.00 (1.00 – 1.01) |
| 763 | Thoracic or lumbosacral neuritis or radiculitis, unspecified | Per88 | 7.2 | 1.00 (0.99 – 1.00) | 2.8E-01 | 12232 | 0.99 (0.99 – 1.00) | 5845 | 1.00 (0.99 – 1.00) |
| 740.1 | Osteoarthritis; localized | AHI 3% | 3.5 | 1.00 (0.99 – 1.00) | 2.8E-01 | 18332 | 1.00 (0.99 – 1.00) | 9404 | 1.00 (0.99 – 1.00) |
| 706.1 | Acne | AHI 3% | 5.6 | 1.00 (1.00 – 1.00) | 2.8E-01 | 18332 | 1.00 (1.00 – 1.01) | 9404 | 1.00 (1.00 – 1.00) |
| 720.1 | Spinal stenosis of lumbar region | AHI 3% | 4.3 | 1.00 (0.99 – 1.00) | 2.8E-01 | 18332 | 1.00 (0.99 – 1.00) | 9404 | 1.00 (0.99 – 1.00) |
| 296.22 | Major depressive disorder | AHI 3% | 22.7 | 1.00 (1.00 – 1.00) | 2.8E-01 | 18332 | 1.00 (1.00 – 1.00) | 9404 | 1.00 (1.00 – 1.00) |
| 280.2 | Iron deficiency anemia secondary to blood loss (chronic) | AHI 3% | 2.3 | 1.00 (0.99 – 1.00) | 2.8E-01 | 18332 | 1.00 (1.00 – 1.01) | 9404 | 1.00 (0.99 – 1.00) |
| 389 | Hearing loss | AHI 3% | 8.9 | 1.00 (1.00 – 1.00) | 2.9E-01 | 18332 | 1.00 (1.00 – 1.01) | 9404 | 1.00 (1.00 – 1.00) |
| 433.21 | Cerebral artery occlusion, with cerebral infarction | Per88 | 4.3 | 1.01 (1.00 – 1.02) | 2.9E-01 | 12232 | 1.00 (0.99 – 1.01) | 5845 | 1.01 (1.00 – 1.02) |
| 727.4 | Ganglion and cyst of synovium, tendon, | AHI 3% | 2.5 | 1.00 (0.99 – 1.00) | 2.9E-01 | 18332 | 1.00 (0.99 – 1.00) | 9404 | 1.00 (0.99 – 1.00) |

| PheCode | Disease | Sleep statistic | Prevalence | Risk ratio | P-value | N | Risk ratio (AHI >15) | N (AHI >15) | Risk ratio (adj sleep disorders) |
|---|---|---|---|---|---|---|---|---|---|
| | and bursa | | | | | | | | |
| 701 | Other hypertrophic and atrophic conditions of skin | AHI 3% | 11.5 | 1.00 (1.00 – 1.00) | 2.9E-01 | 18332 | 1.00 (1.00 – 1.00) | 9404 | 1.00 (1.00 – 1.00) |
| 296.1 | Bipolar | AHI 3% | 3.4 | 1.00 (0.99 – 1.00) | 2.9E-01 | 18332 | 1.00 (0.99 – 1.01) | 9404 | 1.00 (0.99 – 1.00) |
| 550.2 | Diaphragmatic hernia | AHI 3% | 5.5 | 1.00 (0.99 – 1.00) | 2.9E-01 | 18332 | 1.00 (0.99 – 1.00) | 9404 | 1.00 (0.99 – 1.00) |
| 594.1 | Calculus of kidney | AHI 3% | 7.3 | 1.00 (1.00 – 1.00) | 2.9E-01 | 18332 | 1.00 (1.00 – 1.00) | 9404 | 1.00 (1.00 – 1.00) |
| 536.8 | Dyspepsia and other specified disorders of function of stomach | AHI 3% | 3.9 | 1.00 (0.99 – 1.00) | 3.0E-01 | 18332 | 1.00 (0.99 – 1.00) | 9404 | 1.00 (0.99 – 1.00) |
| 598.9 | Other nonspecific findings on examination of urine | Per88 | 3.0 | 0.99 (0.98 – 1.01) | 3.0E-01 | 12232 | 0.99 (0.98 – 1.01) | 5845 | 0.99 (0.98 – 1.01) |
| 008 | Intestinal infection | Per88 | 4.1 | 1.00 (1.00 – 1.01) | 3.0E-01 | 12232 | 1.00 (0.99 – 1.01) | 5845 | 1.00 (1.00 – 1.01) |
| 272.12 | Hyperglyceridemia | Per88 | 2.5 | 1.01 (0.99 – 1.02) | 3.0E-01 | 12232 | 1.00 (0.99 – 1.02) | 5845 | 1.01 (0.99 – 1.02) |
| 496.3 | Bronchiectasis | AHI 3% | 1.2 | 1.00 (0.99 – 1.00) | 3.0E-01 | 18332 | 1.00 (0.99 – 1.00) | 9404 | 1.00 (0.99 – 1.00) |
| 599.5 | Frequency of urination and polyuria | AHI 3% | 9.8 | 1.00 (1.00 – 1.00) | 3.0E-01 | 18332 | 1.00 (1.00 – 1.00) | 9404 | 1.00 (1.00 – 1.00) |
| 741.2 | Stiffness of joint | AHI 3% | 2.5 | 1.00 (0.99 – 1.00) | 3.0E-01 | 18332 | 1.00 (0.99 – 1.00) | 9404 | 1.00 (0.99 – 1.00) |
| 355.1 | Chronic pain syndrome | AHI 3% | 1.5 | 1.00 (1.00 – 1.01) | 3.0E-01 | 18332 | 1.00 (0.99 – 1.01) | 9404 | 1.00 (1.00 – 1.01) |
| 704.1 | Alopecia | Per88 | 5.8 | 1.00 (0.99 – 1.00) | 3.0E-01 | 12232 | 1.00 (0.99 – 1.01) | 5845 | 1.00 (0.99 – 1.00) |
| 504 | Other alveolar and parietoalveolar pneumonopathy | AHI 3% | 1.3 | 1.00 (0.99 – 1.00) | 3.1E-01 | 18332 | 1.00 (0.99 – 1.00) | 9404 | 1.00 (0.99 – 1.00) |
| 475.9 | Postnasal drip | Per88 | 5.0 | 1.00 (1.00 – 1.01) | 3.1E-01 | 12232 | 1.00 (0.99 – 1.01) | 5845 | 1.00 (1.00 – 1.01) |
| 947 | Urticaria | Per88 | 4.8 | 1.00 (0.99 – 1.00) | 3.1E-01 | 12232 | 1.00 (0.98 – 1.01) | 5845 | 1.00 (0.99 – 1.00) |
| 1004 | Other signs and symptoms involving emotional state | AHI 3% | 1.4 | 1.00 (0.99 – 1.00) | 3.1E-01 | 18332 | 1.00 (0.99 – 1.01) | 9404 | 1.00 (0.99 – 1.00) |
| 277.7 | Dysmetabolic syndrome X | AHI 3% | 1.3 | 1.00 (1.00 – 1.01) | 3.1E-01 | 18332 | 1.00 (0.99 – 1.01) | 9404 | 1.00 (1.00 – 1.01) |
| 702.1 | Actinic keratosis | AHI 3% | 5.3 | 1.00 (0.99 – 1.00) | 3.2E-01 | 18332 | 1.00 (1.00 – 1.00) | 9404 | 1.00 (0.99 – 1.00) |
| 110.12 | Althete's foot | Per88 | 2.8 | 1.01 (0.99 – 1.02) | 3.2E-01 | 12232 | 1.00 (0.99 – 1.02) | 5845 | 1.01 (0.99 – 1.02) |
| 470 | Septal Deviations/Turbinate Hypertrophy | AHI 3% | 7.3 | 1.00 (1.00 – 1.00) | 3.2E-01 | 18332 | 1.00 (1.00 – 1.00) | 9404 | 1.00 (1.00 – 1.00) |
| 687.2 | Localized superficial swelling, mass, or lump | AHI 3% | 8.4 | 1.00 (1.00 – 1.00) | 3.2E-01 | 18332 | 1.00 (0.99 – 1.00) | 9404 | 1.00 (1.00 – 1.00) |
| 334 | Degenerative disease of the spinal cord | Per88 | 1.3 | 1.01 (0.99 – 1.03) | 3.2E-01 | 12232 | 1.01 (0.99 – 1.02) | 5845 | 1.01 (0.99 – 1.03) |
| 173 | Neoplasm of uncertain behavior of skin | AHI 3% | 9.4 | 1.00 (1.00 – 1.00) | 3.2E-01 | 18332 | 1.00 (1.00 – 1.01) | 9404 | 1.00 (1.00 – 1.00) |
| 706.1 | Acne | Per88 | 5.6 | 1.00 (0.99 – 1.00) | 3.2E-01 | 12232 | 1.00 (0.99 – 1.01) | 5845 | 1.00 (0.99 – 1.00) |
| 259 | Other endocrine disorders | AHI 3% | 1.7 | 1.00 (0.99 – 1.00) | 3.2E-01 | 18332 | 1.00 (0.99 – 1.01) | 9404 | 1.00 (0.99 – 1.00) |
| 788 | Syncope and collapse | AHI 3% | 8.1 | 1.00 (1.00 – 1.00) | 3.2E-01 | 18332 | 1.00 (1.00 – 1.00) | 9404 | 1.00 (1.00 – 1.00) |
| 805 | Fracture of vertebral column without mention of spinal cord injury | AHI 3% | 1.5 | 1.00 (1.00 – 1.01) | 3.2E-01 | 18332 | 1.00 (0.99 – 1.01) | 9404 | 1.00 (1.00 – 1.01) |
| 773 | Pain in limb | Per88 | 21.2 | 1.00 (1.00 – 1.01) | 3.3E-01 | 12232 | 1.00 (1.00 – 1.01) | 5845 | 1.00 (1.00 – 1.01) |
| 550.5 | Ventral hernia | Per88 | 2.2 | 1.01 (0.99 – 1.02) | 3.3E-01 | 12232 | 1.01 (0.99 – 1.02) | 5845 | 1.01 (0.99 – 1.02) |
| 447 | Other disorders of arteries and arterioles | Per88 | 1.3 | 0.99 (0.97 – 1.01) | 3.3E-01 | 12232 | 1.00 (0.98 – 1.02) | 5845 | 0.99 (0.97 – 1.01) |

| PheCode | Disease | Sleep statistic | Prevalence | Risk ratio | P-value | N | Risk ratio (AHI >15) | N (AHI >15) | Risk ratio (adj sleep disorders) |
|---|---|---|---|---|---|---|---|---|---|
| 356 | Hereditary and idiopathic peripheral neuropathy | AHI 3% | 2.5 | 1.00 (0.99 – 1.00) | 3.3E-01 | 18332 | 1.00 (0.99 – 1.01) | 9404 | 1.00 (0.99 – 1.00) |
| 433.31 | Transient cerebral ischemia | AHI 3% | 4.3 | 1.00 (1.00 – 1.01) | 3.3E-01 | 18332 | 1.00 (1.00 – 1.00) | 9404 | 1.00 (1.00 – 1.01) |
| 110.11 | Dermatophytosis of nail | Per88 | 4.4 | 1.01 (0.99 – 1.02) | 3.4E-01 | 12232 | 1.01 (1.00 – 1.02) | 5845 | 1.01 (0.99 – 1.02) |
| 574.1 | Cholelithiasis | Per88 | 4.6 | 0.99 (0.98 – 1.01) | 3.4E-01 | 12232 | 1.00 (0.99 – 1.01) | 5845 | 0.99 (0.98 – 1.01) |
| 411.8 | Other chronic ischemic heart disease, unspecified | Per88 | 1.5 | 1.01 (0.99 – 1.03) | 3.4E-01 | 12232 | 1.01 (0.99 – 1.02) | 5845 | 1.01 (0.99 – 1.03) |
| 465.4 | Acute laryngitis and tracheitis | AHI 3% | 1.5 | 1.00 (1.00 – 1.01) | 3.4E-01 | 18332 | 1.00 (1.00 – 1.01) | 9404 | 1.00 (1.00 – 1.01) |
| 054 | Herpes simplex | AHI 3% | 4.3 | 1.00 (1.00 – 1.00) | 3.5E-01 | 18332 | 1.00 (0.99 – 1.00) | 9404 | 1.00 (1.00 – 1.00) |
| 573.7 | Abnormal results of function study of liver | Per88 | 3.8 | 1.01 (0.99 – 1.02) | 3.5E-01 | 12232 | 1.01 (1.00 – 1.02) | 5845 | 1.01 (0.99 – 1.02) |
| 272.13 | Mixed hyperlipidemia | Per88 | 6.9 | 1.00 (1.00 – 1.01) | 3.5E-01 | 12232 | 1.00 (0.99 – 1.01) | 5845 | 1.00 (1.00 – 1.01) |
| 530.11 | GERD | AHI 3% | 31.1 | 1.00 (1.00 – 1.00) | 3.5E-01 | 18332 | 1.00 (1.00 – 1.00) | 9404 | 1.00 (1.00 – 1.00) |
| 054 | Herpes simplex | Per88 | 4.3 | 1.00 (0.99 – 1.00) | 3.5E-01 | 12232 | 1.00 (0.98 – 1.01) | 5845 | 1.00 (0.99 – 1.00) |
| 197 | Chemotherapy | Per88 | 3.3 | 1.01 (0.99 – 1.02) | 3.5E-01 | 12232 | 1.00 (0.99 – 1.01) | 5845 | 1.01 (0.99 – 1.02) |
| 512.7 | Shortness of breath | AHI 3% | 17.1 | 1.00 (1.00 – 1.00) | 3.5E-01 | 18332 | 1.00 (1.00 – 1.00) | 9404 | 1.00 (1.00 – 1.00) |
| 365 | Glaucoma | AHI 3% | 5.6 | 1.00 (1.00 – 1.00) | 3.5E-01 | 18332 | 1.00 (0.99 – 1.00) | 9404 | 1.00 (1.00 – 1.00) |
| 246 | Other disorders of thyroid | Per88 | 2.6 | 1.01 (0.99 – 1.02) | 3.6E-01 | 12232 | 1.01 (0.99 – 1.02) | 5845 | 1.01 (0.99 – 1.02) |
| 153.2 | Colon cancer | AHI 3% | 1.1 | 1.00 (1.00 – 1.01) | 3.6E-01 | 18332 | 1.01 (1.00 – 1.02) | 9404 | 1.00 (1.00 – 1.01) |
| 706.8 | Other specified diseases of sebaceous glands | AHI 3% | 8.3 | 1.00 (1.00 – 1.00) | 3.6E-01 | 18332 | 1.00 (1.00 – 1.00) | 9404 | 1.00 (1.00 – 1.00) |
| 388 | Other disorders of ear | AHI 3% | 3.1 | 1.00 (0.99 – 1.00) | 3.6E-01 | 18332 | 1.00 (0.99 – 1.00) | 9404 | 1.00 (0.99 – 1.00) |
| 008 | Intestinal infection | AHI 3% | 4.1 | 1.00 (1.00 – 1.01) | 3.6E-01 | 18332 | 1.00 (1.00 – 1.01) | 9404 | 1.00 (1.00 – 1.01) |
| 197 | Chemotherapy | AHI 3% | 3.3 | 1.00 (1.00 – 1.01) | 3.6E-01 | 18332 | 1.00 (1.00 – 1.01) | 9404 | 1.00 (1.00 – 1.01) |
| 306.9 | Tension headache | AHI 3% | 3.5 | 1.00 (0.99 – 1.00) | 3.7E-01 | 18332 | 1.00 (0.99 – 1.00) | 9404 | 1.00 (0.99 – 1.00) |
| 471 | Nasal polyps | Per88 | 1.9 | 0.99 (0.98 – 1.01) | 3.7E-01 | 12232 | 0.99 (0.97 – 1.01) | 5845 | 0.99 (0.98 – 1.01) |
| 537 | Other disorders of stomach and duodenum | AHI 3% | 7.8 | 1.00 (1.00 – 1.00) | 3.7E-01 | 18332 | 1.00 (1.00 – 1.00) | 9404 | 1.00 (1.00 – 1.00) |
| 345.1 | Epilepsy | Per88 | 1.7 | 0.99 (0.98 – 1.01) | 3.7E-01 | 12232 | 0.99 (0.98 – 1.01) | 5845 | 0.99 (0.98 – 1.01) |
| 599.8 | Other symptoms involving urinary system | Per88 | 3.2 | 1.01 (0.99 – 1.02) | 3.7E-01 | 12232 | 1.00 (0.99 – 1.02) | 5845 | 1.01 (0.99 – 1.02) |
| 561 | Symptoms involving digestive system | Per88 | 3.6 | 1.00 (0.99 – 1.01) | 3.8E-01 | 12232 | 1.00 (0.99 – 1.01) | 5845 | 1.00 (0.99 – 1.01) |
| 287.1 | Spontaneous ecchymoses | AHI 3% | 1.0 | 1.00 (0.99 – 1.00) | 3.8E-01 | 18332 | 1.00 (0.99 – 1.01) | 9404 | 1.00 (0.99 – 1.00) |
| 722.9 | Other and unspecified disc disorder | Per88 | 2.7 | 1.01 (0.99 – 1.02) | 3.8E-01 | 12232 | 1.01 (0.99 – 1.02) | 5845 | 1.01 (0.99 – 1.02) |
| 379.2 | Disorders of vitreous body | AHI 3% | 3.3 | 1.00 (0.99 – 1.00) | 3.8E-01 | 18332 | 1.00 (0.99 – 1.00) | 9404 | 1.00 (0.99 – 1.00) |
| 389.1 | Sensorineural hearing loss | Per88 | 6.0 | 1.00 (0.99 – 1.01) | 3.8E-01 | 12232 | 1.00 (0.99 – 1.01) | 5845 | 1.00 (0.99 – 1.01) |
| 695.8 | Other specified erythematous conditions | Per88 | 1.9 | 0.99 (0.97 – 1.01) | 3.9E-01 | 12232 | 0.99 (0.97 – 1.00) | 5845 | 0.99 (0.97 – 1.01) |
| 251.1 | Hypoglycemia | AHI 3% | 1.6 | 1.00 (1.00 – 1.01) | 3.9E-01 | 18332 | 1.01 (1.00 – 1.01) | 9404 | 1.00 (1.00 – 1.01) |
| 293 | Symptoms involving head and neck | AHI 3% | 1.1 | 1.00 (0.99 – 1.00) | 3.9E-01 | 18332 | 1.00 (0.99 – 1.01) | 9404 | 1.00 (0.99 – 1.00) |

| PheCode | Disease | Sleep statistic | Prevalence | Risk ratio | P-value | N | Risk ratio (AHI >15) | N (AHI >15) | Risk ratio (adj sleep disorders) |
|---|---|---|---|---|---|---|---|---|---|
| 705.8 | Hyperhidrosis | AHI 3% | 3.5 | 1.00 (0.99 – 1.00) | 3.9E-01 | 18332 | 1.00 (0.99 – 1.00) | 9404 | 1.00 (0.99 – 1.00) |
| 591 | Urinary tract infection | Per88 | 8.5 | 1.00 (1.00 – 1.01) | 4.0E-01 | 12232 | 1.00 (0.99 – 1.01) | 5845 | 1.00 (1.00 – 1.01) |
| 374.3 | Ptosis of eyelid | AHI 3% | 1.5 | 1.00 (0.99 – 1.00) | 4.0E-01 | 18332 | 1.00 (0.99 – 1.01) | 9404 | 1.00 (0.99 – 1.00) |
| 735.1 | Flat foot | AHI 3% | 2.0 | 1.00 (0.99 – 1.00) | 4.0E-01 | 18332 | 1.00 (0.99 – 1.00) | 9404 | 1.00 (0.99 – 1.00) |
| 386.2 | Peripheral or central vertigo | Per88 | 2.8 | 0.99 (0.98 – 1.01) | 4.0E-01 | 12232 | 1.00 (0.99 – 1.01) | 5845 | 0.99 (0.98 – 1.01) |
| 395.2 | Nonrheumatic aortic valve disorders | Per88 | 2.9 | 1.01 (0.99 – 1.02) | 4.0E-01 | 12232 | 1.00 (0.99 – 1.01) | 5845 | 1.01 (0.99 – 1.02) |
| 286.81 | Primary hypercoagulable state | Per88 | 1.1 | 0.99 (0.97 – 1.01) | 4.1E-01 | 12232 | 0.99 (0.97 – 1.01) | 5845 | 0.99 (0.97 – 1.01) |
| 379.9 | Pain, swelling or discharge of eye | Per88 | 4.4 | 1.00 (0.99 – 1.01) | 4.1E-01 | 12232 | 1.00 (0.99 – 1.01) | 5845 | 1.00 (0.99 – 1.01) |
| 348.8 | Encephalopathy, not elsewhere classified | Per88 | 1.6 | 1.01 (0.99 – 1.02) | 4.1E-01 | 12232 | 0.99 (0.97 – 1.01) | 5845 | 1.01 (0.99 – 1.02) |
| 722.1 | Displacement of intervertebral disc | Per88 | 4.5 | 1.00 (0.99 – 1.01) | 4.1E-01 | 12232 | 1.00 (0.99 – 1.01) | 5845 | 1.00 (0.99 – 1.01) |
| 356 | Hereditary and idiopathic peripheral neuropathy | Per88 | 2.5 | 1.01 (0.99 – 1.02) | 4.2E-01 | 12232 | 1.00 (0.99 – 1.02) | 5845 | 1.01 (0.99 – 1.02) |
| 565 | Anal and rectal conditions | AHI 3% | 3.8 | 1.00 (1.00 – 1.01) | 4.2E-01 | 18332 | 1.00 (1.00 – 1.01) | 9404 | 1.00 (1.00 – 1.01) |
| 250.41 | Impaired fasting glucose | AHI 3% | 3.7 | 1.00 (1.00 – 1.01) | 4.2E-01 | 18332 | 1.00 (0.99 – 1.00) | 9404 | 1.00 (1.00 – 1.01) |
| 443.9 | Peripheral vascular disease, unspecified | AHI 3% | 1.6 | 1.00 (1.00 – 1.01) | 4.2E-01 | 18332 | 1.00 (0.99 – 1.01) | 9404 | 1.00 (1.00 – 1.01) |
| 475 | Chronic sinusitis | AHI 3% | 6.8 | 1.00 (1.00 – 1.00) | 4.2E-01 | 18332 | 1.00 (0.99 – 1.00) | 9404 | 1.00 (1.00 – 1.00) |
| 562.2 | Diverticulitis | AHI 3% | 3.4 | 1.00 (1.00 – 1.01) | 4.2E-01 | 18332 | 1.00 (0.99 – 1.00) | 9404 | 1.00 (1.00 – 1.01) |
| 727 | Other disorders of synovium, tendon, and bursa | AHI 3% | 2.6 | 1.00 (0.99 – 1.00) | 4.2E-01 | 18332 | 0.99 (0.99 – 1.00) | 9404 | 1.00 (0.99 – 1.00) |
| 261.4 | Vitamin D deficiency | AHI 3% | 11.6 | 1.00 (1.00 – 1.00) | 4.2E-01 | 18332 | 1.00 (1.00 – 1.00) | 9404 | 1.00 (1.00 – 1.00) |
| 277.7 | Dysmetabolic syndrome X | Per88 | 1.3 | 0.99 (0.97 – 1.01) | 4.3E-01 | 12232 | 0.99 (0.97 – 1.02) | 5845 | 0.99 (0.97 – 1.01) |
| 841 | Sprains and strains of back and neck | AHI 3% | 2.3 | 1.00 (1.00 – 1.01) | 4.3E-01 | 18332 | 1.00 (0.99 – 1.01) | 9404 | 1.00 (1.00 – 1.01) |
| 369.5 | Conjunctivitis, infectious | Per88 | 5.4 | 1.00 (0.99 – 1.01) | 4.3E-01 | 12232 | 1.00 (0.99 – 1.01) | 5845 | 1.00 (0.99 – 1.00) |
| 770 | Myalgia and myositis unspecified | Per88 | 6.5 | 1.00 (0.99 – 1.00) | 4.3E-01 | 12232 | 1.00 (0.99 – 1.01) | 5845 | 1.00 (0.99 – 1.00) |
| 599.4 | Urinary incontinence | Per88 | 6.7 | 1.00 (1.00 – 1.01) | 4.3E-01 | 12232 | 1.00 (0.99 – 1.01) | 5845 | 1.00 (1.00 – 1.01) |
| 706.8 | Other specified diseases of sebaceous glands | Per88 | 8.3 | 1.00 (0.99 – 1.00) | 4.3E-01 | 12232 | 1.00 (0.99 – 1.01) | 5845 | 1.00 (0.99 – 1.00) |
| 687.1 | Rash and other nonspecific skin eruption | Per88 | 11.4 | 1.00 (1.00 – 1.01) | 4.3E-01 | 12232 | 1.01 (1.00 – 1.01) | 5845 | 1.00 (1.00 – 1.01) |
| 216 | Benign neoplasm of skin | AHI 3% | 14.0 | 1.00 (1.00 – 1.00) | 4.3E-01 | 18332 | 1.00 (1.00 – 1.00) | 9404 | 1.00 (1.00 – 1.00) |
| 550.2 | Diaphragmatic hernia | Per88 | 5.5 | 1.00 (0.99 – 1.01) | 4.4E-01 | 12232 | 1.00 (0.99 – 1.01) | 5845 | 1.00 (0.99 – 1.01) |
| 078 | Viral warts & HPV | AHI 3% | 4.9 | 1.00 (1.00 – 1.00) | 4.4E-01 | 18332 | 1.00 (1.00 – 1.01) | 9404 | 1.00 (1.00 – 1.00) |
| 979 | Adverse drug events and drug allergies | AHI 3% | 5.7 | 1.00 (1.00 – 1.00) | 4.4E-01 | 18332 | 1.00 (1.00 – 1.00) | 9404 | 1.00 (1.00 – 1.00) |
| 389 | Hearing loss | Per88 | 8.9 | 1.00 (1.00 – 1.01) | 4.4E-01 | 12232 | 1.00 (0.99 – 1.01) | 5845 | 1.00 (1.00 – 1.01) |
| 418 | Nonspecific chest pain | Per88 | 20.0 | 1.00 (1.00 – 1.01) | 4.4E-01 | 12232 | 1.00 (0.99 – 1.00) | 5845 | 1.00 (1.00 – 1.01) |
| 765 | Cervical radiculitis | AHI 3% | 4.4 | 1.00 (0.99 – 1.00) | 4.4E-01 | 18332 | 1.00 (0.99 – 1.00) | 9404 | 1.00 (1.00 – 1.00) |
| 279.2 | Autoimmune disease NEC | AHI 3% | 1.2 | 1.00 (0.99 – 1.00) | 4.4E-01 | 18332 | 1.00 (0.99 – 1.01) | 9404 | 1.00 (0.99 – 1.00) |
| 411.4 | Coronary atherosclerosis | Per88 | 8.1 | 1.00 (0.99 – 1.01) | 4.5E-01 | 12232 | 1.00 (0.99 – 1.01) | 5845 | 1.00 (1.00 – 1.01) |

| PheCode | Disease | Sleep statistic | Prevalence | Risk ratio | P-value | N | Risk ratio (AHI >15) | N (AHI >15) | Risk ratio (adj sleep disorders) |
|---|---|---|---|---|---|---|---|---|---|
| 481 | Influenza | Per88 | 7.0 | 1.00 (1.00 – 1.01) | 4.5E-01 | 12232 | 1.00 (0.99 – 1.01) | 5845 | 1.00 (1.00 – 1.01) |
| 705.8 | Hyperhidrosis | Per88 | 3.5 | 1.00 (0.99 – 1.01) | 4.5E-01 | 12232 | 1.00 (0.99 – 1.02) | 5845 | 1.00 (0.99 – 1.01) |
| 531.2 | Gastric ulcer | AHI 3% | 1.7 | 1.00 (0.99 – 1.00) | 4.5E-01 | 18332 | 1.00 (0.99 – 1.00) | 9404 | 1.00 (0.99 – 1.00) |
| 367.1 | Myopia | AHI 3% | 2.5 | 1.00 (0.99 – 1.00) | 4.5E-01 | 18332 | 1.00 (0.99 – 1.00) | 9404 | 1.00 (0.99 – 1.00) |
| 477 | Epistaxis or throat hemorrhage | AHI 3% | 2.5 | 1.00 (1.00 – 1.01) | 4.5E-01 | 18332 | 1.00 (0.99 – 1.01) | 9404 | 1.00 (1.00 – 1.01) |
| 715 | Other inflammatory spondylopathies | Per88 | 1.0 | 0.99 (0.97 – 1.01) | 4.6E-01 | 12232 | 1.00 (0.98 – 1.02) | 5845 | 0.99 (0.97 – 1.01) |
| 443.1 | Raynaud's syndrome | Per88 | 1.8 | 1.00 (0.98 – 1.01) | 4.6E-01 | 12232 | 0.99 (0.97 – 1.01) | 5845 | 1.00 (0.98 – 1.01) |
| 512.2 | Painful respiration | AHI 3% | 9.5 | 1.00 (1.00 – 1.00) | 4.6E-01 | 18332 | 1.00 (1.00 – 1.00) | 9404 | 1.00 (1.00 – 1.00) |
| 720 | Spinal stenosis | AHI 3% | 4.6 | 1.00 (0.99 – 1.00) | 4.6E-01 | 18332 | 1.00 (1.00 – 1.00) | 9404 | 1.00 (0.99 – 1.00) |
| 053 | Herpes zoster | Per88 | 4.7 | 1.00 (0.99 – 1.01) | 4.6E-01 | 12232 | 1.01 (1.00 – 1.02) | 5845 | 1.00 (0.99 – 1.01) |
| 279.2 | Autoimmune disease NEC | Per88 | 1.2 | 1.01 (0.99 – 1.02) | 4.7E-01 | 12232 | 1.01 (0.99 – 1.03) | 5845 | 1.01 (0.99 – 1.02) |
| 366 | Cataract | AHI 3% | 5.3 | 1.00 (1.00 – 1.00) | 4.7E-01 | 18332 | 1.00 (0.99 – 1.00) | 9404 | 1.00 (1.00 – 1.00) |
| 562.2 | Diverticulitis | Per88 | 3.4 | 1.00 (0.99 – 1.02) | 4.7E-01 | 12232 | 0.99 (0.98 – 1.01) | 5845 | 1.00 (0.99 – 1.02) |
| 351 | Other peripheral nerve disorders | Per88 | 7.5 | 1.00 (1.00 – 1.01) | 4.7E-01 | 12232 | 1.00 (1.00 – 1.01) | 5845 | 1.00 (1.00 – 1.01) |
| 277.51 | Lipoprotein disorders | AHI 3% | 1.8 | 1.00 (1.00 – 1.01) | 4.8E-01 | 18332 | 1.01 (1.00 – 1.01) | 9404 | 1.00 (1.00 – 1.01) |
| 738.4 | Acquired spondylolisthesis | AHI 3% | 3.0 | 1.00 (0.99 – 1.00) | 4.8E-01 | 18332 | 1.00 (0.99 – 1.00) | 9404 | 1.00 (0.99 – 1.00) |
| 367.4 | Presbyopia | Per88 | 2.5 | 1.00 (0.99 – 1.02) | 4.8E-01 | 12232 | 1.01 (1.00 – 1.02) | 5845 | 1.00 (0.99 – 1.02) |
| 781 | Symptoms involving nervous and musculoskeletal systems | AHI 3% | 5.4 | 1.00 (1.00 – 1.00) | 4.8E-01 | 18332 | 1.00 (1.00 – 1.01) | 9404 | 1.00 (1.00 – 1.00) |
| 443.1 | Raynaud's syndrome | AHI 3% | 1.8 | 1.00 (0.99 – 1.00) | 4.8E-01 | 18332 | 1.00 (0.99 – 1.01) | 9404 | 1.00 (0.99 – 1.00) |
| 687.2 | Localized superficial swelling, mass, or lump | Per88 | 8.4 | 1.00 (1.00 – 1.01) | 4.8E-01 | 12232 | 1.00 (0.99 – 1.01) | 5845 | 1.00 (1.00 – 1.01) |
| 429.3 | Symptoms involving cardiovascular system | AHI 3% | 6.9 | 1.00 (1.00 – 1.00) | 4.9E-01 | 18332 | 1.00 (1.00 – 1.01) | 9404 | 1.00 (1.00 – 1.00) |
| 745 | Pain in joint | Per88 | 33.1 | 1.00 (1.00 – 1.00) | 4.9E-01 | 12232 | 1.00 (1.00 – 1.00) | 5845 | 1.00 (1.00 – 1.00) |
| 483 | Acute bronchitis and bronchiolitis | AHI 3% | 5.5 | 1.00 (1.00 – 1.00) | 4.9E-01 | 18332 | 1.00 (1.00 – 1.00) | 9404 | 1.00 (1.00 – 1.00) |
| 703.1 | Ingrowing nail | AHI 3% | 1.9 | 1.00 (0.99 – 1.00) | 4.9E-01 | 18332 | 1.01 (1.00 – 1.01) | 9404 | 1.00 (0.99 – 1.00) |
| 214 | Lipoma | Per88 | 2.6 | 1.00 (0.99 – 1.02) | 4.9E-01 | 12232 | 1.00 (0.99 – 1.01) | 5845 | 1.01 (0.99 – 1.02) |
| 738.4 | Acquired spondylolisthesis | Per88 | 3.0 | 1.00 (0.99 – 1.02) | 4.9E-01 | 12232 | 1.00 (0.99 – 1.02) | 5845 | 1.00 (0.99 – 1.02) |
| 727.1 | Synovitis and tenosynovitis | AHI 3% | 5.9 | 1.00 (1.00 – 1.00) | 4.9E-01 | 18332 | 1.00 (0.99 – 1.00) | 9404 | 1.00 (1.00 – 1.00) |
| 737.3 | Kyphoscoliosis and scoliosis | AHI 3% | 1.8 | 1.00 (1.00 – 1.01) | 4.9E-01 | 18332 | 1.00 (0.99 – 1.01) | 9404 | 1.00 (1.00 – 1.01) |
| 008.5 | Bacterial enteritis | AHI 3% | 3.8 | 1.00 (0.99 – 1.00) | 4.9E-01 | 18332 | 1.00 (0.99 – 1.00) | 9404 | 1.00 (0.99 – 1.00) |
| 571.5 | Other chronic nonalcoholic liver disease | Per88 | 5.1 | 1.00 (0.99 – 1.01) | 4.9E-01 | 12232 | 1.00 (0.99 – 1.01) | 5845 | 1.00 (0.99 – 1.01) |
| 262 | Mineral deficiency NEC | AHI 3% | 3.5 | 1.00 (0.99 – 1.00) | 4.9E-01 | 18332 | 1.00 (0.99 – 1.01) | 9404 | 1.00 (1.00 – 1.00) |
| 389.2 | Conductive hearing loss | Per88 | 1.0 | 0.99 (0.97 – 1.01) | 4.9E-01 | 12232 | 1.00 (0.98 – 1.02) | 5845 | 0.99 (0.97 – 1.01) |
| 380.4 | Impacted cerumen | AHI 3% | 3.7 | 1.00 (1.00 – 1.01) | 4.9E-01 | 18332 | 1.00 (1.00 – 1.01) | 9404 | 1.00 (1.00 – 1.01) |
| 535.2 | Atrophic gastritis | Per88 | 2.7 | 1.00 (0.98 – 1.01) | 5.0E-01 | 12232 | 1.00 (0.98 – 1.01) | 5845 | 1.00 (0.98 – 1.01) |
| 241.2 | Nontoxic multinodular goiter | AHI 3% | 3.0 | 1.00 (1.00 – 1.01) | 5.0E-01 | 18332 | 1.00 (0.99 – 1.01) | 9404 | 1.00 (1.00 – 1.01) |

| PheCode | Disease | Sleep statistic | Prevalence | Risk ratio | P-value | N | Risk ratio (AHI >15) | N (AHI >15) | Risk ratio (adj sleep disorders) |
|---|---|---|---|---|---|---|---|---|---|
| 594.3 | Calculus of ureter | AHI 3% | 1.5 | 1.00 (1.00 – 1.01) | 5.0E-01 | 18332 | 1.00 (1.00 – 1.01) | 9404 | 1.00 (1.00 – 1.01) |
| 760 | Back pain | Per88 | 23.6 | 1.00 (1.00 – 1.01) | 5.0E-01 | 12232 | 1.00 (1.00 – 1.01) | 5845 | 1.00 (1.00 – 1.01) |
| 153.2 | Colon cancer | Per88 | 1.1 | 1.01 (0.99 – 1.03) | 5.1E-01 | 12232 | 1.01 (0.99 – 1.03) | 5845 | 1.01 (0.99 – 1.03) |
| 367.8 | Hypermetropia | AHI 3% | 1.8 | 1.00 (0.99 – 1.00) | 5.1E-01 | 18332 | 1.00 (0.99 – 1.00) | 9404 | 1.00 (0.99 – 1.00) |
| 447.7 | Aortic ectasia | Per88 | 1.3 | 1.01 (0.99 – 1.03) | 5.1E-01 | 12232 | 1.00 (0.99 – 1.02) | 5845 | 1.01 (0.99 – 1.03) |
| 735.1 | Flat foot | Per88 | 2.0 | 1.01 (0.99 – 1.02) | 5.1E-01 | 12232 | 1.00 (0.98 – 1.02) | 5845 | 1.01 (0.99 – 1.02) |
| 217.1 | Nevus, non-neoplastic | AHI 3% | 5.2 | 1.00 (1.00 – 1.00) | 5.2E-01 | 18332 | 1.00 (1.00 – 1.01) | 9404 | 1.00 (1.00 – 1.00) |
| 333.1 | Essential tremor | Per88 | 1.1 | 0.99 (0.97 – 1.02) | 5.2E-01 | 12232 | 1.00 (0.98 – 1.02) | 5845 | 0.99 (0.97 – 1.02) |
| 287.1 | Spontaneous ecchymoses | Per88 | 1.0 | 1.01 (0.99 – 1.02) | 5.2E-01 | 12232 | 1.00 (0.98 – 1.03) | 5845 | 1.01 (0.99 – 1.02) |
| 689 | Disorder of skin and subcutaneous tissue NOS | AHI 3% | 13.7 | 1.00 (1.00 – 1.00) | 5.2E-01 | 18332 | 1.00 (1.00 – 1.00) | 9404 | 1.00 (1.00 – 1.00) |
| 591 | Urinary tract infection | AHI 3% | 8.5 | 1.00 (1.00 – 1.00) | 5.2E-01 | 18332 | 1.00 (1.00 – 1.00) | 9404 | 1.00 (1.00 – 1.00) |
| 530.14 | Reflux esophagitis | AHI 3% | 3.1 | 1.00 (1.00 – 1.01) | 5.2E-01 | 18332 | 1.00 (1.00 – 1.01) | 9404 | 1.00 (1.00 – 1.01) |
| 245.21 | Chronic lymphocytic thyroiditis | Per88 | 1.6 | 1.00 (0.98 – 1.01) | 5.2E-01 | 12232 | 1.00 (0.98 – 1.02) | 5845 | 1.00 (0.98 – 1.01) |
| 941 | Adverse reaction to serum or vaccine | AHI 3% | 2.7 | 1.00 (0.99 – 1.00) | 5.2E-01 | 18332 | 1.00 (0.99 – 1.01) | 9404 | 1.00 (0.99 – 1.00) |
| 579.8 | Nonspecific abnormal findings in stool contents | AHI 3% | 4.7 | 1.00 (1.00 – 1.00) | 5.3E-01 | 18332 | 1.00 (1.00 – 1.01) | 9404 | 1.00 (1.00 – 1.00) |
| 440 | Atherosclerosis | AHI 3% | 1.2 | 1.00 (0.99 – 1.01) | 5.3E-01 | 18332 | 1.00 (1.00 – 1.01) | 9404 | 1.00 (0.99 – 1.01) |
| 350.6 | Disturbances of sensation of smell and taste | Per88 | 1.8 | 0.99 (0.98 – 1.01) | 5.3E-01 | 12232 | 1.00 (0.98 – 1.01) | 5845 | 1.00 (0.98 – 1.01) |
| 286.9 | Abnormal coagulation profile | AHI 3% | 2.8 | 1.00 (0.99 – 1.00) | 5.3E-01 | 18332 | 1.00 (0.99 – 1.01) | 9404 | 1.00 (0.99 – 1.00) |
| 110.2 | Dermatomycoses | Per88 | 1.8 | 1.00 (0.99 – 1.02) | 5.3E-01 | 12232 | 1.00 (0.99 – 1.02) | 5845 | 1.00 (0.99 – 1.02) |
| 433.8 | Late effects of cerebrovascular disease | AHI 3% | 1.4 | 1.00 (1.00 – 1.01) | 5.3E-01 | 18332 | 1.00 (0.99 – 1.01) | 9404 | 1.00 (1.00 – 1.01) |
| 395.1 | Nonrheumatic mitral valve disorders | AHI 3% | 4.0 | 1.00 (1.00 – 1.00) | 5.3E-01 | 18332 | 1.00 (0.99 – 1.00) | 9404 | 1.00 (0.99 – 1.00) |
| 350.6 | Disturbances of sensation of smell and taste | AHI 3% | 1.8 | 1.00 (0.99 – 1.00) | 5.3E-01 | 18332 | 1.00 (0.99 – 1.00) | 9404 | 1.00 (0.99 – 1.00) |
| 333.1 | Essential tremor | AHI 3% | 1.1 | 1.00 (0.99 – 1.01) | 5.3E-01 | 18332 | 1.00 (0.99 – 1.01) | 9404 | 1.00 (0.99 – 1.01) |
| 380.1 | Otitis externa | Per88 | 2.2 | 1.00 (0.99 – 1.02) | 5.3E-01 | 12232 | 1.01 (1.00 – 1.03) | 5845 | 1.00 (0.99 – 1.02) |
| 475 | Chronic sinusitis | Per88 | 6.8 | 1.00 (0.99 – 1.01) | 5.3E-01 | 12232 | 1.00 (0.99 – 1.00) | 5845 | 1.00 (0.99 – 1.01) |
| 473.4 | Voice disturbance | AHI 3% | 4.7 | 1.00 (1.00 – 1.00) | 5.3E-01 | 18332 | 1.00 (1.00 – 1.01) | 9404 | 1.00 (1.00 – 1.00) |
| 550.5 | Ventral hernia | AHI 3% | 2.2 | 1.00 (0.99 – 1.00) | 5.4E-01 | 18332 | 1.00 (0.99 – 1.00) | 9404 | 1.00 (0.99 – 1.00) |
| 348.8 | Encephalopathy, not elsewhere classified | AHI 3% | 1.6 | 1.00 (1.00 – 1.01) | 5.4E-01 | 18332 | 1.00 (1.00 – 1.01) | 9404 | 1.00 (1.00 – 1.01) |
| 593.1 | Gross hematuria | Per88 | 2.7 | 1.00 (0.99 – 1.02) | 5.4E-01 | 12232 | 1.00 (0.99 – 1.02) | 5845 | 1.00 (0.99 – 1.02) |
| 427.21 | Atrial fibrillation | AHI 3% | 9.2 | 1.00 (1.00 – 1.00) | 5.4E-01 | 18332 | 1.00 (1.00 – 1.00) | 9404 | 1.00 (1.00 – 1.00) |
| 577.3 | Cyst and pseudocyst of pancreas | AHI 3% | 1.2 | 1.00 (1.00 – 1.01) | 5.4E-01 | 18332 | 1.01 (1.00 – 1.01) | 9404 | 1.00 (1.00 – 1.01) |
| 531.4 | Peptic ulcer, site unspecified | AHI 3% | 1.3 | 1.00 (0.99 – 1.01) | 5.4E-01 | 18332 | 1.00 (0.99 – 1.01) | 9404 | 1.00 (0.99 – 1.01) |
| 716.9 | Arthropathy NOS | Per88 | 2.2 | 1.00 (0.99 – 1.02) | 5.5E-01 | 12232 | 1.00 (0.99 – 1.02) | 5845 | 1.00 (0.99 – 1.02) |
| 455 | Hemorrhoids | Per88 | 11.2 | 1.00 (0.99 – 1.00) | 5.5E-01 | 12232 | 1.00 (1.00 – 1.01) | 5845 | 1.00 (0.99 – 1.00) |

| PheCode | Disease | Sleep statistic | Prevalence | Risk ratio | P-value | N | Risk ratio (AHI >15) | N (AHI >15) | Risk ratio (adj sleep disorders) |
|---|---|---|---|---|---|---|---|---|---|
| 350.3 | Lack of coordination | Per88 | 1.2 | 0.99 (0.98 – 1.01) | 5.6E-01 | 12232 | 0.99 (0.97 – 1.00) | 5845 | 0.99 (0.98 – 1.01) |
| 696.41 | Psoriasis vulgaris | Per88 | 3.3 | 1.00 (0.98 – 1.01) | 5.6E-01 | 12232 | 1.00 (0.98 – 1.01) | 5845 | 1.00 (0.98 – 1.01) |
| 426.31 | Right bundle branch block | AHI 3% | 2.2 | 1.00 (1.00 – 1.01) | 5.6E-01 | 18332 | 1.00 (0.99 – 1.01) | 9404 | 1.00 (1.00 – 1.01) |
| 742.9 | Other derangement of joint | AHI 3% | 4.8 | 1.00 (1.00 – 1.00) | 5.6E-01 | 18332 | 1.00 (0.99 – 1.00) | 9404 | 1.00 (1.00 – 1.00) |
| 510 | Other diseases of lung | AHI 3% | 3.7 | 1.00 (0.99 – 1.00) | 5.7E-01 | 18332 | 1.00 (0.99 – 1.00) | 9404 | 1.00 (0.99 – 1.00) |
| 574.3 | Cholecystitis without cholelithiasis | AHI 3% | 3.1 | 1.00 (0.99 – 1.00) | 5.7E-01 | 18332 | 1.00 (0.99 – 1.01) | 9404 | 1.00 (0.99 – 1.00) |
| 367.2 | Astigmatism | Per88 | 2.8 | 1.00 (0.99 – 1.01) | 5.7E-01 | 12232 | 1.00 (0.99 – 1.02) | 5845 | 1.00 (0.99 – 1.01) |
| 313.1 | Attention deficit hyperactivity disorder | Per88 | 5.7 | 1.00 (0.99 – 1.00) | 5.7E-01 | 12232 | 0.99 (0.98 – 1.00) | 5845 | 1.00 (0.99 – 1.00) |
| 596 | Other disorders of bladder | Per88 | 2.1 | 1.00 (0.99 – 1.02) | 5.8E-01 | 12232 | 1.00 (0.98 – 1.02) | 5845 | 1.00 (0.99 – 1.02) |
| 592.11 | Acute cystitis | AHI 3% | 2.2 | 1.00 (0.99 – 1.00) | 5.8E-01 | 18332 | 1.00 (0.99 – 1.00) | 9404 | 1.00 (0.99 – 1.00) |
| 471 | Nasal polyps | AHI 3% | 1.9 | 1.00 (0.99 – 1.00) | 5.9E-01 | 18332 | 1.00 (0.99 – 1.00) | 9404 | 1.00 (0.99 – 1.00) |
| 722.9 | Other and unspecified disc disorder | AHI 3% | 2.7 | 1.00 (0.99 – 1.00) | 5.9E-01 | 18332 | 1.00 (0.99 – 1.01) | 9404 | 1.00 (0.99 – 1.00) |
| 695.3 | Rosacea | AHI 3% | 3.5 | 1.00 (0.99 – 1.00) | 5.9E-01 | 18332 | 1.00 (0.99 – 1.00) | 9404 | 1.00 (0.99 – 1.00) |
| 742.9 | Other derangement of joint | Per88 | 4.8 | 1.00 (0.99 – 1.01) | 5.9E-01 | 12232 | 1.00 (0.99 – 1.01) | 5845 | 1.00 (0.99 – 1.01) |
| 599.9 | Other abnormality of urination | Per88 | 4.6 | 1.00 (0.99 – 1.01) | 5.9E-01 | 12232 | 0.99 (0.98 – 1.00) | 5845 | 1.00 (0.99 – 1.01) |
| 244.2 | Acquired hypothyroidism | Per88 | 2.8 | 1.00 (0.99 – 1.02) | 6.0E-01 | 12232 | 1.00 (0.99 – 1.02) | 5845 | 1.00 (0.99 – 1.02) |
| 741.2 | Stiffness of joint | Per88 | 2.5 | 1.00 (0.99 – 1.02) | 6.0E-01 | 12232 | 1.00 (0.99 – 1.02) | 5845 | 1.00 (0.99 – 1.02) |
| 695.3 | Rosacea | Per88 | 3.5 | 1.00 (0.98 – 1.01) | 6.0E-01 | 12232 | 1.01 (0.99 – 1.02) | 5845 | 1.00 (0.98 – 1.01) |
| 766 | Neuralgia, neuritis, and radiculitis NOS | AHI 3% | 3.7 | 1.00 (0.99 – 1.00) | 6.0E-01 | 18332 | 1.00 (0.99 – 1.00) | 9404 | 1.00 (1.00 – 1.00) |
| 557 | Intestinal malabsorption (non-celiac) | Per88 | 1.9 | 1.00 (0.99 – 1.02) | 6.0E-01 | 12232 | 1.00 (0.98 – 1.01) | 5845 | 1.00 (0.99 – 1.02) |
| 327.4 | Insomnia | Per88 | 16.8 | 1.00 (1.00 – 1.00) | 6.0E-01 | 12232 | 1.00 (1.00 – 1.01) | 5845 | 1.00 (1.00 – 1.00) |
| 789 | Nausea and vomiting | Per88 | 13.3 | 1.00 (1.00 – 1.01) | 6.0E-01 | 12232 | 1.00 (1.00 – 1.01) | 5845 | 1.00 (1.00 – 1.01) |
| 262 | Mineral deficiency NEC | Per88 | 3.5 | 1.00 (0.99 – 1.01) | 6.0E-01 | 12232 | 1.01 (0.99 – 1.02) | 5845 | 1.00 (0.99 – 1.01) |
| 946 | Anaphylactic shock NOS | Per88 | 1.9 | 1.00 (0.98 – 1.01) | 6.0E-01 | 12232 | 0.99 (0.97 – 1.01) | 5845 | 1.00 (0.98 – 1.01) |
| 366.2 | Senile cataract | Per88 | 6.6 | 1.00 (0.99 – 1.01) | 6.0E-01 | 12232 | 1.00 (0.99 – 1.01) | 5845 | 1.00 (0.99 – 1.01) |
| 288.1 | Decreased white blood cell count | Per88 | 1.2 | 1.00 (0.99 – 1.02) | 6.0E-01 | 12232 | 1.01 (0.99 – 1.03) | 5845 | 1.00 (0.99 – 1.02) |
| 411.3 | Angina pectoris | AHI 3% | 7.0 | 1.00 (1.00 – 1.00) | 6.1E-01 | 18332 | 1.00 (0.99 – 1.00) | 9404 | 1.00 (1.00 – 1.00) |
| 599.2 | Retention of urine | AHI 3% | 4.0 | 1.00 (0.99 – 1.00) | 6.1E-01 | 18332 | 1.00 (1.00 – 1.01) | 9404 | 1.00 (0.99 – 1.00) |
| 426 | Cardiac conduction disorders | Per88 | 1.2 | 1.01 (0.98 – 1.03) | 6.2E-01 | 12232 | 1.01 (0.99 – 1.03) | 5845 | 1.01 (0.98 – 1.03) |
| 599.5 | Frequency of urination and polyuria | Per88 | 9.8 | 1.00 (0.99 – 1.01) | 6.2E-01 | 12232 | 1.00 (0.99 – 1.00) | 5845 | 1.00 (0.99 – 1.01) |
| 526.41 | Temporomandibular joint disorder, unspecified | Per88 | 1.6 | 1.00 (0.98 – 1.01) | 6.3E-01 | 12232 | 0.99 (0.98 – 1.01) | 5845 | 1.00 (0.98 – 1.01) |
| 263 | Other nutritional deficiency | Per88 | 1.3 | 1.00 (0.99 – 1.02) | 6.3E-01 | 12232 | 1.01 (0.99 – 1.03) | 5845 | 1.00 (0.99 – 1.02) |
| 1004 | Other signs and symptoms involving emotional state | Per88 | 1.4 | 1.00 (0.98 – 1.01) | 6.3E-01 | 12232 | 1.00 (0.98 – 1.02) | 5845 | 1.00 (0.98 – 1.01) |
| 473.4 | Voice disturbance | Per88 | 4.7 | 1.00 (0.99 – 1.01) | 6.3E-01 | 12232 | 1.00 (0.99 – 1.01) | 5845 | 1.00 (0.99 – 1.01) |
| 433.31 | Transient cerebral ischemia | Per88 | 4.3 | 1.00 (0.99 – 1.01) | 6.3E-01 | 12232 | 1.00 (0.99 – 1.01) | 5845 | 1.00 (0.99 – 1.01) |
| 110.12 | Althete's foot | AHI 3% | 2.8 | 1.00 (1.00 – 1.01) | 6.4E-01 | 18332 | 1.00 (0.99 – 1.01) | 9404 | 1.00 (1.00 – 1.01) |

| PheCode | Disease | Sleep statistic | Prevalence | Risk ratio | P-value | N | Risk ratio (AHI >15) | N (AHI >15) | Risk ratio (adj sleep disorders) |
|---|---|---|---|---|---|---|---|---|---|
| 798 | Malaise and fatigue | Per88 | 19.1 | 1.00 (1.00 – 1.00) | 6.4E-01 | 12232 | 1.00 (0.99 – 1.00) | 5845 | 1.00 (1.00 – 1.00) |
| 979 | Adverse drug events and drug allergies | Per88 | 5.7 | 1.00 (0.99 – 1.01) | 6.4E-01 | 12232 | 1.00 (0.99 – 1.01) | 5845 | 1.00 (0.99 – 1.01) |
| 596.5 | Functional disorders of bladder | AHI 3% | 2.1 | 1.00 (0.99 – 1.00) | 6.4E-01 | 18332 | 1.00 (1.00 – 1.01) | 9404 | 1.00 (0.99 – 1.01) |
| 530.11 | GERD | Per88 | 31.1 | 1.00 (1.00 – 1.00) | 6.5E-01 | 12232 | 1.00 (1.00 – 1.00) | 5845 | 1.00 (1.00 – 1.00) |
| 368.2 | Diplopia and disorders of binocular vision | Per88 | 1.9 | 1.00 (0.98 – 1.01) | 6.5E-01 | 12232 | 0.99 (0.97 – 1.01) | 5845 | 1.00 (0.98 – 1.01) |
| 208 | Benign neoplasm of colon | Per88 | 14.6 | 1.00 (0.99 – 1.00) | 6.5E-01 | 12232 | 1.00 (0.99 – 1.00) | 5845 | 1.00 (0.99 – 1.00) |
| 481 | Influenza | AHI 3% | 7.0 | 1.00 (1.00 – 1.00) | 6.5E-01 | 18332 | 1.00 (1.00 – 1.00) | 9404 | 1.00 (1.00 – 1.00) |
| 472 | Chronic pharyngitis and nasopharyngitis | Per88 | 7.9 | 1.00 (0.99 – 1.01) | 6.5E-01 | 12232 | 1.00 (0.99 – 1.01) | 5845 | 1.00 (0.99 – 1.01) |
| 723 | Other disorders of cervical region | AHI 3% | 1.4 | 1.00 (0.99 – 1.00) | 6.5E-01 | 18332 | 1.01 (1.00 – 1.01) | 9404 | 1.00 (0.99 – 1.01) |
| 772.6 | Facial weakness | AHI 3% | 1.4 | 1.00 (0.99 – 1.01) | 6.6E-01 | 18332 | 1.00 (0.99 – 1.01) | 9404 | 1.00 (0.99 – 1.01) |
| 277.5 | Other disorders of lipoid metabolism | Per88 | 2.1 | 1.00 (0.99 – 1.02) | 6.6E-01 | 12232 | 1.01 (0.99 – 1.02) | 5845 | 1.00 (0.99 – 1.02) |
| 289.9 | Abnormality of red blood cells | AHI 3% | 2.5 | 1.00 (1.00 – 1.01) | 6.6E-01 | 18332 | 1.00 (1.00 – 1.01) | 9404 | 1.00 (1.00 – 1.01) |
| 476 | Allergic rhinitis | AHI 3% | 14.6 | 1.00 (1.00 – 1.00) | 6.7E-01 | 18332 | 1.00 (1.00 – 1.00) | 9404 | 1.00 (1.00 – 1.00) |
| 793.2 | Nonspecific abnormal findings on radiological and other examination of other intrathoracic organs (echocardiogram, etc) | AHI 3% | 4.0 | 1.00 (1.00 – 1.00) | 6.7E-01 | 18332 | 1.00 (0.99 – 1.00) | 9404 | 1.00 (1.00 – 1.00) |
| 698 | Pruritus and related conditions | AHI 3% | 9.5 | 1.00 (1.00 – 1.00) | 6.7E-01 | 18332 | 1.00 (1.00 – 1.00) | 9404 | 1.00 (1.00 – 1.00) |
| 300.1 | Anxiety disorder | Per88 | 24.6 | 1.00 (1.00 – 1.00) | 6.7E-01 | 12232 | 1.00 (1.00 – 1.00) | 5845 | 1.00 (1.00 – 1.00) |
| 727.4 | Ganglion and cyst of synovium, tendon, and bursa | Per88 | 2.5 | 1.00 (0.99 – 1.02) | 6.7E-01 | 12232 | 1.01 (0.99 – 1.02) | 5845 | 1.00 (0.99 – 1.02) |
| 440.9 | Atherosclerosis of aorta | Per88 | 1.2 | 1.00 (0.98 – 1.03) | 6.7E-01 | 12232 | 1.01 (0.99 – 1.02) | 5845 | 1.00 (0.98 – 1.03) |
| 374 | Other disorders of eyelids | AHI 3% | 2.7 | 1.00 (1.00 – 1.01) | 6.8E-01 | 18332 | 1.00 (0.99 – 1.00) | 9404 | 1.00 (1.00 – 1.01) |
| 483 | Acute bronchitis and bronchiolitis | Per88 | 5.5 | 1.00 (0.99 – 1.01) | 6.8E-01 | 12232 | 1.00 (0.99 – 1.01) | 5845 | 1.00 (0.99 – 1.01) |
| 250.41 | Impaired fasting glucose | Per88 | 3.7 | 1.00 (0.98 – 1.01) | 6.8E-01 | 12232 | 0.99 (0.98 – 1.01) | 5845 | 1.00 (0.98 – 1.01) |
| 574.3 | Cholecystitis without cholelithiasis | Per88 | 3.1 | 1.00 (0.99 – 1.02) | 6.8E-01 | 12232 | 1.01 (1.00 – 1.02) | 5845 | 1.00 (0.99 – 1.02) |
| 579.8 | Nonspecific abnormal findings in stool contents | Per88 | 4.7 | 1.00 (0.99 – 1.01) | 6.8E-01 | 12232 | 1.00 (0.99 – 1.01) | 5845 | 1.00 (0.99 – 1.01) |
| 593.2 | Microscopic hematuria | Per88 | 4.2 | 1.00 (0.99 – 1.01) | 6.8E-01 | 12232 | 1.00 (0.99 – 1.01) | 5845 | 1.00 (0.99 – 1.01) |
| 454.1 | Varicose veins of lower extremity | Per88 | 3.1 | 1.00 (0.99 – 1.01) | 6.9E-01 | 12232 | 1.00 (0.99 – 1.01) | 5845 | 1.00 (0.99 – 1.01) |
| 277 | Other disorders of metabolism | Per88 | 6.5 | 1.00 (0.99 – 1.01) | 7.0E-01 | 12232 | 1.00 (0.99 – 1.01) | 5845 | 1.00 (0.99 – 1.01) |
| 296.1 | Bipolar | Per88 | 3.4 | 1.00 (0.99 – 1.01) | 7.0E-01 | 12232 | 1.00 (0.99 – 1.01) | 5845 | 1.00 (0.99 – 1.01) |
| 593 | Hematuria | Per88 | 7.9 | 1.00 (0.99 – 1.01) | 7.0E-01 | 12232 | 1.00 (0.99 – 1.00) | 5845 | 1.00 (0.99 – 1.01) |
| 389.2 | Conductive hearing loss | AHI 3% | 1.0 | 1.00 (0.99 – 1.01) | 7.0E-01 | 18332 | 1.00 (0.98 – 1.01) | 9404 | 1.00 (0.99 – 1.01) |
| 766 | Neuralgia, neuritis, and radiculitis NOS | Per88 | 3.7 | 1.00 (0.99 – 1.01) | 7.0E-01 | 12232 | 1.00 (0.99 – 1.01) | 5845 | 1.00 (0.99 – 1.01) |
| 374 | Other disorders of eyelids | Per88 | 2.7 | 1.00 (0.99 – 1.01) | 7.1E-01 | 12232 | 1.00 (0.99 – 1.02) | 5845 | 1.00 (0.99 – 1.01) |
| 740.1 | Osteoarthritis; localized | Per88 | 3.5 | 1.00 (0.99 – 1.02) | 7.1E-01 | 12232 | 1.00 (0.99 – 1.01) | 5845 | 1.00 (0.99 – 1.02) |
| 300.8 | Acute reaction to stress | AHI 3% | 2.2 | 1.00 (1.00 – 1.01) | 7.1E-01 | 18332 | 1.00 (0.99 – 1.01) | 9404 | 1.00 (1.00 – 1.01) |

| PheCode | Disease | Sleep statistic | Prevalence | Risk ratio | P-value | N | Risk ratio (AHI >15) | N (AHI >15) | Risk ratio (adj sleep disorders) |
|---|---|---|---|---|---|---|---|---|---|
| 585.4 | Chronic kidney disease, Stage I or II | Per88 | 2.0 | 1.00 (0.99 – 1.02) | 7.1E-01 | 12232 | 1.00 (0.99 – 1.02) | 5845 | 1.00 (0.99 – 1.02) |
| 599.3 | Dysuria | Per88 | 6.6 | 1.00 (0.99 – 1.01) | 7.2E-01 | 12232 | 1.00 (0.99 – 1.01) | 5845 | 1.00 (0.99 – 1.01) |
| 586.4 | Stricture/obstruction of ureter | AHI 3% | 2.5 | 1.00 (0.99 – 1.00) | 7.2E-01 | 18332 | 1.00 (1.00 – 1.01) | 9404 | 1.00 (0.99 – 1.01) |
| 772.6 | Facial weakness | Per88 | 1.4 | 1.00 (0.98 – 1.02) | 7.2E-01 | 12232 | 1.00 (0.98 – 1.02) | 5845 | 1.00 (0.98 – 1.02) |
| 727.1 | Synovitis and tenosynovitis | Per88 | 5.9 | 1.00 (0.99 – 1.01) | 7.2E-01 | 12232 | 1.00 (0.99 – 1.01) | 5845 | 1.00 (0.99 – 1.01) |
| 465.4 | Acute laryngitis and tracheitis | Per88 | 1.5 | 1.00 (0.98 – 1.01) | 7.2E-01 | 12232 | 1.00 (0.98 – 1.02) | 5845 | 1.00 (0.98 – 1.01) |
| 371.3 | Inflammation of eyelids | AHI 3% | 3.2 | 1.00 (1.00 – 1.00) | 7.2E-01 | 18332 | 1.00 (1.00 – 1.01) | 9404 | 1.00 (1.00 – 1.00) |
| 526 | Diseases of the jaws | Per88 | 1.6 | 1.00 (0.99 – 1.02) | 7.3E-01 | 12232 | 1.00 (0.98 – 1.02) | 5845 | 1.00 (0.99 – 1.02) |
| 286.81 | Primary hypercoagulable state | AHI 3% | 1.1 | 1.00 (0.99 – 1.01) | 7.3E-01 | 18332 | 1.00 (0.99 – 1.01) | 9404 | 1.00 (0.99 – 1.01) |
| 427.61 | Supraventricular premature beats | AHI 3% | 1.8 | 1.00 (0.99 – 1.01) | 7.3E-01 | 18332 | 1.01 (1.00 – 1.01) | 9404 | 1.00 (0.99 – 1.01) |
| 765 | Cervical radiculitis | Per88 | 4.4 | 1.00 (0.99 – 1.01) | 7.3E-01 | 12232 | 1.01 (1.00 – 1.02) | 5845 | 1.00 (0.99 – 1.01) |
| 720 | Spinal stenosis | Per88 | 4.6 | 1.00 (0.99 – 1.01) | 7.3E-01 | 12232 | 1.00 (0.99 – 1.01) | 5845 | 1.00 (0.99 – 1.01) |
| 348.7 | Coma | AHI 3% | 1.5 | 1.00 (0.99 – 1.01) | 7.4E-01 | 18332 | 1.00 (0.99 – 1.00) | 9404 | 1.00 (0.99 – 1.01) |
| 110.13 | Dermatophytosis of the body | AHI 3% | 2.3 | 1.00 (1.00 – 1.01) | 7.4E-01 | 18332 | 1.00 (0.99 – 1.00) | 9404 | 1.00 (1.00 – 1.01) |
| 561 | Symptoms involving digestive system | AHI 3% | 3.6 | 1.00 (1.00 – 1.00) | 7.4E-01 | 18332 | 1.00 (1.00 – 1.01) | 9404 | 1.00 (1.00 – 1.00) |
| 442.1 | Aortic aneurysm | AHI 3% | 1.4 | 1.00 (0.99 – 1.01) | 7.4E-01 | 18332 | 1.00 (0.99 – 1.01) | 9404 | 1.00 (0.99 – 1.01) |
| 285 | Other anemias | AHI 3% | 14.6 | 1.00 (1.00 – 1.00) | 7.4E-01 | 18332 | 1.00 (1.00 – 1.01) | 9404 | 1.00 (1.00 – 1.00) |
| 369.5 | Conjunctivitis, infectious | AHI 3% | 5.4 | 1.00 (1.00 – 1.00) | 7.4E-01 | 18332 | 1.00 (0.99 – 1.00) | 9404 | 1.00 (1.00 – 1.00) |
| 290.1 | Dementias | Per88 | 1.7 | 1.00 (0.98 – 1.02) | 7.4E-01 | 12232 | 1.00 (0.98 – 1.02) | 5845 | 1.00 (0.98 – 1.02) |
| 530.13 | Barrett's esophagus | AHI 3% | 1.9 | 1.00 (0.99 – 1.01) | 7.6E-01 | 18332 | 1.00 (0.99 – 1.00) | 9404 | 1.00 (0.99 – 1.01) |
| 296.2 | Depression | Per88 | 11.9 | 1.00 (0.99 – 1.00) | 7.6E-01 | 12232 | 1.00 (0.99 – 1.01) | 5845 | 1.00 (0.99 – 1.00) |
| 686.1 | Carbuncle and furuncle | AHI 3% | 1.6 | 1.00 (0.99 – 1.01) | 7.6E-01 | 18332 | 1.01 (1.00 – 1.02) | 9404 | 1.00 (0.99 – 1.01) |
| 458.1 | Orthostatic hypotension | Per88 | 1.7 | 1.00 (0.98 – 1.01) | 7.6E-01 | 12232 | 1.00 (0.98 – 1.01) | 5845 | 1.00 (0.98 – 1.01) |
| 426 | Cardiac conduction disorders | AHI 3% | 1.2 | 1.00 (0.99 – 1.01) | 7.7E-01 | 18332 | 1.00 (1.00 – 1.01) | 9404 | 1.00 (0.99 – 1.01) |
| 418 | Nonspecific chest pain | AHI 3% | 20.0 | 1.00 (1.00 – 1.00) | 7.7E-01 | 18332 | 1.00 (1.00 – 1.00) | 9404 | 1.00 (1.00 – 1.00) |
| 565.1 | Anal and rectal polyp | Per88 | 3.8 | 1.00 (0.99 – 1.01) | 7.7E-01 | 12232 | 1.00 (0.99 – 1.01) | 5845 | 1.00 (0.99 – 1.01) |
| 367.1 | Myopia | Per88 | 2.5 | 1.00 (0.99 – 1.01) | 7.7E-01 | 12232 | 1.00 (0.99 – 1.01) | 5845 | 1.00 (0.99 – 1.01) |
| 721.8 | Other allied disorders of spine | Per88 | 2.0 | 1.00 (0.98 – 1.01) | 7.7E-01 | 12232 | 1.00 (0.99 – 1.02) | 5845 | 1.00 (0.98 – 1.01) |
| 599.8 | Other symptoms involving urinary system | AHI 3% | 3.2 | 1.00 (0.99 – 1.00) | 7.8E-01 | 18332 | 1.00 (1.00 – 1.01) | 9404 | 1.00 (1.00 – 1.00) |
| 110.2 | Dermatomycoses | AHI 3% | 1.8 | 1.00 (0.99 – 1.01) | 7.8E-01 | 18332 | 1.00 (0.99 – 1.01) | 9404 | 1.00 (0.99 – 1.01) |
| 274.1 | Gout | Per88 | 4.7 | 1.00 (0.99 – 1.01) | 7.8E-01 | 12232 | 1.00 (0.99 – 1.01) | 5845 | 1.00 (0.99 – 1.01) |
| 465 | Acute upper respiratory infections of multiple or unspecified sites | Per88 | 10.9 | 1.00 (0.99 – 1.01) | 7.8E-01 | 12232 | 1.00 (0.99 – 1.00) | 5845 | 1.00 (0.99 – 1.01) |
| 433.8 | Late effects of cerebrovascular disease | Per88 | 1.4 | 1.00 (0.98 – 1.02) | 7.8E-01 | 12232 | 1.01 (0.99 – 1.02) | 5845 | 1.00 (0.98 – 1.02) |
| 411.1 | Unstable angina (intermediate coronary syndrome) | AHI 3% | 1.7 | 1.00 (0.99 – 1.01) | 7.9E-01 | 18332 | 1.00 (0.99 – 1.00) | 9404 | 1.00 (0.99 – 1.01) |
| 592.11 | Acute cystitis | Per88 | 2.2 | 1.00 (0.99 – 1.02) | 7.9E-01 | 12232 | 1.00 (0.99 – 1.02) | 5845 | 1.00 (0.99 – 1.02) |

| PheCode | Disease | Sleep statistic | Prevalence | Risk ratio | P-value | N | Risk ratio (AHI >15) | N (AHI >15) | Risk ratio (adj sleep disorders) |
|---|---|---|---|---|---|---|---|---|---|
| 470 | Septal Deviations/Turbinate Hypertrophy | Per88 | 7.3 | 1.00 (0.99 – 1.01) | 8.0E-01 | 12232 | 1.00 (0.99 – 1.01) | 5845 | 1.00 (0.99 – 1.01) |
| 594.8 | Renal colic | AHI 3% | 1.4 | 1.00 (0.99 – 1.01) | 8.0E-01 | 18332 | 1.00 (0.99 – 1.01) | 9404 | 1.00 (0.99 – 1.01) |
| 761 | Cervicalgia | AHI 3% | 12.3 | 1.00 (1.00 – 1.00) | 8.0E-01 | 18332 | 1.00 (1.00 – 1.00) | 9404 | 1.00 (1.00 – 1.00) |
| 300.13 | Phobia | Per88 | 2.4 | 1.00 (0.99 – 1.01) | 8.0E-01 | 12232 | 1.00 (0.98 – 1.01) | 5845 | 1.00 (0.99 – 1.01) |
| 272.1 | Hyperlipidemia | Per88 | 30.5 | 1.00 (1.00 – 1.00) | 8.1E-01 | 12232 | 1.00 (0.99 – 1.00) | 5845 | 1.00 (1.00 – 1.00) |
| 574.1 | Cholelithiasis | AHI 3% | 4.6 | 1.00 (1.00 – 1.00) | 8.1E-01 | 18332 | 1.00 (1.00 – 1.01) | 9404 | 1.00 (1.00 – 1.00) |
| 440 | Atherosclerosis | Per88 | 1.2 | 1.00 (0.98 – 1.02) | 8.2E-01 | 12232 | 1.00 (0.99 – 1.02) | 5845 | 1.00 (0.98 – 1.02) |
| 241.1 | Nontoxic uninodular goiter | AHI 3% | 5.6 | 1.00 (1.00 – 1.00) | 8.2E-01 | 18332 | 1.00 (1.00 – 1.00) | 9404 | 1.00 (1.00 – 1.00) |
| 418.1 | Precordial pain | AHI 3% | 3.4 | 1.00 (1.00 – 1.01) | 8.2E-01 | 18332 | 1.00 (1.00 – 1.01) | 9404 | 1.00 (1.00 – 1.01) |
| 440.9 | Atherosclerosis of aorta | AHI 3% | 1.2 | 1.00 (0.99 – 1.01) | 8.2E-01 | 18332 | 1.00 (0.99 – 1.01) | 9404 | 1.00 (0.99 – 1.01) |
| 427.5 | Arrhythmia (cardiac) NOS | AHI 3% | 13.9 | 1.00 (1.00 – 1.00) | 8.3E-01 | 18332 | 1.00 (1.00 – 1.00) | 9404 | 1.00 (1.00 – 1.00) |
| 411.1 | Unstable angina (intermediate coronary syndrome) | Per88 | 1.7 | 1.00 (0.98 – 1.02) | 8.3E-01 | 12232 | 1.00 (0.98 – 1.01) | 5845 | 1.00 (0.98 – 1.02) |
| 350.3 | Lack of coordination | AHI 3% | 1.2 | 1.00 (0.99 – 1.01) | 8.3E-01 | 18332 | 1.00 (1.00 – 1.01) | 9404 | 1.00 (0.99 – 1.01) |
| 819 | Skull and face fracture and other intercranial injury | Per88 | 1.4 | 1.00 (0.99 – 1.02) | 8.4E-01 | 12232 | 1.00 (0.98 – 1.02) | 5845 | 1.00 (0.99 – 1.02) |
| 596 | Other disorders of bladder | AHI 3% | 2.1 | 1.00 (0.99 – 1.01) | 8.4E-01 | 18332 | 1.00 (1.00 – 1.01) | 9404 | 1.00 (0.99 – 1.01) |
| 411.4 | Coronary atherosclerosis | AHI 3% | 8.1 | 1.00 (1.00 – 1.00) | 8.5E-01 | 18332 | 1.00 (1.00 – 1.00) | 9404 | 1.00 (1.00 – 1.00) |
| 350.2 | Abnormality of gait | AHI 3% | 4.7 | 1.00 (1.00 – 1.00) | 8.5E-01 | 18332 | 1.00 (1.00 – 1.01) | 9404 | 1.00 (1.00 – 1.00) |
| 427.8 | Sinoatrial node dysfunction (Bradycardia) | Per88 | 1.1 | 1.00 (0.98 – 1.02) | 8.5E-01 | 12232 | 0.99 (0.97 – 1.01) | 5845 | 1.00 (0.98 – 1.02) |
| 694.2 | Other dyschromia | AHI 3% | 12.9 | 1.00 (1.00 – 1.00) | 8.5E-01 | 18332 | 1.00 (1.00 – 1.00) | 9404 | 1.00 (1.00 – 1.00) |
| 380.4 | Impacted cerumen | Per88 | 3.7 | 1.00 (0.99 – 1.01) | 8.6E-01 | 12232 | 1.00 (0.99 – 1.01) | 5845 | 1.00 (0.99 – 1.01) |
| 561.1 | Diarrhea | AHI 3% | 9.7 | 1.00 (1.00 – 1.00) | 8.6E-01 | 18332 | 1.00 (1.00 – 1.00) | 9404 | 1.00 (1.00 – 1.00) |
| 495 | Asthma | Per88 | 19.3 | 1.00 (1.00 – 1.00) | 8.6E-01 | 12232 | 1.00 (1.00 – 1.01) | 5845 | 1.00 (1.00 – 1.00) |
| 475.9 | Postnasal drip | AHI 3% | 5.0 | 1.00 (1.00 – 1.00) | 8.7E-01 | 18332 | 1.00 (0.99 – 1.00) | 9404 | 1.00 (1.00 – 1.00) |
| 290.1 | Dementias | AHI 3% | 1.7 | 1.00 (0.99 – 1.01) | 8.7E-01 | 18332 | 1.01 (1.00 – 1.01) | 9404 | 1.00 (0.99 – 1.01) |
| 474.2 | Chronic tonsillitis and adenoiditis | Per88 | 2.5 | 1.00 (0.99 – 1.01) | 8.8E-01 | 12232 | 1.00 (0.99 – 1.02) | 5845 | 1.00 (0.99 – 1.01) |
| 427.6 | Premature beats | AHI 3% | 3.4 | 1.00 (1.00 – 1.00) | 8.8E-01 | 18332 | 1.00 (0.99 – 1.00) | 9404 | 1.00 (1.00 – 1.00) |
| 721.8 | Other allied disorders of spine | AHI 3% | 2.0 | 1.00 (0.99 – 1.01) | 8.9E-01 | 18332 | 1.00 (1.00 – 1.01) | 9404 | 1.00 (0.99 – 1.01) |
| 599.2 | Retention of urine | Per88 | 4.0 | 1.00 (0.99 – 1.01) | 8.9E-01 | 12232 | 1.00 (0.99 – 1.01) | 5845 | 1.00 (0.99 – 1.01) |
| 300.11 | Generalized anxiety disorder | Per88 | 7.6 | 1.00 (0.99 – 1.01) | 8.9E-01 | 12232 | 1.00 (0.99 – 1.00) | 5845 | 1.00 (0.99 – 1.01) |
| 772.2 | Spasm of muscle | Per88 | 2.9 | 1.00 (0.99 – 1.01) | 8.9E-01 | 12232 | 1.00 (0.99 – 1.01) | 5845 | 1.00 (0.99 – 1.01) |
| 277.5 | Other disorders of lipoid metabolism | AHI 3% | 2.1 | 1.00 (0.99 – 1.01) | 9.0E-01 | 18332 | 1.00 (1.00 – 1.01) | 9404 | 1.00 (0.99 – 1.01) |
| 772.3 | Muscle weakness | Per88 | 2.7 | 1.00 (0.99 – 1.01) | 9.0E-01 | 12232 | 1.00 (0.98 – 1.01) | 5845 | 1.00 (0.99 – 1.01) |
| 912 | Insect bite | Per88 | 2.1 | 1.00 (0.99 – 1.01) | 9.1E-01 | 12232 | 1.00 (0.99 – 1.02) | 5845 | 1.00 (0.99 – 1.01) |
| 427.8 | Sinoatrial node dysfunction (Bradycardia) | AHI 3% | 1.1 | 1.00 (0.99 – 1.01) | 9.1E-01 | 18332 | 1.00 (1.00 – 1.01) | 9404 | 1.00 (0.99 – 1.01) |

| PheCode | Disease | Sleep statistic | Prevalence | Risk ratio | P-value | N | Risk ratio (AHI >15) | N (AHI >15) | Risk ratio (adj sleep disorders) |
|---|---|---|---|---|---|---|---|---|---|
| 599.3 | Dysuria | AHI 3% | 6.6 | 1.00 (1.00 – 1.00) | 9.1E-01 | 18332 | 1.00 (1.00 – 1.00) | 9404 | 1.00 (1.00 – 1.00) |
| 764 | Sciatica | Per88 | 8.1 | 1.00 (0.99 – 1.01) | 9.1E-01 | 12232 | 1.00 (0.99 – 1.01) | 5845 | 1.00 (0.99 – 1.01) |
| 721.1 | Spondylosis without myelopathy | Per88 | 8.9 | 1.00 (0.99 – 1.01) | 9.1E-01 | 12232 | 1.00 (0.99 – 1.01) | 5845 | 1.00 (0.99 – 1.01) |
| 245.21 | Chronic lymphocytic thyroiditis | AHI 3% | 1.6 | 1.00 (0.99 – 1.00) | 9.2E-01 | 18332 | 1.00 (1.00 – 1.01) | 9404 | 1.00 (0.99 – 1.00) |
| 389.1 | Sensorineural hearing loss | AHI 3% | 6.0 | 1.00 (1.00 – 1.00) | 9.2E-01 | 18332 | 1.00 (1.00 – 1.00) | 9404 | 1.00 (1.00 – 1.00) |
| 764 | Sciatica | AHI 3% | 8.1 | 1.00 (1.00 – 1.00) | 9.2E-01 | 18332 | 1.00 (1.00 – 1.00) | 9404 | 1.00 (1.00 – 1.00) |
| 008.6 | Viral Enteritis | Per88 | 2.3 | 1.00 (0.99 – 1.01) | 9.3E-01 | 12232 | 1.00 (0.98 – 1.01) | 5845 | 1.00 (0.99 – 1.01) |
| 735.3 | Hallux valgus (Bunion) | Per88 | 2.9 | 1.00 (0.99 – 1.01) | 9.3E-01 | 12232 | 1.00 (0.99 – 1.01) | 5845 | 1.00 (0.99 – 1.01) |
| 208 | Benign neoplasm of colon | AHI 3% | 14.6 | 1.00 (1.00 – 1.00) | 9.4E-01 | 18332 | 1.00 (1.00 – 1.00) | 9404 | 1.00 (1.00 – 1.00) |
| 300.3 | Obsessive-compulsive disorders | Per88 | 1.4 | 1.00 (0.99 – 1.01) | 9.4E-01 | 12232 | 0.99 (0.97 – 1.01) | 5845 | 1.00 (0.99 – 1.01) |
| 244.2 | Acquired hypothyroidism | AHI 3% | 2.8 | 1.00 (1.00 – 1.00) | 9.4E-01 | 18332 | 1.00 (1.00 – 1.01) | 9404 | 1.00 (1.00 – 1.00) |
| 296.22 | Major depressive disorder | Per88 | 22.7 | 1.00 (1.00 – 1.00) | 9.5E-01 | 12232 | 1.00 (1.00 – 1.00) | 5845 | 1.00 (1.00 – 1.00) |
| 415.11 | Pulmonary embolism and infarction, acute | AHI 3% | 3.0 | 1.00 (0.99 – 1.00) | 9.5E-01 | 18332 | 1.00 (1.00 – 1.01) | 9404 | 1.00 (0.99 – 1.00) |
| 459.9 | Circulatory disease NEC | AHI 3% | 4.5 | 1.00 (1.00 – 1.00) | 9.5E-01 | 18332 | 1.00 (1.00 – 1.01) | 9404 | 1.00 (1.00 – 1.00) |
| 293 | Symptoms involving head and neck | Per88 | 1.1 | 1.00 (0.98 – 1.02) | 9.5E-01 | 12232 | 1.00 (0.98 – 1.02) | 5845 | 1.00 (0.98 – 1.02) |
| 415 | Pulmonary heart disease | AHI 3% | 1.3 | 1.00 (0.99 – 1.01) | 9.5E-01 | 18332 | 1.00 (1.00 – 1.01) | 9404 | 1.00 (0.99 – 1.01) |
| 008.6 | Viral Enteritis | AHI 3% | 2.3 | 1.00 (0.99 – 1.01) | 9.5E-01 | 18332 | 1.00 (0.99 – 1.01) | 9404 | 1.00 (0.99 – 1.01) |
| 516.1 | Hemoptysis | AHI 3% | 1.5 | 1.00 (0.99 – 1.01) | 9.5E-01 | 18332 | 1.00 (0.99 – 1.01) | 9404 | 1.00 (0.99 – 1.01) |
| 532 | Dysphagia | AHI 3% | 7.4 | 1.00 (1.00 – 1.00) | 9.5E-01 | 18332 | 1.00 (1.00 – 1.00) | 9404 | 1.00 (1.00 – 1.00) |
| 172.22 | Squamous cell carcinoma | Per88 | 2.2 | 1.00 (0.98 – 1.02) | 9.5E-01 | 12232 | 1.00 (0.98 – 1.01) | 5845 | 1.00 (0.98 – 1.02) |
| 172.2 | Other non-epithelial cancer of skin | AHI 3% | 5.7 | 1.00 (1.00 – 1.00) | 9.6E-01 | 18332 | 1.00 (1.00 – 1.00) | 9404 | 1.00 (1.00 – 1.00) |
| 912 | Insect bite | AHI 3% | 2.1 | 1.00 (0.99 – 1.01) | 9.6E-01 | 18332 | 1.00 (1.00 – 1.01) | 9404 | 1.00 (0.99 – 1.01) |
| 345 | Epilepsy, recurrent seizures, convulsions | Per88 | 2.7 | 1.00 (0.99 – 1.01) | 9.6E-01 | 12232 | 1.00 (0.98 – 1.01) | 5845 | 1.00 (0.99 – 1.01) |
| 367.8 | Hypermetropia | Per88 | 1.8 | 1.00 (0.99 – 1.01) | 9.6E-01 | 12232 | 1.00 (0.99 – 1.02) | 5845 | 1.00 (0.99 – 1.01) |
| 596.5 | Functional disorders of bladder | Per88 | 2.1 | 1.00 (0.98 – 1.01) | 9.6E-01 | 12232 | 1.00 (0.98 – 1.01) | 5845 | 1.00 (0.98 – 1.01) |
| 965 | Poisoning by analgesics, antipyretics, and antirheumatics | AHI 3% | 4.3 | 1.00 (1.00 – 1.00) | 9.6E-01 | 18332 | 1.00 (1.00 – 1.01) | 9404 | 1.00 (1.00 – 1.00) |
| 724.8 | Other symptoms referable to back | Per88 | 1.3 | 1.00 (0.98 – 1.02) | 9.6E-01 | 12232 | 1.01 (0.99 – 1.03) | 5845 | 1.00 (0.98 – 1.02) |
| 514.2 | Solitary pulmonary nodule | AHI 3% | 4.2 | 1.00 (1.00 – 1.00) | 9.6E-01 | 18332 | 1.00 (0.99 – 1.00) | 9404 | 1.00 (1.00 – 1.00) |
| 698 | Pruritus and related conditions | Per88 | 9.5 | 1.00 (0.99 – 1.01) | 9.7E-01 | 12232 | 1.00 (0.99 – 1.01) | 5845 | 1.00 (0.99 – 1.01) |
| 464 | Acute sinusitis | AHI 3% | 5.1 | 1.00 (1.00 – 1.00) | 9.7E-01 | 18332 | 1.00 (0.99 – 1.00) | 9404 | 1.00 (1.00 – 1.00) |
| 276.12 | Hyposmolality and/or hyponatremia | AHI 3% | 2.6 | 1.00 (0.99 – 1.01) | 9.7E-01 | 18332 | 1.00 (1.00 – 1.01) | 9404 | 1.00 (1.00 – 1.01) |
| 704 | Diseases of hair and hair follicles | AHI 3% | 3.7 | 1.00 (1.00 – 1.00) | 9.8E-01 | 18332 | 1.00 (1.00 – 1.01) | 9404 | 1.00 (1.00 – 1.00) |
| 259 | Other endocrine disorders | Per88 | 1.7 | 1.00 (0.98 – 1.02) | 9.8E-01 | 12232 | 1.02 (1.00 – 1.04) | 5845 | 1.00 (0.98 – 1.02) |
| 292.1 | Aphasia/speech disturbance | AHI 3% | 2.0 | 1.00 (0.99 – 1.01) | 9.8E-01 | 18332 | 1.00 (1.00 – 1.01) | 9404 | 1.00 (0.99 – 1.01) |
| 375.1 | Dry eyes | Per88 | 4.8 | 1.00 (0.99 – 1.01) | 9.8E-01 | 12232 | 1.00 (0.99 – 1.01) | 5845 | 1.00 (0.99 – 1.01) |
| 563 | Constipation | Per88 | 10.8 | 1.00 (0.99 – 1.01) | 9.9E-01 | 12232 | 1.00 (0.99 – 1.01) | 5845 | 1.00 (0.99 – 1.01) |

| PheCode | Disease | Sleep statistic | Prevalence | Risk ratio | P-value | N | Risk ratio (AHI >15) | N (AHI >15) | Risk ratio (adj sleep disorders) |
|---|---|---|---|---|---|---|---|---|---|
| 427.22 | Atrial flutter | AHI 3% | 2.6 | 1.00 (0.99 – 1.01) | 9.9E-01 | 18332 | 1.00 (1.00 – 1.01) | 9404 | 1.00 (0.99 – 1.01) |
| 386.2 | Peripheral or central vertigo | AHI 3% | 2.8 | 1.00 (1.00 – 1.00) | 9.9E-01 | 18332 | 1.00 (0.99 – 1.00) | 9404 | 1.00 (1.00 – 1.00) |
| 300.3 | Obsessive-compulsive disorders | AHI 3% | 1.4 | 1.00 (0.99 – 1.01) | 9.9E-01 | 18332 | 1.01 (1.00 – 1.01) | 9404 | 1.00 (1.00 – 1.01) |

#### Table S12. Cross-sectional sleep statistic associations with comorbidities sensitivity analyses.

Cross-sectional analyses used the first sleep recording (prioritizing diagnostics, then splits, then home sleep tests) and are ordered by p-values. Primary analyses considered joint-sex PheCodes with >1% prevalence rate and were rank-normalized after adjusting for age, sex, BMI, population, sleep study type, and healthcare encounters. The primary analysis considered all available sleep laboratory data (i.e. including participants being screened for other sleep disorders).

47 non-redundant PheCodes were Bonferroni-level significant (**bolded**) and 117 non-redundant PheCodes were FDR-level significant (in red, uncorrected p-values).

Due to PDF width constraints, sensitivity analyses are presented in two tables, with primary columns repeated (apart from incidence and n):

**Table S11 further includes:**
A sensitivity analysis considering only participants with AHI >15.
A sensitivity analysis further adjusting for an indicator variable for other common sleep disorders (PheCodes 327, 327.1, 327.4, 327.41, 327.5, 327.6, 327.7, 327.71) in the full sleep recording sample.

**Table S12 further includes:**
A sensitivity analysis further adjusting for an indicator variable for common pulmonary disorders (PheCodes 415.1, 415.11, 415.2, 480.1, 480.11, 480.2, 480, 495, 495.2, 496.1, 496.2, 496.21, 496.3, 497, 502, 503, 505, 506, 509.2).
P-values from sleep statistic × sex interaction analyses (not rank-normalized) performed separately. Two Bonferroni-level significant values are displayed in green.
Equivalent sex-stratified results.

| PheCode | Disease | Sleep statistic | Risk ratio | P-value | Risk ratio (adj pulmonary disorders) | Sleep statistic x sex interaction p-value | Risk ratio (Men) | Risk ratio (Women) |
|---|---|---|---|---|---|---|---|---|
| 1013 | Asphyxia and hypoxemia | Per88 | 1.07 (1.06 – 1.07) | **1.1E-70** | 1.07 (1.06 – 1.07) | 2.9E-01 | 1.06 (1.05 – 1.08) | 1.07 (1.06 – 1.08) |
| 496 | Chronic airway obstruction | Per88 | 1.06 (1.05 – 1.06) | **3.2E-45** | 1.05 (1.05 – 1.06) | 9.6E-01 | 1.05 (1.03 – 1.06) | 1.07 (1.06 – 1.08) |
| 509.1 | Respiratory failure | Per88 | 1.07 (1.06 – 1.08) | **7.0E-40** | 1.06 (1.05 – 1.08) | 5.8E-01 | 1.06 (1.04 – 1.07) | 1.07 (1.06 – 1.09) |
| 415.2 | Chronic pulmonary heart disease | Per88 | 1.07 (1.05 – 1.08) | **5.5E-38** | 1.06 (1.05 – 1.07) | 3.4E-01 | 1.06 (1.05 – 1.08) | 1.07 (1.06 – 1.08) |
| 496.21 | Obstructive chronic bronchitis | Per88 | 1.07 (1.06 – 1.09) | **2.4E-34** | 1.07 (1.06 – 1.08) | 3.5E-01 | 1.06 (1.04 – 1.07) | 1.09 (1.07 – 1.10) |
| 508 | Pulmonary collapse; interstitial and compensatory emphysema | Per88 | 1.04 (1.04 – 1.05) | **3.9E-34** | 1.04 (1.03 – 1.05) | 1.0E-01 | 1.03 (1.02 – 1.04) | 1.06 (1.05 – 1.07) |
| 503 | Pulmonary congestion and hypostasis | Per88 | 1.06 (1.05 – 1.07) | **6.8E-31** | 1.05 (1.04 – 1.06) | 1.6E-01 | 1.04 (1.03 – 1.06) | 1.07 (1.06 – 1.08) |
| 496.1 | Emphysema | Per88 | 1.06 (1.05 – 1.07) | **1.1E-30** | 1.06 (1.05 – 1.07) | 3.9E-01 | 1.05 (1.03 – 1.06) | 1.07 (1.06 – 1.09) |

| PheCode | Disease | Sleep statistic | Risk ratio | P-value | Risk ratio (adj pulmonary disorders) | Sleep statistic x sex interaction p-value | Risk ratio (Men) | Risk ratio (Women) |
|---|---|---|---|---|---|---|---|---|
| 512.7 | Shortness of breath | Per88 | 1.02 (1.02 – 1.03) | **8.6E-26** | 1.02 (1.01 – 1.02) | 2.5E-01 | 1.02 (1.01 – 1.03) | 1.02 (1.02 – 1.03) |
| 782.3 | Edema | Per88 | 1.03 (1.03 – 1.04) | **6.7E-25** | 1.03 (1.02 – 1.04) | 9.1E-03 | 1.03 (1.02 – 1.04) | 1.03 (1.02 – 1.04) |
| 428.1 | Congestive heart failure (CHF) NOS | Per88 | 1.04 (1.03 – 1.05) | **1.3E-21** | 1.04 (1.03 – 1.05) | 7.7E-01 | 1.03 (1.02 – 1.04) | 1.05 (1.04 – 1.06) |
| 401.1 | Essential hypertension | AHI 3% | 1.00 (1.00 – 1.01) | **7.0E-20** | 1.00 (1.00 – 1.01) | 7.9E-02 | 1.00 (1.00 – 1.00) | 1.01 (1.00 – 1.01) |
| 512.9 | Other dyspnea | Per88 | 1.02 (1.01 – 1.02) | **5.8E-18** | 1.01 (1.01 – 1.02) | 1.3E-02 | 1.01 (1.01 – 1.02) | 1.02 (1.02 – 1.03) |
| 428.4 | Heart failure with preserved EF [Diastolic heart failure] | Per88 | 1.05 (1.04 – 1.06) | **2.0E-17** | 1.05 (1.04 – 1.06) | 7.2E-01 | 1.04 (1.02 – 1.06) | 1.06 (1.05 – 1.08) |
| 480 | Pneumonia | Per88 | 1.03 (1.02 – 1.03) | **5.6E-17** | 1.02 (1.01 – 1.03) | 3.7E-01 | 1.03 (1.02 – 1.03) | 1.03 (1.02 – 1.04) |
| 505 | Other pulmonary inflamation or edema | Per88 | 1.06 (1.04 – 1.07) | **2.0E-16** | 1.06 (1.04 – 1.07) | 8.1E-01 | 1.05 (1.03 – 1.07) | 1.06 (1.04 – 1.08) |
| 510 | Other diseases of lung | Per88 | 1.04 (1.03 – 1.04) | **2.0E-15** | 1.03 (1.02 – 1.04) | 6.2E-01 | 1.02 (1.01 – 1.04) | 1.04 (1.03 – 1.06) |
| 428.2 | Heart failure NOS | Per88 | 1.05 (1.03 – 1.06) | **1.2E-14** | 1.05 (1.03 – 1.06) | 5.8E-01 | 1.04 (1.02 – 1.05) | 1.05 (1.03 – 1.07) |
| 276.6 | Fluid overload | Per88 | 1.05 (1.04 – 1.06) | **1.3E-14** | 1.05 (1.04 – 1.06) | 5.5E-01 | 1.04 (1.03 – 1.06) | 1.06 (1.04 – 1.08) |
| 340 | Migraine | AHI 3% | 0.99 (0.99 – 0.99) | **5.6E-14** | 0.99 (0.99 – 0.99) | 7.3E-01 | 0.99 (0.99 – 1.00) | 0.99 (0.99 – 0.99) |
| 401.21 | Hypertensive heart disease | Per88 | 1.04 (1.03 – 1.06) | **6.2E-13** | 1.04 (1.03 – 1.06) | 8.3E-01 | 1.03 (1.01 – 1.05) | 1.06 (1.04 – 1.07) |
| 805 | Fracture of vertebral column without mention of spinal cord injury | Per88 | 1.05 (1.03 – 1.06) | **1.2E-12** | 1.05 (1.03 – 1.06) | 5.3E-01 | 1.05 (1.03 – 1.07) | 1.04 (1.02 – 1.06) |
| 458.9 | Hypotension NOS | Per88 | 1.03 (1.02 – 1.04) | **1.5E-12** | 1.03 (1.02 – 1.04) | 6.1E-01 | 1.03 (1.01 – 1.04) | 1.04 (1.03 – 1.05) |
| 401.1 | Essential hypertension | Per88 | 1.01 (1.01 – 1.01) | **2.0E-10** | 1.01 (1.01 – 1.01) | 6.8E-01 | 1.00 (1.00 – 1.01) | 1.01 (1.01 – 1.01) |
| 585.1 | Acute renal failure | Per88 | 1.03 (1.02 – 1.04) | **3.6E-10** | 1.03 (1.02 – 1.04) | 2.8E-01 | 1.02 (1.01 – 1.03) | 1.04 (1.03 – 1.05) |
| 427.7 | Tachycardia NOS | Per88 | 1.02 (1.01 – 1.03) | **5.6E-10** | 1.02 (1.01 – 1.02) | 3.1E-02 | 1.02 (1.01 – 1.03) | 1.02 (1.01 – 1.03) |
| 327.4 | Insomnia | AHI 3% | 0.99 (0.99 – 1.00) | **8.3E-10** | 0.99 (0.99 – 1.00) | 1.0E-01 | 0.99 (0.99 – 1.00) | 0.99 (0.99 – 1.00) |
| 316 | Substance addiction and disorders | Per88 | 1.02 (1.01 – 1.03) | **4.4E-09** | 1.02 (1.01 – 1.03) | 8.9E-01 | 1.02 (1.01 – 1.03) | 1.02 (1.01 – 1.03) |
| 260 | Protein-calorie malnutrition | Per88 | 1.03 (1.02 – 1.04) | **1.6E-08** | 1.03 (1.02 – 1.04) | 7.3E-01 | 1.03 (1.02 – 1.05) | 1.03 (1.01 – 1.04) |
| 250.2 | Type 2 diabetes | AHI 3% | 1.01 (1.00 – 1.01) | **1.8E-08** | 1.01 (1.00 – 1.01) | 8.0E-01 | 1.00 (1.00 – 1.01) | 1.01 (1.00 – 1.01) |
| 339 | Other headache syndromes | AHI 3% | 0.99 (0.99 – 1.00) | **2.2E-08** | 0.99 (0.99 – 1.00) | 5.9E-01 | 0.99 (0.99 – 1.00) | 1.00 (0.99 – 1.00) |
| 317.1 | Alcoholism | Per88 | 1.03 (1.02 – 1.04) | **6.1E-08** | 1.03 (1.02 – 1.04) | 3.2E-01 | 1.03 (1.02 – 1.04) | 1.02 (1.00 – 1.03) |
| 250.2 | Type 2 diabetes | Per88 | 1.01 (1.01 – 1.02) | **1.0E-07** | 1.01 (1.01 – 1.02) | 1.7E-01 | 1.01 (1.00 – 1.02) | 1.01 (1.01 – 1.02) |
| 401.3 | Other hypertensive complications | AHI 3% | 1.02 (1.01 – 1.02) | **1.3E-07** | 1.02 (1.01 – 1.02) | 5.4E-01 | 1.01 (1.00 – 1.02) | 1.02 (1.01 – 1.03) |
| 340.1 | Migraine with aura | AHI 3% | 0.99 (0.99 – 0.99) | **2.0E-07** | 0.99 (0.99 – 0.99) | 3.6E-01 | 0.99 (0.99 – 1.00) | 0.99 (0.98 – 0.99) |
| 504 | Other alveolar and parietoalveolar pneumonopathy | Per88 | 1.04 (1.02 – 1.05) | **2.6E-07** | 1.04 (1.02 – 1.05) | 9.7E-01 | 1.03 (1.01 – 1.05) | 1.05 (1.03 – 1.07) |
| 261 | Vitamin deficiency | AHI 3% | 0.99 (0.98 – 0.99) | **3.7E-07** | 0.99 (0.98 – 0.99) | 3.4E-05 | 1.00 (0.99 – 1.01) | 0.99 (0.98 – 0.99) |
| 505 | Other pulmonary inflamation or edema | AHI 3% | 1.02 (1.01 – 1.03) | **6.5E-07** | 1.02 (1.01 – 1.03) | 4.5E-01 | 1.02 (1.01 – 1.03) | 1.02 (1.01 – 1.03) |
| 797 | Shock | Per88 | 1.04 (1.03 – 1.06) | **1.0E-06** | 1.04 (1.02 – 1.06) | 3.1E-01 | 1.03 (1.01 – 1.05) | 1.05 (1.02 – 1.08) |
| 269 | Proteinuria | AHI 3% | 1.01 (1.01 – 1.02) | **1.3E-06** | 1.01 (1.01 – 1.02) | 4.2E-01 | 1.01 (1.00 – 1.01) | 1.02 (1.01 – 1.02) |
| 585.1 | Acute renal failure | AHI 3% | 1.01 (1.01 – 1.01) | **2.8E-06** | 1.01 (1.01 – 1.01) | 1.6E-01 | 1.01 (1.00 – 1.01) | 1.01 (1.00 – 1.02) |

| PheCode | Disease | Sleep statistic | Risk ratio | P-value | Risk ratio (adj pulmonary disorders) | Sleep statistic x sex interaction p-value | Risk ratio (Men) | Risk ratio (Women) |
|---|---|---|---|---|---|---|---|---|
| 389.4 | Tinnitus | Per88 | 0.98 (0.97 – 0.99) | **3.3E-06** | 0.98 (0.97 – 0.99) | 4.2E-01 | 0.98 (0.97 – 0.99) | 0.98 (0.97 – 0.99) |
| 539 | Bariatric surgery | AHI 3% | 0.99 (0.99 – 1.00) | **6.7E-06** | 0.99 (0.99 – 1.00) | 7.3E-03 | 1.00 (0.99 – 1.00) | 0.99 (0.99 – 1.00) |
| 563 | Constipation | AHI 3% | 0.99 (0.99 – 1.00) | **1.0E-05** | 0.99 (0.99 – 1.00) | 3.2E-01 | 0.99 (0.99 – 1.00) | 0.99 (0.99 – 1.00) |
| 300.9 | Posttraumatic stress disorder | AHI 3% | 0.99 (0.99 – 1.00) | **1.1E-05** | 0.99 (0.99 – 1.00) | 6.2E-01 | 0.98 (0.98 – 0.99) | 0.99 (0.99 – 1.00) |
| 288.2 | Elevated white blood cell count | Per88 | 1.02 (1.01 – 1.03) | **1.2E-05** | 1.02 (1.01 – 1.03) | 6.2E-01 | 1.02 (1.01 – 1.04) | 1.02 (1.01 – 1.04) |
| 425.1 | Primary/intrinsic cardiomyopathies | Per88 | 1.02 (1.01 – 1.04) | **1.7E-05** | 1.02 (1.01 – 1.04) | 7.3E-01 | 1.02 (1.01 – 1.03) | 1.03 (1.01 – 1.04) |
| 503 | Pulmonary congestion and hypostasis | AHI 3% | 1.01 (1.01 – 1.02) | **1.8E-05** | 1.01 (1.01 – 1.02) | 9.2E-01 | 1.01 (1.01 – 1.02) | 1.01 (1.00 – 1.02) |
| 340 | Migraine | Per88 | 0.99 (0.99 – 0.99) | **2.0E-05** | 0.99 (0.98 – 0.99) | 7.0E-01 | 0.99 (0.98 – 1.00) | 0.99 (0.99 – 1.00) |
| 512.8 | Cough | Per88 | 1.01 (1.01 – 1.01) | **2.5E-05** | 1.01 (1.00 – 1.01) | 3.2E-01 | 1.00 (1.00 – 1.01) | 1.01 (1.01 – 1.02) |
| 509.1 | Respiratory failure | AHI 3% | 1.01 (1.01 – 1.02) | **2.6E-05** | 1.01 (1.01 – 1.02) | 5.6E-01 | 1.02 (1.01 – 1.02) | 1.01 (1.00 – 1.02) |
| 428.2 | Heart failure NOS | AHI 3% | 1.01 (1.01 – 1.02) | **2.8E-05** | 1.01 (1.01 – 1.02) | 7.0E-01 | 1.01 (1.00 – 1.02) | 1.01 (1.00 – 1.02) |
| 041 | Bacterial infection NOS | Per88 | 1.02 (1.01 – 1.04) | **3.2E-05** | 1.02 (1.01 – 1.03) | 7.0E-02 | 1.04 (1.02 – 1.05) | 1.02 (1.00 – 1.03) |
| 694.2 | Other dyschromia | Per88 | 0.99 (0.98 – 0.99) | **3.3E-05** | 0.99 (0.98 – 0.99) | 2.8E-01 | 0.99 (0.98 – 1.00) | 0.98 (0.97 – 0.99) |
| 425.1 | Primary/intrinsic cardiomyopathies | AHI 3% | 1.01 (1.01 – 1.01) | **4.0E-05** | 1.01 (1.01 – 1.01) | 8.9E-01 | 1.01 (1.00 – 1.01) | 1.01 (1.00 – 1.02) |
| 136 | Other infectious and parasitic diseases | Per88 | 1.02 (1.01 – 1.02) | **5.4E-05** | 1.01 (1.01 – 1.02) | 1.4E-01 | 1.02 (1.01 – 1.04) | 1.01 (1.00 – 1.02) |
| 290.2 | Delirium due to conditions classified elsewhere | Per88 | 1.04 (1.02 – 1.05) | 7.5E-05 | 1.03 (1.01 – 1.05) | 2.8E-01 | 1.03 (1.01 – 1.05) | 1.04 (1.01 – 1.07) |
| 327.41 | Organic or persistent insomnia | AHI 3% | 0.99 (0.99 – 1.00) | 7.6E-05 | 0.99 (0.99 – 1.00) | 2.9E-01 | 0.99 (0.98 – 1.00) | 0.99 (0.98 – 0.99) |
| 276.14 | Hypopotassemia | Per88 | 1.02 (1.01 – 1.03) | 7.9E-05 | 1.02 (1.01 – 1.03) | 4.4E-01 | 1.02 (1.00 – 1.04) | 1.02 (1.01 – 1.04) |
| 414 | Other forms of chronic heart disease | Per88 | 1.03 (1.01 – 1.04) | 8.4E-05 | 1.03 (1.01 – 1.04) | 1.8E-01 | 1.02 (1.00 – 1.04) | 1.04 (1.01 – 1.06) |
| 745 | Pain in joint | AHI 3% | 1.00 (1.00 – 1.00) | 8.8E-05 | 1.00 (1.00 – 1.00) | 7.1E-03 | 1.00 (1.00 – 1.00) | 1.00 (1.00 – 1.00) |
| 286.9 | Abnormal coagulation profile | Per88 | 1.02 (1.01 – 1.03) | 1.3E-04 | 1.02 (1.01 – 1.03) | 5.8E-02 | 1.00 (0.99 – 1.02) | 1.03 (1.02 – 1.05) |
| 416 | Cardiomegaly | Per88 | 1.03 (1.01 – 1.04) | 1.5E-04 | 1.02 (1.01 – 1.04) | 8.3E-01 | 1.02 (1.01 – 1.04) | 1.03 (1.01 – 1.05) |
| 288.2 | Elevated white blood cell count | AHI 3% | 1.01 (1.00 – 1.01) | 1.7E-04 | 1.01 (1.00 – 1.01) | 1.3E-01 | 1.01 (1.01 – 1.02) | 1.00 (1.00 – 1.01) |
| 261 | Vitamin deficiency | Per88 | 0.97 (0.96 – 0.99) | 2.0E-04 | 0.97 (0.95 – 0.99) | 1.9E-01 | 0.97 (0.92 – 1.03) | 0.98 (0.97 – 1.00) |
| 350.1 | Abnormal involuntary movements | AHI 3% | 0.99 (0.99 – 1.00) | 2.6E-04 | 0.99 (0.99 – 1.00) | 6.9E-01 | 0.99 (0.99 – 1.00) | 0.99 (0.99 – 1.00) |
| 428.3 | Heart failure with reduced EF [Systolic or combined heart failure] | Per88 | 1.03 (1.01 – 1.05) | 2.7E-04 | 1.03 (1.01 – 1.04) | 2.5E-01 | 1.01 (0.99 – 1.04) | 1.05 (1.02 – 1.07) |
| 496.2 | Chronic bronchitis | Per88 | 1.03 (1.02 – 1.05) | 2.9E-04 | 1.03 (1.01 – 1.05) | 6.4E-02 | 1.02 (1.00 – 1.04) | 1.05 (1.02 – 1.08) |
| 274.11 | Gouty arthropathy | AHI 3% | 1.01 (1.00 – 1.02) | 3.2E-04 | 1.01 (1.00 – 1.02) | 1.5E-02 | 1.01 (1.00 – 1.01) | 1.02 (1.00 – 1.03) |
| 598.9 | Other nonspecific findings on examination of urine | AHI 3% | 0.99 (0.99 – 1.00) | 3.2E-04 | 0.99 (0.99 – 1.00) | 1.2E-01 | 1.00 (0.99 – 1.00) | 0.99 (0.98 – 0.99) |
| 278 | Overweight, obesity and other hyperalimentation | Per88 | 0.99 (0.99 – 1.00) | 3.4E-04 | 0.99 (0.99 – 1.00) | 1.0E+00 | 0.99 (0.98 – 1.00) | 0.99 (0.98 – 1.00) |
| 427.9 | Palpitations | AHI 3% | 1.00 (0.99 – 1.00) | 3.8E-04 | 1.00 (0.99 – 1.00) | 6.7E-01 | 1.00 (0.99 – 1.00) | 1.00 (0.99 – 1.00) |
| 250.7 | Diabetic retinopathy | AHI 3% | 1.01 (1.01 – 1.02) | 3.8E-04 | 1.01 (1.01 – 1.02) | 5.8E-01 | 1.02 (1.01 – 1.02) | 1.00 (0.99 – 1.02) |
| 760 | Back pain | AHI 3% | 1.00 (1.00 – 1.00) | 4.1E-04 | 1.00 (1.00 – 1.00) | 7.2E-01 | 1.00 (1.00 – 1.00) | 1.00 (0.99 – 1.00) |
| 726 | Peripheral enthesopathies and allied syndromes | AHI 3% | 1.00 (0.99 – 1.00) | 4.3E-04 | 1.00 (0.99 – 1.00) | 5.1E-01 | 1.00 (0.99 – 1.00) | 0.99 (0.99 – 1.00) |

| PheCode | Disease | Sleep statistic | Risk ratio | P-value | Risk ratio (adj pulmonary disorders) | Sleep statistic x sex interaction p-value | Risk ratio (Men) | Risk ratio (Women) |
|---|---|---|---|---|---|---|---|---|
| 411.3 | Angina pectoris | Per88 | 1.01 (1.01 – 1.02) | 4.4E-04 | 1.01 (1.00 – 1.02) | 8.5E-02 | 1.01 (1.00 – 1.02) | 1.02 (1.01 – 1.03) |
| 250.7 | Diabetic retinopathy | Per88 | 1.03 (1.01 – 1.05) | 5.0E-04 | 1.03 (1.01 – 1.05) | 1.1E-02 | 1.04 (1.02 – 1.06) | 1.01 (0.97 – 1.04) |
| 743.11 | Osteoporosis NOS | Per88 | 1.01 (1.01 – 1.02) | 5.4E-04 | 1.01 (1.00 – 1.02) | 2.6E-01 | 1.02 (1.00 – 1.04) | 1.00 (0.99 – 1.01) |
| 514.2 | Solitary pulmonary nodule | Per88 | 1.02 (1.01 – 1.03) | 5.5E-04 | 1.02 (1.00 – 1.03) | 4.9E-02 | 1.00 (0.99 – 1.02) | 1.03 (1.02 – 1.05) |
| 415 | Pulmonary heart disease | Per88 | 1.03 (1.01 – 1.04) | 5.6E-04 | 1.02 (1.01 – 1.04) | 4.9E-01 | 1.03 (1.00 – 1.06) | 1.03 (1.00 – 1.05) |
| 285 | Other anemias | Per88 | 1.01 (1.00 – 1.01) | 5.7E-04 | 1.01 (1.00 – 1.01) | 3.0E-02 | 1.01 (1.00 – 1.02) | 1.01 (1.00 – 1.02) |
| 278.4 | Abnormal weight gain | AHI 3% | 0.99 (0.99 – 1.00) | 6.2E-04 | 0.99 (0.99 – 1.00) | 2.4E-02 | 0.99 (0.99 – 1.00) | 0.99 (0.99 – 1.00) |
| 797 | Shock | AHI 3% | 1.02 (1.01 – 1.02) | 6.4E-04 | 1.02 (1.01 – 1.02) | 5.1E-01 | 1.01 (1.00 – 1.02) | 1.02 (1.00 – 1.04) |
| 327 | Sleep disorders | AHI 3% | 1.00 (0.99 – 1.00) | 6.7E-04 | 1.00 (0.99 – 1.00) | 1.2E-01 | 1.00 (0.99 – 1.00) | 0.99 (0.99 – 1.00) |
| 395.1 | Nonrheumatic mitral valve disorders | Per88 | 1.02 (1.01 – 1.03) | 7.1E-04 | 1.01 (1.00 – 1.02) | 3.2E-01 | 1.01 (1.00 – 1.03) | 1.02 (1.00 – 1.03) |
| 250.42 | Other abnormal glucose | Per88 | 1.01 (1.00 – 1.02) | 7.3E-04 | 1.01 (1.00 – 1.02) | 2.2E-01 | 1.01 (1.00 – 1.02) | 1.01 (1.00 – 1.02) |
| 530.9 | Heartburn | Per88 | 0.99 (0.98 – 0.99) | 7.4E-04 | 0.98 (0.97 – 0.99) | 6.5E-01 | 0.99 (0.97 – 1.00) | 0.98 (0.97 – 0.99) |
| 840 | Sprains and strains | AHI 3% | 1.00 (0.99 – 1.00) | 7.4E-04 | 1.00 (0.99 – 1.00) | 1.1E-01 | 1.00 (0.99 – 1.00) | 0.99 (0.99 – 1.00) |
| 512.8 | Cough | AHI 3% | 1.00 (1.00 – 1.00) | 7.7E-04 | 1.00 (1.00 – 1.00) | 1.5E-01 | 1.00 (1.00 – 1.00) | 1.00 (1.00 – 1.01) |
| 785 | Abdominal pain | AHI 3% | 1.00 (1.00 – 1.00) | 7.8E-04 | 1.00 (1.00 – 1.00) | 8.0E-01 | 1.00 (1.00 – 1.00) | 1.00 (1.00 – 1.00) |
| 427.3 | Other specified cardiac dysrhythmias | AHI 3% | 0.99 (0.99 – 1.00) | 8.3E-04 | 0.99 (0.99 – 1.00) | 8.7E-01 | 1.00 (0.99 – 1.00) | 1.00 (0.99 – 1.00) |
| 586 | Other disorders of the kidney and ureters | AHI 3% | 1.01 (1.00 – 1.01) | 8.5E-04 | 1.01 (1.00 – 1.01) | 8.4E-01 | 1.01 (1.00 – 1.01) | 1.01 (1.00 – 1.01) |
| 260 | Protein-calorie malnutrition | AHI 3% | 1.01 (1.00 – 1.02) | 9.5E-04 | 1.01 (1.00 – 1.02) | 2.1E-01 | 1.01 (1.00 – 1.02) | 1.00 (1.00 – 1.01) |
| 508 | Pulmonary collapse; interstitial and compensatory emphysema | AHI 3% | 1.01 (1.00 – 1.01) | 1.0E-03 | 1.01 (1.00 – 1.01) | 1.7E-01 | 1.01 (1.00 – 1.01) | 1.00 (1.00 – 1.01) |
| 428.1 | Congestive heart failure (CHF) NOS | AHI 3% | 1.01 (1.00 – 1.01) | 1.0E-03 | 1.01 (1.00 – 1.01) | 3.8E-01 | 1.01 (1.00 – 1.01) | 1.00 (1.00 – 1.01) |
| 558 | Noninfectious gastroenteritis | Per88 | 1.01 (1.01 – 1.02) | 1.1E-03 | 1.01 (1.00 – 1.02) | 4.1E-01 | 1.01 (1.00 – 1.02) | 1.02 (1.01 – 1.03) |
| 585.3 | Chronic renal failure [CKD] | Per88 | 1.02 (1.01 – 1.03) | 1.1E-03 | 1.02 (1.01 – 1.03) | 2.5E-01 | 1.01 (1.00 – 1.02) | 1.02 (1.01 – 1.04) |
| 216 | Benign neoplasm of skin | Per88 | 0.99 (0.99 – 1.00) | 1.2E-03 | 0.99 (0.99 – 1.00) | 6.7E-01 | 0.99 (0.98 – 1.00) | 0.99 (0.98 – 1.00) |
| 427.21 | Atrial fibrillation | Per88 | 1.01 (1.00 – 1.02) | 1.3E-03 | 1.01 (1.00 – 1.02) | 9.8E-02 | 1.01 (1.00 – 1.01) | 1.03 (1.01 – 1.04) |
| 415.11 | Pulmonary embolism and infarction, acute | Per88 | 1.02 (1.01 – 1.03) | 1.3E-03 | 1.01 (1.00 – 1.03) | 5.1E-01 | 1.01 (0.99 – 1.03) | 1.03 (1.01 – 1.04) |
| 271.3 | Intestinal disaccharidase deficiencies and disaccharide malabsorption | AHI 3% | 1.00 (1.00 – 1.01) | 1.3E-03 | 1.00 (1.00 – 1.01) | 9.0E-02 | 1.00 (1.00 – 1.01) | 1.00 (1.00 – 1.01) |
| 519.8 | Other diseases of respiratory system, NEC | Per88 | 1.02 (1.01 – 1.03) | 1.4E-03 | 1.01 (1.00 – 1.02) | 7.2E-01 | 1.01 (1.00 – 1.03) | 1.02 (1.00 – 1.03) |
| 443.9 | Peripheral vascular disease, unspecified | Per88 | 1.03 (1.01 – 1.04) | 1.5E-03 | 1.03 (1.01 – 1.04) | 6.7E-01 | 1.02 (1.00 – 1.04) | 1.03 (1.01 – 1.06) |
| 274.1 | Gout | AHI 3% | 1.01 (1.00 – 1.01) | 1.6E-03 | 1.01 (1.00 – 1.01) | 1.7E-03 | 1.00 (1.00 – 1.01) | 1.02 (1.00 – 1.03) |
| 272.12 | Hyperglyceridemia | AHI 3% | 1.01 (1.00 – 1.01) | 1.6E-03 | 1.01 (1.00 – 1.01) | 8.5E-01 | 1.01 (1.00 – 1.01) | 1.01 (1.00 – 1.02) |
| 357 | Inflammatory and toxic neuropathy | Per88 | 1.01 (1.00 – 1.02) | 1.7E-03 | 1.01 (1.00 – 1.02) | 3.6E-01 | 1.01 (1.00 – 1.02) | 1.01 (1.00 – 1.02) |
| 743.11 | Osteoporosis NOS | AHI 3% | 1.00 (1.00 – 1.01) | 1.8E-03 | 1.00 (1.00 – 1.01) | 7.1E-01 | 1.00 (0.99 – 1.01) | 1.00 (1.00 – 1.00) |
| 560.4 | Other intestinal obstruction | AHI 3% | 1.01 (1.00 – 1.02) | 1.8E-03 | 1.01 (1.00 – 1.02) | 3.7E-01 | 1.01 (1.01 – 1.02) | 1.00 (1.00 – 1.01) |
| 728.7 | Fasciitis | Per88 | 0.98 (0.97 – 0.99) | 1.8E-03 | 0.98 (0.97 – 0.99) | 5.7E-01 | 0.99 (0.98 – 1.01) | 0.98 (0.96 – 0.99) |

| PheCode | Disease | Sleep statistic | Risk ratio | P-value | Risk ratio (adj pulmonary disorders) | Sleep statistic x sex interaction p-value | Risk ratio (Men) | Risk ratio (Women) |
|---|---|---|---|---|---|---|---|---|
| 527.7 | Disturbance of salivary secretion | Per88 | 1.02 (1.01 – 1.03) | 1.9E-03 | 1.02 (1.01 – 1.03) | 2.5E-01 | 1.01 (0.99 – 1.04) | 1.02 (1.01 – 1.04) |
| 585.3 | Chronic renal failure [CKD] | AHI 3% | 1.01 (1.00 – 1.01) | 1.9E-03 | 1.01 (1.00 – 1.01) | 7.5E-01 | 1.00 (1.00 – 1.01) | 1.01 (1.00 – 1.01) |
| 292.3 | Memory loss | AHI 3% | 1.00 (0.99 – 1.00) | 2.0E-03 | 1.00 (0.99 – 1.00) | 7.9E-01 | 1.00 (0.99 – 1.00) | 0.99 (0.99 – 1.00) |
| 250.42 | Other abnormal glucose | AHI 3% | 1.00 (1.00 – 1.01) | 2.1E-03 | 1.00 (1.00 – 1.01) | 3.7E-02 | 1.00 (1.00 – 1.01) | 1.00 (1.00 – 1.01) |
| 228.1 | Hemangioma of skin and subcutaneous tissue | Per88 | 0.98 (0.97 – 0.99) | 2.4E-03 | 0.98 (0.97 – 0.99) | 1.7E-01 | 0.99 (0.97 – 1.00) | 0.99 (0.97 – 1.00) |
| 938.2 | Chronic dermatitis due to solar radiation | Per88 | 0.98 (0.98 – 0.99) | 2.4E-03 | 0.99 (0.98 – 0.99) | 8.5E-01 | 0.99 (0.98 – 1.00) | 0.98 (0.97 – 1.00) |
| 798 | Malaise and fatigue | AHI 3% | 1.00 (1.00 – 1.00) | 2.6E-03 | 1.00 (1.00 – 1.00) | 1.1E-01 | 1.00 (1.00 – 1.00) | 1.00 (0.99 – 1.00) |
| 539 | Bariatric surgery | Per88 | 0.99 (0.98 – 1.00) | 2.6E-03 | 0.99 (0.98 – 1.00) | 5.8E-01 | 0.99 (0.98 – 1.01) | 0.99 (0.98 – 1.00) |
| 427.7 | Tachycardia NOS | AHI 3% | 1.00 (1.00 – 1.01) | 2.7E-03 | 1.00 (1.00 – 1.01) | 5.5E-03 | 1.01 (1.00 – 1.01) | 1.00 (1.00 – 1.01) |
| 272.1 | Hyperlipidemia | AHI 3% | 1.00 (1.00 – 1.00) | 2.8E-03 | 1.00 (1.00 – 1.00) | 4.1E-03 | 1.00 (1.00 – 1.00) | 1.00 (1.00 – 1.01) |
| 532 | Dysphagia | Per88 | 1.01 (1.00 – 1.02) | 3.1E-03 | 1.01 (1.00 – 1.01) | 2.9E-01 | 1.01 (1.00 – 1.02) | 1.01 (1.00 – 1.02) |
| 571.5 | Other chronic nonalcoholic liver disease | AHI 3% | 1.01 (1.00 – 1.01) | 3.3E-03 | 1.01 (1.00 – 1.01) | 1.9E-01 | 1.01 (1.00 – 1.01) | 1.00 (1.00 – 1.01) |
| 456 | Chronic venous insufficiency [CVI] | Per88 | 1.02 (1.01 – 1.03) | 3.3E-03 | 1.02 (1.01 – 1.03) | 6.0E-02 | 1.02 (1.00 – 1.04) | 1.02 (1.00 – 1.03) |
| 300.11 | Generalized anxiety disorder | AHI 3% | 1.00 (0.99 – 1.00) | 3.3E-03 | 1.00 (0.99 – 1.00) | 1.8E-02 | 0.99 (0.99 – 1.00) | 1.00 (1.00 – 1.00) |
| 798.1 | Chronic fatigue syndrome | AHI 3% | 1.00 (0.99 – 1.00) | 3.3E-03 | 1.00 (0.99 – 1.00) | 9.5E-01 | 0.99 (0.99 – 1.00) | 1.00 (0.99 – 1.00) |
| 480.11 | Pneumococcal pneumonia | Per88 | 1.02 (1.01 – 1.04) | 3.5E-03 | 1.02 (1.00 – 1.03) | 2.9E-01 | 1.01 (0.98 – 1.03) | 1.04 (1.02 – 1.06) |
| 291.4 | Specific nonpsychotic mental disorders due to brain damage | Per88 | 0.98 (0.96 – 0.99) | 4.9E-03 | 0.98 (0.96 – 0.99) | 8.2E-01 | 0.98 (0.96 – 1.00) | 0.98 (0.97 – 1.00) |
| 564 | Functional digestive disorders | Per88 | 1.02 (1.01 – 1.03) | 5.4E-03 | 1.02 (1.00 – 1.03) | 4.1E-01 | 1.00 (0.98 – 1.02) | 1.03 (1.01 – 1.05) |
| 480 | Pneumonia | AHI 3% | 1.00 (1.00 – 1.01) | 5.7E-03 | 1.00 (1.00 – 1.01) | 3.2E-01 | 1.01 (1.00 – 1.01) | 1.00 (1.00 – 1.01) |
| 411.2 | Myocardial infarction | Per88 | 1.01 (1.00 – 1.02) | 5.7E-03 | 1.01 (1.00 – 1.02) | 9.2E-01 | 1.01 (1.00 – 1.02) | 1.02 (1.01 – 1.04) |
| 339 | Other headache syndromes | Per88 | 0.99 (0.99 – 1.00) | 5.7E-03 | 0.99 (0.99 – 1.00) | 2.2E-01 | 0.99 (0.98 – 1.00) | 0.99 (0.99 – 1.00) |
| 496.21 | Obstructive chronic bronchitis | AHI 3% | 0.99 (0.98 – 1.00) | 6.2E-03 | 0.99 (0.98 – 1.00) | 1.3E-01 | 0.99 (0.98 – 1.00) | 0.99 (0.97 – 1.00) |
| 287.3 | Thrombocytopenia | Per88 | 1.02 (1.00 – 1.03) | 6.3E-03 | 1.02 (1.00 – 1.03) | 5.1E-01 | 1.02 (1.01 – 1.03) | 1.01 (0.99 – 1.03) |
| 292.2 | Mild cognitive impairment | Per88 | 0.98 (0.96 – 0.99) | 6.9E-03 | 0.98 (0.97 – 1.00) | 4.3E-01 | 0.99 (0.98 – 1.00) | 0.98 (0.95 – 1.00) |
| 375.1 | Dry eyes | AHI 3% | 1.00 (0.99 – 1.00) | 7.0E-03 | 1.00 (0.99 – 1.00) | 4.8E-02 | 1.00 (0.99 – 1.00) | 0.99 (0.99 – 1.00) |
| 575.7 | Other disorders of gallbladder | Per88 | 1.02 (1.01 – 1.03) | 7.2E-03 | 1.02 (1.00 – 1.03) | 9.1E-01 | 1.02 (1.00 – 1.04) | 1.02 (1.00 – 1.04) |
| 276.14 | Hypopotassemia | AHI 3% | 1.01 (1.00 – 1.01) | 7.3E-03 | 1.01 (1.00 – 1.01) | 7.7E-02 | 1.01 (1.00 – 1.02) | 1.00 (1.00 – 1.01) |
| 726.1 | Enthesopathy | AHI 3% | 1.00 (0.99 – 1.00) | 7.7E-03 | 1.00 (0.99 – 1.00) | 2.0E-01 | 0.99 (0.99 – 1.00) | 1.00 (0.99 – 1.00) |
| 302.1 | Decreased libido | Per88 | 0.98 (0.96 – 0.99) | 7.9E-03 | 0.98 (0.96 – 1.00) | 2.4E-01 | 0.98 (0.96 – 1.00) | 0.97 (0.94 – 1.00) |
| 728.7 | Fasciitis | AHI 3% | 1.00 (0.99 – 1.00) | 7.9E-03 | 1.00 (0.99 – 1.00) | 9.2E-01 | 1.00 (0.99 – 1.00) | 0.99 (0.99 – 1.00) |
| 112 | Candidiasis | Per88 | 1.01 (1.00 – 1.02) | 8.7E-03 | 1.01 (1.00 – 1.02) | 3.3E-02 | 1.03 (1.01 – 1.04) | 1.01 (1.00 – 1.02) |
| 528 | Diseases of the oral soft tissues, excluding lesions specific for gingiva and tongue | AHI 3% | 0.99 (0.99 – 1.00) | 8.8E-03 | 0.99 (0.99 – 1.00) | 1.2E-01 | 0.99 (0.99 – 1.00) | 0.99 (0.99 – 1.00) |
| 165.1 | Cancer of bronchus; lung | Per88 | 1.02 (1.01 – 1.04) | 8.8E-03 | 1.02 (1.00 – 1.04) | 3.2E-01 | 0.99 (0.96 – 1.02) | 1.04 (1.02 – 1.06) |
| 261.2 | Vitamin B-complex deficiencies | AHI 3% | 0.99 (0.99 – 1.00) | 8.8E-03 | 0.99 (0.99 – 1.00) | 7.7E-01 | 0.99 (0.99 – 1.00) | 0.99 (0.98 – 1.00) |

| PheCode | Disease | Sleep statistic | Risk ratio | P-value | Risk ratio (adj pulmonary disorders) | Sleep statistic x sex interaction p-value | Risk ratio (Men) | Risk ratio (Women) |
|---|---|---|---|---|---|---|---|---|
| 454.11 | Varicose veins of lower extremity, symptomtic | AHI 3% | 0.99 (0.99 – 1.00) | 9.0E-03 | 0.99 (0.99 – 1.00) | 8.0E-01 | 0.99 (0.98 – 0.99) | 1.00 (0.99 – 1.00) |
| 495 | Asthma | AHI 3% | 1.00 (1.00 – 1.00) | 9.5E-03 | 1.00 (1.00 – 1.00) | 2.0E-01 | 1.00 (0.99 – 1.00) | 1.00 (1.00 – 1.00) |
| 386.9 | Dizziness and giddiness (Light-headedness and vertigo) | Per88 | 0.99 (0.99 – 1.00) | 1.0E-02 | 0.99 (0.99 – 1.00) | 4.6E-01 | 0.99 (0.99 – 1.00) | 0.99 (0.99 – 1.00) |
| 701.2 | Scar conditions and fibrosis of skin | Per88 | 0.99 (0.98 – 1.00) | 1.0E-02 | 0.99 (0.98 – 1.00) | 7.8E-01 | 0.99 (0.98 – 1.00) | 0.99 (0.98 – 1.00) |
| 418.1 | Precordial pain | Per88 | 1.01 (1.00 – 1.02) | 1.1E-02 | 1.01 (1.00 – 1.02) | 3.7E-01 | 1.00 (0.99 – 1.02) | 1.02 (1.01 – 1.04) |
| 340.1 | Migraine with aura | Per88 | 0.99 (0.98 – 1.00) | 1.1E-02 | 0.99 (0.98 – 1.00) | 6.5E-01 | 0.98 (0.96 – 1.01) | 0.99 (0.98 – 1.00) |
| 519.8 | Other diseases of respiratory system, NEC | AHI 3% | 1.01 (1.00 – 1.01) | 1.1E-02 | 1.01 (1.00 – 1.01) | 1.8E-01 | 1.01 (1.00 – 1.01) | 1.00 (1.00 – 1.01) |
| 480.11 | Pneumococcal pneumonia | AHI 3% | 1.01 (1.00 – 1.02) | 1.1E-02 | 1.01 (1.00 – 1.02) | 5.2E-01 | 1.01 (1.00 – 1.02) | 1.01 (1.00 – 1.02) |
| 306 | Other mental disorder | Per88 | 1.01 (1.00 – 1.01) | 1.1E-02 | 1.01 (1.00 – 1.01) | 8.4E-01 | 1.01 (1.00 – 1.02) | 1.00 (1.00 – 1.01) |
| 296 | Mood disorders | AHI 3% | 1.00 (1.00 – 1.01) | 1.1E-02 | 1.00 (1.00 – 1.01) | 3.6E-01 | 1.01 (1.00 – 1.01) | 1.00 (1.00 – 1.01) |
| 458.9 | Hypotension NOS | AHI 3% | 1.01 (1.00 – 1.01) | 1.2E-02 | 1.01 (1.00 – 1.01) | 2.0E-02 | 1.01 (1.00 – 1.01) | 1.00 (0.99 – 1.01) |
| 368 | Visual disturbances | AHI 3% | 1.00 (0.99 – 1.00) | 1.2E-02 | 1.00 (0.99 – 1.00) | 4.4E-01 | 1.00 (0.99 – 1.00) | 0.99 (0.99 – 1.00) |
| 381.1 | Otitis media | Per88 | 1.02 (1.00 – 1.03) | 1.2E-02 | 1.02 (1.00 – 1.03) | 1.6E-01 | 1.02 (1.00 – 1.04) | 1.01 (1.00 – 1.03) |
| 840.3 | Joint/ligament sprain | AHI 3% | 1.00 (0.99 – 1.00) | 1.3E-02 | 1.00 (0.99 – 1.00) | 6.5E-01 | 0.99 (0.99 – 1.00) | 1.00 (0.99 – 1.00) |
| 278.1 | Obesity | AHI 3% | 1.00 (1.00 – 1.00) | 1.3E-02 | 1.00 (1.00 – 1.00) | 5.2E-04 | 1.00 (1.00 – 1.00) | 1.00 (1.00 – 1.00) |
| 411.8 | Other chronic ischemic heart disease, unspecified | AHI 3% | 1.01 (1.00 – 1.02) | 1.4E-02 | 1.01 (1.00 – 1.02) | 8.1E-02 | 1.01 (1.00 – 1.01) | 1.02 (1.01 – 1.04) |
| 770 | Myalgia and myositis unspecified | AHI 3% | 1.00 (0.99 – 1.00) | 1.4E-02 | 1.00 (0.99 – 1.00) | 1.5E-01 | 1.00 (0.99 – 1.01) | 0.99 (0.99 – 1.00) |
| 427.22 | Atrial flutter | Per88 | 1.02 (1.00 – 1.03) | 1.5E-02 | 1.02 (1.00 – 1.03) | 8.0E-02 | 1.01 (1.00 – 1.03) | 1.04 (1.02 – 1.07) |
| 578.9 | Hemorrhage of gastrointestinal tract | Per88 | 1.02 (1.00 – 1.03) | 1.6E-02 | 1.01 (1.00 – 1.03) | 6.1E-01 | 1.01 (1.00 – 1.03) | 1.01 (0.99 – 1.04) |
| 531.4 | Peptic ulcer, site unspecified | Per88 | 1.02 (1.00 – 1.04) | 1.6E-02 | 1.02 (1.00 – 1.04) | 4.5E-01 | 1.02 (1.00 – 1.05) | 1.02 (0.99 – 1.04) |
| 317.1 | Alcoholism | AHI 3% | 1.01 (1.00 – 1.01) | 1.7E-02 | 1.01 (1.00 – 1.01) | 6.5E-01 | 1.00 (1.00 – 1.01) | 1.01 (1.00 – 1.02) |
| 279 | Disorders involving the immune mechanism | Per88 | 1.02 (1.00 – 1.03) | 1.7E-02 | 1.01 (1.00 – 1.03) | 9.5E-01 | 1.02 (1.00 – 1.03) | 1.02 (1.00 – 1.03) |
| 416 | Cardiomegaly | AHI 3% | 1.01 (1.00 – 1.01) | 1.7E-02 | 1.01 (1.00 – 1.01) | 2.1E-02 | 1.00 (1.00 – 1.01) | 1.02 (1.01 – 1.03) |
| 278.11 | Morbid obesity | AHI 3% | 1.00 (1.00 – 1.00) | 1.7E-02 | 1.00 (1.00 – 1.00) | 2.4E-02 | 1.00 (1.00 – 1.01) | 1.00 (1.00 – 1.00) |
| 516.1 | Hemoptysis | Per88 | 1.02 (1.00 – 1.04) | 1.8E-02 | 1.02 (1.00 – 1.03) | 3.2E-01 | 1.03 (1.00 – 1.05) | 1.02 (0.99 – 1.04) |
| 250.1 | Type 1 diabetes | AHI 3% | 1.01 (1.00 – 1.02) | 1.9E-02 | 1.01 (1.00 – 1.02) | 9.4E-02 | 1.00 (0.99 – 1.01) | 1.02 (1.01 – 1.03) |
| 420.2 | Pericarditis | AHI 3% | 1.01 (1.00 – 1.01) | 1.9E-02 | 1.01 (1.00 – 1.01) | 5.5E-01 | 1.01 (1.00 – 1.02) | 1.01 (1.00 – 1.02) |
| 172.21 | Basal cell carcinoma | Per88 | 0.98 (0.97 – 1.00) | 2.0E-02 | 0.98 (0.97 – 1.00) | 5.8E-01 | 0.99 (0.98 – 1.01) | 0.98 (0.96 – 1.00) |
| 452.2 | Deep vein thrombosis [DVT] | AHI 3% | 0.99 (0.99 – 1.00) | 2.1E-02 | 0.99 (0.99 – 1.00) | 4.3E-01 | 0.99 (0.99 – 1.00) | 0.99 (0.99 – 1.00) |
| 564.1 | Irritable Bowel Syndrome | AHI 3% | 1.00 (0.99 – 1.00) | 2.1E-02 | 1.00 (0.99 – 1.00) | 1.3E-01 | 0.99 (0.99 – 1.00) | 1.00 (0.99 – 1.00) |
| 599.9 | Other abnormality of urination | AHI 3% | 1.00 (0.99 – 1.00) | 2.2E-02 | 1.00 (0.99 – 1.00) | 5.7E-02 | 1.00 (0.99 – 1.00) | 1.00 (0.99 – 1.00) |
| 452 | Other venous embolism and thrombosis | AHI 3% | 1.00 (0.99 – 1.00) | 2.2E-02 | 1.00 (0.99 – 1.00) | 1.5E-01 | 1.00 (0.99 – 1.00) | 0.99 (0.99 – 1.00) |
| 427.12 | Paroxysmal ventricular tachycardia | AHI 3% | 1.01 (1.00 – 1.01) | 2.2E-02 | 1.01 (1.00 – 1.01) | 6.3E-01 | 1.01 (1.00 – 1.01) | 1.01 (0.99 – 1.02) |
| 292 | Neurological disorders | Per88 | 0.99 (0.98 – 1.00) | 2.3E-02 | 0.99 (0.98 – 1.00) | 2.9E-01 | 0.99 (0.98 – 1.01) | 0.98 (0.97 – 1.00) |

| PheCode | Disease | Sleep statistic | Risk ratio | P-value | Risk ratio (adj pulmonary disorders) | Sleep statistic x sex interaction p-value | Risk ratio (Men) | Risk ratio (Women) |
|---|---|---|---|---|---|---|---|---|
| 807 | Fracture of ribs | AHI 3% | 1.01 (1.00 – 1.02) | 2.3E-02 | 1.01 (1.00 – 1.02) | 8.7E-01 | 1.01 (1.00 – 1.02) | 1.01 (1.00 – 1.02) |
| 772.3 | Muscle weakness | AHI 3% | 0.99 (0.99 – 1.00) | 2.3E-02 | 0.99 (0.99 – 1.00) | 5.9E-01 | 0.99 (0.98 – 1.00) | 1.00 (0.99 – 1.00) |
| 577.3 | Cyst and pseudocyst of pancreas | Per88 | 1.02 (1.00 – 1.04) | 2.3E-02 | 1.02 (1.00 – 1.03) | 8.8E-01 | 1.02 (1.00 – 1.05) | 1.02 (0.99 – 1.04) |
| 290.2 | Delirium due to conditions classified elsewhere | AHI 3% | 1.01 (1.00 – 1.02) | 2.4E-02 | 1.01 (1.00 – 1.02) | 5.2E-01 | 1.01 (1.00 – 1.01) | 1.01 (1.00 – 1.03) |
| 427.11 | Paroxysmal supraventricular tachycardia | Per88 | 1.01 (1.00 – 1.02) | 2.5E-02 | 1.01 (1.00 – 1.02) | 2.3E-01 | 1.00 (0.98 – 1.02) | 1.02 (1.01 – 1.04) |
| 427.6 | Premature beats | Per88 | 1.01 (1.00 – 1.02) | 2.5E-02 | 1.01 (1.00 – 1.02) | 4.1E-01 | 1.01 (1.00 – 1.02) | 1.01 (0.99 – 1.03) |
| 130.1 | Lyme disease | Per88 | 0.98 (0.96 – 1.00) | 2.5E-02 | 0.98 (0.96 – 1.00) | 5.3E-01 | 0.98 (0.96 – 1.00) | 0.98 (0.96 – 1.01) |
| 474.2 | Chronic tonsillitis and adenoiditis | AHI 3% | 1.01 (1.00 – 1.01) | 2.6E-02 | 1.01 (1.00 – 1.01) | 1.9E-02 | 1.00 (0.99 – 1.01) | 1.01 (1.00 – 1.02) |
| 550.4 | Umbilical hernia | Per88 | 1.01 (1.00 – 1.03) | 2.6E-02 | 1.01 (1.00 – 1.03) | 1.9E-01 | 1.01 (1.00 – 1.03) | 1.01 (0.99 – 1.03) |
| 368 | Visual disturbances | Per88 | 0.99 (0.98 – 1.00) | 2.6E-02 | 0.99 (0.98 – 1.00) | 1.7E-02 | 0.98 (0.97 – 0.99) | 1.00 (0.99 – 1.01) |
| 306.9 | Tension headache | Per88 | 0.99 (0.98 – 1.00) | 2.7E-02 | 0.99 (0.98 – 1.00) | 2.6E-02 | 0.99 (0.97 – 1.01) | 0.99 (0.98 – 1.00) |
| 414 | Other forms of chronic heart disease | AHI 3% | 1.01 (1.00 – 1.01) | 2.7E-02 | 1.01 (1.00 – 1.01) | 6.1E-01 | 1.01 (1.00 – 1.01) | 1.00 (0.99 – 1.01) |
| 338.2 | Chronic pain | AHI 3% | 1.00 (1.00 – 1.00) | 2.7E-02 | 1.00 (1.00 – 1.00) | 4.8E-02 | 1.00 (0.99 – 1.00) | 1.00 (1.00 – 1.00) |
| 782.3 | Edema | AHI 3% | 1.00 (1.00 – 1.01) | 2.8E-02 | 1.00 (1.00 – 1.01) | 5.9E-03 | 1.00 (1.00 – 1.01) | 1.00 (1.00 – 1.01) |
| 496 | Chronic airway obstruction | AHI 3% | 0.99 (0.99 – 1.00) | 2.9E-02 | 0.99 (0.99 – 1.00) | 8.1E-01 | 1.00 (0.99 – 1.00) | 0.99 (0.98 – 1.00) |
| 473 | Diseases of the larynx and vocal cords | AHI 3% | 1.01 (1.00 – 1.01) | 3.0E-02 | 1.01 (1.00 – 1.01) | 3.4E-01 | 1.01 (1.00 – 1.01) | 1.00 (1.00 – 1.01) |
| 465 | Acute upper respiratory infections of multiple or unspecified sites | AHI 3% | 1.00 (1.00 – 1.00) | 3.0E-02 | 1.00 (1.00 – 1.00) | 3.0E-01 | 1.00 (1.00 – 1.00) | 1.00 (1.00 – 1.01) |
| 269 | Proteinuria | Per88 | 1.01 (1.00 – 1.03) | 3.1E-02 | 1.01 (1.00 – 1.03) | 2.5E-01 | 1.02 (1.00 – 1.04) | 1.01 (0.98 – 1.03) |
| 300.4 | Dysthymic disorder | AHI 3% | 1.00 (0.99 – 1.00) | 3.2E-02 | 1.00 (0.99 – 1.00) | 3.8E-02 | 0.99 (0.98 – 1.00) | 1.00 (1.00 – 1.01) |
| 565.1 | Anal and rectal polyp | AHI 3% | 1.00 (1.00 – 1.01) | 3.2E-02 | 1.00 (1.00 – 1.01) | 1.6E-01 | 1.00 (1.00 – 1.01) | 1.01 (1.00 – 1.02) |
| 427.3 | Other specified cardiac dysrhythmias | Per88 | 0.99 (0.98 – 1.00) | 3.2E-02 | 0.99 (0.98 – 1.00) | 1.7E-01 | 0.99 (0.98 – 1.00) | 1.00 (0.99 – 1.01) |
| 041 | Bacterial infection NOS | AHI 3% | 1.01 (1.00 – 1.01) | 3.3E-02 | 1.01 (1.00 – 1.01) | 1.5E-02 | 1.01 (1.01 – 1.02) | 1.00 (0.99 – 1.01) |
| 250.1 | Type 1 diabetes | Per88 | 1.02 (1.00 – 1.04) | 3.4E-02 | 1.02 (1.00 – 1.04) | 1.1E-01 | 1.00 (0.98 – 1.03) | 1.03 (1.01 – 1.06) |
| 772.2 | Spasm of muscle | AHI 3% | 1.00 (0.99 – 1.00) | 3.5E-02 | 1.00 (0.99 – 1.00) | 2.0E-01 | 1.00 (0.99 – 1.00) | 0.99 (0.99 – 1.00) |
| 569 | Other disorders of intestine | Per88 | 1.01 (1.00 – 1.01) | 3.5E-02 | 1.01 (1.00 – 1.01) | 5.7E-01 | 1.01 (1.00 – 1.02) | 1.01 (1.00 – 1.02) |
| 296 | Mood disorders | Per88 | 1.01 (1.00 – 1.01) | 3.5E-02 | 1.01 (1.00 – 1.01) | 8.1E-01 | 1.01 (1.00 – 1.02) | 1.01 (1.00 – 1.02) |
| 473 | Diseases of the larynx and vocal cords | Per88 | 1.01 (1.00 – 1.02) | 3.5E-02 | 1.01 (1.00 – 1.02) | 5.4E-01 | 1.01 (0.99 – 1.03) | 1.01 (1.00 – 1.03) |
| 557 | Intestinal malabsorption (non-celiac) | AHI 3% | 0.99 (0.99 – 1.00) | 3.6E-02 | 0.99 (0.99 – 1.00) | 3.0E-02 | 1.00 (0.99 – 1.01) | 0.99 (0.98 – 1.00) |
| 272.13 | Mixed hyperlipidemia | AHI 3% | 1.00 (1.00 – 1.01) | 3.6E-02 | 1.00 (1.00 – 1.01) | 9.5E-01 | 1.00 (1.00 – 1.01) | 1.00 (1.00 – 1.01) |
| 327.41 | Organic or persistent insomnia | Per88 | 0.99 (0.98 – 1.00) | 3.6E-02 | 0.99 (0.98 – 1.00) | 8.4E-01 | 0.99 (0.98 – 1.01) | 0.98 (0.97 – 1.00) |
| 476 | Allergic rhinitis | Per88 | 0.99 (0.99 – 1.00) | 3.6E-02 | 0.99 (0.99 – 1.00) | 5.5E-01 | 0.99 (0.98 – 1.00) | 1.00 (0.99 – 1.00) |
| 558 | Noninfectious gastroenteritis | AHI 3% | 1.00 (1.00 – 1.01) | 3.6E-02 | 1.00 (1.00 – 1.01) | 1.8E-01 | 1.01 (1.00 – 1.01) | 1.00 (1.00 – 1.01) |
| 477 | Epistaxis or throat hemorrhage | Per88 | 1.01 (1.00 – 1.03) | 3.6E-02 | 1.01 (1.00 – 1.03) | 8.9E-01 | 1.01 (0.99 – 1.03) | 1.02 (1.00 – 1.04) |
| 277.51 | Lipoprotein disorders | Per88 | 0.98 (0.96 – 1.00) | 3.7E-02 | 0.98 (0.96 – 1.00) | 8.8E-01 | 0.97 (0.95 – 1.00) | 0.98 (0.95 – 1.02) |

| PheCode | Disease | Sleep statistic | Risk ratio | P-value | Risk ratio (adj pulmonary disorders) | Sleep statistic x sex interaction p-value | Risk ratio (Men) | Risk ratio (Women) |
|---|---|---|---|---|---|---|---|---|
| 276.6 | Fluid overload | AHI 3% | 1.01 (1.00 – 1.01) | 3.7E-02 | 1.01 (1.00 – 1.01) | 1.4E-01 | 1.01 (1.00 – 1.02) | 1.00 (0.99 – 1.01) |
| 527.7 | Disturbance of salivary secretion | AHI 3% | 1.01 (1.00 – 1.01) | 3.8E-02 | 1.01 (1.00 – 1.01) | 9.8E-01 | 1.01 (1.00 – 1.02) | 1.01 (1.00 – 1.02) |
| 472 | Chronic pharyngitis and nasopharyngitis | AHI 3% | 1.00 (1.00 – 1.01) | 3.9E-02 | 1.00 (1.00 – 1.01) | 9.7E-01 | 1.00 (1.00 – 1.01) | 1.00 (1.00 – 1.01) |
| 550.4 | Umbilical hernia | AHI 3% | 1.01 (1.00 – 1.01) | 4.0E-02 | 1.01 (1.00 – 1.01) | 1.1E-01 | 1.01 (1.00 – 1.01) | 1.00 (0.99 – 1.01) |
| 452.2 | Deep vein thrombosis [DVT] | Per88 | 1.01 (1.00 – 1.02) | 4.2E-02 | 1.01 (1.00 – 1.02) | 3.3E-01 | 1.00 (0.98 – 1.02) | 1.02 (1.01 – 1.04) |
| 704.8 | Other specified diseases of hair and hair follicles | Per88 | 0.99 (0.98 – 1.00) | 4.3E-02 | 0.99 (0.98 – 1.00) | 3.3E-01 | 0.98 (0.97 – 1.00) | 1.00 (0.98 – 1.01) |
| 722.1 | Displacement of intervertebral disc | AHI 3% | 1.00 (0.99 – 1.00) | 4.4E-02 | 1.00 (0.99 – 1.00) | 6.1E-01 | 1.00 (0.99 – 1.00) | 1.00 (0.99 – 1.00) |
| 747.11 | Cardiac shunt/ heart septal defect | AHI 3% | 0.99 (0.99 – 1.00) | 4.5E-02 | 0.99 (0.99 – 1.00) | 9.1E-01 | 0.99 (0.98 – 1.00) | 1.00 (0.99 – 1.01) |
| 789 | Nausea and vomiting | AHI 3% | 1.00 (1.00 – 1.00) | 4.6E-02 | 1.00 (1.00 – 1.00) | 5.8E-03 | 1.00 (1.00 – 1.00) | 1.00 (0.99 – 1.00) |
| 196 | Radiotherapy | AHI 3% | 1.00 (1.00 – 1.01) | 4.6E-02 | 1.00 (1.00 – 1.01) | 3.4E-01 | 1.01 (1.00 – 1.01) | 1.00 (1.00 – 1.01) |
| 427.61 | Supraventricular premature beats | Per88 | 1.02 (1.00 – 1.03) | 4.6E-02 | 1.01 (1.00 – 1.03) | 5.3E-01 | 1.01 (0.99 – 1.03) | 1.02 (0.99 – 1.04) |
| 278.4 | Abnormal weight gain | Per88 | 0.99 (0.98 – 1.00) | 4.8E-02 | 0.99 (0.98 – 1.00) | 9.5E-03 | 1.00 (0.98 – 1.02) | 0.99 (0.98 – 1.00) |
| 452 | Other venous embolism and thrombosis | Per88 | 1.01 (1.00 – 1.02) | 4.8E-02 | 1.01 (1.00 – 1.02) | 3.1E-01 | 1.00 (0.99 – 1.02) | 1.02 (1.00 – 1.03) |
| 292.3 | Memory loss | Per88 | 0.99 (0.98 – 1.00) | 4.8E-02 | 0.99 (0.98 – 1.00) | 7.0E-01 | 0.99 (0.98 – 1.00) | 0.99 (0.98 – 1.00) |
| 687.1 | Rash and other nonspecific skin eruption | AHI 3% | 1.00 (1.00 – 1.00) | 4.9E-02 | 1.00 (1.00 – 1.00) | 2.1E-01 | 1.00 (0.99 – 1.00) | 1.00 (1.00 – 1.00) |
| 455 | Hemorrhoids | AHI 3% | 1.00 (1.00 – 1.00) | 5.0E-02 | 1.00 (1.00 – 1.00) | 7.6E-01 | 1.00 (0.99 – 1.00) | 1.00 (0.99 – 1.00) |
| 401.21 | Hypertensive heart disease | AHI 3% | 1.01 (1.00 – 1.01) | 5.0E-02 | 1.01 (1.00 – 1.01) | 5.8E-01 | 1.00 (1.00 – 1.01) | 1.01 (1.00 – 1.02) |
| 807 | Fracture of ribs | Per88 | 1.02 (1.00 – 1.03) | 5.0E-02 | 1.01 (1.00 – 1.03) | 4.6E-01 | 1.01 (0.98 – 1.03) | 1.02 (1.00 – 1.05) |
| 296.2 | Depression | AHI 3% | 1.00 (1.00 – 1.00) | 5.1E-02 | 1.00 (1.00 – 1.00) | 7.6E-01 | 1.00 (0.99 – 1.00) | 1.00 (1.00 – 1.00) |
| 261.2 | Vitamin B-complex deficiencies | Per88 | 0.99 (0.97 – 1.00) | 5.1E-02 | 0.99 (0.97 – 1.00) | 7.0E-01 | 0.99 (0.97 – 1.02) | 0.98 (0.96 – 1.00) |
| 345.1 | Epilepsy | AHI 3% | 0.99 (0.99 – 1.00) | 5.1E-02 | 0.99 (0.99 – 1.00) | 9.7E-01 | 0.99 (0.98 – 1.00) | 1.00 (0.99 – 1.00) |
| 401.3 | Other hypertensive complications | Per88 | 1.01 (1.00 – 1.03) | 5.2E-02 | 1.01 (1.00 – 1.03) | 6.7E-01 | 1.01 (1.00 – 1.03) | 1.01 (0.99 – 1.03) |
| 292 | Neurological disorders | AHI 3% | 1.00 (0.99 – 1.00) | 5.3E-02 | 1.00 (0.99 – 1.00) | 2.9E-03 | 1.00 (0.99 – 1.00) | 0.99 (0.99 – 1.00) |
| 367.4 | Presbyopia | AHI 3% | 1.00 (0.99 – 1.00) | 5.5E-02 | 1.00 (0.99 – 1.00) | 4.7E-02 | 1.00 (0.99 – 1.00) | 0.99 (0.98 – 1.00) |
| 496.3 | Bronchiectasis | Per88 | 1.02 (1.00 – 1.03) | 5.6E-02 | 1.01 (0.99 – 1.03) | 9.6E-01 | 1.01 (0.99 – 1.03) | 1.02 (1.00 – 1.05) |
| 306 | Other mental disorder | AHI 3% | 1.00 (1.00 – 1.01) | 5.7E-02 | 1.00 (1.00 – 1.01) | 1.9E-01 | 1.00 (1.00 – 1.01) | 1.00 (1.00 – 1.01) |
| 593.2 | Microscopic hematuria | AHI 3% | 1.00 (1.00 – 1.01) | 5.7E-02 | 1.00 (1.00 – 1.01) | 2.8E-01 | 1.00 (1.00 – 1.01) | 1.00 (1.00 – 1.01) |
| 575.7 | Other disorders of gallbladder | AHI 3% | 1.01 (1.00 – 1.01) | 5.8E-02 | 1.01 (1.00 – 1.01) | 7.3E-01 | 1.00 (1.00 – 1.01) | 1.01 (1.00 – 1.01) |
| 530.9 | Heartburn | AHI 3% | 1.00 (0.99 – 1.00) | 6.0E-02 | 1.00 (0.99 – 1.00) | 3.4E-01 | 1.00 (0.99 – 1.00) | 1.00 (0.99 – 1.00) |
| 427.5 | Arrhythmia (cardiac) NOS | Per88 | 1.01 (1.00 – 1.01) | 6.0E-02 | 1.00 (1.00 – 1.01) | 1.2E-01 | 1.00 (0.99 – 1.01) | 1.01 (1.00 – 1.02) |
| 798.1 | Chronic fatigue syndrome | Per88 | 0.99 (0.98 – 1.00) | 6.3E-02 | 0.99 (0.98 – 1.00) | 6.4E-01 | 0.99 (0.98 – 1.01) | 0.99 (0.98 – 1.00) |
| 374.3 | Ptosis of eyelid | Per88 | 0.98 (0.97 – 1.00) | 6.3E-02 | 0.98 (0.96 – 1.00) | 9.4E-01 | 0.97 (0.94 – 1.00) | 0.99 (0.97 – 1.01) |
| 526.41 | Temporomandibular joint disorder, unspecified | AHI 3% | 1.00 (0.99 – 1.00) | 6.4E-02 | 1.00 (0.99 – 1.00) | 7.3E-01 | 0.99 (0.98 – 1.00) | 0.99 (0.99 – 1.00) |
| 781 | Symptoms involving nervous and musculoskeletal systems | Per88 | 1.01 (1.00 – 1.02) | 6.5E-02 | 1.01 (1.00 – 1.01) | 4.4E-01 | 1.01 (0.99 – 1.02) | 1.01 (1.00 – 1.02) |

| PheCode | Disease | Sleep statistic | Risk ratio | P-value | Risk ratio (adj pulmonary disorders) | Sleep statistic x sex interaction p-value | Risk ratio (Men) | Risk ratio (Women) |
|---|---|---|---|---|---|---|---|---|
| 382 | Otalgia | Per88 | 0.99 (0.99 – 1.00) | 6.5E-02 | 0.99 (0.99 – 1.00) | 3.9E-01 | 0.99 (0.98 – 1.01) | 0.99 (0.98 – 1.00) |
| 291.4 | Specific nonpsychotic mental disorders due to brain damage | AHI 3% | 0.99 (0.99 – 1.00) | 6.5E-02 | 0.99 (0.99 – 1.00) | 1.9E-01 | 1.00 (0.99 – 1.00) | 0.99 (0.98 – 1.00) |
| 740.9 | Osteoarthrosis NOS | Per88 | 1.00 (1.00 – 1.01) | 6.5E-02 | 1.00 (1.00 – 1.01) | 7.1E-01 | 1.00 (0.99 – 1.01) | 1.01 (1.00 – 1.01) |
| 426.31 | Right bundle branch block | Per88 | 1.01 (1.00 – 1.03) | 6.6E-02 | 1.01 (1.00 – 1.03) | 8.7E-01 | 1.01 (1.00 – 1.03) | 1.02 (0.99 – 1.05) |
| 402 | Elevated blood pressure reading without diagnosis of hypertension | AHI 3% | 1.00 (1.00 – 1.00) | 6.6E-02 | 1.00 (1.00 – 1.00) | 4.6E-01 | 1.00 (1.00 – 1.00) | 1.00 (1.00 – 1.01) |
| 304 | Adjustment reaction | AHI 3% | 1.00 (0.99 – 1.00) | 6.6E-02 | 1.00 (0.99 – 1.00) | 3.5E-01 | 1.00 (0.99 – 1.00) | 1.00 (1.00 – 1.00) |
| 280.2 | Iron deficiency anemia secondary to blood loss (chronic) | Per88 | 1.01 (1.00 – 1.03) | 6.7E-02 | 1.01 (1.00 – 1.03) | 4.2E-01 | 1.03 (1.01 – 1.05) | 1.01 (1.00 – 1.03) |
| 512.9 | Other dyspnea | AHI 3% | 1.00 (1.00 – 1.00) | 6.7E-02 | 1.00 (1.00 – 1.00) | 8.6E-01 | 1.00 (1.00 – 1.00) | 1.00 (1.00 – 1.00) |
| 726 | Peripheral enthesopathies and allied syndromes | Per88 | 0.99 (0.99 – 1.00) | 6.8E-02 | 0.99 (0.99 – 1.00) | 2.4E-01 | 0.99 (0.98 – 1.00) | 1.00 (0.99 – 1.01) |
| 300.12 | Agorophobia, social phobia, and panic disorder | AHI 3% | 1.00 (0.99 – 1.00) | 6.8E-02 | 1.00 (0.99 – 1.00) | 3.5E-01 | 1.00 (0.99 – 1.00) | 1.00 (0.99 – 1.00) |
| 594.1 | Calculus of kidney | Per88 | 0.99 (0.98 – 1.00) | 6.9E-02 | 0.99 (0.98 – 1.00) | 8.0E-01 | 0.99 (0.98 – 1.00) | 0.99 (0.98 – 1.01) |
| 561.1 | Diarrhea | Per88 | 1.01 (1.00 – 1.01) | 7.0E-02 | 1.00 (1.00 – 1.01) | 6.8E-01 | 1.00 (0.99 – 1.01) | 1.01 (1.00 – 1.01) |
| 715 | Other inflammatory spondylopathies | AHI 3% | 0.99 (0.98 – 1.00) | 7.0E-02 | 0.99 (0.98 – 1.00) | 8.9E-01 | 0.99 (0.98 – 1.00) | 0.99 (0.98 – 1.01) |
| 578.9 | Hemorrhage of gastrointestinal tract | AHI 3% | 1.00 (1.00 – 1.01) | 7.1E-02 | 1.00 (1.00 – 1.01) | 5.8E-01 | 1.00 (1.00 – 1.01) | 1.01 (1.00 – 1.02) |
| 726.2 | Synoviopathy | Per88 | 0.99 (0.97 – 1.00) | 7.2E-02 | 0.99 (0.97 – 1.00) | 4.9E-01 | 0.99 (0.97 – 1.02) | 0.97 (0.95 – 1.00) |
| 345 | Epilepsy, recurrent seizures, convulsions | AHI 3% | 1.00 (0.99 – 1.00) | 7.3E-02 | 1.00 (0.99 – 1.00) | 1.1E-01 | 1.00 (0.99 – 1.00) | 0.99 (0.99 – 1.00) |
| 130.1 | Lyme disease | AHI 3% | 0.99 (0.99 – 1.00) | 7.5E-02 | 0.99 (0.99 – 1.00) | 7.5E-01 | 0.99 (0.99 – 1.00) | 1.00 (0.99 – 1.01) |
| 747.11 | Cardiac shunt/ heart septal defect | Per88 | 1.01 (1.00 – 1.03) | 7.5E-02 | 1.01 (1.00 – 1.03) | 2.8E-01 | 1.03 (1.01 – 1.05) | 1.00 (0.98 – 1.03) |
| 244.4 | Hypothyroidism NOS | Per88 | 1.01 (1.00 – 1.01) | 7.5E-02 | 1.00 (1.00 – 1.01) | 7.2E-01 | 1.00 (0.99 – 1.02) | 1.01 (1.00 – 1.01) |
| 586 | Other disorders of the kidney and ureters | Per88 | 1.01 (1.00 – 1.02) | 7.6E-02 | 1.01 (1.00 – 1.02) | 2.0E-01 | 1.01 (0.99 – 1.02) | 1.01 (0.99 – 1.03) |
| 689 | Disorder of skin and subcutaneous tissue NOS | Per88 | 1.00 (0.99 – 1.00) | 7.7E-02 | 1.00 (0.99 – 1.00) | 1.7E-02 | 0.99 (0.99 – 1.00) | 1.00 (0.99 – 1.01) |
| 720.1 | Spinal stenosis of lumbar region | Per88 | 1.01 (1.00 – 1.02) | 7.9E-02 | 1.01 (1.00 – 1.02) | 4.3E-01 | 1.01 (0.99 – 1.02) | 1.01 (1.00 – 1.03) |
| 454.1 | Varicose veins of lower extremity | AHI 3% | 1.00 (0.99 – 1.00) | 8.1E-02 | 1.00 (0.99 – 1.00) | 8.1E-01 | 0.99 (0.98 – 1.00) | 1.00 (0.99 – 1.00) |
| 564 | Functional digestive disorders | AHI 3% | 1.01 (1.00 – 1.01) | 8.4E-02 | 1.01 (1.00 – 1.01) | 9.3E-01 | 1.01 (1.00 – 1.02) | 1.00 (0.99 – 1.01) |
| 300.8 | Acute reaction to stress | Per88 | 0.99 (0.97 – 1.00) | 8.4E-02 | 0.99 (0.98 – 1.00) | 2.0E-01 | 0.99 (0.96 – 1.02) | 0.99 (0.97 – 1.00) |
| 819 | Skull and face fracture and other intercranial injury | AHI 3% | 0.99 (0.99 – 1.00) | 8.5E-02 | 0.99 (0.99 – 1.00) | 1.2E-01 | 1.00 (0.99 – 1.00) | 0.99 (0.98 – 1.00) |
| 428.3 | Heart failure with reduced EF [Systolic or combined heart failure] | AHI 3% | 1.01 (1.00 – 1.01) | 8.5E-02 | 1.01 (1.00 – 1.01) | 8.8E-01 | 1.00 (1.00 – 1.01) | 1.00 (0.99 – 1.02) |
| 456 | Chronic venous insufficiency [CVI] | AHI 3% | 1.00 (0.99 – 1.00) | 8.7E-02 | 1.00 (0.99 – 1.00) | 2.3E-01 | 0.99 (0.99 – 1.00) | 0.99 (0.99 – 1.00) |
| 427.9 | Palpitations | Per88 | 1.00 (0.99 – 1.00) | 8.8E-02 | 0.99 (0.99 – 1.00) | 3.5E-02 | 0.99 (0.98 – 1.00) | 0.99 (0.99 – 1.00) |
| 433.21 | Cerebral artery occlusion, with cerebral infarction | AHI 3% | 1.00 (1.00 – 1.01) | 8.8E-02 | 1.00 (1.00 – 1.01) | 6.1E-01 | 1.00 (1.00 – 1.01) | 1.00 (1.00 – 1.01) |
| 702.1 | Actinic keratosis | Per88 | 0.99 (0.98 – 1.00) | 8.9E-02 | 0.99 (0.98 – 1.00) | 6.3E-01 | 1.00 (0.98 – 1.01) | 0.99 (0.97 – 1.01) |

| PheCode | Disease | Sleep statistic | Risk ratio | P-value | Risk ratio (adj pulmonary disorders) | Sleep statistic x sex interaction p-value | Risk ratio (Men) | Risk ratio (Women) |
|---|---|---|---|---|---|---|---|---|
| 272.11 | Hypercholesterolemia | Per88 | 1.00 (0.99 – 1.00) | 9.0E-02 | 1.00 (0.99 – 1.00) | 9.0E-01 | 0.99 (0.99 – 1.00) | 1.00 (0.99 – 1.01) |
| 276.12 | Hyposmolality and/or hyponatremia | Per88 | 1.01 (1.00 – 1.02) | 9.0E-02 | 1.01 (1.00 – 1.02) | 3.5E-01 | 1.00 (0.98 – 1.02) | 1.02 (1.00 – 1.04) |
| 290.3 | Other persistent mental disorders due to conditions classified elsewhere | AHI 3% | 0.99 (0.99 – 1.00) | 9.2E-02 | 0.99 (0.99 – 1.00) | 9.4E-01 | 0.99 (0.98 – 1.00) | 1.00 (0.98 – 1.01) |
| 573.7 | Abnormal results of function study of liver | AHI 3% | 1.00 (1.00 – 1.01) | 9.2E-02 | 1.00 (1.00 – 1.01) | 3.3E-01 | 1.00 (1.00 – 1.01) | 1.00 (0.99 – 1.01) |
| 053 | Herpes zoster | AHI 3% | 1.00 (1.00 – 1.01) | 9.4E-02 | 1.00 (1.00 – 1.01) | 1.7E-01 | 1.00 (0.99 – 1.01) | 1.01 (1.00 – 1.01) |
| 773 | Pain in limb | AHI 3% | 1.00 (1.00 – 1.00) | 9.4E-02 | 1.00 (1.00 – 1.00) | 4.0E-01 | 1.00 (1.00 – 1.00) | 1.00 (1.00 – 1.00) |
| 355.1 | Chronic pain syndrome | Per88 | 1.01 (1.00 – 1.03) | 9.5E-02 | 1.01 (1.00 – 1.03) | 1.4E-01 | 1.01 (0.98 – 1.05) | 1.01 (0.99 – 1.03) |
| 280.1 | Iron deficiency anemias, unspecified or not due to blood loss | AHI 3% | 1.00 (0.99 – 1.00) | 9.5E-02 | 1.00 (0.99 – 1.00) | 2.9E-01 | 1.00 (0.99 – 1.00) | 1.00 (1.00 – 1.01) |
| 389.4 | Tinnitus | AHI 3% | 1.00 (0.99 – 1.00) | 9.5E-02 | 1.00 (0.99 – 1.00) | 5.1E-01 | 1.00 (1.00 – 1.00) | 1.00 (0.99 – 1.00) |
| 388 | Other disorders of ear | Per88 | 0.99 (0.98 – 1.00) | 9.6E-02 | 0.99 (0.98 – 1.00) | 1.6E-01 | 0.98 (0.96 – 1.00) | 1.00 (0.98 – 1.01) |
| 690.1 | Seborrheic dermatitis | Per88 | 0.99 (0.98 – 1.00) | 9.8E-02 | 0.99 (0.98 – 1.00) | 5.5E-01 | 1.00 (0.98 – 1.01) | 0.99 (0.97 – 1.01) |
| 459.9 | Circulatory disease NEC | Per88 | 1.01 (1.00 – 1.02) | 9.9E-02 | 1.01 (1.00 – 1.02) | 4.3E-01 | 1.01 (1.00 – 1.02) | 1.01 (1.00 – 1.03) |
| 733 | Other disorders of bone and cartilage | AHI 3% | 1.00 (1.00 – 1.00) | 1.0E-01 | 1.00 (1.00 – 1.00) | 1.0E+00 | 1.00 (1.00 – 1.01) | 1.00 (1.00 – 1.00) |
| 288.1 | Decreased white blood cell count | AHI 3% | 0.99 (0.99 – 1.00) | 1.0E-01 | 0.99 (0.99 – 1.00) | 5.9E-01 | 0.99 (0.98 – 1.00) | 0.99 (0.98 – 1.00) |
| 366 | Cataract | Per88 | 1.01 (1.00 – 1.02) | 1.0E-01 | 1.01 (1.00 – 1.02) | 3.7E-02 | 1.01 (1.00 – 1.02) | 1.00 (0.99 – 1.02) |
| 1013 | Asphyxia and hypoxemia | AHI 3% | 1.00 (1.00 – 1.01) | 1.1E-01 | 1.00 (1.00 – 1.01) | 1.9E-01 | 1.01 (1.00 – 1.02) | 1.00 (0.99 – 1.01) |
| 726.1 | Enthesopathy | Per88 | 0.99 (0.99 – 1.00) | 1.1E-01 | 0.99 (0.99 – 1.00) | 1.3E-01 | 1.00 (0.99 – 1.01) | 0.99 (0.98 – 1.00) |
| 772 | Symptoms of the muscles | Per88 | 0.99 (0.98 – 1.00) | 1.1E-01 | 0.99 (0.98 – 1.00) | 4.6E-01 | 1.00 (0.97 – 1.02) | 0.98 (0.97 – 1.00) |
| 585.4 | Chronic kidney disease, Stage I or II | AHI 3% | 1.01 (1.00 – 1.01) | 1.1E-01 | 1.01 (1.00 – 1.01) | 7.2E-02 | 1.00 (0.99 – 1.01) | 1.01 (1.00 – 1.03) |
| 735.3 | Hallux valgus (Bunion) | AHI 3% | 1.00 (0.99 – 1.00) | 1.1E-01 | 1.00 (0.99 – 1.00) | 3.6E-01 | 1.00 (0.99 – 1.01) | 0.99 (0.99 – 1.00) |
| 381.1 | Otitis media | AHI 3% | 1.00 (1.00 – 1.01) | 1.1E-01 | 1.00 (1.00 – 1.01) | 3.9E-01 | 1.00 (1.00 – 1.01) | 1.00 (0.99 – 1.01) |
| 368.2 | Diplopia and disorders of binocular vision | AHI 3% | 1.00 (0.99 – 1.00) | 1.1E-01 | 1.00 (0.99 – 1.00) | 6.0E-02 | 1.00 (0.99 – 1.01) | 0.99 (0.98 – 1.00) |
| 704.8 | Other specified diseases of hair and hair follicles | AHI 3% | 1.00 (0.99 – 1.00) | 1.1E-01 | 1.00 (0.99 – 1.00) | 2.6E-01 | 1.00 (0.99 – 1.00) | 0.99 (0.99 – 1.00) |
| 690.1 | Seborrheic dermatitis | AHI 3% | 1.00 (0.99 – 1.00) | 1.1E-01 | 1.00 (0.99 – 1.00) | 8.2E-01 | 1.00 (0.99 – 1.00) | 1.00 (0.99 – 1.00) |
| 704.1 | Alopecia | AHI 3% | 1.00 (1.00 – 1.00) | 1.1E-01 | 1.00 (1.00 – 1.00) | 2.1E-01 | 1.00 (1.00 – 1.01) | 1.00 (0.99 – 1.00) |
| 737.3 | Kyphoscoliosis and scoliosis | Per88 | 1.01 (1.00 – 1.02) | 1.1E-01 | 1.01 (1.00 – 1.02) | 1.5E-01 | 1.02 (0.99 – 1.04) | 1.00 (0.99 – 1.02) |
| 946 | Anaphylactic shock NOS | AHI 3% | 1.00 (0.99 – 1.00) | 1.2E-01 | 1.00 (0.99 – 1.00) | 5.2E-01 | 1.00 (0.99 – 1.01) | 0.99 (0.99 – 1.00) |
| 496.2 | Chronic bronchitis | AHI 3% | 0.99 (0.99 – 1.00) | 1.2E-01 | 0.99 (0.99 – 1.00) | 9.2E-01 | 1.00 (0.99 – 1.01) | 0.99 (0.98 – 1.00) |
| 350.1 | Abnormal involuntary movements | Per88 | 0.99 (0.98 – 1.00) | 1.2E-01 | 0.99 (0.98 – 1.00) | 6.1E-01 | 1.00 (0.98 – 1.01) | 1.00 (0.98 – 1.01) |
| 724.8 | Other symptoms referable to back | AHI 3% | 0.99 (0.99 – 1.00) | 1.2E-01 | 0.99 (0.99 – 1.00) | 8.0E-01 | 0.99 (0.98 – 1.00) | 1.00 (0.99 – 1.01) |
| 593 | Hematuria | AHI 3% | 1.00 (1.00 – 1.01) | 1.2E-01 | 1.00 (1.00 – 1.01) | 1.4E-01 | 1.00 (1.00 – 1.00) | 1.00 (1.00 – 1.01) |
| 327.71 | Restless legs syndrome | Per88 | 0.99 (0.98 – 1.00) | 1.2E-01 | 0.99 (0.98 – 1.00) | 7.6E-01 | 1.00 (0.98 – 1.01) | 0.99 (0.98 – 1.00) |
| 594.8 | Renal colic | Per88 | 0.98 (0.96 – 1.00) | 1.2E-01 | 0.98 (0.96 – 1.00) | 5.6E-01 | 0.98 (0.95 – 1.01) | 0.99 (0.96 – 1.02) |
| 740.9 | Osteoarthrosis NOS | AHI 3% | 1.00 (1.00 – 1.00) | 1.2E-01 | 1.00 (1.00 – 1.00) | 5.5E-01 | 1.00 (0.99 – 1.00) | 1.00 (1.00 – 1.00) |

| PheCode | Disease | Sleep statistic | Risk ratio | P-value | Risk ratio (adj pulmonary disorders) | Sleep statistic x sex interaction p-value | Risk ratio (Men) | Risk ratio (Women) |
|---|---|---|---|---|---|---|---|---|
| 136 | Other infectious and parasitic diseases | AHI 3% | 1.00 (1.00 – 1.01) | 1.2E-01 | 1.00 (1.00 – 1.01) | 4.4E-01 | 1.00 (1.00 – 1.01) | 1.00 (1.00 – 1.01) |
| 840.3 | Joint/ligament sprain | Per88 | 0.99 (0.98 – 1.00) | 1.2E-01 | 0.99 (0.98 – 1.00) | 5.9E-01 | 0.99 (0.98 – 1.01) | 0.99 (0.98 – 1.01) |
| 429.3 | Symptoms involving cardiovascular system | Per88 | 1.01 (1.00 – 1.01) | 1.3E-01 | 1.00 (1.00 – 1.01) | 1.4E-01 | 1.00 (0.99 – 1.01) | 1.01 (1.00 – 1.03) |
| 772 | Symptoms of the muscles | AHI 3% | 1.00 (0.99 – 1.00) | 1.3E-01 | 1.00 (0.99 – 1.00) | 2.3E-01 | 0.99 (0.98 – 1.00) | 1.00 (0.99 – 1.00) |
| 941 | Adverse reaction to serum or vaccine | Per88 | 0.99 (0.98 – 1.00) | 1.3E-01 | 0.99 (0.98 – 1.00) | 4.2E-01 | 0.99 (0.97 – 1.01) | 0.99 (0.97 – 1.00) |
| 252.1 | Hyperparathyroidism | AHI 3% | 1.00 (1.00 – 1.01) | 1.3E-01 | 1.00 (1.00 – 1.01) | 3.6E-02 | 1.00 (0.98 – 1.01) | 1.01 (1.00 – 1.02) |
| 564.1 | Irritable Bowel Syndrome | Per88 | 0.99 (0.99 – 1.00) | 1.3E-01 | 0.99 (0.99 – 1.00) | 9.1E-01 | 0.99 (0.98 – 1.01) | 0.99 (0.98 – 1.00) |
| 279 | Disorders involving the immune mechanism | AHI 3% | 1.00 (1.00 – 1.01) | 1.3E-01 | 1.00 (1.00 – 1.01) | 3.8E-02 | 1.01 (1.00 – 1.02) | 1.00 (0.99 – 1.01) |
| 458.1 | Orthostatic hypotension | AHI 3% | 1.00 (0.99 – 1.00) | 1.3E-01 | 0.99 (0.99 – 1.00) | 3.6E-01 | 0.99 (0.98 – 1.00) | 1.00 (0.99 – 1.01) |
| 338.2 | Chronic pain | Per88 | 1.00 (1.00 – 1.01) | 1.3E-01 | 1.00 (1.00 – 1.01) | 2.6E-02 | 1.00 (0.99 – 1.00) | 1.01 (1.00 – 1.01) |
| 173 | Neoplasm of uncertain behavior of skin | Per88 | 0.99 (0.99 – 1.00) | 1.3E-01 | 0.99 (0.99 – 1.00) | 6.5E-01 | 0.99 (0.98 – 1.00) | 1.00 (0.99 – 1.01) |
| 280.1 | Iron deficiency anemias, unspecified or not due to blood loss | Per88 | 1.01 (1.00 – 1.01) | 1.3E-01 | 1.00 (1.00 – 1.01) | 6.4E-01 | 1.01 (0.99 – 1.02) | 1.01 (1.00 – 1.02) |
| 241.2 | Nontoxic multinodular goiter | Per88 | 0.99 (0.98 – 1.00) | 1.3E-01 | 0.99 (0.98 – 1.00) | 7.5E-01 | 0.98 (0.95 – 1.01) | 0.99 (0.97 – 1.00) |
| 447 | Other disorders of arteries and arterioles | AHI 3% | 1.01 (1.00 – 1.01) | 1.3E-01 | 1.01 (1.00 – 1.01) | 7.7E-01 | 1.00 (0.99 – 1.01) | 1.01 (1.00 – 1.02) |
| 217.1 | Nevus, non-neoplastic | Per88 | 0.99 (0.98 – 1.00) | 1.3E-01 | 0.99 (0.98 – 1.00) | 8.9E-02 | 0.98 (0.97 – 1.00) | 1.00 (0.99 – 1.01) |
| 696.41 | Psoriasis vulgaris | AHI 3% | 1.00 (1.00 – 1.01) | 1.3E-01 | 1.00 (1.00 – 1.01) | 2.5E-01 | 1.00 (0.99 – 1.01) | 1.01 (1.00 – 1.02) |
| 733 | Other disorders of bone and cartilage | Per88 | 1.00 (1.00 – 1.01) | 1.4E-01 | 1.00 (1.00 – 1.01) | 5.4E-01 | 1.00 (0.99 – 1.01) | 1.00 (0.99 – 1.01) |
| 302.1 | Decreased libido | AHI 3% | 1.00 (0.99 – 1.00) | 1.4E-01 | 1.00 (0.99 – 1.00) | 5.8E-01 | 0.99 (0.99 – 1.00) | 1.00 (0.99 – 1.01) |
| 426.32 | Left bundle branch block | AHI 3% | 1.01 (1.00 – 1.01) | 1.4E-01 | 1.01 (1.00 – 1.01) | 3.6E-01 | 1.00 (1.00 – 1.01) | 1.01 (1.00 – 1.02) |
| 277 | Other disorders of metabolism | AHI 3% | 1.00 (0.99 – 1.00) | 1.4E-01 | 1.00 (0.99 – 1.00) | 2.8E-02 | 1.00 (0.99 – 1.01) | 1.00 (0.99 – 1.00) |
| 686.1 | Carbuncle and furuncle | Per88 | 1.01 (1.00 – 1.03) | 1.4E-01 | 1.01 (0.99 – 1.03) | 1.1E-01 | 1.00 (0.97 – 1.03) | 1.02 (1.00 – 1.04) |
| 278.11 | Morbid obesity | Per88 | 1.00 (1.00 – 1.01) | 1.5E-01 | 1.00 (1.00 – 1.01) | 2.2E-01 | 1.01 (1.00 – 1.02) | 1.00 (0.99 – 1.01) |
| 427.11 | Paroxysmal supraventricular tachycardia | AHI 3% | 1.00 (1.00 – 1.01) | 1.5E-01 | 1.00 (1.00 – 1.01) | 1.2E-01 | 1.00 (0.99 – 1.01) | 1.00 (1.00 – 1.01) |
| 442.1 | Aortic aneurysm | Per88 | 1.02 (0.99 – 1.04) | 1.5E-01 | 1.01 (0.99 – 1.04) | 8.5E-02 | 0.99 (0.97 – 1.02) | 1.06 (1.03 – 1.08) |
| 196 | Radiotherapy | Per88 | 1.01 (1.00 – 1.02) | 1.5E-01 | 1.01 (1.00 – 1.02) | 8.0E-01 | 1.01 (0.99 – 1.02) | 1.01 (0.99 – 1.03) |
| 701 | Other hypertrophic and atrophic conditions of skin | Per88 | 1.00 (0.99 – 1.00) | 1.5E-01 | 1.00 (0.99 – 1.00) | 3.2E-01 | 0.99 (0.98 – 1.00) | 0.99 (0.98 – 1.00) |
| 426.32 | Left bundle branch block | Per88 | 1.01 (0.99 – 1.03) | 1.5E-01 | 1.01 (0.99 – 1.03) | 3.9E-01 | 1.00 (0.98 – 1.03) | 1.03 (1.00 – 1.06) |
| 537 | Other disorders of stomach and duodenum | Per88 | 0.99 (0.99 – 1.00) | 1.6E-01 | 0.99 (0.99 – 1.00) | 9.2E-01 | 1.00 (0.98 – 1.01) | 1.00 (0.98 – 1.01) |
| 586.4 | Stricture/obstruction of ureter | Per88 | 0.99 (0.97 – 1.00) | 1.6E-01 | 0.99 (0.97 – 1.01) | NA | 1.00 (0.98 – 1.01) | NA |
| 721.1 | Spondylosis without myelopathy | AHI 3% | 1.00 (1.00 – 1.00) | 1.6E-01 | 1.00 (1.00 – 1.00) | 5.3E-01 | 1.00 (0.99 – 1.00) | 1.00 (0.99 – 1.00) |
| 531.2 | Gastric ulcer | Per88 | 1.01 (1.00 – 1.03) | 1.6E-01 | 1.01 (0.99 – 1.03) | 9.3E-01 | 1.00 (0.98 – 1.03) | 1.01 (0.99 – 1.03) |
| 292.2 | Mild cognitive impairment | AHI 3% | 1.00 (0.99 – 1.00) | 1.6E-01 | 1.00 (0.99 – 1.00) | 3.4E-02 | 1.00 (0.99 – 1.00) | 1.00 (0.99 – 1.01) |
| 300.1 | Anxiety disorder | AHI 3% | 1.00 (1.00 – 1.00) | 1.6E-01 | 1.00 (1.00 – 1.00) | 2.2E-03 | 1.00 (1.00 – 1.00) | 1.00 (1.00 – 1.00) |
| 965 | Poisoning by analgesics, antipyretics, and | Per88 | 1.01 (1.00 – 1.02) | 1.6E-01 | 1.00 (1.00 – 1.01) | 5.1E-01 | 1.00 (0.98 – 1.03) | 1.01 (0.99 – 1.02) |

| PheCode | Disease | Sleep statistic | Risk ratio | P-value | Risk ratio (adj pulmonary disorders) | Sleep statistic x sex interaction p-value | Risk ratio (Men) | Risk ratio (Women) |
|---|---|---|---|---|---|---|---|---|
| | antirheumatics | | | | | | | |
| 530.13 | Barrett's esophagus | Per88 | 1.01 (1.00 – 1.03) | 1.6E-01 | 1.01 (1.00 – 1.03) | 1.2E-01 | 1.00 (0.98 – 1.02) | 1.02 (1.00 – 1.05) |
| 251.1 | Hypoglycemia | Per88 | 1.01 (1.00 – 1.03) | 1.6E-01 | 1.01 (0.99 – 1.03) | 1.0E-01 | 1.02 (0.99 – 1.05) | 1.01 (0.99 – 1.03) |
| 599.4 | Urinary incontinence | AHI 3% | 1.00 (0.99 – 1.00) | 1.6E-01 | 1.00 (0.99 – 1.00) | 1.4E-01 | 1.00 (0.99 – 1.00) | 1.00 (0.99 – 1.00) |
| 763 | Thoracic or lumbosacral neuritis or radiculitis, unspecified | AHI 3% | 1.00 (1.00 – 1.00) | 1.7E-01 | 1.00 (1.00 – 1.00) | 9.2E-01 | 1.00 (0.99 – 1.00) | 1.00 (0.99 – 1.00) |
| 300.4 | Dysthymic disorder | Per88 | 0.99 (0.99 – 1.00) | 1.7E-01 | 0.99 (0.99 – 1.00) | 1.7E-01 | 0.98 (0.96 – 1.00) | 1.00 (0.99 – 1.01) |
| 427.12 | Paroxysmal ventricular tachycardia | Per88 | 1.01 (0.99 – 1.03) | 1.7E-01 | 1.01 (0.99 – 1.03) | 2.2E-01 | 1.01 (0.99 – 1.03) | 1.00 (0.97 – 1.03) |
| 727.6 | Rupture of tendon, nontraumatic | Per88 | 0.99 (0.97 – 1.01) | 1.7E-01 | 0.99 (0.97 – 1.01) | 8.1E-01 | 0.98 (0.96 – 1.01) | 0.99 (0.97 – 1.02) |
| 785 | Abdominal pain | Per88 | 1.00 (0.99 – 1.00) | 1.7E-01 | 1.00 (0.99 – 1.00) | 3.4E-01 | 0.99 (0.99 – 1.00) | 1.00 (1.00 – 1.00) |
| 287.3 | Thrombocytopenia | AHI 3% | 1.00 (1.00 – 1.01) | 1.7E-01 | 1.00 (1.00 – 1.01) | 8.0E-01 | 1.00 (1.00 – 1.01) | 1.00 (0.99 – 1.01) |
| 228.1 | Hemangioma of skin and subcutaneous tissue | AHI 3% | 1.00 (0.99 – 1.00) | 1.7E-01 | 1.00 (0.99 – 1.00) | 9.6E-01 | 1.00 (1.00 – 1.00) | 1.00 (0.99 – 1.00) |
| 386.9 | Dizziness and giddiness (Light-headedness and vertigo) | AHI 3% | 1.00 (1.00 – 1.00) | 1.7E-01 | 1.00 (1.00 – 1.00) | 3.0E-02 | 1.00 (1.00 – 1.00) | 1.00 (0.99 – 1.00) |
| 420.2 | Pericarditis | Per88 | 1.01 (0.99 – 1.03) | 1.7E-01 | 1.01 (0.99 – 1.03) | 6.8E-01 | 1.01 (0.98 – 1.03) | 1.02 (0.99 – 1.04) |
| 172.2 | Other non-epithelial cancer of skin | Per88 | 0.99 (0.98 – 1.00) | 1.7E-01 | 0.99 (0.98 – 1.00) | 1.2E-01 | 1.00 (0.98 – 1.01) | 0.99 (0.97 – 1.01) |
| 788 | Syncope and collapse | Per88 | 1.00 (1.00 – 1.01) | 1.7E-01 | 1.00 (1.00 – 1.01) | 9.9E-01 | 1.01 (1.00 – 1.01) | 1.01 (1.00 – 1.02) |
| 172.21 | Basal cell carcinoma | AHI 3% | 1.00 (0.99 – 1.00) | 1.7E-01 | 1.00 (0.99 – 1.00) | 9.7E-01 | 1.00 (0.99 – 1.00) | 1.00 (0.99 – 1.01) |
| 454.11 | Varicose veins of lower extremity, symptomtic | Per88 | 1.01 (1.00 – 1.02) | 1.7E-01 | 1.01 (1.00 – 1.02) | 3.7E-01 | 1.01 (0.99 – 1.04) | 1.01 (0.99 – 1.02) |
| 535.2 | Atrophic gastritis | AHI 3% | 1.00 (0.99 – 1.00) | 1.8E-01 | 1.00 (0.99 – 1.00) | 8.0E-02 | 1.00 (0.99 – 1.00) | 1.00 (0.99 – 1.01) |
| 380.1 | Otitis externa | AHI 3% | 1.00 (0.99 – 1.00) | 1.8E-01 | 1.00 (0.99 – 1.00) | 9.1E-01 | 1.00 (0.99 – 1.00) | 1.00 (0.99 – 1.01) |
| 242 | Thyrotoxicosis with or without goiter | AHI 3% | 1.00 (1.00 – 1.01) | 1.8E-01 | 1.00 (1.00 – 1.01) | 3.0E-01 | 1.01 (1.00 – 1.02) | 1.00 (1.00 – 1.01) |
| 703.1 | Ingrowing nail | Per88 | 1.01 (0.99 – 1.03) | 1.8E-01 | 1.01 (0.99 – 1.03) | 6.2E-01 | 1.00 (0.98 – 1.03) | 1.01 (0.99 – 1.04) |
| 274.11 | Gouty arthropathy | Per88 | 1.01 (0.99 – 1.03) | 1.8E-01 | 1.01 (0.99 – 1.03) | 3.5E-01 | 1.00 (0.98 – 1.02) | 1.03 (0.99 – 1.06) |
| 008.5 | Bacterial enteritis | Per88 | 0.99 (0.98 – 1.00) | 1.9E-01 | 0.99 (0.98 – 1.00) | 8.6E-01 | 1.00 (0.98 – 1.01) | 0.99 (0.98 – 1.00) |
| 304 | Adjustment reaction | Per88 | 1.00 (0.99 – 1.00) | 1.9E-01 | 1.00 (0.99 – 1.00) | 7.4E-01 | 1.00 (0.99 – 1.01) | 0.99 (0.99 – 1.00) |
| 938.2 | Chronic dermatitis due to solar radiation | AHI 3% | 1.00 (0.99 – 1.00) | 1.9E-01 | 1.00 (0.99 – 1.00) | 4.4E-01 | 1.00 (0.99 – 1.00) | 1.00 (0.99 – 1.00) |
| 313.1 | Attention deficit hyperactivity disorder | AHI 3% | 1.00 (1.00 – 1.00) | 1.9E-01 | 1.00 (1.00 – 1.00) | 2.8E-01 | 1.00 (0.99 – 1.00) | 1.00 (0.99 – 1.00) |
| 560.4 | Other intestinal obstruction | Per88 | 1.01 (1.00 – 1.02) | 1.9E-01 | 1.01 (0.99 – 1.02) | 5.8E-03 | 1.03 (1.01 – 1.05) | 0.99 (0.96 – 1.01) |
| 716.9 | Arthropathy NOS | AHI 3% | 1.00 (0.99 – 1.00) | 1.9E-01 | 1.00 (0.99 – 1.00) | 2.2E-02 | 0.99 (0.98 – 1.00) | 1.00 (1.00 – 1.01) |
| 395.2 | Nonrheumatic aortic valve disorders | AHI 3% | 1.00 (1.00 – 1.01) | 1.9E-01 | 1.00 (1.00 – 1.01) | 1.3E-02 | 1.00 (0.99 – 1.01) | 1.01 (1.00 – 1.02) |
| 723 | Other disorders of cervical region | Per88 | 0.99 (0.98 – 1.00) | 1.9E-01 | 0.99 (0.98 – 1.01) | 7.2E-01 | 0.99 (0.96 – 1.02) | 0.99 (0.97 – 1.01) |
| 289.9 | Abnormality of red blood cells | Per88 | 1.01 (1.00 – 1.02) | 1.9E-01 | 1.01 (1.00 – 1.02) | 1.9E-01 | 1.01 (0.99 – 1.03) | 1.01 (0.99 – 1.03) |
| 536.8 | Dyspepsia and other specified disorders of function of stomach | Per88 | 0.99 (0.99 – 1.00) | 1.9E-01 | 0.99 (0.99 – 1.00) | 9.5E-01 | 1.00 (0.99 – 1.02) | 0.99 (0.98 – 1.00) |
| 371.3 | Inflammation of eyelids | Per88 | 0.99 (0.98 – 1.00) | 2.0E-01 | 0.99 (0.98 – 1.01) | 2.7E-01 | 0.98 (0.96 – 0.99) | 1.00 (0.98 – 1.01) |

| PheCode | Disease | Sleep statistic | Risk ratio | P-value | Risk ratio (adj pulmonary disorders) | Sleep statistic x sex interaction p-value | Risk ratio (Men) | Risk ratio (Women) |
|---|---|---|---|---|---|---|---|---|
| 272.11 | Hypercholesterolemia | AHI 3% | 1.00 (1.00 – 1.00) | 2.0E-01 | 1.00 (1.00 – 1.00) | 9.9E-04 | 1.00 (1.00 – 1.00) | 1.00 (1.00 – 1.01) |
| 496.1 | Emphysema | AHI 3% | 1.00 (0.99 – 1.00) | 2.0E-01 | 1.00 (0.99 – 1.00) | 4.6E-01 | 1.00 (0.99 – 1.01) | 0.99 (0.98 – 1.00) |
| 278.1 | Obesity | Per88 | 1.00 (0.99 – 1.00) | 2.0E-01 | 1.00 (0.99 – 1.00) | 3.3E-02 | 1.00 (0.99 – 1.00) | 1.00 (0.99 – 1.00) |
| 300.9 | Posttraumatic stress disorder | Per88 | 1.01 (1.00 – 1.01) | 2.0E-01 | 1.00 (1.00 – 1.01) | 5.0E-01 | 1.01 (0.99 – 1.02) | 1.01 (1.00 – 1.02) |
| 350.2 | Abnormality of gait | Per88 | 0.99 (0.98 – 1.00) | 2.0E-01 | 0.99 (0.98 – 1.00) | 9.6E-01 | 1.00 (0.98 – 1.01) | 0.99 (0.97 – 1.01) |
| 300.12 | Agorophobia, social phobia, and panic disorder | Per88 | 1.01 (1.00 – 1.01) | 2.0E-01 | 1.01 (1.00 – 1.01) | 7.4E-01 | 1.01 (0.99 – 1.02) | 1.01 (1.00 – 1.02) |
| 290.3 | Other persistent mental disorders due to conditions classified elsewhere | Per88 | 0.99 (0.97 – 1.01) | 2.0E-01 | 0.99 (0.97 – 1.01) | 7.5E-01 | 1.00 (0.97 – 1.02) | 0.98 (0.95 – 1.01) |
| 379.9 | Pain, swelling or discharge of eye | AHI 3% | 1.00 (0.99 – 1.00) | 2.0E-01 | 1.00 (0.99 – 1.00) | 5.7E-01 | 1.00 (0.99 – 1.00) | 1.00 (0.99 – 1.00) |
| 078 | Viral warts & HPV | Per88 | 0.99 (0.99 – 1.00) | 2.0E-01 | 1.00 (0.99 – 1.00) | 2.9E-01 | 0.99 (0.98 – 1.01) | 1.00 (0.98 – 1.01) |
| 840 | Sprains and strains | Per88 | 1.00 (0.99 – 1.00) | 2.1E-01 | 0.99 (0.99 – 1.00) | 4.0E-01 | 1.00 (0.99 – 1.01) | 0.99 (0.98 – 1.00) |
| 530.14 | Reflux esophagitis | Per88 | 0.99 (0.98 – 1.00) | 2.1E-01 | 0.99 (0.98 – 1.00) | 3.1E-01 | 0.98 (0.96 – 1.00) | 1.00 (0.99 – 1.02) |
| 327.71 | Restless legs syndrome | AHI 3% | 1.00 (0.99 – 1.00) | 2.1E-01 | 1.00 (0.99 – 1.00) | 2.2E-01 | 1.00 (0.99 – 1.00) | 1.00 (0.99 – 1.00) |
| 316 | Substance addiction and disorders | AHI 3% | 1.00 (1.00 – 1.01) | 2.1E-01 | 1.00 (1.00 – 1.01) | 2.0E-01 | 1.00 (1.00 – 1.01) | 1.00 (1.00 – 1.01) |
| 112 | Candidiasis | AHI 3% | 1.00 (0.99 – 1.00) | 2.1E-01 | 1.00 (0.99 – 1.00) | 1.6E-01 | 1.00 (1.00 – 1.01) | 1.00 (0.99 – 1.00) |
| 110.11 | Dermatophytosis of nail | AHI 3% | 1.00 (0.99 – 1.00) | 2.2E-01 | 1.00 (0.99 – 1.00) | 2.8E-01 | 1.00 (0.99 – 1.00) | 1.00 (0.99 – 1.01) |
| 261.4 | Vitamin D deficiency | Per88 | 1.00 (0.99 – 1.00) | 2.2E-01 | 1.00 (0.99 – 1.00) | 3.8E-01 | 1.00 (0.99 – 1.01) | 1.00 (0.99 – 1.00) |
| 528 | Diseases of the oral soft tissues, excluding lesions specific for gingiva and tongue | Per88 | 1.01 (1.00 – 1.02) | 2.2E-01 | 1.01 (1.00 – 1.02) | 7.3E-01 | 1.00 (0.98 – 1.03) | 1.01 (0.99 – 1.02) |
| 526 | Diseases of the jaws | AHI 3% | 1.00 (0.99 – 1.00) | 2.2E-01 | 1.00 (0.99 – 1.00) | 6.7E-01 | 1.00 (0.99 – 1.01) | 1.00 (0.99 – 1.00) |
| 428.4 | Heart failure with preserved EF [Diastolic heart failure] | AHI 3% | 1.00 (1.00 – 1.01) | 2.2E-01 | 1.00 (1.00 – 1.01) | 5.1E-01 | 1.00 (0.99 – 1.01) | 1.01 (1.00 – 1.02) |
| 300.13 | Phobia | AHI 3% | 1.00 (0.99 – 1.00) | 2.2E-01 | 1.00 (0.99 – 1.00) | 8.2E-01 | 0.99 (0.98 – 1.00) | 1.00 (0.99 – 1.01) |
| 327 | Sleep disorders | Per88 | 1.00 (0.99 – 1.00) | 2.2E-01 | 1.00 (0.99 – 1.00) | 4.4E-01 | 1.00 (0.99 – 1.01) | 0.99 (0.98 – 1.00) |
| 594.3 | Calculus of ureter | Per88 | 0.99 (0.96 – 1.01) | 2.2E-01 | 0.99 (0.96 – 1.01) | 4.2E-01 | 0.99 (0.97 – 1.02) | 0.98 (0.95 – 1.02) |
| 365 | Glaucoma | Per88 | 1.01 (1.00 – 1.01) | 2.2E-01 | 1.01 (1.00 – 1.01) | 5.1E-01 | 1.01 (1.00 – 1.02) | 1.00 (0.99 – 1.02) |
| 415.2 | Chronic pulmonary heart disease | AHI 3% | 1.00 (1.00 – 1.01) | 2.2E-01 | 1.00 (1.00 – 1.01) | 6.6E-01 | 1.01 (1.00 – 1.02) | 1.00 (0.99 – 1.01) |
| 382 | Otalgia | AHI 3% | 1.00 (1.00 – 1.00) | 2.3E-01 | 1.00 (1.00 – 1.00) | 5.9E-01 | 1.00 (0.99 – 1.00) | 1.00 (1.00 – 1.00) |
| 278 | Overweight, obesity and other hyperalimentation | AHI 3% | 1.00 (1.00 – 1.00) | 2.3E-01 | 1.00 (1.00 – 1.00) | 3.6E-01 | 1.00 (1.00 – 1.01) | 1.00 (1.00 – 1.00) |
| 357 | Inflammatory and toxic neuropathy | AHI 3% | 1.00 (0.99 – 1.00) | 2.3E-01 | 1.00 (0.99 – 1.00) | 4.5E-01 | 1.00 (0.99 – 1.00) | 1.00 (0.99 – 1.00) |
| 366.2 | Senile cataract | AHI 3% | 1.00 (0.99 – 1.00) | 2.4E-01 | 1.00 (0.99 – 1.00) | 7.2E-01 | 1.00 (1.00 – 1.00) | 1.00 (0.99 – 1.00) |
| 565 | Anal and rectal conditions | Per88 | 0.99 (0.98 – 1.00) | 2.4E-01 | 0.99 (0.98 – 1.00) | 6.3E-01 | 0.99 (0.98 – 1.01) | 1.00 (0.98 – 1.01) |
| 447.7 | Aortic ectasia | AHI 3% | 1.00 (0.99 – 1.00) | 2.4E-01 | 1.00 (0.99 – 1.00) | 6.2E-02 | 0.99 (0.99 – 1.00) | 1.01 (0.99 – 1.02) |
| 367.2 | Astigmatism | AHI 3% | 1.00 (0.99 – 1.00) | 2.4E-01 | 1.00 (0.99 – 1.00) | 3.1E-01 | 1.00 (0.99 – 1.01) | 0.99 (0.99 – 1.00) |
| 263 | Other nutritional deficiency | AHI 3% | 1.00 (0.99 – 1.00) | 2.4E-01 | 1.00 (0.99 – 1.00) | 5.5E-03 | 1.01 (0.99 – 1.02) | 0.99 (0.98 – 1.00) |
| 761 | Cervicalgia | Per88 | 1.00 (0.99 – 1.00) | 2.4E-01 | 1.00 (0.99 – 1.00) | 6.1E-01 | 0.99 (0.99 – 1.00) | 1.00 (0.99 – 1.00) |

| PheCode | Disease | Sleep statistic | Risk ratio | P-value | Risk ratio (adj pulmonary disorders) | Sleep statistic x sex interaction p-value | Risk ratio (Men) | Risk ratio (Women) |
|---|---|---|---|---|---|---|---|---|
| 379.2 | Disorders of vitreous body | Per88 | 0.99 (0.98 – 1.00) | 2.4E-01 | 0.99 (0.98 – 1.01) | 6.6E-01 | 1.00 (0.99 – 1.02) | 0.98 (0.96 – 1.00) |
| 695.8 | Other specified erythematous conditions | AHI 3% | 1.00 (0.99 – 1.00) | 2.4E-01 | 1.00 (0.99 – 1.00) | 3.9E-01 | 0.99 (0.98 – 1.00) | 1.00 (0.99 – 1.01) |
| 110.13 | Dermatophytosis of the body | Per88 | 0.99 (0.97 – 1.01) | 2.4E-01 | 0.99 (0.97 – 1.01) | 1.5E-01 | 0.99 (0.97 – 1.01) | 0.98 (0.95 – 1.01) |
| 334 | Degenerative disease of the spinal cord | AHI 3% | 1.00 (1.00 – 1.01) | 2.4E-01 | 1.00 (1.00 – 1.01) | 9.6E-01 | 1.00 (0.99 – 1.01) | 1.00 (0.99 – 1.02) |
| 214 | Lipoma | AHI 3% | 1.00 (0.99 – 1.00) | 2.4E-01 | 1.00 (0.99 – 1.00) | 9.3E-01 | 1.00 (0.99 – 1.00) | 1.00 (0.99 – 1.00) |
| 402 | Elevated blood pressure reading without diagnosis of hypertension | Per88 | 1.00 (0.99 – 1.00) | 2.4E-01 | 1.00 (0.99 – 1.00) | 2.8E-01 | 1.00 (0.99 – 1.01) | 0.99 (0.98 – 1.00) |
| 351 | Other peripheral nerve disorders | AHI 3% | 1.00 (1.00 – 1.00) | 2.4E-01 | 1.00 (1.00 – 1.00) | 8.6E-01 | 1.00 (0.99 – 1.00) | 1.00 (1.00 – 1.00) |
| 165.1 | Cancer of bronchus; lung | AHI 3% | 1.00 (1.00 – 1.01) | 2.4E-01 | 1.00 (1.00 – 1.01) | 4.5E-01 | 1.00 (0.99 – 1.01) | 1.01 (1.00 – 1.02) |
| 726.2 | Synoviopathy | AHI 3% | 1.00 (0.99 – 1.00) | 2.5E-01 | 1.00 (0.99 – 1.00) | 3.6E-01 | 0.99 (0.99 – 1.00) | 1.00 (0.99 – 1.01) |
| 569 | Other disorders of intestine | AHI 3% | 1.00 (1.00 – 1.00) | 2.5E-01 | 1.00 (1.00 – 1.00) | 8.0E-01 | 1.00 (1.00 – 1.01) | 1.00 (1.00 – 1.00) |
| 947 | Urticaria | AHI 3% | 1.00 (0.99 – 1.00) | 2.5E-01 | 1.00 (0.99 – 1.00) | 1.4E-01 | 1.00 (1.00 – 1.01) | 1.00 (0.99 – 1.00) |
| 727.6 | Rupture of tendon, nontraumatic | AHI 3% | 1.00 (0.99 – 1.00) | 2.5E-01 | 1.00 (0.99 – 1.00) | 4.1E-01 | 0.99 (0.99 – 1.00) | 1.00 (0.99 – 1.01) |
| 411.2 | Myocardial infarction | AHI 3% | 1.00 (1.00 – 1.01) | 2.5E-01 | 1.00 (1.00 – 1.01) | 3.4E-02 | 1.00 (1.00 – 1.00) | 1.01 (1.00 – 1.02) |
| 242 | Thyrotoxicosis with or without goiter | Per88 | 0.99 (0.98 – 1.01) | 2.5E-01 | 0.99 (0.98 – 1.01) | 4.7E-01 | 1.01 (0.99 – 1.03) | 0.99 (0.97 – 1.00) |
| 704 | Diseases of hair and hair follicles | Per88 | 0.99 (0.98 – 1.00) | 2.5E-01 | 1.00 (0.98 – 1.01) | 7.5E-02 | 1.00 (0.98 – 1.01) | 0.99 (0.97 – 1.00) |
| 241.1 | Nontoxic uninodular goiter | Per88 | 0.99 (0.99 – 1.00) | 2.5E-01 | 1.00 (0.99 – 1.00) | 9.2E-01 | 0.99 (0.97 – 1.01) | 0.99 (0.98 – 1.00) |
| 252.1 | Hyperparathyroidism | Per88 | 1.01 (0.99 – 1.03) | 2.6E-01 | 1.01 (0.99 – 1.03) | 7.9E-01 | 1.01 (0.98 – 1.03) | 1.01 (0.99 – 1.03) |
| 701.2 | Scar conditions and fibrosis of skin | AHI 3% | 1.00 (1.00 – 1.00) | 2.6E-01 | 1.00 (1.00 – 1.00) | 7.0E-01 | 1.00 (0.99 – 1.00) | 1.00 (1.00 – 1.01) |
| 246 | Other disorders of thyroid | AHI 3% | 1.00 (1.00 – 1.01) | 2.6E-01 | 1.00 (1.00 – 1.01) | 1.5E-01 | 1.00 (0.99 – 1.01) | 1.00 (1.00 – 1.01) |
| 348.7 | Coma | Per88 | 1.01 (0.99 – 1.02) | 2.6E-01 | 1.01 (0.99 – 1.02) | 8.7E-01 | 1.00 (0.97 – 1.02) | 1.02 (1.00 – 1.04) |
| 841 | Sprains and strains of back and neck | Per88 | 0.99 (0.98 – 1.01) | 2.6E-01 | 0.99 (0.98 – 1.01) | 3.2E-01 | 0.98 (0.96 – 1.01) | 0.99 (0.98 – 1.01) |
| 593.1 | Gross hematuria | AHI 3% | 1.00 (1.00 – 1.01) | 2.6E-01 | 1.00 (1.00 – 1.01) | 1.6E-02 | 1.00 (1.00 – 1.01) | 1.00 (0.99 – 1.01) |
| 727 | Other disorders of synovium, tendon, and bursa | Per88 | 0.99 (0.98 – 1.01) | 2.6E-01 | 0.99 (0.98 – 1.01) | 9.0E-01 | 0.99 (0.97 – 1.01) | 0.99 (0.97 – 1.01) |
| 793.2 | Nonspecific abnormal findings on radiological and other examination of other intrathoracic organs (echocardiogram, etc) | Per88 | 1.01 (1.00 – 1.02) | 2.7E-01 | 1.01 (1.00 – 1.02) | 4.6E-01 | 1.00 (0.98 – 1.02) | 1.01 (1.00 – 1.03) |
| 244.4 | Hypothyroidism NOS | AHI 3% | 1.00 (1.00 – 1.00) | 2.7E-01 | 1.00 (1.00 – 1.00) | 3.7E-01 | 1.00 (0.99 – 1.00) | 1.00 (1.00 – 1.00) |
| 271.3 | Intestinal disaccharidase deficiencies and disaccharide malabsorption | Per88 | 1.00 (1.00 – 1.01) | 2.7E-01 | 1.00 (1.00 – 1.01) | 2.6E-01 | 1.00 (1.00 – 1.01) | 1.00 (1.00 – 1.01) |
| 172.22 | Squamous cell carcinoma | AHI 3% | 1.00 (0.99 – 1.00) | 2.7E-01 | 1.00 (0.99 – 1.00) | 7.4E-01 | 1.00 (0.99 – 1.00) | 1.00 (0.99 – 1.01) |
| 292.1 | Aphasia/speech disturbance | Per88 | 0.99 (0.98 – 1.01) | 2.8E-01 | 0.99 (0.98 – 1.00) | 6.4E-02 | 0.98 (0.96 – 1.00) | 1.00 (0.98 – 1.02) |
| 464 | Acute sinusitis | Per88 | 0.99 (0.99 – 1.00) | 2.8E-01 | 0.99 (0.98 – 1.00) | 2.4E-01 | 1.00 (0.98 – 1.02) | 0.99 (0.98 – 1.00) |
| 512.2 | Painful respiration | Per88 | 1.00 (1.00 – 1.01) | 2.8E-01 | 1.00 (1.00 – 1.01) | 3.3E-01 | 1.01 (1.00 – 1.02) | 1.00 (0.99 – 1.01) |
| 763 | Thoracic or lumbosacral neuritis or radiculitis, unspecified | Per88 | 1.00 (0.99 – 1.00) | 2.8E-01 | 1.00 (0.99 – 1.00) | 8.1E-01 | 0.99 (0.98 – 1.01) | 0.99 (0.98 – 1.01) |

| PheCode | Disease | Sleep statistic | Risk ratio | P-value | Risk ratio (adj pulmonary disorders) | Sleep statistic x sex interaction p-value | Risk ratio (Men) | Risk ratio (Women) |
|---|---|---|---|---|---|---|---|---|
| 740.1 | Osteoarthritis; localized | AHI 3% | 1.00 (0.99 – 1.00) | 2.8E-01 | 1.00 (0.99 – 1.00) | 4.5E-01 | 0.99 (0.99 – 1.00) | 1.00 (0.99 – 1.01) |
| 706.1 | Acne | AHI 3% | 1.00 (1.00 – 1.00) | 2.8E-01 | 1.00 (1.00 – 1.00) | 7.2E-02 | 1.00 (0.99 – 1.00) | 1.00 (1.00 – 1.00) |
| 720.1 | Spinal stenosis of lumbar region | AHI 3% | 1.00 (0.99 – 1.00) | 2.8E-01 | 1.00 (0.99 – 1.00) | 3.9E-01 | 1.00 (0.99 – 1.00) | 1.00 (0.99 – 1.01) |
| 296.22 | Major depressive disorder | AHI 3% | 1.00 (1.00 – 1.00) | 2.8E-01 | 1.00 (1.00 – 1.00) | 3.3E-02 | 1.00 (1.00 – 1.00) | 1.00 (1.00 – 1.00) |
| 280.2 | Iron deficiency anemia secondary to blood loss (chronic) | AHI 3% | 1.00 (0.99 – 1.00) | 2.8E-01 | 1.00 (0.99 – 1.00) | 8.7E-03 | 1.01 (1.00 – 1.02) | 0.99 (0.99 – 1.00) |
| 389 | Hearing loss | AHI 3% | 1.00 (1.00 – 1.00) | 2.9E-01 | 1.00 (1.00 – 1.00) | 3.4E-02 | 1.00 (1.00 – 1.00) | 1.01 (1.00 – 1.01) |
| 433.21 | Cerebral artery occlusion, with cerebral infarction | Per88 | 1.01 (1.00 – 1.02) | 2.9E-01 | 1.01 (0.99 – 1.02) | 9.3E-01 | 1.01 (0.99 – 1.02) | 1.01 (0.99 – 1.03) |
| 727.4 | Ganglion and cyst of synovium, tendon, and bursa | AHI 3% | 1.00 (0.99 – 1.00) | 2.9E-01 | 1.00 (0.99 – 1.00) | 9.2E-01 | 1.00 (0.99 – 1.01) | 1.00 (0.99 – 1.00) |
| 701 | Other hypertrophic and atrophic conditions of skin | AHI 3% | 1.00 (1.00 – 1.00) | 2.9E-01 | 1.00 (1.00 – 1.00) | 9.5E-03 | 1.00 (1.00 – 1.01) | 1.00 (1.00 – 1.00) |
| 296.1 | Bipolar | AHI 3% | 1.00 (0.99 – 1.00) | 2.9E-01 | 1.00 (0.99 – 1.00) | 7.8E-03 | 1.00 (0.99 – 1.00) | 1.00 (0.99 – 1.01) |
| 550.2 | Diaphragmatic hernia | AHI 3% | 1.00 (0.99 – 1.00) | 2.9E-01 | 1.00 (0.99 – 1.00) | 3.1E-01 | 1.00 (0.99 – 1.00) | 1.00 (0.99 – 1.00) |
| 594.1 | Calculus of kidney | AHI 3% | 1.00 (1.00 – 1.00) | 2.9E-01 | 1.00 (1.00 – 1.00) | 5.3E-01 | 1.00 (1.00 – 1.01) | 1.00 (1.00 – 1.01) |
| 536.8 | Dyspepsia and other specified disorders of function of stomach | AHI 3% | 1.00 (0.99 – 1.00) | 3.0E-01 | 1.00 (0.99 – 1.00) | 5.4E-01 | 1.00 (1.00 – 1.01) | 0.99 (0.99 – 1.00) |
| 598.9 | Other nonspecific findings on examination of urine | Per88 | 0.99 (0.98 – 1.01) | 3.0E-01 | 0.99 (0.98 – 1.01) | 1.5E-01 | 0.99 (0.96 – 1.01) | 1.00 (0.98 – 1.01) |
| 008 | Intestinal infection | Per88 | 1.00 (1.00 – 1.01) | 3.0E-01 | 1.00 (1.00 – 1.01) | 6.1E-02 | 0.99 (0.97 – 1.01) | 1.01 (1.00 – 1.02) |
| 272.12 | Hyperglyceridemia | Per88 | 1.01 (0.99 – 1.02) | 3.0E-01 | 1.01 (0.99 – 1.02) | 2.6E-01 | 1.01 (1.00 – 1.03) | 0.98 (0.95 – 1.01) |
| 496.3 | Bronchiectasis | AHI 3% | 1.00 (0.99 – 1.00) | 3.0E-01 | 1.00 (0.99 – 1.00) | 6.6E-01 | 1.00 (0.99 – 1.01) | 0.99 (0.98 – 1.01) |
| 599.5 | Frequency of urination and polyuria | AHI 3% | 1.00 (1.00 – 1.00) | 3.0E-01 | 1.00 (1.00 – 1.00) | 6.7E-01 | 1.00 (1.00 – 1.00) | 1.00 (1.00 – 1.01) |
| 741.2 | Stiffness of joint | AHI 3% | 1.00 (0.99 – 1.00) | 3.0E-01 | 1.00 (0.99 – 1.00) | 9.7E-01 | 1.00 (0.99 – 1.00) | 1.00 (0.99 – 1.01) |
| 355.1 | Chronic pain syndrome | AHI 3% | 1.00 (1.00 – 1.01) | 3.0E-01 | 1.00 (1.00 – 1.01) | 9.0E-01 | 1.00 (0.99 – 1.01) | 1.00 (0.99 – 1.01) |
| 704.1 | Alopecia | Per88 | 1.00 (0.99 – 1.00) | 3.0E-01 | 1.00 (0.99 – 1.00) | 3.2E-01 | 1.00 (0.99 – 1.01) | 0.99 (0.99 – 1.00) |
| 504 | Other alveolar and parietoalveolar pneumonopathy | AHI 3% | 1.00 (0.99 – 1.00) | 3.1E-01 | 1.00 (0.99 – 1.00) | 4.6E-01 | 0.99 (0.98 – 1.01) | 1.00 (0.99 – 1.01) |
| 475.9 | Postnasal drip | Per88 | 1.00 (1.00 – 1.01) | 3.1E-01 | 1.00 (0.99 – 1.01) | 5.8E-01 | 1.00 (0.99 – 1.02) | 1.00 (0.99 – 1.01) |
| 947 | Urticaria | Per88 | 1.00 (0.99 – 1.00) | 3.1E-01 | 0.99 (0.99 – 1.00) | 6.0E-01 | 0.99 (0.97 – 1.01) | 1.00 (0.99 – 1.01) |
| 1004 | Other signs and symptoms involving emotional state | AHI 3% | 1.00 (0.99 – 1.00) | 3.1E-01 | 1.00 (0.99 – 1.00) | 4.7E-01 | 1.00 (0.99 – 1.01) | 0.99 (0.98 – 1.00) |
| 277.7 | Dysmetabolic syndrome X | AHI 3% | 1.00 (1.00 – 1.01) | 3.1E-01 | 1.00 (1.00 – 1.01) | 2.1E-02 | 1.01 (1.00 – 1.02) | 1.00 (0.99 – 1.01) |
| 702.1 | Actinic keratosis | AHI 3% | 1.00 (0.99 – 1.00) | 3.2E-01 | 1.00 (0.99 – 1.00) | 4.0E-01 | 1.00 (1.00 – 1.00) | 1.00 (0.99 – 1.01) |
| 110.12 | Althete's foot | Per88 | 1.01 (0.99 – 1.02) | 3.2E-01 | 1.01 (0.99 – 1.02) | 1.9E-01 | 1.01 (0.99 – 1.02) | 1.00 (0.98 – 1.02) |
| 470 | Septal Deviations/Turbinate Hypertrophy | AHI 3% | 1.00 (1.00 – 1.00) | 3.2E-01 | 1.00 (1.00 – 1.00) | 9.6E-01 | 1.00 (1.00 – 1.01) | 1.00 (1.00 – 1.00) |
| 687.2 | Localized superficial swelling, mass, or lump | AHI 3% | 1.00 (1.00 – 1.00) | 3.2E-01 | 1.00 (1.00 – 1.00) | 5.2E-02 | 1.00 (1.00 – 1.00) | 1.00 (0.99 – 1.00) |

| PheCode | Disease | Sleep statistic | Risk ratio | P-value | Risk ratio (adj pulmonary disorders) | Sleep statistic x sex interaction p-value | Risk ratio (Men) | Risk ratio (Women) |
|---|---|---|---|---|---|---|---|---|
| 334 | Degenerative disease of the spinal cord | Per88 | 1.01 (0.99 – 1.03) | 3.2E-01 | 1.01 (0.99 – 1.02) | 4.9E-01 | 1.01 (0.99 – 1.03) | 1.01 (0.98 – 1.04) |
| 173 | Neoplasm of uncertain behavior of skin | AHI 3% | 1.00 (1.00 – 1.00) | 3.2E-01 | 1.00 (1.00 – 1.00) | 8.3E-01 | 1.00 (1.00 – 1.01) | 1.00 (1.00 – 1.00) |
| 706.1 | Acne | Per88 | 1.00 (0.99 – 1.00) | 3.2E-01 | 1.00 (0.99 – 1.00) | 2.7E-01 | 0.99 (0.98 – 1.01) | 1.00 (0.99 – 1.01) |
| 259 | Other endocrine disorders | AHI 3% | 1.00 (0.99 – 1.00) | 3.2E-01 | 1.00 (0.99 – 1.00) | 3.7E-02 | 1.00 (0.99 – 1.01) | 0.99 (0.98 – 1.00) |
| 788 | Syncope and collapse | AHI 3% | 1.00 (1.00 – 1.00) | 3.2E-01 | 1.00 (1.00 – 1.00) | 4.1E-02 | 1.00 (1.00 – 1.00) | 1.00 (0.99 – 1.00) |
| 805 | Fracture of vertebral column without mention of spinal cord injury | AHI 3% | 1.00 (1.00 – 1.01) | 3.2E-01 | 1.00 (1.00 – 1.01) | 6.2E-01 | 1.00 (0.99 – 1.01) | 1.00 (0.99 – 1.01) |
| 773 | Pain in limb | Per88 | 1.00 (1.00 – 1.01) | 3.3E-01 | 1.00 (1.00 – 1.01) | 4.3E-01 | 1.00 (0.99 – 1.01) | 1.00 (1.00 – 1.01) |
| 550.5 | Ventral hernia | Per88 | 1.01 (0.99 – 1.02) | 3.3E-01 | 1.01 (0.99 – 1.02) | 5.6E-01 | 1.02 (1.00 – 1.04) | 1.00 (0.98 – 1.02) |
| 447 | Other disorders of arteries and arterioles | Per88 | 0.99 (0.97 – 1.01) | 3.3E-01 | 0.99 (0.97 – 1.01) | 9.2E-01 | 0.98 (0.95 – 1.01) | 1.00 (0.97 – 1.03) |
| 356 | Hereditary and idiopathic peripheral neuropathy | AHI 3% | 1.00 (0.99 – 1.00) | 3.3E-01 | 1.00 (0.99 – 1.00) | 3.8E-01 | 1.00 (0.99 – 1.00) | 1.00 (0.99 – 1.01) |
| 433.31 | Transient cerebral ischemia | AHI 3% | 1.00 (1.00 – 1.01) | 3.3E-01 | 1.00 (1.00 – 1.01) | 7.2E-01 | 1.00 (0.99 – 1.00) | 1.00 (1.00 – 1.01) |
| 110.11 | Dermatophytosis of nail | Per88 | 1.01 (0.99 – 1.02) | 3.4E-01 | 1.01 (1.00 – 1.02) | 3.8E-01 | 1.00 (0.99 – 1.02) | 1.01 (0.99 – 1.03) |
| 574.1 | Cholelithiasis | Per88 | 0.99 (0.98 – 1.01) | 3.4E-01 | 0.99 (0.98 – 1.00) | 3.0E-01 | 1.01 (0.99 – 1.02) | 0.99 (0.98 – 1.00) |
| 411.8 | Other chronic ischemic heart disease, unspecified | Per88 | 1.01 (0.99 – 1.03) | 3.4E-01 | 1.01 (0.99 – 1.03) | 7.7E-01 | 1.01 (0.99 – 1.03) | 1.02 (0.98 – 1.06) |
| 465.4 | Acute laryngitis and tracheitis | AHI 3% | 1.00 (1.00 – 1.01) | 3.4E-01 | 1.00 (1.00 – 1.01) | 9.2E-02 | 1.01 (1.00 – 1.02) | 1.00 (0.99 – 1.01) |
| 054 | Herpes simplex | AHI 3% | 1.00 (1.00 – 1.00) | 3.5E-01 | 1.00 (1.00 – 1.00) | 2.4E-01 | 1.00 (1.00 – 1.01) | 1.00 (0.99 – 1.00) |
| 573.7 | Abnormal results of function study of liver | Per88 | 1.01 (0.99 – 1.02) | 3.5E-01 | 1.00 (0.99 – 1.02) | 6.3E-01 | 1.01 (0.99 – 1.02) | 1.00 (0.98 – 1.02) |
| 272.13 | Mixed hyperlipidemia | Per88 | 1.00 (1.00 – 1.01) | 3.5E-01 | 1.00 (0.99 – 1.01) | 4.7E-01 | 1.01 (1.00 – 1.02) | 1.00 (0.98 – 1.02) |
| 530.11 | GERD | AHI 3% | 1.00 (1.00 – 1.00) | 3.5E-01 | 1.00 (1.00 – 1.00) | 5.9E-01 | 1.00 (1.00 – 1.00) | 1.00 (1.00 – 1.00) |
| 054 | Herpes simplex | Per88 | 1.00 (0.99 – 1.00) | 3.5E-01 | 1.00 (0.99 – 1.00) | 6.6E-02 | 1.01 (1.00 – 1.03) | 0.99 (0.98 – 1.00) |
| 197 | Chemotherapy | Per88 | 1.01 (0.99 – 1.02) | 3.5E-01 | 1.00 (0.99 – 1.02) | 5.3E-02 | 0.99 (0.97 – 1.01) | 1.02 (1.00 – 1.03) |
| 512.7 | Shortness of breath | AHI 3% | 1.00 (1.00 – 1.00) | 3.5E-01 | 1.00 (1.00 – 1.00) | 3.4E-04 | 1.00 (1.00 – 1.01) | 1.00 (0.99 – 1.00) |
| 365 | Glaucoma | AHI 3% | 1.00 (1.00 – 1.00) | 3.5E-01 | 1.00 (1.00 – 1.00) | 2.1E-01 | 1.00 (1.00 – 1.01) | 1.00 (0.99 – 1.00) |
| 246 | Other disorders of thyroid | Per88 | 1.01 (0.99 – 1.02) | 3.6E-01 | 1.01 (0.99 – 1.02) | 6.7E-01 | 1.01 (0.98 – 1.04) | 1.01 (0.99 – 1.02) |
| 153.2 | Colon cancer | AHI 3% | 1.00 (1.00 – 1.01) | 3.6E-01 | 1.00 (1.00 – 1.01) | 1.9E-01 | 1.00 (0.99 – 1.01) | 1.01 (1.00 – 1.03) |
| 706.8 | Other specified diseases of sebaceous glands | AHI 3% | 1.00 (1.00 – 1.00) | 3.6E-01 | 1.00 (1.00 – 1.00) | 6.2E-01 | 1.00 (1.00 – 1.00) | 1.00 (0.99 – 1.00) |
| 388 | Other disorders of ear | AHI 3% | 1.00 (0.99 – 1.00) | 3.6E-01 | 1.00 (0.99 – 1.00) | 4.7E-01 | 1.00 (0.99 – 1.01) | 1.00 (0.99 – 1.00) |
| 008 | Intestinal infection | AHI 3% | 1.00 (1.00 – 1.01) | 3.6E-01 | 1.00 (1.00 – 1.01) | 1.5E-01 | 1.00 (1.00 – 1.01) | 1.00 (0.99 – 1.00) |
| 197 | Chemotherapy | AHI 3% | 1.00 (1.00 – 1.01) | 3.6E-01 | 1.00 (1.00 – 1.01) | 1.2E-01 | 1.00 (0.99 – 1.00) | 1.01 (1.00 – 1.01) |
| 306.9 | Tension headache | AHI 3% | 1.00 (0.99 – 1.00) | 3.7E-01 | 1.00 (0.99 – 1.00) | 8.7E-01 | 1.00 (0.99 – 1.01) | 1.00 (0.99 – 1.00) |
| 471 | Nasal polyps | Per88 | 0.99 (0.98 – 1.01) | 3.7E-01 | 0.99 (0.97 – 1.01) | 4.2E-01 | 1.00 (0.98 – 1.02) | 0.98 (0.96 – 1.01) |
| 537 | Other disorders of stomach and duodenum | AHI 3% | 1.00 (1.00 – 1.00) | 3.7E-01 | 1.00 (1.00 – 1.00) | 8.1E-02 | 1.00 (1.00 – 1.01) | 1.00 (0.99 – 1.00) |
| 345.1 | Epilepsy | Per88 | 0.99 (0.98 – 1.01) | 3.7E-01 | 0.99 (0.98 – 1.01) | 9.0E-02 | 0.98 (0.96 – 1.00) | 1.00 (0.98 – 1.02) |
| 599.8 | Other symptoms involving urinary system | Per88 | 1.01 (0.99 – 1.02) | 3.7E-01 | 1.01 (0.99 – 1.02) | 9.7E-01 | 1.01 (0.99 – 1.02) | 1.01 (0.99 – 1.02) |

| PheCode | Disease | Sleep statistic | Risk ratio | P-value | Risk ratio (adj pulmonary disorders) | Sleep statistic x sex interaction p-value | Risk ratio (Men) | Risk ratio (Women) |
|---|---|---|---|---|---|---|---|---|
| 561 | Symptoms involving digestive system | Per88 | 1.00 (0.99 – 1.01) | 3.8E-01 | 1.00 (0.99 – 1.01) | 9.0E-01 | 1.00 (0.99 – 1.02) | 0.99 (0.97 – 1.00) |
| 287.1 | Spontaneous ecchymoses | AHI 3% | 1.00 (0.99 – 1.00) | 3.8E-01 | 1.00 (0.99 – 1.00) | 7.2E-02 | 1.00 (0.99 – 1.02) | 0.99 (0.98 – 1.00) |
| 722.9 | Other and unspecified disc disorder | Per88 | 1.01 (0.99 – 1.02) | 3.8E-01 | 1.01 (0.99 – 1.02) | 9.9E-01 | 1.01 (1.00 – 1.03) | 1.00 (0.97 – 1.02) |
| 379.2 | Disorders of vitreous body | AHI 3% | 1.00 (0.99 – 1.00) | 3.8E-01 | 1.00 (0.99 – 1.00) | 8.7E-01 | 1.00 (0.99 – 1.01) | 0.99 (0.99 – 1.00) |
| 389.1 | Sensorineural hearing loss | Per88 | 1.00 (0.99 – 1.01) | 3.8E-01 | 1.00 (0.99 – 1.01) | 1.9E-01 | 0.99 (0.98 – 1.00) | 1.01 (0.99 – 1.02) |
| 695.8 | Other specified erythematous conditions | Per88 | 0.99 (0.97 – 1.01) | 3.9E-01 | 0.99 (0.97 – 1.01) | 9.1E-01 | 0.98 (0.94 – 1.01) | 1.00 (0.97 – 1.02) |
| 251.1 | Hypoglycemia | AHI 3% | 1.00 (1.00 – 1.01) | 3.9E-01 | 1.00 (1.00 – 1.01) | 1.4E-02 | 1.01 (1.00 – 1.03) | 0.99 (0.99 – 1.00) |
| 293 | Symptoms involving head and neck | AHI 3% | 1.00 (0.99 – 1.00) | 3.9E-01 | 1.00 (0.99 – 1.00) | 1.1E-01 | 0.99 (0.98 – 1.00) | 1.00 (0.99 – 1.01) |
| 705.8 | Hyperhidrosis | AHI 3% | 1.00 (0.99 – 1.00) | 3.9E-01 | 1.00 (0.99 – 1.00) | 2.9E-01 | 1.00 (0.99 – 1.00) | 1.00 (0.99 – 1.00) |
| 591 | Urinary tract infection | Per88 | 1.00 (1.00 – 1.01) | 4.0E-01 | 1.00 (1.00 – 1.01) | 9.7E-01 | 0.99 (0.98 – 1.01) | 1.01 (1.00 – 1.01) |
| 374.3 | Ptosis of eyelid | AHI 3% | 1.00 (0.99 – 1.00) | 4.0E-01 | 1.00 (0.99 – 1.00) | 2.3E-01 | 1.00 (0.99 – 1.01) | 1.00 (0.99 – 1.01) |
| 735.1 | Flat foot | AHI 3% | 1.00 (0.99 – 1.00) | 4.0E-01 | 1.00 (0.99 – 1.00) | 8.6E-01 | 1.00 (0.99 – 1.01) | 1.00 (0.99 – 1.01) |
| 386.2 | Peripheral or central vertigo | Per88 | 0.99 (0.98 – 1.01) | 4.0E-01 | 0.99 (0.98 – 1.01) | 7.5E-01 | 0.99 (0.97 – 1.01) | 1.00 (0.98 – 1.02) |
| 395.2 | Nonrheumatic aortic valve disorders | Per88 | 1.01 (0.99 – 1.02) | 4.0E-01 | 1.01 (0.99 – 1.02) | 1.6E-01 | 1.00 (0.98 – 1.01) | 1.02 (1.00 – 1.05) |
| 286.81 | Primary hypercoagulable state | Per88 | 0.99 (0.97 – 1.01) | 4.1E-01 | 0.99 (0.97 – 1.01) | 7.1E-02 | 0.97 (0.93 – 1.00) | 1.00 (0.98 – 1.03) |
| 379.9 | Pain, swelling or discharge of eye | Per88 | 1.00 (0.99 – 1.01) | 4.1E-01 | 0.99 (0.99 – 1.00) | 1.8E-01 | 0.98 (0.96 – 1.00) | 1.00 (0.99 – 1.01) |
| 348.8 | Encephalopathy, not elsewhere classified | Per88 | 1.01 (0.99 – 1.02) | 4.1E-01 | 1.00 (0.99 – 1.02) | 3.9E-02 | 0.99 (0.97 – 1.01) | 1.03 (1.00 – 1.05) |
| 722.1 | Displacement of intervertebral disc | Per88 | 1.00 (0.99 – 1.01) | 4.1E-01 | 1.00 (0.99 – 1.01) | 9.9E-02 | 0.99 (0.98 – 1.00) | 1.00 (0.99 – 1.02) |
| 356 | Hereditary and idiopathic peripheral neuropathy | Per88 | 1.01 (0.99 – 1.02) | 4.2E-01 | 1.01 (0.99 – 1.02) | 2.1E-02 | 1.01 (1.00 – 1.03) | 0.99 (0.97 – 1.01) |
| 565 | Anal and rectal conditions | AHI 3% | 1.00 (1.00 – 1.01) | 4.2E-01 | 1.00 (1.00 – 1.01) | 3.4E-03 | 1.00 (1.00 – 1.01) | 1.00 (0.99 – 1.00) |
| 250.41 | Impaired fasting glucose | AHI 3% | 1.00 (1.00 – 1.01) | 4.2E-01 | 1.00 (1.00 – 1.01) | 8.6E-01 | 1.00 (0.99 – 1.00) | 1.01 (1.00 – 1.01) |
| 443.9 | Peripheral vascular disease, unspecified | AHI 3% | 1.00 (1.00 – 1.01) | 4.2E-01 | 1.00 (1.00 – 1.01) | 7.0E-01 | 1.00 (0.99 – 1.01) | 1.00 (0.99 – 1.02) |
| 475 | Chronic sinusitis | AHI 3% | 1.00 (1.00 – 1.00) | 4.2E-01 | 1.00 (1.00 – 1.00) | 2.4E-01 | 1.00 (1.00 – 1.00) | 1.00 (0.99 – 1.00) |
| 562.2 | Diverticulitis | AHI 3% | 1.00 (1.00 – 1.01) | 4.2E-01 | 1.00 (1.00 – 1.01) | 5.1E-01 | 1.00 (0.99 – 1.00) | 1.00 (1.00 – 1.01) |
| 727 | Other disorders of synovium, tendon, and bursa | AHI 3% | 1.00 (0.99 – 1.00) | 4.2E-01 | 1.00 (0.99 – 1.00) | 5.5E-01 | 1.00 (0.99 – 1.01) | 1.00 (0.99 – 1.00) |
| 261.4 | Vitamin D deficiency | AHI 3% | 1.00 (1.00 – 1.00) | 4.2E-01 | 1.00 (1.00 – 1.00) | 5.0E-01 | 1.00 (1.00 – 1.01) | 1.00 (1.00 – 1.00) |
| 277.7 | Dysmetabolic syndrome X | Per88 | 0.99 (0.97 – 1.01) | 4.3E-01 | 0.99 (0.97 – 1.01) | 9.6E-02 | 0.99 (0.95 – 1.03) | 0.99 (0.97 – 1.02) |
| 841 | Sprains and strains of back and neck | AHI 3% | 1.00 (1.00 – 1.01) | 4.3E-01 | 1.00 (1.00 – 1.01) | 4.0E-01 | 1.01 (1.00 – 1.02) | 1.00 (0.99 – 1.01) |
| 369.5 | Conjunctivitis, infectious | Per88 | 1.00 (0.99 – 1.01) | 4.3E-01 | 0.99 (0.98 – 1.00) | 2.1E-01 | 1.00 (0.99 – 1.02) | 0.99 (0.98 – 1.00) |
| 770 | Myalgia and myositis unspecified | Per88 | 1.00 (0.99 – 1.00) | 4.3E-01 | 1.00 (0.99 – 1.00) | 2.9E-01 | 1.01 (0.99 – 1.02) | 0.99 (0.98 – 1.00) |
| 599.4 | Urinary incontinence | Per88 | 1.00 (1.00 – 1.01) | 4.3E-01 | 1.00 (1.00 – 1.01) | 8.7E-01 | 1.00 (0.98 – 1.02) | 1.00 (0.99 – 1.01) |
| 706.8 | Other specified diseases of sebaceous glands | Per88 | 1.00 (0.99 – 1.00) | 4.3E-01 | 1.00 (0.99 – 1.00) | 5.8E-01 | 1.00 (0.99 – 1.01) | 1.00 (0.98 – 1.01) |
| 687.1 | Rash and other nonspecific skin eruption | Per88 | 1.00 (1.00 – 1.01) | 4.3E-01 | 1.00 (1.00 – 1.01) | 8.8E-01 | 1.00 (0.99 – 1.01) | 1.00 (1.00 – 1.01) |
| 216 | Benign neoplasm of skin | AHI 3% | 1.00 (1.00 – 1.00) | 4.3E-01 | 1.00 (1.00 – 1.00) | 4.7E-01 | 1.00 (1.00 – 1.00) | 1.00 (1.00 – 1.00) |
| 550.2 | Diaphragmatic hernia | Per88 | 1.00 (0.99 – 1.01) | 4.4E-01 | 1.00 (0.99 – 1.01) | 9.7E-01 | 1.00 (0.98 – 1.01) | 1.00 (0.98 – 1.01) |

| PheCode | Disease | Sleep statistic | Risk ratio | P-value | Risk ratio (adj pulmonary disorders) | Sleep statistic x sex interaction p-value | Risk ratio (Men) | Risk ratio (Women) |
|---|---|---|---|---|---|---|---|---|
| 078 | Viral warts & HPV | AHI 3% | 1.00 (1.00 – 1.00) | 4.4E-01 | 1.00 (1.00 – 1.00) | 4.6E-01 | 1.00 (1.00 – 1.00) | 1.00 (0.99 – 1.00) |
| 979 | Adverse drug events and drug allergies | AHI 3% | 1.00 (1.00 – 1.00) | 4.4E-01 | 1.00 (1.00 – 1.00) | 9.4E-01 | 1.00 (1.00 – 1.01) | 1.00 (1.00 – 1.01) |
| 389 | Hearing loss | Per88 | 1.00 (1.00 – 1.01) | 4.4E-01 | 1.00 (1.00 – 1.01) | 6.6E-01 | 1.00 (0.99 – 1.01) | 1.00 (0.99 – 1.02) |
| 418 | Nonspecific chest pain | Per88 | 1.00 (1.00 – 1.01) | 4.4E-01 | 1.00 (1.00 – 1.00) | 3.9E-02 | 1.00 (0.99 – 1.00) | 1.00 (1.00 – 1.01) |
| 765 | Cervical radiculitis | AHI 3% | 1.00 (0.99 – 1.00) | 4.4E-01 | 1.00 (0.99 – 1.00) | 3.0E-01 | 1.00 (0.99 – 1.00) | 1.00 (0.99 – 1.01) |
| 279.2 | Autoimmune disease NEC | AHI 3% | 1.00 (0.99 – 1.00) | 4.4E-01 | 1.00 (0.99 – 1.00) | 4.2E-01 | 1.00 (0.99 – 1.01) | 1.00 (0.99 – 1.01) |
| 411.4 | Coronary atherosclerosis | Per88 | 1.00 (0.99 – 1.01) | 4.5E-01 | 1.00 (0.99 – 1.01) | 2.2E-01 | 1.00 (0.99 – 1.01) | 1.01 (1.00 – 1.03) |
| 481 | Influenza | Per88 | 1.00 (1.00 – 1.01) | 4.5E-01 | 1.00 (0.99 – 1.01) | 8.5E-01 | 1.00 (0.99 – 1.01) | 1.01 (1.00 – 1.02) |
| 705.8 | Hyperhidrosis | Per88 | 1.00 (0.99 – 1.01) | 4.5E-01 | 1.00 (0.99 – 1.01) | 9.2E-01 | 1.00 (0.99 – 1.02) | 1.01 (0.99 – 1.02) |
| 531.2 | Gastric ulcer | AHI 3% | 1.00 (0.99 – 1.00) | 4.5E-01 | 1.00 (0.99 – 1.00) | 1.3E-01 | 0.99 (0.99 – 1.00) | 1.00 (0.99 – 1.01) |
| 367.1 | Myopia | AHI 3% | 1.00 (0.99 – 1.00) | 4.5E-01 | 1.00 (0.99 – 1.00) | 5.9E-02 | 1.00 (1.00 – 1.01) | 0.99 (0.99 – 1.00) |
| 477 | Epistaxis or throat hemorrhage | AHI 3% | 1.00 (1.00 – 1.01) | 4.5E-01 | 1.00 (1.00 – 1.01) | 2.6E-01 | 1.00 (0.99 – 1.00) | 1.01 (1.00 – 1.02) |
| 715 | Other inflammatory spondylopathies | Per88 | 0.99 (0.97 – 1.01) | 4.6E-01 | 0.99 (0.97 – 1.01) | 8.4E-01 | 0.98 (0.95 – 1.02) | 1.00 (0.97 – 1.03) |
| 443.1 | Raynaud's syndrome | Per88 | 1.00 (0.98 – 1.01) | 4.6E-01 | 0.99 (0.98 – 1.01) | 4.2E-01 | 0.98 (0.95 – 1.00) | 1.00 (0.98 – 1.01) |
| 512.2 | Painful respiration | AHI 3% | 1.00 (1.00 – 1.00) | 4.6E-01 | 1.00 (1.00 – 1.00) | 9.5E-02 | 1.00 (1.00 – 1.01) | 1.00 (0.99 – 1.00) |
| 720 | Spinal stenosis | AHI 3% | 1.00 (0.99 – 1.00) | 4.6E-01 | 1.00 (0.99 – 1.00) | 9.2E-03 | 0.99 (0.99 – 1.00) | 1.00 (1.00 – 1.01) |
| 053 | Herpes zoster | Per88 | 1.00 (0.99 – 1.01) | 4.6E-01 | 1.00 (0.99 – 1.01) | 1.9E-01 | 1.00 (0.99 – 1.02) | 1.00 (0.99 – 1.02) |
| 279.2 | Autoimmune disease NEC | Per88 | 1.01 (0.99 – 1.02) | 4.7E-01 | 1.00 (0.99 – 1.02) | 2.2E-01 | 1.02 (1.00 – 1.05) | 1.00 (0.98 – 1.02) |
| 366 | Cataract | AHI 3% | 1.00 (1.00 – 1.00) | 4.7E-01 | 1.00 (1.00 – 1.00) | 9.1E-01 | 1.00 (0.99 – 1.00) | 1.00 (0.99 – 1.00) |
| 562.2 | Diverticulitis | Per88 | 1.00 (0.99 – 1.02) | 4.7E-01 | 1.00 (0.99 – 1.02) | 8.7E-02 | 1.00 (0.98 – 1.02) | 1.00 (0.98 – 1.02) |
| 351 | Other peripheral nerve disorders | Per88 | 1.00 (1.00 – 1.01) | 4.7E-01 | 1.00 (1.00 – 1.01) | 2.6E-01 | 1.00 (0.99 – 1.02) | 1.00 (0.99 – 1.01) |
| 277.51 | Lipoprotein disorders | AHI 3% | 1.00 (1.00 – 1.01) | 4.8E-01 | 1.00 (1.00 – 1.01) | 8.1E-01 | 1.00 (0.99 – 1.01) | 1.00 (0.99 – 1.02) |
| 738.4 | Acquired spondylolisthesis | AHI 3% | 1.00 (0.99 – 1.00) | 4.8E-01 | 1.00 (0.99 – 1.00) | 1.3E-01 | 1.00 (0.99 – 1.00) | 1.00 (0.99 – 1.01) |
| 367.4 | Presbyopia | Per88 | 1.00 (0.99 – 1.02) | 4.8E-01 | 1.00 (0.99 – 1.02) | 1.4E-01 | 1.01 (0.99 – 1.03) | 1.00 (0.98 – 1.01) |
| 781 | Symptoms involving nervous and musculoskeletal systems | AHI 3% | 1.00 (1.00 – 1.00) | 4.8E-01 | 1.00 (1.00 – 1.00) | 2.5E-01 | 1.00 (1.00 – 1.01) | 0.99 (0.99 – 1.00) |
| 443.1 | Raynaud's syndrome | AHI 3% | 1.00 (0.99 – 1.00) | 4.8E-01 | 1.00 (0.99 – 1.00) | 6.1E-01 | 1.00 (0.99 – 1.00) | 1.00 (0.99 – 1.00) |
| 687.2 | Localized superficial swelling, mass, or lump | Per88 | 1.00 (1.00 – 1.01) | 4.8E-01 | 1.00 (1.00 – 1.01) | 1.5E-02 | 1.00 (0.99 – 1.02) | 1.00 (0.99 – 1.01) |
| 429.3 | Symptoms involving cardiovascular system | AHI 3% | 1.00 (1.00 – 1.00) | 4.9E-01 | 1.00 (1.00 – 1.00) | 6.5E-01 | 1.00 (1.00 – 1.01) | 1.00 (1.00 – 1.01) |
| 745 | Pain in joint | Per88 | 1.00 (1.00 – 1.00) | 4.9E-01 | 1.00 (1.00 – 1.00) | 6.6E-01 | 1.00 (0.99 – 1.00) | 1.00 (0.99 – 1.00) |
| 483 | Acute bronchitis and bronchiolitis | AHI 3% | 1.00 (1.00 – 1.00) | 4.9E-01 | 1.00 (1.00 – 1.00) | 1.4E-01 | 1.00 (0.99 – 1.00) | 1.00 (1.00 – 1.01) |
| 703.1 | Ingrowing nail | AHI 3% | 1.00 (0.99 – 1.00) | 4.9E-01 | 1.00 (0.99 – 1.00) | 9.8E-01 | 1.00 (0.99 – 1.01) | 0.99 (0.98 – 1.00) |
| 214 | Lipoma | Per88 | 1.00 (0.99 – 1.02) | 4.9E-01 | 1.00 (0.99 – 1.02) | 9.9E-01 | 1.00 (0.98 – 1.02) | 1.00 (0.98 – 1.02) |
| 738.4 | Acquired spondylolisthesis | Per88 | 1.00 (0.99 – 1.02) | 4.9E-01 | 1.00 (0.99 – 1.02) | 2.6E-01 | 0.99 (0.97 – 1.01) | 1.01 (0.99 – 1.03) |
| 727.1 | Synovitis and tenosynovitis | AHI 3% | 1.00 (1.00 – 1.00) | 4.9E-01 | 1.00 (1.00 – 1.00) | 6.5E-01 | 1.00 (0.99 – 1.00) | 1.00 (1.00 – 1.00) |

| PheCode | Disease | Sleep statistic | Risk ratio | P-value | Risk ratio (adj pulmonary disorders) | Sleep statistic x sex interaction p-value | Risk ratio (Men) | Risk ratio (Women) |
|---|---|---|---|---|---|---|---|---|
| 737.3 | Kyphoscoliosis and scoliosis | AHI 3% | 1.00 (1.00 – 1.01) | 4.9E-01 | 1.00 (1.00 – 1.01) | 2.9E-02 | 0.99 (0.98 – 1.00) | 1.00 (1.00 – 1.01) |
| 008.5 | Bacterial enteritis | AHI 3% | 1.00 (0.99 – 1.00) | 4.9E-01 | 1.00 (0.99 – 1.00) | 4.6E-01 | 1.00 (0.99 – 1.00) | 1.00 (1.00 – 1.01) |
| 571.5 | Other chronic nonalcoholic liver disease | Per88 | 1.00 (0.99 – 1.01) | 4.9E-01 | 1.00 (0.99 – 1.01) | 4.3E-01 | 1.01 (0.99 – 1.02) | 1.00 (0.98 – 1.01) |
| 262 | Mineral deficiency NEC | AHI 3% | 1.00 (0.99 – 1.00) | 4.9E-01 | 1.00 (0.99 – 1.00) | 1.9E-01 | 1.00 (0.99 – 1.01) | 1.00 (0.99 – 1.00) |
| 389.2 | Conductive hearing loss | Per88 | 0.99 (0.97 – 1.01) | 4.9E-01 | 0.99 (0.97 – 1.01) | 3.9E-01 | 0.98 (0.95 – 1.02) | 1.00 (0.97 – 1.03) |
| 380.4 | Impacted cerumen | AHI 3% | 1.00 (1.00 – 1.01) | 4.9E-01 | 1.00 (1.00 – 1.01) | 9.1E-02 | 1.00 (0.99 – 1.00) | 1.00 (1.00 – 1.01) |
| 535.2 | Atrophic gastritis | Per88 | 1.00 (0.98 – 1.01) | 5.0E-01 | 0.99 (0.98 – 1.01) | 9.8E-03 | 0.98 (0.97 – 1.00) | 1.00 (0.99 – 1.02) |
| 241.2 | Nontoxic multinodular goiter | AHI 3% | 1.00 (1.00 – 1.01) | 5.0E-01 | 1.00 (1.00 – 1.01) | 9.9E-01 | 1.00 (0.99 – 1.01) | 1.00 (1.00 – 1.01) |
| 594.3 | Calculus of ureter | AHI 3% | 1.00 (1.00 – 1.01) | 5.0E-01 | 1.00 (1.00 – 1.01) | 2.7E-01 | 1.01 (1.00 – 1.01) | 0.99 (0.98 – 1.01) |
| 760 | Back pain | Per88 | 1.00 (1.00 – 1.01) | 5.0E-01 | 1.00 (1.00 – 1.00) | 6.9E-01 | 1.00 (0.99 – 1.01) | 1.00 (1.00 – 1.01) |
| 153.2 | Colon cancer | Per88 | 1.01 (0.99 – 1.03) | 5.1E-01 | 1.01 (0.99 – 1.03) | 8.6E-01 | 1.00 (0.97 – 1.03) | 1.00 (0.97 – 1.04) |
| 367.8 | Hypermetropia | AHI 3% | 1.00 (0.99 – 1.00) | 5.1E-01 | 1.00 (0.99 – 1.00) | 2.2E-01 | 0.99 (0.99 – 1.00) | 1.00 (0.99 – 1.01) |
| 447.7 | Aortic ectasia | Per88 | 1.01 (0.99 – 1.03) | 5.1E-01 | 1.01 (0.99 – 1.03) | 3.2E-02 | 1.00 (0.98 – 1.02) | 1.04 (1.01 – 1.07) |
| 735.1 | Flat foot | Per88 | 1.01 (0.99 – 1.02) | 5.1E-01 | 1.00 (0.99 – 1.02) | 7.2E-01 | 1.00 (0.98 – 1.03) | 1.01 (0.99 – 1.03) |
| 217.1 | Nevus, non-neoplastic | AHI 3% | 1.00 (1.00 – 1.00) | 5.2E-01 | 1.00 (1.00 – 1.00) | 9.6E-01 | 1.00 (1.00 – 1.01) | 1.00 (1.00 – 1.01) |
| 333.1 | Essential tremor | Per88 | 0.99 (0.97 – 1.02) | 5.2E-01 | 0.99 (0.97 – 1.01) | 3.1E-01 | 1.00 (0.97 – 1.03) | 0.98 (0.94 – 1.02) |
| 287.1 | Spontaneous ecchymoses | Per88 | 1.01 (0.99 – 1.02) | 5.2E-01 | 1.00 (0.99 – 1.02) | 2.9E-01 | 1.01 (0.97 – 1.05) | 1.01 (0.99 – 1.03) |
| 689 | Disorder of skin and subcutaneous tissue NOS | AHI 3% | 1.00 (1.00 – 1.00) | 5.2E-01 | 1.00 (1.00 – 1.00) | 3.4E-01 | 1.00 (1.00 – 1.00) | 1.00 (1.00 – 1.00) |
| 591 | Urinary tract infection | AHI 3% | 1.00 (1.00 – 1.00) | 5.2E-01 | 1.00 (1.00 – 1.00) | 1.8E-06 | 1.00 (1.00 – 1.01) | 1.00 (0.99 – 1.00) |
| 530.14 | Reflux esophagitis | AHI 3% | 1.00 (1.00 – 1.01) | 5.2E-01 | 1.00 (1.00 – 1.01) | 2.0E-01 | 1.00 (0.99 – 1.00) | 1.01 (1.00 – 1.01) |
| 245.21 | Chronic lymphocytic thyroiditis | Per88 | 1.00 (0.98 – 1.01) | 5.2E-01 | 0.99 (0.98 – 1.01) | 3.7E-01 | 0.97 (0.93 – 1.01) | 1.00 (0.98 – 1.02) |
| 941 | Adverse reaction to serum or vaccine | AHI 3% | 1.00 (0.99 – 1.00) | 5.2E-01 | 1.00 (0.99 – 1.00) | 5.9E-01 | 1.00 (0.99 – 1.01) | 1.00 (0.99 – 1.00) |
| 579.8 | Nonspecific abnormal findings in stool contents | AHI 3% | 1.00 (1.00 – 1.00) | 5.3E-01 | 1.00 (1.00 – 1.00) | 8.2E-01 | 1.00 (0.99 – 1.00) | 1.00 (0.99 – 1.00) |
| 440 | Atherosclerosis | AHI 3% | 1.00 (0.99 – 1.01) | 5.3E-01 | 1.00 (0.99 – 1.01) | 4.2E-01 | 1.00 (0.99 – 1.01) | 1.01 (1.00 – 1.02) |
| 350.6 | Disturbances of sensation of smell and taste | Per88 | 0.99 (0.98 – 1.01) | 5.3E-01 | 0.99 (0.98 – 1.01) | 4.2E-01 | 1.00 (0.98 – 1.02) | 0.99 (0.97 – 1.02) |
| 286.9 | Abnormal coagulation profile | AHI 3% | 1.00 (0.99 – 1.00) | 5.3E-01 | 1.00 (0.99 – 1.00) | 7.5E-01 | 1.00 (0.99 – 1.00) | 1.00 (0.99 – 1.01) |
| 110.2 | Dermatomycoses | Per88 | 1.00 (0.99 – 1.02) | 5.3E-01 | 1.01 (0.99 – 1.02) | 7.7E-01 | 1.00 (0.98 – 1.02) | 1.01 (0.99 – 1.03) |
| 433.8 | Late effects of cerebrovascular disease | AHI 3% | 1.00 (1.00 – 1.01) | 5.3E-01 | 1.00 (1.00 – 1.01) | 9.2E-01 | 1.00 (1.00 – 1.01) | 1.00 (0.99 – 1.02) |
| 395.1 | Nonrheumatic mitral valve disorders | AHI 3% | 1.00 (1.00 – 1.00) | 5.3E-01 | 1.00 (0.99 – 1.00) | 7.4E-01 | 1.00 (1.00 – 1.00) | 1.00 (0.99 – 1.00) |
| 350.6 | Disturbances of sensation of smell and taste | AHI 3% | 1.00 (0.99 – 1.00) | 5.3E-01 | 1.00 (0.99 – 1.00) | 7.9E-01 | 1.00 (0.99 – 1.01) | 1.00 (0.99 – 1.01) |
| 333.1 | Essential tremor | AHI 3% | 1.00 (0.99 – 1.01) | 5.3E-01 | 1.00 (0.99 – 1.01) | 3.9E-01 | 1.00 (0.99 – 1.01) | 1.00 (0.98 – 1.01) |
| 380.1 | Otitis externa | Per88 | 1.00 (0.99 – 1.02) | 5.3E-01 | 1.00 (0.99 – 1.02) | 8.9E-01 | 1.00 (0.98 – 1.02) | 1.01 (0.99 – 1.03) |
| 475 | Chronic sinusitis | Per88 | 1.00 (0.99 – 1.01) | 5.3E-01 | 0.99 (0.99 – 1.00) | 5.3E-01 | 1.00 (0.98 – 1.01) | 1.00 (0.99 – 1.01) |
| 473.4 | Voice disturbance | AHI 3% | 1.00 (1.00 – 1.00) | 5.3E-01 | 1.00 (1.00 – 1.00) | 4.2E-01 | 1.00 (1.00 – 1.01) | 1.00 (0.99 – 1.00) |
| 550.5 | Ventral hernia | AHI 3% | 1.00 (0.99 – 1.00) | 5.4E-01 | 1.00 (0.99 – 1.00) | 1.9E-01 | 1.00 (0.99 – 1.00) | 1.00 (0.99 – 1.01) |

| PheCode | Disease | Sleep statistic | Risk ratio | P-value | Risk ratio (adj pulmonary disorders) | Sleep statistic x sex interaction p-value | Risk ratio (Men) | Risk ratio (Women) |
|---|---|---|---|---|---|---|---|---|
| 348.8 | Encephalopathy, not elsewhere classified | AHI 3% | 1.00 (1.00 – 1.01) | 5.4E-01 | 1.00 (1.00 – 1.01) | 3.6E-01 | 1.00 (0.99 – 1.01) | 1.00 (0.99 – 1.01) |
| 593.1 | Gross hematuria | Per88 | 1.00 (0.99 – 1.02) | 5.4E-01 | 1.00 (0.99 – 1.02) | 6.2E-01 | 1.00 (0.98 – 1.02) | 1.01 (0.99 – 1.03) |
| 427.21 | Atrial fibrillation | AHI 3% | 1.00 (1.00 – 1.00) | 5.4E-01 | 1.00 (1.00 – 1.00) | 1.7E-01 | 1.00 (1.00 – 1.00) | 1.00 (1.00 – 1.01) |
| 577.3 | Cyst and pseudocyst of pancreas | AHI 3% | 1.00 (1.00 – 1.01) | 5.4E-01 | 1.00 (1.00 – 1.01) | 3.7E-01 | 1.00 (0.99 – 1.02) | 1.00 (0.99 – 1.01) |
| 531.4 | Peptic ulcer, site unspecified | AHI 3% | 1.00 (0.99 – 1.01) | 5.4E-01 | 1.00 (0.99 – 1.01) | 7.9E-01 | 0.99 (0.98 – 1.00) | 1.00 (0.99 – 1.01) |
| 716.9 | Arthropathy NOS | Per88 | 1.00 (0.99 – 1.02) | 5.5E-01 | 1.00 (0.99 – 1.02) | 6.2E-01 | 1.01 (0.98 – 1.03) | 1.00 (0.98 – 1.03) |
| 455 | Hemorrhoids | Per88 | 1.00 (0.99 – 1.00) | 5.5E-01 | 1.00 (0.99 – 1.00) | 7.8E-02 | 1.00 (0.99 – 1.01) | 0.99 (0.98 – 1.00) |
| 350.3 | Lack of coordination | Per88 | 0.99 (0.98 – 1.01) | 5.6E-01 | 0.99 (0.98 – 1.01) | 8.7E-01 | 1.00 (0.98 – 1.02) | 0.99 (0.97 – 1.02) |
| 696.41 | Psoriasis vulgaris | Per88 | 1.00 (0.98 – 1.01) | 5.6E-01 | 0.99 (0.98 – 1.01) | 3.7E-01 | 1.00 (0.98 – 1.01) | 0.99 (0.97 – 1.01) |
| 426.31 | Right bundle branch block | AHI 3% | 1.00 (1.00 – 1.01) | 5.6E-01 | 1.00 (1.00 – 1.01) | 3.9E-01 | 1.00 (1.00 – 1.01) | 1.00 (0.99 – 1.01) |
| 742.9 | Other derangement of joint | AHI 3% | 1.00 (1.00 – 1.00) | 5.6E-01 | 1.00 (1.00 – 1.00) | 1.2E-01 | 1.00 (0.99 – 1.00) | 1.00 (1.00 – 1.01) |
| 510 | Other diseases of lung | AHI 3% | 1.00 (0.99 – 1.00) | 5.7E-01 | 1.00 (0.99 – 1.00) | 7.6E-01 | 1.00 (0.99 – 1.00) | 1.00 (0.99 – 1.01) |
| 574.3 | Cholecystitis without cholelithiasis | AHI 3% | 1.00 (0.99 – 1.00) | 5.7E-01 | 1.00 (0.99 – 1.00) | 4.2E-02 | 1.00 (0.99 – 1.01) | 1.00 (0.99 – 1.00) |
| 367.2 | Astigmatism | Per88 | 1.00 (0.99 – 1.01) | 5.7E-01 | 1.00 (0.99 – 1.01) | 7.0E-01 | 1.00 (0.99 – 1.02) | 1.00 (0.99 – 1.02) |
| 313.1 | Attention deficit hyperactivity disorder | Per88 | 1.00 (0.99 – 1.00) | 5.7E-01 | 1.00 (0.99 – 1.00) | 1.4E-01 | 0.99 (0.98 – 1.00) | 1.00 (0.99 – 1.01) |
| 596 | Other disorders of bladder | Per88 | 1.00 (0.99 – 1.02) | 5.8E-01 | 1.01 (0.99 – 1.02) | 5.5E-02 | 0.99 (0.98 – 1.01) | 1.03 (1.00 – 1.05) |
| 592.11 | Acute cystitis | AHI 3% | 1.00 (0.99 – 1.00) | 5.8E-01 | 1.00 (0.99 – 1.00) | 3.2E-01 | 1.00 (0.99 – 1.01) | 1.00 (0.99 – 1.00) |
| 471 | Nasal polyps | AHI 3% | 1.00 (0.99 – 1.00) | 5.9E-01 | 1.00 (0.99 – 1.00) | 1.1E-01 | 1.00 (1.00 – 1.01) | 0.99 (0.98 – 1.00) |
| 722.9 | Other and unspecified disc disorder | AHI 3% | 1.00 (0.99 – 1.00) | 5.9E-01 | 1.00 (0.99 – 1.00) | 2.2E-01 | 1.00 (0.99 – 1.01) | 0.99 (0.99 – 1.00) |
| 695.3 | Rosacea | AHI 3% | 1.00 (0.99 – 1.00) | 5.9E-01 | 1.00 (0.99 – 1.00) | 9.8E-01 | 1.00 (1.00 – 1.01) | 1.00 (0.99 – 1.00) |
| 742.9 | Other derangement of joint | Per88 | 1.00 (0.99 – 1.01) | 5.9E-01 | 1.00 (0.99 – 1.01) | 9.7E-01 | 1.00 (0.99 – 1.02) | 0.99 (0.98 – 1.01) |
| 599.9 | Other abnormality of urination | Per88 | 1.00 (0.99 – 1.01) | 5.9E-01 | 1.00 (0.99 – 1.01) | 5.4E-01 | 0.99 (0.98 – 1.01) | 1.00 (0.99 – 1.02) |
| 244.2 | Acquired hypothyroidism | Per88 | 1.00 (0.99 – 1.02) | 6.0E-01 | 1.00 (0.99 – 1.02) | 1.7E-01 | 0.99 (0.96 – 1.02) | 1.01 (0.99 – 1.03) |
| 741.2 | Stiffness of joint | Per88 | 1.00 (0.99 – 1.02) | 6.0E-01 | 1.00 (0.99 – 1.02) | 3.4E-01 | 1.02 (1.00 – 1.03) | 0.99 (0.97 – 1.01) |
| 695.3 | Rosacea | Per88 | 1.00 (0.98 – 1.01) | 6.0E-01 | 1.00 (0.99 – 1.01) | 9.2E-02 | 1.01 (0.99 – 1.03) | 0.98 (0.97 – 1.00) |
| 766 | Neuralgia, neuritis, and radiculitis NOS | AHI 3% | 1.00 (0.99 – 1.00) | 6.0E-01 | 1.00 (0.99 – 1.00) | 5.5E-01 | 1.00 (0.99 – 1.00) | 1.00 (1.00 – 1.01) |
| 557 | Intestinal malabsorption (non-celiac) | Per88 | 1.00 (0.99 – 1.02) | 6.0E-01 | 1.00 (0.99 – 1.01) | 7.9E-01 | 1.00 (0.98 – 1.03) | 1.00 (0.99 – 1.02) |
| 327.4 | Insomnia | Per88 | 1.00 (1.00 – 1.00) | 6.0E-01 | 1.00 (1.00 – 1.00) | 1.8E-01 | 1.00 (0.99 – 1.00) | 1.00 (0.99 – 1.00) |
| 789 | Nausea and vomiting | Per88 | 1.00 (1.00 – 1.01) | 6.0E-01 | 1.00 (0.99 – 1.00) | 9.3E-01 | 1.00 (0.99 – 1.01) | 1.00 (1.00 – 1.01) |
| 262 | Mineral deficiency NEC | Per88 | 1.00 (0.99 – 1.01) | 6.0E-01 | 1.00 (0.99 – 1.01) | 6.2E-01 | 1.01 (0.99 – 1.03) | 1.00 (0.99 – 1.02) |
| 946 | Anaphylactic shock NOS | Per88 | 1.00 (0.98 – 1.01) | 6.0E-01 | 0.99 (0.98 – 1.01) | 7.9E-01 | 0.98 (0.95 – 1.01) | 1.01 (0.99 – 1.02) |
| 366.2 | Senile cataract | Per88 | 1.00 (0.99 – 1.01) | 6.0E-01 | 1.00 (0.99 – 1.01) | 3.4E-01 | 1.01 (1.00 – 1.02) | 1.00 (0.98 – 1.01) |
| 288.1 | Decreased white blood cell count | Per88 | 1.00 (0.99 – 1.02) | 6.0E-01 | 1.00 (0.99 – 1.02) | 1.9E-01 | 1.00 (0.98 – 1.03) | 1.01 (0.99 – 1.03) |
| 411.3 | Angina pectoris | AHI 3% | 1.00 (1.00 – 1.00) | 6.1E-01 | 1.00 (1.00 – 1.00) | 2.7E-01 | 1.00 (1.00 – 1.00) | 1.00 (0.99 – 1.01) |
| 599.2 | Retention of urine | AHI 3% | 1.00 (0.99 – 1.00) | 6.1E-01 | 1.00 (0.99 – 1.00) | 4.7E-02 | 1.00 (1.00 – 1.01) | 1.00 (0.99 – 1.00) |

| PheCode | Disease | Sleep statistic | Risk ratio | P-value | Risk ratio (adj pulmonary disorders) | Sleep statistic x sex interaction p-value | Risk ratio (Men) | Risk ratio (Women) |
|---|---|---|---|---|---|---|---|---|
| 426 | Cardiac conduction disorders | Per88 | 1.01 (0.98 – 1.03) | 6.2E-01 | 1.01 (0.98 – 1.03) | 6.5E-01 | 1.00 (0.97 – 1.02) | 1.03 (0.99 – 1.06) |
| 599.5 | Frequency of urination and polyuria | Per88 | 1.00 (0.99 – 1.01) | 6.2E-01 | 1.00 (0.99 – 1.01) | 8.3E-01 | 1.00 (0.99 – 1.01) | 1.00 (0.99 – 1.01) |
| 526.41 | Temporomandibular joint disorder, unspecified | Per88 | 1.00 (0.98 – 1.01) | 6.3E-01 | 1.00 (0.98 – 1.01) | 3.4E-01 | 0.99 (0.96 – 1.02) | 1.00 (0.98 – 1.01) |
| 263 | Other nutritional deficiency | Per88 | 1.00 (0.99 – 1.02) | 6.3E-01 | 1.00 (0.99 – 1.02) | 8.4E-02 | 1.04 (1.02 – 1.07) | 0.98 (0.96 – 1.01) |
| 1004 | Other signs and symptoms involving emotional state | Per88 | 1.00 (0.98 – 1.01) | 6.3E-01 | 1.00 (0.98 – 1.01) | 5.4E-01 | 0.99 (0.97 – 1.01) | 1.00 (0.98 – 1.02) |
| 473.4 | Voice disturbance | Per88 | 1.00 (0.99 – 1.01) | 6.3E-01 | 1.00 (0.99 – 1.01) | 6.2E-01 | 1.00 (0.99 – 1.02) | 1.00 (0.99 – 1.02) |
| 433.31 | Transient cerebral ischemia | Per88 | 1.00 (0.99 – 1.01) | 6.3E-01 | 1.00 (0.99 – 1.01) | 7.4E-01 | 1.00 (0.99 – 1.01) | 1.01 (0.99 – 1.02) |
| 110.12 | Althete's foot | AHI 3% | 1.00 (1.00 – 1.01) | 6.4E-01 | 1.00 (1.00 – 1.01) | 9.8E-01 | 1.00 (0.99 – 1.01) | 1.00 (0.99 – 1.01) |
| 798 | Malaise and fatigue | Per88 | 1.00 (1.00 – 1.00) | 6.4E-01 | 1.00 (0.99 – 1.00) | 3.3E-01 | 1.00 (0.99 – 1.00) | 1.00 (1.00 – 1.01) |
| 979 | Adverse drug events and drug allergies | Per88 | 1.00 (0.99 – 1.01) | 6.4E-01 | 1.00 (0.99 – 1.01) | 8.8E-01 | 1.00 (0.98 – 1.01) | 1.01 (1.00 – 1.02) |
| 596.5 | Functional disorders of bladder | AHI 3% | 1.00 (0.99 – 1.00) | 6.4E-01 | 1.00 (0.99 – 1.00) | 7.5E-01 | 1.00 (0.99 – 1.01) | 1.00 (0.99 – 1.01) |
| 530.11 | GERD | Per88 | 1.00 (1.00 – 1.00) | 6.5E-01 | 1.00 (1.00 – 1.00) | 7.6E-01 | 1.00 (1.00 – 1.01) | 1.00 (1.00 – 1.00) |
| 368.2 | Diplopia and disorders of binocular vision | Per88 | 1.00 (0.98 – 1.01) | 6.5E-01 | 1.00 (0.98 – 1.01) | 4.2E-01 | 1.00 (0.98 – 1.02) | 1.00 (0.98 – 1.02) |
| 208 | Benign neoplasm of colon | Per88 | 1.00 (0.99 – 1.00) | 6.5E-01 | 1.00 (0.99 – 1.00) | 4.2E-01 | 1.00 (0.99 – 1.01) | 0.99 (0.98 – 1.00) |
| 481 | Influenza | AHI 3% | 1.00 (1.00 – 1.00) | 6.5E-01 | 1.00 (1.00 – 1.00) | 9.4E-01 | 1.00 (1.00 – 1.01) | 1.00 (1.00 – 1.01) |
| 472 | Chronic pharyngitis and nasopharyngitis | Per88 | 1.00 (0.99 – 1.01) | 6.5E-01 | 1.00 (0.99 – 1.01) | 9.0E-01 | 1.00 (0.99 – 1.01) | 1.00 (0.99 – 1.01) |
| 723 | Other disorders of cervical region | AHI 3% | 1.00 (0.99 – 1.00) | 6.5E-01 | 1.00 (0.99 – 1.00) | 4.3E-02 | 0.99 (0.98 – 1.01) | 1.00 (0.99 – 1.01) |
| 772.6 | Facial weakness | AHI 3% | 1.00 (0.99 – 1.01) | 6.6E-01 | 1.00 (0.99 – 1.01) | 3.0E-01 | 1.00 (0.99 – 1.01) | 1.00 (0.99 – 1.01) |
| 277.5 | Other disorders of lipoid metabolism | Per88 | 1.00 (0.99 – 1.02) | 6.6E-01 | 1.00 (0.99 – 1.02) | 5.0E-01 | 1.01 (0.98 – 1.03) | 1.00 (0.98 – 1.03) |
| 289.9 | Abnormality of red blood cells | AHI 3% | 1.00 (1.00 – 1.01) | 6.6E-01 | 1.00 (1.00 – 1.01) | 4.9E-02 | 1.01 (1.00 – 1.01) | 1.00 (0.99 – 1.00) |
| 476 | Allergic rhinitis | AHI 3% | 1.00 (1.00 – 1.00) | 6.7E-01 | 1.00 (1.00 – 1.00) | 3.5E-01 | 1.00 (1.00 – 1.00) | 1.00 (1.00 – 1.00) |
| 793.2 | Nonspecific abnormal findings on radiological and other examination of other intrathoracic organs (echocardiogram, etc) | AHI 3% | 1.00 (1.00 – 1.00) | 6.7E-01 | 1.00 (1.00 – 1.00) | 9.9E-01 | 1.00 (0.99 – 1.01) | 1.00 (0.99 – 1.00) |
| 698 | Pruritus and related conditions | AHI 3% | 1.00 (1.00 – 1.00) | 6.7E-01 | 1.00 (1.00 – 1.00) | 8.3E-01 | 1.00 (1.00 – 1.01) | 1.00 (1.00 – 1.00) |
| 300.1 | Anxiety disorder | Per88 | 1.00 (1.00 – 1.00) | 6.7E-01 | 1.00 (1.00 – 1.00) | 5.1E-01 | 1.00 (0.99 – 1.01) | 1.00 (1.00 – 1.01) |
| 727.4 | Ganglion and cyst of synovium, tendon, and bursa | Per88 | 1.00 (0.99 – 1.02) | 6.7E-01 | 1.00 (0.99 – 1.02) | 1.6E-01 | 1.01 (0.99 – 1.04) | 1.00 (0.98 – 1.02) |
| 440.9 | Atherosclerosis of aorta | Per88 | 1.00 (0.98 – 1.03) | 6.7E-01 | 1.00 (0.98 – 1.03) | 1.5E-01 | 0.99 (0.97 – 1.02) | 1.01 (0.98 – 1.05) |
| 374 | Other disorders of eyelids | AHI 3% | 1.00 (1.00 – 1.01) | 6.8E-01 | 1.00 (1.00 – 1.01) | 7.2E-01 | 1.00 (0.99 – 1.01) | 1.00 (1.00 – 1.01) |
| 483 | Acute bronchitis and bronchiolitis | Per88 | 1.00 (0.99 – 1.01) | 6.8E-01 | 1.00 (0.99 – 1.01) | 8.6E-01 | 1.00 (0.98 – 1.01) | 1.01 (0.99 – 1.02) |
| 250.41 | Impaired fasting glucose | Per88 | 1.00 (0.98 – 1.01) | 6.8E-01 | 1.00 (0.99 – 1.01) | 3.9E-01 | 0.99 (0.97 – 1.01) | 1.00 (0.98 – 1.01) |
| 574.3 | Cholecystitis without cholelithiasis | Per88 | 1.00 (0.99 – 1.02) | 6.8E-01 | 1.00 (0.99 – 1.01) | 1.5E-02 | 0.98 (0.96 – 1.00) | 1.02 (1.01 – 1.03) |
| 579.8 | Nonspecific abnormal findings in stool contents | Per88 | 1.00 (0.99 – 1.01) | 6.8E-01 | 1.00 (0.99 – 1.01) | 9.6E-01 | 1.00 (0.98 – 1.01) | 1.00 (0.98 – 1.01) |
| 593.2 | Microscopic hematuria | Per88 | 1.00 (0.99 – 1.01) | 6.8E-01 | 1.00 (0.99 – 1.01) | 6.0E-01 | 1.00 (0.99 – 1.02) | 1.00 (0.98 – 1.01) |

| PheCode | Disease | Sleep statistic | Risk ratio | P-value | Risk ratio (adj pulmonary disorders) | Sleep statistic x sex interaction p-value | Risk ratio (Men) | Risk ratio (Women) |
|---|---|---|---|---|---|---|---|---|
| 454.1 | Varicose veins of lower extremity | Per88 | 1.00 (0.99 – 1.01) | 6.9E-01 | 1.00 (0.99 – 1.01) | 3.1E-01 | 0.99 (0.97 – 1.02) | 1.00 (0.99 – 1.02) |
| 277 | Other disorders of metabolism | Per88 | 1.00 (0.99 – 1.01) | 7.0E-01 | 1.00 (0.99 – 1.01) | 3.7E-03 | 1.01 (1.00 – 1.03) | 0.99 (0.98 – 1.00) |
| 296.1 | Bipolar | Per88 | 1.00 (0.99 – 1.01) | 7.0E-01 | 1.00 (0.99 – 1.01) | 2.0E-02 | 0.98 (0.96 – 1.00) | 1.02 (1.00 – 1.03) |
| 593 | Hematuria | Per88 | 1.00 (0.99 – 1.01) | 7.0E-01 | 1.00 (0.99 – 1.01) | 9.2E-01 | 1.00 (0.99 – 1.01) | 1.00 (0.99 – 1.01) |
| 389.2 | Conductive hearing loss | AHI 3% | 1.00 (0.99 – 1.01) | 7.0E-01 | 1.00 (0.99 – 1.01) | 3.3E-01 | 0.99 (0.98 – 1.01) | 1.00 (0.99 – 1.02) |
| 766 | Neuralgia, neuritis, and radiculitis NOS | Per88 | 1.00 (0.99 – 1.01) | 7.0E-01 | 1.00 (0.99 – 1.01) | 1.0E+00 | 1.00 (0.98 – 1.01) | 1.00 (0.98 – 1.01) |
| 374 | Other disorders of eyelids | Per88 | 1.00 (0.99 – 1.01) | 7.1E-01 | 1.00 (0.99 – 1.01) | 7.5E-01 | 1.00 (0.99 – 1.02) | 1.00 (0.98 – 1.02) |
| 740.1 | Osteoarthritis; localized | Per88 | 1.00 (0.99 – 1.02) | 7.1E-01 | 1.00 (0.99 – 1.02) | 6.5E-01 | 1.00 (0.98 – 1.02) | 1.00 (0.98 – 1.02) |
| 300.8 | Acute reaction to stress | AHI 3% | 1.00 (1.00 – 1.01) | 7.1E-01 | 1.00 (1.00 – 1.01) | 9.5E-01 | 1.00 (0.99 – 1.01) | 1.00 (0.99 – 1.01) |
| 585.4 | Chronic kidney disease, Stage I or II | Per88 | 1.00 (0.99 – 1.02) | 7.1E-01 | 1.00 (0.98 – 1.02) | 6.8E-01 | 1.00 (0.98 – 1.02) | 0.99 (0.96 – 1.03) |
| 599.3 | Dysuria | Per88 | 1.00 (0.99 – 1.01) | 7.2E-01 | 1.00 (0.99 – 1.01) | 3.1E-01 | 0.99 (0.98 – 1.01) | 1.01 (1.00 – 1.02) |
| 586.4 | Stricture/obstruction of ureter | AHI 3% | 1.00 (0.99 – 1.00) | 7.2E-01 | 1.00 (0.99 – 1.00) | NA | 1.00 (0.99 – 1.00) | NA |
| 772.6 | Facial weakness | Per88 | 1.00 (0.98 – 1.02) | 7.2E-01 | 0.99 (0.98 – 1.01) | 3.8E-01 | 1.00 (0.98 – 1.03) | 0.99 (0.96 – 1.02) |
| 727.1 | Synovitis and tenosynovitis | Per88 | 1.00 (0.99 – 1.01) | 7.2E-01 | 1.00 (0.99 – 1.01) | 1.4E-01 | 1.00 (0.98 – 1.02) | 1.00 (0.98 – 1.01) |
| 465.4 | Acute laryngitis and tracheitis | Per88 | 1.00 (0.98 – 1.01) | 7.2E-01 | 0.99 (0.98 – 1.01) | 9.5E-01 | 1.00 (0.97 – 1.02) | 1.00 (0.97 – 1.02) |
| 371.3 | Inflammation of eyelids | AHI 3% | 1.00 (1.00 – 1.00) | 7.2E-01 | 1.00 (1.00 – 1.00) | 1.3E-01 | 1.00 (0.99 – 1.01) | 1.00 (0.99 – 1.00) |
| 526 | Diseases of the jaws | Per88 | 1.00 (0.99 – 1.02) | 7.3E-01 | 1.00 (0.99 – 1.02) | 9.5E-01 | 1.00 (0.97 – 1.03) | 1.00 (0.99 – 1.02) |
| 286.81 | Primary hypercoagulable state | AHI 3% | 1.00 (0.99 – 1.01) | 7.3E-01 | 1.00 (0.99 – 1.01) | 9.9E-01 | 1.00 (0.98 – 1.01) | 1.00 (0.99 – 1.01) |
| 427.61 | Supraventricular premature beats | AHI 3% | 1.00 (0.99 – 1.01) | 7.3E-01 | 1.00 (0.99 – 1.01) | 5.7E-01 | 1.00 (0.99 – 1.01) | 1.00 (0.99 – 1.01) |
| 765 | Cervical radiculitis | Per88 | 1.00 (0.99 – 1.01) | 7.3E-01 | 1.00 (0.99 – 1.01) | 3.0E-01 | 0.99 (0.98 – 1.01) | 1.01 (1.00 – 1.02) |
| 720 | Spinal stenosis | Per88 | 1.00 (0.99 – 1.01) | 7.3E-01 | 1.00 (0.99 – 1.01) | 1.9E-01 | 0.99 (0.97 – 1.00) | 1.01 (0.99 – 1.02) |
| 348.7 | Coma | AHI 3% | 1.00 (0.99 – 1.01) | 7.4E-01 | 1.00 (0.99 – 1.01) | 5.6E-01 | 1.00 (0.99 – 1.01) | 1.00 (0.99 – 1.01) |
| 110.13 | Dermatophytosis of the body | AHI 3% | 1.00 (1.00 – 1.01) | 7.4E-01 | 1.00 (1.00 – 1.01) | 1.3E-01 | 1.00 (0.99 – 1.00) | 1.01 (0.99 – 1.02) |
| 561 | Symptoms involving digestive system | AHI 3% | 1.00 (1.00 – 1.00) | 7.4E-01 | 1.00 (1.00 – 1.00) | 7.4E-01 | 1.00 (1.00 – 1.01) | 1.00 (0.99 – 1.00) |
| 442.1 | Aortic aneurysm | AHI 3% | 1.00 (0.99 – 1.01) | 7.4E-01 | 1.00 (0.99 – 1.01) | 9.5E-02 | 1.00 (0.99 – 1.01) | 1.01 (1.00 – 1.03) |
| 285 | Other anemias | AHI 3% | 1.00 (1.00 – 1.00) | 7.4E-01 | 1.00 (1.00 – 1.00) | 2.2E-04 | 1.00 (1.00 – 1.01) | 1.00 (1.00 – 1.00) |
| 369.5 | Conjunctivitis, infectious | AHI 3% | 1.00 (1.00 – 1.00) | 7.4E-01 | 1.00 (1.00 – 1.00) | 2.7E-01 | 1.00 (1.00 – 1.01) | 1.00 (0.99 – 1.00) |
| 290.1 | Dementias | Per88 | 1.00 (0.98 – 1.02) | 7.4E-01 | 1.00 (0.98 – 1.02) | 7.4E-01 | 1.00 (0.98 – 1.02) | 1.00 (0.98 – 1.04) |
| 530.13 | Barrett's esophagus | AHI 3% | 1.00 (0.99 – 1.01) | 7.6E-01 | 1.00 (0.99 – 1.01) | 7.2E-01 | 1.00 (0.99 – 1.01) | 1.00 (0.99 – 1.01) |
| 296.2 | Depression | Per88 | 1.00 (0.99 – 1.00) | 7.6E-01 | 1.00 (0.99 – 1.00) | 9.8E-01 | 1.00 (0.99 – 1.01) | 1.00 (0.99 – 1.01) |
| 686.1 | Carbuncle and furuncle | AHI 3% | 1.00 (0.99 – 1.01) | 7.6E-01 | 1.00 (0.99 – 1.01) | 2.9E-01 | 0.99 (0.98 – 1.00) | 1.01 (1.00 – 1.02) |
| 458.1 | Orthostatic hypotension | Per88 | 1.00 (0.98 – 1.01) | 7.6E-01 | 1.00 (0.98 – 1.01) | 2.2E-01 | 1.00 (0.98 – 1.02) | 1.00 (0.98 – 1.02) |
| 426 | Cardiac conduction disorders | AHI 3% | 1.00 (0.99 – 1.01) | 7.7E-01 | 1.00 (0.99 – 1.01) | 2.9E-01 | 1.00 (0.99 – 1.01) | 1.00 (0.99 – 1.02) |
| 418 | Nonspecific chest pain | AHI 3% | 1.00 (1.00 – 1.00) | 7.7E-01 | 1.00 (1.00 – 1.00) | 9.9E-01 | 1.00 (1.00 – 1.00) | 1.00 (1.00 – 1.00) |
| 565.1 | Anal and rectal polyp | Per88 | 1.00 (0.99 – 1.01) | 7.7E-01 | 1.00 (0.99 – 1.01) | 3.5E-01 | 1.00 (0.98 – 1.01) | 1.00 (0.98 – 1.01) |

| PheCode | Disease | Sleep statistic | Risk ratio | P-value | Risk ratio (adj pulmonary disorders) | Sleep statistic x sex interaction p-value | Risk ratio (Men) | Risk ratio (Women) |
|---|---|---|---|---|---|---|---|---|
| 367.1 | Myopia | Per88 | 1.00 (0.99 – 1.01) | 7.7E-01 | 1.00 (0.99 – 1.01) | 9.8E-01 | 1.00 (0.98 – 1.02) | 1.00 (0.98 – 1.01) |
| 721.8 | Other allied disorders of spine | Per88 | 1.00 (0.98 – 1.01) | 7.7E-01 | 1.00 (0.99 – 1.01) | 4.0E-01 | 1.00 (0.98 – 1.02) | 0.99 (0.97 – 1.01) |
| 599.8 | Other symptoms involving urinary system | AHI 3% | 1.00 (0.99 – 1.00) | 7.8E-01 | 1.00 (0.99 – 1.00) | 6.4E-03 | 1.00 (1.00 – 1.01) | 1.00 (0.99 – 1.00) |
| 110.2 | Dermatomycoses | AHI 3% | 1.00 (0.99 – 1.01) | 7.8E-01 | 1.00 (0.99 – 1.01) | 9.5E-02 | 1.00 (0.99 – 1.00) | 1.01 (1.00 – 1.02) |
| 274.1 | Gout | Per88 | 1.00 (0.99 – 1.01) | 7.8E-01 | 1.00 (0.99 – 1.01) | 3.6E-01 | 0.99 (0.98 – 1.01) | 1.02 (1.00 – 1.04) |
| 465 | Acute upper respiratory infections of multiple or unspecified sites | Per88 | 1.00 (0.99 – 1.01) | 7.8E-01 | 1.00 (0.99 – 1.00) | 3.2E-01 | 1.00 (0.99 – 1.01) | 1.00 (0.99 – 1.01) |
| 433.8 | Late effects of cerebrovascular disease | Per88 | 1.00 (0.98 – 1.02) | 7.8E-01 | 1.00 (0.98 – 1.02) | 3.6E-01 | 1.00 (0.98 – 1.02) | 1.01 (0.98 – 1.04) |
| 411.1 | Unstable angina (intermediate coronary syndrome) | AHI 3% | 1.00 (0.99 – 1.01) | 7.9E-01 | 1.00 (0.99 – 1.01) | 3.3E-01 | 1.00 (0.99 – 1.00) | 1.01 (1.00 – 1.02) |
| 592.11 | Acute cystitis | Per88 | 1.00 (0.99 – 1.02) | 7.9E-01 | 1.00 (0.99 – 1.02) | 2.5E-01 | 0.99 (0.96 – 1.01) | 1.01 (0.99 – 1.03) |
| 470 | Septal Deviations/Turbinate Hypertrophy | Per88 | 1.00 (0.99 – 1.01) | 8.0E-01 | 1.00 (0.99 – 1.00) | 4.0E-01 | 1.00 (0.99 – 1.01) | 1.00 (0.99 – 1.01) |
| 594.8 | Renal colic | AHI 3% | 1.00 (0.99 – 1.01) | 8.0E-01 | 1.00 (0.99 – 1.01) | 4.1E-01 | 1.00 (0.99 – 1.01) | 1.01 (0.99 – 1.02) |
| 761 | Cervicalgia | AHI 3% | 1.00 (1.00 – 1.00) | 8.0E-01 | 1.00 (1.00 – 1.00) | 3.2E-01 | 1.00 (1.00 – 1.00) | 1.00 (0.99 – 1.00) |
| 300.13 | Phobia | Per88 | 1.00 (0.99 – 1.01) | 8.0E-01 | 1.00 (0.99 – 1.01) | 2.1E-01 | 1.01 (0.99 – 1.04) | 0.99 (0.98 – 1.01) |
| 272.1 | Hyperlipidemia | Per88 | 1.00 (1.00 – 1.00) | 8.1E-01 | 1.00 (1.00 – 1.00) | 1.1E-01 | 1.00 (0.99 – 1.00) | 1.00 (1.00 – 1.01) |
| 574.1 | Cholelithiasis | AHI 3% | 1.00 (1.00 – 1.00) | 8.1E-01 | 1.00 (1.00 – 1.00) | 2.5E-01 | 1.00 (1.00 – 1.01) | 1.00 (0.99 – 1.00) |
| 440 | Atherosclerosis | Per88 | 1.00 (0.98 – 1.02) | 8.2E-01 | 1.00 (0.98 – 1.02) | 1.9E-01 | 0.98 (0.95 – 1.01) | 1.03 (1.01 – 1.06) |
| 241.1 | Nontoxic uninodular goiter | AHI 3% | 1.00 (1.00 – 1.00) | 8.2E-01 | 1.00 (1.00 – 1.00) | 1.9E-01 | 1.00 (0.99 – 1.01) | 1.00 (0.99 – 1.00) |
| 418.1 | Precordial pain | AHI 3% | 1.00 (1.00 – 1.01) | 8.2E-01 | 1.00 (1.00 – 1.01) | 8.3E-01 | 1.00 (0.99 – 1.01) | 1.00 (0.99 – 1.01) |
| 440.9 | Atherosclerosis of aorta | AHI 3% | 1.00 (0.99 – 1.01) | 8.2E-01 | 1.00 (0.99 – 1.01) | 4.5E-01 | 1.00 (0.99 – 1.01) | 1.01 (1.00 – 1.02) |
| 427.5 | Arrhythmia (cardiac) NOS | AHI 3% | 1.00 (1.00 – 1.00) | 8.3E-01 | 1.00 (1.00 – 1.00) | 4.4E-01 | 1.00 (1.00 – 1.00) | 1.00 (1.00 – 1.00) |
| 411.1 | Unstable angina (intermediate coronary syndrome) | Per88 | 1.00 (0.98 – 1.02) | 8.3E-01 | 1.00 (0.98 – 1.02) | 4.5E-01 | 1.00 (0.97 – 1.02) | 1.01 (0.98 – 1.05) |
| 350.3 | Lack of coordination | AHI 3% | 1.00 (0.99 – 1.01) | 8.3E-01 | 1.00 (0.99 – 1.01) | 5.5E-01 | 1.00 (0.99 – 1.01) | 0.99 (0.98 – 1.01) |
| 819 | Skull and face fracture and other intercranial injury | Per88 | 1.00 (0.99 – 1.02) | 8.4E-01 | 1.00 (0.98 – 1.01) | 1.2E-01 | 1.01 (0.99 – 1.03) | 0.99 (0.96 – 1.01) |
| 596 | Other disorders of bladder | AHI 3% | 1.00 (0.99 – 1.01) | 8.4E-01 | 1.00 (0.99 – 1.01) | 8.8E-01 | 1.00 (0.99 – 1.01) | 1.00 (0.99 – 1.01) |
| 411.4 | Coronary atherosclerosis | AHI 3% | 1.00 (1.00 – 1.00) | 8.5E-01 | 1.00 (1.00 – 1.00) | 2.6E-01 | 1.00 (1.00 – 1.00) | 1.00 (1.00 – 1.01) |
| 350.2 | Abnormality of gait | AHI 3% | 1.00 (1.00 – 1.00) | 8.5E-01 | 1.00 (1.00 – 1.00) | 1.4E-01 | 1.00 (1.00 – 1.01) | 1.00 (0.99 – 1.00) |
| 427.8 | Sinoatrial node dysfunction (Bradycardia) | Per88 | 1.00 (0.98 – 1.02) | 8.5E-01 | 1.00 (0.98 – 1.02) | 6.4E-02 | 0.99 (0.97 – 1.01) | 1.03 (1.00 – 1.06) |
| 694.2 | Other dyschromia | AHI 3% | 1.00 (1.00 – 1.00) | 8.5E-01 | 1.00 (1.00 – 1.00) | 5.8E-01 | 1.00 (1.00 – 1.00) | 1.00 (1.00 – 1.00) |
| 380.4 | Impacted cerumen | Per88 | 1.00 (0.99 – 1.01) | 8.6E-01 | 1.00 (0.99 – 1.01) | 7.3E-01 | 1.00 (0.99 – 1.01) | 1.00 (0.99 – 1.02) |
| 561.1 | Diarrhea | AHI 3% | 1.00 (1.00 – 1.00) | 8.6E-01 | 1.00 (1.00 – 1.00) | 6.3E-01 | 1.00 (1.00 – 1.00) | 1.00 (0.99 – 1.00) |
| 495 | Asthma | Per88 | 1.00 (1.00 – 1.00) | 8.6E-01 | 0.99 (0.98 – 0.99) | 3.5E-01 | 1.00 (0.99 – 1.00) | 1.00 (1.00 – 1.01) |
| 475.9 | Postnasal drip | AHI 3% | 1.00 (1.00 – 1.00) | 8.7E-01 | 1.00 (1.00 – 1.00) | 7.6E-01 | 1.00 (0.99 – 1.00) | 1.00 (1.00 – 1.01) |

| PheCode | Disease | Sleep statistic | Risk ratio | P-value | Risk ratio (adj pulmonary disorders) | Sleep statistic x sex interaction p-value | Risk ratio (Men) | Risk ratio (Women) |
|---|---|---|---|---|---|---|---|---|
| 290.1 | Dementias | AHI 3% | 1.00 (0.99 – 1.01) | 8.7E-01 | 1.00 (0.99 – 1.01) | 8.4E-01 | 1.00 (1.00 – 1.01) | 1.00 (0.99 – 1.01) |
| 474.2 | Chronic tonsillitis and adenoiditis | Per88 | 1.00 (0.99 – 1.01) | 8.8E-01 | 1.00 (0.99 – 1.01) | 1.8E-01 | 0.99 (0.97 – 1.01) | 1.01 (1.00 – 1.02) |
| 427.6 | Premature beats | AHI 3% | 1.00 (1.00 – 1.00) | 8.8E-01 | 1.00 (1.00 – 1.00) | 2.1E-01 | 1.00 (1.00 – 1.01) | 1.00 (0.99 – 1.01) |
| 721.8 | Other allied disorders of spine | AHI 3% | 1.00 (0.99 – 1.01) | 8.9E-01 | 1.00 (0.99 – 1.01) | 5.9E-01 | 1.00 (0.99 – 1.01) | 1.00 (0.99 – 1.01) |
| 599.2 | Retention of urine | Per88 | 1.00 (0.99 – 1.01) | 8.9E-01 | 1.00 (0.99 – 1.01) | 6.8E-01 | 1.00 (0.99 – 1.01) | 1.01 (0.99 – 1.03) |
| 300.11 | Generalized anxiety disorder | Per88 | 1.00 (0.99 – 1.01) | 8.9E-01 | 1.00 (0.99 – 1.01) | 1.8E-01 | 1.00 (0.99 – 1.01) | 1.00 (1.00 – 1.01) |
| 772.2 | Spasm of muscle | Per88 | 1.00 (0.99 – 1.01) | 8.9E-01 | 1.00 (0.99 – 1.01) | 9.6E-02 | 1.00 (0.99 – 1.02) | 1.00 (0.98 – 1.01) |
| 277.5 | Other disorders of lipoid metabolism | AHI 3% | 1.00 (0.99 – 1.01) | 9.0E-01 | 1.00 (0.99 – 1.01) | 7.5E-03 | 1.01 (1.00 – 1.01) | 0.99 (0.98 – 1.00) |
| 772.3 | Muscle weakness | Per88 | 1.00 (0.99 – 1.01) | 9.0E-01 | 1.00 (0.99 – 1.01) | 2.2E-01 | 1.00 (0.98 – 1.02) | 0.99 (0.98 – 1.01) |
| 912 | Insect bite | Per88 | 1.00 (0.99 – 1.01) | 9.1E-01 | 1.00 (0.99 – 1.01) | 8.6E-01 | 1.00 (0.97 – 1.03) | 0.99 (0.97 – 1.01) |
| 427.8 | Sinoatrial node dysfunction (Bradycardia) | AHI 3% | 1.00 (0.99 – 1.01) | 9.1E-01 | 1.00 (0.99 – 1.01) | 5.4E-01 | 1.00 (0.99 – 1.01) | 1.00 (0.99 – 1.02) |
| 599.3 | Dysuria | AHI 3% | 1.00 (1.00 – 1.00) | 9.1E-01 | 1.00 (1.00 – 1.00) | 7.4E-03 | 1.00 (1.00 – 1.01) | 1.00 (0.99 – 1.00) |
| 764 | Sciatica | Per88 | 1.00 (0.99 – 1.01) | 9.1E-01 | 1.00 (0.99 – 1.01) | 4.3E-01 | 1.00 (0.99 – 1.01) | 1.00 (0.99 – 1.01) |
| 721.1 | Spondylosis without myelopathy | Per88 | 1.00 (0.99 – 1.01) | 9.1E-01 | 1.00 (0.99 – 1.01) | 6.8E-01 | 1.00 (0.99 – 1.01) | 1.00 (0.99 – 1.01) |
| 245.21 | Chronic lymphocytic thyroiditis | AHI 3% | 1.00 (0.99 – 1.00) | 9.2E-01 | 1.00 (0.99 – 1.00) | 6.6E-01 | 0.99 (0.98 – 1.00) | 1.00 (0.99 – 1.01) |
| 389.1 | Sensorineural hearing loss | AHI 3% | 1.00 (1.00 – 1.00) | 9.2E-01 | 1.00 (1.00 – 1.00) | 3.3E-01 | 1.00 (1.00 – 1.00) | 1.00 (1.00 – 1.01) |
| 764 | Sciatica | AHI 3% | 1.00 (1.00 – 1.00) | 9.2E-01 | 1.00 (1.00 – 1.00) | 1.5E-01 | 1.00 (1.00 – 1.01) | 1.00 (0.99 – 1.00) |
| 008.6 | Viral Enteritis | Per88 | 1.00 (0.99 – 1.01) | 9.3E-01 | 1.00 (0.99 – 1.01) | 9.0E-01 | 1.01 (0.99 – 1.04) | 0.99 (0.98 – 1.01) |
| 735.3 | Hallux valgus (Bunion) | Per88 | 1.00 (0.99 – 1.01) | 9.3E-01 | 1.00 (0.99 – 1.01) | 8.3E-01 | 0.99 (0.97 – 1.02) | 1.00 (0.98 – 1.01) |
| 208 | Benign neoplasm of colon | AHI 3% | 1.00 (1.00 – 1.00) | 9.4E-01 | 1.00 (1.00 – 1.00) | 3.3E-01 | 1.00 (1.00 – 1.00) | 1.00 (1.00 – 1.00) |
| 300.3 | Obsessive-compulsive disorders | Per88 | 1.00 (0.99 – 1.01) | 9.4E-01 | 1.00 (0.99 – 1.01) | 1.8E-01 | 1.00 (0.98 – 1.01) | 1.01 (0.99 – 1.02) |
| 244.2 | Acquired hypothyroidism | AHI 3% | 1.00 (1.00 – 1.00) | 9.4E-01 | 1.00 (1.00 – 1.00) | 3.7E-01 | 0.99 (0.98 – 1.00) | 1.00 (1.00 – 1.01) |
| 296.22 | Major depressive disorder | Per88 | 1.00 (1.00 – 1.00) | 9.5E-01 | 1.00 (1.00 – 1.00) | 1.0E-01 | 1.00 (0.99 – 1.00) | 1.00 (1.00 – 1.01) |
| 415.11 | Pulmonary embolism and infarction, acute | AHI 3% | 1.00 (0.99 – 1.00) | 9.5E-01 | 1.00 (0.99 – 1.00) | 4.2E-01 | 1.00 (0.99 – 1.01) | 1.00 (0.99 – 1.01) |
| 459.9 | Circulatory disease NEC | AHI 3% | 1.00 (1.00 – 1.00) | 9.5E-01 | 1.00 (1.00 – 1.00) | 8.3E-01 | 1.00 (1.00 – 1.00) | 1.00 (0.99 – 1.01) |
| 293 | Symptoms involving head and neck | Per88 | 1.00 (0.98 – 1.02) | 9.5E-01 | 1.00 (0.98 – 1.02) | 5.7E-01 | 0.99 (0.97 – 1.02) | 1.00 (0.98 – 1.03) |
| 415 | Pulmonary heart disease | AHI 3% | 1.00 (0.99 – 1.01) | 9.5E-01 | 1.00 (0.99 – 1.01) | 4.3E-03 | 1.01 (1.00 – 1.02) | 0.99 (0.98 – 1.00) |
| 008.6 | Viral Enteritis | AHI 3% | 1.00 (0.99 – 1.01) | 9.5E-01 | 1.00 (0.99 – 1.01) | 8.2E-01 | 1.00 (0.99 – 1.01) | 1.00 (0.99 – 1.01) |
| 516.1 | Hemoptysis | AHI 3% | 1.00 (0.99 – 1.01) | 9.5E-01 | 1.00 (0.99 – 1.01) | 4.7E-01 | 1.00 (0.99 – 1.01) | 1.00 (0.99 – 1.01) |
| 532 | Dysphagia | AHI 3% | 1.00 (1.00 – 1.00) | 9.5E-01 | 1.00 (1.00 – 1.00) | 8.1E-01 | 1.00 (1.00 – 1.01) | 1.00 (0.99 – 1.00) |
| 172.22 | Squamous cell carcinoma | Per88 | 1.00 (0.98 – 1.02) | 9.5E-01 | 1.00 (0.98 – 1.01) | 2.6E-01 | 0.99 (0.98 – 1.01) | 1.02 (1.00 – 1.05) |
| 172.2 | Other non-epithelial cancer of skin | AHI 3% | 1.00 (1.00 – 1.00) | 9.6E-01 | 1.00 (1.00 – 1.00) | 3.8E-01 | 1.00 (1.00 – 1.01) | 1.00 (0.99 – 1.00) |
| 912 | Insect bite | AHI 3% | 1.00 (0.99 – 1.01) | 9.6E-01 | 1.00 (0.99 – 1.01) | 9.5E-01 | 1.00 (0.99 – 1.01) | 1.00 (0.99 – 1.01) |
| 345 | Epilepsy, recurrent seizures, convulsions | Per88 | 1.00 (0.99 – 1.01) | 9.6E-01 | 1.00 (0.99 – 1.01) | 8.1E-02 | 1.00 (0.98 – 1.01) | 1.00 (0.99 – 1.02) |
| 367.8 | Hypermetropia | Per88 | 1.00 (0.99 – 1.01) | 9.6E-01 | 1.00 (0.99 – 1.01) | 7.0E-01 | 0.99 (0.97 – 1.02) | 1.01 (0.99 – 1.03) |

| PheCode | Disease | Sleep statistic | Risk ratio | P-value | Risk ratio (adj pulmonary disorders) | Sleep statistic x sex interaction p-value | Risk ratio (Men) | Risk ratio (Women) |
|---|---|---|---|---|---|---|---|---|
| 596.5 | Functional disorders of bladder | Per88 | 1.00 (0.98 – 1.01) | 9.6E-01 | 1.00 (0.98 – 1.01) | 8.2E-01 | 1.01 (0.99 – 1.03) | 0.99 (0.97 – 1.01) |
| 965 | Poisoning by analgesics, antipyretics, and antirheumatics | AHI 3% | 1.00 (1.00 – 1.00) | 9.6E-01 | 1.00 (1.00 – 1.00) | 3.7E-01 | 1.00 (1.00 – 1.01) | 1.00 (0.99 – 1.00) |
| 724.8 | Other symptoms referable to back | Per88 | 1.00 (0.98 – 1.02) | 9.6E-01 | 1.00 (0.98 – 1.02) | 8.3E-01 | 0.99 (0.96 – 1.02) | 1.01 (0.99 – 1.03) |
| 514.2 | Solitary pulmonary nodule | AHI 3% | 1.00 (1.00 – 1.00) | 9.6E-01 | 1.00 (1.00 – 1.00) | 3.5E-01 | 1.00 (0.99 – 1.00) | 1.00 (0.99 – 1.01) |
| 698 | Pruritus and related conditions | Per88 | 1.00 (0.99 – 1.01) | 9.7E-01 | 1.00 (0.99 – 1.01) | 7.9E-01 | 0.99 (0.98 – 1.01) | 1.00 (1.00 – 1.01) |
| 464 | Acute sinusitis | AHI 3% | 1.00 (1.00 – 1.00) | 9.7E-01 | 1.00 (1.00 – 1.00) | 3.6E-01 | 1.00 (0.99 – 1.01) | 1.00 (0.99 – 1.00) |
| 276.12 | Hyposmolality and/or hyponatremia | AHI 3% | 1.00 (0.99 – 1.01) | 9.7E-01 | 1.00 (0.99 – 1.01) | 3.5E-01 | 1.00 (0.99 – 1.00) | 1.00 (0.99 – 1.01) |
| 704 | Diseases of hair and hair follicles | AHI 3% | 1.00 (1.00 – 1.00) | 9.8E-01 | 1.00 (1.00 – 1.00) | 6.4E-03 | 1.00 (1.00 – 1.01) | 0.99 (0.99 – 1.00) |
| 259 | Other endocrine disorders | Per88 | 1.00 (0.98 – 1.02) | 9.8E-01 | 1.00 (0.98 – 1.02) | 5.3E-02 | 1.02 (1.00 – 1.05) | 0.97 (0.95 – 1.00) |
| 292.1 | Aphasia/speech disturbance | AHI 3% | 1.00 (0.99 – 1.01) | 9.8E-01 | 1.00 (0.99 – 1.01) | 1.5E-01 | 1.00 (0.99 – 1.01) | 1.00 (0.99 – 1.01) |
| 375.1 | Dry eyes | Per88 | 1.00 (0.99 – 1.01) | 9.8E-01 | 1.00 (0.99 – 1.01) | 4.0E-01 | 1.00 (0.99 – 1.02) | 0.99 (0.98 – 1.01) |
| 563 | Constipation | Per88 | 1.00 (0.99 – 1.01) | 9.9E-01 | 1.00 (0.99 – 1.00) | 5.1E-01 | 1.00 (0.99 – 1.01) | 1.00 (1.00 – 1.01) |
| 427.22 | Atrial flutter | AHI 3% | 1.00 (0.99 – 1.01) | 9.9E-01 | 1.00 (0.99 – 1.01) | 6.0E-01 | 1.00 (1.00 – 1.01) | 1.00 (0.99 – 1.01) |
| 386.2 | Peripheral or central vertigo | AHI 3% | 1.00 (1.00 – 1.00) | 9.9E-01 | 1.00 (1.00 – 1.00) | 6.1E-01 | 1.00 (0.99 – 1.01) | 1.00 (0.99 – 1.01) |
| 300.3 | Obsessive-compulsive disorders | AHI 3% | 1.00 (0.99 – 1.01) | 9.9E-01 | 1.00 (0.99 – 1.01) | 9.0E-01 | 1.00 (0.99 – 1.01) | 1.00 (0.99 – 1.01) |
